# The prevalence of cardiovascular disease risk factors among adults living in extreme poverty: a cross-sectional analysis of 105 nationally representative surveys with 33 million participants

**DOI:** 10.1101/2022.10.08.22280861

**Authors:** Pascal Geldsetzer, Rebecca L. Tisdale, Lisa Stehr, Felix Michalik, Julia Lemp, Krishna K. Aryal, Albertino Damasceno, Corine Houehanou, Jutta Mari Adelin Jørgensen, Nuno Lunet, Mary Mayige, Sahar Saeedi Moghaddam, Joseph Kibachio Mwangi, Christian Bommer, Maja-Emilia Marcus, Michaela Theilmann, Rifat Atun, Justine Ina Davies, David Flood, Jennifer Manne-Goehler, Jacqueline Seiglie, Till Bärnighausen, Sebastian Vollmer

**Affiliations:** Department of Medicine, Division of Primary Care and Population Health, Stanford University School of Medicine; Stanford, CA, USA; Chan Zuckerberg Biohub; San Francisco, CA, USA; Veterans Affairs Palo Alto Health Care System; Palo Alto, CA, USA; Department of Health Policy, Stanford University School of Medicine, and Center for Health Policy, Freeman Spogli Institute for International Studies, Stanford University; Stanford, CA, USA; Heidelberg Institute of Global Health, Faculty of Medicine and University Hospital, Heidelberg University, Heidelberg, Germany; Department for International Development/Nepal Health Sector Programme 3/Monitoring Evaluation and Operational Research, Abt Associates, Kathmandu, Nepal; Department of Public and Forensic Health Sciences and Medical Education, Faculty of Medicine, University of Porto, Porto, Portugal; Laboratory of Epidemiology of Chronic and Neurological Diseases, Faculty of Health Sciences, University of Abomey-Calavi, Cotonou, Benin; Institute of Global Health, Dept of Public Health and Epidemiology, Copenhagen University, Denmark; National Institute for Medical Research, Dar es Salaam, Tanzania; Non-Communicable Diseases Research Center, Endocrinology and Metabolism Population Sciences Institute, Tehran University of Medical Sciences, Tehran, Iran; Division of Non-Communicable Diseases, Ministry of Health, Nairobi, Kenya; Centre for Modern Indian Studies, University of Goettingen, Göttingen, Germany; Department of Economics, University of Goettingen, Göttingen, Germany; Department of Global Health and Population, Harvard T. H. Chan School of Public Health, Boston, MA, USA; Department of Global Health and Social Medicine, Harvard Medical School, Harvard University, Boston, MA, USA; Institute of Applied Health Research, University of Birmingham, Birmingham, United Kingdom; MRC/Wits Rural Public Health and Health Transitions Research Unit, School of Public Health, University of the Witwatersrand, Johannesburg, South Africa; Centre for Global Surgery, Department of Global Health, Stellenbosch University, Cape Town, South Africa; University of Michigan, Ann Arbor, MI, USA; Division of Infectious Diseases, Brigham and Women’s Hospital, Boston, MA, USA; Medical Practice Evaluation Center, Massachusetts General Hospital, Harvard Medical School, Boston, MA, USA; Diabetes Unit, Massachusetts General Hospital, Boston, MA, USA; Department of Medicine, Harvard Medical School, Boston, MA, USA; Harvard Center for Population and Development Studies, Cambridge, MA, USA

## Abstract

**Background:** Historically, the international development community has often held the view that those living in extreme poverty (at less than $1.90/day) are likely to have a low prevalence of cardiovascular disease (CVD) risk factors due to calorie scarcity, a largely plant-based diet, and physical labor. Evidence on CVD risk factor prevalence among adults living below the World Bank’s international line for extreme poverty globally is sparse because studies have used measures of socioeconomic status that are not comparable across study populations and countries. For adults living in extreme poverty, this study aimed to determine i) the prevalence of each of five key CVD risk factors, ii) how the prevalence of these CVD risk factors varies across and within countries, and iii) the level of treatment coverage with statin, antihypertensive, and diabetes therapy.

**Methods:** We pooled individual-level data from 105 nationally representative household surveys with physical measurements of hypertension, diabetes, overweight, obesity, and dyslipidemia, as well as self-reported smoking status, from 78 countries that represent 85% of people living in extreme poverty globally. Those living in extreme poverty were defined by ordering participants according to a continuous household wealth index or household income value, and then applying World Bank estimates on the proportion of each country’s population that is living in extreme poverty. We used modified Poisson regression models to examine how CVD risk factor prevalence among those in extreme poverty varied by age, sex, educational attainment, and rural versus urban residency. We also calculated the proportion of participants with diabetes or hypertension who self-reported the use of blood pressure- or blood glucose-lowering medication, respectively; the proportion with hypertension who achieved blood pressure control; and the proportion recommended for statin use according to WHO guidelines who reported taking a statin.

**Results:** Of 32,695,579 participants, 7,922,289 were living in extreme poverty (<$1.90 per day), 15,986,099 on less than $3.20/day, and 23,466,879 on less than $5.50/day. Among those living in extreme poverty (<$1.90 per day), the age-standardized prevalence was 17.5% (95% CI: 16.7% – 18.3%) for hypertension, 4.0% (95% CI: 3.6% – 4.5%) for diabetes, 10.6% (95% CI: 9.0% – 12.3%) for current smoking, 3.1% (95% CI: 2.8% – 3.3%) for obesity, and 1.4% (95% CI: 0.9% – 1.9%) for dyslipidemia. In most countries in our analysis, the prevalence of these risk factors among those living in extreme poverty was not notably lower than in the total population. Hypertension treatment and control, diabetes treatment, and statin use were low across all poverty levels in low-income countries, while in lower and upper middle-income countries individuals living at more extreme levels of poverty had a lower probability of reporting the use of these medications and achieving hypertension control.

**Conclusions:** We found a high prevalence of CVD risk factors among adults living below the World Bank’s international line for extreme poverty, along with low statin use and low treatment rates for diabetes and hypertension. Our detailed analysis and comparison across poverty levels could inform equity discussions for resource allocation and the design of effective interventions.

## Background

The prevalence of cardiovascular disease (CVD) risk factors among those living in extreme poverty in low- and middle-income countries (LMICs) has often been assumed to be low ^1,2^. Historically, this subpopulation has been thought to take in fewer calories and have correspondingly lower body mass index ^3–7^, consume a largely plant-based diet ^3,7–11^ and earn income through occupations associated with higher physical activity ^3,6,12,13^. All of these lifestyle patterns decrease the risk of developing CVD and its risk factors ^14^.

Understanding the extent to which this assumption of a low prevalence of CVD risk factors among those in extreme poverty holds true is important for setting priorities within health policy and care delivery, both for equity and effectiveness. From an equity perspective, if CVD risk factors mostly affect wealthier population groups in LMICs, then investing in programs aimed at preventing and treating CVD instead of focusing on those conditions that disproportionately affect those in extreme poverty could further worsen health inequalities by wealth. Regarding effectiveness, understanding the prevalence of CVD risk factors in different population groups is essential for the tailoring of appropriate interventions. For instance, if the prevalence of treatable CVD risk factors, such as diabetes and hypertension, is high among those living in extreme poverty but their engagement with the healthcare system for these conditions is low, then programs that identify ways to engage this population in screening and long-term care for these conditions will be needed.

Despite the importance of this information for guiding health policy and healthcare delivery, to date little data from nationally representative surveys on CVD risk factor prevalence among adults living in extreme poverty exist ^15,16^. The few studies that do exist employ one of two types of measures of poverty. A subset of the literature relies on relative measures of poverty, particularly household wealth quintiles ^15,16^ or tertiles ^17,18^. These measures are specific to the population in which they are calculated because they are relative categories (i.e., the absolute difference in the degree of poverty between a given set of household wealth quintiles will vary across populations) and based on dwelling characteristics and household ownership of durable goods that are inherently context-specific ^19^. They, thus, have limited value in informing CVD risk factor prevalence among those living below an absolute threshold of poverty, such as the World Bank’s international poverty line for extreme poverty ^20^. A second body of literature uses educational attainment as a measure of poverty ^15–17,21–23^. This approach has two critical limitations. First and foremost, education is an imperfect proxy for poverty. For instance, a given individual’s education is unlikely to change later in life ^24^, yet that individual’s economic status could change significantly over the decades after leaving school. Second, quality of education varies widely across and within countries in LMICs ^25,26^, such that years of schooling or a given school degree hold little information outside of specific settings.

This study overcomes these limitations of sparse nationally representative data and poverty measures that are not comparable across settings. We have brought together and harmonized 105 nationally representative surveys with (except for smoking) physical measurements on five major CVD risk factors (diabetes, dyslipidemia, hypertension, obesity, and smoking) from 78 LMICs. This is by far the largest and most comprehensive analysis of the relationship between poverty and CVD risk factors to date. The countries included in our dataset are estimated to be home to 85% of individuals living in extreme poverty worldwide ^27,28^, and represent 53% of the global population as well as 64% of the global population living in LMICs ^28^. By harnessing the national representativeness of our data, we employ a consistent definition of extreme poverty across countries such that we are, for the first time, able to provide estimates of CVD risk factor prevalence among adults living in extreme poverty globally. Specifically, for adults living in extreme poverty in our study countries, this study aimed to determine i) the prevalence of each of five key CVD risk factors (and how these prevalence estimates compare to population groups with higher levels of income), ii) how the prevalence of these CVD risk factors varies across and within countries, and iii) the level of treatment coverage with statin, antihypertensive, and diabetes therapy.

## Methods

### Data sources

To identify relevant surveys, we first searched the Stepwise Approach to Surveillance (STEPS) database of the World Health Organization (WHO) and systematically searched for surveys conducted by the Demographic and Health Surveys (DHS) program and the WHO Global Adult Tobacco Survey (GATS) initiative (Supplementary Materials and Methods 1). STEPS, DHS, and GATS surveys each utilize standardized questionnaires. Our search was limited to countries designated LMICs at the time of data collection based on the World Bank income classification ^29^. For inclusion, we required that surveys: (1) be conducted during or after 2005; (2) be nationally representative with a response rate >50%; (3) provide individual-level data; (4) measured at least one of the following risk factors using the measure in parentheses: hypertension (blood pressure), diabetes (fasting blood glucose or HbA1c), overweight/obesity (height, weight), smoking (self-report via questionnaire), or dyslipidemia (total or LDL cholesterol); and (5) include a measurement of household income (self-reported household income or a household wealth index based on questionnaire responses to a household’s ownership of selected assets, materials used for housing construction, and types of water access and sanitation facilities) (Supplementary Materials and Methods 1). If an eligible STEPS, DHS, or GATS dataset was not available for an LMIC, or we could not gain access to the individual-level data, we conducted a systematic search with Google to identify eligible surveys for the given country (Supplementary Materials and Methods 1).

Our search returned 54 STEPS surveys, 103 DHS surveys, 30 GATS surveys, and 9 surveys via the Google search that met our inclusion criteria. When more than one survey measured a CVD risk factor in a given country, we used the survey that was conducted more recently for that CVD risk factor. If survey collection years were identical, we used the survey that sampled a wider age range of participants. Through this process (detailed in Figures S1-S4), we arrived at a total of 105 surveys for our analysis, of which 36 were STEPS surveys, 51 DHS surveys, 12 GATS surveys and six other types of surveys.

The sampling strategy used by each survey included in our analysis is detailed in Supplementary Materials and Methods 2.

### Definition of poverty

The World Bank publishes annual data on the estimated proportion of the population in each country that is living below each of three poverty lines, defined by an income (all in 2011 purchasing-power-parity [PPP] adjusted dollars) of $1.90 per day (the “international poverty line”), $3.20 per day, and $5.50 per day ^20^. Our analysis focuses on those living in extreme poverty, which is generally defined by the international poverty line of $1.90 per day ^20^. However, to enable comparison with other income groups in LMICs, we also examine the prevalence of CVD risk factors for the population with an income below each $3.20 and $5.50 per day, as well as those with an income greater than $5.50 per day. For simplicity, we refer to these categorizations as “poverty levels”.

Sixty-seven surveys included a continuous household wealth index. This index was computed from a principal component analysis of participants’ answers to questions on key household dwelling characteristics and household ownership of durable goods (i.e., goods that are not completely consumed in one use). Most STEPS surveys instead asked participants for their household’s income, allowing participants to provide the income as a yearly, monthly, or weekly figure.

Given that the surveys included in our analysis were all nationally representative, we used the distribution of household wealth or income in each survey dataset to define which participants fell below each poverty level. Specifically, we first ordered participants in each dataset from “poorest” to “richest” by their value of the (continuous) household wealth index or income. We then used the World Bank’s estimate for the proportion of the population in the given country (for the year of the survey’s data collection) that fell below each poverty level (the “poverty headcount ratio”) to select a given proportion (after applying sampling weights) of the participants with the lowest household wealth or income values. For example, in a dataset with 5,000 participants for a country with a poverty headcount ratio of 10% for extreme poverty ($1.90 per day) in the year of the survey’s data collection, we determined that the 500 participants with the lowest household wealth or income value were living in extreme poverty. Details on the household wealth and income measures available in each survey, and our calculation to assign a poverty level to each participant, are provided in Table S13 and Supplementary Materials and Methods 5.4.

### Definition of CVD risk factors and statin use

#### Hypertension

Of the 50 surveys that we used to estimate hypertension prevalence, 37 surveys used a digital upper arm meter for blood pressure (BP) measurements, one survey a digital wrist meter, and one a manual mercury sphygmomanometer (Supplementary Materials and Methods 3.1, Table S2). For 11 countries, the BP measurement device was not stated in the survey documentation. BP was measured three times in 46 countries, three countries undertook two measurements, and one country measured twice inducing a third measurement if the first two differed by more than 10 mmHg. Based on established usage in LMICs and following the recommendations of the International Society of Hypertension, we defined hypertension as systolic BP ≥140 mmHg, diastolic BP ≥90 mmHg, or reporting to be taking medication to lower BP ^30^. If three BP measurements were available, the mean of the last two was employed, and averages were used for surveys with two measurements.

#### Diabetes

Of the 42 surveys that we used to estimate diabetes prevalence, 30 surveys used a point-of-care fasting capillary glucose measurement, three surveys used a laboratory-based assessment of fasting plasma glucose, and nine surveys measured HbA1c (Supplementary Materials and Methods 3.2, Table S3). Following clinical lab standards and published guidelines ^31^, we used plasma equivalents for our analysis by multiplying capillary glucose values by 1.11 in those eight surveys that did not provide plasma equivalents. Based on the WHO diagnostic criteria, respondents were diagnosed with diabetes if they had either (1) a fasting plasma glucose of 7.0 mmol/l (126 mg/dl) or higher, (2) a plasma glucose of 11.1 mmol/l (200 mg/dl) or higher with no fasting, (3) HbA1c records of 6.5% or higher, or (4) reported to take medication for diabetes ^32^. In the 17 surveys for which information on fasting status was missing, we assumed participants to be fasting, thereby complying with the fasting instructions that were given in all surveys.

#### Overweight and obesity

Height and weight were measured by trained personnel in all 72 surveys that we used to estimate overweight and obesity prevalence. Details on height and weight measurements for each survey are given in Supplementary Materials and Methods 3.3, and Table S4. Following WHO classifications, we defined overweight as a body mass index (BMI) of 25.0 to <30.0 kg/m^2^ and obesity as a BMI ≥30.□kg/m^2 33^.

#### Smoking

In the 73 surveys that we used to estimate smoking prevalence, we defined current smoking as stating to be a current smoker of cigarettes or other smoked tobacco products. The specific questions used to define smoking in each survey are detailed in Supplementary Materials and Methods 3.4. and Table S5.

#### Dyslipidemia

Cholesterol measurements were taken using point-of-care devices (see Supplementary Materials and Methods 3.5 and Table S6 for details). We used total cholesterol and low-density lipoprotein (LDL) to determine dyslipidemia. Of the 32 surveys that we used to estimate dyslipidemia prevalence, five had measured both total and LDL cholesterol while the remaining 27 surveys only measured total cholesterol. In the 27 surveys that did not measure LDL but did measure total cholesterol, triglycerides, and high-density lipoprotein (HDL), we used the Friedewald equation to estimate LDL for each participant ^34^. Following the National Cholesterol Education Program’s Adult Treatment Panel III (ATP III) guidelines ^35^, we defined dyslipidemia as (1) having a total cholesterol above 240 mg/dL, (2) self-reporting the use of lipid-lowering medication, or (3) having an LDL higher than 160 mg/dL. More details on the measurement of blood lipids and definition of dyslipidemia in each survey are provided in Supplementary Materials and Methods 3.5. and 5.1.

#### Statin use

Nineteen surveys collected data on statin use. The questions used to determine statin use and CVD history in each survey are detailed in Supplementary Materials and Methods 3.8.

### Statistical analysis

We used age-standardized sampling weights for all prevalence estimates, weighting to the age structure of the WHO standard reference population ^36^. When providing estimates across all included countries, by World Bank country income group, or by world region, we additionally weighted countries according to the proportion of all individuals globally living below the given poverty level who reside in the given country (as estimated using the World Bank’s poverty headcount ratio and the United Nation’s population size estimates ^27,37^). All analyses were restricted to non-pregnant participants aged 15 years or older.

Our analysis had four steps. First, we estimated the prevalence of each CVD risk factor by poverty level (income<$1.90/day, <$3.20/day, <$5.50/day, >$5.50/day, or total population) and World Bank country income group in the year of the survey’s data collection (low-income, lower middle-income, or upper middle-income) ^29^.

Second, to determine how the prevalence of each CVD risk factor varied by individual-level characteristics among those living in extreme poverty, we regressed each CVD risk factor onto age, sex, educational attainment, and rural versus urban residency (both jointly in covariate-adjusted regression and separately in covariate-unadjusted regressions). We used Poisson regression models with a robust error structure, country-level fixed effects, and restricted cubic splines with five knots for age (placed at the 5^th^, 27.5^th^, 50^th^, 72.5^th^, and 95^th^ percentile). Education was categorized as “no formal schooling”, “primary education” (≤ grade 6), and “secondary education (grade 7 to 12) or further”, using local education classifications whenever available, and years of education in cases where categorical variables were unavailable. More detail on the creation of the education variable is provided in Supplementary Materials and Methods 4 and Tables S10-S11. Standard errors were adjusted for clustering at the level of the primary sampling unit (usually a village or neighborhood) and household.

Third, we examined how the prevalence of each CVD risk factor among those living in extreme poverty varied by a country’s level of economic development (as estimated using GDP per capita in the year of the survey’s data collection ^38^) and a country’s “poverty intensity”. We use the term “poverty intensity” in this manuscript to refer to the mean daily income (in 2011 PPP-adjusted dollars) among those living in extreme poverty, as estimated by the World Bank for the year of the survey’s data collection ^39^. It is, thus, a measure of the depth of poverty among those below the poverty line of $1.90 per day. To do so, we plotted the prevalence of each CVD risk factor in a country against the country’s GDP per capita and the country’s poverty intensity among those living in extreme poverty.

Fourth, to ascertain the level of treatment coverage for CVD risk reduction among those living in extreme poverty, we calculated the proportion of participants that self-reported the use of medication to lower BP or blood glucose levels among those with hypertension and diabetes, respectively. In addition, among those with hypertension who reported to be taking BP-lowering medication, we computed the proportion that had a systolic BP <140 mmHg and a diastolic BP <90 mmHg (henceforth referred to as “controlled hypertension”). Lastly, we calculated treatment coverage with statin medication following the WHO guidelines for recommended statin use ^40^. To do so, we computed the proportion of participants: i) with a history of CVD (see Supplementary Materials and Methods 3.8 for the questions used in each survey to ascertain CVD history) who reported to be taking a statin medication (statin use for secondary prevention), and ii) who were 40 years or older with either a diagnosis of diabetes or a ten-year CVD risk greater than 20% (as per the 2019 WHO laboratory-based risk equations ^41^; statin use for primary prevention). More detail on the analyses for this fourth step is provided in Supplementary Materials and Methods 5.3.

This analysis was considered exempt for not-human-subjects research by the institutional review boards at the Stanford University School of Medicine and the Heidelberg University Medical Faculty.

### Data availability

This study includes individual-level data from 105 surveys. Data are publicly available for 102 of these surveys. For data that are not publicly accessible and for which we have arranged specific data-use agreements, we are unable to share these data given the terms of our agreements. All data management and analysis code will be posted in a public repository upon acceptance of the manuscript.

## Results

### Sample characteristics

Our analysis included 105 surveys with a total sample size of 32,695,579 participants (**Table 1**). The survey-level median age of participants was 35.0 years with a survey-level interquartile range of 32.0 to 39.0 years. Of these 32,695,579 participants, 7,922,289 were living in extreme poverty (<$1.90 per day), 15,986,099 on less than $3.20/day, and 23,466,879 on less than $5.50/day. The median survey response rate was 96.3%, ranging from 91.0% to 98.8%.

**Table 1.**
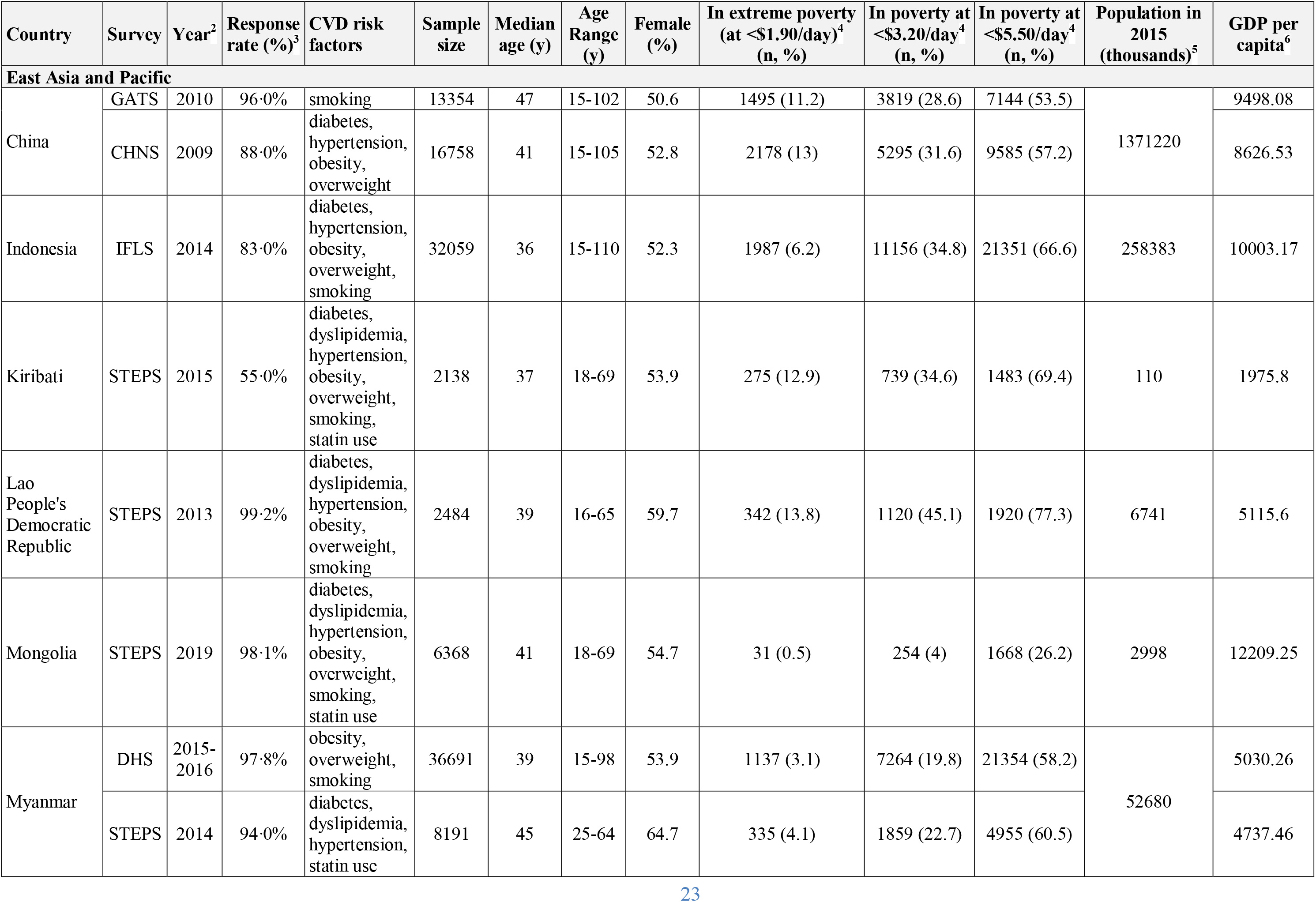
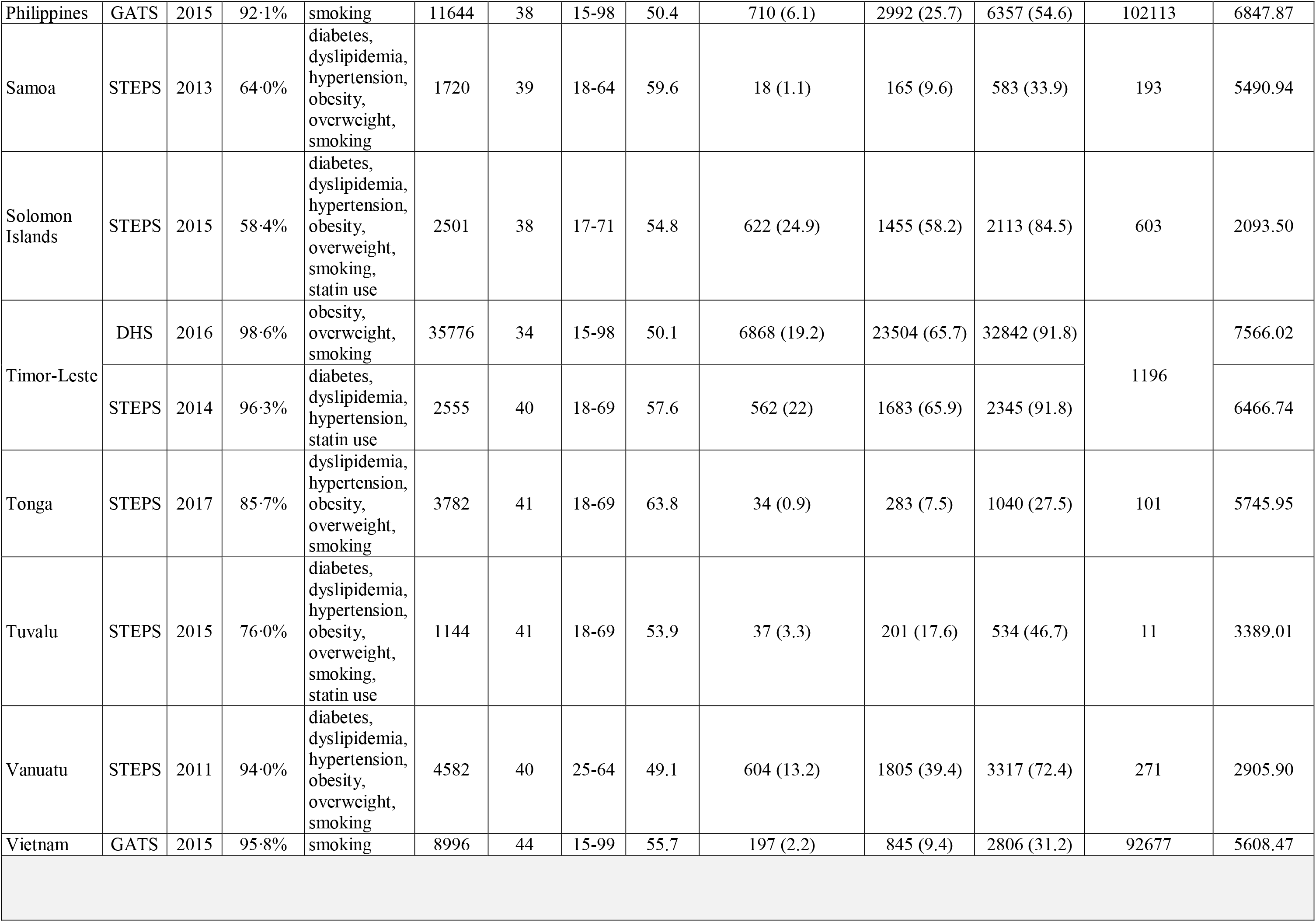
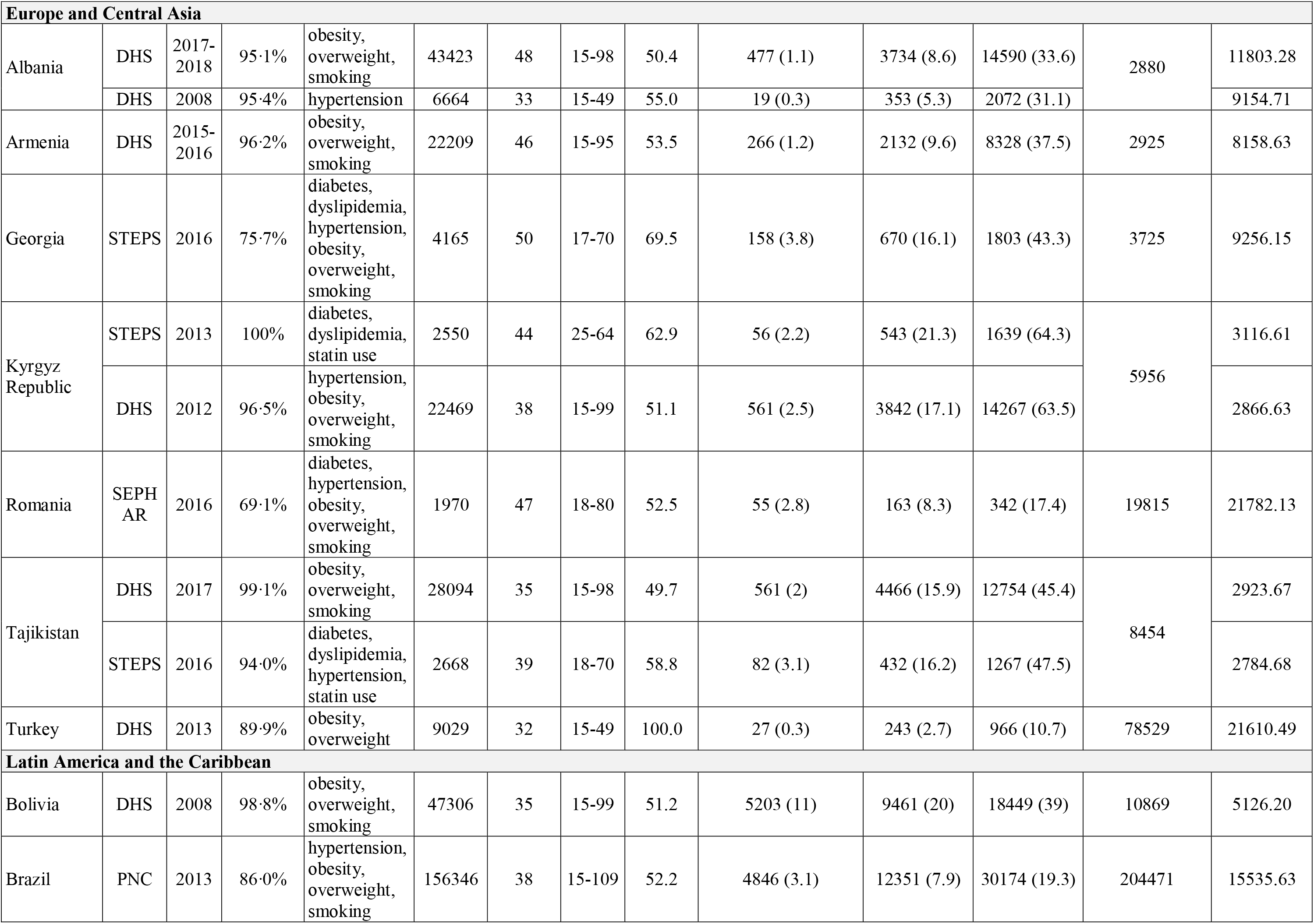
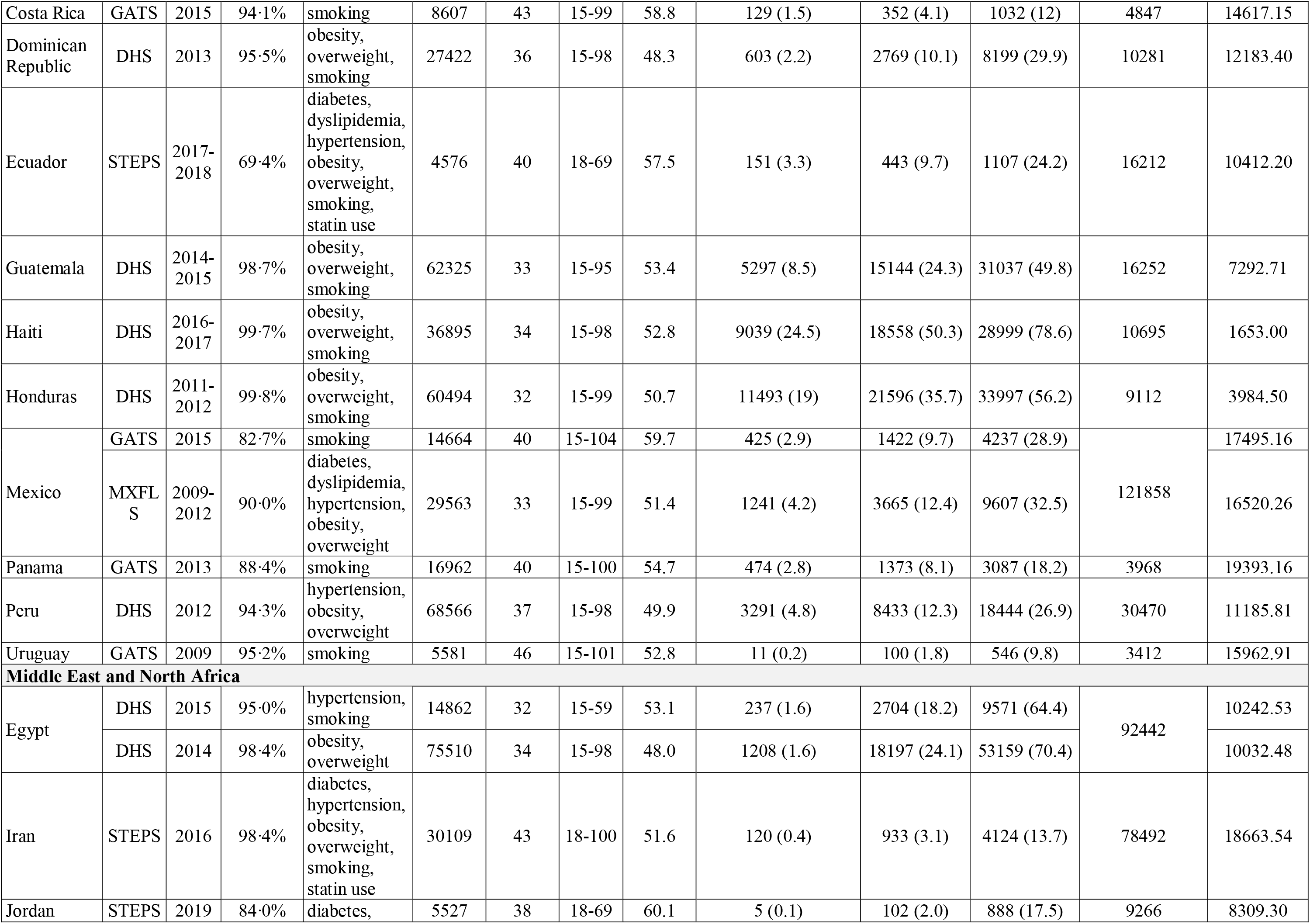
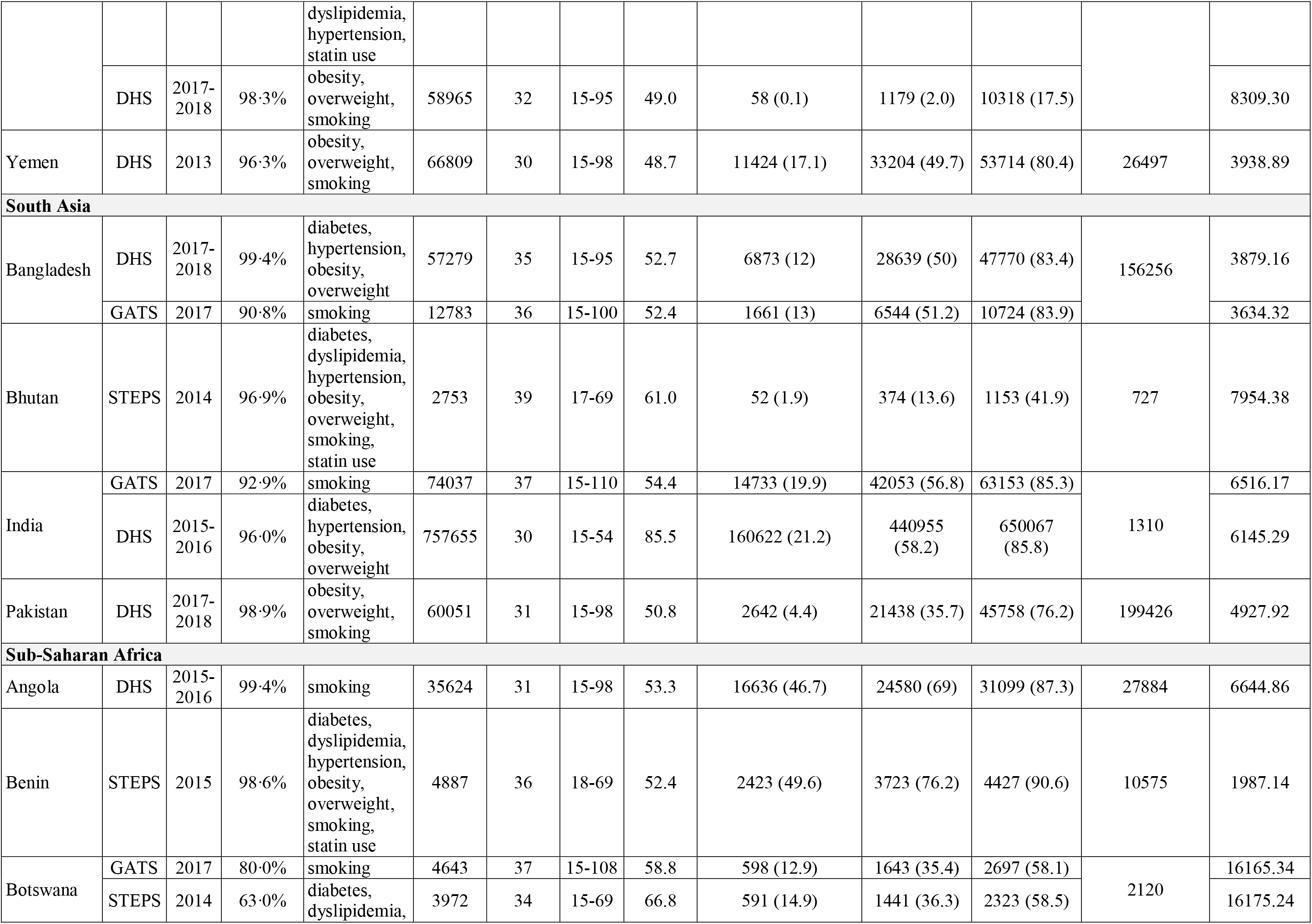
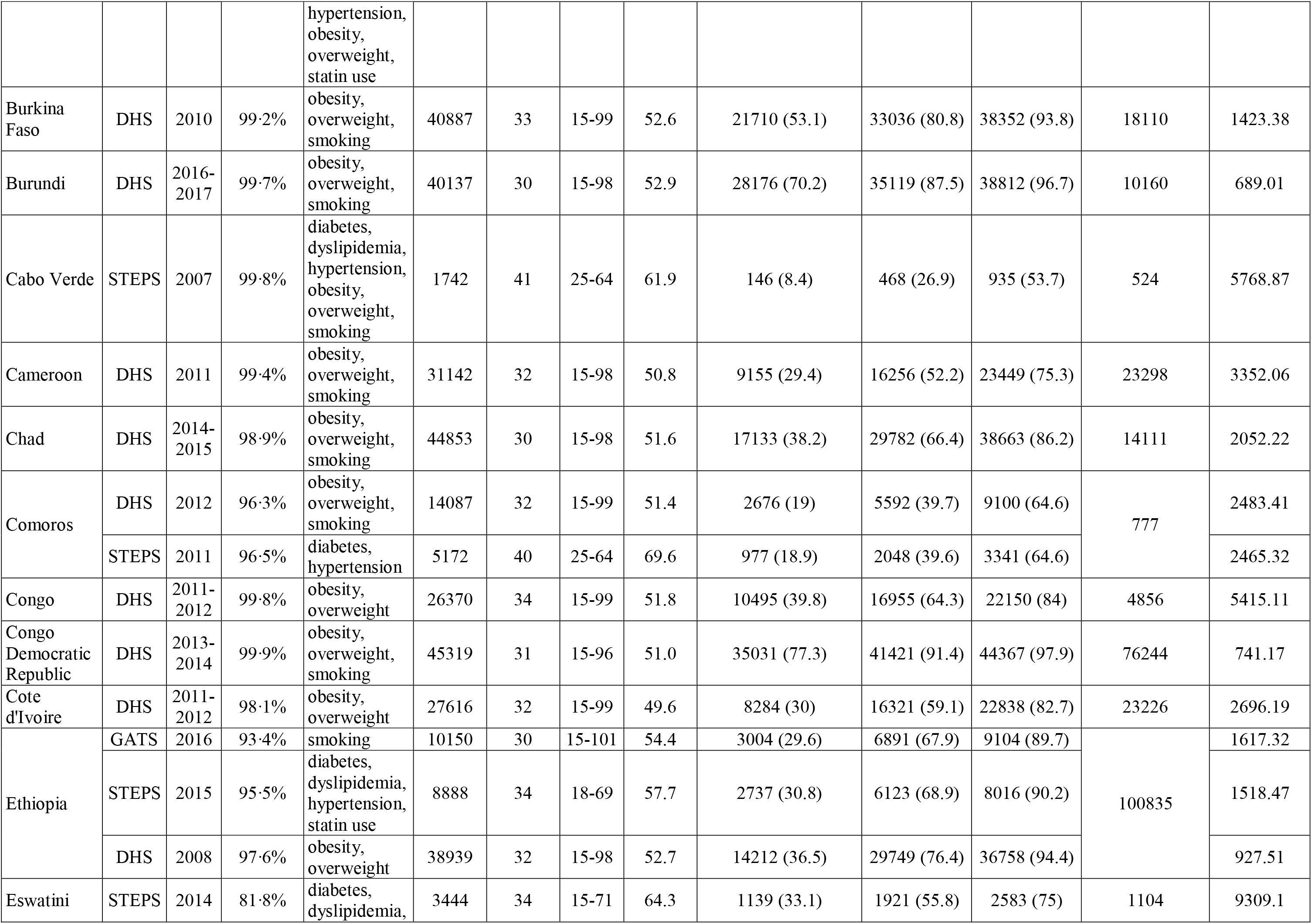
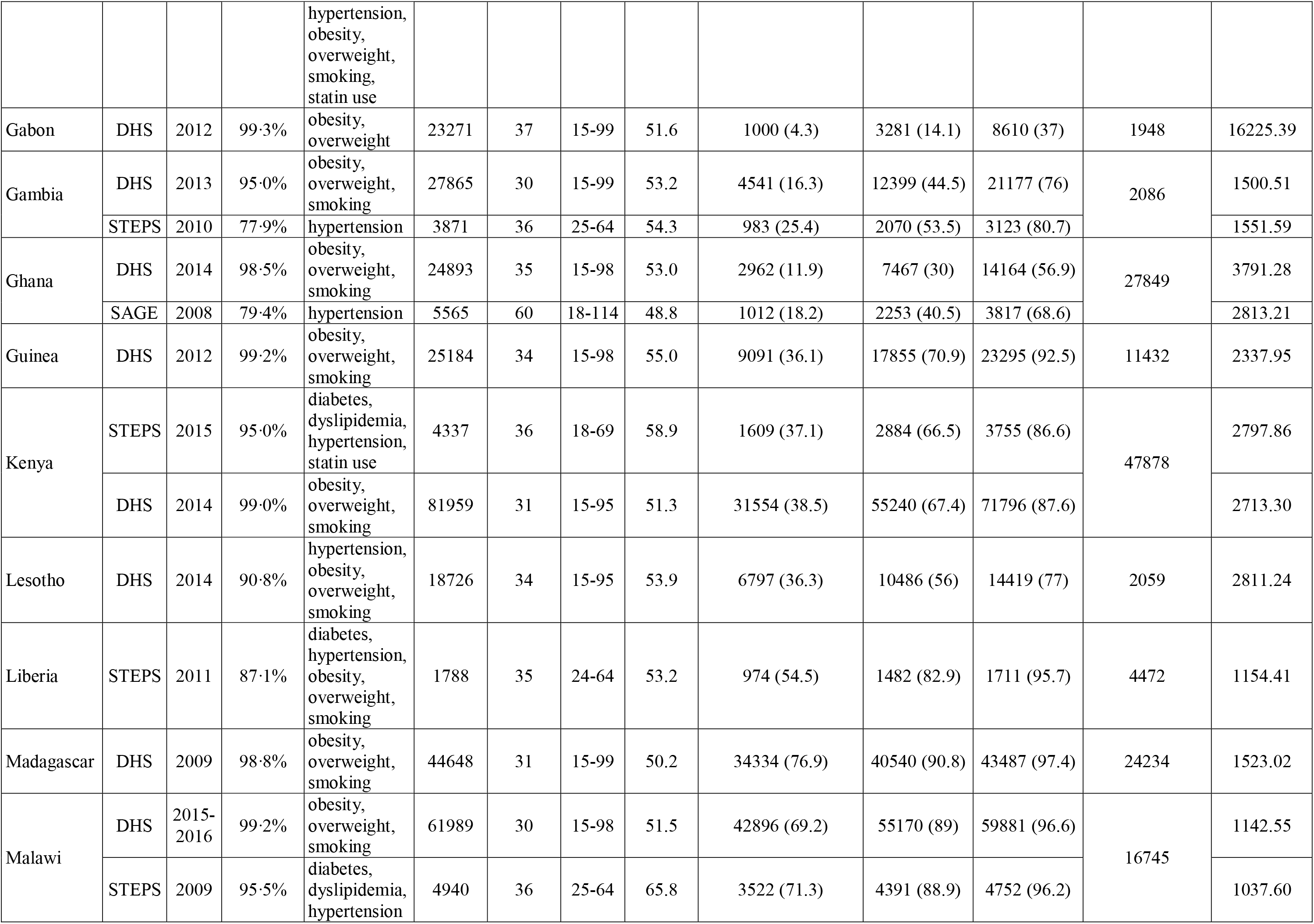
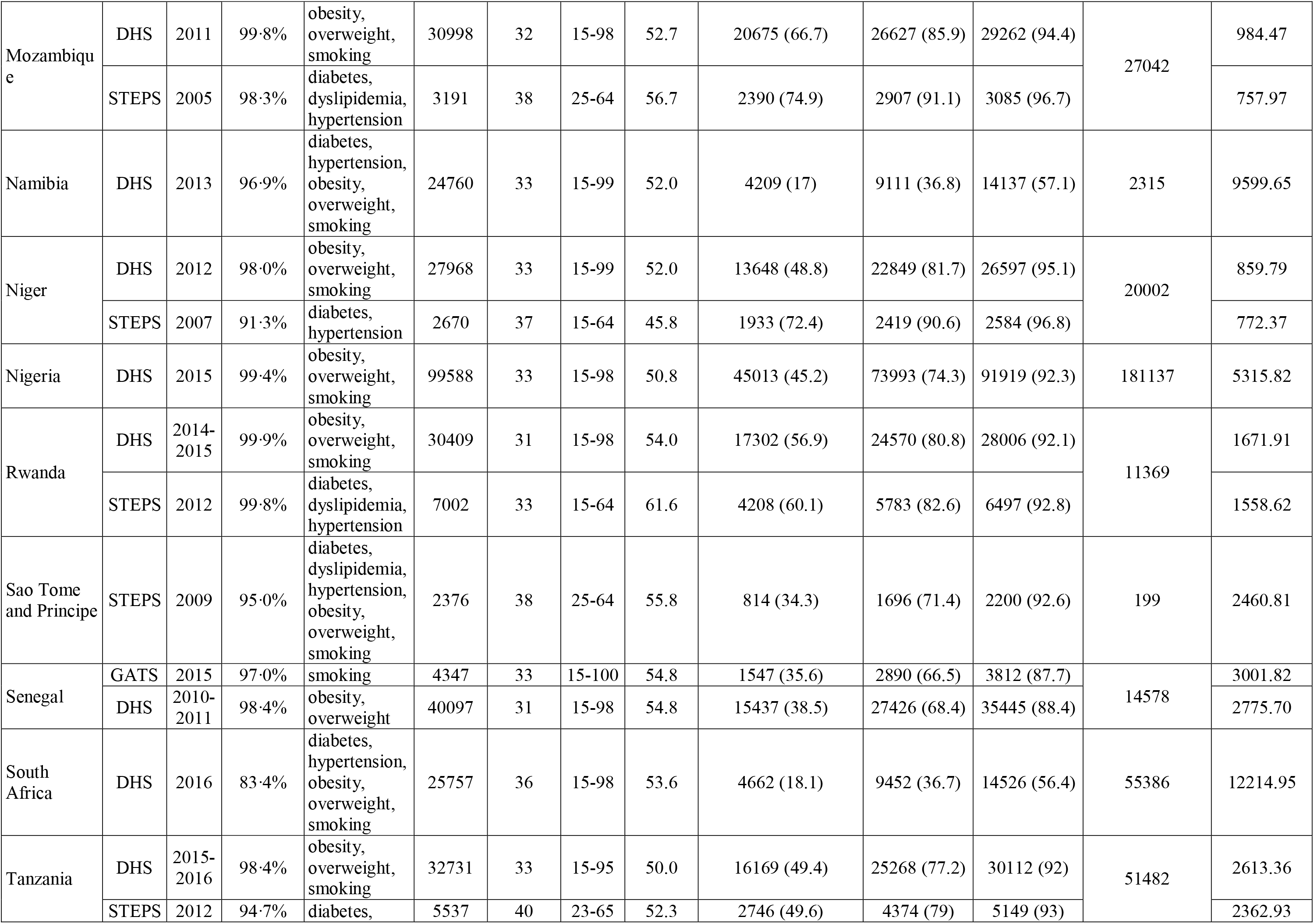
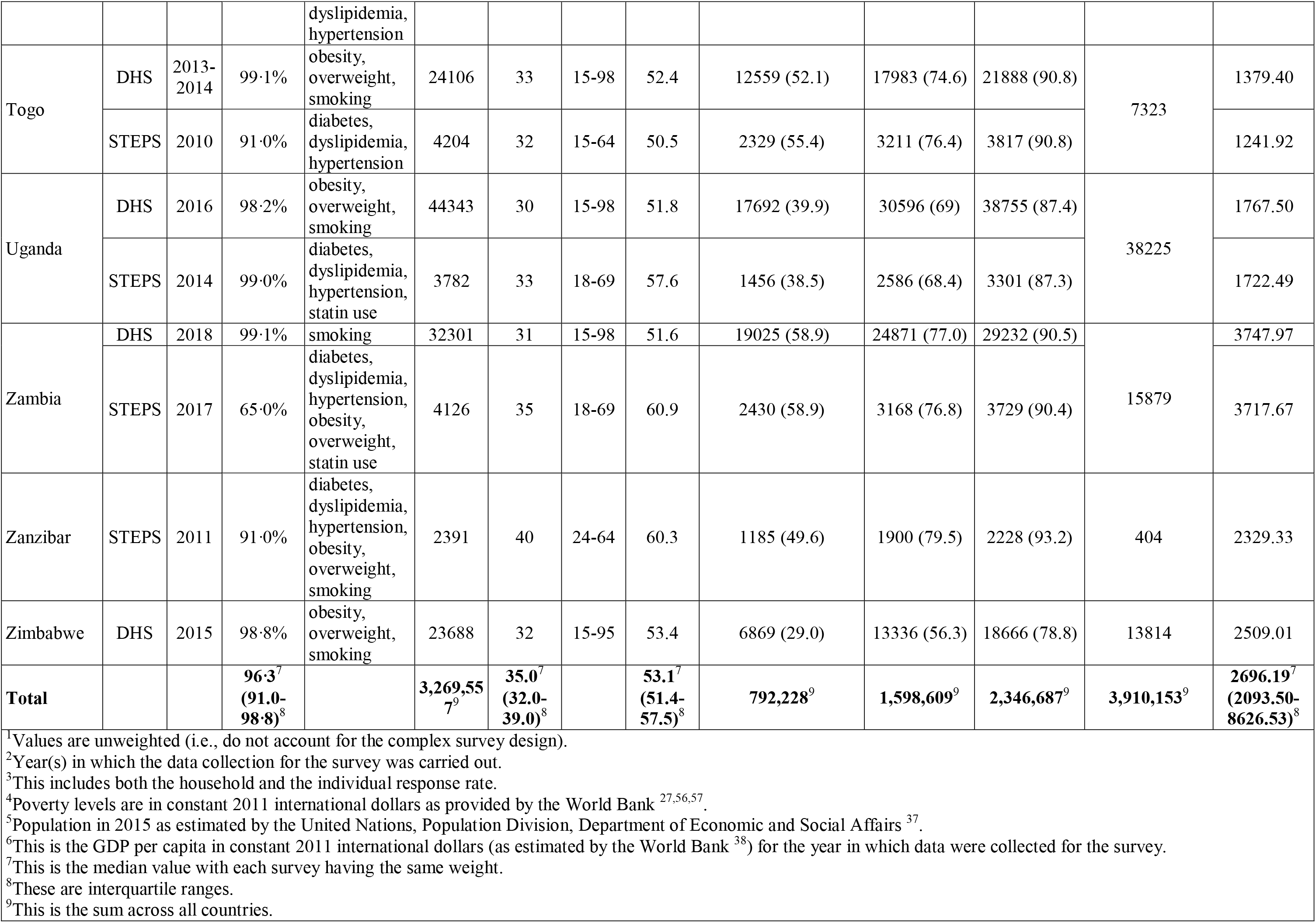
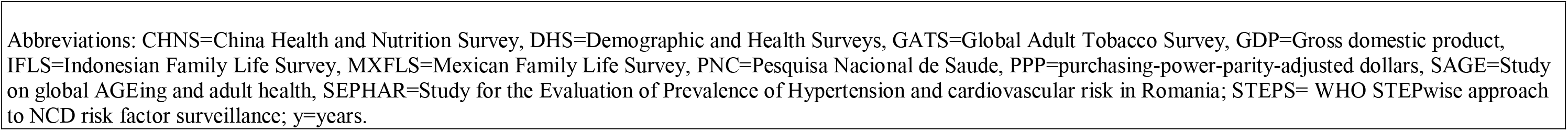
Survey characteristics by region and county^1^

### Prevalence of CVD risk factors among adults living in extreme poverty

Across all our study countries, the age-standardized prevalence of each CVD risk factor among those living in extreme poverty (<$1.90 per day) was 17.5% (95% CI: 16.7% – 18.3%) for hypertension, 4.0% (95% CI: 3.6% – 4.5%) for diabetes, 10.6% (95% CI: 9.0% – 12.3%) for current smoking, 3.1% (95% CI: 2.8% – 3.3%) for obesity, and 1.4% (95% CI: 0.9% – 1.9%) for dyslipidemia (Table S14).

Stratified by World Bank country income category, **Figure 1** compares the prevalence of each CVD risk factor among those living in extreme poverty (<$1.90 per day) to the prevalence among the entire population, as well as those living below $3.20 per day, below $5.50 per day, and above $5.50 per day. With the exception of current smoking, obesity, and dyslipidemia in lower middle-income countries, the prevalence of each CVD risk factor among those living in extreme poverty (<$1.90 per day) was not notably lower than in the total population, and frequently even exceeded the prevalence in the total population. The prevalence of *hypertension* was above ten percent among each population income group in Figure 1 in low-income, lower middle-income, and upper middle-income countries, with relative differences between these population groups being comparatively small. Similarly, differences in the prevalence of *diabetes* across population income groups did not exceed six percentage points. A clear gradient wherein those with higher incomes had a higher prevalence of diabetes was only apparent in lower middle-income countries. The prevalence of *current smoking* was low across population income groups in low-income countries, and high across population income groups in upper middle-income countries. As for diabetes, we observed an income gradient in the prevalence of current smoking in lower middle-income countries wherein individuals with higher incomes were more likely to smoke. The prevalence of *obesity* displayed a similarly positive income gradient across all World Bank country categories. The prevalence of *dyslipidemia* was below three percent among each population income group in low-income countries and above ten percent among each population income group in upper middle-income countries. In lower middle-income countries, the prevalence of dyslipidemia had a strong positive income gradient.

**Fig. 1.**
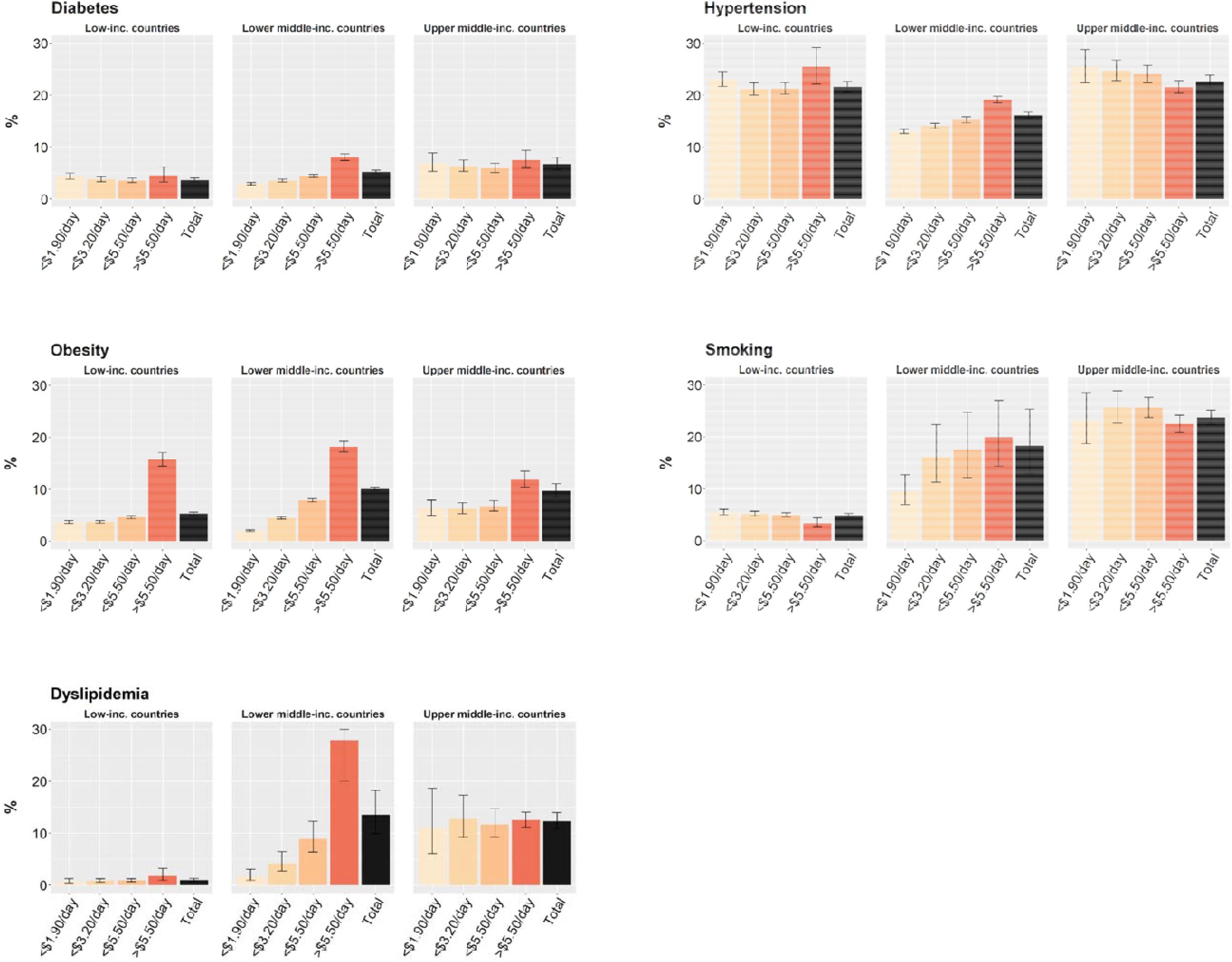
Age-standardized prevalence (in percent) of CVD risk factors by poverty level in low-income, lower middle-income, and upper middle-income countries Figure 1 depicts age-standardized prevalence of CVD risk factors for those with income <$1.90/day, <$3.20/day, <$5.50/day, and ≥$5.50/day for diabetes, hypertension, obesity, smoking, and dyslipidemia. For dyslipidemia, the upper bound of the 95% CI for those with income ≥$5.50/day in lower middle-income countries is 37.4%. Abbreviations: inc.=income

### Variation in CVD risk factor prevalence among those living in extreme poverty by sex, education, and rural-urban residency

Among participants living in extreme poverty, only diabetes was associated with differential prevalence by rurality, with urban dwellers at higher risk (RR: 1.47 [95% CI: 1.08 – 2.00]) (**Figure 2**). When stratifying those living in extreme poverty by sex, men had a slightly higher prevalence of hypertension (RR: 1.09 [95% CI: 1.01 – 1.17]), a substantially lower prevalence of obesity (RR: 0.36 [95% CI: 0.24 – 0.54]), and a far higher prevalence of smoking (RR: 13.16 [95% CI: 10.37 – 16.70]) than women. No significant differences in CVD risk factor prevalence were seen by education level except for smoking, which was more common among those in extreme poverty with a high school education or above compared to those with no schooling (RR: 1.45 [95% CI: 1.22 – 1.74]).

**Fig. 2.**
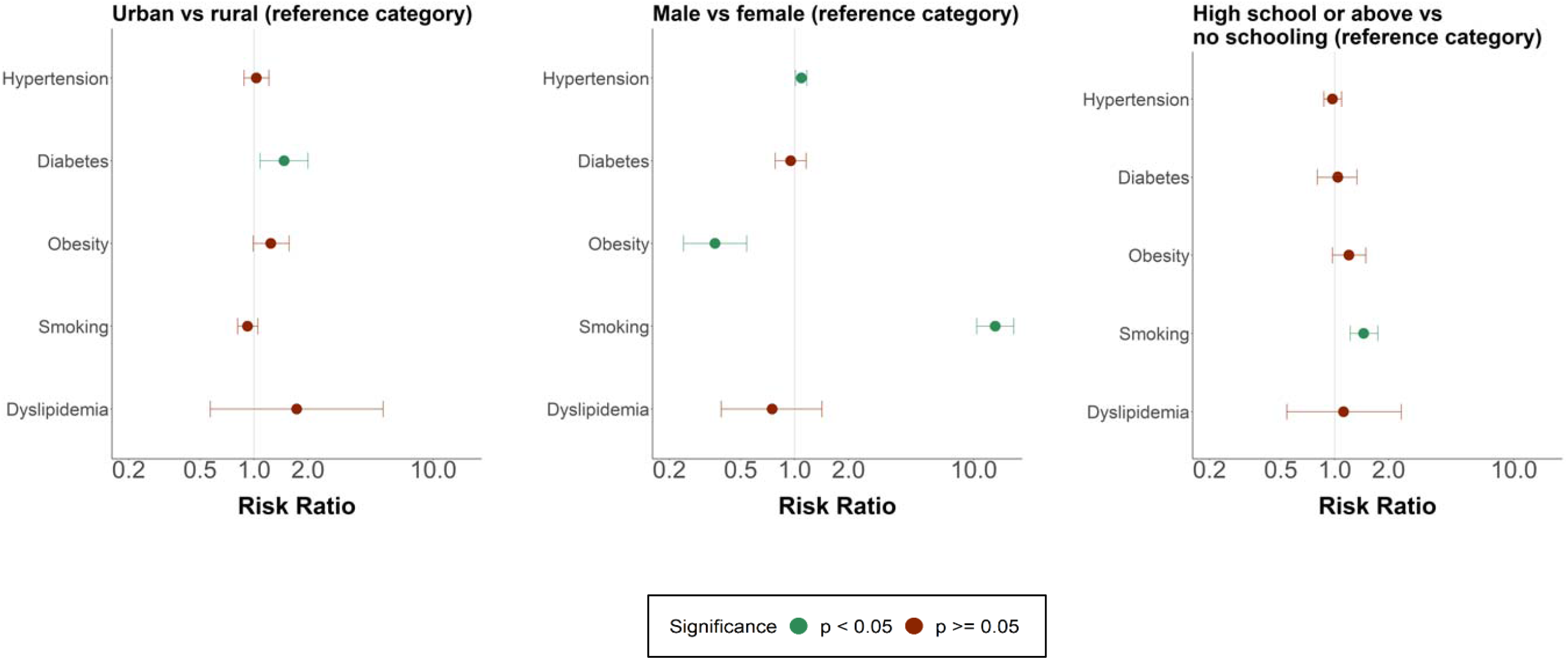
Variation in CVD risk factor prevalence among those living in extreme poverty (<$1.90/day) by rural-urban residency, sex, and education Figure 2 depicts variation in CVD risk factors among those living in extreme poverty with stratification by rurality, sex, and level of education. The risk ratio is plotted on a logarithmic scale.

### Association of CVD risk factor prevalence among those living in extreme poverty with a country’s economic development and intensity of poverty

The prevalence of overweight and obesity among those living in extreme poverty (<$1.90 per day) was positively associated with a country’s level of economic development (GDP per capita) as well as the mean daily income of those living in extreme poverty in a country (i.e., the less “intense” the mean level of poverty among those living in extreme poverty in a country, the higher was the prevalence of overweight and obesity among those living in extreme poverty) (**Figure 3**). A country’s economic development and poverty intensity was not significantly correlated with the prevalence of hypertension and diabetes among those living in extreme poverty. Current smoking was positively associated with a country’s mean daily income among those living in extreme poverty. Analogous analyses as in Figure 3, but for those living below $3.20 per day and $5.50 per day, are shown in Figures S11-S13, and demonstrated similar patterns.

**Fig. 3.**
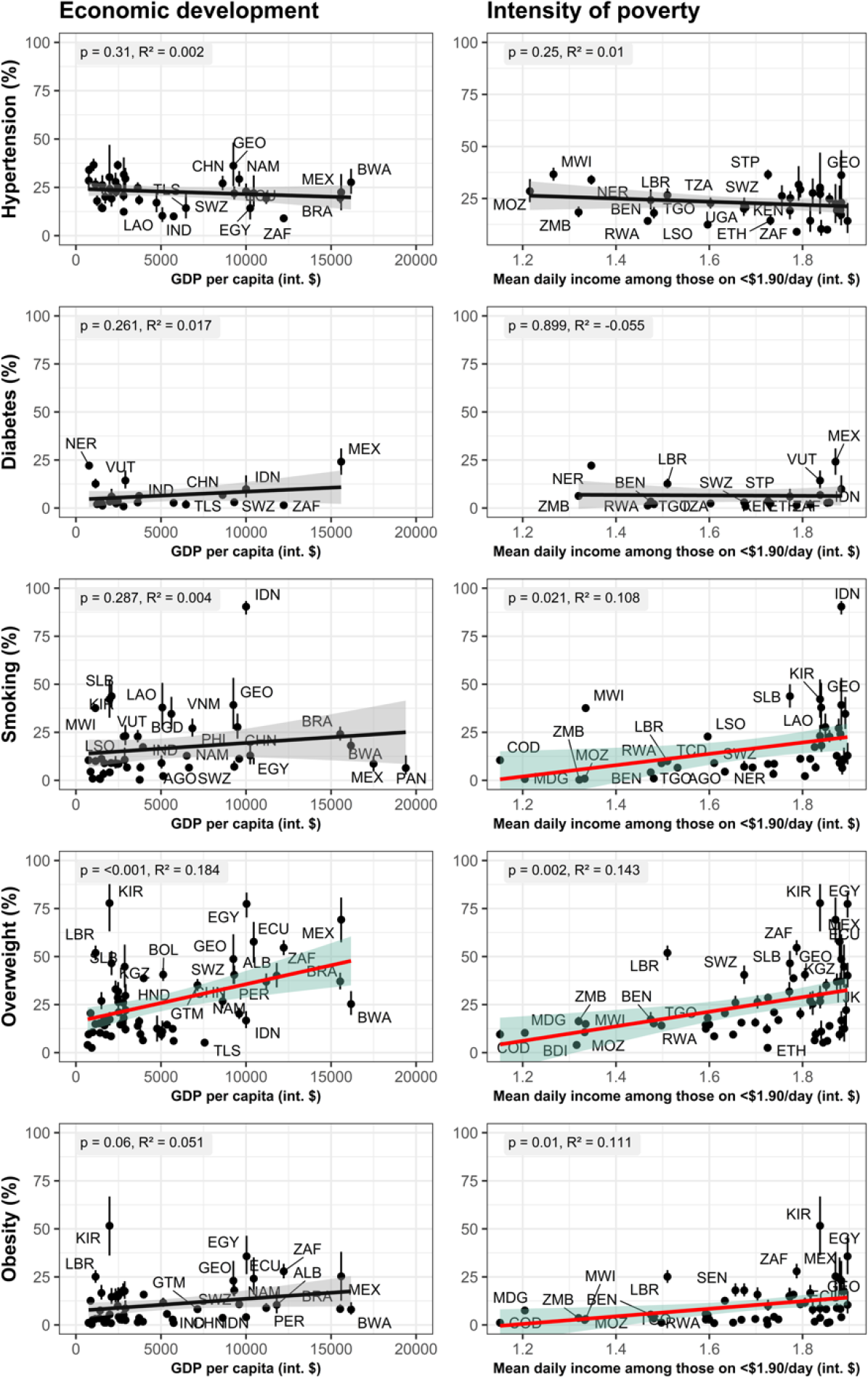
Associations of CVD risk factor prevalence among those living in extreme poverty (<$1.90/day) with countries’ GDP per capita and poverty intensity^1,2,3,4^ ^1^Regression lines in red and confidence intervals in green indicate trends that are statistically significant (p<0.05). ^2^The diagonal lines (in black or red) depict ordinary least-squares regressions (with each country having the same weight) of the CVD risk factor prevalence among those living in extreme poverty (<$1.90/day) in a country onto countries’ GDP per capita or mean daily income among those living on <$1.90/day. The R^2^ and p-values shown in the grey boxes refer to these ordinary least-squares regressions. ^3^The grey or green boundary depict the point-wise 95% prediction interval and the vertical bars are 95% confidence intervals around point estimates. ^4^GDP per capita and the mean daily income among those living on <$1.90/day in a country was taken from the World Bank for the year of the survey’s data collection ^38,39^. Abbreviations: GDP=Gross Domestic Product; int.=international. Country abbreviations: AGO=Angola; ALB=Albania; BDI=Burundi; BEN=Benin; BOL=Bolivia; BRA=Brazil; BWA=Botswana; CHN=China; COD=Congo Democratic Republic; ECU=Ecuador; EGY=Egypt; ETH=Ethiopia; GEO=Georgia; GTM=Guatemala; HND=Honduras; IND=India; IDN=Indonesia; KEN=Kenya; KGZ=Kyrgyz Republic; KIR=Kiribati; LAO=Laos; LBR=Liberia; LSO=Lesotho; MDG=Madagascar; MEX=Mexico; MOZ=Mozambique; MWI=Malawi; NAM=Namibia; NER=Niger; PAN=Panama; PER=Peru; PHL=Philippines; RWA=Rwanda; STP=São Tomé and Principe; SLB=Solomon Islands; SWZ=Eswatini; TCD=Chad; TGO=Togo; TJK=Tajikistan; TLS=Timor-Leste; TZA=Tanzania; VNM=Vietnam; VUT=Vanuatu; ZAF=South Africa; ZMB=Zimbabwe Figure 3 depicts associations of CVD risk factor prevalence among those living in extreme poverty (<$1.90/day) with countries’ GDP per capita and poverty intensity.

### Treatment of CVD risk among those living in extreme poverty

Among those living in extreme poverty (<$1.90/day) who had hypertension, 15.2% (95% CI: 13.3% – 17.1%) reported taking BP-lowering medication and 5.7% (95% CI: 4.7% – 6.7%) had achieved hypertension control (i.e., systolic BP <140 mmHg and diastolic BP <90 mmHg). 19.7% (95% CI: 15.3% – 24.1%) of those living in extreme poverty (<$1.90/day) with diabetes reported taking blood glucose-lowering medication. Among those living in extreme poverty (<$1.90/day) who should be taking a statin for secondary prevention of CVD according to WHO guidelines, only 1.1% (95% CI: 0.4% – 1.9%) were doing so. In low-income countries, hypertension treatment and control, diabetes treatment, and statin use were low among each poverty level (**Figure 4**). In lower middle-income countries, individuals living at more extreme levels of poverty had a consistently lower probability of being on treatment for their hypertension and diabetes, achieving hypertension control, and using statins for primary or secondary prevention of CVD. These gradients by poverty level were less pronounced in upper middle-income countries, where instead the difference between those at any poverty level of less than <$5.50/day versus living on >$5.50/day tended to be the largest.

**Fig. 4.**
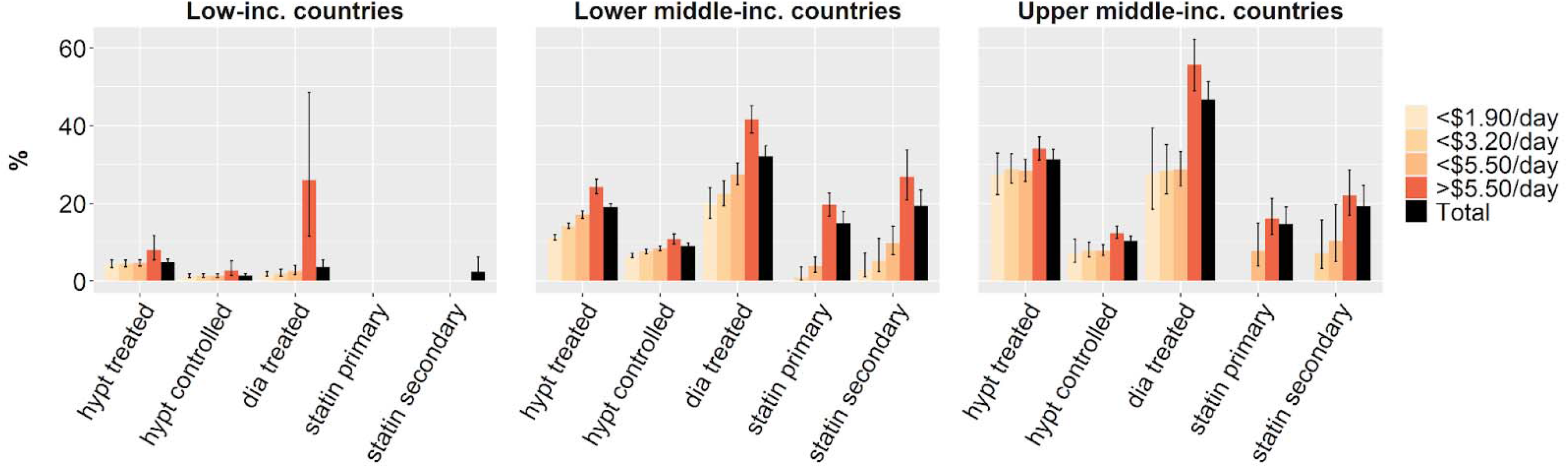
Hypertension treatment and control, diabetes treatment, and statin use, by individuals’ poverty level and World Bank country income group^1,2,3^ ^1^Prevalences were not calculated for samples with a denominator of less than 50 individuals. This was the case for statin use for both primary and secondary prevention of CVD in low-income countries (except for the total population for secondary prevention), statin use for primary prevention of 5 CVD among those living on <$1.90/day in lower middle-income countries, statin use for primary prevention of CVD among those living on <$1.90/day and <$3.20/day in upper middle-income countries, and statin use for secondary prevention of CVD among those living on <$1.90/day in upper middle-income countries. ^2^All prevalence estimates are age-standardized. ^3^”Statin primary” and “statin secondary” refer to statin use for primary and secondary prevention of CVD, respectively. 10 Abbreviations: inc=income; dia=diabetes; hypt=hypertension. Figure 4 depicts hypertension diagnosis and treatment, diabetes treatment, and statin use by poverty level and World Bank country income group.

## Discussion

Our analysis of nationally representative survey data on CVD risk factors from 78 LMICs that are home to the vast majority (an estimated 85% ^27,28^) of individuals living in extreme poverty globally generally found a high prevalence of CVD risk factors among this population group. In fact, in most countries included in our analysis, the prevalence of these risk factors among those living in extreme poverty was not notably lower than in the total population. Our findings thus contradict the common assumption that the environment (e.g., food scarcity ^3–7^) and lifestyles (e.g., more physical labor ^3,6,12,13^) of those living in extreme poverty in LMICs results in a minimal prevalence of CVD risk factors among this group. At the same time, we find marked positive income gradients in rates of evidence-based treatment for these conditions, which further increases the risk of CVD events, such as myocardial infarctions and strokes, among those in extreme poverty relative to wealthier individuals in LMICs.

In 2015, The Lancet NCDI Poverty Commission was founded in the conviction that “non-communicable diseases and injuries (NCDIs) are an important, yet an under-recognized and poorly-understood contributor to the death and suffering” of the poorest billion in the world ^42^. Focusing largely on the political, economic, and financing aspects of addressing NCDIs in LMICs ^42^, the commission restricted its quantitative analyses on the prevalence of NCDIs and its risk factors among the poorest billion merely to data from seven Health and Demographic Surveillance Sites (each representative for populations ranging in size from 31,000 to 129,000 ^43^) in five countries in sub-Saharan Africa. In what is, to our knowledge, the first study to estimate CVD risk factor prevalence among adults living in extreme poverty using multi-country nationally representative data, this research thus provides a crucial empirical foundation for the future work of this commission. At its inception, the commission hypothesized that NCDIs among those in extreme poverty are more likely to be the result of infections and harmful environments (e.g., exposure to environmental toxins) than behavioral risk factors ^44^. Although we did not examine infectious and environmental risk factors, we found a comparatively high prevalence of five major CVD risk factors, for which behavioral choices are thought to play an important causal role ^14,45,46^, among adults in extreme poverty.

In addition to our findings on the overall prevalence of CVD risk factors among those living in extreme poverty, our detailed analyses of how CVD risk factor prevalence and treatment coverage vary by individual-level characteristics among this population group could inform the effective targeting and tailoring of appropriate interventions and policies to reduce CVD risk in this vulnerable group. However, because risk factors may not map perfectly or predictably onto resultant CVD outcomes, such as because of competing risk from infectious causes of death, our data are only suggestive of the CVD burden of those living in extreme poverty as compared to that of wealthier population groups. They cannot on their own establish whether investments in treating CVD risk factors in LMICs would be equitable. Nevertheless, we would like to highlight five points that we believe are important when considering the equity implications of investing in reducing CVD risk in LMICs. First, we found that those living in extreme poverty in LMICs not only have a high prevalence of CVD risk factors but are generally also least likely to be taking CVD risk-reducing medications. Second, it is likely that this vulnerable segment of society has the least access to high-quality care for CVD events in most settings ^47,48^. Third, those living in extreme poverty experience far greater wage losses relative to their household spending from an illness episode (including for CVD) than do wealthier population groups ^49^. Fourth, we found that the prevalence of overweight and obesity among those living in extreme poverty are positively associated with a country’s GDP per capita and the mean daily income among those living in extreme poverty. It is, therefore, likely that as LMICs develop economically, overweight and obesity prevalence among the poorest segments of their societies will increase and, at least partially as a result of this rise in unhealthy weight, the prevalence of diabetes, dyslipidemia, and hypertension will also grow. Lastly, relative to its disease burden, care and prevention for CVD, and NCDs more broadly, are currently vastly underfunded compared to infectious diseases and maternal and child health ^50,51^.

Our study has several additional limitations. First, while the countries included in our sample comprised 86% of the current global population of LMICs (as well as 85% of the global population) estimated to be living in extreme poverty ^27,37^, they are not a probabilistic subset of countries but were instead selected due to their data availability. They may, thus, not fully represent all LMICs or adults living in extreme poverty globally. Second, the surveys were conducted in different years, limiting comparability among them (although a comparison of countries was not the primary objective of our study) and generalizability to the present. They do, however, constitute the most recent nationally representative data available. Third, we did not have detailed consumption data for our surveys, but had to rely instead on household wealth indices or data on household income to define extreme poverty. In a subset of 38 surveys in which we did not have a household wealth index, we used self-reported household income to define poverty. This is a limitation in so far as households in LMICs commonly have multiple income sources (complicating calculations for participants) and varying income between different years ^52^. In addition, we were unable to adjust household income based on household size in this subset of 38 surveys due to a lack of consistent information on the number of household members and their age across surveys. In 19 of these 38 surveys, the survey for which we had to use total household income was only used for a subset of the five CVD risk factors examined in this analysis. Fourth, we did not have sufficiently detailed and homogeneous data to be able to use multi-dimensional measures of poverty, such as the multi-dimensional poverty index ^53^. Fifth, hypertension and diabetes were assessed in one-off measurements rather than the multiple measurements that are generally recommended for a clinical diagnosis of these conditions ^54,55^. Lastly, although the primary objective of our analysis was not to compare countries (or surveys) with each other, it is possible that the variation in the questionnaires and methodology for measuring hypertension, diabetes, and dyslipidemia across our surveys introduced some level of bias in our analyses of how CVD risk factor prevalence among those living in extreme poverty is associated with countries’ GDP per capita and poverty intensity. However, the three main sources of data used in this analysis (the DHS, STEPS, and GATS) each used a standardized questionnaire and approach to measuring CVD risk factors.

In sum, this study demonstrates that CVD risk factors affect individuals across the full socioeconomic spectrum, including those living in extreme poverty, within countries at all levels of economic development. Although we did not directly assess CVD morbidity and mortality nor the disease burden from non-CVD causes, our findings of a high CVD risk factor prevalence along with a low coverage with CVD risk-reducing medications among those living in extreme poverty suggest that equity concerns are likely misplaced in justifying the current vast underfunding of CVD prevention and care compared to other health areas in LMICs ^50,51^. Our unique effort of collating nationally representative data on CVD risk factors from 78 LMICs and applying a consistent definition of poverty across surveys can inform not only resource allocation decisions but also the design of appropriate preventive or care interventions.

## Data Availability

All data produced are available online at

https://www.who.int/teams/noncommunicable-diseases/surveillance/systems-tools/steps

https://dhsprogram.com/Data/

https://www.who.int/teams/noncommunicable-diseases/surveillance/systems-tools/global-adult-tobacco-survey

https://data.worldbank.org/indicator/

## Acknowledgements

We would like to thank all participants in the household surveys that have been harmonized and analyzed in this study.

## Funding

Chan Zuckerberg Biohub (PG)

Veterans Affairs Office of Academic Affairs Advanced Fellowship in Health Services Research (RLT)

Alexander-von-Humboldt university professorship through the Alexander von Humboldt Foundation (TB)

## Author contributions

Conceptualization: PG, TB, SV

Methodology: PG, LS, FM, MM, MT, TB, SV

Visualization: PG, RLT, LS, FM

Funding acquisition: PG, TB

Project administration: PG, RLT, LS

Supervision: PG

Writing – original draft: PG, RLT, LS

Writing – review & editing: PG, RLT, LS, FM, JL, KKA, AD, CH, JMAJ, NL, MM, SSM, KJM, MM, MT, JID, DF, JMG, JS, TB, SV

## Competing interests

The authors declare that they have no competing interests.

## Materials and correspondence

Correspondence and materials requests should be addressed to the corresponding author (PG).

## Supplementary materials

Materials and Methods

Supplementary Text

Figs. S1 to S26

Tables S1 to S51

References

## Notes

### Competing Interest Statement

The authors have declared no competing interest.

### Author Declarations

The study used survey data from the Stepwise Approach to Surveillance (STEPS) database (https://www.who.int/teams/noncommunicable-diseases/surveillance/systems-tools/steps), the Demographic and Health Surveys (DHS) (https://dhsprogram.com/Data/), and the WHO Global Adult Tobacco Survey (GATS) initiative (https://www.who.int/teams/noncommunicable-diseases/surveillance/systems-tools/global-adult-tobacco-survey). In addition, data from the World Bank (https://data.worldbank.org/indicator/) were used.

