## Supplementary appendix for "The prevalence of cardiovascular disease risk factors among adults living in extreme poverty: a cross-sectional analysis of 105 nationally representative surveys with 33 million participants"

### Table of Contents

**Materials and Methods S1: Search Methods ... 5**

***1.1 General approach to search*** ... *5*

***1.2 Inclusion criteria for a survey*** ... *5*

***1.3 Countries included in search*** ... *5*

**Materials and Methods S2: Country categories and country-specific sampling methods ... 6**

Table S1. Country categories (World Bank classification) (*29*) ... 6

***2.1. Country-Specific Sampling Methods*** ... *8*

**Materials and Methods S3: Country-specific Measurement Methods ... 125**

***3.1 Hypertension*** ... *125*
Table S2. Hypertension measurement by country ... 125

***3.2. Diabetes*** ... *138*
Table S3. Diabetes measurement by country ... 138

***3.3. Height/Weight*** ... *152*

***3.4. Tobacco use*** ... *168*
Table S5. Tobacco use questions by country ... 168

***3.5. Lipids*** ... *170*
Table S6. Lipid measurement by country ... 170

***3.6. Hypertension medication use*** ... *178*
Table S7. Consistency of phrasing across surveys (Blood pressure medication) ... 178

***3.7. Diabetes medication use*** ... *179*
Table S8. Consistency of phrasing across surveys (Diabetes medication) ... 179

***3.8. Statin use*** ... *181*
Table S9. Phrasing across STEPS surveys for statin use and CVD history ... 181

**Materials and Methods S4: Computation of education categories ... 182**

Table S10. Summary of categorization of educational attainment from survey-specific categories to three-level study variable (DHS, STEPS and GATS) ... 182
Table S11. Categorization of educational attainment from survey-specific categories to three-level study variable, by country ... 183

**Materials and Methods S5: Statistical Analysis ... 193**

***5.1. Cleaning and definition of risk factors*** ... *193*

***5.2. Regression analyses*** ... *194*

***5.3. Computation of the care cascade for hypertension, diabetes and statins*** ... *195*
*Hypertension* ... 195
*Diabetes* ... 195
*Statins* ... 195

### Materials and Methods S1: Search Methods

#### *1.1 General approach to search*

This project was conducted under the auspices of the GHP Project on Access to Care for Cardiometabolic Diseases (HPACC), a collaboration of researchers from the Department of Global Health and Population at the Harvard T.H. Chan School of Public Health and partners worldwide (*58*). We obtained datasets through a systematic request approach, searching for WHO STEPS, GATS and DHS datasets first and complementing these by a Google search, as detailed in other HPACC publications (e.g. (*59*)).

#### *1.2 Inclusion criteria for a survey*

1) Based on the World Bank income classification, the country was an upper-middle, lower-middle, or low-income country at the time of data collection;
2) The survey was conducted during or after 2005; if multiple surveys collected the same risk factor in one country, we used the survey that was conducted more recently. If survey collection years were identical or close, we used the survey with a wider age range;
3) The survey data was made available at the individual level;
4) The survey contained measures of at least one of the following risk factors: hypertension (blood pressure), diabetes (fasting blood glucose or HbA1c), overweight/obesity (height, weight), smoking (self-report via questionnaire), or dyslipidemia (total or LDL cholesterol);
5) The survey included a measurement of household income (self-reported household income or a household wealth index based on questionnaire responses to a household's ownership of selected assets, materials used for housing construction, and types of water access and sanitation facilities) and the poverty headcount ratio was available for the respective country;
6) The survey was nationally representative with a response rate of over 50%.

#### *1.3 Countries included in search*

Afghanistan, Albania, Algeria, American Samoa, Angola, Argentina, Armenia, Azerbaijan, Bangladesh, Belarus, Belize, Bolivia, Bosnia and Herzegovina, Brazil, Bulgaria, Burundi, Cameroon, Central African Republic, Chad Chile, China, Colombia, Congo, Cook Islands, Côte d'Ivoire, Cuba, Democratic People's Republic of Korea, Democratic Republic of the Congo, Djibouti, Dominica, Dominican Republic, Ecuador, Egypt, El Salvador, Equatorial Guinea, Eritrea, Fiji, Gabon, Ghana, Guatemala, Guinea, Guinea-Bissau, Haiti, Honduras, India, Indonesia, Jamaica, Kazakhstan, Kenya, Kyrgyz Republic, Lao People's Democratic Republic, Lesotho, Libya, Macedonia, Madagascar, Malaysia, Maldives, Mali, Marshall Islands, Mauritania, Mauritius, Mexico, Micronesia (Federated States of), Montenegro, Morocco, Mozambique, Namibia, Nicaragua, Niger, Nigeria, Occupied Palestinian Territory, Pakistan, Panama, Peru, Philippines, Romania, Russia, Saint Lucia, Samoa, São Tomé and Principe, Senegal, Serbia, Solomon Islands, Somalia, South Africa, South Sudan, Sudan, Suriname, Syrian Arab Republic, Tajikistan, Thailand, Tunisia, Turkey, Turkmenistan, Ukraine, Venezuela, Yemen, Zambia, Zimbabwe

Overall the STEPS, DHS, GATS and Google search process yielded 105 datasets included in our study (see Figs. S1-4).

**Date of search:** January 2020

### Materials and Methods S2: Country categories and country-specific sampling methods

Table S1. Country categories (World Bank classification) (*29*)

| World Bank income classification | Country |
|---|---|
| Low income | Yemen<br>Burkina Faso<br>Burundi<br>Chad<br>Congo Democratic Republic<br>Ethiopia<br>Gambia<br>Guinea<br>Liberia<br>Madagascar<br>Malawi<br>Mozambique<br>Niger<br>Rwanda<br>Togo<br>Uganda |
| Lower middle income | Indonesia<br>Kiribati<br>Lao People's Democratic Republic<br>Mongolia<br>Myanmar<br>Philippines<br>Samoa<br>Solomon Islands<br>Timor-Leste<br>Vanuatu<br>Vietnam<br>Kyrgyz Republic<br>Tajikistan<br>Bolivia<br>Haiti<br>Honduras<br>Egypt<br>Iran<br>Bangladesh<br>Bhutan<br>India |

| | |
|---|---|
| | Pakistan<br>Angola<br>Benin<br>Cabo Verde<br>Cameroon<br>Comoros<br>Congo<br>Côte d'Ivoire<br>Eswatini<br>Ghana<br>Kenya<br>Lesotho<br>Nigeria<br>São Tomé and Principe<br>Senegal<br>Tanzania<br>Zambia<br>Zanzibar[1]<br>Zimbabwe |
| Upper middle income | China<br>Tonga<br>Tuvalu<br>Albania<br>Armenia<br>Georgia<br>Romania<br>Turkey<br>Brazil<br>Costa Rica<br>Dominican Republic<br>Ecuador<br>Guatemala<br>Mexico<br>Panama<br>Peru<br>Jordan<br>Botswana<br>Gabon<br>Namibia<br>South Africa |
| High income | Uruguay |

[1]Zanzibar has its own Ministry of Health and was therefore included in our analysis.

#### *2.1. Country-Specific Sampling Methods*

*Note: In order to ensure accuracy in reporting, sampling methods are pasted verbatim from specified sources.*

**Albania DHS 2017-2018**

"The ADHS surveys were done on a nationally representative sample that was representative at the prefecture level as well by rural and urban areas. A total of 715 enumeration areas (EAs) were selected as sample clusters, with probability proportional to each prefecture's population size. The sample design called for 24 households to be randomly selected in every sampling cluster, regardless of its size, but some of the EAs contained fewer than 24 households. In these EAs, all households were included in the survey. The EAs are considered the sample's primary sampling unit (PSU). The team of interviewers updated and listed the households in the selected EAs. Upon arriving in the selected clusters, interviewers spent the first day of fieldwork carrying out an exhaustive enumeration of households, recording the name of each head of household and the location of the dwelling. The listing was done with tablet PCs, using a digital listing application. When Interviewers completed their respective sections of the EA, they transferred their files into the supervisor's tablet PC, where the information was automatically compiled into a single file in which all households in the EA were entered. The software and field procedures were designed to ensure there were no duplications or omissions during the household listing process. The supervisor used the software in his tablet to randomly select 24 households for the survey from the complete list of households. All women aged 15-49 who were usual residents of the selected households or who slept in the households the night before the survey were eligible for individual interviews with the full Woman's Questionnaire. Women aged 50-59 were also interviewed, but with an abbreviated questionnaire that left out all questions related to reproductive health and mother and child health. A 50% subsample was selected for the survey of men. Every man aged 15-59 who was a usual resident of or had slept in the household the night before the survey was eligible for an individual interview in these households."

Source: *Institute of Statistics, Institute of Public Health, and ICF. 2018. Albania Demographic and Health Survey 2017-18. Tirana, Albania: Institute of Statistics, Institute of Public Health, and ICF. Available at: https://dhsprogram.com/pubs/pdf/FR348/FR348.pdf*

**Albania DHS 2008-2009**

"Data collection was conducted from 28 October, 2008 to 26 April, 2009 using a nationally representative sample of almost 9,000 households. All women age 15-49 in these households and all men age 15-49 in half of the households were eligible to be individually interviewed. In addition to the data collected through interviews with these women and men, capillary blood samples were collected from all children age 6-59 months and all eligible women and men age 15-49 for anaemia testing. All children under five years of age and eligible women and men age 15-49 were weighed and measured to assess their nutritional status. Finally, blood pressure (BP) was measured for eligible women and men in the households selected for the men's interview to estimate the prevalence of hypertension in the adult population. [...]

The 2008-09 Albania Demographic and Health Survey is based on a representative probability sample of almost 9,000 households. This sample was selected in such a manner as to allow separate urban and rural, as well as regional-level estimates for key population and health indicators, e.g., fertility, contraceptive prevalence, and infant mortality for children under five.

The 2008-09 ADHS utilized a two-stage sample design. The first stage involved selection of a sample of primary sampling units (PSUs) from the PSUs used for the 2008 Living Standards Measurement Study (LSMS). In total, 450 PSUs were selected for the ADHS sample, including 245 urban PSUs and 205 rural PSUs, covering 4 geographic domains—mountains, central, coastal, and urban Tirana. A listing of each of the selected PSUs was carried out in preparation for the LSMS. The ADHS survey selected 20 households from the updated household listing in each PSU, excluding those households selected for the LSMS. In two PSUs, numbers 27 (13 households) and 172 (17 households), there were less than 20 households in the re-listed PSU—all households were selected in those cases. In a further 6 PSUs there were less than 20 households after the LSMS households were excluded. In these PSUs some of the households from the LSMS sample were included to bring the number of households selected up to 20. After selection of the households, the sample selection forms were printed and the list of selected households was adapted for use in a Personal Digital Assistant (PDA). All women age 15-49 in the total sample of households, and all men age 15-49 in the sub-sample of half of the households, who were either usual residents of the households or visitors present in the household on the night before the survey were eligible to be interviewed."

Source: *Institute of Statistics, Institute of Public Health [Albania] and ICF Macro. 2010. Albania Demographic and Health Survey 2008-09. Tirana, Albania: Institute of Statistics, Institute of Public Health and ICF Macro. Available at: https://dhsprogram.com/pubs/pdf/FR230/FR230.pdf*

**Angola DHS 2015-2016**
The original report is in Portugese. We translated the sampling strategies using Google translate.

"The 2015-2016 IIMS sample was selected from the results base and from the mapping of the General Population and Housing Census (RGPH) of Angola, carried out by INE in 2014, and guarantees a representation at national, provincial, urban level and rural, as well as in terms of sociodemographic characteristics such as sex, age groups, education level and socioeconomic quintiles of the population. The sample is stratified by province and urban / rural area with three stages of selection. In the first stage, 627 primary sampling units were selected with probability proportional to the size, the size measure being the number of households in each stratum within each province. In the second stage, 26 households were selected with equal probabilities in the primary urban and rural sampling units. This selection was made after a previous listing of households. Based on this procedure, 16,302 households were selected for the survey. The sample covers only the population residing in households, excluding households and their members residing in collective residences, such as hotels, hospitals, military barracks, student residences, etc., and the homeless. In each province, 33 primary sampling units were selected, with the exception of Luanda, with 66."

Source: *Instituto Nacional de Estatística - INE/Angola, Ministério da Saúde - MINSA/Angola, Ministério do Planeamento e do Desenvolvimento Territorial (MINPLAN) and ICF. 2017. Inquérito de Indicadores Múltiplos e de Saúde em Angola 2015-2016. Luanda, Angola e Rockville, Maryland, EUA: INE, MINSA, MINPLAN and ICF. Available at: https://dhsprogram.com/pubs/pdf/FR327/FR327.pdf*

**Armenia DHS 2015-2016**
"The sampling frame used for the 2015-16 ADHS is the Armenia Population and Housing Census, which was conducted in Armenia in 2011 (APHC 2011). The sampling frame is a complete list of enumeration areas (EAs) covering the whole country, a total number of 11,571 EAs, provided by the National Statistical Service (NSS) of Armenia, the implementing agency for the 2015-16 ADHS. This EA frame was created from the census data base by summarizing the households down to EA level. A representative probability sample of 8,749 households was selected for the 2015-16 ADHS sample. The sample was selected in two stages. In the first stage, 313 clusters (192 in urban areas and 121 in rural areas) were selected from a list of EAs in the sampling frame. In the second stage, a complete listing of households was carried out in each selected cluster. Households were then systematically selected for participation in the survey. [...] Because of the approximately equal sample size in each marz, the sample is not self-weighting at the national level, and weighting factors have been calculated, added to the data file, and applied so that results are representative at the national level.

All women age 15-49 who were either permanent residents of the households in the 2015-16 ADHS sample or visitors present in the household on the night before the survey were eligible to be interviewed. Interviews were completed with 6,116 women. In addition, in a subsample of one-half of all households selected for the survey, all men age 15-49 were eligible to be interviewed if they were either permanent residents or visitors present in the household on the night before the survey. Interviews were completed with 2,755 men. Height and weight information was collected from eligible women age 15-49 and children age 0-59 months in all households."

Source: *National Statistical Service [Armenia], Ministry of Health [Armenia], and ICF. 2017. Armenia Demographic and Health Survey 2015-16. Rockville, Maryland, USA: National Statistical Service, Ministry of Health, and ICF. Available at: https://dhsprogram.com/pubs/pdf/FR325/FR325.pdf*

**Bangladesh DHS 2017-2018**
"The sample for the BDHS 2017-18 is nationally representative and covers the entire population living in non-institutional housing units in the country. Bangladesh consists of eight administrative departments: Barishal, Chattogram, Dhaka, Khulna, Mymensingh, Rajshahi, Rangpur and Sylhet. Each urban area is divided into areas, which are further subdivided into each area. The area is divided into Union Parishades (UPs) and territories within the United States. These subdivisions can divide the country as a whole into urban and urban areas.

The survey is based on a two-stage stratified sample of households. In the First Stage, 675 EAs (250 in urban areas and 425 in rural areas) were selected with probability proportional to EA size. The sample in that stage was drawn by BBS, following the specifications provided by ICF that include cluster allocation and instructions on sample selection. A complete household listing operation was then carried out in all selected EAs to provide a sampling frame for the second-stage selection of households. In the second stage of sampling, a systematic sample of an average of 30 households per EA was selected to provide statistically reliable estimates of key demographic and health variables for the country as a whole, for urban and rural areas separately, and for each of the eight divisions.

Based on this design, 20,250 residential buildings were selected. Completed interviews were expected from around 20,100 unmarried women aged 15 to 49 years. In addition, in a subsample of a quarter of households (approx. 7-8 households per EA), all married women 50 and older, unmarried women 18 and older, and men 18 and older were weighed and measured their height. [...] The survey was successfully conducted in 672 clusters after eliminating three clusters (one in the city and two in the countryside) that were completely eroded by the floods. These clusters were located in Dhaka (an urban cluster), Rajshahi (a rural cluster) and Rangpur (a rural cluster). A total of 20,160 households were selected for the survey."

Source: *National Institute of Population Research and Training (NIPORT), and ICF. 2020. Bangladesh Demographic and Health Survey 2017-18. Dhaka, Bangladesh, and Rockville, Maryland, USA: NIPORT and ICF. Available at: https://dhsprogram.com/pubs/pdf/FR344/FR344.pdf*

**Bangladesh GATS 2017**
"A multi-stage, geographically clustered sample design was used to produce nationally representative data. A total of 14,880 households were sampled. One individual was randomly chosen from each selected household to participate in the survey."

Source: *no report available. Sampling information obtained from the fact sheet. Available at: https://www.who.int/tobacco/surveillance/survey/gats/fact-sheet-gats-bangladesh-2017.pdf?ua=1*

**Benin STEPS 2015**
The original report is in French. We translated the sampling strategies using Google translate.

“The study was conducted using a three-stage random sampling technique. The sampling frame was provided by the National Institute of Statistics and Economic Analysis (INSAE) from the data of the fourth General Census of Population and Housing (RGPH4) in Benin in 2013. The first stage consisted of the random selection of 260 Enumeration Areas (EAs); The second stage consisted in drawing lots for 20 households per EA; The third step consisted in randomly selecting one individual per household retained according to the Kish method recommended by the WHO for the STEPS survey.”

Source: *Rapport final de l’enquête pour la surveillance des facteurs de risque des maladies non transmissibles par l'approche ‘’STEPSwise’’ de l'OMSENQUETE ‘’STEPS 2015’’ au Bénin. Available at: https://www.who.int/ncds/surveillance/steps/benin/en/.*

**Bhutan STEPS 2014**

"To achieve a nationally representative sample, a multistage sampling method was used to select enumeration areas, households and eligible participants at each of the selected households in three stages. The 2005 National Census was chosen as the basis for the sampling frame, with "Geogs" (blocks) in rural areas and towns in urban areas forming the primary sampling units (PSUs). Since the population distribution for urbanicity is 70:30 (rural:urban), 63 PSUs in rural and 14 PSUs in urban areas were chosen. PSUs were selected through the probability proportionate to size (PPS) sampling using the number of households in each PSU. Two secondary sampling units (SSUs) for every rural PSU and 4 SSUs for every urban PSU were selected. This led to the selection of 126 SSUs from rural and 56 SSUs from urban areas. This was also carried out by PPS sampling, using the number of households in each SSU. A total of 16 households from each SSU (both rural and urban) were selected using systematic random sampling. The sampling frame for this was the list of households with a unique identification number (ID) developed by the enumerators for the survey. At the household level, the Kish sampling method was used to randomly select one eligible member (aged 18–69 years) of the household for the survey. The Kish method ranks eligible household members in order of decreasing age, starting with males and then females, and randomly selects a respondent using the automated program for Kish selection in the handheld personal digital assistant (PDA)."

Source: *National survey for noncommunicable disease risk factors and mental health using approach WHO Steps Approach in Bhutan – 2014. Available at: http://www.who.int/chp/steps/bhutan/en/.*
*Additional reference: World Health Organization Regional Office for South-East Asia. National survey for noncommunicable disease risk factors and mental health using WHO STEPS approach in Bhutan—2014. Geneva: World Health Organization; 2014.*

**Bolivia DHS 2008**
The original report is in Spanish. We translated the sampling strategies using Google translate.

"*Sample Design*
As in the previous ENDSAS, the sample design for the ENDSA 2008 provides a nationally probabilistic sample, which is stratified, by conglomerate and in two stages. In the first stage, 1,000 conglomerates of households were selected throughout the country - called Primary Sampling Units (UPM) - with probability proportional to their size. In a second stage, a fixed number of 20 households were selected within each UPM selected in the first stage. In each department, the UPMS of the sample framework were stratified according to the categorization of the population sizes. The urban residence area is defined as all the towns of 2,000 inhabitants and more. The stratification defined in the sample framework is also implicitly applied to ENDSA 2008. The units of analysis are women of childbearing age and their corresponding children under five years of age located in the selected households; in addition to men aged 15 to 64 found in a subsample of these same homes.

*Objectives of the Sample Design*
The 2008 ENDSA sample was designed to obtain estimates of the various indicators with acceptable levels of confidence at different levels of geographical disaggregation. These levels are:

- Country total
- Total for each of the two areas of residence: urban and rural
- Total for each of the three geographical regions: Altiplano, Valle and Llano
- Total for each of the nine departments

In relation to maternal mortality, the estimation of the maternal mortality ratio with the assigned size of 20,000 households is only presented for the total of the country and even in this case it is considered relatively inaccurate.

*Sampling frame*
The sample framework used for the design and selection of the ENDSA 2008 sample was provided by the National Statistics Institute, which has been prepared with information and material from the last National Population and Housing Census of 2001. The 1,000 Units Sampling Primary selected from this framework for ENDSA 2008 were updated in terms of the number and location of resident households. Each UPM has a complete identification by means of an assigned code, in addition to having a perfectly delimited map."

Source: *Coa, R, Ochoa, LH & ICF International. Bolivia Encuesta Nacional de Demografía y Salud 2008. Available at: https://dhsprogram.com/pubs/pdf/FR228/FR228[08Feb2010].pdf.*

**Botswana GATS 2017**
"The sampling frame for GATS Botswana was delineated from the 2011 Population and Housing Census. During the 2011 Population and Housing census, Botswana was divided into census districts. Therefore, stratified random sampling was used and the sampling units were stratified by census districts. The 26 census districts in Botswana thus served as the strata. However, during the analysis, the separate domains of estimation were urban and rural areas where semi-urban areas in the country were split into either urban or rural depending on the ratio of urban EAs to rural EAs in a district. Mostly urban districts, (i.e. more than 50% of urban EAs), fell within the urban stratum and the same logic with the rural stratum. Thus, the urban stratum was made up of 10 urban districts, whereas the rural stratum was comprised of 16 rural districts, as illustrated by Table 1 below. The Primary Sampling Units (PSUs) were the Enumeration Areas (EAs) whereas Secondary Sampling Units (SSUs) and Tertiary Sampling Units (TSUs) were households and GATS-eligible household residents, respectively.

The 2011 Population and Housing census defined the Enumeration Area (EA) as the smallest geographic unit, which represents an average workload for an enumerator over a specified period. The EA was established using the average number of dwelling units because a dwelling unit in Botswana has identifiable boundaries. A dwelling unit in its traditional sense consists of one or more structures or buildings, permanent or temporary, usually surrounded by a fence/wall or something to mark its boundaries. In some areas, the huts and/or other structures may not be fenced, but they are usually in very close proximity. Dwelling units in Botswana have target addresses, which provide access to the household(s). [...]

A stratified three-stage probability sampling design was used to select the sample for this survey. The sample was first allocated proportionately across the census districts as shown by Table 1 above and the first stage was the selection of EAs (PSUs). In this survey, the measure of size was the number of households in a district as per the 2011 Population and Housing Census. Probability Proportional to Size (PPS) systematic sampling method with a random start was then applied to select the EAs and fifteen (15) households per EA. Although the EAs were established using the average number of dwelling units and the average size of an EA was approximately 120-150 dwelling units, a list of all EAs in Botswana with corresponding sizes (i.e. number of households) were created during the mapping and listing exercise. The cumulative population sizes of the EAs were also established and this was divided by the calculated sample size to determine the Sampling Interval (SI). A random number between 1 and SI was then selected and subsequent sampling numbers were calculated. Sampling numbers were then compared with the cumulated sizes and the PSUs selected were the first whose cumulated sizes were greater or equal to the sampling numbers.

The second stage of sampling was selection of households (SSUs). Fresh lists of households within the selected EAs were prepared by Botswana GATS team (more specifically the supervisors and IT staff ) during households mapping and listing. Mapping and listing were conducted prior to deployment of the interviewers and this helped in the preparation of case files. Statistics Botswana also provided technical support during mapping and listing of households. New household listing was used to update the household information in selected enumeration areas and to take care of the expected variation in both the average size and differential growth of the EAs since the last mapping was done in 2011. Both occupied and non-occupied households were listed in this survey and the listing followed the GATS Household Listing manual. Households within a dwelling unit were listed independently provided that people living in those households do not make common provision for food or other essentials for living together. Otherwise, a dwelling unit with more than one household, in which people living in it make common provision for food or other essentials for living together, was considered as one household. Systematic random sampling with a random start was then applied to select households from the roster of households prepared. The systematic selection interval was determined as the number of occupied households on the EA listing form divided by the number of households selected per EA, rounded to the nearest integer.

The third stage of sampling was the selection of eligible individuals residing in selected households (TSUs). A roster of individuals, aged 15 years and above within the selected household, was created and one individual, aged 15 years or older within the selected household, was then randomly selected from the roster to administer the questionnaire."

Source: *Botswana Ministry of Health and Wellness: Global Adult Tobacco Survey: Report 2017. Available at: https://www.afro.who.int/sites/default/files/2021-01/GATS%20Botswana%202017%20Report.pdf*

**Botswana STEPS 2014**

"Botswana has a population of over 2 million with 27 districts and 4,845 enumeration areas and sample size of 300 enumeration areas with a target population of 6,400 people was systematically drawn from a pool of the whole enumeration areas. Against the identified enumeration areas numbers of households were listed and proportion of participants was calculated from the total sample size required for the country. Finally a computer generated random number was drawn to go into specific households in that specific enumeration area and at the end eligible participants residing in the household were listed into the electronic hand held data assistant (PDA) and at the end a name was picked automatically to participate in the survey."

<u>Source:</u> *Republic of Botswana - Ministry of Health and WHO : Botswana STEPS survey report on Non Communicable Diseases Risk Factors.2015. Available at: https://extranet.who.int/ncdsmicrodata/index.php/catalog/318*

**Brazil Pesquisa Nacional de Saúde 2013**
The text below was translated from:
https://www.pns.icict.fiocruz.br/index.php?pag=planoamostragem

"The Master Sample is a set of units of areas that are selected to meet various surveys of the IBGE Integrated System of Household Searches (SIPD). These units are considered primary sampling units (PSUs) in the sample planning of each of the surveys that use the Master Sample, such as PNS. The sampling plan consists of the stratification of the UPAs and selection of these units with probability proportional to the size, given by the number of permanent private households (DPPs).

The register for selection of the Master Sample was a file containing information from the Demographic Census 2010 on the census tracts of the geographic scope, whose limits are defined in the Operational Geographic Base 2010, totaling 316574 sectors. A sector or set of sectors with at least 60 DPPs was defined as UPA, with the exception of a few units, because it was not possible to aggregate sectors in some municipalities.

The stratification of the UPAs obeys four different criteria: administrative, including the division of the UF into capital, rest of the Metropolitan Region (RM) or Integrated Region of Economic Development - RIDE, and rest of the UF; geographical subdivision, which subdivides capitals and other large municipalities into more strata; situation that involves rural / urban categorization; and the statistician in order to improve the accuracy of the estimates.

As part of the SIPD, the sampling design of the PNS followed, in part, the sampling design of the Master Sample, especially with regard to the stratification of the UPAs.

The PNS sample is by clusters in three stages of selection:

- 1st stage: selection with probability proportional to the size (given by the number of DPPs in each unit) of the UPAs sub-sample in each stratum of the Master Sample;
- 2nd stage: selection by simple random sample of households in each UPA selected in the first stage;
- 3rd stage: selection by simple random sampling of the adult (person aged 18 years or older) among all adult residents of the household.

The PNS will integrate the SIPD, which will make it possible to relate the information collected with other researches, such as the PNAD and the Household Budget Survey (POF) at different levels of geographic aggregation."

Sources: *Pesquisa Nacional de Saúde. Plano de Amostragem. Published 2010. https://www.pns.icict.fiocruz.br/index.php?pag=planoamostragem.*
*Instituto Brasileiro de Geografia e Estatística (IBGE). Pesquisa Nacional de Saúde 2013. ftp://ftp.ibge.gov.br/PNS/2013/pns2013.pdf. Published 2014.*

**Burkina Faso DHS 2010**
The original report is in French. We translated the sampling strategies using Google translate.

"The 2010 EDSBF-MICS IV covered the population of individuals residing in ordinary households across the country. A national sample of approximately 15,000 households was selected. The sample is stratified to provide an adequate representation of urban and rural settings as well as the 13 areas of study (made up of the 13 administrative regions) for which estimates are available for all key indicators. This is a stratified, two-degree drawn area sample. [...]

- At the first stage, clusters or enumeration areas (EAs) over the whole of the national territory were drawn from the list of EAs established during the General Population and Housing Census (RGPH) of 2006. Overall, 574 clusters, including 176 in urban areas and 398 in rural areas, were selected by carrying out a systematic selection with probability proportional to the size, the size of the EA being the number of households.
- A count of households in each of these 574 clusters provided a list of households from which a second-stage sample of households was drawn with a systematic drawing at equal probability.
- All women aged 15-49 usually living in selected households, or present the night before the survey, were eligible to be surveyed. In addition, in a subsample of one in two households, men aged 15-59 were also surveyed. In this same subsample of households, all women eligible for the survey as well as all children aged 6 to 59 months were to be measured and weighed in order to assess their nutritional status.
- The 574 clusters selected in the sample could be surveyed with the exception of one cluster in the Sahel region. A total of 14,947 households were selected and, of these, 14,536 households were identified and were occupied at the time of the survey. Of these 14,536 households, 14,424 were successfully interviewed, for a response rate of 99%, as shown in Table 1.1."

Source: *Institut National de la Statistique et de la Démographie - INSD/Burkina Faso and ICF International. 2012. Enquête Démographique et de Santé et à Indicateurs Multiples du Burkina Faso 2010. Calverton, Maryland, USA: INSD and ICF International. Available at: https://dhsprogram.com/pubs/pdf/FR256/FR256.pdf*

**Burundi DHS 2016-2017**
The original report is in French. We translated the sampling strategies using Google translate.

"The EDSB-III, 2016-2017 aims to produce representative results at the level of the entire country, urban and rural areas, Bujumbura-Mairie and other provinces of Burundi. To do this, the national territory was divided into 18 study areas corresponding to the 18 provinces and, in each study area (except Bujumbura Mairie which has no rural part), two strata were created: the stratum urban and rural. The sample was drawn stratum by stratum. Thus, the sample of the EDSB-III, 2016-2017 is based on a stratified area survey and drawn at two stages.

At the first stage, 554 Primary Survey Units (PSU) or clusters were drawn from the list Enumeration Areas (ZD) established during the General Population and Housing Census (RGPH) carried out in 2008, by carrying out a systematic selection with probability proportional to the size, the size of the UPS being the number of households. An enumeration of households in each of these clusters provided a list of households from which was drawn, at the second stage, a sample of 30 households per cluster, both in urban and rural areas with a systematic selection from equal probability. A total of 16,620 households (3,180 in urban areas in 106 clusters and 13,440 in rural areas in 448 clusters) were selected.

All women aged 15-49 usually living in selected households or present the night before the survey were eligible to be surveyed. In addition, in a sub-sample of one in two households, all men aged 15 -59 years old were eligible to be surveyed, women (15-49 years old) and children under five years old were eligible for the various biological tests and measures."

Source: *Ministère à la Présidence chargé de la Bonne Gouvernance et du Plan Burundi - MPBGP, Ministère de la Santé Publique et de la Lutte contre le Sida Burundi - MSPLS, Institut de Statistiques et d'Études Économiques du Burundi - ISTEEBU, et ICF. 2017. Troisième Enquête Démographique et de Santé au Burundi. Bujumbura, Burundi : ISTEEBU, MSPLS, and ICF. Available at: https://dhsprogram.com/pubs/pdf/FR335/FR335.pdf*

**Cabo Verde STEPS 2007**
"The STEPS survey in Cape Verde was a population-based survey of adults aged 25-64. A cluster sample design was used to produce representative data for that age range in Cape Verde. A total of 1,762 adults participated in the Cape Verde STEPS survey.

**Sampling procedure:** A multi-stage cluster sample of households. One individual within the age range of the survey was selected per household. Analysis weights were calculated by taking the inverse of the probability of selection of each participant. These weights were adjusted for differences in the age-sex composition of the sample population as compared to the target population.
Different weight variables are available per Step:

wStep1 - for interview data
wStep2 - for physical measures
wStep3 - for biochemical measures

This allows for differences in the weight calculation for each Step of the survey as the age-sex composition of the respondents to each Step can differ slightly due to refusal or drop out. Additionally, some countries perform subsampling for Step 2 and/or Step 3. When no subsampling is done and response rates do not differ across Steps of the survey, the 3 weight variables will be the same."

Source: *no report available. Sampling information obtained from the fact sheet. Available at: https://extranet.who.int/ncdsmicrodata/index.php/catalog/319/related_materials. Additional information is available from WHO NCD Microdata Repository. Available at:https://extranet.who.int/ncdsmicrodata/index.php/catalog/319/study-description#page=sampling&tab=study-desc*

**Cameroon DHS 2018**
The original report is in French. We translated the sampling strategies using Google translate.

"The DCAT-V sample is a random, stratified, 2-stage sample. The primary survey unit is the ZD as defined in the RGPH of 2005. The sample is distributed in such a way as to guarantee an adequate representation of the urban and rural areas as well as of the 12 areas of study which are made up of the regions of Adamaoua, Center (without Yaoundé), East, Far North, Littoral (without Douala), North, North-West, West, Sud and South-West and the cities of Douala and Yaoundé. Apart from the cities of Yaoundé and Douala which are entirely in an urban environment, each of the 10 remaining study areas is separated into urban and rural parts to form sampling strata, i.e. each type of residence has to be within the study area constitutes a sampling stratum. Thus, 22 sampling strata were created and the sample was drawn independently from each stratum. In the first stage, 470 EAs were drawn with a probability proportional to their size, the size being the number of households residing in the EA . Note that before sorting the first stage EAs, the sampling frame was sorted according to the field of study and the area of residence (urban / rural) to form the sampling strata; EAs were then sorted according to administrative units below the region, that is, departments and arrondissements. This operation introduces an implicit stratification at the level of all administrative units located below the region with a sample allocation proportional to the size of the unit.

After the selection of the primary units and before the main survey, a household enumeration and a map update were carried out in each selected EA. The enumeration consisted of visiting each selected ZD to establish a site plan and an access plan to the ZD; to make a quick visit to each resident household to check the status of the housing and note the name of the head of household and the address of the household in order to establish a complete list of the households enumerated for each unit. This list of households was used as a sampling frame for the selection of second-stage households. Before the household enumeration, large EAs exceeding 300 households were divided into segments of which only one segment was retained in the sample. This last step is not considered a degree of selection because the segmentation is only intended to limit the work of enumeration within the EA. Thus, within the framework of the ESDC-V, a cluster corresponds to an EA or part of an EA for those which are very large (more than 300 households) and of which only a part has been selected.

At the second stage, in each of the clusters selected at the first stage, a fixed number of 28 households were selected with a systematic selection with equal probability, from the lists newly established at the time of the enumeration. [...] In total, 470 ZD were selected, of which 245 in urban areas and 225 in rural areas. In addition, 13,160 households were selected in total, of which 6,860 in urban areas and 6,300 in rural areas."

Source: *Institut National de la Statistique/INS et ICF. 2020. Enquête Démographique et de Santé du Cameroun 2018. Yaoundé, Cameroun et Rockville, Maryland, USA : INS et ICF. Available at: https://dhsprogram.com/pubs/pdf/FR360/FR360.pdf*

**Chad DHS 2014-2015**
The original report is in French. We translated the sampling strategies using Google translate.

“The 2014-2015 DHS-MICS is designed to produce representative results at the level of the whole country, at the urban and rural level separately, at the city level of N'Djaména, and at the regional level. from Chad. To do this, the national territory was divided into 21 fields of study corresponding to the 22 regions and the city of N'Djaména and, in each field of study (except N'Djaména), two strata were created: the urban and rural stratum. The sample was drawn stratum by stratum. Thus, the 2014-2015 DHS-MICS sample is based on a stratified and two-stage area survey.

At the first stage, 626 Primary Survey Units (UPS) or clusters were drawn from the list of Enumeration Areas (ZD) established during the General Population and Housing Census (RGPH) carried out in 2009 by INSEED, by carrying out a systematic selection with probability proportional to the size, the size of the UPS being the number of households. A count of the households in each of these clusters provided a list of households from which was drawn, in the second stage, a sample of 25 households per cluster in the urban area, and 30 households per cluster in the rural area with a systematic drawing with equal probability.

A total of 17,965 households (4,075 in urban areas in 163 clusters and 13,890 in rural areas in 463 clusters) were selected. All women aged 15-49 usually living in selected households, or present the night before the survey, were eligible to be surveyed. In addition, in a subsample of one in three households, all men aged 15-59 were eligible to be surveyed. [...]

In two-thirds of households not selected for the male survey, all women and all children under five were eligible to be measured and weighed to assess their nutritional status. Finally, in this sub-sample, questionnaires on mosquito nets, disability, tobacco and alcohol consumption, child labor and discipline were asked.[...] During the 2014-2015 DHS-MICS, among the 626 clusters selected, 624 clusters in the sample were surveyed. A total of 17,892 households were selected and, among them, 17,422 households were identified at the time of the survey.”

Source: *Institut National de la Statistique des Études Économiques et Démographiques - INSEED/Tchad, Ministère de la Santé Publique - MSP/Tchad et ICF International, 2014-2015. Enquête Démographique et de Santé et à Indicateurs Multiples (EDS-MICS 2014-2015). Rockville, Maryland, USA: INSEED, MSP et ICF International. Available at: https://dhsprogram.com/pubs/pdf/FR317/FR317.pdf*

**China GATS 2010**

"A multistage stratified cluster sampling design was applied to the survey (see Appendix B for details). The country was divided into six regions: North, Northeast, East, Center and South, Southwest and Northwest. Each of the six regions was divided into urban and rural areas, making 12 strata in total. In the first stage, urban districts or rural counties / cities were selected at the county level, using the probability proportional to sample size (PPS) from the 2007 household registration database administered by the Ministry of Public Security of China. The measure of size (MOS) was the number of households. In the second stage, two urban neighborhood communities or rural villages were selected from each of the primary sampling units using the PPS method. The selected secondary sampling unit was divided into segments of about 1,000 households (using mapping and listing to determine the number) and one segment was randomly selected. In the third stage, 75 households were selected through simple random sampling from each selected segment. In the last stage, the individual was randomly selected from each participating household using simple random sampling."

Source: *Ministry of Health and the Chinese Center for Disease Control and Prevention: Global Adult Tobacco Survey (GATS). China 2010 country report. Available at: https://www.who.int/tobacco/surveillance/survey/gats/en_gats_china_report.pdf?ua=1*

**China Health and Nutrition Survey 2009**

"The China Health and Nutrition Survey is a longitudinal study across 228 communities within nine provinces of China. Surveys began in 1989, with subsequent surveys every 2–4 years, for a total of nine rounds between 1989 and 2011. The China Health and Nutrition Survey was designed to provide representation of rural, urban and suburban areas varying substantially in geography, economic development, public resources and health indicators,[13] and it is the only large-scale,

longitudinal study of its kind in China. The original survey in 1989 used a multistage, random cluster design in eight provinces (Liaoning, Jiangsu, Shandong, Henan, Hubei, Hunan, Guangxi and Guizhou) to select a stratified probability sample; a ninth province, Heilongjiang, was added in 1997 using a similar sampling strategy. Essentially, two cities (one large and one small city— usually the provincial capital and a lower income city) and four counties (stratified by income: one high, one low and two middle income counties) were selected in each province. Within cities, two urban and two suburban communities were selected; within counties, one community in the capital city and three rural villages were chosen. Twenty households per community were then selected for participation. The study met the standards for the ethical treatment of participants and was approved by the Institutional Review Boards of the University of North Carolina at Chapel Hill and the Institute of Nutrition and Food Safety, Chinese Center for Disease Control and Prevention."

Source: *Attard, Samantha M.; Herring, Amy H.; Wang, Huiling; Howard, Annie Green; Thompson, Amanda L.; Adair, Linda S.; Mayer-Davis, Elizabeth J.; & Gordon-Larsen, Penny. (2015). Implications of Iron Deficiency/Anemia on the Classification of Diabetes Using HbA1c. Nutrition & Diabetes, 5, e166*

**Comoros DHS 2012**
The original report is in French. We translated the sampling strategies using Google translate.

"The 2012 ESDC-MICS II sample is a nationally representative sample, based on a two-stage stratified cluster survey. The 2012 ESDC-MICS II sample is a representative sample of households. As with all past DHS surveys, the EDSC-MICS II comprised 5 fields of study consisting of each of the three main islands (regions) of the country (Mwali, Ndzuwani, Ngazidja), urban and rural areas.

At the first stage, 252 clusters were selected, and drawn in proportion to their size, from the list of Enumeration Areas (EA) established during the General Population and Housing Census (RGPH) of 2003 and, in part, updated in 2012 during mapping work. All selected clusters were subjected to a complete household enumeration. In the second stage, households were drawn, with equal probability, from the list of households established during the enumeration operation. The number of households drawn is 20 in each urban or rural cluster.

In total, 5,041 households were selected for the household survey. All selected household members were identified using the household questionnaire. All women aged 15 to 49 (usual residents or visitors) who were in these households were interviewed individually. In addition, these women, as well as all children under the age of 5, were measured and weighed. Similarly, in one in two households, all men aged 15-59 residing in these selected households were also interviewed individually."

Source: *Direction Générale de la Statistique et de la Prospective - DGSP/Comores and ICF International. 2014. Enquête Démographique et de Santé et à Indicateurs Multiples aux Comores 2012. Rockville, MD 20850, USA: DGSP and ICF International. Available at: https://dhsprogram.com/pubs/pdf/FR278/FR278.pdf*

**Comoros STEPS 2011**
The original report is in French. We translated the sampling strategies using Google translate.

"The STEPS survey on risk factors for chronic diseases in the Union of the Comoros took place from January to March 2011. This study has undertaken Step 1, Step 2 and Step 3. Indeed, socio-demographic and behavioral measures were collected in Step 1. Physical measures such as height, weight and tension were collected in Step 2 and biochemical measurements were collected to assess the levels of blood glucose and cholesterol levels in Step 3. The STEPS survey conducted in Comoros Union is a survey of general population, targeting adults aged 25 to 64 years. A stratified survey was used to produce representative data for this age group. A total of 5556 adults aged 25 to 64 participated in the STEPS survey on a sample of 5760 people representing an overall response rate of 96.5%."

Source: *WHO: Non Communicable Diseases Risk Factors: Union des Comores. STEPS 2011 - Note de synthèse. Available at: http://www.who.int/chp/steps/comoros/en/.*

**Congo DHS 2011-2012**

The original report is in French. We translated the sampling strategies using Google translate.

"The DCAT-II sample is nationally representative. It is based on a two-stage stratified cluster survey. Unlike the ESDC-I which had divided the country into four fields of study (Brazzaville, Pointe-Noire, the South and the North), the ESDC-II retained as its field of study, each of the twelve departments of the countries (Kouilou, Niari, Lékoumou, Bouenza, Pool, Plateaux, Cuvette, Cuvette-Ouest, Sangha, Likouala, Brazzaville and Pointe-Noire). Three departments comprising urban and rural areas (Niari, Bouenza and Sangha) were subdivided into urban and rural to form 15 strata and the sample was drawn independently in each stratum. Given the very large differences in the size of the different departments, a self-weighted sample would not have made it possible to obtain enough cases in the less populated departments. We therefore oversampled the small departments and under-sampled the most populous departments. The data were then weighted to give each department its real weight in the population of the Congo.

At the first stage, 384 clusters were drawn, in proportion to their size, from the list of Enumeration Areas (Enumeration Areas) established during the 2007 RGPH. All the selected clusters were subject to an exhaustive enumeration of households. . In the second stage, households were drawn, with equal probability, from the list of households established during the enumeration operation. The number of households drawn is 24 in each urban cluster and 34 in each rural cluster. In total, 11,727 households were selected for the household survey.

All selected household members were recorded in the household questionnaire. All women aged 15 to 49 (usual residents or visitors) who were in these households were interviewed individually. In addition, these women, along with all children under the age of five, were measured and weighed, and in one in two households provided a blood sample to estimate the prevalence of anemia. In addition, in one in two sampled households, all men aged 15 to 59 (usual residents or visitors) were also interviewed individually."

Source: *Centre Nationale de la Statistique et des Études Économiques - CNSEE/Congo and ICF International. 2013. Enquête Démographique et de Santé du Congo (EDSC-II) 2011-2012. Calverton, Maryland, USA: CNSEE and ICF International. Available at: https://dhsprogram.com/pubs/pdf/FR267/FR267.pdf*

**Congo Democratic Republic DHS 2013-2014**
The original report is in French. We translated the sampling strategies using Google translate.

"The objective of the EDS-RDC II is to produce representative results at the level of the whole country, at the level of urban and rural areas separately, and at the level of each of the eleven provinces. In addition, for most DHS-RDC II indicators, the results are representative at the level of each of the twenty-six new provinces, with each new province being an area of study. To do this, the national territory was divided into twenty-six areas of study and, in each area of study, three strata were created: the stratum of statutory towns, the stratum of cities and the stratum of rural areas. The sample was drawn stratum by stratum. Thus, the DHS-RDC II sample is based on an area survey, stratified at the level of primary units and drawn at several stages.

For the strata of statutory towns and cities, a number of neighborhoods were drawn, in the first degree, from the exhaustive list of all city or city neighborhoods. At the second stage, 34 households were drawn from the neighborhoods selected at the first stage. For the strata of the rural environment, the sector or chiefdom was drawn in the first degree and one or more villages in the sectors and chiefdoms drawn in the first degree in the second degree. In the third stage, 34 households were drawn in the villages drawn in the second stage. In the first stage, the selection was carried out with probability proportional to the population size of the sampling units, while in the second and third stages, a systematic selection was carried out with equal probability. The EDS-RDC II sampling frame is the general population census of 1984. It has been partially updated on several occasions by administrative censuses and recently in the context of the presidential and legislative elections of 2011. This sampling frame is the same as that used by the Multiple Indicator Cluster Surveys (MICS) of 2010 and by the 1-2-3 survey (Employment, Informal Sector and Household Consumption) of 2012, carried out by the Institute National Statistics (INS).

The final sampling unit chosen is the cluster (neighborhood or village) and a total of 540 clusters were drawn. A count of the households in each of these units provided an exhaustive list of households from which a sample of households was drawn. The cluster corresponds to the district or the village when the size of this one, in households, does not exceed 500. Beyond this number, the district or the sampled village was subdivided into several segments among which only one was. retained in the sample. In contrast, small villages have been regrouped. A total of 18,360 households (5,474 urban in 161 clusters and 12,886 rural in 379 clusters) were drawn.

All women aged 15-49 usually living in selected households, or present the night before the survey, were eligible to be surveyed. In addition, in a subsample of one in two households (see Figure 1), all men aged 15-59 were eligible to be surveyed. [...] In this same sub-sample, all women as well as all children under the age of five were eligible to be measured and weighed to assess their nutritional status."

Source: *Ministère du Plan et Suivi de la Mise en œuvre de la Révolution de la Modernité - MPSMRM/Congo, Ministère de la Santé Publique - MSP/Congo and ICF International. 2014. Enquête Démographique et de Santé en République Démocratique du Congo 2013-2014. Rockville, Maryland, USA: MPSMRM, MSP, and ICF International. Available at: https://dhsprogram.com/pubs/pdf/FR300/FR300.pdf*

**Costa Rica GATS 2015**
"A multi-stage stratified cluster design was used to obtain nationally representative data. Survey information was collected using electronic handheld devices. A total of 9,680 households were sampled, and one individual was randomly selected from each participating household to complete the survey. There were a total of 8,607 individual completed interviews."

Source: *no report available. Sampling information obtained from executive summary. Available at: https://www.who.int/tobacco/surveillance/survey/gats/cri_executive_summary_en.pdf?ua=1*

**Cote d'Ivoire DHS 2011-12**
The original report is in French. We translated the sampling strategies using Google translate.

"*Sampling*
The 2011-2012 EDS-MICS targets the population of individuals residing in ordinary households across the country. A national sample of 10,413 households was selected. The sample was stratified so as to provide an adequate representation of the urban and rural areas as well as of the eleven fields of study, corresponding to the ten former administrative regions and to the city of Abidjan, for which we have an estimate for all key indicators.

The EDS-MICS 2011-2012 sample was a two-stage stratified area sample. In the first stage, census clusters or districts (CD) were drawn throughout the national territory from the list of CDs from the 1998 General Census of Population and Housing (RGPH). the age of this database, a complete update of the clusters, was carried out. This updating made it possible to obtain a complete list of households residing in the cluster and this list served as a sampling frame to draw households in the second degree. Overall, 352 clusters, including 161 in urban areas and 191 in rural areas, were selected by carrying out a systematic draw with probability proportional to size; the size of the CD being the number of households.

In the second degree of drawing and according to the place of residence, a fixed number of households was drawn in each cluster counted by a systematic drawing with equal probability. The average number of households drawn from a cluster is 27 in urban areas and 32 in rural areas.

All women aged 15-49 usually living in selected households, or present the night before the survey, were eligible to be surveyed. In addition, in a subsample of one in two households, all men aged 15-59 were also surveyed. In this subsample, all women and men eligible for the individual survey were also eligible for the HIV test. In addition, in this sub-sample of households, all women and men eligible for the survey and all children aged 6-59 months were offered an anemia test. In addition, in this sub-sample of households, all women eligible for the survey as well as all children under the age of 5 were eligible to be measured and weighed to determine their nutritional status. Finally, in households in the same sub-sample, all children under the age of five and all pregnant women were eligible for the parasitaemia test. [...]

It appears that out of the 352 clusters selected in the 2011-2012 EDS-MICS, 351 could have been investigated, one being inaccessible. A total of 10,413 households were selected and, among them, 9,873 occupied households were identified at the time of the survey. Among these 9,873 households, 9,686 could be successfully surveyed, representing a response rate of 98% (Table 1). The household response rate is slightly higher in rural areas (99%) than in urban areas (97%).

In the 9,686 households surveyed, 10,848 women aged 15-49 were identified as being eligible for the individual survey, and for 10,060 of them, the interview was successfully conducted, representing a rate of 93% response. The men's survey was carried out in every second household: 5,677 men were eligible and 5,135 were interviewed (response rate of 91%)."

Source: *Institut National de la Statistique – Cote d'Ivoire & ICF International. Enquête Démographique et de Santé et à Indicateurs Multiples 2011-2012. Available at: https://dhsprogram.com/pubs/pdf/FR272/FR272.pdf.*

**Dominican Republic DHS 2013**
The original report is in Spanish. We translated the sampling strategies using Google translate.

"*1.2.1 Objectives of the sample design*
ENDESA 2013 was designed to obtain reliable estimates on a series of indicators relevant for the country as a whole, by urban and rural area as independent domains, and for each of the nine health regions of the country. However, depending on the indicator, analysis domains apply more restricted, especially the case of HIV prevalence. The study universes are made up of homes, women aged 15-49, their children under 5 years old, and by men 15-59 years old.

*Sample Design*
The sample design implemented by ENDESA-2013 was probabilistic, by conglomerate, stratified and bi-etipaic. The set of census supervision areas was used as a sample framework extracted from the Population and Housing Census of 2010, prepared by the National Statistics Office (ONE). These areas constitute the Primary Sampling Units (UPMs) (see Appendix A for more details). As a first stage in the design, the UPMs were stratified by province and area of residence. In this stage, 524 UPMs were selected, with probability proportional to their size (PPT). In the map 1.1 the geographical distribution of the selected UPMs is presented. These areas were subjected to a cartographic update process, in order to take into account possible changes within the same. In the second stage, 25 households were selected in each of the updated UPMs, following a random-systematic process. In each of the selected households, they were considered eligible for the survey all women aged 15 to 49 years and all men aged 15 to 59 years regulars of those homes or who slept in them the night before. For the application of the Questionnaire of Morbidity and Health Expenditures, a subsample was taken, randomly taking one in two homes already selected in the second stage. [...]

To ensure that the level of accuracy of the survey is comparable between regions, the sample considers a 'power' affixation between regions and between areas of residence within each region. On the base from a sample of 25 households within each cluster, the sample selected 524 Enumeration Areas (AE), 364 in urban areas and 160 in rural areas. The sample was selected so as to guarantee a sufficient number of cases in each domain, ranging from 1,150 households in region IV to 2,000 households in the region 0, as shown in Table A.3 of Annex A. This sample allocation would result in a number enough of women and men aged 15-49 in each region, as shown in Table A.4 of the Annex TO. It should be noted that the new health regionalization that includes the Azua province in region VI and not in region I as in ENDESA-2007. The rest of the regions did not undergo changes."

<u>Source:</u> *Centro de Estudios Sociales y Demográficos (CESDEM) & ICF International. República Dominicana Encuesta Demográfica y de Salud 2013. Published 2014. Available at:https://dhsprogram.com/publications/publication-FR292-DHS-Final-Reports.cfm*

**Ecuador STEPS 2018**
The original report is in Spanish. We translated the sampling strategies using Google translate.

"The STEPS sample design used probabilistic sampling techniques in order to guarantee the geographic and study domain representativeness of the survey, and to calculate expansion factors and errors associated with sampling. The target population or universe of study included the total of adults aged 18 to 69, disaggregated by men and women, residing in the territory of Ecuador, except Galapagos. According to the INEC population projection, it included 10,249,369 people. The observation unit and elementary unit of analysis were the people between 18 and 69 years of the Ecuadorian territory, except Galapagos.

The sampling frame for the STEPS Survey was defined from the Sampling Frame for household surveys of the National Institute of Statistics and Censuses - INEC. However, due to the scope of the inter-institutional cooperation agreement MSP-INEC, the cartographic information for the gathering of information was restricted to that used for the Population and Housing Census - CPV 2010. The delimited frame contains mainly identification variables; location variables; stratification variables; and, design and control variables of the selection units.

*Type and design stages of the sample.* The STEPS sample was selected following a probabilistic element sampling scheme with the following three selection stages: i) first stage: selection of Primary Sampling Units (PSU) by stratum; ii) second stage: selection of 12 occupied dwellings within each PSU selected in the first stage; and, iii) third stage: selection of 1 person between 18 and 69 years old per household. [...]

*Sample selection.* The selection of the PSUs, according to the established size, was carried out independently in a random manner in each of the strata. Twelve dwellings were also randomly selected from each previously selected cluster. From the second survey period, given the high rates of change in occupation, 16 dwellings were chosen per cluster, to counteract this effect. The change affected the 230 remaining conglomerates, giving a total of 6,680 homes to be surveyed. Finally, a list was made of the eligible persons within each dwelling, randomly selecting one of them. [...]

*Post-stratification.* Considering that the sample was selected from the INEC sampling frame for household surveys, which is made up mostly of information dating from 2010 and that the information gathering period is 2018, there is a substantial difference in the constitution of the study domains. In order to solve this drawback, a method called post-stratification is used. The post-stratification method allows an adjustment to the expansion factors by incorporating auxiliary information. In this case, the auxiliary information used comes from the INEC population projections, giving us the possibility of adjusting the population totals investigated to those resulting from the population projections. Therefore, two poststrates are defined: men and women between 18 and 69 years old. These coincide with the study domains of the sample."

Source: *Ministerio de Salud Pública: Encuesta STEPS Ecuador 2018. Vigilancia de enfermedades no transmisibles y factores de riesgo. INFORME DE RESULTADOS Available at: https://extranet.who.int/ncdsmicrodata/index.php/catalog/774*

**Egypt DHS 2015**
"The 2015 EHIS took advantage of the sample developed for the ever-married women survey component of the 2014 EDHS. The 2014 EDHS was implemented in a total of 842 primary sampling units (PSUs) selected from 25 governorates. The frame for selection of these units was a list of all shiakhas and villages in Egypt. This list was obtained from the Central Agency for Public Mobilization and Statistics (CAPMAS) and updated as necessary to reflect any recent changes.

For the EHIS, a sub-sample of 614 PSUs (shiakhas/villages) was selected from the 842 PSUs included in the 2014 EDHS sample. The household listing prepared during the 2014 EDHS for these PSUs was used to select the household sample for the 2015 EHIS. The selection was conducted in such a way that the EHIS household sample was totally independent of the 2014 EDHS sample, i.e., no household was included in both samples. It was expected that approximately 28,500 individuals age 6 months to 59 years, eligible for the 2015 EHIS testing and interviews, would be identified in the selected households.

The sample for the 2015 EHIS was designed to provide estimates of the key health indicators that the survey was designed to measure including the prevalence of hepatitis B and C for the country as a whole and for six major subdivisions (Urban Governorates, urban Lower Egypt, rural Lower Egypt, urban Upper Egypt, rural Upper Egypt, and Frontier Governorates). The sample also allows for estimates of some key indicators at the governorate level."

Source: *Ministry of Health and Population/Egypt, El-Zanaty and Associates/Egypt, and ICF International. 2015. Egypt Health Issues Survey 2015. Cairo, Egypt: Ministry of Health and Population/Egypt and ICF International. Available at: https://dhsprogram.com/pubs/pdf/FR313/FR313.pdf*

**Egypt DHS 2014**

"The sample for the 2014 EDHS was selected in four stages. A list of shiakhas/towns constituted the primary sampling frame for urban areas, and a list of villages served as the frame for rural areas. The Central Agency of Public Mobilization and Statistics (CAPMAS) updated these lists, which had been originally prepared for the 2006 census, to reflect the situation in 2013.

In order to provide for implicit geographic stratification, the lists of shiakhas/towns and villages in each governorate were arranged in serpentine order according to their location from north to south within the governorate. During the first stage selection, a total of 926 primary sampling units (481 shiakhas/towns and 445 villages) were chosen for the 2014 EDHS sample with probability proportional to size.

The second stage of selection involved several steps. First, for each of the primary sampling units (PSU), maps were obtained and divided into parts of roughly equal size, with the number of parts determined by dividing the population in the shiakha or village by 5,000. One to three parts were then selected systematically from each PSU, depending on the size of the shiakha or village. Three parts were selected in shiakhas/villages with a population of 100,000 or more, and two parts were selected in shiakhas/villages with populations between 20,000 and 100,000. In the remaining smaller shiakhas/villages, one part was selected.

A quick count was carried out in the selected parts in each PSU to provide the information needed to divide the parts into a number of segments of roughly equal size. Due to security issues, the quick count operation could not be undertaken in North and South Sinai, and, thus, the 42 clusters selected in those governorates were not included in the 2014 Egypt DHS. Because the populations of those governorates comprise less than 1 percent of Egypt's total population, their exclusion does not affect national estimates. However, because they comprise two of the five Frontier Governorates, information that is presented in this report for the Frontier Governorates is not comparable to results in prior EDHS surveys in which all five Frontier Governorates were surveyed.

After the quick count was completed, two to three segments were selected from each PSU. In large shiakhas/towns and villages where there were two or three parts, one segment was chosen from each part. In small shiakhas/towns and villages where only one part had been selected, two segments were chosen from that part. A household listing was obtained for each segment. Using the household lists, a systematic random sample of 29,471 households was chosen for the 2014 EDHS. During the survey, usual household members and visitors who were present in the household during the night before the survey visit were identified and listed in the household questionnaire. All ever-married women 15-49 included in that list were eligible for the individual survey interview."

Source: *Ministry of Health and Population [Egypt], El-Zanaty and Associates [Egypt], and ICF International. 2015. Egypt Demographic and Health Survey 2014. Cairo, Egypt and Rockville, Maryland, USA: Ministry of Health and Population and ICF International. Available at: https://dhsprogram.com/pubs/pdf/FR302/FR302.pdf*

**Eswatini STEPS 2014**
"A Multi-stage cluster sampling design was applied. The survey covered all the four regions of the country. The size of the country and the distances between the regions and communities made it possible for the survey to sample a population representing all the 4 regions. The Multi-stage sampling procedure was implemented in the following procedural steps:

*Stage 1:* All four regions were included as a sampling frame of our Primary Sampling Unit (PSU). The number of the PSUs at this stage ensured precision in the survey estimates and as a result 216 PSUs were selected using probability proportional to size sampling.

*Stage 2:* The second stage of cluster sampling procedure entailed listing, sorting and random systematic sampling of the Secondary Sampling Units (Households) within the PSUs selected in stage 1 where 20 households were selected from each PSU. Based on census data, only households with eligible participants were systematically sampled through random systematic sampling.

*Stage 3:* At this level, all the eligible participants within a household were sequentially listed into the PDAs and only one participant per household was randomly sampled using KISH method built into the PDAs. The KISH method is a widely used technique that uses a pre-assigned table of random numbers to identify the person to be interviewed."

Source: *Swaziland Government. Ministry of Health: WHO STEPS. Noncommunicable Disease Risk Factor Surveillance Report Swaziland 2014. Available at: http://www.who.int/chp/steps/swaziland/en/.*

**Ethiopia GATS 2016**

"The Ethiopia GATS 2016 was a household survey designed to collect nationally representative data on Ethiopians age 15 years or older. The survey used a standardized questionnaire, and multistage geographically stratified cluster sample design, data collection, and management procedures. Electronic handheld devices were used for data collection and management. One individual was randomly chosen from each selected household to participate in the survey."

Source: *no report available. Sampling information obtained from the executive summary. Available at: https://www.who.int/tobacco/surveillance/survey/gats/eth-executive-summary-2016.pdf?ua=1*

**Ethiopia DHS 2016**

"The sampling frame used for the 2016 EDHS is the Ethiopia Population and Housing Census (PHC), which was conducted in 2007 by the Ethiopia Central Statistical Agency. The census frame is a complete list of 84,915 enumeration areas (EAs) created for the 2007 PHC. An EA is a geographic area covering on average 181 households. The sampling frame contains information about the EA location, type of residence (urban or rural), and estimated number of residential households. With the exception of EAs in six zones of the Somali region, each EA has accompanying cartographic materials. These materials delineate geographic locations, boundaries, main access, and landmarks in or outside the EA that help identify the EA. In Somali, a cartographic frame was used in three zones where sketch maps delineating the EA geographic boundaries were available for each EA; in the remaining six zones, satellite image maps were used to provide a map for each EA.

Administratively, Ethiopia is divided into nine geographical regions and two administrative cities. The sample for the 2016 EDHS was designed to provide estimates of key indicators for the country as a whole, for urban and rural areas separately, and for each of the nine regions and the two administrative cities. The 2016 EDHS sample was stratified and selected in two stages. Each region was stratified into urban and rural areas, yielding 21 sampling strata. Samples of EAs were selected independently in each stratum in two stages. Implicit stratification and proportional allocation were achieved at each of the lower administrative levels by sorting the sampling frame within each sampling stratum before sample selection, according to administrative units in different levels, and by using a probability proportional to size selection at the first stage of sampling.

In the first stage, a total of 645 EAs (202 in urban areas and 443 in rural areas) were selected with probability proportional to EA size (based on the 2007 PHC) and with independent selection in each sampling stratum. A household listing operation was carried out in all of the selected EAs from September to December 2015. The resulting lists of households served as a sampling frame for the selection of households in the second stage. Some of the selected EAs were large, consisting of more than 300 households. To minimise the task of household listing, each large EA selected for the 2016 EDHS was segmented. Only one segment was selected for the survey with probability proportional to segment size. Household listing was conducted only in the selected segment; that is, a 2016 EDHS cluster is either an EA or a segment of an EA.

In the second stage of selection, a fixed number of 28 households per cluster were selected with an equal probability systematic selection from the newly created household listing. All women age 15-49 and all men age 15-59 who were either permanent residents of the selected households or visitors who stayed in the household the night before the survey were eligible to be interviewed. In half of the selected households, all women age 15-49 were eligible for the FGM/C module, and only one woman per household was selected for the domestic violence module. In all of the selected households, height and weight measurements were collected from children age 0-59 months, women age 15-49, and men age 15-59."

Source: *Central Statistical Agency/CSA/Ethiopia and ICF. 2016. Ethiopia Demographic and Health Survey 2016. Addis Ababa, Ethiopia, and Rockville, Maryland, USA: CSA and ICF. Available at: https://dhsprogram.com/pubs/pdf/FR328/FR328.pdf*

**Ethiopia STEPS 2015**

"In this study, a mix of sampling approach namely stratified, three-stage cluster sampling, simple random sampling and Kish method were employed to select the study settings and the study participants. The sampling frame was based on the population and housing census conducted for Ethiopia in 2007 (CSA, 2008). There are 11 regions including the two city administrations (Addis Ababa and Dire Dawa). Each region is divided into administrative zones. The two city administrations are divided into sub-cities. The administrative zones in the nine regions and sub-cities in the two city administrations are subdivided into districts or 'Woreda'. The districts 'Woredas' are also further divided into 'Kebele'. The kebeles are the smallest administrative units with clear geographic jurisdiction in Ethiopia. Within Kebeles, there are Enumeration Areas (EAs) which are delineated by the Central Statistical Agency (CSA) of Ethiopia. Therefore, Enumeration Areas (EAs) were considered as the primary sampling units for this survey. According to the 2007 population and housing census, there were a total of 15,837 Kebeles in Ethiopia i.e. 14,364 in rural and 1,473 in urban kebeles(12). Taking into account the cost of the study and the level of precision, 20 households per EA and one eligible individual from each household with a total of 513 EAs were covered nationwide. Stratifying the sampling design by place of residence we allocated about 404 EAs for rural and the remaining 109 to urban area. [...] As shown in the table 2.1 above, the Primary Sampling Units (PSUs) were the EAs both in rural and urban settings. At the first stage, 513 PSUs (404 rural and 109 urban) were selected with probability proportionate to size. This is followed by a random selection of secondary sampling units (SSUs) per selected PSU in the second stage. The Secondary Sampling Units (SSUs) were the households. The total number of EA in 2007 was 82,037. The Enumeration areas essentially comprise on average 100 households both in urban and rural settings. Twenty households were selected from each EA using systematic sampling. Thus, a total of 10,260 households were selected from the 513 EAs (20 households per EA). The sampling interval was determined by dividing the total number of households in the selected EA by 20. Prior to sampling, supervisors and data collectors visited the selected EAs and conducted a fresh listing of all households in that EA in consultation with local health workers and any other active member who have a good understanding of the local context. In the third stage, eligible individuals were selected from household using Kish method. Only one eligible participant (an adult age 15-69 years) in the selected household was enrolled in the survey. Using the Kish method, eligible participants in each household were ranked in order of decreasing age, starting with men followed by women."

Source: *Ethiopian Public Health Institute 2016: Ethiopia STEPS report on risk factors for non-communicable diseases and prevalence of selected NCDs. Available at: https://extranet.who.int/ncdsmicrodata/index.php/catalog/794/download/5522*

**Gabon DHS 2012**
The original report is in French. We translated the sampling strategies using Google translate.

"The 2012 EDSG-II targets the population of individuals residing in ordinary households across the country. A national sample of around 10,000 households was selected. The sample is stratified so as to provide an adequate representation of urban and rural environments as well as the ten areas of study corresponding to the nine provinces (including the Estuary without Libreville and the Ogooué Maritime without Port-Gentil) and to the cities grouped together. Libreville and Port-Gentil, for which an estimate is available for all key indicators.

The EDSG-II sample is a stratified, two-stage drawn area sample. At the first stage, clusters or Enumeration Sectors (EAs) were drawn across the entire national territory from the list of EAs established for the 2003 General Population and Housing Census (RGPH 2003). Overall, 336 clusters, including 183 in urban areas and 153 in rural areas, were selected by carrying out a systematic selection with probability proportional to size, the size of the EA being the number of households. A count of the households in each of these clusters provided a list of households from which a second-stage sample of households was drawn with a systematic drawing at equal probabilities.

All women aged 15-49 usually living in selected households, or having spent the night before the survey, were eligible to be surveyed. In addition, in a sub-sample of two out of three households, 5,880 men aged 15-59 were also surveyed. [...] In addition, in this same sub-sample, all women as well as all children under the age of five were eligible to be measured and weighed in order to assess their nutritional status."

Source: *Direction Générale de la Statistique - DGS/Gabon and ICF International. 2013. Enquête Démographique et de Santé du Gabon 2012. Calverton, Maryland, and Libreville, Gabon: DGS/Gabon and ICF International. Available at: https://dhsprogram.com/pubs/pdf/FR276/FR276.pdf*

**Gambia DHS 2013**
"The Gambia is divided into eight Local Government Areas. In turn, each LGA is subdivided into districts and each district (with the exception of Banjul) into settlements. An enumeration area (EA) is a geographic section delineated so that a team of enumerators can easily cover it during a census. In the case of The Gambia, an EA can be a settlement, a cluster of small settlements, or part of a large settlement. The 2013 GDHS sample was designed to produce reliable estimates of the most important variables for the country as a whole, for urban and rural areas, and for each of the municipalities and LGAs. The sampling frame used for the 2013 GDHS was the latest population and housing census, conducted in 2003 (census data were provided by the Gambia Bureau of Statistics). The frame excluded individuals living in collective housing units such as hotels, hospitals, work camps, prisons, and boarding schools.

The 2013 GDHS sample was a stratified sample selected in two stages. Stratification was done by dividing each LGA into urban and rural areas (except Banjul and Kanifing, which are entirely urban settlements), achieving a total of 14 sampling strata. In the first stage, 281 EAs were selected with probability proportional to size and with independent selection in each sampling stratum. These EAs constituted the primary sampling units (PSUs). After selection of the EAs and before the main fieldwork, a household listing operation was carried out in all of the selected EAs. The listing operation consisted of visiting each of the 281 selected EAs, drawing a location map and detailed sketch map, and recording on the household listing forms all structures found in the EA, as well as all residential households within these structures (including the address and name of the household head). The resulting list of households served as the sampling frame for the selection of households in the second stage of sampling.

In the second stage, 25 households per EA were selected via equal probability systematic selection. All women age 15-49 who were usual household members or who spent the night before the survey in the selected households were eligible for individual interviews. A subsample of one in every two sampled households was selected for the male survey (all men age 15-59 who were usual household members or who spent the night before the survey in the household were eligible for individual interviews) as well as for collection of blood samples for HIV, anaemia, and malaria testing."

Source: *The Gambia Bureau of Statistics - GBOS and ICF International. 2014. The Gambia Demographic and Health Survey 2013. Banjul, The Gambia: GBOS and ICF International. Available at: https://dhsprogram.com/pubs/pdf/FR289/FR289.pdf*

**Gambia STEPS 2010**
"The STEPS survey in The Gambia was a population-based survey of adults aged 25-64. Probability proportional to size sampling was used in both the first and second stage of sampling to produce representative data for that age range in The Gambia. A total of 5,280 people were targeted for the study but only a total of 4,111 participated.

*Sampling procedure:* A multi-stage cluster sample of households. One individual within the age range of the survey was selected per household. Analysis weights were calculated by taking the inverse of the probability of selection of each participant. These weights were adjusted for differences in the age-sex composition of the sample population as compared to the target population.

Different weight variables are available per Step:
- wStep1 - for interview data
- wStep2 - for physical measures

This allows for differences in the weight calculation for each Step of the survey as the age-sex composition of the respondents to each Step can differ slightly due to refusal or drop out. Additionally, some countries perform subsampling for Step 2. When no subsampling is done and response rates do not differ across Steps of the survey, the 2 weight variables will be the same."

Source: *no report available. Sampling information obtained from the fact sheet. Available at: https://extranet.who.int/ncdsmicrodata/index.php/catalog/616/related_materials. Further information from WHO NCD Microdata Repository. Available at:: https://extranet.who.int/ncdsmicrodata/index.php/catalog/616/study-description#page=sampling&tab=study-desc*

**Georgia STEPS 2016**
"The STEPS survey of non-communicable disease (NCD) risk factors in Georgia was carried out from June 2016 to September 2016. Georgia carried out Step 1, Step 2 and Step 3. Socio demographic and behavioural information was collected in Step 1. Physical measurements such as height, weight and blood pressure were collected in Step 2. Biochemical measurements were collected to assess blood glucose and cholesterol levels in Step 3. The survey was a population-based survey of adults aged 18-69. A Multi-stage cluster sampling design was used to produce representative data for that age range in Georgia. A total of 5554 adults participated in the survey. The overall response rate was 75.7%. The following equipment was used for the biological measurements: Samsung Galaxy Tab 4OS; Cardiochek PA; Growth Management Scale (330 HRS BMI); BMI calculator Seca 491; ergonomic Seca 201; test panels (Chol/HDL/Glu for Cardiocheck PA); and the blood pressure monitor Bosch Sohn Medicus UNO."

Source: *National Center for Disease Control and Public Health. Georgia STEPS Survey 2016 Fact Sheet. Available at: http://www.who.int/chp/steps/georgia/en/.*

**Ghana DHS 2014**

"The sampling frame used for the 2014 GDHS is an updated frame from the 2010 Ghana Population and Housing Census provided by the Ghana Statistical Service (GSS 2013b). The sampling frame excluded nomadic and institutional populations such as persons in hotels, barracks, and prisons. The 2014 GDHS followed a two-stage sample design and was intended to allow estimates of key indicators at the national level as well as for urban and rural areas and each of Ghana's 10 administrative regions. The first stage involved selecting sample points (clusters) consisting of enumeration areas (EAs) delineated for the 2010 PHC. A total of 427 clusters were selected, 216 in urban areas and 211 in rural areas. The second stage involved the systematic sampling of households. A household listing operation was undertaken in all the selected EAs in January-March 2014, and households to be included in the survey were randomly selected from the list. About 30 households were selected from each cluster to constitute the total sample size of 12,831 households. Because of the approximately equal sample sizes in each region, the sample is not self-weighting at the national level, and weighting factors have been added to the data file so that the results will be proportional at the national level.

All women age 15-49 who were either permanent residents of the selected households or visitors who stayed in the household the night before the survey were eligible to be interviewed and have their blood pressure measured. In half of the households, all men age 15-59 who were either permanent residents of the selected households or visitors who stayed in the households the night before the survey were eligible to be interviewed. In addition, in the subsample of households selected for the male survey:

- blood pressure measurements were performed among eligible men who consented to being tested; [...]
- height and weight information was collected from eligible women, men, and children age 0-59 months."

<u>Source:</u> *Ghana Statistical Service - GSS, Ghana Health Service - GHS, and ICF International. 2015. Ghana Demographic and Health Survey 2014. Rockville, Maryland, USA: GSS, GHS, and ICF International. Available at: https://dhsprogram.com/pubs/pdf/FR307/FR307.pdf.*

**Ghana Study on global AGEing and adult health (SAGE) 2008-2009**

"The sampling method used for the Ghana SAGE Wave 1 was based on the design for the World Health Survey, 2003, in which the primary sampling units (PSUs) were stratified by region and location (urban/rural). Selection of the PSUs was based on proportional allocation by size. Each enumeration area (EA) was selected independently within each stratum. In the WHS/SAGE Wave 0, a total of 6 000 households were to be interviewed and therefore 300 EAs were selected nationwide. Twenty households were to be randomly selected in each EA using systematic sampling. The number of EAs per region was based on the population size of the region. For SAGE Wave 1, a total of 5 000 50+ respondents and 1 000 18–49-year-old respondents were required and therefore 250 EAs out of the 298 EAs of the WHS/ SAGE Wave 0 were used based on the availability of respondents aged 50+ years within the EAs.

Enumeration areas with no 50+ individuals were not included. Within each EA, 20 households with one or more 50+ individuals and four households with members aged 18–49 were to be selected. All respondents aged 50+ within households with over 50s from the WHS were automatically selected and additional households with members aged 50+ years were randomly selected to make a total of 20 households for each EA. The four households of the 18–49 years age group were randomly selected from the WHS/SAGE Wave 0 households list per EA. All the 50+ year olds within the selected households were to be interviewed together with the four identified under-50 respondents. Field work and data entry were undertaken between May 2007 and June 2008."

Source: *Biritwum R, Mensah G, Yawson A, Minicuci N. Study on global AGEing and adult health (SAGE) -Wave 1: The Ghana National Report. University of Ghana Medical School, Department of Community Health. July 2013.*

**Guatemala DHS 2014-2015**
The original report is in Spanish. We translated the sampling strategies using Google translate.

"Guatemala is divided into 22 departments that constitute the main domains of study and analysis of the survey. Each department is divided into rural and urban strata, although these are considered as domains of analysis only at the national level. The sampling frame of the survey is based on the INE master sample, drawn from the data and cartographic material of the 2002 census, the most recent.

Since the objectives of the survey are similar to those of the ENSMI 2008-2009, the initial budget was based on the selection of a sample of around 22,000 households. As the results of the survey would be tabulated at the departmental level, it was decided to select a minimum of 900 households in each department. In the ENSMI 2008-2009, 30 households were selected in each sector of the sample, but in this survey the number was reduced to 26 households and the number of sectors of the sample of each department was increased to make the sample more efficient. For each department, except Guatemala, the number of sectors in the sample was increased to 36, distributed proportionally in the urban and rural strata of the department. With the selection of 26 households for each sector of the sample, a sample of 936 households resulted for the majority of departments, which is slightly larger than the ENSMI 2008-2009 sample (900 households). For the department of Guatemala the number of sectors was increased to 48 for the municipality of Guatemala and 60 for the rest of the department, with a corresponding increase in the number of households in the sample to 1,248 and 1,560 respectively.

A multi-stage stratified selection procedure was used with the sectors as primary units. In the first stage, the INE selected the sectors of the master sample in a systematic way and with proportional probabilities to its size within each department, rural and urban area, and socioeconomic level (NSE). For the second stage, the 4,020 sectors of the INE master sample were explicitly stratified by department and rural and urban area, and an implicit stratification was used by socio-economic level. Given the sample size of the ENSMI 2014-2015 (864 sectors), it was not possible to consider these three stratification variables and only the first two variables (area and department) were considered. To somehow include stratification by socioeconomic stratum (NSE), before making the selection of the sectors of the sample, within each area and department the sectors were ordered according to this variable by NSE, and then a procedure was applied of systematic sampling with equal probability of selection. In each of the 864 sectors selected, the list of their homes and the cartographic material that place them were updated. Then, in the last stage of sampling, this information was used to systematically and with equal probability select 26 households from the updated list of households for interviews of women aged 15 to 49 years. Finally, one of every two households (13 households) were randomly and systematically selected from these households, where interviews of men aged 15 to 59 years were conducted. These then constitute a subsample of households of half of the total selected households.

In order for the estimates of the survey to be representative of the entire population and its different strata, a weighting of the data was necessary. Weights were calculated for the total number of households surveyed, which were adjusted according to the response rate. Specific weights were also calculated for the subsample of households in which the men's interviews were conducted. For the data collected in the women's questionnaire specific weights were calculated based on household weights, according to the response rate of women. In the case of men, specific weights were calculated that consider the total household weights, the subsample household weights and the response rate of men. […]

Table 1.2 shows the results of the sample implementation for households and eligible people. It is observed that, of a total of 22,308 selected households, 21,662 were employed and of these 21,383 were interviewed successfully, which gives a response rate of 99 percent. In this group of households interviewed, 26,767 eligible women were found, of which 97 percent were interviewed. In the case of men, interviews were conducted in one in two households, 11,930 were found eligible, 93 percent of them were interviewed."

Source: *Ministerio de Salud Pública y Asistencia Social – MSPAS/Guatemala, Instituto Nacional de Estadística – INE/Guatemala, Secretarío de Planificación y Programación del la Presidencia & ICF international. VI Encuesta Nacional de Salud Materno Infantil 2014-2015. Available at: https://dhsprogram.com/pubs/pdf/FR318/FR318.pdf.*

**Guinea DHS 2018**
The original report is in French. We translated the sampling strategies using Google translate.

"The EDSV 2018 aims to produce representative results at the level of the whole country, for the city of Conakry, for the Other Cities, for the urban and rural areas, and at the level of the administrative regions of Guinea. To do this, the national territory was divided into 8 study areas corresponding to the 7 administrative regions and the Capital Conakry. In each area of study (except the city of Conakry which has no rural part), two strata were created: the urban strata and the rural strata. A total of 15 sampling strata were created.

The first stage sample will be drawn independently in each stratum, and the second stage sample will be drawn independently in each primary unit drawn at the first stage. The sampling frame retained for the EDSV 2018 is the database of the General Housing Population Census which took place in 2014 (RGPH, 2014). The National Institute of Statistics (INS) has a computer file containing 9,679 ZD. In this file, each EA appears with its identifiers (region, prefecture, sub-prefecture, and identification code), its size in households and inhabitants, and its type of place of residence (urban or rural). Each EA has a base map that shows the boundaries of the EA, the names of localities belonging to the EA, and the access roads. The average size of EAs is households in rural areas, with an overall average of 153 households per EA.

The 2018 EDSV sample was drawn, stratum by stratum, by a stratified and 2-degree area survey. By deciding to select 20 households per cluster, in the first degree, 401 EAs were drawn with a probability proportional to the size, the size being the number of households residing in the EA. Among the 401 ZD drawn, 138 are located in urban areas and 263 in rural areas. Table 1.1 shows the results of the interviews. Among the 8,020 households drawn, 2,760 are from urban areas and 5,260 are from rural areas. Before drawing the first stage EAs, the sampling frame was sorted according to the administrative units within each stratum, that is to say, prefecture and sub-prefecture, then the number of the EA. This operation will introduce an implicit stratification at the level of all administrative units below the region with a sample allocation proportional to their size. This will provide better representation of the sample at the level of each region and at the national level.

After the selection of the primary units and before the main survey, a household enumeration and an update of the map were carried out in each selected EA. This operation made it possible to obtain a complete list of occupied households in each EA drawn and which served as a basis for the selection of second-degree households. Then, at the second stage, a sample of 20 households was drawn by cluster in both urban and rural areas with a systematic selection at equal probability."

Source: *Institut National de la Statistique/INS et ICF. 2018. Enquête Démographique et de Santé en Guinée 2018. Conakry, Guinée, et Rockville, Maryland, USA : INS et ICF. Available at: https://dhsprogram.com/pubs/pdf/FR353/FR353.pdf*

**Haiti DHS 2017**
The original report is in French. We translated the sampling strategies using Google translate.

"A national sample of 13,546 households was selected, of which 13,405 were interviewed. The sample is stratified so as to provide an adequate representation of the urban and rural areas as well as of the 11 fields of study, corresponding to the 10 departments, and to the Metropolitan Area. The EMMUS-VI (Sixth Mortality, Morbidity and Service Use Survey) sample is a stratified area sample, drawn in two stages. In the first stage, 450 Enumeration Sections (SDE) or clusters were drawn across the national territory, with a systematic drawing with probability proportional to the size. The size of an SDE is the number of households residing in the SDE. The sampling frame used for EMMUS-VI is the 2003 General Population and Housing Census (RGPH 2003), with a partial update in 2011 by the IHSI. After the EDS was drawn and before the main survey, a household count and an update of the map of each selected EDS were carried out. In the second stage, a sample of households was drawn with a systematic drawing with equal probability from the newly established list in the enumeration. All women aged 15-49 usually living in selected households, or present the night before the survey, were eligible to be surveyed. In two thirds of households, the survey was also carried out among men aged 15-64. In a third of households, women aged 50-64 and men aged 35-64 were also eligible, but only for certain aspects of the survey."

Source: *Enquête Mortalité, Morbidité et Utilisation des Services - EMMUS-VI 2016-2017 Pétion-Ville, Haïti, et Rockville, Maryland, USA : IHE et ICF. Available at: https://dhsprogram.com/pubs/pdf/FR326/FR326.pdf*

**Honduras DHS 2011-2012**
The original report is in Spanish. We translated the sampling strategies using Google translate.

“For the design and selection of the ENDESA 2011-2012 sample, a computerized archive of the sectors and segments for each department prepared with information from the last National Census of Population and Housing of the year 2001. Each segment and / or sector has a complete identification that corresponds to the department and municipality codes where they are located. Also took I finished a cartographic survey of each unit, which allowed a quick update of homes in the segments that were selected for the sample. Sample design provides a sample probabilistic nationwide, which is stratified and two-stages. Within each department the sectors censuses were stratified in areas of urban and rural residence, and additionally in each department They were stratified according to their municipalities.

With the objective of selecting a total of 23,000 households, survey information was used ENDESA 2005-2006, as an aid instrument to estimate certain design parameters. With this Information was estimated that there would be a total of approximately 23,468 full interviews of women. The total of selected households was distributed by departments but not proportionally to their size population but to be able to meet the required objectives of the survey. The final number of households is adjusted to have a minimum of 1,100 homes per department except for Bay Islands and Thanks to God, in which a size of 600 homes was determined. As the allocation of households by department does not it was made proportionally to its corresponding percentage of population within the country, the total sample it is not self-weighted and therefore weighting factors were needed in the final elaboration of the tabulations These factors carry the sample not proportional to the estimated participation of the departments within the total population. The units of analysis are women of childbearing age from 15 to 49 years and children under five years; and men of childbearing age 15 to 59 years located in the households interviewed. [...]

The ENDESA 2011-2012 sample was designed to provide analysis at different levels of geographical disaggregation, such as the area of residence of the population (urban or rural); of the regions Departmental health and metropolitan areas of the departments of Cortés and Francisco Morazán. The Population universe for the individual women questionnaire of ENDESA 2011-2012 was defined as the Female population 15 to 49 years of age from across the country. The population interviewed with the individual questionnaire was identified in 21,362 selected occupied households, where the household questionnaire and the men's questionnaire were also applied in a subsample.”

Source: *Secretaría de Salud, Instituto Nacional de Estadística & ICF International. Honduras Encuesta Nacional de Salud y Demografía y Salud - ENDESA 2011-2012. Available at: https://dhsprogram.com/pubs/pdf/FR274/FR274.pdf.*

**India GATS 2016-2017**
"A multi-stage sample design was used for both rounds of GATS. From each of the sampled households, one household member 15 years of age or older was randomly selected for an individual interview."

Source: *no report available. Sampling information obtained from the fact sheet. Available at: https://www.who.int/tobacco/surveillance/survey/gats/GATS_India_2016-17_FactSheet.pdf?ua=1*

**India DHS 2015-2016**
"Decisions about the overall sample size required for NFHS-4 were guided by several considerations, paramount among which was the need to produce indicators at the district, state/union territory (UT), and national levels, as well as separate estimates for urban and rural areas in the 157 districts that have 30-70 percent of the population living in urban areas as per the 2011 census, with a reasonable level of precision. In addition, the NFHS-4 sample was designed to be able to produce separate estimates for slum and non-slum areas in eight cities (Chennai, Delhi, Hyderabad, Indore, Kolkata, Meerut, Mumbai, and Nagpur), and to provide general population estimates of HIV prevalence for women and men for India as a whole, for urban and rural areas of India, and for 11 groups of states/union territories. [...] To achieve a representative sample of 15 percent of households, NFHS-4 conducted interviews in every alternate selected household in 30 percent of the selected clusters. In all, 28,586 Primary Sampling Units (PSUs) were selected across the country in NFHS-4, of which fieldwork was completed in 28,522 clusters. The NFHS-4 sample is a stratified two-stage sample. The 2011 census served as the sampling frame for the selection of PSUs. PSUs were villages in rural areas and Census Enumeration Blocks (CEBs) in urban areas. PSUs with fewer than 40 households were linked to the nearest PSU. Within each rural stratum, villages were selected from the sampling frame with probability proportional to size (PPS). In each stratum, six approximately equal substrata were created by crossing three substrata, each created based on the estimated number of households in each village, with two substrata, each created based on the percentage of the population belonging to scheduled castes and scheduled tribes (SCs/STs).

Within each explicit sampling stratum, PSUs were sorted according to the literacy rate of women age 6+ years. The final sample PSUs were selected with PPS sampling. In urban areas, CEB information was obtained from the Office of the Registrar General and Census Commissioner, New Delhi. CEBs were sorted according to the percentage of the SC/ST population in each CEB, and sample CEBs were selected with PPS sampling. In every selected rural and urban PSU, a complete household mapping and listing operation was conducted prior to the main survey. Selected PSUs with an estimated number of at least 300 households were segmented into segments of approximately 100-150 households. Two of the segments were randomly selected for the survey using systematic sampling with probability proportional to segment size. Therefore, an NFHS-4 cluster is either a PSU or a segment of a PSU. In the second stage, in every selected rural and urban cluster, 22 households were randomly selected with systematic sampling."

Source: *International Institute for Population Sciences & ICF International. National Family and Health Survey (NFHS-4) 2015-16. Available at: https://dhsprogram.com/pubs/pdf/FR339/FR339.pdf.*

**Indonesia Family Life Survey 2014-2015**
"Because it is a longitudinal survey, IFLS5 drew its sample from IFLS1, IFLS2, IFLS2+, IFLS3 and IFLS4. The IFLS1 sampling scheme stratified on provinces and urban/rural location, then randomly sampled within these strata (see Frankenberg and Karoly, 1995, for a detailed description). Provinces were selected to maximize representation of the population, capture the cultural and socioeconomic diversity of Indonesia, and be cost-effective to survey given the size and terrain of the country. For mainly cost-effectiveness reasons, 14 of the then existing 27 provinces were excluded. The resulting sample included 13 of Indonesia's 27 provinces containing 83% of the population: four provinces on Sumatra (North Sumatra, West Sumatra, South Sumatra, and Lampung), all five of the Javanese provinces (DKI Jakarta, West Java, Central Java, DI Yogyakarta, and East Java), and four provinces covering the remaining major island groups (Bali, West Nusa Tenggara, South Kalimantan, and South Sulawesi).

Within each of the 13 provinces, enumeration areas (EAs) were randomly chosen from a nationally representative sample frame used in the 1993 SUSENAS, a socioeconomic survey of about 60,000 households. The IFLS randomly selected 321 enumeration areas in the 13 provinces, over-sampling urban EAs and EAs in smaller provinces to facilitate urban-rural and Javanese–non-Javanese comparisons.

Within a selected EA, households were randomly selected based upon 1993 SUSENAS listings obtained from regional BPS office. A household was defined as a group of people whose members reside in the same dwelling and share food from the same cooking pot (the standard BPS definition). Twenty households were selected from each urban EA, and 30 households were selected from each rural EA. This strategy minimized expensive travel between rural EAs while balancing the costs of correlations among households. For IFLS1 a total of 7,730 households were sampled to obtain a final sample size goal of 7,000 completed households. This strategy was based on BPS experience of about 90% completion rates. In fact, IFLS1 exceeded that target and interviews were conducted with 7,224 households in late 1993 and early 1994. In IFLS1 it was determined to be too costly to interview all household members, so a sampling scheme was used to randomly select several members within a household to provide detailed individual information."

Source: *Strauss, J, Witoelar F, and Sikoki, B. The Fifth Wave of the Indonesia Family Life Survey (IFLS5): Overview and Field Report. March 2016. WR-1143/1-NIA/NICHD. Available at: https://www.rand.org/content/dam/rand/pubs/working_papers/WR1100/WR1143z1/RAND_WR1143z1.pdf*

**Iran STEPS 2016**
"The sampling part, which includes determining the sample size and the cluster head, belongs to the pre-study phase and was planned in the form of a specific protocol for sample size and statistical sampling. All experts in the quality control team supervised the finding of samples and cluster heads.
In order to estimate the prevalence rate of the risk factors for non-communicable diseases in the country in 1395, a sampling method proportionate to the population was used, which is a common approach in survey studies. Therefore, the selected sample size was proportionated to the population of that province. On the other hand, for estimating the prevalence of the risk factors in the province, in order to be on the safe side, the smallest sample size for achieving the predicted rates was calculated at 95%. This rate was equal to 384 samples, which was selected as the smallest sample size in the least populated province, Ilam. The required sample size for other provinces was therefore calculated according to the population of that province proportionate to the population of the reference province, Ilam. Besides, to control the non-response error, 10% was added to the calculated sample size in each province.
In order to decrease costs and increase efficiency, for provinces with 800 samples or more, weights were given to their samples. Weight-giving is an effective method used in surveys in order to decrease the sample size. This was achieved in the selected provinces by considering the calculated sample size as half and the sampling weight as double. The total sample size was calculated to be 30150 and to achieve this sample size, sampling from 3015 clusters was required."

Source: *Non Communicable Diseases Research Center, Endocrinology and Metabolism Research Institute of Tehran University of Medical Sciences: Atlas of non-communicable diseases risk-factors surveillance in the islamic republic of Iran. STEPs 2016.*
*Available at: https://www.who.int/ncds/surveillance/steps/STEPS_2016_Atlas_EN.pdf?ua=1*

**Jordan STEPS 2019**

"3.4.1 Sampling of Jordanians

A national cross-sectional survey was conducted adopting a two-stage stratified-cluster sampling design. The margin error was (5%) and the confidence level was set at 95%. The Jordan Population and Housing Census 2015 was used as a sampling frame for Jordanians. A sample of 3000 households was randomly drawn to represent the Jordanian population. It was designed in a probability proportional to size (PPS) way to provide valid and reliable survey estimates across the entire Kingdom of Jordan - rural and urban areas, the twelve governorates and the smaller communities within. The sample also ensured reliable estimates in terms of geographical distribution, where Jordan was divided into three regions; north, centre, and south, also at governorate level. The north of Jordan covered Ajloun, Irbid, Jerash, and Mafraq, the centre region covered Amman, Balqa, Madaba, and Zarqa, and the south region covered Aqaba, Karak, Ma'an, and Tafieleh. Furthermore, each governorate was subdivided into area units called census blocks, which were the Primary Sampling Units (PSU-Blocks) for this survey (on average a PSU comprises 50-70 households). The PSU-Blocks were then regrouped to form clusters. From each PSU, eight households were randomly drawn with an equal probability systematic selection. A household was defined as a group of people living in the same dwelling space who eat meals together, acknowledging the authority of a man or a woman as the head of the household. After the household selection and obtaining the permission of household residents to participate in the survey, all the eligible household members were entered into the STEPs program, which ran a random selection to choose one member from each household. The sample size from each governorate was proportionate to the population's size

3.4.2 Sampling of Syrians

Sampling of Syrian refugees took place in four governorates; Amman, Zarqa, Irbid and Mafraq, where over than 90% of Syrian refugees reside in Jordan. The sample size was proportional to the population size in each of the governorate, i.e., the bigger the population, the bigger the sample size selected (table, 3). To update the sampling frame for Syrians, a prelisting of households took place where Syrian households were visited and entered a listing frame (the same DOS listing questionnaire was used). This sampling frame enabled weight calculation and the selection of the final Syrians sample. This was followed by determining the sampling blocks (PSU) which were 375, then from each block, 8 households were randomly selected to fulfill the pre-calculated sample size of 3000."

Source: *Centre of Strategic Studies - University of Jordan, Jordan Ministry of Health, World Health Organization: Jordan National Stepwise Survey (STEPs) for Noncommunicable Diseases Risk Factors 2019. STEPs Technical Report. Available at: https://extranet.who.int/ncdsmicrodata/index.php/catalog/853/download/6010*

**Jordan DHS 2017-2018**

"The sampling frame used for the 2017-18 JPFHS is based on Jordan's Population and Housing Census (JPHC) frame for 2015. The current survey is designed to produce results representative of the country as a whole, of urban and rural areas separately, of three regions, of 12 administrative governorates, and of three national groups: Jordanians, Syrians, and a group combined from various other nationalities.

The 12 governorates are grouped into three regions—the North region (Irbid, Jarash, Ajloun, and Mafraq), the Central region (Amman, Zarqa, Balqa, and Madaba), and the South region (Karak, Tafiela, Ma'an, and Aqaba). Each of the governorates of Jordan is divided into progressively smaller districts, sub-districts, localities, areas, and sub-areas. In addition to these administrative units, in the recent population census each sub-area was divided into convenient area units called census blocks. An electronic file contains a complete list of all census blocks, with information on households, populations, and geographical locations of each block. The census blocks are regrouped to form a general statistical unit of moderate size, called a cluster, which is widely used in various surveys as the primary sampling unit (PSU).

The sample for the 2017-18 JPFHS is a stratified sample selected in two stages from the 2015 census frame. Stratification was achieved by separating each governorate into urban and rural areas. Each of the Syrian camps in the governorates of Zarqa and Mafraq formed its own sampling stratum. In total, 26 sampling strata were constructed. Samples were selected independently in each sampling stratum, through a two-stage selection process, according to the sample allocation. Before the sample selection, the sampling frame was sorted by district and sub-district within each sampling stratum. By using a probability-proportional-to-size selection for the first stage of selection, an implicit stratification and proportional allocation were achieved at each of the lower administrative levels.

In the first stage, 970 clusters were selected with probability proportional to cluster size, with the cluster size being the number of residential households enumerated in the 2015 JPHC. The sample allocation took into account the precision consideration at the governorate level and at the level of each of the three special domains. After selection of PSUs and clusters, a household listing operation was carried out in all selected clusters. The resulting household lists served as the sampling frame for selecting households in the second stage. A fixed number of 20 households per cluster were selected with an equal probability systematic selection from the newly created household listing.

All ever-married women age 15-49 who were either residents of the selected households or visitors who stayed in the households the night before the survey were eligible for an interview (Figure 1.1). [...] Also, in one in every two of the households in this subsample (i.e., 25% of all households), all men age 15-59 who were usual residents of the selected households or who stayed in the households the night before the survey were eligible for an interview."

Source: *Department of Statistics/DOS and ICF. 2019. Jordan Population and Family and Health Survey 2017-18. Amman, Jordan, and Rockville, Maryland, USA: DOS and ICF. Available at: https://dhsprogram.com/pubs/pdf/FR346/FR346.pdf*

**Kenya STEPS 2015**
"The 2015 Kenya STEPs survey was a national cross-sectional household survey designed to provide estimates for indicators on risk factors for non-communicable diseases for persons age 18 – 69 years. The sample was designed with a sample size of 6,000 individuals to allow national estimates by sex (male and female) and residence (urban and rural areas). The survey used the fifth National Sample Surveys and Evaluation Programme (NASSEP V) master sample frame that was developed and maintained by KNBS. The frame was developed using the Enumeration Areas (EAs) generated from the 2009 Kenya Population and Housing Census to form 5,360 clusters split into four equal sub-samples. A three-stage cluster sample design was adopted for the survey involving selection of clusters, households and eligible individuals. In the first stage, 200 clusters (100 urban and 100 rural) were selected from one sub-sample of NASSEP V frame. A uniform sample of 30 households from the listed households in each cluster was selected in the second stage of sampling. The last stage of sampling was done using Personal Digital Assistants (PDAs) at the time of survey, where one individual was randomly selected from all eligible listed household members using a programmed KISH method of sampling."

Source: *Ministry of Health Kenya, Division of Noncommunicable Diseases, Kenya National Bureau of Statistics, & World Health Organization. WHO: Kenya STEPwise Survey for Non Communicable Diseases Risk Factors 2015 Report. Available at: http://www.who.int/chp/steps/Kenya_2015_STEPS_Report.pdf?ua=1.*

**Kenya DHS 2014**

"The sample for the 2014 KDHS was drawn from a master sampling frame, the Fifth National Sample Survey and Evaluation Programme (NASSEP V). This is a frame that the KNBS currently operates to conduct household-based surveys throughout Kenya. Development of the frame began in 2012, and it contains a total of 5,360 clusters split into four equal subsamples. These clusters were drawn with a stratified probability proportional to size sampling methodology from 96,251 enumeration areas (EAs) in the 2009 Kenya Population and Housing Census. The 2014 KDHS used two subsamples of the NASSEP V frame that were developed in 2013. Approximately half of the clusters in these two subsamples were updated between November 2013 and September 2014. Kenya is divided into 47 counties that serve as devolved units of administration, created in the new constitution of 2010. During the development of the NASSEP V, each of the 47 counties was stratified into urban and rural strata; since Nairobi county and Mombasa county have only urban areas, the resulting total was 92 sampling strata.

The 2014 KDHS was designed to produce representative estimates for most of the survey indicators at the national level, for urban and rural areas separately, at the regional (former provincial1) level, and for selected indicators at the county level. In order to meet these objectives, the sample was designed to have 40,300 households from 1,612 clusters spread across the country, with 995 clusters in rural areas and 617 in urban areas. Samples were selected independently in each sampling stratum, using a two-stage sample design. In the first stage, the 1,612 EAs were selected with equal probability from the NASSEP V frame. The households from listing operations served as the sampling frame for the second stage of selection, in which 25 households were selected from each cluster.

The interviewers visited only the preselected households, and no replacement of the preselected households was allowed during data collection. The Household Questionnaire and the Woman's Questionnaire were administered in all households, while the Man's Questionnaire was administered in every second household. Because of the non-proportional allocation to the sampling strata and the fixed sample size per cluster, the survey was not self-weighting. The resulting data have, therefore, been weighted to be representative at the national, regional, and county levels."

Source: *Kenya National Bureau of Statistics, Ministry of Health/Kenya, National AIDS Control Council/Kenya, Kenya Medical Research Institute, National Council for Population and Development/Kenya, and ICF International. 2015. Kenya Demographic and Health Survey 2014. Rockville, MD, USA: Kenya National Bureau of Statistics, Ministry of Health/Kenya, National AIDS Control Council/Kenya, Kenya Medical Research Institute, National Council for Population and Development/Kenya, and ICF International. Available at: https://dhsprogram.com/pubs/pdf/FR308/FR308.pdf*

**Kiribati STEPS 2015**
"The second Kiribati STEPS Survey was a population-based survey of 18-69 year olds. The decision was to use three age groups: 18-29, 30-44, 45-69 years for men and women using the following corrections:

• Design Effect of 1.0 (clustering at village and household level)

• 95% confidence interval; p value .05

• 0.7% response rate

• Baseline prevalence percentage indicator: 0.5

• FPC – not applicable

• 6 age-sex groups (18-29 years, 30-44 years, 45-69 years)

As STEPS is intended to be nationally representative, a multi-stage cluster sampling method was used. The STEPS sampling spreadsheet was completed using the most recent census information (2012). The sample was selected in two stages assuming no replacement. At the first stage, a sample of Enumeration Areas (Islands and villages) from each stratum using probability proportional to size (PPS) sampling was selected. In the second stage, a fixed number of households from each selected Enumeration Area using systematic sampling was selected. The third stage of sampling selection was done at the household level using the KISH method.

The sampling identified that data collection would be needed on the following islands: Makin, Butaritari, Marakei, Abaiang, North Tarawa, South Tarawa,Betio, Maiana, Abemama, Kuria, Aranuka, Nonouti, Tabiteuea North, Tabiteuea South, Arorae, Tabuaeran and Kiritimati. Further details in Annex 3."

Source: *Ministry of Health and Medical Services and WHO Western Pacific Region: Kiribati NCD risk factors STEPS report 2015-2016 - in collaboration with the WHO. Available at: https://extranet.who.int/ncdsmicrodata/index.php/catalog/724*

**Kyrgyz Republic STEPS 2013**
*"Sampling procedure:* A multi-stage cluster sample of households. One individual within the age range of the survey was selected per household.
Analysis weights were calculated by taking the inverse of the probability of selection of each participant. These weights were adjusted for differences in the age-sex composition of the sample population as compared to the target population. Different weight variables are available per Step:

wStep1 - for interview data
wStep2 - for physical measures
wStep3 - for biochemical measures

This allows for differences in the weight calculation for each Step of the survey as the age-sex composition of the respondents to each Step can differ slightly due to refusal or drop out."

Source: *no report or fact sheet available. Sampling information obtained from WHO NCD Microdata Repository. Available at: https://extranet.who.int/ncdsmicrodata/index.php/catalog/271/study-description#page=overview&tab=study-desc*

**Kyrgyz Republic DHS 2012**

"A representative probability sample of 8,216 households was selected for the 2012 KgDHS. The sample was selected in two stages. In the first stage, 316 clusters were selected from the sampling frame, which is a complete list of enumeration areas created for the 2009 Population and Housing Census. In the second stage, before the main survey, a household listing operation was conducted in each of the selected clusters. Households were then randomly selected from the newly updated listing for participation in the survey.

All women age 15-49 who either were permanent residents of the households in the 2012 KgDHS sample or were visitors present in the household on the night before the survey were eligible to be interviewed. Interviews were completed with 8,208 women. In addition, in a subsample of one-third of the households selected for the survey, all men age 15-49 were eligible to be interviewed if they were either permanent residents or visitors present in the household on the night before the survey. Interviews were completed with 2,413 men."

Source: *National Statistical Committee of the Kyrgyz Republic (NSC), Ministry of Health [Kyrgyz Republic], and ICF International. 2013. Kyrgyz Republic Demographic and Health Survey 2012. Bishkek, Kyrgyz Republic, and Calverton, Maryland, USA: NSC, MOH, and ICF International. Available at: https://dhsprogram.com/pubs/pdf/FR283/FR283.pdf*

**Lao PDR STEPS 2013**
*"Sampling procedure:* A multi-stage cluster sample of households. One individual within the age range of the survey was selected per household.
Analysis weights were calculated by taking the inverse of the probability of selection of each participant. These weights were adjusted for differences in the age-sex composition of the sample population as compared to the target population. Different weight variables are available per Step:

- wStep1 - for interview data
- wStep2 - for physical measures
- wStep3 - for biochemical measures

This allows for differences in the weight calculation for each Step of the survey as the age-sex composition of the respondents to each Step can differ slightly due to refusal or drop out. Additionally, some countries perform subsampling for Step 2 and/or Step 3. When no subsampling is done and response rates do not differ across Steps of the survey, the 3 weight variables will be the same."

Source: *no report or fact sheet available. Sampling information obtained from WHO NCD Microdata Repository. Available at:: https://extranet.who.int/ncdsmicrodata/index.php/catalog/588/study-description#page=sampling&tab=study-desc*

**Lesotho DHS 2014**

"The sampling frame used for the 2014 LDHS is an updated frame from the 2006 Lesotho Population and Housing Census (PHC) provided by the Lesotho Bureau of Statistics (BOS). The sampling frame excluded nomadic and institutional populations such as persons in hotels, barracks, and prisons. The 2014 LDHS followed a two-stage sample design and was intended to allow estimates of key indicators at the national level as well as in urban and rural areas, four ecological zones,[1] and each of Lesotho's 10 districts. The first stage involved selecting sample points (clusters) consisting of enumeration areas (EAs) delineated for the 2006 PHC. A total of 400 clusters were selected, 118 in urban areas and 282 in rural areas. The second stage involved systematic sampling of households. A household listing operation was undertaken in all of the selected EAs in July 2014, and households to be included in the survey were randomly selected from these lists. About 25 households were selected from each sample point, for a total sample size of 9,942 households. Because of the approximately equal sample sizes in each district, the sample is not self-weighting at the national level, and weighting factors have been added to the data file so that the results will be proportional at the national level. All women age 15-49 who were either permanent residents of the selected households or visitors who stayed in the household the night before the survey were eligible to be interviewed. In half of the households, all men age 15-59 who were either permanent residents of the selected households or visitors who stayed in the household the night before the survey were eligible to be interviewed. In the subsample of households selected for the male survey, blood pressure measurements and anaemia testing were performed among eligible women and men who consented to being tested. With the parent's or guardian's consent, children age 6-59 months were also tested for anaemia. In the same subsample of households, blood specimens were collected for laboratory testing of HIV from eligible women and men who consented; height and weight were measured for eligible women, men, and children age 0-59 months; and mid-upper-arm circumference (MUAC) measurements were collected for children age 6-59 months."

Footnotes: "1 Lowlands, Foothills, Mountains, and Senqu River Valley. 2 Butha-Buthe, Leribe, Berea, Maseru, Mafeteng, Mohale's Hoek, Quthing, Qacha's Nek, Mokhotlong, and Thaba-Tseka. 3 One rural EA was inadvertently dropped from the sample. After the fieldwork was completed, it was determined that the EA had not been visited."

Source: *Ministry of Health, The DHS Program & ICF International. 2016. Lesotho Demographic and Health Survey 2014. Available at: https://www.dhsprogram.com/pubs/pdf/FR309/FR309.pdf.*

**Liberia STEPS 2011**
"Random multi-cluster sampling method was used to collect data during this survey in 5 of the 15 counties of Liberia with the district serving as the primary sampling unit. Different sampling frames were designed and used at the district (Primary Sampling Unit-PSU), Chiefdoms (Secondary Sampling Unit-SSU) and household levels. Households listing generated from the 2008 National Population Census was used, and in each household, the list of individuals' resident was obtained and the Kish Method was used. Kish Method is a household sampling technique developed by WHO for STEPS. The field team selected households by using nutrition sampling method (throwing a pencil to get a selected direction). When the household enumeration sampling point is established, the interviewer counts all the households and using interval sample to get the household number. In each household, one person was selected using the Kish method."

Source: *Ministry of Health and Social Welfare: Liberia Chronic Disease Risk Factor Surveillance. Available at: http://www.who.int/chp/steps/Liberia_2011_STEPS_Report.pdf?ua=1.*

**Madagascar DHS 2008-2009**
The original report is in French. We translated the sampling strategies using Google translate.

"The fourth Madagascar Demographic and Health Survey (EDSMD-IV) targets a national sample of approximately 18,000 women aged 15 to 49 and 9,000 men aged 15 to 59. The EDSMD-IV sample is an area sample, stratified and drawn at 2 degrees. The primary sampling unit is the cluster or enumeration area (EA) as defined during the census mapping activities carried out in 2008 in preparation for the next General Population and Housing Census. Each of the 22 regions was divided into urban and rural parts to form, with the city of Antananarivo, 45 strata. The sample was drawn independently from each stratum. In the first degree, 600 clusters were drawn with a probability proportional to the size. Size is the estimated number of households in the cluster during the census mapping operation. A count of the households in each selected cluster yielded a list of households, from which the sample households were drawn in the second stage. Each woman, aged 15 to 49, identified in the households surveyed was also surveyed using the Woman questionnaire. In a sub-sample of 50% of the selected households, all men aged 15 to 59 were also interviewed. In addition, it was also in this subsample of households that anthropometric measurements were taken and a blood sample was taken for hemoglobin and syphilis tests."

Source: *Institut National de la Statistique - INSTAT/Madagascar and ICF Macro. 2010. Enquête Démographique et de Santé de Madagascar 2008-2009. Antananarivo, Madagascar: INSTAT and ICF Macro. Available at: https://dhsprogram.com/pubs/pdf/FR236/FR236.pdf*

**Malawi DHS 2015-2016**

"The sampling frame used for the 2015-16 MDHS is the frame of the Malawi Population and Housing Census (MPHC), conducted in Malawi in 2008, and provided by the Malawi National Statistical Office (NSO). The census frame is a complete list of all census standard enumeration areas (SEAs) created for the 2008 MPHC. A SEA is a geographic area that covers an average of 235 households. The sampling frame contains information about the SEA location, type of residence (urban or rural), and the estimated number of residential households. Administratively, Malawi is divided into 28 districts. The sample for the 2015-16 MDHS was designed to provide estimates of key indicators for the country as a whole, for urban and rural areas separately, and for each of the 28 districts. Indicators will also be shown for the Northern, Central, and Southern regions of the country.

The 2015-16 MDHS sample was stratified and selected in two stages. Each district was stratified into urban and rural areas; this yielded 56 sampling strata. Samples of SEAs were selected independently in each stratum in two stages. Implicit stratification and proportional allocation were achieved at each of the lower administrative levels by sorting the sampling frame within each sampling stratum before sample selection, according to administrative units in different levels, and by using a probability proportional to size selection at the first stage of sampling.

In the first stage, 850 SEAs, including 173 SEAs in urban areas and 677 in rural areas, were selected with probability proportional to the SEA size and with independent selection in each sampling stratum. The SEA size is the number of residential households in the SEA as defined in the 2008 MPHC. A household listing operation was implemented in all the selected SEAs during August-October 2016. The resulting lists of households served as a sampling frame for the selection of households in the second stage. Some of the selected SEAs were large. To minimise the task of household listing, each large SEA (more than 250 households) selected for the 2015-16 MDHS was segmented. One segment was selected for the survey with probability proportional to the segment size. A household listing was conducted only in the selected segment. Thus, in the 2015-16 MDHS, a cluster is either an SEA or a segment of an SEA.

In the second stage of selection, a fixed number of 30 households per urban cluster and 33 per rural cluster were selected with an equal probability systematic selection from the newly created household listing. All women age 15-49 who were either permanent residents of the selected households or visitors who stayed in the household the night before the survey were eligible to be interviewed. In one-third of all sampled households, all men age 15-54, including both usual residents and other persons who stayed in the household the night before the interview, were eligible for individual interview.[...] Height and weight information was also collected from eligible women and from children age 0-59 months."

Source: *National Statistical Office - NSO/Malawi and ICF. 2017. Malawi Demographic and Health Survey 2015-16. Zomba, Malawi: NSO and ICF. Available at: https://dhsprogram.com/pubs/pdf/FR319/FR319.pdf*

**Malawi STEPS 2009**
"This survey was designed to obtain data that would be a representation of the whole Malawi. To achieve this, a multi-stage sampling method was used to select enumeration areas (EAs), households and eligible participants (three stages).

*Stage 1: Selection of enumeration areas (EAs):*
Sampling frame: Administratively, Malawi is divided into twenty-eight districts. In turn, each district is subdivided into smaller administrative units. Each administrative unit is sub-divided into enumeration areas (EAs) by the National Statistical Office (NSO). Enumeration areas are classified as urban or rural. Each EA has a sketch map drawn by NSO. The sketch map shows the EA boundaries, location of buildings, and other landmarks. The list of EAs in Malawi from the latest population and housing census conducted in June 2008 was obtained from NSO. This list was used as a sampling frame for random selection of EAs for the NCD STEPs Survey as described below.

Number of enumeration areas selected: In accordance with WHO STEPS Manual Part 2 Section 2, the recommended number of participants to be selected at each primary sampling unit (in our case in each EA) was 30-50 (WHO 2005). For this survey it was decided that at least 40 participants in each enumeration area were to be recruited. Given that the estimated required sample size was 5,760 (including the 20% non-response rate), the total number of EAs selected was therefore 5,760/40= 144. Thus a total of 144 EAs were randomly selected from the list of all EAs in Malawi.

Sampling method for EA selection: Probability Proportional to Size (PPS) sampling method was used to randomly sample the 144 EAs from the whole country as follows: The EAs in Microsoft Excel® were first sorted in descending order of population (largest to the smallest).

- Then the total population of all EAs in Malawi were calculated.
- A column of cumulative total population of EAs was then created.
- Then the sampling interval was calculated by dividing total population by 144 (total number of EAs to be selected).
- Random number (the seed) was generated by computer in the excel® using the formula RANDBETWEEN (sampling interval).
- The EA whose cumulative total contained the seed was the first to be selected.
- The 2nd, 3rd, ........, 144th EA was selected by systematically adding the random number to the sampling interval. [...]

*Stage 2: Selection of households*
The EA sampling frame obtained from NSO had information on the total number of households in each EA. Forty households were to be selected from each EA (as described above under of number of EAs to be selected). The sampling interval for household selection in each EA was therefore determined by dividing total EA population by 40. Systematic sampling method (every nth household) was then used to randomly select the required 40 households.

*Stage 3: Selection of eligible participants at household level*
Only one eligible participant (an adult aged 25-64 years) in the selected households was enrolled in the survey. In households with more than one eligible participant, Kish Method sampling method was used to randomly select one. By Kish Method, eligible participants in each household were ranked in order of decreasing age, starting with males then females."

Source: *Ministry of Health and WHO: Malawi National STEPS survey for chronic non-communicable diseases and their risk factors. Final report. Available at: https://extranet.who.int/ncdsmicrodata/index.php/catalog/435*

**Mexico GATS 2015**

*"Target population:* The 2015 GATS Mexico was a nationally representative house-hold survey of the population 15 years of age and older. The survey was conducted in both urban and rural areas. Eligible persons are citizens and non-citizens resident in the country and have lived in the country for, at least six months within the year prior to the survey. The definition for the target population was consistent with the 2009 GATS Mexico. Individuals aged 15 years and older were excluded from the survey if they were non-citizen visitors, or citizens who lived inside military bases, hospitals, prisons, nurseries or any other institutionalized residence.

*Sampling criteria:* In urban areas, Area Geoestadística Básica or Basic Geostatisti-cal Area – (AGEB) from the Mexican National Population and Housing Census 2010 (30) lists were used as a primary sampling unit, which provides lists of urban blocks and also area maps. In rural areas, the same census location list was used as part of the clusters housing frame. The population areas excluded from AGEB's were included in the location lists for each municipality, to achieve a complete coverage for all the eligible population and meet the sampling criteria.

*Sampling methodology:* A three-stage stratified cluster probability sampling scheme was developed and adjusted for non-response using non-response rates by gender and residence obtained from 2009 GATS Mexico. The 2015 GATS Mexico sample was an indepen-dent sample of adults aged 15 years and older. The reporting domain was by residence and gender, as presented in table A. [...]

*Stratification:* The sample design considered two levels of stratifications, the first one based on the locality size, and the second one based on the identification of tobacco consumption patterns at a state level, per gender and residence. For the first stratification, in urban areas (same as the 2009 GATS Mexico), the following was used: a) urban Area Geoestadística Básica or Basic Geostatistical Area (AGEBs) in localities with more than 100, 000 inhabitants, b) suburban AGEB's in localities with 2,500 and less than 100,000 inhabitants. In rural areas, localities showed wide variability in sizes. There are localities with hundreds or even thousands of in-habitants; however, there are also localities with less than 10 inhabitants. Stratification, in this case, was by: a) Large localities from 100 to 2,500 inhabitants, and b) small localities with less than 100 inhabitants. Second, stratification was built with the Ward hierarchical clustered method with Euclidean distances between states, us-ing tobacco consumption prevalence by gender and residence from the 2006 Encuesta Nacional de Salud y Nutrición survey (the same survey estimates used for the 2009 GATS Mexico). The stratum according to tobacco consumption prevalence are shown in table C. Stratification considered 20 stratums, 10 for urban areas (2 for residence per 5 for tobacco use prevalence); and 10 for rural areas (also 2 for rural per 5 for tobacco use prevalence. [...]

*Final sample:* The original sample distribution was 9,900 households for urban areas and 7,860 households for rural. A total planned sample of 17,765 households was selected for the survey. Initially, in the urban stratum, 223 PSUs were selected. However, two of the selected 223 PSUs were inaccessible. An additional four PSUs from Mexico City and four from Baja California were selected to bring the total to 229 PSUs with complete infor-mation. In the suburban stratum, 108 PSUs were selected and visited. In the rural stratum, 258 PSUs were selected in large localities with only 254 providing complete information, and 30 PSUs were selected in small localities with only 26 providing complete information."

Source: *Pan American Health Organization, Instituto Nacional de Salud Pública. Global Adult Tobacco Survey. Mexico 2015. Cuernavaca, Mexico: INSP/PAHO, 2017. Available at: https://www.who.int/tobacco/surveillance/survey/gats/mex-report-2015-english.pdf?ua=*

**Mexico Family Life Survey 2009-12**

"The design of the first round, the baseline survey (MxFLS-1), was undertaken by the National Institute of Statistics and Geography (INEGI, per its name in Spanish). The baseline sample is probabilistic, stratified, multi-staged, and independent at every phase of the study. The population is comprised by Mexican households in 2002. Primary sampling units were selected under criterions of national, urban-rural and regional representation on pre-established demographic and economic variables. Regional definitions are in accordance with the National Development Plan 2000-2006.

Currently, the MxFLS contains information for a 10-year period, collected in three rounds: 2002 (MxFLS-1), 2005-2006 (MxFLS-2) and 2009-2012 (MxFLS-3). Future rounds have been programmed in order to have a database that allows studying efficiently the well-being of the Mexican population at different moments in time. The first round or baseline survey (MXFLS-1), implemented in 2002, collected information on a sample of 35,000 individuals from 8,400 households in 150 communities throughout the country. The second (MxFLS-2) and third round (MxFLS-3) were conducted during 2005-2006 and 2009-2012, respectively. Given the longitudinal design of the survey, the MxFLS-2 and MxFLS-3 aimed to relocate and re-interview the sample of the MxFLS-1—including those individuals who migrated within Mexico or emigrated to the United States of America—and to interview the individuals or households that grew out from previous samples. The MxFLS-2 and MxFLS-3 relocated and re-interviewed almost 90 percent of the original sampled households. A primary goal of the Mexican Family Life Survey (MxFLS) is to create a longitudinal and multi-thematic database. On the one hand, the longitudinal design allows a long term tracking of individuals regardless of changes in residence and new household formations (split-offs). On the other hand, the multi-thematic design allows collecting— with a single tool—a wide range of socioeconomic and demographic indicators of the Mexican population. The first round of the survey (MxFLS-1) took place during 2002 reaching a sample of 8,400 households (35,000 individuals) in 150 urban and rural communities throughout the country. The second (MxFLS-2) and third round (MxFLS-3) were conducted during 2005-2006 and 2009- 2012, respectively. Given the longitudinal design of the survey, the MxFLS-2 and MxFLS-3 aimed to relocate and re-interview the sample of the MxFLS- 1—including those individuals who migrated within Mexico or emigrated to the Unites States—and to interview the individuals or households that grew out from previous samples. The MxFLS-2 and MxFLS-3 relocated and re- interviewed almost 90 percent of the original sampled households."

Source: *The Mexican Family Life Survey website. http://www.ennvih-mxfls.org/english/introduccion.html*

**Mongolia STEPS 2019**
"A multistage stratified sampling design was used to produce representative data for that age range in Mongolia."

Source: *National Center for Disease Control and Public Health. Mongolia STEPS Survey 2019 Fact Sheet. Available at: https://extranet.who.int/ncdsmicrodata/index.php/catalog/836/related-materials*

**Mozambique DHS 2011**
The original report is in Portugese. We translated the sampling strategies using Google translate.

"The IDS-2011 comprises a probabilistic, stratified and multi-stage sample, selected from the Data and Cartography of the III General Census of Population and Housing, carried out by INE in 2007. The sample allows to obtain accurate estimates at national, urban and rural levels, regional and provincial. The sample covers only the population residing in households. Households and respective members residing in collective residences, such as hotels, hospitals, military barracks, student homes, etc., and homeless people, which together make up 3.3% of the country's total population, were excluded from the sample.

In the first stage, 611 UPA (Primary Sampling Units) were selected with probability proportional to the size, with the number of households in each stratum within each province being measured in size. In the second sample stage, 20 households in the urban UPA and 25 households in the rural UPA were selected with equal probability. This selection was made after a previous listing of households. Finally, in the third stage, an exhaustive sampling was performed, that is, demographic and health data were collected for all women aged 15-49 and children under 5 found in the selected households.

In each selected enumeration area, a list of structures was made and their classification into residential and non-residential. Only residential structures were used for sample selection in each enumeration area. In each province 51-55 UPA were selected, with the exception of Nampula (60 UPA), Zambézia and Sofala (58 UPA each), Maputo Province (61 UPA) and Maputo City (65 UPA), due to the greater variability in social characteristics -demographic and low coverage indicators for women 15-49 years old and children under 5 years old for these provinces. Appendix A is dedicated to the detailed description of the sample design methodology, including its breakdown by domain of analysis and procedures for selection at each sampling step.

For the identification of the limits of the enumeration areas (UPA) in the field, the Global Differential Positioning System (DGPS) was used, where control points were introduced and united to show the AE limit in the DGPS. After identifying the AE boundaries on the ground and next to the Guide, the inquirer proceeded with a listing based on the DGPS, where each listed household was represented by a pair of coordinates and an identification number on the DGPS."

Source: *Ministerio da Saude - MISAU/Moçambique, Instituto Nacional de Estatística - INE/Moçambique and ICF International. Moçambique Inquérito Demográfico e de Saúde 2011. Calverton, Maryland, USA: MISA/Moçambique, INE/Moçambique and ICF International. Available at: https://dhsprogram.com/pubs/pdf/FR266/FR266.pdf*

**Mozambique STEPS 2005**

"For the present community-based cross-sectional study, a sample of adults aged 25 to 64 years was assembled using the sampling frame of the 1997 census, which was designed to be representative at a national level and by place of residence (urban or rural). Ninety-five geographical clusters were selected, among which all of the households were listed and 25 randomly selected and visited. All of the eligible subjects in the same household were invited for the study. Fifty-five subjects refused to participate, and 3323 were evaluated between September and November 2005.

Subjects were evaluated following the World Health Organization Stepwise Approach to Chronic Disease Risk Factor Surveillance (STEPS), which included a questionnaire on sociodemographic and behavioral factors (including smoking and drinking habits), medical and health history, and physical measurements (including blood pressure, weight, height, and waist circumference), using standardized methods. The World Health Organization STEPS instrument for non-communicable disease risk factors (core and expanded version 2.18) was used for data collection, after translation to Portuguese."

*Source: Albertino Damasceno, Ana Azevedo, Carla Silva-Matos, António Prista, Domingos Diogo, Nuno Lunet (2009). Hypertension Prevalence, Awareness, Treatment, and Control in Mozambique Urban/Rural Gap During Epidemiological Transition. Hypertension. 54: 77-83.*

**Myanmar DHS 2015-2016**

"The sampling frame consisted of 76,990 primary sampling units (PSUs) across the country. A PSU is either a census enumeration area (EA) or a ward or village tract in a sensitive area not enumerated during the census. Each PSU had cartographic materials that delineated its geographic location, boundaries, main access points, and landmarks. The sampling frame contained information about each PSU's administrative subordinations (state or region and district), the type of residence (urban or rural), and the estimated number of residential households. The sampling frame excluded institutional populations, such as persons in hotels, barracks, and prisons, but included those from internally-displaced population camps. A master sample was created under the aegis of the Department of Population within the Ministry of Labor, Immigration and Population. The sample was based on the 2014 census frame, which is used to coordinate household-based surveys conducted in Myanmar, including the current 2015-16 MDHS. The master sample is a large, nationally representative sample consisting of 4,000 PSUs drawn from the entire census frame; these can be used for sub-selecting multi-stage household-based survey samples. The master sample is large enough to provide design flexibility for the various household-based surveys in Myanmar. The master sample is a stratified sample, selected with probability proportional to size (PPS).

Stratification is achieved by separating each state or region into urban and rural areas, each of which formed a separate sampling stratum. In total, 30 sampling strata were created. Samples were selected independently in each sampling stratum. Implicit stratification and proportional allocation were achieved at each of the lower administrative unit levels. This was done before sample selection by sorting the sampling frame within the explicit stratum, according to administrative unit, and by using a PPS selection procedure. The 2015-16 MDHS followed a stratified two-stage sample design and was intended to allow estimates of key indicators at the national level, in urban and rural areas, and for each of the seven States and eight Regions of Myanmar. The first stage involved selecting sample points (clusters) consisting of EAs or ward/village tracts. A total of 442 clusters (123 urban and 319 rural) were selected from the master sample.

At the second stage, a fixed number of 30 households was selected from each of the selected clusters (a total of 13,260 households), using equal probability systematic sampling. For the clusters, which were completely enumerated during the population census, the census household listings were taken as the base and updated in the field by the household listing teams. These updated lists were used for selecting the sample households. For the clusters that were not enumerated or partially enumerated during the census, an independent household listing operation was carried out. Because of the non-proportional sample allocation, the sample was not a self-weighting sample. Weighting factors had to be calculated, added to the data file, and applied so that results are representative at the national as well as regional level.

All women age 15-49 who were either permanent residents of the selected households or visitors who stayed in the households the night before the survey were eligible to be interviewed. In half of the selected households (every second household), all men age 15-49 who were either residents or visitors who stayed in the household the night before the survey were eligible to be interviewed. During the course of the fieldwork, 4 clusters were identified as insecure and were replaced with other clusters in the vicinity. In addition, 1 urban cluster had to be dropped due to worsening security. Overall, the survey was successfully carried out in 441 clusters."

Source: *Ministry of Health and Sports (MoHS) and ICF. 2017. Myanmar Demographic and Health Survey 2015-16. Nay Pyi Taw, Myanmar, and Rockville, Maryland USA: Ministry of Health and Sports and ICF. Available at: https://dhsprogram.com/pubs/pdf/FR324/FR324.pdf*

**Myanmar STEPS 2014**
"To achieve a nationally representative sample, a multi-stage sampling method was used to select townships, wards and villages, households and eligible participants at each of the selected households.

*Stage 1: Selection of primary sampling units (PSUs)*
Administratively, Myanmar is divided into 330 townships. A township is subdivided into wards for urban settings and village tracts and then villages for rural settings. The list of townships has been used as the sampling frame at the first stage of sampling. Townships form the Primary Sampling Units (PSUs). Out of the total 330 PSUs, 52 PSUs were selected using Probability Proportionate to Size of population in each PSU (PPS).

*Stage 2: Selection of Secondary Sampling Units (SSUs)*
From each selected PSU (township), 6 SSUs (wards and villages) were chosen using probability proportionate to population size, totaling 312 SSUs for the whole country.

*Stage 3: Selection of eligible participants at household level*
From each selected SSU (ward/village), 30 households were selected using systematic random sampling. The sampling frame for this sampling is the list of households with unique identification number (ID) developed from a recent listing of households available from the Basic Health Staff.

*Stage 4: Selection of eligible participants at household level*
One eligible participant (aged between 25 and 64 years) in the selected households was recruited for the survey. The Kish sampling method was used to randomly select one eligible member of the household. Using the Kish Method, eligible participants (adults aged 25 to 64 years) in each household were ranked in order of 8 decreasing age, starting with males then females, then randomly selected using the automated program for Kish selection in the handheld PDA. Each PSU (township) was estimated to contribute 180 participants, totaling 9,360 participants for 52 selected townships for the whole country. In actual study, the total sample size was 8757 participants."

Source: *Myanmar Diabetes Association, Department of Public Health and Department of Medical Research: Report on National Survey of diabetes mellitus and risk factors for non-communicable diseases in Myanmar (2014). Available at: https://www.who.int/ncds/surveillance/steps/myanmar/en/*

**Namibia DHS 2013**
"The sample for the 2013 NDHS was a stratified sample selected in two stages. In the first stage, 554 EAs were selected with a stratified probability proportional to size within the sampling frame. The EA size is the number of households residing in the EA and recorded in the 2011 NPHC. Stratification was achieved by separating each region into urban and rural areas. Therefore, the 13 regions were stratified into 26 sampling strata: 13 rural strata, and 13 urban strata. Samples were selected independently in each stratum, with a predetermined number of EAs selected as shown in Table A.3. Implicit stratification with proportional allocation was achieved at each of the lower administrative unit levels by sorting the sampling frame before the sample selection. Sorting was done according to the constituency and the EA code within a sampling stratum, and by using a probability proportional-to-size selection procedure.

After the selection of EAs and before the main survey, a household listing operation was carried out in all selected EAs, and the resulting lists of households served as a sampling frame for the selection of households in the second stage. Some of the selected EAs may large. To limit the amount of work done to list each household, selected EAs with more than 200 households were segmented by the listing team in the field before the household listing. Only one segment was selected for the survey, with probability proportional to the segment size. Household listing was conducted only in the selected segment (see detailed instructions for segmentation in the DHS Manual for Household Listing). So a 2013 NDHS cluster is either an EA or a segment of an EA. In the second-stage selection, a fixed number of 20 households was selected in every urban cluster and rural cluster, by equal probability systematic sampling. A spreadsheet indicating the selected household numbers for each cluster was prepared. The survey interviewers interviewed only the pre-selected households. To prevent bias, no replacements and no changes of the pre-selected households were allowed in the implementing stages. In half of the selected households where there was no male survey, all women age 15-49 were interviewed; in the other half of the selected households where there was a male survey, all males and females age 15-64 were interviewed."

Source: *The Namibian Ministry of Health and Social Services (MoHSS) & ICF International. 2014. The Namibia Demographic and Health Survey 2013. Available at: https://dhsprogram.com/pubs/pdf/fr298/fr298.pdf.*

**Niger DHS 2012**
The original report is in French. We translated the sampling strategies using Google translate.

"The EDSN-MICS IV 2012 sample is a nationally representative sample of households, based on a stratified two-stage cluster survey. As with all past DHSs, the EDSN-MICS IV comprises 10 fields of study made up of each of the eight regions of the country (Agadez, Diffa, Dosso, Maradi, Tahoua, Tillabéri, Zinder and Niamey), the urban environment and the rural environment.

In the first stage, 480 clusters were selected, and drawn in proportion to their size, from the list of Enumeration Areas (EA) established during the General Population and Housing Census (RGPH) of 2001. All the clusters Selected clusters were the subject of an exhaustive enumeration of households. In the second stage, households were drawn, with equal probability, from the list of households established during the enumeration operation. The number of households drawn is 25 in each urban or rural cluster. Due to insecurity in the Agadez region, three and a half clusters were abandoned, but this did not affect the region's performance. All selected household members were identified using the household questionnaire.

All women aged 15 to 49 (usual residents or visitors) who were in these households were interviewed individually. These women, along with all children under the age of five, were measured and weighed in one in two households."

Source: *Institut National de la Statistique - INS/Niger and ICF International. 2013. Enquête Démographique et de Santé et à Indicateurs Multiples du Niger 2012. Calverton, Maryland, USA: INS/Niger and ICF International. Available at: https://dhsprogram.com/pubs/pdf/FR277/FR277.pdf*

**Niger STEPS 2007**
The original report is in French. We translated the sampling strategies using Google translate.

"In the first stage, the EAs were taken from those delimited in the Third General Census of Population and Housing. The EAs were drawn up systematically and independently in each stratum. For each sample, the cumulative size of the population of each stratum was first calculated in the sampling frame. The Polling interval was then calculated as follows: I =M/a, rounded to the nearest whole, where M is the population of the stratum according to the sampling frame and the number of EAs to be drawn in the stratum. The series of survey numbers R, R + I, R + 2I, R + 3I, etc. was calculated, where R is the random number between 1 and I. Each survey number was then compared to the cumulative headcount column. The first EA that was drawn is the first EA on the list whose cumulative workforce is equal to or greater than the first survey number. The second EA that was drawn is the one that follows (after the first EA that has been drawn) whose cumulative number is equal to or greater than the second survey number, and so on. .[...] 30 households were systematically drawn by EA. The step is therefore equal to the number of households in the EA divided by 304. In the household, a person meeting the age criterion was interviewed, that is to say between the ages of 15 and 64. In the event that the quota for the 55-64 age group is difficult to reach, two participants have been selected in households where there is an adult aged 55 to 65, this in accordance with the Kish method."

Source: *République du Niger - Ministere de la santé publique. Direction générale de la santé publique et direction de la lutte contre la maladie et les endémies: Mesure des facteurs de risque des maladies non transmissibles Au Niger (Approche Step"wise" de l'OMS). Available at: https://www.who.int/ncds/surveillance/steps/niger/en/*

**Nigeria DHS 2018**

"The sampling frame used for the 2018 NDHS is the Population and Housing Census of the Federal Republic of Nigeria (NPHC), which was conducted in 2006 by the National Population Commission. Administratively, Nigeria is divided into states. Each state is subdivided into local government areas (LGAs), and each LGA is divided into wards. In addition to these administrative units, during the 2006 NPHC each locality was subdivided into convenient areas called census enumeration areas (EAs). The primary sampling unit (PSU), referred to as a cluster for the 2018 NDHS, is defined on the basis of EAs from the 2006 EA census frame. Although the 2006 NPHC did not provide the number of households and population for each EA, population estimates were published for 774 LGAs. A combination of information from cartographic material demarcating each EA and the LGA population estimates from the census was used to identify the list of EAs, estimate the number of households, and distinguish EAs as urban or rural for the survey sample frame. Before sample selection, all localities were classified separately into urban and rural areas based on predetermined minimum sizes of urban areas (cut-off points); consistent with the official definition in 2017, any locality with more than a minimum population size of 20,000 was classified as urban.

The sample for the 2018 NDHS was a stratified sample selected in two stages. Stratification was achieved by separating each of the 36 states and the Federal Capital Territory into urban and rural areas. In total, 74 sampling strata were identified. Samples were selected independently in every stratum via a two-stage selection. Implicit stratification were achieved at each of the lower administrative levels by sorting the sampling frame before sample selection according to administrative order and by using a probability proportional to size selection during the first sampling stage.

In the first stage, 1,400 EAs were selected with probability proportional to EA size. EA size was the number of households in the EA. A household listing operation was carried out in all selected EAs, and the resulting lists of households served as a sampling frame for the selection of households in the second stage. In the second stage's selection, a fixed number of 30 households was selected in every cluster through equal probability systematic sampling, resulting in a total sample size of approximately 42,000 households. The household listing was carried out using tablets, and random selection of households was carried out through computer programming. The interviewers conducted interviews only in the pre-selected households. To prevent bias, no replacements and no changes of the pre-selected households were allowed in the implementing stages.

Due to the non-proportional allocation of the sample to the different states and the possible differences in response rates, sampling weights were calculated, added to the data file, and applied so that the results would be representative at the national level as well as the domain level. Because the 2018 NDHS sample was a two-stage stratified cluster sample selected from the sampling frame, sampling weights were calculated based on sampling probabilities separately for each sampling stage and for each cluster.

The 2018 NDHS included all women age 15-49 in the sample households. Those who were either permanent residents of the selected households or visitors who stayed in the households the night before the survey were eligible to be interviewed. The men's survey was conducted in one-third of the sample households, and all men age 15-59 in these households were included (Figure 1.1). [...] Similarly, biomarker information was collected only in those households selected for the men's survey. The biomarkers included in this survey were height and weight for women age 15-49 [...].

The survey was successfully carried out in 1,389 clusters after 11 clusters with deteriorating law-and-order situations during fieldwork were dropped. These areas were in Zamfara (4 clusters), Lagos (1 cluster), Katsina (2 clusters), Sokoto (3 clusters), and Borno (1 cluster).In the case of Borno, 11 of the 27 LGAs were dropped due to high insecurity, and therefore the results might not represent the entire state."

Source: *National Population Commission - NPC/Nigeria and ICF. 2019. Nigeria Demographic and Health Survey 2018. Abuja, Nigeria, and Rockville, Maryland, USA: NPC and ICF. Available at: https://dhsprogram.com/pubs/pdf/FR359/FR359.pdf*

**Pakistan DHS 2017-2018**

"The sampling frame used for the 2017-18 PDHS is a complete list of enumeration blocks (EBs) created for the Pakistan Population and Housing Census 2017, which was conducted from March To May 2017. The Pakistan Bureau of Statistics (PBS) supported the sample design of the survey and worked in close coordination with NIPS. The 2017-18 PDHS represents the population of Pakistan including Azad Jammu and Kashmir (AJK) and the former Federally Administrated Tribal Areas (FATA), which were not included in the 2012-13 PDHS1. The results of the 2017-18 PDHS are representative at the national level and for the urban and rural areas separately. The survey estimates are also representative for the four provinces of Punjab, Sindh, Khyber Pakhtunkhwa, and Balochistan; for two regions including AJK and Gilgit Baltistan (GB); for Islamabad Capital Territory (ICT);and for FATA. In total, there are 13 second-level survey domains.

The 2017-18 PDHS followed a stratified two-stage sample design. The stratification was achieved by separating each of the eight regions into urban and rural areas. In total, 16 sampling strata were created. Samples were selected independently in every stratum through a two-stage selection process. Implicit Stratification and proportional allocation were achieved at each of the lower administrative levels by sorting the sampling frame within each sampling stratum before sample selection, according to administrative units at different levels, and by using a probability-proportional-to-size selection at the first stage of sampling.

The first stage involved selecting sample points (clusters) consisting of EBs. EBs were drawn with a probability proportional to their size, which is the number of households residing in the EB at the time of the census. A total of 580 clusters were selected.

The second stage involved systematic sampling of households. A household listing operation was undertaken in all of the selected clusters,and a fixed number of 28 households per cluster was selected with an equal probability systematic selection process, for a total sample size of approximately 16,240households. The household selection was carried out centrally at the NIPS data processing office. The Survey teams only interviewed the pre-selected households. To prevent bias, no replacements and no changes to the pre-selected households were allowed at the implementing stages.

Due to non-proportional sample allocation, the sample was not self-weighting. Weighting Factors have been calculated, added to the data file, and applied so that results are representative at the national level for Pakistan (including FATA and ICT Islamabad) and separately for Azad Jammu and Kashmir and Gilgit Baltistan. The 2017-18 PDHS included all ever-married women age 15-49. Those who were either permanent residents of the selected households or visitors who stayed in the households the night before the survey were eligible to be interviewed. The Survey of men was conducted in one-third of the sample households,and all ever-married men age 15-49 in these households were included. [...] Similarly, height and weight information was collected from eligible women age 15-49 and children age 0-59 months only in those households selected for the man's survey.

The survey was successfully carried out in 561 clusters, after dropping 19 clusters due to security concerns during the fieldwork. These clusters were in Balochistan (1), FATA (2), GilgitBaltistan (6), Khyber Pakhtunkhwa (4), Azad Jammu and Kashmir (1), Punjab (2), Sindh (1) and ICT Islamabad (2 restricted areas)."

Source: *National Institute of Population Studies - NIPS/Pakistan and ICF. 2019. Pakistan Demographic and Health Survey 2017-18. Islamabad, Pakistan, and Rockville, Maryland, USA: NIPS and ICF. Available at: https://dhsprogram.com/pubs/pdf/FR354/FR354.pdf*

**Panama GATS 2013**
"The GATS sampling frame included information obtained from the 2010 National Census of Population and Housing as a reference, with a total of 21,222 primary sampling units (PSUs), each containing 38 and more occupied households, distributed as follows by number of PSUs: urban, 13,926; rural, 6,354; and indigenous, 1,053, representing 2,691,551 people aged 15 years or over. The survey relied on a representative sample, selected randomly, which considered 959 PSUs that were distributed in 14 health regions of the country, and a sampling size of 19,603 households, which was split almost evenly by gender. GATS Panama 2013 used a multistage probability sample of adults aged ≥15 years as the sampling frame. A stratified, cluster sample design was employed to select the households to be included in the survey.

Sampling stages:

*First Stage:* This involved the selection of primary sampling units (PSUs) having 38 or more households or more using three strata: urban, rural, and indigenous. In this stage, the PSUs of the indigenous strata were classified in two categories as (a) self-represented (with a probability equal to 1 of being included in the sample) or (b) not self-represented. All the PSUs with a population of fewer than 60 inhabitants were included in the indigenous sample of the GATS.

*Second Stage:* This involved random selection of households in selected PSUs.

*Third Stage:* In this stage an individual was randomly selected from the selected households. As established in the study methodology, selection of these single sampling units involved consideration of gender parity.

The sample, which included 16,962 adults, was treated in such a way as to be applicable to the 2,691,551 Panamanians aged 15 years or over."

Source: *Ministerio de Salud. Instituto Conmemorativo Gorgas de Estudios de Salud: Global Adult Tobacco Survey: Country report 2013. Available at: https://www.who.int/tobacco/surveillance/survey/gats/pan_report2013.pdf?ua=1*

**Peru DHS 2012**

"The Peruvian Encuesta Demografica y de Salud Familiar was a cross-sectional, multistage survey based on a probabilistic cluster sampling design. The survey took place between March and December of 2012 and was performed by the Instituto Nacional de Estadistica e Informatica (INEI). The sample population were females of childbearing age (15 and 49 years). The sample was selected in two stages. The first stage utilized information from the Population and Household Census 2007 and entailed the random selection of clusters as the primary sampling unit (UPM) with probability proportional to their population size. Each UPM had an average size of 120 households. In the second stage, individual households were selected as the secondary sampling unit (USM). Prior to the second stage, a cartographic update and registry of the households within each cluster was performed. Out of the 28,376 households selected, the interview was performed in 27,488 households, yielding a response rate of 99 percent. Out of 24,552 eligible women, 23,888 were interviewed, yielding a response rate of 97.3 percent."

Source: *Instituto Nacional de Estadística e Informática. 2013. Encuesta Demográfica y de Salud Familiar 2012. Available at: https://dhsprogram.com/pubs/pdf/FR284/FR284.pdf. Blood pressure measurement details were obtained directly from the Oficina Tecnica de Difusion at the Instituto Nacional de Estadistica e Informatica on April 11th, 2019.*

**Philippines GATS 2015**
"The GATS Philippines 2015 used the Philippines 2003 Master Sample (MS) created for the Philippines Statistics Authority (PSA)'s household-based surveys with some modifications to conform to GATS protocol on sampling design. One of the four replicates of the MS was used for GATS.

*Stratification:* Within each region, the first level of stratification was by province, highly urbanized cities (HUCs) and independent component cities (ICCs). This stratification is helpful for making separate design-based estimates for provinces, HUCs and ICCs. However, the estimates at this level generally will have low reliability. Implicit stratification was also applied within the final explicit strata using the following variables: proportion of strong horses; agricultural indicators; and per capita income (i.e.total income of the municipality divided by its total population).

*Stages of Selection:* The 2003 MS was based on a three-staged sample design with barangays or combination of small contiguous barangays within the same municipality as primary sampling units (PSU). The average PSU size or the average number of households in a PSU is 923. For the GATS Philippines 2015, an additional stage of selection was added to select one eligible respondent per household.

At the first stage of selection, where there were 16,579 primary sampling units (PSUs) across the country, a sample of 2,826 PSUs was drawn using probability proportional to the estimated size (PPES), the measure of size being the number of households from 2000 Census of Population and Housing. From the full set of 2,826 sampled PSUs, four replicates are formed by randomly assigning any number from one to four to each PSU. PSUs with the same number were grouped together to form one replicate. For the 2015 Philippine GATS, replicate four is selected.

In the second stage selection, within each selected PSU, enumeration areas (EAs) were selected with probability proportional to the number of households in the EA. An EA was defined as an area with discernible boundaries consisting of approximately 350 contiguous households. For the GATS Philippines 2015, there were 794 sample EAs in replicate four.

In the third stage, within each sampled EA,16 to 18 housing units (HUs) on the average were selected using systematic sampling. For operational considerations, at most 30 HUs were selected per sample EA. All households in the sample HUs were interviewed except for HUs with more than three households. In those cases, only three households were selected. For the GATS Philippines 2015, all households listed in the sample HU were interviewed. However, if additional households were found during the actual visit, random selection process was applied to reselect a sample household within the sample housing unit. Finally, from each sampled household, one eligible respondent from the household members 15 years old and over, regardless of sex, was randomly selected for interview using the GATS Individual Questionnaire."

Source: *Department of Health and Philippine Statistics Authority: Global Adult Tobacco Survey: Philippines Country Report 2015. Available at: https://www.who.int/tobacco/surveillance/survey/gats/phl_country_report.pdf?ua=1*

**Romania Study for the Evaluation of Prevalence of Hypertension and Cardiovascular Risk (SEPHAR II)**

"Sampling was performed by a multi-stratified procedure, leading to the selection of a representative sample of 1942 adults. Subject selection followed the principle of equality of chances of being enrolled in the study, regardless of the size of the place of residency.

Stratification criteria for sample selection were:

- territorial regions (Romania's territory was divided into 7 regions plus the capital city Bucharest, based on the National Statistics Institute recommendations: the North-East region, the South-East region, the South region, the South-West region, the West region, the North-West region, the Central region and the Bucharest region);
- locality type (cities with over 200 000 inhabitants, cities with 50 000–200 000 inhabitants, cities with less than 50 000 inhabitants, Commune);
- gender (male and female);
- age groups (18–24 years, 25–34 years, 35–44 years, 45–54 years, 55–64 years, 65–80 years).

In the first stage of selection, the adult population weighted average was calculated for each region and each district, and, based on this, the number of adult persons from each region/district was calculated from the working sample of 1942 subjects.

In the second stage of selection, the number of localities of a certain size from which the subjects were later selected was established for each district. This number was directly proportional to the population in the respective district. A random selection of a certain locality in a certain category was done using a computer software (generation of random numbers). The selected localities represent the interview centers where the study was to take place. The weighted average of the specific locality population in the district was calculated, and, based on this, the number of people selected to participate in the study.

The third stage of selection consisted of distribution by gender of adult people selected from each locality, using Romania's population gender distribution according to the 2002 census (F : M = 51.25% vs. 48.75%) and the fourth stage of selection consisted of distribution by age of male and female adult people selected from each locality, using Romania's population age distribution according to the 2002 census."

Source: *Dorobantu M, Tautu OF, Darabont R, Ghiorghe S, Badila E, Dana M, Dobreanu M, Baila I, Rutkowski M, Zdrojewski T. Objectives and methodology of Romanian SEPHAR II Survey. Project for comparing the prevalence and control of cardiovascular risk factors in two East-European countries: Romania and Poland. Arch Med Sci. 2015 Aug 12;11(4):715-23.*

**Rwanda DHS 2014-2015**
"The sampling frame used for the 2014-15 RDHS was the 2012 Rwanda Population and Housing Census (RPHC). The sampling frame consisted of a list of enumeration areas (EAs) covering the entire country, provided by the National Institute of Statistics of Rwanda, the implementing agency for the RDHS. An EA is a natural village or part of a village created for the 2012 RPHC; these areas served as counting units for the census.

The 2014-15 RDHS followed a two-stage sample design and was intended to allow estimates of key indicators at the national level as well as for urban and rural areas, five provinces, and each of Rwanda's 30 districts (for some limited indicators). The first stage involved selecting sample points (clusters) consisting of EAs delineated for the 2012 RPHC. A total of 492 clusters were selected, 113 in urban areas and 379 in rural areas.

The second stage involved systematic sampling of households. A household listing operation was undertaken in all of the selected EAs from July 7 to September 6, 2014, and households to be included in the survey were randomly selected from these lists. Twenty-six households were selected from each sample point, for a total sample size of 12,792 households. However, during data collection, one of the households was found to actually be two households, which increased the total sample to 12,793. Because of the approximately equal sample sizes in each district, the sample is not self-weighting at the national level, and weighting factors have been added to the data file so that the results will be proportional at the national level.

All women age 15-49 who were either permanent residents of the household or visitors who stayed in the household the night before the survey were eligible to be interviewed. In half of the households, all men age 15-59 who either were permanent household residents or were visiting the night before the survey were eligible to be interviewed. [...] Height and weight information was collected from eligible women, and children (age 0-5) in the same subsample. [...] Height and weight information was collected from eligible men."

Source: *National Institute of Statistics of Rwanda, Ministry of Finance and Economic Planning/Rwanda, Ministry of Health/Rwanda, and ICF International. 2015. Rwanda Demographic and Health Survey 2014-15. Kigali, Rwanda: National Institute of Statistics of Rwanda, Ministry of Finance and Economic Planning/Rwanda, Ministry of Health/Rwanda, and ICF International. Available at: https://dhsprogram.com/pubs/pdf/FR316/FR316.pdf*

**Rwanda STEPS 2012**

"Multistage cluster sampling was used to select these participants from the population based on information from the last census. The three levels of clustering were: 1. Random selection of a statistical enumeration area (as defined by NISR) 2. Random selection of a household within the enumeration area 3. Random selection of an individual within the household.

*Selection of Enumeration Areas:* Administratively, Rwanda is divided into thirty districts. In turn, each district is subdivided into sectors. Each sector is sub-divided into cells and then into villages. Villages are synonymous with enumeration area's (EAs) in Rwanda and there are a total of 14,953 EAs in Rwanda. A total of 180 EA's (or 1.2%) were randomly selected from this total using a probability proportional to size method that gives those EA's with more people living in them a higher chance of being selected. In this way, the representativeness of the selected EAs is maximized.

*Selection of households*: Forty households were randomly selected from within each of the selected EAs from a list of households supplied by NISR based on the most recent census. Selection of eligible participants: One eligible participant (an adult aged 15-64 years) was randomly selected from within each household using the Kish sampling method which is built into the PDAs used in the survey."

Source: *Republic of Rwanda. Ministry of Health: Rwanda Non-communicable Diseases Risk Factors Report November 2015. Available at: https://www.who.int/ncds/surveillance/steps/rwanda/en/.*

**Samoa STEPS 2013**
"A multi-stage cluster sample design was used. A total of 1766 adults participated in the survey. Analysis weights were calculated by taking the inverse of the probability of selection of each participant. These weights were adjusted for differences in the age-sex composition of the sample population as compared to the target population. Different weight variables are available per Step:

wStep1 - for interview data
wStep2 - for physical measures
wStep3 - for biochemical measures

This allows for differences in the weight calculation for each Step of the survey as the age-sex composition of the respondents to each Step can differ slightly due to refusal or drop out. Additionally, some countries perform subsampling for Step 2 and/or Step 3. When no subsampling is done and response rates do not differ across Steps of the survey, the 3 weight variables will be the same."

Source: *no report available. Sampling information obtained from the fact sheet. Available at: https://extranet.who.int/ncdsmicrodata/index.php/catalog/707/related_materials and WHO NCD Microdata Repository. Available at: https://extranet.who.int/ncdsmicrodata/index.php/catalog/707/study-description#page=sampling&tab=study-desc*

**São Tomé and Principe STEPS 2008**

"The São Tomé and Principe STEPS survey is a general population survey targeting adults aged 25 to 64. A cluster selection was used to produce representative data for this age group in São Tomé and Príncipe. A total of 2,457 adults participated in the São Tomé and Principe STEPS survey.

A multi-stage cluster sampling design was used to produce representative data.

Analysis weights reflect adjustments for differences in the age-sex composition of the sample population as compared to the target population. Different weight variables are available per Step:

wStep1 - for interview data

wStep2 - for physical measures

wStep3 - for biochemical measures

This allows for differences in the weight calculation for each Step of the survey as the age-sex composition of the respondents to each Step can differ slightly due to refusal or drop out."

Source: *no report available. Sampling information obtained from the fact sheet. Available at: https://extranet.who.int/ncdsmicrodata/index.php/catalog/735/related_materials and WHO NCD Microdata Repository. Available at: https://extranet.who.int/ncdsmicrodata/index.php/catalog/735/study-description#page=sampling&tab=study-desc*

**Senegal GATS 2015**
"A multi-stage, geographically clustered sample design was used to produce nationally representative data. A total of 4,514 households were sampled and one individual was randomly selected from each participating household to complete the survey."

Source: *no report available. Sampling information obtained from the fact sheet. Available at: https://www.who.int/tobacco/surveillance/survey/gats/sen-factsheet-2015.pdf?ua=1*

**Senegal DHS 2010-2011**

“To achieve the objectives of this survey, a stratified national sample of 8,232 households was drawn, with the expectation that approximately 15,044 women age 15-49 and 4,429 men age 15-59 in these households would be interviewed. Blood samples were drawn from one-third of the total sample, 4,104 menand 5,326 women. The sampling procedure used for the EDS-MICS 2010-11 is a two-stage stratified, randomly drawn sample. The primary sampling unit, also called a cluster, is the census district (CD) or a section of a CD when the district is very large. The urban and rural sections of each region each correspond to a sampling stratum. A total of 28 strata were created.

The first-stage sample was drawn independently in each stratum while the second-stage sample was independently derived within each primary unit in the first stage. [...] In the first sampling stage, 391 clusters (147 clusters in urban areas and 244 rural clusters) were selected by conducting a systematic sampling with probability proportional to size, with the size of the CD being the number of households. An enumeration of households in each of these clusters yielded the list of households from which a sample of 21 households was drawn in the second stage through a systematic drawing with equal probability.

All women age 15-49 ordinarily residing in or visiting the household the night before the survey were identified in these households, and were interviewed individually. Within each cluster, among the 21 households selected for the women’s survey, 8 households were selected for the men’s survey. All men age 15-59 in these selected households were eligible for the men’s survey. [...] Moreover, in this same sub-sample of households all men and the eligible women, as well as all children under age 5, were selected to be measured and weighed in order to determine their nutritional status.“

Source: *Agence Nationale de la Statistique et de la Démographie - ANSD/Sénégal and ICF International. 2012. Enquête Démographique et de Santé à Indicateurs Multiples Sénégal (EDS-MICS) 2010-2011. Calverton, Maryland, USA: ANSD and ICF International. https://dhsprogram.com/pubs/pdf/FR258/FR258_English.pdf*

**Solomon Islands STEPS 2015**

"A multi-stage cluster sample design was used to produce representative data. Analysis weights were calculated by taking the inverse of the probability of selection of each participant. These weights were adjusted for differences in the age-sex composition of the sample population as compared to the target population. Different weight variables are available per Step:

wStep1 - for interview data
wStep2 - for physical measures
wStep3 - for biochemical measures

This allows for differences in the weight calculation for each Step of the survey as the age-sex composition of the respondents to each Step can differ slightly due to refusal or drop out."

Source: *no report or fact sheet available. Sampling information obtained from WHO NCD Microdata Repository. Available at: : https://extranet.who.int/ncdsmicrodata/index.php/catalog/710/study-description#page=overview&tab=study-desc*

**South Africa DHS 2016**

"[A] nationally representative sample of 15,000 dwelling units (DUs) was selected; all residential households in the selected DUs were eligible to be included in the survey. In all sampled households, all women age 15-49 who are usual members of the households or who stayed in the households on the night before the interview were eligible for interviews. In addition, in a subsample of the DUs (every second DU), all men age 15-59 who are usual members of the households or who stayed in the households on the night before the interview were eligible for interviews. In this same subsample, all women and men age 15 and older who are usual members of the selected households and those who spent the night before the survey in the selected households were eligible to complete a module on adult health and to have biomarker measurements and tests. […]

The sampling frame used for the SADHS 2016 is the Master Sample Frame (MSF) prepared by Statistics South Africa (Stats SA); the MSF was compiled from the Census 2011 frame. The latter is a list of 103,576 enumeration areas (EAs) that cover the whole country. An EA is a geographic area consisting of a convenient number of DUs that serve as counting units for the census. The MSF is a list of 71,241 primary sampling units (PSUs). For each PSU, the MSF contains information about location (province, district, and municipality), type of residence (urban, traditional, or farm), and estimated number of residential households.

An MSF PSU can be an EA, a group of small EAs, or part of an EA. In preparation for the MSF, out-of- scope EAs were removed from the frame. These out-of-scope areas were defined as institutional EAs and EAs that had zero DU counts at the time of the Census 2011. Furthermore, EAs with a very small number of households were excluded from the frame, which has been adjusted for during the sampling weight adjustment for under-coverage. Households in the excluded EAs accounted for less than 1% of households in the population. Moreover, all of the identified large EAs were conceptually split into one or more PSUs of equal size depending on the number of DUs in the EA. Finally, small EAs with DU counts of between 20 and 99 were pooled with neighbouring EAs that had the same geographical characteristics to form a new PSU.

Administratively, South Africa is divided into nine provinces; each province is subdivided into districts, and each district is subdivided into municipalities. […] In South Africa, about 68.3% of households are in urban areas and 31.7% in non-urban areas. […] There are a total of 71,241 PSUs in the MSF, 45,651 in urban areas, 22,214 in traditional areas, and 3,376 in farms. The average PSU size is 210 households; urban and farm PSUs are larger in size (averages of 224 and 237 households, respectively), whereas traditional PSUs have a smaller size (an average of 178 households).

The sample for the SADHS 2016 is a stratified sample selected in two stages from the MSF. Stratification was achieved by separating each province into urban, traditional, and farm areas. In total, 26 sampling strata were created (since there are no traditional areas in Western Cape). Samples were selected independently in each sampling stratum by a two-stage selection. Implicit stratification and proportional allocation were achieved at each of the lower administrative levels within a given sampling stratum by sorting the sampling frame according to administrative units at different levels in each stratum and using probability proportional to size selection at the first stage of sampling.

In the first stage, 750 PSUs were selected (468 in urban areas, 224 in traditional areas, and 58 in farm areas) with probability proportional to PSU size and with independent selection in each sampling stratum […]. A listing operation was carried out in all of the selected PSUs, and the resulting lists of DUs served as a sampling frame for the selection of DUs in the second stage. Before the listing activities, informal or congested PSUs were identified so that a segmentation process could be administered. These PSUs were divided into segments of about 20 DUs each, with only one segment selected at random for the survey. Therefore, a cluster in the SADHS 2016 was either a PSU or a segment of a PSU.

In the second stage of selection, a fixed number of 20 DUs per cluster was selected with an equal probability systematic selection from the newly created household listing. In segmented PSUs, if the segment contained 20 DUs or fewer, all DUs in the selected segment were eligible for the survey. In segments with more than 20 DUs, 20 DUs were randomly selected and were eligible for the survey. The survey interviewer interviewed only the households in the pre-selected DUs. No replacements and no changes of the pre-selected DUs were allowed in the implementing stages in order to prevent bias.

[…] To ensure that the survey precision is comparable across provinces, a power allocation was used to allocate between provinces and between different types of residence within each province. The survey was expected to be conducted in about 15,000 residential households, 9,360 in urban areas, 4,480 in traditional areas, and 1,160 in farms. The sample was expected to result in about 10,335 completed interviews with women age 15-49 and about 4,573 completed interviews with men age 15-59."

Source: *National Department of Health & ICF International. South Africa Demographic and Health Survey 2016. Available at: https://www.dhsprogram.com/publications/publication-fr337-dhs-final-reports.cfm.*

**Tajikistan DHS 2017**

"The sampling frame used for the 2017 TjDHS is the 2010 Tajikistan Population and Housing Censusconducted by the SA. Administratively, Tajikistan is divided into five regions: Dushanbe, Districts of Republican Subordination (DRS), Sughd, Khatlon, and Gorno-Badakhshan Autonomous Oblast (GBAO). Each region is subdivided into urban and rural areas. The country is divided into districts distributed over the country's regions. Each district is further divided into census divisions, which are subdivided into instruction areas. Each instruction area is divided into urban enumeration areas (EAs) or rural villages.

The sampling frame of the 2017 TjDHSis a list of EAs and natural villages covering all urban and rural areas of the country, with the primary sampling units (PSUs) being EAs in urban areas and natural villages in rural areas. An EA is a geographical area, usually a city block, consisting of the minimum number of households required for efficient counting; each EA serves as a counting unit for the population census. The sample was designed to yield representative results for the urban and rural areas separately, and for each of the four administrative regions and Dushanbe. In addition, as in the previous TjDHS survey, the sample was designed to allow certain indicators to be presented for the 12 districts in Khatlon covered under the Feed the Future program (FTF); these 12 districts have been combined as a single FTF domain. The sampling frame excluded institutional populations such as persons in hotels, barracks, and prisons. The 2017 TjDHS followed a stratified two-stage sample design.

The first stage involved selecting sample PSUs (clusters) with a probability proportional to their size within each sampling stratum. A total of 366 clusters were selected, 166 in urban areas and 200 in rural areas. The second stage involved systematic sampling of households. A household listing operation was undertaken in all of the selected clusters, and a fixed number of 22 households was selected from each cluster with an equal probability systematic selection process, for a total sample of just over 8,000 households.

All women age 15-49 who were either permanent residents of the selected households or visitors who stayed in the households the night before the survey were eligible to be interviewed. [...] Height and weight information was collected from eligible women age 15-49 and children age 0-59 months in all households."

Source: *Statistical Agency under the President of the Republic of Tajikistan, Ministry of Health and Social Protection of Population of the Republic of Tajikistan, and ICF. 2018. Tajikistan Demographic and Health Survey 2017. Dushanbe, Republic of Tajikistan, and Rockville, Maryland, USA: Statistical Agency under the President of the Republic of Tajikistan (SA), Ministry of Health and Social Protection of Population of the Republic of Tajikistan (MOHSP), and ICF. Available at: https://dhsprogram.com/pubs/pdf/FR341/FR341.pdf*

**Tajikistan STEPS 2016**
"A multi-stage cluster sample of households. One individual within the age range of the survey was selected per household.
Analysis weights were calculated by taking the inverse of the probability of selection of each participant. These weights were adjusted for differences in the age-sex composition of the sample population as compared to the target population. Different weight variables are available per Step:

wStep1 - for interview data
wStep2 - for physical measures
wStep3 - for biochemical measures

This allows for differences in the weight calculation for each Step of the survey as the age-sex composition of the respondents to each Step can differ slightly due to refusal or drop out."

Source: *report not available. Sampling information obtained from WHO NCD Microdata Repository. Available at: : https://extranet.who.int/ncdsmicrodata/index.php/catalog/270/study-description#page=sampling&tab=study-desc*

**Tanzania DHS 2015-2016**

"The sample design for the 2015-16 TDHS-MIS was done in two stages and was intended to provide estimates for the entire country, for urban and rural areas in Tanzania Mainland, and for Zanzibar. For specific indicators such as contraceptive use, the sample design allowed the estimation of indicators for each of the 30 regions (25 regions from Tanzania Mainland and 5 regions from Zanzibar). The first stage involved selecting sample points (clusters), consisting of enumeration areas ( EAs ) delineated for the 2012 Tanzania Population and Housing Census. A total of 608 clusters were selected.

In the second stage, a systematic selection of households was involved. A complete households listing was carried out for all 608 selected clusters prior to the fieldwork. From the list, 22 households were then systematically selected from each cluster, yielding a representative probability sample of 13,376 households for the 2015-16 TDHS-MIS. To estimate geographic differentials for certain demographic indicators, Tanzania was divided into nine geographic zones. Although these zones are not official administrative areas, this classification system is also used by the Reproductive and Child Health Section of the MoHCDGEC. Grouping the regions into zones allowed a relatively large number of people in the denominator and a reduced sampling error. Note that the zones, defined below, differ slightly from the zones used in previous DHS surveys. Therefore, comparisons across the zones and from survey to survey should be made with caution. The zones are as follows:

- Western zone: Tabora, Kigoma
- Northern zone: Kilimanjaro, Tanga, Arusha
- Central zone: Dodoma, Singida, Manyara
- Southern Highlands zone : Iringa, Njombe, Ruvuma
- Southern zone: Lindi, Mtwara
- South West Highlands zone : Mbeya, Rukwa, Katavi
- Lake zone: Kagera, Mwanza, Geita, Mara, Simiyu, Shinyanga
- Eastern zone: Dar es Salaam, Pwani, Morogoro
- Zanzibar: Kaskazini Unguja, Kusini Unguja, Mjini Magharibi, Kaskazini Pemba, Kusini Pemba

All women age 15-49 who were either usual residents or visitors in the household on the night before the survey were included in the 2015-16 TDHS-MIS and were eligible to be interviewed. In a subsample of one-third of all the households selected for the survey, all men age 15-49 were eligible to be interviewed if they were either usual residents or visitors in the household on the night before the survey."

Source: *Ministry of Health, Community Development, Gender, Elderly and Children - MoHCDGEC/Tanzania Mainland, Ministry of Health - MoH/Zanzibar, National Bureau of Statistics - NBS/Tanzania, Office of Chief Government Statistician - OCGS/Zanzibar, and ICF. 2016. Tanzania Demographic and Health Survey and Malaria Indicator Survey (TDHS-MIS) 2015-16. Dar es Salaam/Tanzania: MoHCDGEC, MoH, NBS, OCGS, and ICF. Available at: https://dhsprogram.com/pubs/pdf/FR321/FR321.pdf*

**Tanzania STEPS 2012**

"The STEPS survey in the United Republic of Tanzania was a population-based survey of adults aged 25-64. The study used both multistage cluster and random probability sampling procedures. Fifty of 119 total districts were randomly selected as primary sampling units (PSUs). Within these PSUs, enumeration areas (EAs) of > 50 households were randomly selected. Any EA with < 50 households was merged with a neighboring EA. Within the EAs, households were randomly selected from a list of all eligible households in the EA. A total of 5762 adults participated in the Tanzania STEPS survey. Within each selected household, the Kish method was used to select the STEPS participant. This procedure was followed until the predetermined sample was obtained for the enumeration area."

Source: *Ministry of Health and Social Welfare and National Institute for Medical Research: Tanzania STEPS survey report. Available at: http://www.who.int/chp/steps/UR_Tanzania_2012_STEPS_Report.pdf?ua=1*

**Timor-Leste DHS 2016**

"The sampling frame used for the 2016 TLDHS is the 2015 Timor-Leste Population and Housing Census (2015 TLPHC) provided by the Timor-Leste GDS. The sampling frame is a complete list of enumeration areas (EAs) created for the 2015 population census. In the 2015 TLPHC, there are an average of 89 households per EA. The sampling frame contains information about the administrative unit, the type of residence, the number of residential households, and the male and female population in each of the EAs.

There are five geographic regions in Timor-Leste, and these are subdivided into 12 municipalities and special administrative region (SAR) of Oecussi. The 2016 TLDHS sample was designed to produce reliable estimates of indicators for the country as a whole, for urban and rural areas, and for each of the 13 municipalities. A representative probability sample of approximately 12,000 households was drawn; the sample was stratified and selected in two stages. In the first stage, 455 EAs were selected with probability proportional to EA size from the 2015 TLPHC: 129 EAs in urban areas and 326 EAs in rural areas. In the second stage, 26 households were randomly selected within each of the 455 EAs; the sampling frame for this household selection was the 2015 TLPHC household listing available from the census database. It was decided not to conduct a standard DHS household listing operation because the 2015 TLPHC listing was less than a year old and there were constraints on the survey's funding and timeline.

In the list of households provided by the 2015 TLPHC, each dwelling was identified by a unique number, its GIS coordinates, and a computerized map indicating the dwelling's position. At the time of fieldwork, GDS also provided the names of the household heads for the selected households. These data were uploaded to the tablet computers used for data collection to assist survey teams in locating the selected households. Interviewers only contacted pre-selected households. The sample design and sample size calculations took into consideration anticipated rates of non-response at the household and individual levels. No replacements or changes of the pre-selected households were allowed in order to prevent bias. Because of the non-proportional sample allocation to the sampling strata and the fixed sample size per cluster, the survey is not self-weighting. The resulting data have, therefore, been weighted to be representative at the national, urban/rural, and municipality levels.

All selected households were eligible for an interview with the Household Questionnaire. All women age 15-49 and children age 0-59 months who were either permanent residents of the selected households or visitors who stayed in the household the night before the survey were eligible for anthropometric measurements, and the women were eligible for individual interview. In one-third of the sampled households, all men age 15-59, including both usual residents and visitors who stayed in the household the night before the interview, were eligible for individual interview. In the subsample of households selected for the men's interview, women age 15-49, men age 15-49, and children age 0-59 months were eligible for anthropometric measurements."

<u>Source:</u> *General Directorate of Statistics (GDS), Ministry of Health and ICF. 2018. Timor-Leste Demographic and Health Survey 2016. Dili, Timor-Leste and Rockville, Maryland, USA: GDF and ICF. Available at: https://dhsprogram.com/pubs/pdf/FR329/FR329.pdf*

**Timor-Leste STEPS 2014**
"Note: Data from Census 2010 were used for all sampling considerations. Even though planning and mapping for 2015 Census is ongoing, data from the Census will only be available after July 2015.

*STEP 1: Selection of Enumeration Area*

1. List of EA with number of HH by district for Census 2010 was obtained from the Directorate of Statistics. There are 1826 EAs in Timor-Leste. Out of these, 150 EAs were selected.
2. The number of EAs to be selected from each district was based on their proportion in the country's population as per Census 2010.
3. The numbers of Households (HH) per EAs varied from 0 to more than 300. Therefore, probability proportion to size (PPS) was used.
4. For each district, the EAs were arranged in ascending order of HH size.
5. Sampling interval was obtained by dividing the total number of HH in the district by the number of EA to be selected from that district.
6. A random number was generated between one and the sampling interval for that district, using tools available at random.org.
7. The EA where that random number fell was the first EA to be selected.
8. Subsequently, the sampling interval was added to the random number and the EA where this new number fell was selected. For the next number, the sampling interval was added to the number and so on, till the population of HH was exhausted or target number of EA achieved.
9. This was done separately for each district.
10. The final list was compiled and had 150 EAs. These are spread over about 125 sucos.

*STEP 2. Selection of Households in an Enumeration Area*
Listing the house numbers to be visited

1. It was decided to use the 2010 HH size of each EA. Based on past experience, it was expected that the increase would be on an average about 4–5%.
2. The list of households to be selected by enumerators was decided centrally.
3. Sampling interval was calculated by dividing the total number of households in the EA by 18.
4. The first HH number was selected randomly by reading the last two digits of a currency note. If the number represented by the two digits was more than 18, the last digit was taken into consideration. For each EA, a different currency note was used. This could also be done it by using the tool at random.org. or by draw of lots.
5. The subsequent HH are identified by adding the sampling interval as was done for selection of EA."

Source: *Ministry of Health Timor-Leste & World Health Organization Regional Office for South-East Asia. National Survey for Non-Communicable Disease Risk Factors and Injuries: Using WHO STEPS approach in Timor-Leste – 2014. Available at: https://www.who.int/ncds/surveillance/steps/Timor-Leste_2014_STEPS_Report.pdf.*

**Togo DHS 2013-2014**
The original report is in French. We translated the sampling strategies using Google translate.

"The EDST-III aims to produce representative results at the level of the whole country, at the level of urban and rural areas separately, at the level of the city of Lomé, and at the level of each of the five regions of Togo. To do this, the national territory was divided into six study areas corresponding to the five regions and the city of Lomé and, in each study area (except Lomé), two strata were created: the urban environment stratum. and that of rural areas. The sample has been drawn stratum by stratum. Thus, the EDST-III sample is based on a two-stage stratified and drawn area survey.

At the first stage, 330 Primary Survey Units (UPS) or clusters were drawn from the list of Enumeration Areas (EA) established during the General Population and Housing Census (RGPH) carried out in 2010 by the DGSCN, by carrying out a systematic selection with probability proportional to the size, the size of the UPS being the number of households. A count of the households in each of these clusters provided a list of households from which was drawn, in the second stage, a sample of 30 households per cluster with a systematic drawing at equal probability.

A total of 9,899 households (3,840 urban in 128 clusters and 6,059 rural in 202 clusters) were selected. All women aged 15-49 usually living in selected households, or present the night before the survey, were eligible to be surveyed. In addition, in a subsample of one in two households, all men aged 15-59 were eligible to be surveyed. [...] In this same sub-sample, all women as well as all children under the age of five were eligible to be measured and weighed to assess their nutritional status."

Source: *Ministère de la Planification, du Développement et de l'Aménagement du Territoire - MPDAT/Togo, Ministère de la Santé - MS/Togo and ICF International, 2015. Enquête Démographique et de Santé au Togo 2013-2014. Rockville,Maryland, USA: MPDAT, MS and ICF International. Available at: https://dhsprogram.com/pubs/pdf/FR301/FR301.pdf*

**Togo STEPS 2010**
The original report is in French. We translated the sampling strategies using Google translate.

"Those included in this survey are male or female subjects, living in urban or rural areas, aged 15 to 64 on the day of the survey, residing in the enumeration area for at least 6 months and having given their informed consent to participate in this study. [...] Three hundred clusters were randomly selected in a systematic draw with probability proportional to the size of the cluster (number of households) in the 4620 areas of enumeration of the DGSCN (General Directorate of Statistics and National Accounts) sampling frame. In order to obtain the 4,800 households at the rate of 1 individual / household, 16 households per cluster were randomly selected at the second stage of survey. In each of the selected households, one individual was selected as a survey participant via the Kish Method. A household was defined as the group of persons, who regularly share the main meal (regardless of their relationship). Households were not replaced in the event of a refusal or two unsuccessful visits to the eligible person selected by Kish's method. If the selected person was unwell or not present at the time of the interview, the investigators either tried to find a new appointment or searched for the respondent."

Source: *Service de Maladies Non Transmissibles de la Division de l'épidémiologie: Rapport final de l'enquête STEPS Togo 2010. Available at: http://www.who.int/chp/steps/2010STEPS_Report_Togo_FR.pdf?ua=1.*

**Tonga STEPS 2017**
"Multi-stage clustering sample design was used to produce representative data for the targeted age range. Analysis weights were calculated by taking the inverse of the probability of selection of each participant. These weights were adjusted for differences in the age-sex composition of the sample population as compared to the target population. Different weight variables are available per Step:

wStep1 - for interview data
wStep2 - for physical measures
wStep3 - for biochemical measures

This allows for differences in the weight calculation for each Step of the survey as the age-sex composition of the respondents to each Step can differ slightly due to refusal or drop out."

Source: *no report or fact sheet available. Sampling information obtained from WHO NCD Microdata Repository. Available at: : https://extranet.who.int/ncdsmicrodata/index.php/catalog/713/study-description#page=sampling&tab=study-desc*

**Turkey DHS 2013**

"In the selection of the TDHS-2013 sample, a weighted, multi-stage, stratified cluster sampling approach was used. Sample selection for the TDHS-2013 was undertaken in two stages. The first stage of selection included the selection of blocks as primary sampling units from each strata and this task was requested from the TURKSTAT. The frame for the block selection was prepared using information on the population sizes of settlements obtained from the 2012 Address Based Population Registration System. Settlements with a population of 10,000 and more were defined as "urban", while settlements with populations less than 10,000 were considered "rural" for purposes of the TDHS-2013 sample design. Systematic selection was used for selecting the blocks; thus settlements were given selection probabilities proportional to their sizes. Therefore more blocks were sampled from larger settlements.

The second stage of sample selection involved the systematic selection of a fixed number of households from each block, after block lists were obtained from TURKSTAT and were updated through a field operation; namely the listing and mapping fieldwork. Twenty-five households were selected as a cluster from urban blocks, and 18 were selected as a cluster from rural blocks. The total number of households selected in TDHS-2013 is 14,490.

The total number of clusters in the TDHS-2013 was set at 642. Block level household lists, each including approximately 100 households, were provided by TURKSTAT, using the National Address Database prepared for municipalities. The block lists provided by TURKSTAT were updated during the listing and mapping activities.

All women at ages 15-49 who usually live in the selected households and/or were present in the household the night before the interview were regarded as eligible for the Women's Questionnaire and were interviewed. All analysis in this report is based on de facto women."

Source: *Hacettepe University Institute of Population Studies, 2014, 2013 Turkey Demographic and Health Survey. Hacettepe University Institute of Population Studies, T.R. Ministry of Development and TÜBITAK, Ankara, Turkey. Available at: https://dhsprogram.com/pubs/pdf/FR352/FR352.pdf*

**Tuvalu STEPS 2015**
"The Tuvalu STEPS Survey was a population based cross-sectional survey of 18-69 year olds. Analysis weights were calculated by taking the inverse of the probability of selection of each participant. These weights were adjusted for differences in the age-sex composition of the sample population as compared to the target population. Different weight variables are available per Step:

wStep1 - for interview data
wStep2 - for physical measures
wStep3 - for biochemical measures

This allows for differences in the weight calculation for each Step of the survey as the age-sex composition of the respondents to each Step can differ slightly due to refusal or drop out. Additionally, some countries perform subsampling for Step 2 and/or Step 3. When no subsampling is done and response rates do not differ across Steps of the survey, the 3 weight variables will be the same."

Source: *no report or fact sheet available. Sampling information obtained from WHO NCD Microdata Repository. Available at: : https://extranet.who.int/ncdsmicrodata/index.php/catalog/639/study-description#page=overview&tab=study-desc*

**Uganda DHS 2016**

"The sampling frame used for the 2016 UDHS is the frame of the Uganda National Population and Housing Census (NPHC), conducted in 2014;the sampling frame was provided by the Uganda Bureau of Statistics. The census frame is a complete list of all census enumeration areas (EAs) created for the 2014 NPHC. In Uganda, an EA is a geographic area that covers an average of 130 households. The sampling frame contains information about EA location, type of residence (urban or rural), and the estimated number of residential households. At the time of the NPHC, Uganda was divided administratively into 112 districts, which were grouped for this survey into 15 regions. The sample for the 2016 UDHS was designed to provide estimates of key indicators for the country as a whole, for urban and rural areas separately, and for each of the 15 regions. Estimates are also presented for three special areas: the Lake Victoria islands, the mountain districts, and greater Kampala.

The 2016 UDHS regions include the following districts:

- South Central: Butambala, Gomba, Mpigi, Bukomansimbi, Kalangala, Kalungu, Lwengo, Lyantonde, Masaka, Rakai, Sembabule, and Wakiso
- North Central: Buikwe, Buvuma, Kayunga, Kiboga, Kyankwanzi, Luwero, Mityana, Mubende, Mukono, Nakaseke, and Nakasongola
- Kampala: Kampala
- Busoga: Bugiri, Namutumba, Buyende, Iganga, Jinja, Kaliro, Kamuli, Luuka, Mayuge, and Namayingo
- Bukedi: Budaka, Butaleja, Kibuku, Pallisa, Tororo, and Busia
- Bugisu: Bulambuli, Kapchorwa, Kween, Bududa, Manafwa, Mbale, Sironko, and Bukwo
- Teso:Amuria, Bukedea, Katakwi, Kumi, Ngora, Soroti, Kaberamaido, and Serere
- Karamoja:Abim, Amudat, Kaabong, Kotido, Moroto, Nakapiripirit, and Napak
- Lango:Alebtong, Amolatar, Dokolo, Lira, Otuke, Apac, Kole, and Oyam
- Acholi:Agago, Amuru, Gulu, Lamwo, Pader, Kitgum, and Nwoya
- West Nile: Adjumani, Arua, Koboko, Maracha, Moyo, Nebbi, Yumbe, and Zombo
- Bunyoro: Buliisa, Hoima, Kibaale, Kiryandongo, and Masindi
- Tooro: Bundibugyo, Kabarole, Kasese, Ntoroko, Kyenjojo, Kamwenge, and Kyegegwa
- Kigezi: Kabale, Kisoro, Kanungu, and Rukungiri
- Ankole: Buhweju, Bushenyi, Ibanda, Isingiro, Kiruhura, Mbarara, Mitooma, Ntungamo, Rubirizi, and Sheema.

The 2016 UDHS special areas include the following:

- Islands: islands and shoreline areas in Kalangala, Mayuge, Buvuma, Namayingo, Rakai, Mukono,and Wakiso districts.
- Mountains: Bundibugyo, Kasese, Ntoroko, Bukwo, Bulambuli, Kapchorwa, Kween, Kisoro, Sironko, Mbale, and Kaabong districts
- Greater Kampala: Kampala district and urban areas in Mukono and Wakiso districts.

The 2016 UDHS sample was stratified and selected in two stages. In the first stage, 697 EAs were selected from the 2014 Uganda NPHC: 162 EAs in urban areas and 535 in rural areas. One cluster from Acholi subregion was eliminated because of land disputes.

Households constituted the second stage of sampling. A listing of households was compiled in each of the 696 accessible selected EAs from April to October 2016, with some listing overlapping with fieldwork. Maps were drawn for each of the sampled clusters and all of the listed households. The listing excluded institutional living arrangements such as army barracks, hospitals, police camps, and boarding schools. To minimise the task of household listing, each large EA (i.e., more than 300 households) selected for the 2016 UDHS was segmented. Only one segment was selected for the survey with probability proportional to segment size,and the household listing was conducted only in the selected segment. Thus, a 2016 UDHS cluster is either an EA or a segment of an EA.

In total, a representative sample of 20,880 households (30 per EA or EA segment) was randomly selected for the 2016 UDHS. The allocation of the sample EAs featured a power allocation with a small adjustment because a proportional allocation would not have met the minimum number of clusters per survey domain required for a DHS survey. The sample EAs were selected independently from each stratum using probability proportional to size. The 20,880 selected households resulted in 18,506 women successfully interviewed, with an average of 1,200 complete interviews per domain. All women age 15-49 who were either permanent residents of the selected households or visitors who stayed in the household the night before the survey were eligible to be interviewed. In one-third of the sampled households, all men age 15-54, including both usual residents and visitors who stayed in the household the night before the interview, were eligible for individual interviews. [...] Height and weight information was also collected from eligible women and men, as well as children age 0-59 months."

Source*: Uganda Bureau of Statistics - UBOS and ICF. 2018. Uganda Demographic and Health Survey 2016. Kampala, Uganda and Rockville, Maryland, USA: UBOS and ICF. Available at: https://dhsprogram.com/pubs/pdf/FR333/FR333.pdf*

**Uganda STEPS 2014**

"A two stage sampling design was used to draw the sample. At the first stage, Enumeration Areas (EAs) were drawn with Probability Proportional to Size (PPS), and at the second stage, households which were the ultimate sampling units were drawn using Simple Random Sampling (SRS). A total of 350 EAs were selected from 2014 Uganda Population and Housing Census Mapping Frame. At the EA level, the target was 14 households.

*Sample frame:* The 2014 Uganda NCD survey used a sampling frame of the 2014 Population Census Mapping listing provided by the Uganda Bureau of Statistics (UBOS). The UBOS has an electronic file consisting of 78,950 Enumeration Areas (EAs) created for the 2014 Population and Housing Census. An EA is a geographic area consisting of a convenient number of dwelling units that serve as counting units for the census. Tables A.1 provides information on the distribution of EAs and households in the sampling frame by region and residence. The table shows that among the 78,950 EAs, 13,087 (22%) are in urban areas and 65,863 (78%) are in rural areas. The average size of an EA, measured in number of households, is 95 in an urban EA and 77 in a rural EA, with an overall average of 79.

*Sample size:* The size required for the sample was determined by taking into consideration several factors:-the three most important being (i) the degree of precision (reliability) desired for the survey estimates, (ii) the cost and operational limitations, and (iii) the efficiency of the design. The Uganda NCD STEP 1 and 2 complete with completed information were 3987 and 3720 with all the three STEPS 1,2 & 3."

Source: *Ministry of Health: Non-communicable disease risk factor baseline survey. Uganda 2014 report. Available at: https://extranet.who.int/ncdsmicrodata/index.php/catalog/633*

**Uruguay GATS 2009**
“A multi-stage, geographically clustered sample design was used to produce nationally representative data. One individual was randomly chosen from each selected household to participate in the survey.”

Source: *no report available. Sampling information obtained from the fact sheet. Available at:* https://www.who.int/tobacco/surveillance/fact_sheet_of_gats_uruguay_2010.pdf?ua=1

**Vanuatu STEPS 2011**
"The survey used a cluster sampling design where the primary sampling unit was enumeration area (EA) and the secondary sampling unit was households. All 6 provinces in Vanuatu were included in the survey. One hundred and thirteen (113) EAs were randomly selected proportion to the size of the EA from a total of 411 EAs. Forty four (44) households were then randomly selected in each EA proportional to the number of households in each EA. The selection of participants within each household was done using the Kish method. The total number of households selected by combined Enrolment Areas was 4,972."

Source: *Vanuatu Ministry of Health and WHO Western Pacifics Region: Vanuatu NCD Risk Factors STEPS Report. Available at: https://extranet.who.int/ncdsmicrodata/index.php/catalog/714*

**Vietnam GATS 2015**
*"Sample size:* 8,200. The number was calculated according to standardized GATS protocol, which is required to obtain reliable estimates of the key variables of gender and urban or rural area.

*Sampling framework* (master sample): The sample frame for GATS 2015 was a cluster frame made by the General Statistics Office (GSO) based on the frame of Population and Housing Census 2009 and updated with 2014 data. Based on the Population and Housing Census data 2009, GSO prepared a 15% master sample to serve as a future national survey sampling frame. The master sample contains 25,500 enumeration areas (EAs) from 706 of the 708 districts of Viet Nam (two island districts were excluded from the GSO master sample frame).

*Sampling method* of GATS is two-stage stratified sampling. The master sample frame of GSO was divided by two stratification variables: 1. urbanization (1 = urban; 2 = rural) and district group (1 = district, town, or city of a province; 2 = plain or coastal district; 3 = mountainous or island district), which means that the master sample frame was divided into six sample frames or six strata:

1. Districts, towns, or cities of a province in an urban area.
2. Plain or coastal districts in an urban area.
3. Mountainous or island districts in an urban area.
4. Districts, towns, or cities of a province in a rural area.
5. Plain or coastal districts in a rural area.
6. Mountainous or island districts in a rural area.

*Sampling stages:*

- In the first stage of sampling, the primary sampling unit (PSU) was an enumeration area (EA).
- In the second stage, households in the EAs were selected; then individuals from each household were selected for interview.

*Selection of EAs:* The number of households in each EA in urban areas was 133 and in the rural was 120; 10% of households in each EA was selected. Thus, 13 households in each EA in urban areas and 12 households in each EA in rural areas were selected. To ensure the number of individuals to be interviewed in the urban was the same as the number interviewed in the rural area (4,100 each), the number of EAs selected in the urban was 4,100/13 = 315 EAs and number of EAs selected in the rural area was 4,100/12 = 342 EAs. The number of EAs allocated for each stratum was estimated on the basis of the probability-proportional-to-size (PPS) sampling method by using number of households in each stratum and total number of EAs selected in the urban (U) and in the rural (R) areas [...]

*Selection of households:* From the list all households in each EA, households were selected by using a systematic random sampling method (13 households in each EA in urban areas and 12 households in each EA in rural areas plus the nonresponse rate from previous survey GATS in 2010. Thus, the number of households selected in each urban EA was 15 and in each rural EA was 14). The total number of households for GATS 2015 was 9,513 (4,725 in urban areas and 4,788 in rural areas). Selection of individuals for interview (from each selected household): all household members were listed and one eligible household member from each selected household was then randomly chosen for the interview. Note that the current design and the design in which EAs were sampled directly from the master sample were analogous. The selection probability of an eligible individual was calculated as a product of selection probability for each stage."

Source: *Ministry of Health of Vietnam, Hanoi Medical University, General Statistics Office, Centers for disease control and prevention and WHO: Global Adult Tobacco Survey - Vietnam 2015. https://www.who.int/tobacco/surveillance/survey/gats/vietnam-country-report-2015.pdf?ua=1*

**Yemen DHS 2013**

"To have enough cases to report on key indicators in each of the 21 reporting domains, the smallest governorates in terms of population were oversampled while the largest were undersampled. The 2004 General Population Housing and Establishment Census was used as the sampling frame. During the 2004 census, the country was divided into areas convenient for data collection called census enumeration areas (EAs). The primary sampling unit (PSU), referred to as a cluster for the 2013 YNHDS, is defined on the basis of EAs from the 2004 EA census frame. The 2013 YNHDS sample was selected using a stratified two-stage cluster design consisting of 800 clusters, with 213 in urban areas and 587 in rural areas.

A complete listing of households and a mapping exercise were carried out for each cluster from November 10 to November 30, 2012, with the resulting lists of households serving as the sampling frame for the selection of households in the second stage. All households were listed. In each rural cluster, one household was randomly selected. This household and the next 24 households on the list together constituted the household sample for each of the 587 rural clusters; in urban clusters, the 25 households were randomly selected. The total of 800 clusters was estimated to yield a sample of 20,000 households at the national level. However, for security reasons, ten clusters were not listed. All ever-married and never-married women age 15-49 in each selected household were eligible to be interviewed."

Source: *Ministry of Public Health and Population (MOPHP), Central Statistical Organization (CSO) [Yemen], Pan Arab Program for Family Health (PAPFAM), and ICF International. 2015. Yemen National Health and Demographic Survey 2013. Rockville, Maryland, USA: MOPHP, CSO, PAPFAM, and ICF International. Available at: https://dhsprogram.com/pubs/pdf/FR296/FR296.pdf*

**Zambia DHS 2018**

"The sampling frame used for the 2018 ZDHS is the Census of Population and Housing (CPH) of the Republic of Zambia, conducted in 2010 by ZamStats. Zambia is divided into 10 provinces. Each province is subdivided into districts, each district into constituencies, and each constituency into wards. In addition to these administrative units, during the 2010 CPH each ward was divided into convenient areas called census supervisory areas (CSAs), and in turn each CSA was divided into enumeration areas (EAs). An enumeration area is a geographical area assigned to an enumerator for the purpose of conducting a census count; according to the Zambian census frame,each EA consists of an average of 110 households.

The current version of the EA frame for the 2010 CPH was updated to accommodate some changes in districts and constituencies that occurred between 2010 and 2017. The list of EAs incorporates census information on households and population counts. Each EA has a cartographic map delineating its boundaries, with identification information and a measure of size, which is the number of residential households enumerated in the 2010 CPH. This list of EAs was used as the sampling frame for the 2018 ZDHS. The 2018 ZDHS followed a stratified two-stage sample design. The first stage involved selecting sample points (clusters) consisting of EAs. EAs were selected with a probability proportional to their size within each sampling stratum. A total of 545 clusters were selected. The second stage involved systematic sampling of households. A household listing operation was undertaken in all of the selected clusters. During the listing, an average of 133 households were found in each cluster, from which a fixed number of 25 households were selected through an equal probability systematic selection process, to obtain a total sample size of 13,625 households. Results from this sample are representative at the national, urban and rural, and provincial levels. All women age 15-49 and men age 15-59 who were either permanent residents of the selected households or visitors who stayed in the households the night before the survey were eligible to be interviewed."

Source: *Zambia Statistics Agency, Ministry of Health (MOH) Zambia, and ICF. 2019. Zambia Demographic and Health Survey 2018. Lusaka, Zambia, and Rockville, Maryland, USA: Zambia Statistics Agency, Ministry of Health, and ICF. Available at: https://dhsprogram.com/pubs/pdf/FR361/FR361.pdf*

**Zambia STEPS 2017**

"To ensure that the sample reflected the entire country of Zambia, a multi-stage cluster sampling technique was used to select a nationally representative sample of adults in Zambia aged 18 to 69 years. It was decided to utilize the household listing from the Zambia Population Based HIV Impact Assessment (ZAMPHIA) - a household-based national survey that was conducted between March and August 2016 in order to measure the status of Zambia's national HIV response. ZAMPHIA offered the most pragmatic up to date and accessible national household listing to be used as the sampling frame for this survey. The ZAMPHIA survey included 60,581 households drawn from 1,103 clusters referred to in this report as standard enumeration area (SEA) (Table 2.4.1). Thus the sample drawn for the STEPS survey was a subsample of the households selected for the ZAMPHIA survey. In the first stage of sampling, SEAs were selected from each province using probability proportional to size (PPS). In the second stage, 15 households in rural SEAs and 20 households in urban SEAs were selected systematically using appropriate sampling interval based on the number of households in that SEA. These households constituted the final list of households for the STEPS survey prepared for the field investigators (FI). In the third stage, while the FI approached the household and sought consent, all eligible members in the household were entered into the Android-based devise used for the survey. The device then selected one member from the eligible members using a simple random sampling technique. The selected member was then interviewed having gone through the ethical process of consent after being provided with information on the survey. If the selected member was not available, a scheduled visit was made. If the selected member could not be reached after two scheduled visits he or she was considered as non-response. There was no replacement strategy so as to maintain the integrity and representativeness of the sample."

Source: *Ministry of Health, Republic of Zambia: Zambia STEPS for non communicable diseases risk factors. Zambia Report for 2017 Available at: https://extranet.who.int/ncdsmicrodata/index.php/catalog/620*

**Zanzibar STEPS 2011**
"The survey took place in June and July 2011, followed by data cleaning and analysis. [...]

*Sampling Frame:* The target population is the entire population in Zanzibar whereby the whole of Zanzibar was selected as the survey site, and hence all districts included. The total population is estimated to be 1.2 million distributed unevenly between 10 districts. The sampling frame represented the entire population in Zanzibar.

*Sampling Technique:* The sampling strategy used is a multi-stage cluster sampling with stratification. The ten districts are considered as different strata, and the total number of primary sampling units, PSU, is allocated proportionately across all strata. Each district is divided into smaller clusters. These clusters are the geographical and administrative units called Shehia. The Shehia are divided into smaller clusters called zones (also called mitaa, vitongoji, or vijiji) which typically consist of 100-300 households. Zones smaller than that were merged to make up one larger cluster, and zones much larger were split in smaller clusters.

At the first stage clusters were selected using Simple Random Selection, SRS, from the list of clusters (Shehia) within each district. At the second stage clusters (zones) were randomly selected using probability proportionate to size (PPS). At the third stage households were randomly selected from the household lists provided by the administrative leader of the Shehia.

The two last stages of sampling was done using the software STEPSsampling.xls from WHO. Finally participants were selected from the household using Kish method. The household lists were complete and included households with no eligible participants for the survey. Therefore an extra 7 households were sampled at third stage in each cluster for replacement in case a selected household had no eligible participants and had to be changed. This was done before data collectors went to the cluster. Resources allowed for 100 PSU which was why 2800/100 = 28 households were selected from each PSU (and disproportionate from each SSU)."

Source: *Ministry of Health Zanzibar: NCD Survey Report. Main findings from the National Non-communicable Disease Risk Factor Survey 2011. Available at: https://extranet.who.int/ncdsmicrodata/index.php/catalog/621*

**Zimbabwe DHS 2015**

"The 2015 ZDHS sample was designed to yield representative information for most indicators for the country as a whole, for urban and rural areas, and for each of Zimbabwe's ten provinces: Manicaland, Mashonaland Central, Mashonaland East, Mashonaland West, Matabeleland North, Matabeleland South, Midlands, Masvingo, Harare, and Bulawayo. The 2012 Zimbabwe Population Census was used as the sampling frame for the 2015 ZDHS.

Administratively, each province in Zimbabwe is divided into districts, and each district is divided into smaller administrative units called wards. During the 2012 Zimbabwe Population Census, each ward was subdivided into convenient areas, which are called census enumeration areas (EAs). The 2015 ZDHS sample was selected with a stratified, two-stage cluster design, with EAs as the sampling units for the first stage. The 2015 ZDHS sample included 400 EAs—166 in urban areas and 234 in rural areas.

The second stage of sampling included the listing exercises for all households in the survey sample. A complete listing of households was conducted for each of the 400 selected EAs in March 2015. Maps were drawn for each of the clusters and all private households were listed. The listing excluded institutional living arrangements such as army barracks, hospitals, police camps, and boarding schools. A representative sample of 11,196 households was selected for the 2015 ZDHS.

Women age 15-49 and men age 15-54 who were either permanent residents of the selected households or visitors who stayed in the household the night before the survey were eligible for interviewing."

<u>Source:</u> *Zimbabwe National Statistics Agency & ICF International. Zimbabwe Demographic and Health Survey 2015. Available at: https://dhsprogram.com/pubs/pdf/FR322/FR322.pdf.*

### Materials and Methods S3: Country-specific Measurement Methods

#### *3.1 Hypertension*

Blood pressure was measured with a digital upper arm meter in 37 countries. One survey applied a digital wrist meter, while one used a manual mercury sphygmomanometer. For 11 countries, the measurement device could not be determined due to missing survey reports or, if available, a lack of specification.

Table S2. Hypertension measurement by country

| Measurement device | Country | Number of measure-ments | Interval between measurements |
|---|---|---|---|
| Omron M6 Comfort and Omron HEM-711ac; Life Source Model UA-789 for individuals with a wide upper arm circumference | Albania | 3 | 10 minutes |
| Life Source UA-767 Plus Digital Monitor | Bangladesh | 3 | 10 minutes |
| Boso-Medicus Uno | Benin | 3 | 3 minutes |
| Boso Medicus Control | Bhutan | 3 | 3 minutes |
| Not specified | Botswana | 3 | Not specified |
| G-TECH Model MA 100 | Brazil | 3 | 2 minutes |
| Not specified | Cabo Verde | 3 | Not specified |
| Manual mercury sphygmomanometer | China | 3 | 10 minutes |
| Digital upper arm meter (model not specified) | Comoros | 2 | 5-10 minutes |
| Digital upper arm meter (model not specified) | Ecuador | 3 | Not specified |
| Life Source Digital Monitors Model UA-767V; Life Source Digital Monitors Model UA-789 for individuals with a wide upper arm circumference | Egypt | 3 | 10 minutes |
| Boso Medicus PC (model not specified) | Eswatini | 3 | 3-5 minutes |
| Boso-Medicus Uno | Ethiopia | 3 | 3 minutes |
| Not specified | Gambia | 3 | Not specified |
| Boso Medicus Uno | Georgia | 3 | 3 minutes |
| Boso Medistar Wrist Blood Pressure Monitor Model S | Ghana | 3 | 1 minute |
| Omron Digital Monitor HEM-8712 | India | 3 | 5 minutes |

| | | | |
|---|---|---|---|
| Omron Digital Monitor HEM-7203 | Indonesia | 3 | First measurement taken at beginning of interview, subsequent two taken during the course of the interview |
| Beurer medical, type: BM 20 | Iran | 3 | 5 minutes |
| Omron M3 Digital Monitor | Jordan | 3 | 3 minutes |
| Omron M2 Digital Monitor | Kenya | 3 | 3-5 minutes |
| Omron M4 Digital Monitor | Kiribati | 3 | 2-3 minutes |
| Digital upper arm meter (brand and model not specified) | Kyrgyz Republic | 3 | 10 minutes |
| Not specified | Lao PDR | 3 | Not specified |
| Omron M3W Digital Monitor | Lesotho | 3 | 10 minutes |
| Omron digital upper arm meter (model not specified) | Liberia | 3 | Not specified |
| Omron Digital Monitor M4 - I | Malawi | 3 | 3-5 minutes |
| Omron digital upper arm meter (model not specified) | Mexico | 2 | Not specified |
| Not specified | Mongolia | 3 | Not specified |
| Omron 3 Series Digital Monitor | Mozambique | 2 with a third measurement if the first two differed by >10 mmHg | 1 minute |
| Boso Medicus automatic digital blood pressure monitor | Myanmar | 3 | 5 minutes |
| Life Source Digital Monitor Model UA-767 | Namibia | 3 | Not specified |
| Omron (model not specified) | Niger | 3 | Not specified |
| Omron Digital Monitor HEM-7113 | Peru | 2 | 2 minutes |
| A&D Medical UA-767PC Automatic Monitor | Romania | 3 | 1 minute |
| Omron M4 Digital Automatic Blood Pressure Monitor | Rwanda | 3 | 2-3 minutes |
| Not specified | Samoa | 3 | Not specified |
| Not specified | São Tomé and Principe | 3 | Not specified |

| | | | |
|---|---|---|---|
| Not specified | Solomon Islands | 3 | Not specified |
| Omron 1300 Digital Monitor | South Africa | 3 | 3 minutes or more |
| Not specified | Tajikistan | 3 | Not specified |
| Omron digital upper arm meter (model not specified) | Tanzania | 3 | Not specified |
| Omron digital upper arm meter (model not specified) | Timor-Leste | 3 | 2 minutes |
| Omron digital upper arm meter (model not specified) | Togo | 3 | 5 minutes |
| Not specified | Tonga | 3 | Not specified |
| Not specified | Tuvalu | 3 | Not specified |
| Boso Medicus Uno | Uganda | 3 | 3-5 minutes |
| Omron M4 | Vanuatu | 3 | 2-3 minutes |
| automated blood pressure machine (model not specified) | Zambia | 3 | 3-5 minutes |
| Omron M2 Digital Monitor | Zanzibar | 3 | 5 minutes |

*In the following, please note: In order to ensure accuracy in reporting, blood pressure measurement methods are pasted verbatim from specified sources.*

**Albania DHS 2008-2009**
"Blood pressure was measured for all women and men interviewed in the households selected for the men's survey (half of all households). Interviewers were provided with the Omron HEM-711ac blood pressure monitor - a fully automatic, digital oscillometric blood pressure measuring device with automatic upper-arm inflation and automatic pressure release. Three measurements of systolic and diastolic blood pressure (measured in millimetres of mercury (mmHg)) were taken during the survey interview with an interval of at least 10 minutes between measurements. At the end of the interview, respondents were provided information about their average blood pressure reading and advised to see a medical provider for further screening if that reading fell outside normal ranges according to internationally recommended guidelines (WHO, 1999)."
*Source: Institute of Statistics, Institute of Public Health [Albania] and ICF Macro. 2010. Albania Demographic and Health Survey 2008-09. Tirana, Albania: Institute of Statistics, Institute of Public Health and ICF Macro. Available at: https://dhsprogram.com/pubs/pdf/FR230/FR230.pdf*

**Bangladesh DHS 2017-2018**
"Blood pressure (BP) measures and blood specimens for glucose testing were collected from all women and men age 18 and above (regardless of marital status) in the subsample of one-fourth of the households. All women and men identified as eligible for blood pressure measurements and glucose testing were contacted and had the test explained to them. Blood pressure measures and blood specimens for glucose testing were taken for those who consented. [...] One male health technician and one female technician from each team were engaged for biomarker testing.

The LIFE SOURCE® UA-767 Plus BP monitor was used to measure blood pressure. This automatic device includes separate cuffs (small, medium, and large). Two health technicians were trained per team according to the recommended protocol. Three blood pressure measurements were taken at intervals of approximately 10 minutes. The average of the second and third measurements was used to report respondents' blood pressure values."
*Source: National Institute of Population Research and Training (NIPORT), and ICF. 2020. Bangladesh Demographic and Health Survey 2017-18. Dhaka, Bangladesh, and Rockville, Maryland, USA: NIPORT and ICF. Available at: https://dhsprogram.com/pubs/pdf/FR344/FR344.pdf*

**Benin STEPS 2015**
The original report is in French. We translated the information using Google translate.

"Blood pressure was measured in subjects seated for at least 15 minutes (without crossing your legs), bladder empty, no coffee or any other arousal. The participant was not to speak during the measurement, the participant's elbow supported with the left arm placed on the table, palm facing up, cuff of suitable size placed above the elbow so that the band is positioned 1 - 2 cm in height. above the bend in the elbow. Three (03) measurements were taken with a 3 min rest for the participant between measurements. All the measurements were recorded in the PDA. The third measurement was recorded on the participant's measurement results sheet."
*Source: Rapport final de l'enquête pour la surveillance des facteurs de risque des maladies non transmissibles par l'approche ''STEPSwise'' de l'OMSENQUETE ''STEPS 2015'' au Bénin. Available at: https://www.who.int/ncds/surveillance/steps/benin/en/.*

**Bhutan STEPS 2014**
No further information available

**Botswana STEPS 2014**
No further information available

**Brazil Pesquisa Nacional de Saúde 2013**
The paper is in Portugese. We translated the Information using Google translate.

BP was measured by a trained team, using a calibrated digital device. Three BP measurements were made, with two-minute intervals between them. The measurements were then entered into a smartphone. The mean arterial BP between the second and third measurements was used for the present study. [...] Routines for measuring BP consisted of a protocol that included being at rest, emptying the bladder, not drinking or smoking during the 30-minute period, not performing physical activities during the one-hour period prior to the measurement, measuring in a sitting position, having rested at least five minutes before, among others. Further details can be seen in other studies. Considering that the BP may rise at the time of measurement, it is recommended that it be validated by repeated measurements, on two or more occasions, for the diagnosis of AH. However, in this study, the individual who had BP ≥ 140/90 mmHg in a single measurement was considered hypertensive due to the impossibility of further measures for the adopted sample."

*Source: Malta, Deborah & Gonçalves, Renata & Machado, Ísis & Freitas, Maria & Azeredo, Cimar & Szwarcwald, Célia. (2018). Prevalência da hipertensão arterial segundo diferentes critérios diagnósticos, Pesquisa Nacional de Saúde. Revista Brasileira de Epidemiologia. 21. 10.1590/1980-549720180021.supl.1.*

**Cabo Verde STEPS 2007**
No further information available

**China Health and Nutrition Survey 2009**
"After at least a 10-min rest, arterial BP (Korotkoff phase I for systolic and phase V for diastolic pressure) was measured on the right arm in the sitting position using a mercury sphygmomanometer with the cuff maintained at the heart level. BP was measured three times on one visit, and the mean of three readings was used in the analysis."
*Source: Liang, Y., Liu, R., Du, S., & Qiu, C. (2014). Trends in incidence of hypertension in Chinese adults, 1991-2009: the China Health and Nutrition Survey. International journal of cardiology, 175(1), 96–101. https://doi.org/10.1016/j.ijcard.2014.04.258*

**Comoros STEPS 2011**
No further information available

**Ecuador STEPS 2018**
The original report is in Spanish. We translated information using Google translate.

"The STEPS Survey required the following equipment and supplies: [...] v) digital arm blood pressure monitor with universal cuff."
*Source: Ministerio de Salud Pública: Encuesta STEPS Ecuador 2018. Vigilancia de enfermedades no transmisibles y factores de riesgo. INFORME DE RESULTADOS Available at: https://extranet.who.int/ncdsmicrodata/index.php/catalog/774*

**Egypt DHS 2015**
"Blood pressure measurements were taken for all individuals age 15-59 who consented to the procedure. The measurements were taken at three points during the interview by interviewers using fully automatic digital blood pressure monitors with upper arm automatic inflation. Prior to taking the first blood pressure reading, the interviewer measured the respondent's arm circumference in order to determine the appropriate cuff size to use. Respondents were asked for information about recent intake of food and caffeinated beverages as these factors can influence blood pressure readings. Respondents were given information on their blood pressure at the end of the interview."
*Source: Ministry of Health and Population/Egypt, El-Zanaty and Associates/Egypt, and ICF International. 2015. Egypt Health Issues Survey 2015. Cairo, Egypt: Ministry of Health and Population/Egypt and ICF International. Available at: https://dhsprogram.com/pubs/pdf/FR313/FR313.pdf*

**Eswatini STEPS 2014**
"Blood pressure measurements were taken using battery powered digital blood pressure machine Boso Medicus. Three readings were taken 3-5 minutes apart and the third reading was then recorded."

*Source: Swaziland Government. Ministry of Health: WHO STEPS. Noncommunicable Disease Risk Factor Surveillance Report Swaziland 2014. Available at: http://www.who.int/chp/steps/swaziland/en/.*

**Ethiopia STEPS 2015**

"Blood pressure and heart rate measurements were taken three times on the right arm of the survey
participants in a sitting position, using a Boso-Medicus Uno instrument with a universal cuff and automatic blood pressure and heart rate monitor. The mean of three measurements was taken for analysis. The measurements were taken after the participant had rested for 15 minutes, and each with three minutes of rest between the measurements (maximum deviation of cuff pressure measurement ± 3 mmHg, and of
pulse rate display ± 5%)."

*Source: Ethiopian Public Health Institute 2016: Ethiopia STEPS report on risk factors for non-communicable diseasesand prevalence of selected NCDs. Available at: https://extranet.who.int/ncdsmicrodata/index.php/catalog/794/download/5522*

**Gambia STEPS 2010**

No further information available

**Georgia STEPS 2016**

The original report is in Georgian. We translated the information using Google translate.

"Blood pressure monitors were used: Bosch and Sohn Medicus UNO. Blood pressure was measured for each respondent. The heart rate was measured along with the diastolic (DBP) and systolic blood pressure (SBP) numbers displayed on the pressure gauge. The data used in the data analysis is the arithmetic mean of the indicators."

*Source: National Center for Disease Control and Public Health. Georgia STEPS survey 2016. Final report. Available at: https://extranet.who.int/ncdsmicrodata/index.php/catalog/223*

**Ghana Study on global AGEing and adult health (SAGE) 2008-2009**

"Blood pressure was measured three times on the right arm/wrist of the seated respondent using a wrist blood pressure monitor. Out of three measurements, an average of the latter two measurements was used as the blood pressure value in this analysis."

*Source: Wu, F., Guo, Y., Chatterji, S. et al. Common risk factors for chronic non-communicable diseases among older adults in China, Ghana, Mexico, India, Russia and South Africa: the study on global AGEing and adult health (SAGE) wave 1. BMC Public Health 15, 88 (2015). https://doi.org/10.1186/s12889-015-1407-0*

**India DHS 2015-2016**

"Blood pressure was measured for eligible women age 15-49 and (in the state module subsample of households only) eligible men age 15-54, using an Omron Blood Pressure Monitor to determine the prevalence of hypertension. Blood pressure measurements for each respondent were taken three times with an interval of five minutes between readings. Respondents whose average systolic blood pressure (SBP) was >140 mm Hg or average diastolic blood pressure (DBP) was >90 mm Hg were considered to have elevated blood pressure readings and they were encouraged to see a doctor for a full evaluation."

*Source: International Institute for Population Sciences & ICF International. National Family and Health Survey (NFHS-4) 2015-16. Available at: https://dhsprogram.com/pubs/pdf/FR339/FR339.pdf.*

**Indonesia Family Life Survey 2014-2015**
"The measurements started with blood pressure (members 15 and older), taken three times on alternate arms. [...] An Omron meter, HEM-7203 was used for taking blood pressure. Most of the time a normal sized cuff was used. Large cuffs were available is needed."
*Source: Strauss, J, Witoelar F, and Sikoki, B. The Fifth Wave of the Indonesia Family Life Survey (IFLS5): Overview and Field Report. March 2016. WR-1143/1-NIA/NICHD. Available at: https://www.rand.org/content/dam/rand/pubs/working_papers/WR1100/WR1143z1/RAND_WR1143z1.pdf*

**Iran STEPS 2016**
"The physical evaluations included measurements of height, weight, waist size, hip size, blood pressure, pulse and step counting, which was conducted by female questioners for women and male questioners for men after the participants' physical readiness and by using similar tools available in step 2 packages previously provided for the questioning team. [...] the questioning teams were obliged to use tools included in the package for step 2 provided for them by the central headquarters. Furthermore, the accuracy of the pressure gauges was checked according to step 2's protocol. In case of any failure in the equipment, supple-mentary equipment was used and the faulty ones were sent to the central headquarters to be exchanged."
*Source: Non Communicable Diseases Research Center, Endocrinology and Metabolism Research Institute of Tehran University of Medical Sciences: Atlas of non-communicable diseases risk-factors surveillance in the islamic republic of Iran. STEPs 2016. Available at: https://www.who.int/ncds/surveillance/steps/STEPS_2016_Atlas_EN.pdf?ua=1*

**Jordan STEPS 2019**
"Omron M3 devices were used to measure blood pressure. [...] After completing STEP 1, the data collector resumed with taking the physical measurements of STEP 2. This entailed taking blood pressure measurements (3 times, at a 3-minute interval), the weight, height, waist and hip circumference. Blood pressure measurements were taken using digitalized blood pressure monitors."
*Source: Centre of Strategic Studies - University of Jordan, Jordan Ministry of Health, World Health Organization: Jordan National Stepwise Survey (STEPs) for Noncommunicable Diseases Risk Factors 2019. STEPs Technical Report. Available at: https://extranet.who.int/ncdsmicrodata/index.php/catalog/853/download/6010 and WHO NCD Microdata Repository. Available at: https://extranet.who.int/ncdsmicrodata/index.php/catalog/853/study-description#metadata-data_collection*

**Kenya STEPS 2015**
"STEP 2 included Physical Measurements on Blood Pressure, Heart Rate, Height, Weight, and Waist & Hip Circumference. [...] Blood pressure and pulse rate was taken three times using an automated blood pressure measuring instrument (OMRON®)."

*Source: Ministry of Health Kenya, Division of Noncommunicable Diseases, Kenya National Bureau of Statistics, & World Health Organization. WHO: Kenya STEPwise Survey for Non Communicable Diseases Risk Factors 2015 Report. Available at: http://www.who.int/chp/steps/Kenya_2015_STEPS_Report.pdf?ua=1.*

**Kiribati STEPS 2015**
"The OMRON M4 Digital Automatic Blood Pressure Monitor was used to measure resting blood pressure. Blood pressure was measured three times - the first reading followed by two more measurements taken in 2-3 minute intervals. The three readings of the blood pressure were recorded, and the average of the second and third readings was used in the analysis."
*Source: Ministry of Health and Medical Services and WHO Western Pacific Region: Kiribati NCD risk factors STEPS report 2015-2016 - in collaboration with the WHO. Available at: https://extranet.who.int/ncdsmicrodata/index.php/catalog/724*

**Kyrgyz Republic DHS 2012**
"To measure blood pressure, the survey interviewers were provided with a fully automatic, digital device with automatic upper-arm inflation and automatic pressure release. Interviewers were trained in the use of this device according to the manufacturer's recommended protocol. Three measurements of systolic and diastolic blood pressure (measured in millimeters of mercury [mmHg]) were taken during the survey interview, with an interval of at least 10 minutes between measurements. The average of the second and third measurements was used to classify individuals with respect to hypertension, following internationally recommended categories (WHO, 1999). Individuals were classified as hypertensive if their systolic blood pressure exceeded 140 mmHg or if their diastolic blood pressure exceeded 90 mmHg. Elevated blood pressure was classified as mild, moderate, or severe according to the cut-off points recommended by the World Health Organization and the National Institutes of Health (WHO, 1999; NIH, 1997)."
*Source: National Statistical Committee of the Kyrgyz Republic (NSC), Ministry of Health [Kyrgyz Republic], and ICF International. 2013. Kyrgyz Republic Demographic and Health Survey 2012. Bishkek, Kyrgyz Republic, and Calverton, Maryland, USA: NSC, MOH, and ICF International. Available at: https://dhsprogram.com/pubs/pdf/FR283/FR283.pdf*

**Lao PDR STEPS 2013**
No further information available

**Lesotho DHS 2014**
"During the individual interview, three blood pressure measurements were taken from consenting women age 15-49 and men age 15-59 using Omron M3W blood pressure monitors. Measurements were taken at intervals of 10 minutes or more. The average of the second and third measurements was used to classify the respondent with respect to hypertension, according to internationally recommended categories (WHO 1999; NIH 1997). The results, as well as information about the symptoms of high blood pressure and ways in which it can be prevented, were provided to the respondent via the Blood Pressure Findings Report Form and Brochure."
*Source: Ministry of Health, The DHS Program & ICF International. 2016. Lesotho Demographic and Health Survey 2014. Available at: https://www.dhsprogram.com/pubs/pdf/FR309/FR309.pdf.*

**Liberia STEPS 2011**
"This survey was designed based on the WHO STEPS Survey approach. The survey utilize the STEPS procedures for calculating sample size at the primary (district-PSU) and secondary (Chiefdom-SSU) levels, the Kish Method for household level sampling, a generic questionnaire that are adaptable by countries and Personal Digital Assistance (PDAs) that are approved by WHO for data collection and submission including blood pressure monitors (omron), blood pressure monitor cuffs and weighing scales."
Source: *Ministry of Health and Social Welfare: Liberia Chronic Disease Risk Factor Surveillance. Available at: http://www.who.int/chp/steps/Liberia_2011_STEPS_Report.pdf?ua=1.*

**Malawi STEPS 2009**
"Blood pressure measurements were taken using battery powered digital blood pressure machine (Omron® M4-I). Three readings were taken 3-5 minutes apart. During the analysis the average of the last two readings was as the final blood pressure leading."
*Source: Ministry of Health and WHO: Malawi National STEPS survey for chronic non-communicable diseases and their risk factors. Final report. Available at: https://extranet.who.int/ncdsmicrodata/index.php/catalog/435*

**Mexico Family Life Survey 2009-12**
"In each wave of the survey, a physical health assessment is conducted as part of the individual interview. In the baseline, height, weight, waist circumference, hip circumference and blood pressure were measured along with hemoglobin, which was measured in the home with the Hemocue photometer. These measures were repeated in the first and second follow-ups."
*Source: The Biomarker Network. Available at: https://gero.usc.edu/cbph/network/studies-with-biomarkers/the-mexican-family-life-survey-mxfls/*

**Mongolia STEPS 2019**
No further information available

**Mozambique STEPS 2005**
No further information available

**Myanmar STEPS 2014**
"Blood pressure measurements were taken using automatic digital blood pressure monitor (Boso Medicus) with universal cuff size. Before taking the measurements, participants were asked to sit quietly and rest for 15 minutes with legs uncrossed. The sphygmomanometer cuff was placed on the left arm while the participant rested their forearm on a table with the palm facing upward. Three readings were taken 5 minutes apart. During the analysis the average of the last two readings were taken."
*Source: Myanmar Diabetes Association, Department of Public Health and Department of Medical Research: Report on National Survey of diabetes mellitus and risk factors for non-communicable diseases in Myanmar (2014). Available at: https://www.who.int/ncds/surveillance/steps/myanmar/en/*

**Namibia DHS 2013**

"In the 2013 NDHS, blood pressure measurements (systolic and diastolic) were carried out among women and men age 35-64 to assess the prevalence of high blood pressure among adults. The measurements were not used for diagnostic purposes. Rather, respondents who had an abnormal measurement were informed of their blood pressure level and advised to visit a health facility for evaluation. Blood pressure was measured using the Life Source UA-767 Plus digital device with automatic upper-arm inflation and automatic pressure release. Interviewers were trained in the use of this device according to the manufacturer's recommended protocol. Three blood pressure measurements were taken, and the first measurement was discarded. The average of the last two measurements was reported as the blood pressure reading in millimetres of mercury (mmHg)."
*Source: The Namibia Ministry of Health and Social Services (MoHSS) & ICF International. 2014. The Namibia Demographic and Health Survey 2013. Available at: https://dhsprogram.com/pubs/pdf/fr298/fr298.pdf.*

**Niger STEPS 2007**
No further information available

**Peru DHS 2012**
No further information available

**Romania: Study for the Evaluation of Prevalence of Hypertension and Cardiovascular Risk (SEPHAR II)**
"Blood pressure (BP) measurements were taken using an automatic oscillometric blood pressure measuring device certified by AAMI (Association for the Advancement of Medical Instrumentation), European Society of Hypertension (ESH) and the British Hypertension Society (BHS), model A&D UA 95 Plus. The use of other devices was forbidden. Before performing BP measurements, arm circumference was measured (using a tailor's tape measure with a maximum deviation of 0.5 cm, at the widest level of the arm) in order to choose the right cuff type (if the arm circumference was more than 32 cm, a cuff for obese people was used, and if the arm circumference was equal to or less than 32 cm, a standard cuff was used). The GP took one BP measurement at both arms and after that took two additional measurements at the arm with the highest BP value on the first measurement, at time intervals of at least 1 min.

Blood pressure measuring conditions respected the recommendations of the European Society of Hypertension. Thirty minutes before BP measurements the patient was not allowed to smoke, drink coffee or do exercise. Blood pressure measurements and thus the questionnaire were not applied to subjects under the obvious influence of alcohol consumption. In this situation, the physician scheduled a new meeting with the subject. [...]

*Delivering patient results and procedure in case of significant abnormalities*
Each individual tested in the study received from their field GP written information regarding their blood pressure values. If diagnosed with arterial hypertension, the patient was informed that they would have to report to their primary care physician. If very high BP values were detected, carrying a high risk for acute complications (above 210/130 mm Hg), the patient was referred to an emergency care center."
*Dorobantu, M., Tautu, O. F., Darab2ont, R., Ghiorghe, S., Badila, E., Dana, M., Dobreanu, M., Baila, I., Rutkowski, M., & Zdrojewski, T. (2015). Objectives and methodology of Romanian*

*SEPHAR II Survey. Project for comparing the prevalence and control of cardiovascular risk factors in two East-European countries: Romania and Poland. Archives of medical science : AMS, 11(4), 715–723. https://doi.org/10.5114/aoms.2015.53290*

**Rwanda STEPS 2012**
"Survey staff conducted the physical measurements following the recommended STEPwise protocols. The OMRON M4 Digital Automatic Blood Pressure Monitor was used to measure resting blood pressure. Blood pressure was measured three times; the first reading followed by two more measurements taken with 2-3 minute intervals. The three readings of the blood pressure were recorded, and the average of the second and third readings was used in the analysis."
*Source: Republic of Rwanda. Ministry of Health: Rwanda Non-communicable Diseases Risk Factors Report November 2015. Available at: https://www.who.int/ncds/surveillance/steps/rwanda/en/.*

**Samoa STEPS 2013**
No further information available

**São Tomé and Principe STEPS 2008**
No further information available

**Solomon Islands STEPS 2015**
No further information available

**South Africa DHS 2016**
"Three blood pressure measurements were taken from consenting women and men age 15 and older using Omron 1300 digital blood pressure monitors. Measurements were taken at intervals of 3 minutes or more. For the purpose of returning the result to the respondent, the third measurement was used to classify the respondent with respect to hypertension, according to internationally recommended categories (WHO 1999). Respondents who were informed that they had high blood pressure were provided with a written referral to a health facility for further management."
*Source: National Department of Health & ICF International. South Africa Demographic and Health Survey 2016. Available at: https://www.dhsprogram.com/publications/publication-fr337-dhs-final-reports.cfm.*

**Tajikistan STEPS 2016**
No further information available

**Tanzania STEPS 2012**
"Blood pressure was taken using an electronic sphygmomanometer (OMRON®). Blood pressure was taken at rest, with patient seated and the arm elevated at the level of the heart. All physical measurements were repeated 3 times."
*Source: Ministry of Health and Social Welfare and National Institute for Medical Research: Tanzania STEPS survey report. Available at: http://www.who.int/chp/steps/UR_Tanzania_2012_STEPS_Report.pdf?ua=1*

**Timor-Leste STEPS 2014**
"Blood pressure was measured using digital sphygmomanometers on the left arm, while the participants were in sitting position with the hand resting on the armrest of the chair or some objects; after ensuring that they had rested for 10 minutes. A second and third reading was taken after two minutes' resting interval (Figure 2.4) and the mean of the last two measurements of blood pressure used for analysis."
*Source: Ministry of Health Timor-Leste & World Health Organization Regional Office for South-East Asia. National Survey for Non-Communicable Disease Risk Factors and Injuries: Using WHO STEPS approach in Timor-Leste – 2014. Available at: https://www.who.int/ncds/surveillance/steps/Timor-Leste_2014_STEPS_Report.pdf.*

**Togo STEPS 2010**
No further information available

**Tonga STEPS 2017**
No further information available

**Tuvalu STEPS 2015**
No further information available

**Uganda STEPS 2014**
"Blood pressure measurements were taken on the left arm with the participant in the sitting position using battery powered digital blood pressure machine (Boso Medicus Uno®). Three readings were taken 3-5 minutes apart. [...] During the analysis the average of the last two readings was as the final blood pressure reading and heart rate respectively."
*Source: Ministry of Health: Non-communicable disease risk factor baseline survey. Uganda 2014 report. Available at: https://extranet.who.int/ncdsmicrodata/index.php/catalog/633*

**Vanuatu STEPS 2011**
"Trained survey staff conducted the physical measurements following the recommended STEPwise protocols. The OMRON M4 Digital Automatic Blood Pressure Monitor was used to measure resting blood pressure. Blood pressure was measured three times; the first reading followed by two more measurements taken with 2-3 minute intervals in between. The three readings of the blood pressure were recorded, and the average of the second and third readings was used in the analysis."
*Source: Vanuatu Ministry of Health and WHO Western Pacifics Region: Vanuatu NCD Risk Factors STEPS Report. Available at: https://extranet.who.int/ncdsmicrodata/index.php/catalog/714*

**Zambia STEPS 2017**
"According to the STEPS protocol, blood pressure measurements were taken using an automated blood pressure machine. Three readings were taken 3-5 minutes apart with the last two readings averaged to estimate the respondent's blood pressure."

*Source: Ministry of Health, Republic of Zambia: Zambia STEPS for non communicable diseases risk factors. Zambia Report for 2017 Available at: https://extranet.who.int/ncdsmicrodata/index.php/catalog/620*

**Zanzibar STEPS 2011**

"Three readings of BP and heart rate after 15 min of rest was done using an electronic BP meter (Omron) with a middle sized cuff on left arm. The mean of the second and third reading was used for analysis. The participants which could not have their BP measured due to upper arm circumference size, was recorded as out of range. Measurement between 40/30 mmHg and 300/200 mmHg was recorded. Previous history of hypertension, as well as antihypertensive medication used and lifestyle advises given by health care worker, was recorded before BP measurement."

*Source: Ministry of Health Zanzibar: NCD Survey Report. Main findings from the National Non-communicable Disease Risk Factor Survey 2011. Available at: https://extranet.who.int/ncdsmicrodata/index.php/catalog/621*

#### *3.2. Diabetes*

In three countries, biomarkers were measured via blood samples sent to a laboratory; in 30 countries, biomarkers were measured with a point-of-care device. Nine countries collected glycated hemoglobin A1c (HbA1c) only, while six of these did not specify a HbA1c measurement device.

Table S3. Diabetes measurement by country

| Measurement device | Country | Post Hoc Adjustment[1] |
|---|---|---|
| *Point-of-care fasting capillary glucose* | | |
| HemoCue® Glucose 201 Analyzer (HemoCue, Brea, California, USA) | Bangladesh, Namibia, Tanzania | None |
| CardioCheck® PA (pts Diagnostics, Indianapolis, Indiana, USA) | Benin, Bhutan, Eswatini, Ethiopia, Jordan, Kenya, Kiribati, Lao PDR, Mongolia, Rwanda, Solomon Islands, Timor-Leste, Uganda, Zambia | None |
| Accutrend® Plus (Roche, Basel, Switzerland) | Liberia, Malawi, Mozambique, São Tomé and Principe, Togo, Vanuatu, Zanzibar | Multiplied by 1.11 |
| Accu-Chek Active® (Roche, Basel, Switzerland) | Niger, Samoa | None |
| Accu-Chek Performa® (Roche, Basel, Switzerland) | Tuvalu | None |
| multiCare-in© (Biochemical Systems International, Arezzo, Italy) | Georgia | None |
| FreeStyle Optium H glucometer | India | Multiplied by 1.11 |
| SD LipidoCare Analyzer | Myanmar | None |
| *Laboratory-based Assessment of Fasting Plasma Glucose* | | |
| Hitachi 7600 modular chemistry analyzer (Hitachi, Tokyo, Japan) | China | N/A |
| Cobas 6000 analyzer (Roche Diagnostics, Indianapolis, Indiana, USA) | Romania | N/A |
| Cobas C311 auto-analyzer (Roche Diagnostics, Indianapolis, Indiana, USA) | Iran | N/A |
| *Glycated hemoglobin A1c (HbA1c)* | | |
| Dried blood spots using the Hemocue system | Indonesia | N/A |
| Point-of-care In2ItTM device by Bio-Rad | Mexico | N/A |

| | | |
|---|---|---|
| Dried blood spots using blood chemistry analyzer (not specified) | South Africa | N/A |
| Not specified | Botswana, Cabo Verde, Comoros, Ecuador, Kyrgyz Republic, Tajikistan | |

[1]This refers to the conversion from capillary to plasma equivalents.

*<u>In the following, please note:</u> In order to ensure accuracy in reporting, blood glucose measurement methods are pasted verbatim from specified sources.*

**Bangladesh DHS 2017-2018**

"Blood pressure (BP) measures and blood specimens for glucose testing were collected from all women and men age 18 and above (regardless of marital status) in the subsample of one-fourth of the households. All women and men identified as eligible for blood pressure measurements and glucose testing were contacted and had the test explained to them. Blood pressure measures and blood specimens for glucose testing were taken for those who consented. Those consenting to glucose testing were asked to fast overnight for 8 hours and were visited early the next day (the second day of fieldwork) to ensure that they had not broken their fast before the test. Individuals failing to take the test for any reason were requested again to fast overnight and take the test on the following day (the third day of fieldwork). Collection of blood specimens for glucose testing required two visits to a household in most cases. One male health technician and one female technician from each team were engaged for biomarker testing. [...]
The HemoCue 201 RT analyzer was used for measurement of blood glucose. Capillary whole blood was obtained from the middle or ring finger after respondents had fasted overnight. The first two drops were wiped away, and the third drop was taken for measurement. Blood glucose was measured in millimoles per liter (mmol/L). World Health Organization cut-off points were used for fasting plasma blood glucose measurements. Individuals with fasting plasma glucose values of7.0 mmol/L or above are classified as having diabetes."
*Source: National Institute of Population Research and Training (NIPORT), and ICF. 2020. Bangladesh Demographic and Health Survey 2017-18. Dhaka, Bangladesh, and Rockville, Maryland, USA: NIPORT and ICF. Available at: https://dhsprogram.com/pubs/pdf/FR344/FR344.pdf*

**Benin STEPS 2015**

The original report is in French. We translated the information using Google translate.

"The morning of the second day was devoted first to the fasting blood glucose and cholesterol tests by the team's qualified health worker (Laboratory Technician or Nurse) who pricked participants in the finger with a sterile disposable needle, then took a small drop of blood to do the tests in front of the participant and give the results on the spot. The rest of this day and the third are devoted to revisits, then the cycle at the end of each day, the data collection supervisor checked in with the team leader to ensure the completeness of the data, the correct completion of the questionnaires, the completeness of missing data and the reliability of the biological tests."
*Source: Rapport final de l'enquête pour la surveillance des facteurs de risque des maladies non transmissibles par l'approche ''STEPSwise'' de l'OMSENQUETE ''STEPS 2015'' au Bénin. Available at: https://www.who.int/ncds/surveillance/steps/benin/en/.*

**Bhutan STEPS 2014**
"Immediately after the training, survey teams were allocated to the chiwog/enumeration areas where they would go to conduct the survey. Each team administered the STEP 1 (Questionnaire) and STEP 2 (Physical measurements) on the first visit to a household. The participants were then asked to fast overnight i.e. not consume any food or drinks (except water) after 10 p.m. the previous night until the blood sample was collected in the morning. [...] Participants were asked to go to the testing centre set up by the survey team (located in the vicinity) the next morning. Here the blood samples were taken and the urine samples delivered to the survey team."
*Source: National survey for noncommunicable disease risk factors and mental health using approach WHO Steps Approach in Bhutan – 2014. Available at: http://www.who.int/chp/steps/bhutan/en/.*
*Additional reference: World Health Organization Regional Office for South-East Asia. National survey for noncommunicable disease risk factors and mental health using WHO STEPS approach in Bhutan—2014. Geneva: World Health Organization; 2014.*

**Botswana STEPS 2014**
No further information available

**Cabo Verde STEPS 2007**
No further information available

**China Health and Nutrition Survey 2009**
"After an overnight fast, blood was collected by venipuncture (12 ml). Whole blood was immediately centrifuged and serum tested for glucose using a Hitachi 7600 analyzer using a glucose oxidase phenol 4-aminoantipyrine peroxidase kit (GOD-PAP; Randox, Crumlin, UK). HbA1c was measured at a central lab in each province with a high-performance liquid chromatography system (D10 HLC, Bio-Rad, Hercules, CA, USA; PDQ HPLC, Primus, Kansas City, MO, USA; Model HLC-723 G7, Tosoh Corporation, Tokyo, Japan). All the samples were calibrated to the D10 HLC from Bio-Rad."
*Source: Attard, S., Herring, A., Wang, H. et al. Implications of iron deficiency/anemia on the classification of diabetes using HbA1c. Nutr & Diabetes 5, e166 (2015). https://doi.org/10.1038/nutd.2015.16*

**Comoros STEPS 2011**
No further information available

**Ecuador STEPS 2018**
The original report is in Spanish. We translated the information using Google translate.

"The STEPS Survey required the following equipment and supplies: [...] vi) portable equipment for measuring glucose and total cholesterol in capillary blood with its respective detector or chip and strip;"
*Source: Ministerio de Salud Pública: Encuesta STEPS Ecuador 2018. Vigilancia de enfermedades no transmisibles y factores de riesgo. INFORME DE RESULTADOS Available at: https://extranet.who.int/ncdsmicrodata/index.php/catalog/774*

**Eswatini STEPS 2014**
"Fasting blood glucose and total cholesterol comprised the targeted biochemical measures used to measure health risk for NCDs. On the first day of the survey after completion of STEP1 and STEP2, participants were asked to fast overnight of that day. i.e. people were asked not to consume any food except for clear water after taking dinner on that night until the survey team came in the morning of the following day (day 2). People in the selected EA were seen in their various homesteads where a finger prick was done using a CarioChekPA test system and a drop of blood was tested for glucose and total cholesterol. Those that complied with advice (fasting overnight) were eligible for testing."
*Source: Swaziland Government. Ministry of Health: WHO STEPS. Noncommunicable Disease Risk Factor Surveillance Report Swaziland 2014. Available at: http://www.who.int/chp/steps/swaziland/en/.*

**Ethiopia STEPS 2015**
"Laboratory tests were performed for blood glucose, total cholesterol and HDL cholesterol using CardioCheck PA Analyser and for Triglycerides levels using Cobas Integra 400 Plus (Roche Diagnostics
GmbH, Mannheim, Germany) clinical chemistry analyser. Concentrations of glucose, total cholesterol and HDL cholesterol were measured in capillary blood the next day after STEPS 1 and 2 of the data collection. Capillary blood tests were performed for all survey respondents using a CardioChek PA Analyser, after fasting. Laboratory test results were assessed and categorized according to the definition."
*Source: Ethiopian Public Health Institute 2016: Ethiopia STEPS report on risk factors for non-communicable diseasesand prevalence of selected NCDs. Available at: https://extranet.who.int/ncdsmicrodata/index.php/catalog/794/download/5522*

**Georgia STEPS 2016**
"The blood glucose levels of the study participants were assessed using capillary blood and a rapid test. Individuals who had received breakfast were excluded from the test."
*Source: National Center for Disease Control and Public Health. Georgia STEPS Survey 2016 Fact Sheet. Available at: http://www.who.int/chp/steps/georgia/en/.*

**India DHS 2015-2016**
"Random blood glucose was measured using a finger-stick blood specimen for eligible women age 15-49 and (in the state module subsample of households only) eligible men age 15-54 using the FreeStyle Optium H glucometer with glucose test strips. A referral form to a health facility for additional medical evaluation was provided for any respondent with a random blood glucose level of >200 mg/dl."
*Source: International Institute for Population Sciences & ICF International. National Family and Health Survey (NFHS-4) 2015-16. Available at: https://dhsprogram.com/pubs/pdf/FR339/FR339.pdf.*

**Indonesia Family Life Survey 2014-2015**
"In IFLS4, a random sample of IFLS1 dynastic households were chosen to get dried blood spots. In IFLS5 we continued to collect dried blood spots on the same respondents sampled in IFLS4. In principle these blood spots can be analyzed for many different purposes. Our aim was to retest

for C-reactive protein, a measure of body inflammation, which has been shown to be correlated with adult chronic diseases, including heart disease and HbA1c, new to IFLS5, a longer run measure of diabetes. The blood spots were collected in conjunction with using the Hemocue system to measure blood hemoglobin. The first drops of blood were used with the hemoglobin and after drops were put onto the Whatman cards. Hand warmers were used prior to the prick to increase blood flow. The Whatman cards were allowed to dry for at least 4 hours in the household on a special drying rack and then put into a small ziplocked bag, together with a desicant. The ziplock bags with desicants helped to keep the blood samples dry. The samples were kept in a cooler with a plastic cooler bag until they could be mailed back to headquarters in Yogyakarta. Usually dbs cards were mailed twice per week. The mailings were in special envelopes that were insulated in order to keep the cards cool. The mail reached headquarters usually within two days, where they were put into a deep freezer (kept at -40C) for storage until they were assayed. Each ziplock bag had a label with the household and person identification numbers, age and sex. Each card had the person id."
*Source: Strauss, J, Witoelar F, and Sikoki, B. The Fifth Wave of the Indonesia Family Life Survey (IFLS5): Overview and Field Report. March 2016. WR-1143/1-NIA/NICHD. Available at: https://www.rand.org/content/dam/rand/pubs/working_papers/WR1100/WR1143z1/RAND_WR1143z1.pdf*

**Iran STEPS 2016**
"[...] as separation equipment was not available in rural areas, a type of sampling vacuum tube for the glucose biochemical test was selected so that it wouldn't lose any amount of blood sugar during transportation to the chosen lab in the town. The tube used for this purpose was the sodium fluoride tube. Three tubes, the sodium fluoride tube, the lithium Heparin tube and EDTA tube were used in this study. The sodium fluoride tube 6cc was used for the glucose biochemical test, the lithium Heparin tube 9cc for the Total-Cholesterol‹ HDL-C‹ ALT and Triglyceride tests, and the EDTA 6cc was used for the HbA1c test. As the sodium fluoride tubes were not available in the country, the importing company was asked to obtain and provide them.To start with step 3 of the survey, there had to be a procedure to transport the samples to universities which involved purchasing equipment such as vacuum tubes, connecting tubes, urine containers, needles, Parafilm, sampler tips and holders. For the tubes to be sent to the universities and returned safely to the Non-Communi-cable Diseases Research Center, a special box was designed that could hold all the tubes used in this study. The design of the box caused a suitable and accurate transportation of the tubes (sending and returning them) along with the possibility of sending the individual samples separately. Furthermore, to decrease probable errors, a process was designed in which each individual's sample was given a package with a pre-defined specific barcode (the cluster's code). All the contents of this package had a unique label containing the individual's code, the university's name, town's name, and the type of sample on them. Then, labels were prepared to give each indi-vidual a cluster and a box code. To prepare over 31050 labels, PVC rolls (to prevent the labels from coming off the tubes during transportation) and suitable resins (to prevent the writings on the tubes from being deleted) were used. After preparing labels and purchasing the tubes, the boxes for each individual were prepared and labeled and eventually the boxes prepared for each cluster were sent to the relating university. It must be noted that all procedures including labeling, putting caps on tubes and preparing the boxes were done in sterile conditions. Other equipment such as sampler tips, needles and holders were packaged and sent to

the relating university in the number required. Based on the conducted need analysis and the project management protocol, the required number was considered 2% higher than the defined number in case of probable problems.[...] In order to do biochemical tests on the collected samples, it was necessary to purchase special kits for the au-to-analyzer device and therefore lengthy negotiations were done with the dealer company which eventually caused the kits to be delivered to the Non-Communicable Diseases Research Center in two parts. For a suitable transfer of the samples from rural areas to the town and from the town to the Non-Communicable Diseases Research Center (the study's central headquarters), the transferring conditions underwent various tests (picture 8-2). For a suitable transportation, the two parameters, the allowable time for transfer and the transfer temperature, were studied. After studying past studies conducted in the central headquarters, reviewing articles and conducting numerous analyses, the optimum temperature came as to be 0 to 4 degrees Celsius and the maximum transfer time from sampling to delivery to the Non-Communicable Diseases Research Center (central headquarters) was decided to be 18 hours (from the moment sampling started until delivery to the central head-quarters). All samples were sent either by vaccine-transportation vehicles or airplane (depending on the distance and the samples' conditions) and were delivered to the central headquarters in less than 18 hours. Eventually, 20560 biological samples were received by the study's central headquarters and all biochemical tests were done on the samples having good quality and quantity.

*8.2 biochemical tests on blood and urine*
Following receiving the samples and forms, the cluster codes on forms were checked by Lims software and in case of any error, the cluster codes were corrected. After inspecting the samples, biochemical tests were done by Cobas C311 auto-analyzer and using Roche kits. In order to ensure the correctness of the results achieved through the auto-analyzer device, the control of each test was checked and recorded daily (Picture 8-3) and in case the device was out of the intended range, a sample calibrator was placed and the control sample was checked again. After ensuring the correct reading of the device, the sample reception routine started on that day. On average, the urine sodium calibration was done three times a day and the calibration for other tests were done once in two weeks. The glucose biochemical test was done on the sample plasma taken from the sodium fluoride vacuum tube, the Chol, Trig, HbA1c and ALT biochemical tests on the sample plasma taken from the lithium Heparin vacuum tube, the HbA1c on the full blood sample (EDTA) and eventually, the sodium and creatinine tests were done on the urine sample separately for each individual. At the end of each working day, the test results were transferred from the auto-analyzer to the server using LABIT software and for a higher assurance, a paper sample of all the test results were archived and backed up on a flash disk and an external hard disk at the end of that very day."
*Source: Non Communicable Diseases Research Center, Endocrinology and Metabolism Research Institute of Tehran University of Medical Sciences: Atlas of non-communicable diseases risk-factors surveillance in the islamic republic of Iran. STEPs 2016. Available at: https://www.who.int/ncds/surveillance/steps/STEPS_2016_Atlas_EN.pdf?ua=1*

**Jordan STEPS 2019**
"At the end of STEP 2 participants were instructed to fast overnight (not to consume any food or drinks except water until the morning of the following day), and to collect a urine sample in the

evening before fasting (this was for participants from Amman only). One day following the visit for STEP 1 and 2, the
team of medical doctors from the MOH visited the households who consented to conducting STEP 3 to take blood samples (all over Jordan) and collect urine samples (Amman only). In case the household did not respond to the first visit of STEP 3 another visit was made. If on the second visit no one responded, the household was recorded as missing for STEP 3 only. Similarly, if the participant forgot to fast overnight a second visit was scheduled for the next day. However, on the second attempt, if the person forgot to fast again, the team collected the blood sample, indicating that it was a non-fasting sample. Blood glucose and cholesterol were measured using a cardio check Analyzer PA."
*Source: Centre of Strategic Studies - University of Jordan, Jordan Ministry of Health, World Health Organization: Jordan National Stepwise Survey (STEPs) for Noncommunicable Diseases Risk Factors 2019. STEPs Technical Report. Available at: https://extranet.who.int/ncdsmicrodata/index.php/catalog/853/download/6010*

**Kenya STEPS 2015**
"STEP 3 included Biochemical Measurements on Blood Glucose and Blood Lipids. A point-of-care instrument (CardiocheckPA analyzer®) from PTS diagnostics was used for Blood Glucose and Blood Lipids."
*Source: Ministry of Health Kenya, Division of Noncommunicable Diseases, Kenya National Bureau of Statistics, & World Health Organization. WHO: Kenya STEPwise Survey for Non Communicable Diseases Risk Factors 2015 Report. Available at: http://www.who.int/chp/steps/Kenya_2015_STEPS_Report.pdf?ua=1.*

**Kiribati STEPS 2015**
"The survey included taking blood and urine samples. To measure fasting blood glucose and total cholesterol, participants fasted from 10:00pm the previous night until 7:00am the following morning. Capillary blood samples were drawn using the finger prick method; [...] All women of child-bearing age (18-49 years old) had capillary blood samples drawn using the finger prick method to measure haemoglobin using Hemocue 301."
*Source: Ministry of Health and Medical Services and WHO Western Pacific Region: Kiribati NCD risk factors STEPS report 2015-2016 - in collaboration with the WHO. Available at: https://extranet.who.int/ncdsmicrodata/index.php/catalog/724*

**Kyrgyz Republic STEPS 2013**
No further information available

**Lao PDR STEPS 2013**
"Cardiochek devices used for glucose and cholesterol measurements."
*Source: No report or fact sheet available. Information obtained from WHO NCD Microdata Repository. Available at: https://extranet.who.int/ncdsmicrodata/index.php/catalog/588/study-description#page=data_collection&tab=study-desc*

**Liberia STEPS 2011**
"The biochemical measurement in this survey includes the collection of blood sample from adults to measure a type of sugar in the body call glucose. There are several different types of

blood glucose tests available but this survey used the fasting blood sugar (FBS), which measures blood glucose after you have not eaten for at least 8 hours. It is often the first test done to check for pre-diabetes and diabetes."
*Source: Ministry of Health and Social Welfare: Liberia Chronic Disease Risk Factor Surveillance. Available at: http://www.who.int/chp/steps/Liberia_2011_STEPS_Report.pdf?ua=1.*

**Malawi STEPS 2009**
"On the first day of the survey after STEP 1 and part of STEP 2, participants were asked to starve overnight of that day. i.e. people were asked not to consume any food except for clear water after taking super/dinner of that day until the survey team came again in morning of the following day (day 2). People in the selected EA converged at the agreed place in the EA where finger blood samples for biochemistry tests were taken. Those that complied with advice (starving overnight) were eligible for finger prick blood sample collection. Total cholesterol and fasting blood glucose were measured using Accutrend®Plus machines."
*Source: Ministry of Health and WHO: Malawi National STEPS survey for chronic non-communicable diseases and their risk factors. Final report. Available at: https://extranet.who.int/ncdsmicrodata/index.php/catalog/435*

**Mexico Family Life Survey 2009-12**
"In each wave of the survey, a physical health assessment is conducted as part of the individual interview. [...] In the first follow-up, total cholesterol and (nonfasted) glucose were measured in the home. [...] In addition, HDL, total cholesterol and, for a sub-sample, HbA1c are measured in the home. HDL and total cholesterol are measured with the CardioChek Analyzer and HbA1c with the Bio-Rad In2It Analyzer. The CRP was measured using blood spots collected in the second follow-up."
*Source: The Biomarker Network. Available at: https://gero.usc.edu/cbph/network/studies-with-biomarkers/the-mexican-family-life-survey-mxfls/*

**Mongolia STEPS 2019**
"CardioChek PA was used for blood glucose and cholesterol measurements."
*Source: WHO NCD Microdata Repository. Available at: https://extranet.who.int/ncdsmicrodata/index.php/catalog/836/study-description#metadata-data_collection*

**Mozambique STEPS 2005**
"Accutrend was used for blood glucose and cholesterol measurements."
*Source: WHO NCD Microdata Repository. Available at: https://extranet.who.int/ncdsmicrodata/index.php/catalog/726/study-description#page=data_collection&tab=study-desc*

**Myanmar STEPS 2014**
"On the day of the survey when STEP 1 and STEP 2 have been finished, participants were asked to fast overnight i.e. people were be asked not to consume any food or drinks after 10 p.m. at night, except water, until the morning of the following day. Participants were asked to go to the designated testing centre the next morning where capillary blood was be taken by finger prick for rapid test. Those participants that complied with the fasting advice were eligible for blood

sample collection. Blood glucose, cholesterol, triglycerides, HDL and LDL were measured using SD LipidoCare Analyzer onsite, which requires a finger-prick blood draw to measure glucose and blood lipids. Then participants were be given glucose drink of monohydrous glucose 82.5 g (equivalent of anhydrous glucose 75 g) dissolved in 100-150 ml of drinking water and the subjects were requested to remain in the survey site for two hours. Postprandial plasma glucose was tested two hours after the glucose drink. SD LipidoCare Analyzer gives the blood glucose result as plasma equivalent value although it used capillary whole blood."
*Source: Myanmar Diabetes Association, Department of Public Health and Department of Medical Research: Report on National Survey of diabetes mellitus and risk factors for non-communicable diseases in Myanmar (2014). Available at: https://www.who.int/ncds/surveillance/steps/myanmar/en/*

**Namibia DHS 2013**
"In the 2013 NDHS, blood glucose testing was conducted to estimate the prevalence of diabetes mellitus type 2 among women and men age 35-64. After an overnight fast, a blood sample was obtained from respondents by a finger prick, and the blood was tested using the HemoCue Glucose 201 RT system (HemoCue Ab, Angelholm, Sweden) to determine the blood glucose level. Blood glucose levels were recorded as millimoles per litre (mmol/L) and compared with the World Health Organization's cutoffs to classify the prevalence of diabetes among adult women and men."
*Source: The Namibia Ministry of Health and Social Services (MoHSS) & ICF International. 2014. The Namibia Demographic and Health Survey 2013. Available at: https://dhsprogram.com/pubs/pdf/fr298/fr298.pdf.*

**Niger STEPS 2007**
"Accu-Chek Active devices were used for blood glucose measurements."
*Source: WHO NCD Microdata Repository. Available at: https://extranet.who.int/ncdsmicrodata/index.php/catalog/736/study-description#page=data_collection&tab=study-desc*

**Romania Study for the Evaluation of Prevalence of Hypertension and Cardiovascular Risk (SEPHAR II)**
"Blood samples were collected at the investigator GP's office or at the subject's home if he/she was not able to walk. Prior to taking a blood sample, the nurse made sure that the subject had not eaten for the past 8–14 h or that he/she had not drunk any sweet drinks or drinks of any caloric value in the past 8–14 h. The nurse used a preassembled blood draw kit containing a 22G needle, a vacutainer kit, 8 ml vacuum type tube with separator gel for biochemistry, a 2 ml vacuum type tube with EDTA anticoagulant for glycated hemoglobin and a 6 ml vacuum type tube with sodium fluoride glycolytic inhibitor for blood glucose.

The nurse performed the following actions: confirmed the subject's personal data, took samples of venous blood with the subject in a sitting position with a maximum duration of stasis of 30 s, transferred urine from the receptacle it was collected in by the subject to the vacuum transport tube, filled out the referral form to the laboratory (filling in the time and date of sample collection), marked all the test tubes with stickers containing the subject's individual study code, prepared the material for transportation and transported the material to the central laboratory.

*Central laboratory*
Laboratory testing was carried out after the materials were delivered to the central laboratory.
[...]
The test results were electronically delivered to the study organizers, who entered them in the database.

*Delivering patient results and procedure in case of significant abnormalities*
Laboratory results were communicated to individuals by mail, with a proviso that they would be completed with a delay of up to 6 weeks after actual collection of samples. A physician assessed results outside the normal range; if he considered that the respective result required diagnostic evaluation or emergency therapy (e.g. significant hypokalemia), the patient was informed of this fact by telephone, along with sending their results by mail. People with fasting glucose ≥ 126 mg/dl were additionally referred for a fasting plasma glucose test in the local laboratory; those with blood glucose of 100–125 mg/dl received additional referral for a 75 g glucose oral tolerance test"
*Dorobantu, M., Tautu, O. F., Darabont, R., Ghiorghe, S., Badila, E., Dana, M., Dobreanu, M., Baila, I., Rutkowski, M., & Zdrojewski, T. (2015). Objectives and methodology of Romanian SEPHAR II Survey. Project for comparing the prevalence and control of cardiovascular risk factors in two East-European countries: Romania and Poland. Archives of medical science : AMS, 11(4), 715–723. https://doi.org/10.5114/aoms.2015.53290*

**Rwanda STEPS 2012**
"Participants provided separate consent for participating in step 3. Participants fasted from 10:00pm in the evening until measurements were taken the following morning. Capillary blood samples were drawn using the finger prick method. Total cholesterol, HDL and fasting blood glucose were measured using CardioChekPA (Glucose, Cholesterol, HDL) as per STEPs protocol. For every tenth finger-prick test performed, a venous sample was taken for quality control at the National Reference Laboratory. These quality control samples were withdrawn from the database and thus fewer people (n=6651) completed this step compared to steps 2 and 3. Participants who were identified as being at high risk of developing, or with, advanced chronic conditions were referred for a follow-up clinical examination."
*Source: Republic of Rwanda. Ministry of Health: Rwanda Non-communicable Diseases Risk Factors Report November 2015. Available at: https://www.who.int/ncds/surveillance/steps/rwanda/en/.*

**Samoa STEPS 2013**
"Accuchek was used to obtain blood glucose and cholesterol measurements."
*Source: WHO NCD Microdata Repository. Available at: https://extranet.who.int/ncdsmicrodata/index.php/catalog/707/study-description#page=data_collection&tab=study-desc*

**São Tomé and Principe STEPS 2008**
"Accutrend devices were used for biochemical measurements."
*Source: WHO NCD Microdata Repository. Available at: https://extranet.who.int/ncdsmicrodata/index.php/catalog/735/study-description#page=data_collection&tab=study-desc*

**Solomon Islands STEPS 2015**

"Cardiochek was used for blood glucose and cholesterol measurements."

*Source: NCD Microdata Repository: Solomon Islands – STEPS 2015. Available at: https://extranet.who.int/ncdsmicrodata/index.php/catalog/710/study-description#page=data_collection&tab=study-desc*

**South Africa DHS 2016**

"Nurses collected finger-prick blood specimens for laboratory HbA1c and HIV testing of women and men age 15 and older who consented. The protocol for blood specimen collection and analysis was based on the anonymous linked protocol developed by The DHS Program. This protocol allows for merging of test results with the sociodemographic data collected in the individual questionnaires after removal of all information that could potentially identify an individual. Nurses explained the procedure, the confidentiality of the data, and the fact that the test results would not be made available to respondents. Blood for HbA1cand HIV testing was collected on a filter paper card. The card was preprinted with five circles, each of which could hold approximately 75 µlof blood and the first of which had been treated with a reagent required for HbA1ctesting. If a respondent consented to both HbA1c and HIV testing, five blood spots from the finger prick were collected on the filter paper card, to which a barcode label unique to the respondent was affixed. Duplicate barcodes were attached to the Biomarker Questionnaire, one to indicate that the respondent had consented to HbA1ctesting and another to indicate that the respondent had consented to HIV testing. A fourth copy of the same barcode was affixed to the dried blood spot (DBS) transmittal sheet to track the blood samples from the field to the laboratory. [...]

If the respondent consented only to HbA1c testing, a single blood drop was collected on the appropriate pretreated circle of the filter paper card to which the barcode label was affixed, and duplicate barcode labels were attached to the Biomarker Questionnaire and the DBS transmittal sheet. Blood samples were dried overnight and packaged for storage the following morning. Samples were periodically collected from the field and transported to the Global Clinical and Viral Laboratory (GCVL) in Durban. Upon arrival at GCVL, each blood sample was logged into the CSPro HIV Test Tracking System database, given a laboratory number, and stored at -20C until tested. The HbA1c and HIV testing protocols stipulated that blood could be tested only after questionnaire data collection had been completed, data had been verified and cleaned, and all unique identifiers other than the anonymous barcode number had been removed from the data file. To measure HbA1c, a common method was adapted for use with DBS specimens. Specifically, using a blood chemistry analyser, total haemoglobin concentration was measured by a colorimetric method monitoring the change in absorbance at 410 nm. HbA1c concentration was measured by a turbidimetric immunoinhibition method monitoring the change in absorbance at 340nm. HbA1c concentration is expressed as a percentage of total haemoglobin."

*Source: National Department of Health & ICF International. South Africa Demographic and Health Survey 2016. Available at: https://www.dhsprogram.com/publications/publication-fr337-dhs-final-reports.cfm.*

**Tajikistan STEPS 2016**

No further information available

**Tanzania STEPS 2012**

"In addition to the questionnaires and physical examination, participant were invited for fasting blood glucose and cholesterol measurements. Participants were instructed to fast for at least 8 hours before the test. All tests were performed by a trained technician. Blood was taken by finger prick, and measured using Hemoque® 201 analyzer."
*Source: Ministry of Health and Social Welfare and National Institute for Medical Research: Tanzania STEPS survey report. Available at: http://www.who.int/chp/steps/UR_Tanzania_2012_STEPS_Report.pdf?ua=1*

**Timor-Leste STEPS 2014**
"STEP 3 included biochemical measurements including fasting blood glucose and cholesterol were done by dry chemistry method using CardioChek devices (Figure 2.5). All the measurements were taken at the house of the participant."
*Source: Ministry of Health Timor-Leste & World Health Organization Regional Office for South-East Asia. National Survey for Non-Communicable Disease Risk Factors and Injuries: Using WHO STEPS approach in Timor-Leste – 2014. Available at: https://www.who.int/ncds/surveillance/steps/Timor-Leste_2014_STEPS_Report.pdf.*

**Togo STEPS 2010**
The original report is in French. We translated the details on measurement using Google translate.

"Data collection was done in selected households in accordance with STEPS recommendations. The data for Step 1 and 2 were collected on the first day and that of Step 3 the following day in the same household by the health worker (nurse, medical assistant, student) in medicine, Laborantin) of the binomial. For the Step 3 data collection, participants were instructed to be fasting. In addition, the international unit used for the Glycemia and the total cholesterol level in Step 3 was the Milligram / deciliter (mg / dl). For the STEP 3 measurements, the STEPS Togo site used the recommended Accutrend Plus device by WHO. The confidence interval for the blood sugar measurement of this device was between 20 and 600 mg / dl and that of the total cholesterol level was between 150 and 600 mg / dl."
*Source: Service de Maladies Non Transmissibles de la Division de l'épidémiologie: Rapport final de l'enquête STEPS Togo 2010. Available at: http://www.who.int/chp/steps/2010STEPS_Report_Togo_FR.pdf?ua=1.*

**Tuvalu STEPS 2015**
"Accutrend Plus meter and Accu-chek Performa were used for cholesterol and glucose measurements."
*Source: WHO NCD Microdata Repository. Available at: https://extranet.who.int/ncdsmicrodata/index.php/catalog/639/study-description#page=data_collection&tab=study-desc*

**Uganda STEPS 2014**
"Participants were informed that STEP 1 and STEP 2 would be done on day 1 and STEP 3 would be done on day 2. Participants were requested to fast overnight after STEP 1 and STEP 2 and not to indulge in exercise or smoking in the morning of the STEP 3. Participants in the selected EA converged at the agreed place in the EA where finger blood samples for biochemistry tests were taken. Most of the participants had a very short distance to walk, and if substantial distance was

envisaged, they came on a motorcycle (Boda-Boda) or were collected by the survey team vehicle. Only those who reported compliance with the procedures for STEP 3 (overnight 8-hr fast, no exercise in the morning of the study and smoking) were eligible for finger prick blood sample collection. Total cholesterol, High Density Lipoprotein Cholesterol (HDL-C) and fasting blood glucose were measured using CardioChek® PA meter."
*Source: Ministry of Health: Non-communicable disease risk factor baseline survey. Uganda 2014 report. Available at: https://extranet.who.int/ncdsmicrodata/index.php/catalog/633*

**Vanuatu STEPS 2011**
"The survey included assessments of fasting blood glucose and fasting total cholesterol. Participants fasted from 10:00pm the previous night. Testing commenced at 6.00am the following morning, when capillary blood samples were drawn using the method of finger prick.

Accutrend devices were used for blood glucose and cholesterol measurements."
*Source: Vanuatu Ministry of Health and WHO Western Pacifics Region: Vanuatu NCD Risk Factors STEPS Report. Available at: https://extranet.who.int/ncdsmicrodata/index.php/catalog/714 and WHO NCD Microdata Repository. Available at: https://extranet.who.int/ncdsmicrodata/index.php/catalog/714/study-description#page=data_collection&tab=study-desc*

**Zambia STEPS 2017**
"STEP 3 was done in the morning of the following day in most cases. However in some places modifications were made so that participants were prepared beforehand through local leaders and community health workers, who were contact before the research teams arrived. Eligible members from sampled families were asked to come to a central location on a named day and time. Participants were told not to eat until they were seen by the research team. Once on site, the research team explained the purpose of the study to sampled families. Prior to entering the names of those eligible, selection was done and urine was collected immediately after validating that it wasn't the first time to pass urine that morning. For those who had fasted, glucose measurements were taken the same day while those who had not fasted, glucose measurements were done the following morning. [...] Blood glucose levels were measured after at least 10 hours of fasting. Testing was performed using a portable rapid diagnostic device (Cardiochek™) machine which used test strips for both blood glucose and lipid profile (total Cholesterol and HDL Cholesterol). Blood sample was collected via a minimally invasive figure prick. "
*Source: Ministry of Health, Republic of Zambia: Zambia STEPS for non communicable diseases risk factors. Zambia Report for 2017 Available at: https://extranet.who.int/ncdsmicrodata/index.php/catalog/620*

**Zanzibar STEPS 2011**
"Biochemical tests for glucose, cholesterol and triglyceride were performed on a different day than the interview and anthropometric measurements due to fasting requirements. A point-of-care instrument (Accutrend Plus from Roche) was used in order to be able to provide results to the participants on the same day of their attendance, and to reduce suspicion and rumour about blood sampling. The instrument was calibrated daily before use.

**Blood sugar:**

The participants were during the first day also asked questions about their history of diabetes and treatment for diabetes. A reported diagnosis of diabetes was recorded if the fasting blood sugar (FBG) had been measured and a doctor or health worker had told the participant that they had diabetes. Participants with a reported diagnosis of diabetes were asked if they were currently receiving insulin, oral drugs or had received special dietary and lifestyle advices from a doctor or health care worker. Whole blood capillary glucose was measured for each available fasting participant to assess the diabetes status of the surveyed population, the total proportion of participants with raised fasting blood glucose was calculated and was categorised as belonging to either of the following three categories (based on IDF criterions):

- Diabetes: Previously diagnosed by a health care worker and currently on medication OR FBG ≥ 6.1 mmol/L without being on treatment
- Pre-diabetes: Impaired Fasting Glucose with FBG ≥ 5.6 and <6.1 mmol/L
- Normal blood glucose levels FBG below 5.6 mmol/L

During analysis those who had un-recordable high or low measurements were included with highest recordable value (33.3 mmol/L) respectively lowest (1.1 mmol/L) in order to include these in the analysis."

*Source: Ministry of Health Zanzibar: NCD Survey Report. Main findings from the National Non-communicable Disease Risk Factor Survey 2011. Available at: https://extranet.who.int/ncdsmicrodata/index.php/catalog/621*

#### *3.3. Height/Weight*

Table S4. Height/weight measurement by country[1]

| Measurement | Country |
|---|---|
| *Weight* | |
| Mechanical needle scales | Madagascar |
| Heavy precision scales | Mozambique |
| Digital scale, not otherwise specified | Guatemala, Turkey |
| Portable digital scale, not otherwise specified | Ecuador |
| Pre-calibrated weighing scale (bathroom scale), not otherwise specified | Eswatini |
| Calibrated digital floor scale, not otherwise specified | China |
| Camry model EB1003 scale | Indonesia |
| MOMERT weighing machine | Zanzibar |
| SECA electronic scale, not otherwise specified | Burundi, Cameroon, Guinea, Jordan, Kenya, Kiribati, Rwanda, Zimbabwe |
| SECA lightweight mother-infant scales with digital screens, not otherwise specified | Armenia, Ethiopia, Gambia, Myanmar |
| SECA 213 portable stadiometers | South Africa |
| SECA 491 scale | Georgia |
| SECA 872 digital scale | Yemen |
| SECA 874 scale | India, Kyrgyz Republic |
| SECA 878 lightweight electronic scale with digital screen | Bangladesh, Malawi, Nigeria, Tajikistan, Uganda |
| SECA 881 scale with digital screen | Albania |
| Siltec PS500L | Vanuatu |
| Tanita HD 95 electronic scale | Romania |
| Not specified | Benin, Bhutan, Bolivia, Botswana, Brazil, Burkina Faso, Cabo Verde, Chad Comoros, Congo, Democratic Republic of the Congo, |

| | |
|---|---|
| | Côte d'Ivoire, Dominican Republic, Egypt, Gabon, Ghana, Haiti, Honduras, Iran, Lao PDR, Lesotho, Liberia, Mexico, Mongolia, Namibia, Niger, Pakistan, Peru, Samoa, São Tomé and Principe, Senegal, Solomon Islands, Tanzania, Timor-Leste, Togo, Tonga, Tuvalu, Zambia |
| *Height* | |
| Marked wooden scale | Zanzibar |
| Measuring board | Gambia, Yemen, Turkey |
| Measuring boards provided by UNICEF | Armenia |
| Measuring rods | Burundi, Guinea |
| Graduated measuring boards | Cameroon |
| Portable measuring device | Romania |
| Portable mechanical wall height rod | Ecuador |
| Portable stadiometer | China |
| Stadiometers | Eswatini |
| Frankfort plane | Liberia |
| SECA stadiometer | Kiribati |
| SECA 213 stadiometer | India, Indonesia, South Africa |
| SECA 214 stadiometer | Albania |
| SECA Leicester Height Measure | Vanuatu |
| Shorr Productions measuring boards/ShorrBoard® | Bangladesh, Ethiopia, Jordan, Kenya, Kyrgyz Republic, Malawi, Myanmar, Nigeria, Rwanda, Tajikistan, Uganda, Zimbabwe |
| Shorr altimeter | Mozambique |
| InLab S50 | Mongolia |
| Not specified | Benin, Bhutan, Bolivia, Botswana, Brazil, Burkina Faso, Cabo Verde, Chad, Comoros, Congo, Congo Democratic Republic, Côte d'Ivoire, Dominican Republic, Egypt, Gabon, Georgia, Ghana, Guatemala, Haiti, Honduras, |

| | |
|---|---|
| | Iran, Lao PDR, Lesotho, Madagascar, Mexico, Namibia, Niger, Pakistan, Peru, Samoa, São Tomé and Principe, Senegal, Solomon Islands, Tanzania, Timor-Leste, Togo, Tonga, Tuvalu, Zambia |

[1]Height and weight measurements were available in 72 countries. For 38 countries, weight measurement devices were not specified; for 39 countries, height measurement devices were not specified.

*In the following, please note: In order to ensure accuracy in reporting, anthropometric measurement methods are pasted verbatim from specified sources.*

**Albania DHS 2017-2018**

"Weight and height measures were collected for all children listed in the Household Questionnaire born since January 2003 and all women age 15-49. In households selected for the Men's Questionnaire (half of all households), all men age 15-49 were also measured. Weight data were obtained using Seca 881 scales with digital screens, as recommended by UNICEF, and were capable of collecting mother-and-baby measurements as well as individual measurements. For measuring height (or length), two separate pieces of equipment were used. Seca 416 Infantometres were used to measure recumbent length of infants and children less than 24 months of age. For children 24 months of age or older, as well as adult women and men, Sec 214 Stadiometres were used. "

*Source: Institute of Statistics, Institute of Public Health [Albania] and ICF Macro. 2010. Albania Demographic and Health Survey 2008-09. Tirana, Albania: Institute of Statistics, Institute of Public Health and ICF Macro. Available at: https://dhsprogram.com/pubs/pdf/FR230/FR230.pdf*

**Armenia DHS 2015-2016**

"In all households, height and weight measurements were recorded for children age 0-59 months and women age 15-49 years. Weight measurements were obtained using lightweight, electronic Seca scales with a digital screen and the mother/child function. Height measurements were carried out with measuring boards provided by UNICEF."

*Source: National Statistical Service [Armenia], Ministry of Health [Armenia], and ICF. 2017. Armenia Demographic and Health Survey 2015-16. Rockville, Maryland, USA: National Statistical Service, Ministry of Health, and ICF. Available at: https://dhsprogram.com/pubs/pdf/FR325/FR325.pdf*

**Bangladesh DHS 2017-2018**

"Height and weight measures were collected for children under age 5 and ever-married women age 15-49 in all of the households sampled. Also, in the subsample of one-fourth of the households, these measures were obtained for never-married women age 18 and above, ever-married women age 50 and above,and men age 18 and above. Weight measurements were obtained using lightweight, electronic SECA 878 scales with a digital screen and the mother and child function. Height measurements were carried out with measuring boards made by Shorr Productions. [...] One male health technician and one female technician from each team were deployed to take both measurements."

*Source: National Institute of Population Research and Training (NIPORT), and ICF. 2020. Bangladesh Demographic and Health Survey 2017-18. Dhaka, Bangladesh, and Rockville, Maryland, USA: NIPORT and ICF. Available at: https://dhsprogram.com/pubs/pdf/FR344/FR344.pdf*

**Benin STEPS 2015**
No further information available

**Bhutan STEPS 2014**
No further information available

**Bolivia DHS 2008**
No further information available

**Botswana STEPS 2014**
No further information available

**Brazil Pesquisa Nacional de Saúde 2013**
The original report is in Portugese. We translated the information using Google translate.

"The procedures for anthropometric and blood pressure measurements and the training of the field team were developed in conjunction with the Laboratory of Nutritional Assessment of Populations - Lanpopda, Faculty of Public Health, University of São Paulo - USP. In addition, criteria were developed for the establishment of biologically improbable measures. For anthropometric measures, the same procedures as the Family Budget Survey - POF 2008-2009, carried out by IBGE in partnership with the Ministry of Health, were followed."
*Source: Pesquisa Nacional de Saúde. Plano de Amostragem. Published 2010. https://www.pns.icict.fiocruz.br/index.php?pag=planoamostragem.*

**Burkina Faso DHS 2010**
No further information available

**Burundi DHS 2016-2017**
The original report is in French. We translated the information using Google translate.

"Height and weight were taken for women aged 15-49 and children under 5 in all households selected for the male survey. Weight measurements were obtained using electronic scales (SECA) while height measurements were taken using measuring rods: children under the age of two were measured lying down while older were measured in the standing position"
*Source: Institut National de la Statistique et de la Démographie - INSD/Burkina Faso and ICF International. 2012. Enquête Démographique et de Santé et à Indicateurs Multiples du Burkina Faso 2010. Calverton, Maryland, USA: INSD and ICF International. Available at: https://dhsprogram.com/pubs/pdf/FR256/FR256.pdf*

**Cabo Verde STEPS 2007**
No further information available

**Cameroon DHS 2018**
The original report is in French. We translated the information using Google translate.

"In the sub-sample of households not selected for the male survey, the weight and height of women aged 15-49 and children under 5 were measured using electronic scales (SECA) and graduated measuring boards. These measurements, recorded in the Biomarkers questionnaire (used in the sub-sample of households not selected for the male survey) were used to assess their nutritional status."
*Source: Institut National de la Statistique/INS et ICF. 2020. Enquête Démographique et de Santé du Cameroun 2018. Yaoundé, Cameroun et Rockville, Maryland, USA : INS et ICF. Available at: https://dhsprogram.com/pubs/pdf/FR360/FR360.pdf*

**Chad DHS 2014-2015**
The original report is in French. We translated the information using Google translate.

"Finally, the household questionnaire was also used to record the results of anthropometric measurements (weight and height) and blood samples taken for the HIV test."
*Source: Institut National de la Statistique des Études Économiques et Démographiques - INSEED/Tchad, Ministère de la Santé Publique - MSP/Tchad et ICF International, 2014-2015. Enquête Démographique et de Santé et à Indicateurs Multiples (EDS-MICS 2014-2015). Rockville, Maryland, USA: INSEED, MSP et ICF International. Available at: https://dhsprogram.com/pubs/pdf/FR317/FR317.pdf*

**China Health and Nutrition Survey 2009**
"Height was measured to the nearest 0.1 cm without shoes using a portable stadiometer, and weight was measured to the nearest 0.1 kg with lightweight clothing on a calibrated digital floor scale."
*Source: Liang, Y., Liu, R., Du, S., & Qiu, C. (2014). Trends in incidence of hypertension in Chinese adults, 1991-2009: the China Health and Nutrition Survey. International journal of cardiology, 175(1), 96–101. https://doi.org/10.1016/j.ijcard.2014.04.258*

**Comoros DHS 2012**
No further information available

**Congo DHS 2011-2012**
The original report is in French. We translated the information using Google translate.

"The training for the main survey took place from August 8 to September 8, 2011. One hundred and five people with at least a high school diploma completed the training. This included not only theoretical lectures on interviewing and information recording techniques and exercises on how to complete the questionnaires, but also field practice in non-sample areas and practice exercises of the surveyors, anthropometric measurements and anemia test in integrated health centers in Brazzaville.[...] It should be noted that anthropometric data were entered daily in the field in the Nutrisurvey or ENA (Emergency Nutrition Assessment) software. This made it possible to detect and correct the flags (aberrant or inconsistent data) during the collection."
*Source: Centre Nationale de la Statistique et des Études Économiques - CNSEE/Congo and ICF International. 2013. Enquête Démographique et de Santé du Congo (EDSC-II) 2011-2012.*

*Calverton, Maryland, USA: CNSEE and ICF International. Available at: https://dhsprogram.com/pubs/pdf/FR267/FR267.pdf*

**Congo Democratic Republic DHS 2013-2014**
The original report is in French. We translated the information using Google translate.

"Finally, the household questionnaire was also used to record the results of anthropometric measurements (weight and height), hemoglobin and malaria tests, and blood samples taken for the HIV test and vaccination coverage tests."
*Source: Ministère du Plan et Suivi de la Mise en œuvre de la Révolution de la Modernité - MPSMRM/Congo, Ministère de la Santé Publique - MSP/Congo and ICF International. 2014. Enquête Démographique et de Santé en République Démocratique du Congo 2013-2014. Rockville, Maryland, USA: MPSMRM, MSP, and ICF International. Available at: https://dhsprogram.com/pubs/pdf/FR300/FR300.pdf*

**Côte d'Ivoire DHS 2011-12**
The original report is in French. We translated the information using Google translate.

"At the end of the training, 3 permanent supervisors, 18 agents responsible for sampling and taking anthropometric measurements, 18 female controllers, 18 male team leaders, 18 interviewers and 36 interviewers were selected. [...] The household questionnaire also made it possible to collect information on the characteristics of the household [...]. This questionnaire was also used to record the results of anthropometric measurements (weight and height), hemoglobin (Hb), malaria tests of eligible persons."
*Source: Institut National de la Statistique – Cote d'Ivoire & ICF International. Enquête Démographique et de Santé et à Indicateurs Multiples 2011-2012. Available at: https://dhsprogram.com/pubs/pdf/FR272/FR272.pdf.*

**Dominican Republic DHS 2013**
No further information available

**Ecuador STEPS 2018**
The original report is in Spanish. We translated the information using Google translate.

"The STEPS Survey required the following equipment and supplies: [...] ii) portable digital scale; iii) portable mechanical wall height rod, iv) tape measure [...]."
*Source: Ministerio de Salud Pública: Encuesta STEPS Ecuador 2018. Vigilancia de enfermedades no transmisibles y factores de riesgo. INFORME DE RESULTADOS Available at: https://extranet.who.int/ncdsmicrodata/index.php/catalog/774*

**Egypt DHS 2014**
No further information available

**Eswatini STEPS 2014**
"Height was measured using the stadiometers. Height was measured without the participant wearing foot or headgear.Before the reading was taken,the respondent was requested to have his

feet together, heels against the back board, knees straight, and to look straight ahead. Height was recorded in centimetres. Weight measurements were taken on a pre-calibrated weighing scale (bathroom scale).The scales were calibrated daily using a known weight (1kg packet of sugar). Participants were weighed dressed in light clothing and barefooted. Measurements were taken to the nearest 0.1 kg."
*Source: Swaziland Government. Ministry of Health: WHO STEPS. Noncommunicable Disease Risk Factor Surveillance Report Swaziland 2014. Available at: http://www.who.int/chp/steps/swaziland/en/.*

**Ethiopia DHS 2016**
"Height and weight measurements were carried out on women age 15-49, men age 15-59, and children under age 5 in all selected households. Weight measurements were obtained using lightweight SECA mother-infant scales with a digital screen designed and manufactured under the guidance of UNICEF. Height measurements were carried out using a Shorr measuring board."
*Source: Central Statistical Agency/CSA/Ethiopia and ICF. 2016. Ethiopia Demographic and Health Survey 2016. Addis Ababa, Ethiopia, and Rockville, Maryland, USA: CSA and ICF. Available at: https://dhsprogram.com/pubs/pdf/FR328/FR328.pdf*

**Gabon DHS 2012**
No further information available

**Gambia DHS 2013**
"Height and weight measurements were carried out on women age 15-49 and children age 0-59 months in half of the households selected for the male survey. Weight measurements were obtained using lightweight SECA mother-infant scales with digital screens, designed and manufactured under the guidance of UNICEF. Height measurements were carried out using a measuring board."
*Source: The Gambia Bureau of Statistics - GBOS and ICF International. 2014. The Gambia Demographic and Health Survey 2013. Banjul, The Gambia: GBOS and ICF International. Available at: https://dhsprogram.com/pubs/pdf/FR289/FR289.pdf*

**Georgia STEPS 2016**
The original report is in Georgian. We translated the information using Google translate.

"The second step was to measure height, weight, hip circumference (measurement of ergonomic dimension with Seca 201), determination of pulse and blood pressure readings. It is noteworthy that pregnant patients were measured by height, weight, hip circumference: Growth Management Scale (330 HRS BMI) is used for weight and height estimation, using standard Seca 491 calculators."
*Source: National Center for Disease Control and Public Health. Georgia STEPS survey 2016. Final report. Available at: https://extranet.who.int/ncdsmicrodata/index.php/catalog/223*

**Ghana DHS 2014**
"In the subsample of households selected for the male survey, height and weight measurements were recorded for children age 0-59 months, women age 15-49, and men age 15-59."

*Source: Ghana Statistical Service - GSS, Ghana Health Service - GHS, and ICF International. 2015. Ghana Demographic and Health Survey 2014. Rockville, Maryland, USA: GSS, GHS, and ICF International. Available at: https://dhsprogram.com/pubs/pdf/FR307/FR307.pdf.*

**Guatemala DHS 2014-2015**
The original report is in Spanish. We translated the information using Google translate.

"The ENSMI 2014-2015 includes anthropometric measurements of weight and length in girls and boys under 24 months; weight and height in girls and boys from 24 to 60 months; and weight and height in women 15 to 49 years of age. To guarantee the quality of the measurements, training and anthropometric standardization were carried out for the group of anthropometrists in the use of the equipment and in the correct technique to perform the measurements. Theoretical and practical activities were carried out in the classroom, and practical application activities were carried out in two health services. After the training, the anthropometric standardization process was carried out, with the Habicht method, through which the staff developed the ability to take measurements of weight, height and length with accuracy and precision. The anthropometric standardization process was carried out in the nurseries of the Society for the Protection of the Child. [...] Eac(*58*, *59*)h work group consisted of a supervisor or group supervisor, a field editor, an anthropometrist, five interviewers, and four interviewers. Each of the groups was in charge of three vehicles for mobilization in the sample sectors, digital scale, height / infantometer, GPS, HemoCue and cell phone."
*Source: Ministerio de Salud Pública y Asistencia Social – MSPAS/Guatemala, Instituto Nacional de Estadística – INE/Guatemala, Secretarío de Planificación y Programación del la Presidencia & ICF international. VI Encuesta Nacional de Salud Materno Infantil 2014-2015. Available at: https://dhsprogram.com/pubs/pdf/FR318/FR318.pdf.*

**Guinea DHS 2018**
The original report is in French. We translated the information using Google translate.

"Height and weight were taken for women aged 15-49 and children under 5 in all households selected for the male survey. Weight measurements were obtained using electronic scales (SECA) while height measurements were taken using measuring rods."
*Source: Institut National de la Statistique/INS et ICF. 2018. Enquête Démographique et de Santé en Guinée 2018. Conakry, Guinée, et Rockville, Maryland, USA : INS et ICF. Available at: https://dhsprogram.com/pubs/pdf/FR353/FR353.pdf*

**Haiti DHS 2017**
The original report is in French. We translated the information using Google translate.

"Height and weight were recorded for children under five in all households; in two out of three households, these parameters were also recorded among women aged 15-49. In the other subsample of one in three households, weight and height were also recorded for men and women aged 35-64 and children aged 5-14."
*Source: Enquête Mortalité, Morbidité et Utilisation des Services - EMMUS-VI 2016-2017 Pétion-Ville, Haïti, et Rockville, Maryland, USA : IHE et ICF. Available at: https://dhsprogram.com/pubs/pdf/FR326/FR326.pdf*

**Honduras DHS 2011-2012**
No further information available

**India DHS 2015-2016**
"Height and weight were measured for children age 0-59 months, women age 15-49, and (in the state module subsample of households only) men age 15-54. The Seca 874 digital scale was used to measure the weight of children and adults. The height of adults and children age 24-59 months was measured with the Seca 213 stadiometer. The Seca 417 infantometer was used to measure the recumbent length of children under two years or less than 85 cm."
*Source: International Institute for Population Sciences & ICF International. National Family and Health Survey (NFHS-4) 2015-16. Available at: https://dhsprogram.com/pubs/pdf/FR339/FR339.pdf.*

**Indonesia Family Life Survey 2014-2015**
"Heights were measured using a Seca plastic height board, model 213. This measured heights to the nearest millimeter. Weights were taken using a Camry model EB1003 scale. This measured to the nearest tenth of a kilogram."
*Source: Strauss, J, Witoelar F, and Sikoki, B. The Fifth Wave of the Indonesia Family Life Survey (IFLS5): Overview and Field Report. March 2016. WR-1143/1-NIA/NICHD. Available at: https://www.rand.org/content/dam/rand/pubs/working_papers/WR1100/WR1143z1/RAND_WR1143z1.pdf*

**Iran STEPS 2016**
"The physical evaluations included measurements of height, weight, waist size, hip size, blood pressure, pulse and step counting, which was conducted by female questioners for women and male questioners for men after the participants' physical readiness and by using similar tools available in step 2 packages previously provided for the questioning team. Compatibility with the listed items in the relating protocol was carefully ensured by the supervising teams from the national and central headquarters as well as supervisors from the universities headquarters, who attended the venues. It is remarkable to mention that each time before weighing the individuals, the accuracy of the scales used was controlled by the proof one-kilogram weight available. Also, the questioning teams were obliged to use tools included in the package for step 2 provided for them by the central headquarters. Furthermore, the accuracy of the pressure gauges was checked according to step 2's protocol. In case of any failure in the equipment, supple-mentary equipment was used and the faulty ones were sent to the central headquarters to be exchanged."
*Source: Non Communicable Diseases Research Center, Endocrinology and Metabolism Research Institute of Tehran University of Medical Sciences: Atlas of non-communicable diseases risk-factors surveillance in the islamic republic of Iran. STEPs 2016. Available at: https://www.who.int/ncds/surveillance/steps/STEPS_2016_Atlas_EN.pdf?ua=1*

**Jordan DHS 2017-2018**
"Height and weight measurements were recorded for children age 0-59 months and for women age 15-49. [...]

As mentioned in Chapter 1, anthropometric data were collected during the 2017-18 JPFHS for children under age 5 and women age 15-49 in all households; Shorr height boards and electronic Seca weight scales were used in gathering these data."
*Source: Department of Statistics/DOS and ICF. 2019. Jordan Population and Family and Health Survey 2017-18. Amman, Jordan, and Rockville, Maryland, USA: DOS and ICF. Available at: https://dhsprogram.com/pubs/pdf/FR346/FR346.pdf*

**Kenya DHS 2014**
"To derive these indices, the 2014 KDHS took height and weight measurements among women age 15-49. Weight measurements were made using an electronic scale (SECA scale). Standing height measurements were made using height/length (Shorr) boards. Respondents for whom there was no information on height and/or weight and those who were pregnant or had given birth in the two months preceding the survey were excluded from the analysis."
*Source: Kenya National Bureau of Statistics, Ministry of Health/Kenya, National AIDS Control Council/Kenya, Kenya Medical Research Institute, National Council for Population and Development/Kenya, and ICF International. 2015. Kenya Demographic and Health Survey 2014. Rockville, MD, USA: Kenya National Bureau of Statistics, Ministry of Health/Kenya, National AIDS Control Council/Kenya, Kenya Medical Research Institute, National Council for Population and Development/Kenya, and ICF International. Available at: https://dhsprogram.com/pubs/pdf/FR308/FR308.pdf*

**Kiribati STEPS 2015**
"Survey staff obtained physical measurements following the recommended STEPwise protocols. Height and weight were measured once using the Seca Stadiometer to the nearest whole centimetre and the Seca scales to the nearest 0.1 kg, respectively. Participants were measured without shoes and wearing only light clothing."
*Source: Ministry of Health and Medical Services and WHO Western Pacific Region: Kiribati NCD risk factors STEPS report 2015-2016 - in collaboration with the WHO. Available at: https://extranet.who.int/ncdsmicrodata/index.php/catalog/724*

**Kyrgyz Republic DHS 2012**
"ICF staff trained the 18 health technicians on anthropometric measurements (height and weight) and anemia testing (hemoglobin). Anthropometry was conducted using the SECA 874 digital scale (weight) and the Shorr board (height), and hemoglobin testing was conducted using the HemoCue photometer system (Hb 201+)."
*Source: National Statistical Committee of the Kyrgyz Republic (NSC), Ministry of Health [Kyrgyz Republic], and ICF International. 2013. Kyrgyz Republic Demographic and Health Survey 2012. Bishkek, Kyrgyz Republic, and Calverton, Maryland, USA: NSC, MOH, and ICF International. Available at: https://dhsprogram.com/pubs/pdf/FR283/FR283.pdf*

**Lao PDR STEPS 2013**
No further information available

**Lesotho DHS 2014**

"Height and weight measurements were recorded for children age 0-59 months, women age 15-49, and men age 15-59."
*Source: Ministry of Health, The DHS Program & ICF International. 2016. Lesotho Demographic and Health Survey 2014. Available at: https://www.dhsprogram.com/pubs/pdf/FR309/FR309.pdf.*

**Liberia STEPS 2011**
"The WHO STEPS instrument was used to collect data on selected socio-demographic characteristics and lifestyle behaviors including, physical activity, and physical measurements of weight, height, waist and hip circumference, as well as blood pressure. Weight and height were measured with participants standing without shoes and wearing light clothing. Participants stood upright with the head in Frankfort plane for height measurement. Height was recorded to the nearest 0.5 cm, and weight was recorded to the nearest 100g."
*Source: Ministry of Health and Social Welfare: Liberia Chronic Disease Risk Factor Surveillance. Available at: http://www.who.int/chp/steps/Liberia_2011_STEPS_Report.pdf?ua=1.*

**Madagascar DHS 2008-2009**
The original report is in French. We translated the information using Google translate.

"As during the DHS previously carried out in Madagascar, the EDSMD-IV assessed the nutritional status of women aged 15-49 and children under five using anthropometric measurements. For logistical reasons, the EDSMD-IV could not use electronic scales with digital display for weight gain, but mechanical needle scales. These scales did not make it possible to obtain sufficiently precise weights for the children and, after verification of the data collected, the data relating to the weight of the children were not validated and are therefore not presented in this report: only l The nutritional indicator based on children's height was analyzed. On the other hand, data on the weight of women could be used to assess their nutritional status (see Chapter 11)."
*Source: Institut National de la Statistique - INSTAT/Madagascar and ICF Macro. 2010. Enquête Démographique et de Santé de Madagascar 2008-2009. Antananarivo, Madagascar: INSTAT and ICF Macro. Available at: https://dhsprogram.com/pubs/pdf/FR236/FR236.pdf*

**Malawi DHS 2015-2016**
"Weight was measured with an electronic SECA 878 flat scale designed for mobile use. For the weighing of very young children, the mother or caretaker was weighed first. [...] Height was measured with a Shorr Board® measuring board."
*Source: National Statistical Office - NSO/Malawi and ICF. 2017. Malawi Demographic and Health Survey 2015-16. Zomba, Malawi: NSO and ICF. Available at: https://dhsprogram.com/pubs/pdf/FR319/FR319.pdf*

**Mexico Family Life Survey 2009-12**
"In each wave of the survey, a physical health assessment is conducted as part of the individual interview. In the baseline, height, weight, waist circumference, hip circumference and blood pressure were measured along with hemoglobin, which was measured in the home with the Hemocue photometer. These measures were repeated in the first and second follow-ups. [...] In the second follow-up, sitting height was assessed."

*Source: The Biomarker Network: The Mexican Family Life Survey. Available at: https://gero.usc.edu/cbph/network/studies-with-biomarkers/the-mexican-family-life-survey-mxfls/*

**Mongolia STEPS 2019**
"InLab S50 was used for height measurements using ultrasonic waves."
*Source: WHO NCD Microdata Repository. Available at: https://extranet.who.int/ncdsmicrodata/index.php/catalog/836/study-description#metadata-data_collection*

**Mozambique DHS 2011**
The original report is in Portugese. We translated the information using Google translate.

"Nutritional status was assessed in all women aged 15 to 49 years. All women in this age group were measured, using Shorr type altimeters, designed especially for fieldwork, and heavy precision scales."
*Source: Ministerio da Saude - MISAU/Moçambique, Instituto Nacional de Estatística - INE/Moçambique and ICF International. Moçambique Inquérito Demográfico e de Saúde 2011. Calverton, Maryland, USA: MISA/Moçambique, INE/Moçambique and ICF International. Available at: https://dhsprogram.com/pubs/pdf/FR266/FR266.pdf*

**Myanmar DHS 2015-2016**
"Height and weight measurements were recorded for children age 0-59 months and women age 15-49. [...] Measurements were taken using measuring boards specially made by Shorr Productions for use in survey settings and lightweight SECA scales with digital screens."
*Source: Ministry of Health and Sports (MoHS) and ICF. 2017. Myanmar Demographic and Health Survey 2015-16. Nay Pyi Taw, Myanmar, and Rockville, Maryland USA: Ministry of Health and Sports and ICF. Available at: https://dhsprogram.com/pubs/pdf/FR324/FR324.pdf*

**Namibia DHS 2013**
No further information available

**Niger DHS 2012**
No further information available

**Nigeria DHS 2018**
"Height and weight measurements were recorded for children age 0-59 months and women age 15-49. The 2018 NDHS included quality assurance procedures to improve anthropometry data quality. [...] Laboratory scientists and nurses were trained to measure the height and weight of children and adults. [...] older children and adults were measured standing up (height). Weight measurements were taken using SECA scales with a digital display (model number SECA 878U). Height and length were measured with a ShorrBoard® measuring board."
*Source: National Population Commission - NPC/Nigeria and ICF. 2019. Nigeria Demographic and Health Survey 2018. Abuja, Nigeria, and Rockville, Maryland, USA: NPC and ICF. Available at: https://dhsprogram.com/pubs/pdf/FR359/FR359.pdf*

**Pakistan DHS 2017-2018**

“In a subsample of the households selected for the man's survey, the 2017-18 PDHS recorded height and weight measurements for children age 0-59 months and women age 15-49. Two enumerators in each field team were assigned to jointly take measurements. In contrast to the data collection procedure for the household and individual interviews,data related to anthropometry was initially recorded on the Biomarker Questionnaireand subsequently entered into interviewers' tablet computers.”
*Source: National Institute of Population Stud(41, 58–63)ies - NIPS/Pakistan and ICF. 2019. Pakistan Demographic and Health Survey 2017-18. Islamabad, Pakistan, and Rockville, Maryland, USA: NIPS and ICF. Available at: https://dhsprogram.com/pubs/pdf/FR354/FR354.pdf*

**Peru DHS 2012**
No further information available

**Romania Study for the Evaluation of Prevalence of Hypertension and Cardiovascular Risk (SEPHAR II)**
“The anthropometric measurements were made using the following devices:

- weight – using an approved electronic scale (model Tanita HD 95), with a maximum deviation of 0.1 kg, with the subject wearing light clothes (without outer garments and without shoes),
- height – using a portable measuring device with a maximum deviation of 0.5 cm,
- waist circumference, hip circumference and arm circumference – using a tailor's measuring tool, with a maximum deviation of 0.5 cm.”

*Source: Dorobantu M, Tautu OF, Darabont R, et al. Objectives and methodology of Romanian SEPHAR II Survey. Project for comparing the prevalence and control of cardiovascular risk factors in two East-European countries: Romania and Poland. Arch Med Sci. 2015;11(4):715-723. doi:10.5114/aoms.2015.53290*

**Rwanda DHS 2014-2015**
“In the all of the households not selected for the male survey, height and weight measurements were recorded for children age 0-5, women age 15-49, and men age 15-59. Height and weight information was collected from eligible men in half of households selected for male survey. [...] Weight measurements were taken using a lightweight electronic SECA scale designed and manufactured under the guidance of the United Nations Children’s Fund (UNICEF). The scale allowed for the weighing of very young children through an automatic mother-child adjustment that eliminated the mother’s weight while she was standing on the scale with her baby. Height measurements were carried out using a Shorr measuring board also produced under the guidance of UNICEF.”
*Source: National Institute of Statistics of Rwanda, Ministry of Finance and Economic Planning/Rwanda, Ministry of Health/Rwanda, and ICF International. 2015. Rwanda Demographic and Health Survey 2014-15. Kigali, Rwanda: National Institute of Statistics of Rwanda, Ministry of Finance and Economic Planning/Rwanda, Ministry of Health/Rwanda, and ICF International. Available at: https://dhsprogram.com/pubs/pdf/FR316/FR316.pdf*

**Samoa STEPS 2013**

No further information available

**São Tomé and Principe STEPS 2008**
No further information available

**Senegal DHS 2010-2011**
No further information available

**Solomon Islands STEPS 2015**
No further information available

**South Africa DHS 2016**
"Seca 878 digital scales, Seca 417 infantometers (for children under age 2), and Seca 213 portable stadiometers (for children age 2 and older and for adults) were used for these measurements. In addition, waist circumference was measured for women and men using a Seca 201 measuring tape."
*Source: National Department of Health & ICF International. South Africa Demographic and Health Survey 2016. Available at: https://www.dhsprogram.com/publications/publication-fr337-dhs-final-reports.cfm.*

**Tajikistan DHS 2017**
"Anthropometry:In all households, height and weight measurements were recorded for children age 0-59 months and women age 15-49. Weight measurements were obtained using lightweight, electronic SECA 878 scales with a digital screen and the mother and child function. Height measurements were carried out with measuring boards made by Shorr Productions. Children younger than age 24 months were measured while lying down (recumbent) on the board, while standing height was measured for older children."
*Source: Statistical Agency under the President of the Republic of Tajikistan, Ministry of Health and Social Protection of Population of the Republic of Tajikistan, and ICF. 2018. Tajikistan Demographic and Health Survey 2017. Dushanbe, Republic of Tajikistan, and Rockville, Maryland, USA: Statistical Agency under the President of the Republic of Tajikistan (SA), Ministry of Health and Social Protection of Population of the Republic of Tajikistan (MOHSP), and ICF. Available at: https://dhsprogram.com/pubs/pdf/FR341/FR341.pdf*

**Tanzania DHS 2015-2016**
"Height and weight measurements were recorded for children under age 5 and women age 15-49."
*Source: Ministry of Health, Community Development, Gender, Elderly and Children - MoHCDGEC/Tanzania Mainland, Ministry of Health - MoH/Zanzibar, National Bureau of Statistics - NBS/Tanzania, Office of Chief Government Statistician - OCGS/Zanzibar, and ICF. 2016. Tanzania Demographic and Health Survey and Malaria Indicator Survey (TDHS-MIS) 2015-16. Dar es Salaam/Tanzania: MoHCDGEC, MoH, NBS, OCGS, and ICF. Available at: https://dhsprogram.com/pubs/pdf/FR321/FR321.pdf*

**Timor-Leste DHS 2016**
"The 2016 TLDHS conducted anthropometry measurement and anemia testing. Women age 15-49 years and children age 0-59 months were eligible for anthropometry measurement in all households. In one-third of the sampled households, men age 15-59 were also eligible for anthropometry measurement."
*Source: General Directorate of Statistics (GDS), Ministry of Health and ICF. 2018. Timor-Leste Demographic and Health Survey 2016. Dili, Timor-Leste and Rockville, Maryland, USA: GDF and ICF. Available at: https://dhsprogram.com/pubs/pdf/FR329/FR329.pdf*

**Togo DHS 2013-2014**
No further information available

**Tonga STEPS 2017**
No further information available

**Turkey DHS 2013**
"In order to assess women's nutritional status, women were weighed and their heights measured using the same equipment used to obtain children's measurements (i.e., an electronic scale and wooden height board)."
*Source: Hacettepe University Institute of Population Studies, 2014, 2013 Turkey Demographic and Health Survey. Hacettepe University Institute of Population Studies, T.R. Ministry of Development and TÜBITAK, Ankara, Turkey. Available at: https://dhsprogram.com/pubs/pdf/FR352/FR352.pdf*

**Tuvalu STEPS 2015**
No further information available

**Uganda DHS 2016**
"Anthropometry.Height and weight measurements were recorded for children age 0-59 months, women age 15-49, and men age 15-54. [...] Weight was measured with an electronic SECA 878 flat scale designed for mobile use. [...] Height was measured with a Shorr Board® measuring board."
*Source: Uganda Bureau of Statistics - UBOS and ICF. 2018. Uganda Demographic and Health Survey 2016. Kampala, Uganda and Rockville, Maryland, USA: UBOS and ICF. Available at: https://dhsprogram.com/pubs/pdf/FR333/FR333.pdf*

**Vanuatu STEPS 2011**
"Height and weight were measured once using the Seca Leicester Height Measure to the nearest 0.1 cm and the Siltec PS500L to the nearest 0.1 kg, respectively. Participants were measured without shoes and wearing only light clothing."
*Source: Vanuatu Ministry of Health and WHO Western Pacifics Region: Vanuatu NCD Risk Factors STEPS Report. Available at: https://extranet.who.int/ncdsmicrodata/index.php/catalog/714*

**Yemen DHS 2013**

“In all households, height and weight measurements were recorded for children age 0-59 months and women age 15-49 years. In addition, mid-upper-arm circumference was also measured among women age 15-49. [...] Weight measurements were obtained using a SECA 872 digital scale, designed for weighing children and adults. Height measurements were carried out using locally manufactured measuring boards.”
*Source: Ministry of Public Health and Population (MOPHP), Central Statistical Organization (CSO) [Yemen], Pan Arab Program for Family Health (PAPFAM), and ICF International. 2015. Yemen National Health and Demographic Survey 2013. Rockville, Maryland, USA: MOPHP, CSO, PAPFAM, and ICF International. Available at: https://dhsprogram.com/pubs/pdf/FR296/FR296.pdf*

**Zambia STEPS 2017**
No further information available

**Zanzibar STEPS 2011**
No further information available

**Zimbabwe DHS 2015**
“SECA digital electronic scales were used to measure weight; ShorrBoards® were used to measure height and length.”
*Source: Zimbabwe National Statistics Agency and ICF International. 2016. Zimbabwe Demographic and Health Survey 2015: Final Report. Rockville, Maryland, USA: Zimbabwe National Statistics Agency (ZIMSTAT) and ICF International.*

#### *3.4. Tobacco use*

73 countries in total provided data on self-reported tobacco use that were included in our analysis.

Table S5. Tobacco use questions by country

| Country | Questions on tobacco use |
|---|---|
| Albania, Burkina Faso, Chad, Comoros, Congo Democratic Republic, Dominican Republic, Gambia, Ghana, Guatemala, Honduras, Kenya, Lesotho, Mozambique, Myanmar, Niger, Rwanda, Togo | Q1: Do you currently smoke cigarettes? Do you currently smoke or use any type of tobacco? What other type of tobacco do you currently smoke or use? (pipes, cigars, snuff?) |
| Bolivia, Kyrgyz Republic, Madagascar, Namibia | Q2: Do you currently smoke cigarettes? |
| Benin, Bhutan[1], Cabo Verde, Ecuador, Eswatini, Georgia, Kiribati, Lao PDR, Liberia, Mongolia, Samoa, São Tomé and Principe, Solomon Islands, Tonga, Tuvalu, Zanzibar | Q3: Do you currently smoke any tobacco products such as cigarettes, cigars or pipes? |
| Vanuatu | Q4: Do you currently smoke any tobacco products? |
| Armenia, Burundi, Guinea, Haiti, Jordan[1], Malawi, Nigeria, Pakistan, Tajikistan, Tanzania, Timor-Leste, Uganda, Zambia, Zimbabwe | Q5: Do you currently smoke cigarettes or other tobacco products every day, on some days, or not at all? Do you currently smoke or use any other type of tobacco every day, some days, or not at all? |
| Angola, Botswana, Cameroon, Egypt, India[2], South Africa | Q6: Do you currently smoke cigarettes or other tobacco products every day, on some days, or not at all? |
| China | Q7: Do you currently smoke tobacco on a daily basis, less than daily, or not at all? On average, how many of the following products do you currently smoke each day? Also, let me know if you smoke the product, but not every day |
| Mexico, Panama, Philippines | Q8: I would now like to ask you some questions about smoking tobacco, including cigarettes, cigars, pipe. Please do not answer about smokeless tobacco and other ways of getting nicotine such as electronic cigarettes at this time. Do you currently smoke tobacco on a daily basis, less than daily, or not at all? |
| Indonesia | Q9: Have you ever chewed tobacco, smoked a pipe, smoked self-rolled cigarettes, or smoked cigarettes/cigars? Do you still have the habit or have |

| | |
|---|---|
| | you totally quit? |
| Bangladesh, Brazil, Costa Rica, Ethiopia, Iran, Romania, Senegal, Uruguay, Vietnam, Yemen | Unknown[3] |

[1]Questions included additional country-specific tobacco products for Bhutan (bidis) and Jordan (nargila).
[2]No report available for the survey year (2017). Question was obtained from the most recent questionnaire available (2009).
[3]No survey report or questionnaire available.

**Definition of smokers and non-smokers**

Respondents were classified as current smokers if they either i) currently use any type of tobacco product, such as cigarettes, cigars or pipes (Q1, Q2, Q3, Q4), ii) use tobacco products daily or sporadically (Q5, Q6, Q7, Q8) or iii) ever consumed a tobacco product and still maintain this habit (Q9). Respondents answering “no” to either Q1, Q2, Q3, or Q4 as well as participants stating no use of any tobacco products at all in Q5, Q6, Q7, or Q8 were labeled as non-smokers. Accordingly, participants who had either never used a tobacco product or quit usage (Q9) were considered non-smokers.

***3.5. Lipids***

In 27 countries, biomarkers were measured with a point-of-care device; in five countries, the biomarker measurement method could not be identified.

Table S6. Lipid measurement by country

| Measurement device | Country |
|---|---|
| *Point-of-care measurement* | |
| Accutrend Plus | Malawi, Mozambique, São Tomé and Principe, Tanzania, Togo, Tuvalu, Vanuatu, Zanzibar |
| Accu-Chek Active® (Roche, Basel, Switzerland) | Samoa |
| CardioCheck PA | Benin, Bhutan, Eswatini, Jordan, Kenya, Kiribati, Lao PDR, Rwanda, Solomon Islands, Timor-Leste, Tonga, Uganda, Zambia |
| Cobas Integra 400 Plus (Roche, Mannheim, Germany) | Ethiopia |
| multiCare-in© (Biochemical Systems International, Arezzo, Italy) | Georgia |
| SD LipidoCare Analyzer | Mongolia, Myanmar |
| Point-of-care In2ItTM device by Bio-Rad | Mexico |
| Unknown | Botswana, Cabo Verde, Ecuador, Kyrgyz Republic, Tajikistan |

*In the following, please note: In order to ensure accuracy in reporting, lipid measurement methods are pasted verbatim from specified sources.*

**Benin STEPS 2015**

"**Equipment:**

Electronic device for the determination of capillary glycemia and total cholesterol + Triglycerides brand CardioChek ® P.A with MEMO chips and suitable strips. […] The actual survey took place from October 19 to December 30, 2015. During data collection, the first day was devoted to administering the questionnaire and taking physical measurements. An appointment was then made for the next morning at the respondent's home or sometimes in the nearest care unit or another appropriate place for fasting blood sugar and cholesterol tests. […] The morning of the second day was first devoted to fasting blood sugar and cholesterol tests by the team's qualified health worker (Laboratory Technician or Nurse) who pricked the participants' fingers with a single use, sterile needle, and then took a small drop of blood to do the tests in front of the participant and give the results on the spot. The rest of this day and the third are devoted to revisits, then the cycle begins again."

*Source: Rapport final de l'enquête pour la surveillance des facteurs de risque des maladies non transmissibles par l'approche ''STEPSwise'' de l'OMSENQUETE ''STEPS 2015'' au Bénin. Available at: https://www.who.int/ncds/surveillance/steps/benin/en/.*

**Bhutan STEPS 2014**

"Immediately after the training, survey teams were allocated to the chiwog/enumeration areas where they would go to conduct the survey. Each team administered the STEP 1 (Questionnaire) and STEP 2 (Physical measurements) on the first visit to a household. The participants were then asked to fast overnight i.e. not consume any food or drinks (except water) after 10 p.m. the previous night until the blood sample was collected in the morning. […] Participants were asked to go to the testing centre set up by the survey team (located in the vicinity) the next morning. Here the blood samples were taken and the urine samples delivered to the survey team."

*Source: National survey for noncommunicable disease risk factors and mental health using approach WHO Steps Approach in Bhutan – 2014. Available at: http://www.who.int/chp/steps/bhutan/en/.*

*Additional reference: World Health Organization Regional Office for South-East Asia. National survey for noncommunicable disease risk factors and mental health using WHO STEPS approach in Bhutan—2014. Geneva: World Health Organization; 2014.*

**Botswana STEPS 2014**

No further information available

**Cabo Verde STEPS 2007**

No further information available

**Ecuador STEPS 2018**

The original report is in Spanish. We translated the information using Google translate.

"The STEPS Survey required the following equipment and supplies:[...] vi) portable equipment for measuring glucose and total cholesterol in capillary blood with its respective detector or chip and strip; vii) lancet and micropipette."

*Source: Ministerio de Salud Publica: Encuesta STEPS Ecuador 2018. Vigilancia de enfermedades no transmisibles y factores de riesgo. INFORME DE RESULTADOS Available at: https://extranet.who.int/ncdsmicrodata/index.php/catalog/774*

**Eswatini STEPS 2014**

"On the first day of the survey after completion of STEP 1 and STEP 2, participants were asked to fast overnight of that day. i.e. people were asked not to consume any food except for clear water after taking dinner on that night until the survey team came in the morning of the following day (day 2). People in the selected EA were seen in their various homesteads where a finger prick was done using a CardioChek PA test system and a drop of blood was tested for glucose and total cholesterol. Those that complied with advice (fasting overnight) were eligible for testing."

*Source: Swaziland Government. Ministry of Health: WHO STEPS. Noncommunicable Disease Risk Factor Surveillance Report Swaziland 2014. Available at: http://www.who.int/chp/steps/swaziland/en/.*

**Ethiopia STEPS 2015**

"Laboratory tests were performed for blood glucose, total cholesterol and HDL cholesterol using CardioCheck PA Analyser and for Triglycerides levels using Cobas Integra 400 Plus (Roche Diagnostics
GmbH, Mannheim, Germany) clinical chemistry analyser. Concentrations of glucose, total cholesterol and HDL cholesterol were measured in capillary blood the next day after STEPS 1 and 2 of the data collection. Capillary blood tests were performed for all survey respondents using a CardioChek PA Analyser, after fasting. Laboratory test results were assessed and categorized according to the definition."
*Source: Ethiopian Public Health Institute 2016: Ethiopia STEPS report on risk factors for non-communicable diseasesand prevalence of selected NCDs. Available at: https://extranet.who.int/ncdsmicrodata/index.php/catalog/794/download/5522*

**Georgia STEPS 2016**
The original report is in Georgian. We translated the information using Google translate.

"Also, with the help of rapid tests, the content of total cholesterol and high-density lipoproteins in the blood was determined."
*Source: National Center for Disease Control and Public Health. Georgia STEPS survey 2016. Final report. Available at: https://extranet.who.int/ncdsmicrodata/index.php/catalog/223*

**Jordan 2019**
"At the end of STEP 2 participants were instructed to fast overnight (not to consume any food or drinks except water until the morning of the following day), and to collect a urine sample in the evening before fasting (this was for participants from Amman only). One day following the visit for STEP 1 and 2, the
team of medical doctors from the MOH visited the households who consented to conducting STEP 3 to take blood samples (all over Jordan) and collect urine samples (Amman only). In case the household did not respond to the first visit of STEP 3 another visit was made. If on the second visit no one responded, the household was recorded as missing for STEP 3 only. Similarly, if the participant forgot to fast overnight a second visit was scheduled for the next day. However, on the second attempt, if the person forgot to fast again, the team collected the blood sample, indicating that it was a non-fasting sample. Blood glucose and cholesterol were measured using a cardio check Analyzer PA."
*Source: Centre of Strategic Studies - University of Jordan, Jordan Ministry of Health, World Health Organization: Jordan National Stepwise Survey (STEPs) for Noncommunicable Diseases Risk Factors 2019. STEPs Technical Report. Available at: https://extranet.who.int/ncdsmicrodata/index.php/catalog/853/download/6010*

**Kenya STEPS 2015**
"In this survey respondents had their blood glucose levels measured after at least 12 hours of fasting. Testing was performed using a Cardiocheck machine which used test strips for both blood glucose and lipid profile (total Cholesterol and HDL Cholesterol). Blood sample was collected via a minimally invasive figure prick."
*Source: Ministry of Health Kenya, Division of Noncommunicable Diseases, Kenya National Bureau of Statistics, & World Health Organization. WHO: Kenya STEPwise Survey for Non Communicable Diseases Risk Factors 2015 Report. Available at: http://www.who.int/chp/steps/Kenya_2015_STEPS_Report.pdf?ua=1.*

**Kiribati STEPS 2015/16**
"In general, the survey personnel obtained informed consent from survey participants, gave fasting instructions to those participating in STEP 3, and made appointment times for those who consented to participate in the survey. Survey personnel conducted STEP 1 (questionnaire) at home if the participant was willing; if not, it will follow STEPS 2 and 3, which was done at a central location in each village on the second (or third) morning. […] The survey included taking blood and urine samples. To measure fasting blood glucose and total cholesterol, participants fasted from 10:00pm the previous night until 7:00am the following morning. Capillary blood samples were drawn using the finger prick method; and the Cardiochek used to measure total cholesterol, HDL and glucose in samples."
*Source: Ministry of Health and Medical Services and WHO Western Pacific Region: Kiribati NCD risk factors STEPS report 2015-2016 - in collaboration with the WHO. Available at: https://extranet.who.int/ncdsmicrodata/index.php/catalog/724*

**Kyrgyz Republic STEPS 2013**
No further information available

**Lao PDR STEPS 2013**
"Cardiochek devices used for glucose and cholesterol measurements."
*Source: No report or fact sheet available. Information obtained from WHO NCD Microdata Repository. Available at: https://extranet.who.int/ncdsmicrodata/index.php/catalog/588/study-description#page=data_collection&tab=study-desc*

**Malawi STEPS 2009**
"On the first day of the survey after STEP 1 and part of STEP 2, participants were asked to starve overnight of that day. i.e. people were asked not to consume any food except for clear water after taking super/dinner of that day until the survey team came again in morning of the following day (day 2). People in the selected EA converged at the agreed place in the EA where finger blood samples for biochemistry tests were taken. Those that complied with advice (starving overnight) were eligible for finger prick blood sample collection. Total cholesterol and fasting blood glucose were measured using Accutrend®Plus machines."
*Source: Ministry of Health and WHO: Malawi National STEPS survey for chronic non-communicable diseases and their risk factors. Final report. Available at: https://extranet.who.int/ncdsmicrodata/index.php/catalog/435*

**Mexico Family Life Survey 2009-12**
"In each wave of the survey, a physical health assessment is conducted as part of the individual interview. [...] In the first follow-up, total cholesterol and (nonfasted) glucose were measured in the home. In the second follow-up, sitting height was assessed. In addition, HDL, total cholesterol and, for a sub-sample, HbA1c are measured in the home. HDL and total cholesterol are measured with the CardioChek Analyzer and HbA1c with the Bio-Rad In2It Analyzer. The CRP was measured using blood spots collected in the second follow-up."
*Source: The Biomarker Network. Available at: https://gero.usc.edu/cbph/network/studies-with-biomarkers/the-mexican-family-life-survey-mxfls/*

**Mongolia STEPS 2019**
"LipidoCare was used for cholesterol and triglyceride measurements."
*Source: WHO NCD Microdata Repository. Available at: https://extranet.who.int/ncdsmicrodata/index.php/catalog/836/study-description#metadata-data_collection*

**Mozambique STEPS 2005**
"Accutrend was used for blood glucose and cholesterol measurements."
*Source: WHO NCD Microdata Repository. Available at: https://extranet.who.int/ncdsmicrodata/index.php/catalog/726/study-description#page=data_collection&tab=study-desc*

**Myanmar STEPS 2014**
"Fasting blood glucose, 2 hour blood glucose, total blood cholesterol, triglycerides, HDL and LDL cholesterol were determined using SD LipidoCare Analyzer. There were 18 data collection teams. In each data collection team, there were 6 members i.e. 1 team leader (medical doctor) for overall management and glucose loading and testing samples for blood glucose and lipids, 2 team members for face-face interview, 2 team members for measuring height and weight and 1 helper for registering and arranging participants. A 5-day training workshop for the survey data collection teams was conducted at University of Medicine (2),Yangon on 11-15 August 2014. […]

The training workshop included sessions on the […] use of SD LipiddoCare Analyzer for blood glucose and lipids, emergency management and referral of critically high blood glucose level for medical doctors in the data collection teams and quality control of all field processes. Each team were provided with a field kit containing: […] devices and test strips for STEP 3 (plus lancets, swabs and sharp containers, gloves, pipettes) and glucose packs for oral glucose tolerance tests. […] On the day of the survey when STEP 1 and STEP 2 have been finished, participants were asked to fast overnight i.e. people were be asked not to consume any food or drinks after 10 p.m. at night, except water, until the morning of the following day.

Participants were asked to go to the designated testing centre the next morning where capillary blood was be taken by finger prick for rapid test. Those participants that complied with the fasting advice were eligible for blood sample collection. Blood glucose, cholesterol, triglycerides, HDL and LDL were measured using SD LipidoCare Analyzer onsite, which requires a finger-prick blood draw to measure glucose and blood lipids."
*Source: Myanmar Diabetes Association, Department of Public Health and Department of Medical Research: Report on National Survey of diabetes mellitus and risk factors for non-communicable diseases in Myanmar (2014). Available at: https://www.who.int/ncds/surveillance/steps/myanmar/en/*

**Rwanda STEPS 2012**
"Participants provided separate consent for participating in step 3. Participants fasted from 10:00 pm in the evening until measurements were taken the following morning. Capillary blood samples were drawn using the finger prick method. Total cholesterol, HDL and fasting blood glucose were measured using CardioChekPA (Glucose, Cholesterol, HDL) as per STEPs

protocol. For every tenth finger-prick test performed, a venous sample was taken for quality control at the National Reference Laboratory. These quality control samples were withdrawn from the database and thus fewer people (n=6651) completed this step compared to steps 2 and 3. Participants who were identified as being at high risk of developing, or with, advanced chronic conditions were referred for a follow-up clinical examination."
*Source: Republic of Rwanda. Ministry of Health: Rwanda Non-communicable Diseases Risk Factors Report November 2015. Available at: https://www.who.int/ncds/surveillance/steps/rwanda/en/.*

**Samoa STEPS 2013**
"Accuchek was used to obtain blood glucose and cholesterol measurements"
*Source: WHO NCD Microdata Repository. Available at: https://extranet.who.int/ncdsmicrodata/index.php/catalog/707/study-description#page=data_collection&tab=study-desc*

**São Tomé and Principe STEPS 2008**
"Accutrend devices were used for biochemical measurements."
*Source: WHO NCD Microdata Repository. Available at: https://extranet.who.int/ncdsmicrodata/index.php/catalog/735/study-description#page=data_collection&tab=study-desc*

**Solomon Islands STEPS 2015**
**"**Cardiochek was used for blood glucose and cholesterol measurements."
*Source: WHO NCD Microdata Repository. Available at: https://extranet.who.int/ncdsmicrodata/index.php/catalog/710/study-description#page=data_collection&tab=study-desc*

**Tajikistan STEPS 2016**
No further information available

**Tanzania STEPS 2012**
"In addition to the questionnaires and physical examination, participant were invited for fasting blood glucose and cholesterol measurements. Participants were instructed to fast for at least 8 hours before the test. All tests were performed by a trained technician. Blood was taken by finger prick, and measured using Hemoque® 201 analyzer. Lipid measurement for Total Cholesterol and Triglycerides were done using Accutrend® Cholesterol machine."
*Source: Ministry of Health and Social Welfare and National Institute for Medical Research: Tanzania STEPS survey report. Available at: http://www.who.int/chp/steps/UR_Tanzania_2012_STEPS_Report.pdf?ua=1*

**Timor-Leste STEPS 2014**
"STEP 3 included biochemical measurements including fasting blood glucose and cholesterol were done by dry chemistry method using CardioCheck devices (Figure 2.5). All the measurements were taken at the house of the participant. […] Day 1 – Survey of the suco for verification of the number of households, calculation of sampling interval, approaching households, taking consent from the selected individuals, interviewing for the STEP 1 and STEP

2, informing the respondents to fast for next day. Day 2 and 3 – Morning fasting samples to be taken from the respondents (by trained enumerators only) whose interviews were completed the previous day. The remaining respondents were interviewed and asked to be fasting for the next day to collect blood samples. The data collection effectively took 45 days "
*Source: Ministry of Health Timor-Leste & World Health Organization Regional Office for South-East Asia. National Survey for Non-Communicable Disease Risk Factors and Injuries: Using WHO STEPS approach in Timor-Leste – 2014. Available at: https://www.who.int/ncds/surveillance/steps/Timor-Leste_2014_STEPS_Report.pdf.*

**Togo STEPS 2010**
The original report is in French. We translated the information using Google translate.

"Data collection was done in selected households in accordance with STEPS recommendations. The data for Step 1 and 2 were collected on the first day and that of Step 3 the following day in the same household by the health worker (nurse, medical assistant, student) in medicine, Laborantin) of the binomial. For the Step 3 data collection, participants were instructed to be fasting. In addition, the international unit used for the Glycemia and the total cholesterol level in Step 3 was the Milligram / deciliter (mg / dl). For the STEP 3 measurements, the STEPS Togo site used the recommended Accutrend Plus device by WHO. The confidence interval for the blood sugar measurement of this device was between 20 and 600 mg / dl and that of the total cholesterol level was between 150 and 600 mg / dl."
*Source: Service de Maladies Non Transmissibles de la Division de l'épidémiologie: Rapport final de l'enquête STEPS Togo 2010. Available at: http://www.who.int/chp/steps/2010STEPS_Report_Togo_FR.pdf?ua=1.*

**Tonga STEPS 2017**
"Cardiochek PA devices were used for the blood measures in Step 3."
*Source: WHO NCD Microdata Repository. Available at: https://extranet.who.int/ncdsmicrodata/index.php/catalog/713/study-description#page=data_collection&tab=study-desc*

**Tuvalu: STEPS 2015**
"Accutrend Plus meter and Accu-chek Performa were used for cholesterol and glucose measurements."
*Source: WHO NCD Microdata Repository. Available at:https://extranet.who.int/ncdsmicrodata/index.php/catalog/639/study-description#page=data_collection&tab=study-desc*

**Uganda STEPS 2014**
"Participants were informed that STEP 1 and STEP 2 would be done on day 1 and STEP 3 would be
done on day 2. Participants were requested to fast overnight after STEP 1 and STEP 2 and not to indulge in exercise or smoking in the morning of the STEP 3. Participants in the selected EA converged at the agreed place in the EA where finger blood samples for biochemistry tests were taken. Most of the participants had a very short distance to walk, and if substantial distance was envisaged, they came on a motorcycle (Boda-Boda) or were collected by the survey team vehicle. Only those who reported compliance with the procedures for STEP 3 (overnight 8-hr

fast, no exercise in the morning of the study and smoking) were eligible for finger prick blood sample collection. Total cholesterol, High Density Lipoprotein Cholesterol (HDL-C) and fasting blood glucose were measured using CardioChek® PA meter."
*Source: Ministry of Health: Non-communicable disease risk factor baseline survey. Uganda 2014 report. Available at: https://extranet.who.int/ncdsmicrodata/index.php/catalog/633*

**Vanuatu STEPS 2011**
"The survey included assessments of fasting blood glucose and fasting total cholesterol. Participants fasted from 10:00pm the previous night. Testing commenced at 6.00am the following morning, when capillary blood samples were drawn using the method of finger prick.

Accutrend devices were used for blood glucose and cholesterol measurements."
*Source: Vanuatu Ministry of Health and WHO Western Pacifics Region: Vanuatu NCD Risk Factors STEPS Report. Available at: https://extranet.who.int/ncdsmicrodata/index.php/catalog/714 and WHO NCD Microdata Repository. Available at: https://extranet.who.int/ncdsmicrodata/index.php/catalog/714/study-description#page=data_collection&tab=study-desc*

**Zambia STEPS 2017**
"STEP 3 included blood chemistry rapid diagnostic tests to assess fasting blood glucose and total cholesterol. This was done by the use of Cardio-Check spot testing equipment. […] STEP 3 was done in the morning of the following day in most cases. However in some places modifications were made so that participants were prepared beforehand through local leaders and community health workers, who were contact before the research teams arrived. Eligible members from sampled families were asked to come to a central location on a named day and time. Participants were told not to eat until they were seen by the research team. Once on site, the research team explained the purpose of the study to sampled families. Prior to entering the names of those eligible, selection was done and urine was collected immediately after validating that it wasn't the first time to pass urine that morning. For those who had fasted, glucose measurements were taken the same day while those who had not fasted, glucose measurements were done the following morning."
*Source: Ministry of Health, Republic of Zambia: Zambia STEPS for non communicable diseases risk factors. Zambia Report for 2017 Available at: https://extranet.who.int/ncdsmicrodata/index.php/catalog/620*

**Zanzibar STEPS 2011**
"Equipment and Reagents Used : [...] AccuChek and AccuTrend Plus (Roche) machines with their corresponding test strips (for measurement of blood glucose, cholesterol and triglycerides). [...] Analyses for blood lipids were conducted on fasting participants only. The cut-off for raised total cholesterol was set at 5.0 mmol/L. The cut-off for raised triglycerides was set at 2.0 mmol/L."
*Source: Ministry of Health Zanzibar: NCD Survey Report. Main findings from the National Non-communicable Disease Risk Factor Survey 2011. Available at: https://extranet.who.int/ncdsmicrodata/index.php/catalog/621*

#### *3.6. Hypertension medication use*

Information on blood pressure medication was available for 47 countries in total.

Table S7. Consistency of phrasing across surveys (Blood pressure medication)

| Country | Questions on anti-hypertensive medication |
|---|---|
| Benin, Bhutan, Botswana, Ecuador, Eswatini, Georgia, Kenya, Kiribati, Myanmar, Solomon Islands, Tajikistan, Timor-Leste, Tonga, Tuvalu, Uganda, Zambia | In the past 2 weeks, have you taken any medicine for your high blood pressure prescribed by a doctor or other healthcare professional? |
| Cabo Verde[1], Comoros, Gambia, Lao PDR, Liberia, Malawi, Mozambique, Niger, Rwanda, Samoa, São Tomé and Principe, Tanzania, Togo, Vanuatu, Zanzibar | Are you currently receiving any of the following treatments/advice for high blood pressure prescribed by a doctor or other health worker? Drugs (medication) that you have taken in the past two weeks |
| Bangladesh, Namibia, Peru | Are you currently receiving any of the following treatment/advice by a doctor or other health worker to control your blood pressure? Prescribed medication |
| Brazil | In the last two weeks, you took medication because of high blood pressure (high blood pressure)? |
| Albania, Egypt, India, Kyrgyz Republic, Lesotho, South Africa | To lower your hypertension or high blood pressure, are you now taking prescribed medication?<br>What type of medication are you taking? |
| China | Are you currently taking anti-hypertension drugs? |
| Indonesia | In order to deal with [hypertension] are you currently taking prescribed medication on a weekly basis?<br>Are you now taking the following treatments to treat [hypertension] and its complications? |
| Mexico | Do you currently take medicines regularly/ follow a treatment for: Hypertension |
| Ghana, Iran, Romania | Unknown (not specified in report/questionnaire not available) |

[1]No country-specific questionnaire available. Question is derived from generic STEPS questionnaire.

Overall, hypertension medication questions are highly similar. Most surveys asked either for medication in the last two weeks or current usage. We thus assumed that these negligible differences in phrasing of the time frame did not affect our analysis.

#### *3.7. Diabetes medication use*

Information on diabetes medication was available for 44 countries in total.

Table S8. Consistency of phrasing across surveys (Diabetes medication)

| Country | Questions on diabetes medication |
|---|---|
| Bhutan, Botswana, Eswatini, Kenya, Kiribati, Kyrgyz Republic, Lesotho, Mongolia, Peru, Solomon Islands, South Africa, Tajikistan, Timor-Leste, Togo, Tuvalu, Uganda, Zambia | In the past two weeks, have you taken any drugs (medication) for diabetes prescribed by a doctor or other health worker? |
| Cabo Verde, Comoros, Ethiopia, Jordan, Lao PDR, Liberia, Malawi, Mozambique, Rwanda, Samoa, São Tomé and Principe, Tanzania, Vanuatu, Zanzibar | Are you currently receiving any of the following treatments for diabetes prescribed by a doctor or other health worker as well as any advice? 1. Insulin, 2. Oral medication, 3. [...] |
| Bangladesh, Namibia, Niger | Are you currently receiving any of the following treatment/advice by a doctor or other health worker for your high blood glucose or diabetes? Prescribed medication such as insulin? |
| Benin, Ecuador, Georgia, Myanmar | In the past 2 weeks, have you taken any medicine for your diabetes prescribed by a doctor or other healthcare professional? Are you currently taking insulin for your diabetes prescribed by a doctor or other healthcare professional? |
| China | Did you use any of these treatment methods? 1. Oral medicine 2. Insulin, [...] |
| Indonesia | In order to deal with [diabetes] are you currently taking prescribed medication on a weekly basis? Are you now taking the following treatments to treat [diabetes] and its complications? |
| Mexico | Do you currently take medicines regularly/follow a treatment for: [...] Diabetes? |
| India[1], Iran[2], Romania | Unknown |

[1]From the survey report as well as the question labeling of the data, it appears that these questions were taken directly from the DHS questionnaire, which asks: "Are you taking medication to control your high blood sugar or diabetes?" (DHS7-Module-NCD Questionnaire; 06 June 2016)

[2] From the survey report as well as the question labeling of the data, it appears that these questions were taken directly from the generic STEPS questionnaire, which asks: "In the past two weeks, have you taken any drugs (medication) for diabetes prescribed by a doctor or other health worker?" (Generic STEPS Questionnaire, Version 3.1)

As with hypertension, diabetes medication questions are highly similar. Most surveys asked either for medication in the last two weeks or current usage. Thus we assumed that these negligible differences in phrasing of the time frame did not affect our analysis. One exception is

China; more details on how we handled this inconsistency in phrasing are outlined in eMethods 5.

#### *3.8. Statin use*

Information on statin use was available for 19 countries. The questioning for statin use and CVD history was consistent among all STEPS surveys used in our analysis and based on the generic version of the WHO STEPS questionnaire (v3.3), available at: https://www.who.int/ncds/surveillance/steps/instrument/STEPS_Instrument_V3.2.pdf

Table S9. Phrasing across STEPS surveys for statin use and CVD history

| **Variable** | **Questions in STEPS survey** |
|---|---|
| Statin use | Are you currently taking statins (Lovastatin/Simvastatin/Atorvastain or any other statin) regularly to prevent or treat heart disease? |
| CVD history | Have you ever had a heart attack or chest pain from heart disease (angina) or a stroke (cerebrovascular accident or incident)? |

However, survey questions did not account for details regarding CVD history. As such, we could not differentiate between heart disease and strokes, whether strokes were ischemic or hemorrhagic or distinguish if ischemic heart disease derived from previous myocardial infarction and/or chronic angina. In addition, information on statin use and CVD history stems from self-reports. However, self-reports of CVD history and medication have been found to provide a high degree of accuracy and reliability in similar studies (*60*).

### Materials and Methods S4: Computation of education categories

We established three categories for educational attainment, namely ‘no formal schooling‘, ‘primary education’ (≤ grade 6), ‘secondary education (grade 7 to 12) or further‘. The standard DHS asks for five different education levels, including ‘No education’, ‘Incomplete primary’, ‘Complete primary’, ‘Incomplete secondary’, ‘Complete secondary’ and ‘Higher’. The standard STEPS questionnaire covers the following education levels: ‘No formal schooling’, ‘Less than primary school’, ‘Primary school completed’, ‘Secondary school completed’, ‘High school completed’, ‘College/University completed’ and ‘Postgraduate degree’. In the standard GATS questionnaire respondents could choose their education levels from the eight categories ‘No formal schooling’, ‘Less than primary school completed’, ‘Primary school completed’, ‘Less than secondary school completed’, ‘Secondary school completed’, ‘High school completed’, ‘College/University completed’ and ‘Postgraduate degree completed’. Based the classifications outlined, we developed our classification from the standard questionnaires as follows:

Table S10. Summary of categorization of educational attainment from survey-specific categories to three-level study variable (DHS, STEPS and GATS)

| Survey | No formal schooling | Primary education’ (≤ grade 6) | | Secondary education (grade 7 to 12) or further | | | |
|---|---|---|---|---|---|---|---|
| **DHS** | “No education“ | “Incomplete primary“ | “Complete primary“ | “Incomplete secondary“ | “Complete secondary“ | “Higher“ | |
| **STEPS** | “No formal schooling“ | “Less than primary school“ | “Primary school completed“ | “Secondary school completed“ | “High school completed“ | “College/University completed“ | “Postgraduate degree“ |
| **GATS** | “No formal schooling“ | “Less than primary school completed“ | “Primary school completed“ | “Less than secondary school completed“ | “Secondary school completed“ | “College/University completed“ | “Postgraduate degree completed“ |

Six surveys did not employ either the DHS, STEPS or GATS questionnaires. 104 out of 105 countries collected education levels of survey participants. We used completed years of education in Iran, since this survey did not provide any education categories. Details on the computation of our education categories for all countries are provided in Table S11 below.

Table S11. Categorization of educational attainment from survey-specific categories to three-level study variable, by country

| Educational attainment categories | | | | | | |
|---|---|---|---|---|---|---|
| **Country** | **No formal schooling** | **Primary education' (≤ grade 6)** | | **Secondary education (grade 7 to 12) or further** | | |
| **Albania 2017-2018** | "No education" | "Incomplete primary" | "Complete primary" | "Incomplete secondary" | "Complete secondary" | "Higher" |
| **Albania 2008-2009** | "No education" | "Incomplete primary" | "Complete primary" | "Incomplete secondary" | "Complete secondary" | "Higher" |
| **Angola** | "No education" | "Incomplete primary" | "Complete primary" | "Incomplete secondary" | "Complete secondary" | "Higher" |
| **Armenia** | "No education" | "Incomplete primary" | "Complete primary" | "Incomplete secondary" | "Complete secondary" | "Higher" |
| **Bangladesh 2017-2018** | "No education" | "Incomplete primary" | "Complete primary" | "Incomplete secondary" | "Complete secondary" | "Higher" |
| **Bangladesh 2017** | "No formal schooling" | "Less than primary school" | "Primary school completed" | "Less than secondary school" | "Secondary school complete" | "Above" |
| **Benin** | "No formal schooling" | "Less than primary school" | "Primary school completed" | "End of secondary, Level 1" | "End of secondary, Level 2" | "College/University completed", "Postgraduate degree" |
| **Bhutan** | "No formal schooling" | "Less than primary school" | "Primary school completed" | "Class 10 completed" | "Class 12 completed" | "College/University completed", "Postgraduate degree" |
| **Bolivia** | "No education" | "Incomplete primary" | "Complete primary" | "Incomplete secondary" | "Complete secondary" | "Higher" |
| **Botswana 2017** | "No formal schooling" | "Less than primary school" | "Primary school completed" | "Less than (Junior) secondary school completed" | "Secondary (Junior) school completed", "Less than (Senior) secondary school completed", "High (Senior) school completed" | "College/University completed", "Postgraduate degree completed" |

| | | | | | | |
|---|---|---|---|---|---|---|
| **Botswana 2014** | "No formal schooling" | "Less than primary school" | "Primary school completed" | "Jnr. Secondary school completed", "Snr. Secondary school completed" | "High school completed" | "Tertiary school completed", "College/University completed", "Postgraduate degree" |
| **Brazil** | – | "Preparation course for EJA (Youth and Adult Education)", "Primary School Education (4 Years)", "Primary School Education (Antigo ginaso)" | "Primary School Degree for Older People", "8 Years of Primary School" | "Primary and Secondary Education" | "3 Years of Secondary School", "Secondary School Degree for Older People" | "Bachelor's Degree", "Master's Degree", "PhD" |
| **Burkina Faso** | "No education" | "Incomplete primary" | "Complete primary" | "Incomplete secondary" | "Complete secondary" | "Higher" |
| **Burundi** | "No education" | "Incomplete primary" | "Complete primary" | "Incomplete secondary" | "Complete secondary" | "Higher" |
| **Cabo Verde** | "No formal schooling" | "Less than primary school" | "Primary school completed" | "Class 10 completed" | "Class 12 completed" | "College/University completed", "Postgraduate degree" |
| **Cameroon** | "No education" | "Incomplete primary" | "Complete primary" | "Incomplete secondary" | "Complete secondary" | "Higher" |
| **Chad** | "No education" | "Incomplete primary" | "Complete primary" | "Incomplete secondary" | "Complete secondary" | "Higher" |
| **China 2010** | "No formal schooling" | "Less than primary school completed" | "Primary school completed" | "Less than secondary school completed" | "Secondary school completed", "High school completed/Technical secondary school" | "College/University completed", "Postgraduate degree completed" |
| **China 2009** | "No school completed" | "1-4 years of primary school" | "Graduated from primary school" | "Lower middle school degree" | "Upper middle school degree" | "Technical or vocational degree", "University or college degree", "Master's degree or higher" |

| | | | | | | |
|---|---|---|---|---|---|---|
| **Comoros 2012** | “No education” | “Incomplete primary” | ”Complete primary” | ”Incomplete secondary” | ”Complete secondary” | ”Higher” |
| **Comoros 2011** | “No formal schooling” | “Less than primary school” | “Primary school completed” | “Secondary school completed” | “High school completed” | “College/University completed”, “Postgraduate degree” |
| **Congo** | “No education” | “Incomplete primary” | ”Complete primary” | ”Incomplete secondary” | ”Complete secondary” | ”Higher” |
| **Congo Democratic Republic** | “No education” | “Incomplete primary” | ”Complete primary” | ”Incomplete secondary” | ”Complete secondary” | ”Higher” |
| **Costa Rica** | “No formal schooling” | “Less than primary school completed” | “Primary school completed” | “Less than secondary school completed” | “Secondary school completed” | “College/University completed”, “Postgraduate degree completed” |
| **Côte d'Ivoire** | “No education” | “Incomplete primary” | ”Complete primary” | ”Incomplete secondary” | ”Complete secondary” | ”Higher” |
| **Dominican Republic** | “No education” | “Incomplete primary” | ”Complete primary” | ”Incomplete secondary” | ”Complete secondary” | ”Higher” |
| **Ecuador** | “No formal schooling” | “Literacy center”, “Basic education” | “Primary school” | “Secondary school”, “Middle education/Baccalaureate” | “Non-University Superior” | “University Superior”, “Postgraduate degree” |
| **Egypt 2015** | “No education” | “Incomplete primary” | ”Complete primary” | ”Incomplete secondary” | ”Complete secondary” | ”Higher” |
| **Egypt 2014** | “No education” | “Incomplete primary” | ”Complete primary” | ”Incomplete secondary” | ”Complete secondary” | ”Higher” |
| **Eswatini** | “No formal schooling” | “Less than primary school” | “Primary school completed” | “Secondary school completed” | “High school completed” | “College/University completed”, “Postgraduate degree” |
| **Ethiopia 2016 (GATS)** | “No formal schooling” | “Less than primary school completed” | “Primary school completed” | “Less than secondary school completed” | “Secondary school completed” | “College/University completed”, “Postgraduate degree completed” |
| **Ethiopia 2016 (DHS)** | “No education” | “Incomplete primary” | ”Complete primary” | ”Incomplete secondary” | ”Complete secondary” | ”Higher” |

| | | | | | | |
|---|---|---|---|---|---|---|
| **Ethiopia 2015 (STEPS)** | “No formal schooling” | “Less than primary school completed” | “Primary school completed” | “Less than secondary school completed” | “Secondary school completed” | “College/University completed”, “Postgraduate degree completed” |
| **Gabon** | “No education” | “Incomplete primary” | ”Complete primary” | ”Incomplete secondary” | ”Complete secondary” | ”Higher” |
| **Gambia 2013** | “No education” | “Incomplete primary” | ”Complete primary” | ”Incomplete secondary” | ”Complete secondary” | ”Higher” |
| **Gambia 2010** | “No formal schooling” | “Pre-primary/nursery completed” | “Primary/lower basic completed” | “Middle/upper basic completed” | “Secondary technical completed”, “Senior secondary completed”, “High school completed”, “Vocational completed” | “College completed”, “University completed” |
| **Georgia** | “No formal schooling” | “Less than primary school” | “Primary school completed” | “Secondary school completed” | “High school completed” | “College/University completed”, “Postgraduate degree” |
| **Ghana 2014** | “No education” | “Incomplete primary” | ”Complete primary” | ”Incomplete secondary” | ”Complete secondary” | ”Higher” |
| **Ghana 2007-2008** | “No formal schooling” | “Less than primary school” | “Primary school completed” | “Secondary school completed” | “High school (or equivalent) completed” | “College/University completed”, “Postgraduate degree” |
| **Guatemala** | “No education” | “Incomplete primary” | ”Complete primary” | ”Incomplete secondary” | ”Complete secondary” | ”Higher” |
| **Guinea** | “No education” | “Incomplete primary” | ”Complete primary” | ”Incomplete secondary” | ”Complete secondary” | ”Higher” |
| **Haiti** | “No education” | “Incomplete primary” | ”Complete primary” | ”Incomplete secondary” | ”Complete secondary” | ”Higher” |
| **Honduras** | “No education” | “Incomplete primary” | ”Complete primary” | ”Incomplete secondary” | ”Complete secondary” | ”Higher” |

| | | | | | | |
|---|---|---|---|---|---|---|
| **India 2017** | "No formal schooling" | "Less than primary" | "Primary but less than secondary" | "Secondary and above" | | |
| **India 2015-2016** | "Illiterate" "Literate without Formal education" | "Below Primary" | "Primary" | "Middle", "Secondary/Matric Class-X" | "Hr. Secondary", "Sr.Secondary", "Pre-University", "Class XII" | "Graduate/B.Tech/ BBA/MVVS/Equivalent", "Post Graduate/ M. Tech/ MBA/ MD/ Equivalent or higher", "Non-Technical/Technical diploma or certificate not equivalent to degree" |
| **Indonesia**[1] | "No formal schooling", "Kindergarten" | "Adult Education A, not completed", "Islamic elementary school (Madrasah), not completed" | "Adult Education A, completed", "Islamic elementary school (Madrasah), completed" | "Junior High General", "Junior High Vocational", "Junior/High School (Madrasah) ", "Adult Education B", "Senior High General, not completed", "Senior High Vocational, not completed", "Adult Education C, not completed", "Islamic Senior High School (Madrasah), not completed" | "Senior High General, completed", "Senior High Vocational, completed", "Adult Education C, completed", "Islamic Senior High School (Madrasah), completed" | "College", "University (Bachelor, Master, Doctorate)", "Open University" |
| **Iran** | Years of education = 0 | Years of education = 1-4 | Years of education = 5 | Years of education = 6-12 | Years of education = 13 | Years of education ≥14 |
| **Jordan 2019** | "No formal schooling" | "Primary school completed" | | "Secondary school completed" | "Post-Secondary school completed" | "College/University completed", "Postgraduate degree" |
| **Jordan 2017-2018** | "No education" | "Incomplete primary" | "Complete primary" | "Incomplete secondary" | "Complete secondary" | "Higher" |
| **Kenya 2015** | "No formal schooling" | "Less than primary school" | "Primary school completed" | "Secondary school completed" | "High school completed" | "College/University completed", "Postgraduate degree" |
| **Kenya 2014** | "No education" | "Incomplete primary" | "Complete primary" | "Incomplete secondary" | "Complete secondary" | "Higher" |
| **Kiribati** | "No formal schooling" | "Less than primary school" | "Primary school completed" | "Junior secondary school completed" | "Senior secondary school completed" | "College/University completed", "Postgraduate degree" |

| | | | | | | |
|---|---|---|---|---|---|---|
| **Kyrgyz Republic 2013** | "No formal schooling" and years of education = 0 | "No formal schooling" and years of education >0 | "Primary school competed (3rd year)" | "Secondary school completed (9th year)" | "High school completed (11th year)" | "University/Postgraduate degree" |
| **Kyrgyz Republic 2012** | "No education" | "Incomplete primary" | "Complete primary" | "Incomplete secondary" | "Complete secondary" | "Higher" |
| **Laos** | "No formal schooling" | "Less than primary school" | "Primary school completed" | "Secondary school completed" | "High school completed" | "College/University completed", "Postgraduate degree" |
| **Lesotho** | "No education" | "Incomplete primary" | "Complete primary" | "Incomplete secondary" | "Complete secondary" | "Higher" |
| **Liberia** | "No formal schooling" | "Less than primary school" | "Primary school completed" | "Junior high completed" | "Senior high completed" | "College/University completed", "Postgraduate degree" |
| **Madagascar** | "No education" | "Incomplete primary" | "Complete primary" | "Incomplete secondary" | "Complete secondary" | "Higher" |
| **Malawi 2015-2016** | "No education" | "Incomplete primary" | "Complete primary" | "Incomplete secondary" | "Complete secondary" | "Higher" |
| **Malawi 2009** | Years of education = 0 | Years of education = 1-7 | Years of education = 8 | Years of education = 9-11 | Years of education = 12 | Years of education ≥13 |
| **Mexico 2015** | "Without formal schooling" | "Primary" | | "Secondary", "Technical or trade secondary", "Normal basic" | "Preparatory or vocational", "Technical high school", "Technical or trade degree", "Normal upper level" | "Undergraduate", "Masters/Doctorate" |
| **Mexico 2009-2012** | "Without instruction", "Preschool", "Kindergarten" | "Elementary, year 1-5" | "Elementary, year 6" | "Secondary School", "Open Secondary school" | "High school", "Open High school", "Basic normal" | "College", "Graduate" |
| **Mongolia** | "No formal schooling" | "Less than primary school" | "Primary school completed" | "Secondary school completed" | "High school completed" | "Technical school/college completed", "University completed", "Postgraduate degree (Master/PhD)" |

| | | | | | | |
|---|---|---|---|---|---|---|
| **Mozambique 2011** | "No formal schooling" | "Incomplete primary (EP1" | "Primary school (EP1)", "Primary school (EP2)" | "Secondary school" | "Pre-University" | "University" |
| **Mozambique 2005** | "No formal schooling" | "Less than primary school" | "Primary school completed" | "Secondary school completed" | "High school completed" | "College/University completed", "Postgraduate degree" |
| **Myanmar 2015-2016** | "No education" | "Incomplete primary" | "Complete primary" | "Incomplete secondary" | "Complete secondary" | "Higher" |
| **Myanmar 2014** | "No formal schooling" | "Preschool to 3rd standard" | "4th to 7th standard" | "8th to 9th standard" | "10th standard completed" | "College/University completed", "Postgraduate degree" |
| **Namibia** | "No education" | "Incomplete primary" | "Complete primary" | "Incomplete secondary" | "Complete secondary" | "Higher" |
| **Niger 2012** | "No education" | "Incomplete primary" | "Complete primary" | "Incomplete secondary" | "Complete secondary" | "Higher" |
| **Niger 2007** | "No formal schooling" | "Incomplete primary" | "Complete primary" | "Complete secondary", "Koranic school" | "Complete High school" | "College/University", "Postgraduate degree" |
| **Nigeria** | "No education" | "Incomplete primary" | "Complete primary" | "Incomplete secondary" | "Complete secondary" | "Higher" |
| **Pakistan** | "No education" | "Incomplete primary" | "Complete primary" | "Incomplete secondary" | "Complete secondary" | "Higher" |
| **Panama** | "No formal schooling", "Special education" | "Less than primary school completed" | "Primary school completed" | "Less than secondary school completed" | "Secondary school completed", "Vocational", "Superior, no university" | "College/University completed", "Postgraduate degree completed" |
| **Peru** | "No education" | "Incomplete primary" | "Complete primary" | "Incomplete secondary" | "Complete secondary" | "Higher" |
| **Philippines** | "No formal schooling" | "Elementary undergraduate" | "Elementary graduate" | "High school undergraduate" | "High school graduate" | "Post secondary years 1, 2 or 3", "College undergraduate", "College graduate", "Postgraduate degree completed" |

| | | | | | | |
|---|---|---|---|---|---|---|
| **Romania** | “No Education” | “Primary school” | | “Secondary school” | | “Higher Education” |
| **Rwanda 2014-2015** | “No education” | “Incomplete primary” | “Complete primary” | “Incomplete secondary” | “Complete secondary” | “Higher” |
| **Rwanda 2012** | “No formal schooling” | “Less than primary school” | “Primary school completed” | “Incomplete secondary” | | “College/University completed”, “Postgraduate degree completed” |
| **Samoa** | “No formal schooling” | “Incomplete primary” | “Complete primary” | “Complete secondary” | “Complete High school” | “College/University”, “Postgraduate degree” |
| **São Tomé and Principe** | “No formal schooling” | “Less than primary school” | “Primary school completed” | “Complete secondary” | “Complete High school” | “College/University”, “Postgraduate degree” |
| **Senegal 2015**[2] | “No formal schooling“ | “Less than primary school complete“ | “Primary school completed“ | “Less than secondary school completed“ | “Secondary school completed“ | “College/University completed“, “Postgraduate degree completed“ |
| **Senegal 2010-2011** | “No education” | “Incomplete primary” | “Complete primary” | “Incomplete secondary” | “Complete secondary” | “Higher” |
| **Solomon Islands** | “No formal schooling” | “Less than primary school” | “Primary school completed” | “Complete secondary” | “Complete High school” | “College/University”, “Postgraduate degree” |
| **South Africa** | “No education” | “Incomplete primary” | “Complete primary” | “Incomplete secondary” | “Complete secondary” | “Higher” |
| **Tajikistan 2017** | “No education” | “Incomplete primary” | “Complete primary” | “Incomplete secondary” | “Complete secondary” | “Higher” |
| **Tajikistan 2016** | “No formal schooling” | “Primary school completed” and years of education <4 | “Primary school completed”, and years of education = 4 | “Secondary school completed” | “High school completed” | “College/University completed”, “Postgraduate degree” |
| **Tanzania 2015-2016** | “No education” | “Incomplete primary” | “Complete primary” | “Incomplete secondary” | “Complete secondary” | “Higher” |
| **Tanzania 2012** | “No formal schooling” | “Less than primary school” | “Primary school completed” | “Secondary school completed” | “High school completed” | “College/University completed”, “Postgraduate degree” |
| **Timor-Leste 2016** | “No education” | “Incomplete primary” | “Complete primary” | “Incomplete secondary” | “Complete secondary” | “Higher” |

| | | | | | | |
|---|---|---|---|---|---|---|
| **Timor-Leste 2014** | "No formal schooling" | "Less than primary school (grade 6 not completed)" | "Primary school completed (grade 6 completed)" | "Pre-Secondary school completed (grade 9 completed)" | "Secondary school completed (grade 12 completed)" | "College Diploma completed (1-3 years)", "College/University completed (4-6 years)", "Postgraduate degree" |
| **Togo 2013-2014** | "No education" | "Incomplete primary" | "Complete primary" | "Incomplete secondary" | "Complete secondary" | "Higher" |
| **Togo 2010** | "No formal schooling" | "Less than primary school" | "Primary school completed" | "Lower secondary education completed (college)" | "Upper secondary education completed (lycée or equivalent)" | "Higher schooling, university" |
| **Tonga** | "Pre-school" | "Primary" | "Lower secondary" | "Upper secondary" | "Technical and vocational" | "Tertiary" |
| **Turkey** | "No education" | "Incomplete primary" | "Complete primary" | "Incomplete secondary" | "Complete secondary" | "Higher |
| **Tuvalu** | "No formal schooling" | "Less than primary school" | "Primary school completed" | "Secondary school completed" | "High school completed" | "College/University completed", "Postgraduate degree" |
| **Uganda 2016** | "No education" | "Incomplete primary" | "Complete primary" | "Incomplete secondary" | "Complete secondary" | "Higher" |
| **Uganda 2014** | "No formal schooling" | Years of education = 1-5 | "Primary school completed" | "O-Level completed", "Junior Secondary completed" | "A-Level completed" | "University/higher institutions completed", "Postgraduate degree" |
| **Uruguay** | "No formal schooling" | "Less than primary school completed" | "Primary school completed" | "Less than secondary school completed" | "Secondary school completed" | "College/University completed", "Postgraduate degree completed" |
| **Vanuatu** | "No formal schooling" | "Less than primary school" | "Primary school completed" | "Secondary school completed" and years of education <12 | "Secondary school completed" and years of education ≥12" | "College/University completed", "Postgraduate degree" |
| **Vietnam** | "No formal education" | "Not having completed primary education" | "Having completed primary education" | "Having completed basic secondary school" | "Having completed secondary school" | "Graduated university/college/specialized secondary education", "Having been post-graduated" |

| | | | | | | |
|---|---|---|---|---|---|---|
| **Yemen** | Not asked | | | | | |
| **Zambia 2018** | "No education" | "Incomplete primary" | "Complete primary" | "Incomplete secondary" | "Complete secondary" | "Higher" |
| **Zambia 2017** | "No formal schooling" | "Less than primary school" | "Primary school completed" | "Junior secondary school completed" | "Secondary Higher school completed" | "College/University completed", "Postgraduate degree" |
| **Zanzibar** | "No formal schooling" | "Less than primary school" | "Primary school completed (P7)" | "Secondary school completed (form four)" | "High school completed (form six)" | "College/University completed", "Postgraduate degree" |
| **Zimbabwe** | "No education" | "Incomplete primary" | "Complete primary" | "Incomplete secondary" | "Complete secondary" | "Higher" |

[1]The IFLS (Indonesia) included two additional categories: Islamic Schools ("Pesantren") and schools for individuals with a disability. As we could not determine how these correspond to the general school system, respondents of both education categories were excluded from analyses involving education (103 out of a total of 32467 participants).
[2]No country-specific questionnaire available. Education categories were thus derived from the GATS standard questionnaire.

### Materials and Methods S5: Statistical Analysis

#### *5.1. Cleaning and definition of risk factors*

*Lipids*

Dyslipidemia can be determined from total cholesterol, LDL cholesterol, triglycerides and HDL cholesterol. As the relevance and efficacy of triglycerides and HDL cholesterol remain debated (*35*), we decided to limit our analysis on two dyslipidemia biomarkers, namely total cholesterol and LDL cholesterol. However, triglycerides and HDL cholesterol measurements were used for the calculation of LDL cholesterol and therefore included in the data cleaning process outlined below.

Regarding lower cut-offs, we followed a conservative exclusion procedure that accounts for measurement ranges of the most commonly applied point-of-care measurement devices and excluded values below 3 mg/dL for total cholesterol, triglycerides, and HDL cholesterol (*61*). We determined upper cut-offs by again considering measurement ranges of point-of-care devices and following thresholds that have proved reasonable in other studies (*62*). Consequently, total cholesterol values above 300 mg/dL, triglycerides values larger than 600 mg/dL and HDL cholesterol measures greater than 100 mg/dL were excluded during the data cleaning process. LDL cholesterol was labeled N/A in the few cases where our calculated values turned out to be negative.

Following the outlined cleaning process we assessed LDL cholesterol values from the Friedewald equation:

$$\text{LDL (mg/dL)} = \text{TC (mg/dL)} - \text{HDL (mg/dL)} - \text{TG (mg/dL)} / 5$$

Based on the ATP III guidelines, participants were classified to have dyslipidemia if their calculated LDL cholesterol level was higher than 4.14 mmol/L or when they reported to use medication for dyslipidemia. In contrast to LDL cholesterol, total cholesterol measurements can be directly used to determine dyslipidemia and respondents with a total cholesterol level above 6.21 mmol/L (*35*) or those using medication to lower cholesterol levels were considered to suffer from dyslipidemia.

Cleaning and definition of remaining risk factors is described in the main manuscript methods section.

#### *5.2. Regression analyses*

For interpretation, regression tables report risk ratios (RR), average marginal effects (AME), and their respective p-values, which we assume to indicate statistical significance when below 0.05. These indicators can be interpreted as follows: RR is a relative ratio that shows how many times more probable the occurrence of a risk factor is among groups with the respective individual level characteristic compared to the reference group. A value above 1 indicates a greater risk and a value below 1 suggests a reduced risk for this group. RRs of 1 or close to 1 can be interpreted as both groups having a similar risk to retain the risk factor. RRs allow easier interpretation compared with other ratios (notably Odds Ratio), which drove our decision to use Poisson regression instead of logistic regression in our analysis. The average marginal effect (AME) shows the average absolute change in prevalence when the predictor (individual-level characteristic) changes by one unit. We assumed that regression categories (covariates, poverty level) needed a minimum sample size of 20 to yield valid results. Hence, if e.g., one poverty level had less than 20 male adults with obesity, regressions were not conducted for sex for that particular group. The place of residence variable (urban/rural) was missing for Kiribati (2015), Myanmar (2014), Samoa (2013), Solomon Islands (2015), Timor-Leste (2014), Tonga (2017), Tuvalu (2015), Vanuatu (2011), Albania (2008-2009), Tajikistan (2016), Ecuador (2018), Mexico (2009-2012), Egypt (2015), Botswana (2014), Cabo Verde (2007), Comoros (2011), Eswatini (2014), Liberia (2011), Niger (2007), Rwanda (2012) and São Tomé and Principe (2008). Education was missing for Yemen (2013).

#### *5.3. Computation of the care cascade for hypertension, diabetes and statins*

*Hypertension*

Our cascade model used respondents diagnosed with hypertension as a fixed denominator for both stages. The numerator is the proportion of participants with hypertension that reached the respective stage, given that they achieved the previous stage in step 2. In other words, if a respondent said that s/he does not use any anti-hypertensive medication, the answer for step 2 was set to "no". For both steps, answers were treated as "no" if respondents did not know the answer or refused to give any answer at all.

Step 1: Is using blood pressure lowering medication ("Hypt treated")
Questions regarding the use of anti-hypertensive medication were asked in 47 countries. All countries except for three asked for the current drug use, by explicitly asking for either current use or usage within the last two weeks. For Ghana, Iran and Romania, the specific questions could not be determined as the survey questionnaires were unavailable. Although often asked in surveys, lifestyle advice (e.g., exercise, diet, and weight loss) was not included in our variable. A detailed list of the respective questions is provided in Table S7.

Step 2: Is using blood pressure lowering medication and has normal BP ("Hypt controlled")
This step included all respondents diagnosed with hypertension that reported the use of anti-hypertensive medication and had normal BP levels (systolic BP <140 mmHg and a diastolic BP <90 mmHg). This information was available for 50 countries.

*Diabetes*

We followed the same procedure to determine the share of diabetics that uses medication to lower blood glucose levels.
Step 1: Is using medication for diabetes ("Dia treated")

Questions on medication use were asked in 44 countries. For three of these (India, Iran, Romania), specific questions could not be determined since the survey questionnaire was not available. For the remaining countries, all except for China explicitly asked for current medication use, by asking respondents whether they currently use medication or used medication within the last two weeks. In China, the survey asked respondents: "Did you use any of these treatment methods? [Oral medication, insulin,...]" allowing the indication of current or past drug use. We constructed our variable "Dia treated" to include both oral medication and insulin but no lifestyle advice.

*Statins*

Questions on statin use were asked in 19 countries. According to WHO guidelines, statin use is recommended as a measure of secondary prevention for individuals with a self-reported history of CVD ("statin secondary"). For individuals without a prior CVD history, statin use as primary prevention is suggested for individuals aged 40 or above who have either been diagnosed with diabetes or who have a 10-year CVD risk higher than 20% (based on 2019 WHO laboratory-based risk equations) (*41*) ("statin primary"). The 10-year CVD risk factor is determined by considering the factors age, smoking status, systolic blood pressure, history of diabetes and total cholesterol and was calculated with the *whoishRisk* package in R (*63*). Consequently, samples for the calculation of statin use varied depending on whether we calculated the outcome for primary or secondary prevention, their respective definition is outlined in Table S12 below.

Table S12: Definitions of outcome denominators for statin use*

| **Outcome** | **Denominator** |
|---|---|
| Secondary prevention | Number of non-pregnant adults ages 40-69 years who self report prior CVD |
| Primary prevention | Number of non-pregnant adults ages 40-69 years without a history of self-reported CVD and either (1) a history of self-reported diabetes or (2) 10-year CVD risk >20% using the 2019 WHO laboratory-based risk equations |

*Definitions followed the indicator recommended in the WHO NCD Global Monitoring Framework (*64*), WHO HEARTS Technical Package for CVD Management in Primary Health Care (*65*), and the WHO-PEN clinical guidelines (*40*).

Diabetes was defined as either (1) a self-reported diagnosis provided by the respondent, (2) a self-reported use of diabetes medication, or (3) high glucose levels during biochemical measurements of the STEPS surveys, for which we applied the same definition as for our diabetes risk factor outlined in the methods section of the manuscript. We used demographic weights, corresponding to Step 1 weights in STEP surveys, to calculate the secondary prevention outcome and biomarker-based weights (Step 3 weights) for the calculation of 10-year CVD risk scores.

We plotted percentages with 95% confidence intervals for the proportion of individuals using anti-hypertensive medication ("Hypt treated"), with controlled hypertension ("Hypt Controlled"), the percentage using drugs to lower blood glucose levels ("Dia treated"), and statins for secondary and primary prevention of CVD ("Statin secondary", "Statin primary") in Figure 4 in the manuscript. To highlight differences in the care continuum, percentages were stratified by poverty level (poverty headcount ratio <$1.90/day; <$3.20/day; <$5.50/day; >$5.50/day and general LMIC population as well as extremely poor vs. non-poor) and age group (15-24, 25-34, 35-44, 45-54, 66-64, 65+). In addition, we created two separate plots for females and males (Figures S14-18). Prevalences for all cascade steps and poverty levels are outlined further in Tables S19-21.

#### *5.4. Computation of wealth measures*

Sixty-seven surveys included a continuous wealth index. This index was created following the standard approach of DHS surveys, which uses information about dwelling characteristics and household ownership of durable goods (i.e., goods that are not consumed in one use) (*66*). For this approach, each dwelling characteristic and durable good is labeled with a binary indicator. After conducting a principal component analysis (PCA) on these binary indicator variables, the first unrotated principal component is used for the wealth index (*67*). Thirty-three surveys provided continuous income measures. Participants could self-report their continuous income for the year preceding data collection on a weekly, monthly, or yearly basis. We used income categories or income quintiles when participants were unable to estimate their exact earnings and instead provided estimates in pre-coded income groups. Income categories were used when participants were asked to estimate their average earnings in four pre-coded income categories given in local currencies, which was the case for the Tonga STEPS 2017 survey. Four surveys provided income quintiles. In these surveys, income estimates were given in five pre-coded income groups ranging from 1 (lowest wealth quintile covering the poorest 20% of the sample) to 5 (highest wealth quintile covering the wealthiest 20% of the sample). Table S13 below shows all wealth measures that were used for each survey.

*Definition of poverty and creation of poverty levels*

We used the World Bank's poverty headcount ratio, an annual estimate of the proportion of a country's population living below the national poverty line(s), to create four poverty levels. Poverty headcount ratio thresholds are defined by an income (all in 2011 purchasing-power-parity adjusted dollars) of $1.90 per day, $3.20 per day, and $5.50 per day (*27,56,57,68*). Even though our analysis is focused on the prevalence of CVD risk factors among the extremely poor (those living below the international poverty line of $1.90 per day), we also determined populations with an income below $3.20 per day, $5.50 per day, and above $5.50 per day to allow comparisons between different poverty levels within LMICs. If poverty headcount ratios were missing for a survey year, we imputed them with the Kalman filter imputation method (*69*).

Given that all surveys used in our analysis provide nationally representative data, we used the distribution of wealth or income in each survey dataset to determine the share of participants falling below each poverty threshold ($1.90/day, $3.20/day, $5.50/day, above $5.50/day). First, we ordered participants within each survey by their wealth or income from 'poorest' to 'richest'. We then applied the WB poverty headcount ratio of the respective survey year to determine the proportion of participants with the lowest wealth or income that fell below each poverty headcount ratio threshold. For example, assuming a country with the poverty headcount ratios of 10% ($1.90/day), 20% ($3.20/day) and 40% ($5.50/day) and a survey dataset of 5,000 participants, our classification would yield 500 participants with the lowest income in the lowest poverty level ($1.90/day), 1,000 participants in the $3.20/day poverty level, 2,000 participants in the $5.50/day poverty level, and 3,000 participants in the 'above $5.50/day' group.

Table S13. Wealth measures by country

| Wealth measure | Country |
|---|---|
| Wealth index | Albania (2008/2009), Albania (2017/2018), Angola, Armenia, Bangladesh (2017), Bangladesh (2017/2018), Bolivia, Botswana (2017), Brazil, Burkina Faso, Burundi, Cameroon, Chad, China (2010), Comoros (2012), Congo Democratic Republic, Congo, Costa Rica, Côte d'Ivoire, Dominican Republic, Egypt (2014), Egypt (2015), Ethiopia (2016), Gabon, Gambia (2013), Ghana (2007/2008), Ghana (2014), Guatemala, Guinea, Haiti, Honduras, India (2015/2016), India (2017), Indonesia, Iran, Jordan (2017/2018), Kenya (2014), Kenya (2015), Kyrgyz Republic (2012), Lesotho, Madagascar, Malawi (2015/2016), Mexico (2015), Mozambique (2011), Myanmar (2015/2016), Namibia, Niger (2012), Nigeria, Pakistan, Panama, Peru, Philippines, Rwanda (2014/2015), Senegal (2010/2011), Senegal (2015), South Africa, Tajikistan (2017), Tanzania (2015/2016), Timor-Leste (2016), Togo (2013/2014), Turkey, Uganda (2016), Uruguay, Vietnam, Yemen, Zambia (2018), Zimbabwe |
| Continuous income | Benin, Bhutan, Botswana (2014), Cabo Verde, China (2009), Comoros (2011), Ecuador, Eswatini, Gambia (2010), Georgia, Kiribati, Kyrgyz Republic (2013), Lao PDR, Liberia, Malawi (2009), Mexico (2009-2012), Mozambique (2005), Myanmar (2014), Niger (2007), Romania, Rwanda (2012), Samoa, Sao Tome and Principe, Solomon Islands, Tanzania (2012), Tajikistan (2016), Timor-Leste (2014), Togo (2010), Tuvalu, Uganda (2014), Vanuatu, Zambia (2017), Zanzibar |
| Wealth quintiles | Botswana (2017), Ethiopia (2015), Jordan (2019), Mongolia |
| Income categories | Tonga |

### Detailed Results

Figure S1: Flowchart for inclusion of STEPS surveys

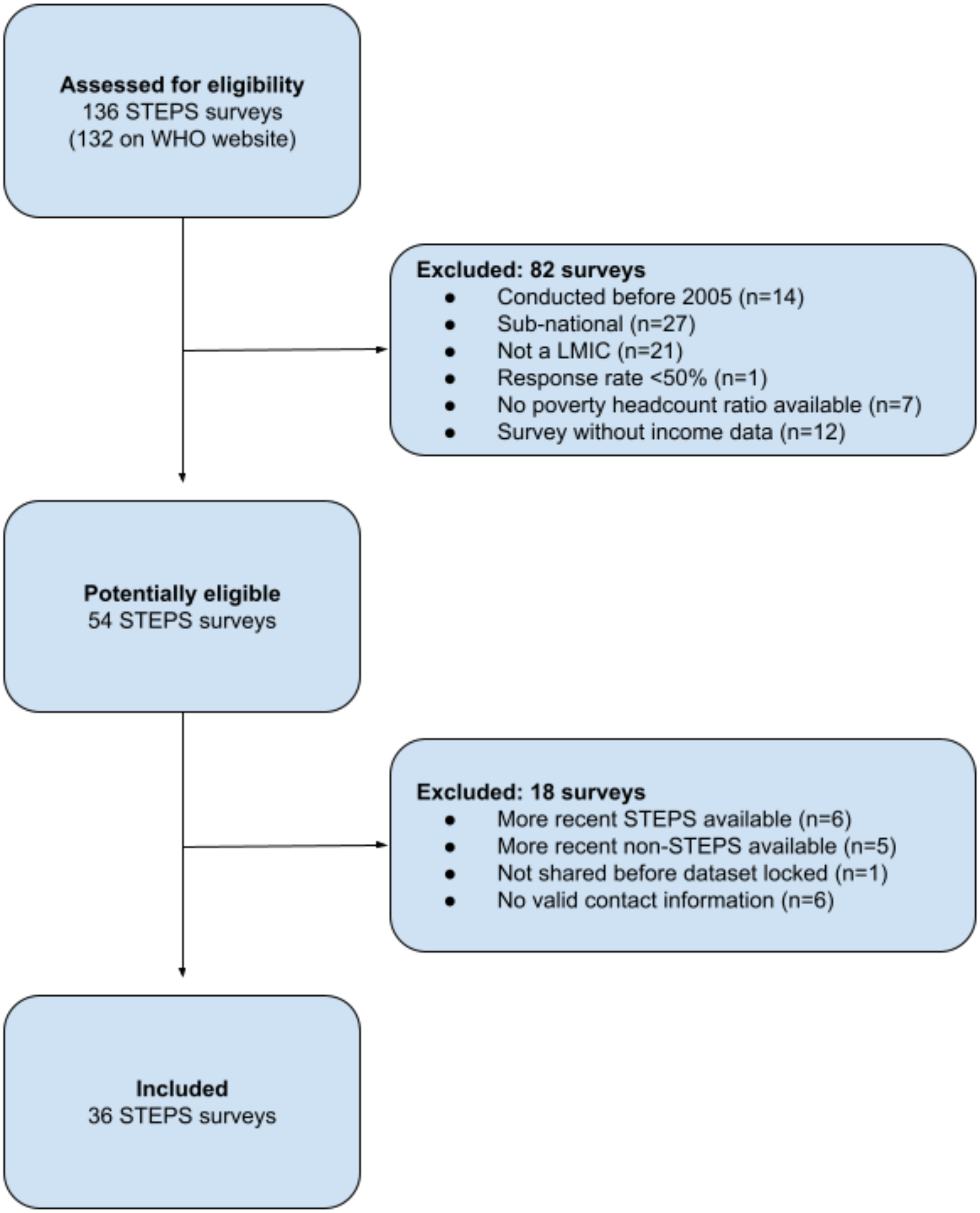

Figure S2: Flowchart for inclusion of DHS surveys

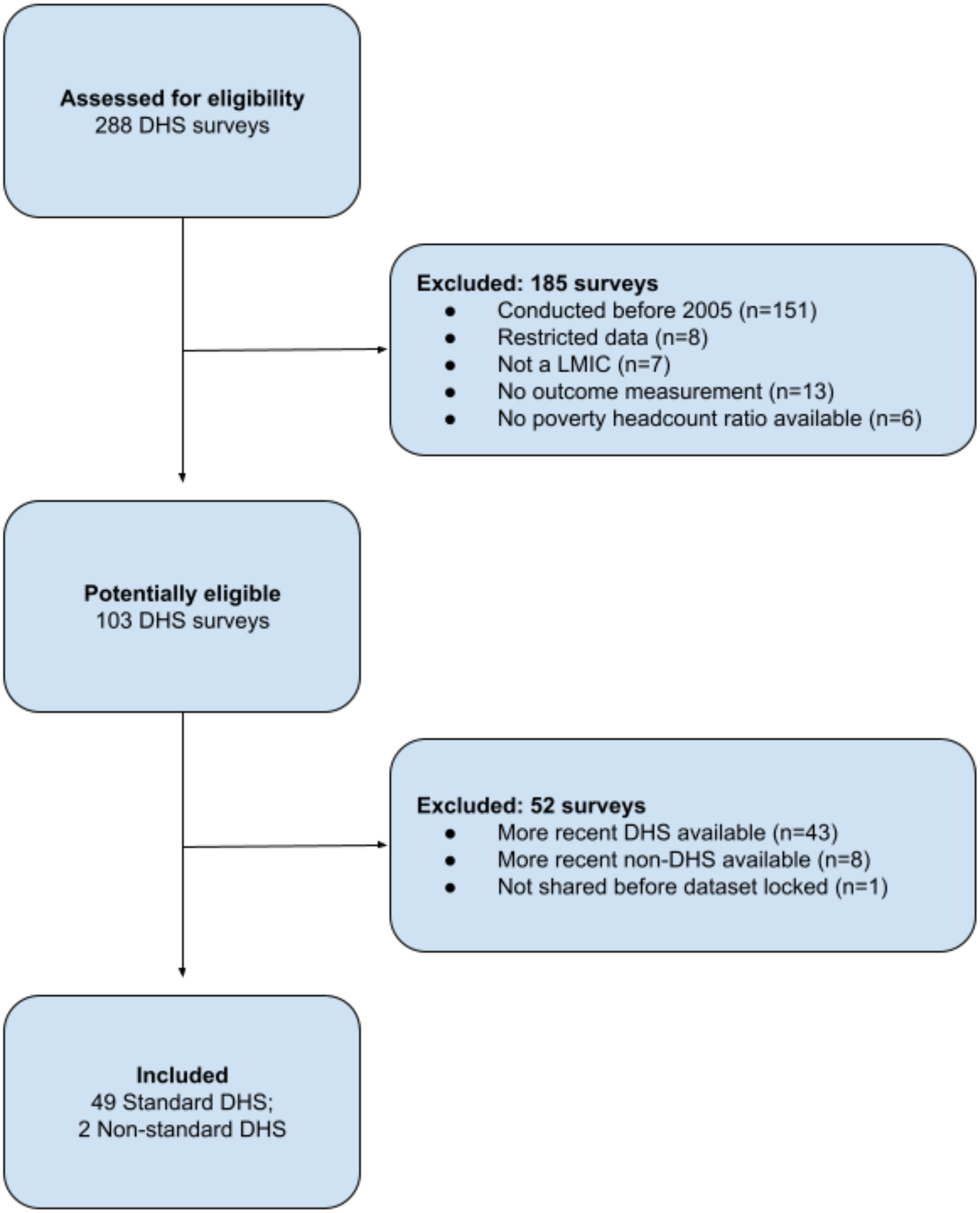

Figure S3: Flowchart for inclusion of GATS surveys

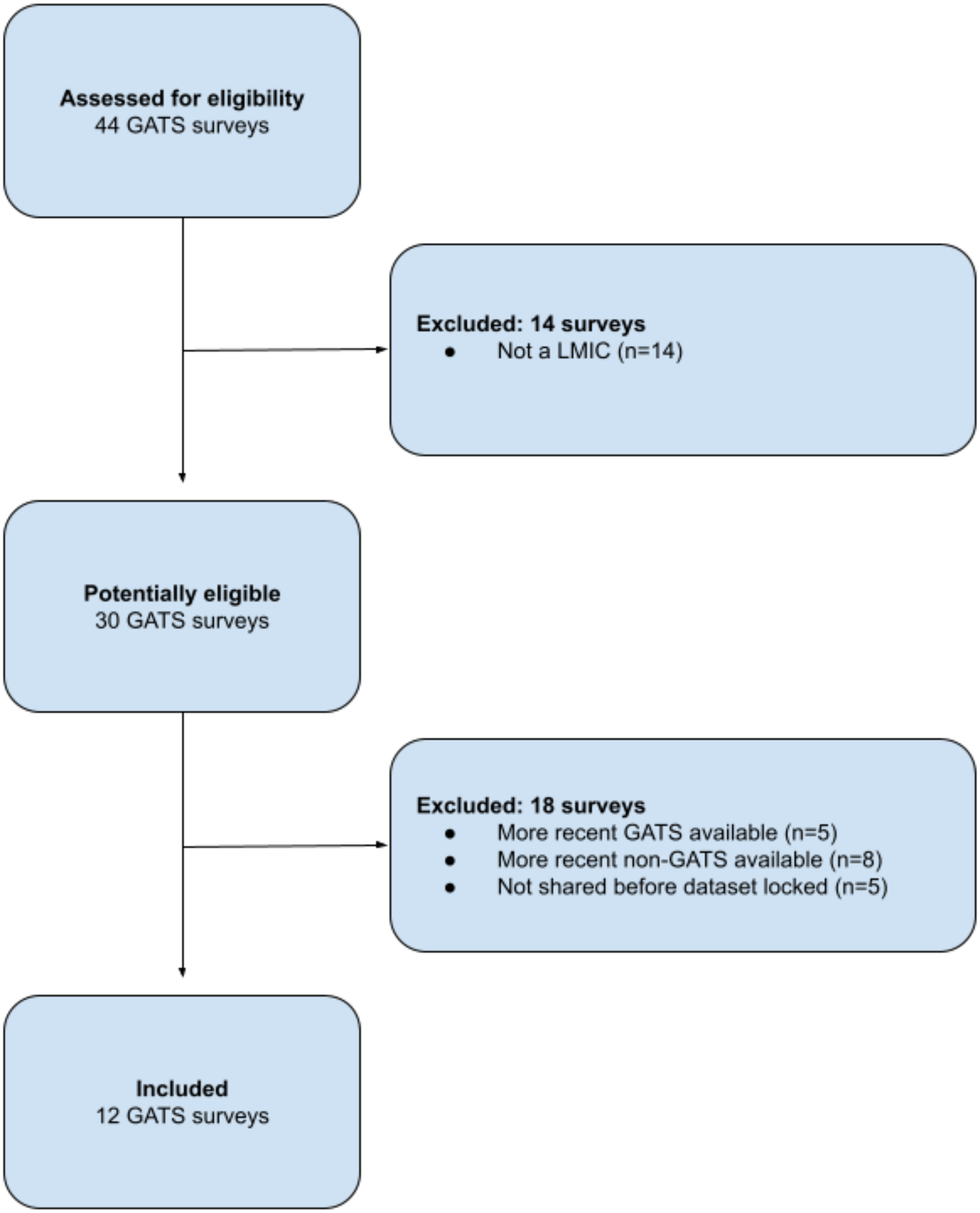

Figure S4: Flowchart for inclusion of surveys from Google search

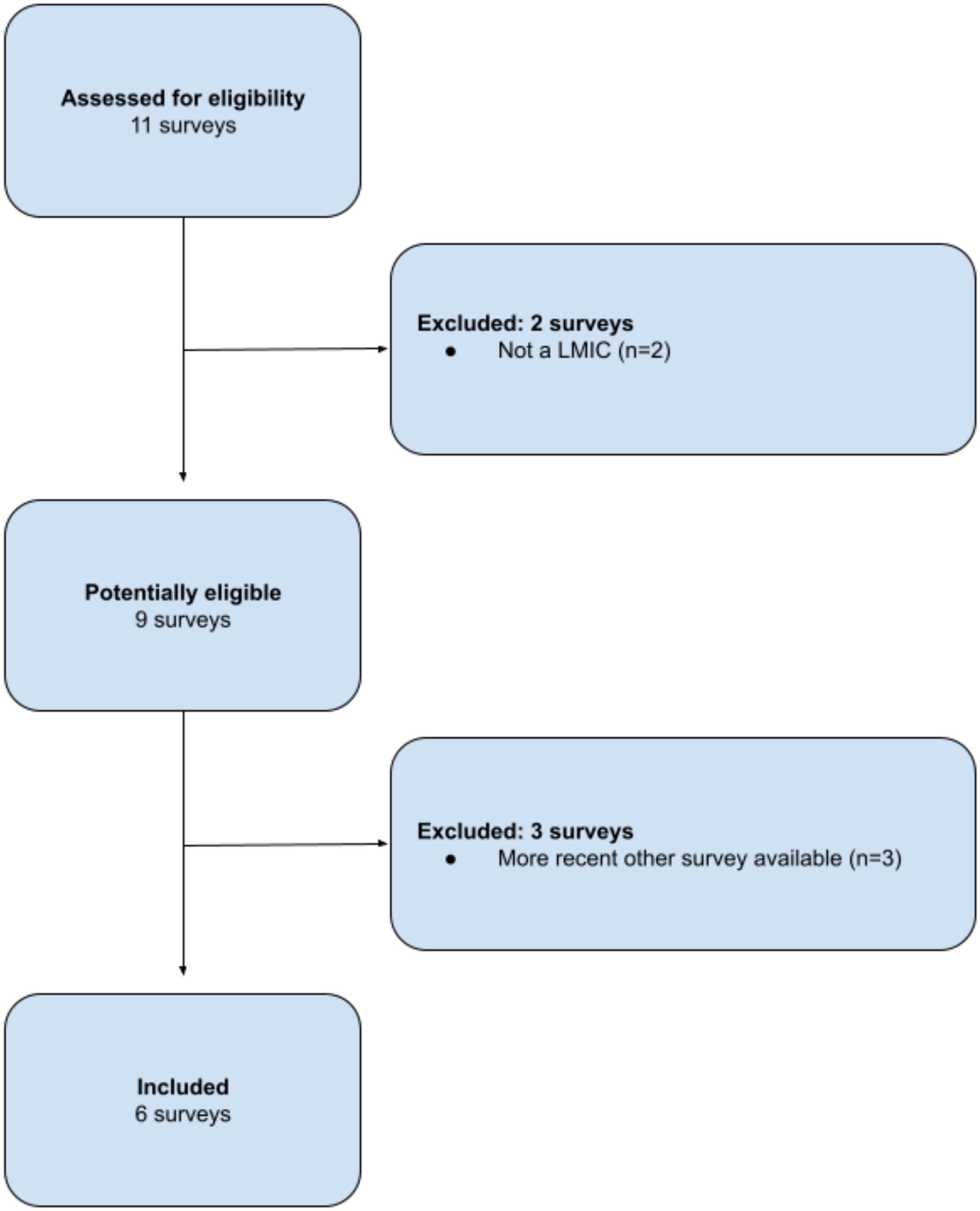

Table S14: Prevalences by 10-year age groups and poverty level

| Poverty level | Hypertension | | Diabetes | | Overweight | | Obesity | | Smoking | | Dyslipidemia | |
|---|---|---|---|---|---|---|---|---|---|---|---|---|
| | Prevalence | Obs. | Prevalence | Obs. | Prevalence | Obs. | Prevalence | Obs. | Prevalence | Obs. | Prevalence | Obs. |
| ***All age groups*** | | | | | | | | | | | | |
| **Total** | 19.1 (18.6-19.7) | 1071000 | 5.7 (5.2-6.2) | 900986 | 31.4 (30.3-32.3) | 1283946 | 9.6 (9.1-10.1) | 1283946 | 19.2 (16.2-22.2) | 916495 | 8.3 (6.4-10.2) | 70591 |
| **<$1.90/day** | 17.5 (16.7-18.3) | 205029 | 4.0 (3.6-4.5) | 193554 | 13.3 (12.6-14.1) | 265232 | 3.1 (2.8-3.3) | 265232 | 10.6 (9.0-12.3) | 192046 | 1.4 (0.9-1.9) | 16851 |
| **<$3.20/day** | 17.3 (16.7-17.9) | 533696 | 4.2 (3.9-4.5) | 502414 | 18.8 (18.2-19.4) | 623345 | 4.7 (4.5-4.9) | 623345 | 15.9 (12.8-18.9) | 373299 | 2.7 (1.9-3.5) | 31303 |
| **<$5.50/day** | 18.2 (17.6-18.8) | 804359 | 4.8 (4.5-5.1) | 738802 | 25.5 (24.9-26.1) | 935766 | 7.3 (7.0-7.6) | 935766 | 17.9 (14.3-21.4) | 568738 | 5.1 (3.8-6.4) | 46179 |
| **>$5.50/day** | 20.8 (20.0-21.6) | 266641 | 7.6 (6.5-8.7) | 162184 | 42.8 (41.1-44.6) | 348180 | 14.2 (13.2-15.2) | 348180 | 21.2 (18.7-23.7) | 347757 | 17.0 (12.6-21.4) | 24412 |
| ***15-24*** | | | | | | | | | | | | |
| **Total** | 5.7 (5.4-6.1) | 294096 | 2.5 (2.2-2.8) | 269746 | 14.4 (13.6-15.2) | 408375 | 3.7 (3.5-4.0) | 408375 | 11.7 (9.5-14.2) | 255756 | 0.8 (0.5-1.4) | 8557 |
| **<$1.90/day** | 6.1 (5.4-6.8) | 62004 | 2.1 (1.9-2.4) | 60418 | 6.4 (5.8-7.0) | 92051 | 1.6 (1.5-1.8) | 92051 | 5.2 (4.1-6.5) | 62074 | 0.3 (0.1-0.7) | 2666 |
| **<$3.20/day** | 5.7 (5.3-6.1) | 167747 | 2.2 (1.9-2.4) | 163064 | 8.3 (7.9-8.7) | 216857 | 1.9 (1.7-2.0) | 216857 | 8.6 (6.6-11.1) | 117149 | 0.4 (0.2-0.8) | 4388 |
| **<$5.50/day** | 5.8 (5.5-6.1) | 244522 | 2.4 (2.1-2.6) | 233848 | 11.3 (10.8-11.9) | 316799 | 2.8 (2.6-3.0) | 316799 | 10.2 (8.0-13) | 172933 | 0.4 (0.2-0.7) | 5844 |
| **>$5.50/day** | 5.5 (4.8-6.4) | 49574 | 2.8 (2.2-3.5) | 35898 | 22.9 (21.4-24.6) | 91576 | 6.4 (5.8-7.0) | 91576 | 14.4 (11.9-17.3) | 82823 | 2.7 (1.4-5.0) | 2713 |
| ***25-34*** | | | | | | | | | | | | |
| **Total** | 11.7 (11.2-12.1) | 289977 | 3.3 (2.9-3.6) | 255617 | 32.9 (31.8-34.0) | 371313 | 10.5 (10.0-11.0) | 371313 | 17.7 (14.8-21.1) | 233439 | 4.2 (2.3-7.5) | 19085 |
| **<$1.90/day** | 12.2 (11.5-13.0) | 58727 | 2.6 (2.3-2.9) | 56042 | 12.0 (11.2-12.7) | 79091 | 2.8 (2.5-3.1) | 79091 | 9.7 (8.0-11.6) | 52270 | 0.4 (0.2-0.8) | 4978 |
| **<$3.20/day** | 12.0 (11.5-12.5) | 150341 | 2.6 (2.4-2.9) | 143497 | 19.3 (18.6-20.0) | 182410 | 5.0 (4.7-5.3) | 182410 | 14.2 (11.2-17.7) | 99233 | 1.2 (0.7-2.2) | 9067 |
| **<$5.50/day** | 12.1 (11.7-12.6) | 225807 | 3.0 (2.8-3.2) | 211677 | 27.8 (27.0-28.5) | 274715 | 8.5 (8.1-8.9) | 274715 | 16.1 (12.8-19.9) | 149929 | 2.0 (1.2-3.3) | 12984 |
| **>$5.50/day** | 10.7 (9.8-11.7) | 64170 | 4.0 (3.2-4.9) | 43940 | 43.9 (41.9-45.9) | 96598 | 14.8 (13.7-15.9) | 96598 | 20.4 (17.5-23.5) | 83510 | 11.4 (5-23.8.0) | 6101 |
| ***35-44*** | | | | | | | | | | | | |
| **Total** | 19.8 (19.0-20.6) | 259070 | 5.5 (5.1-6.0) | 222520 | 40.4 (39.0-41.8) | 309371 | 13.6 (12.8-14.4) | 309371 | 22.6 (19.5-26.0) | 190686 | 9.0 (6.9-11.5) | 17171 |
| **<$1.90/day** | 18.9 (17.8-20.0) | 50051 | 4.4 (3.6-5.2) | 47887 | 17.7 (16.3-19.1) | 63409 | 4.0 (3.5-4.6) | 63409 | 12.5 (10.7-14.7) | 39267 | 1.4 (0.7-2.8) | 3758 |

| | | | | | | | | | | | | |
|---|---|---|---|---|---|---|---|---|---|---|---|---|
| **<$3.20/day** | 19.4 (18.6-20.2) | 128906 | 4.9 (4.3-5.4) | 122682 | 26.1 (25.1-27.1) | 149314 | 6.9 (6.4-7.5) | 149314 | 19.1 (16.0-22.6) | 76794 | 2.8 (1.8-4.2) | 7345 |
| **<$5.50/day** | 20.2 (19.4-21.0) | 194850 | 5.2 (4.8-5.7) | 181564 | 34.4 (33.4-35.4) | 224920 | 10.8 (10.2-11.4) | 224920 | 21.3 (17.8-25.3) | 117792 | 5.8 (4.2-8.0) | 11111 |
| **>$5.50/day** | 19.1 (17.8-20.6) | 64220 | 6.1 (5.2-7.2) | 40956 | 51.0 (48.3-53.6) | 84451 | 18.5 (17.0-20.2) | 84451 | 24.4 (21.5-27.7) | 72894 | 17.6 (13.1-23.2) | 6060 |
| ***45-54*** | | | | | | | | | | | | |
| **Total** | 31.2 (30.0-32.4) | 153018 | 9.1 (7.9-10.4) | 119777 | 43.0 (41.2-44.7) | 149469 | 13.4 (12.4-14.4) | 149469 | 28.9 (25.1-33.0) | 123895 | 17.7 (14.5-21.3) | 13925 |
| **<$1.90/day** | 31.6 (29.1-34.2) | 25904 | 5.2 (4.1-6.6) | 23847 | 24.2 (21.7-26.8) | 27057 | 5.3 (4.3-6.5) | 27057 | 20.1 (17.0-23.6) | 22074 | 2.7 (1.6-4.6) | 2861 |
| **<$3.20/day** | 29.9 (28.3-31.5) | 67259 | 6.2 (5.3-7.2) | 61594 | 28.4 (26.9-30.0) | 65546 | 7.5 (6.7-8.3) | 65546 | 27.0 (22.6-31.8) | 44626 | 6.8 (4.2-10.7) | 5585 |
| **<$5.50/day** | 31.2 (29.8-32.7) | 104435 | 7.6 (6.8-8.5) | 92618 | 36.3 (34.8-37.9) | 101073 | 10.4 (9.6-11.2) | 101073 | 28.9 (24.3-34.0) | 70350 | 12.8 (9.4-17.1) | 8647 |
| **>$5.50/day** | 31.2 (29.5-33.0) | 48583 | 11.3 (9.3-13.6) | 27159 | 52.0 (49.1-54.8) | 48396 | 17.4 (15.8-19.2) | 48396 | 28.9 (25.6-32.4) | 53545 | 25.4 (18.8-33.3) | 5278 |
| ***55-64*** | | | | | | | | | | | | |
| **Total** | 42.2 (40.3-44.2) | 44360 | 11.7 (10.3-13.3) | 22497 | 38.6 (36.2-41.0) | 27589 | 8.6 (7.6-9.7) | 27589 | 30.5 (26.7-34.6) | 64655 | 21.1 (17.2-25.8) | 9598 |
| **<$1.90/day** | 39.4 (35.1-43.9) | 5534 | 11.7 (8.9-15.2) | 3990 | 28.8 (23.9-34.4) | 2288 | 5.4 (3.4-8.6) | 2288 | 31.2 (26.8-36.0) | 9273 | 7.5 (4.4-12.5) | 2132 |
| **<$3.20/day** | 41.9 (38.7-45.1) | 12222 | 9.4 (7.8-11.3) | 8243 | 30.2 (26.8-33.8) | 5730 | 5.4 (4.2-7.0) | 5730 | 37.0 (31.4-42.8) | 20475 | 9.5 (6.5-13.7) | 3997 |
| **<$5.50/day** | 41.4 (39.0-43.8) | 21371 | 9.6 (8.3-11.0) | 13352 | 31.6 (29.1-34.4) | 11320 | 5.5 (4.6-6.7) | 11320 | 35.9 (30.4-41.7) | 33478 | 16.3 (12.8-20.7) | 6195 |
| **>$5.50/day** | 43.3 (40.6-45.9) | 22989 | 14.9 (12.6-17.4) | 9145 | 46.5 (43.3-49.8) | 16269 | 12.2 (10.7-13.8) | 16269 | 25.5 (22.9-28.3) | 31177 | 30.0 (21.1-40.8) | 3403 |
| ***>65*** | | | | | | | | | | | | |
| **Total** | 55.4 (52.9-57.8) | 30479 | 14.2 (12.6-16.0) | 10829 | 35.4 (32.9-37.9) | 17829 | 8.8 (7.7-10.1) | 17829 | 22.9 (19.9-26.2) | 48064 | 12.8 (8.4-18.9) | 2255 |
| **<$1.90/day** | 55.7 (50.8-60.5) | 2809 | 11.1 (8.1-15.1) | 1370 | 27.1 (22.4-32.3) | 1336 | 6.3 (4.2-9.4) | 1336 | 28.7 (24.4-33.5) | 7088 | 4.7 (2.0-10.4) | 456 |
| **<$3.20/day** | 53.0 (49.4-56.5) | 7221 | 11.9 (9.7-14.4) | 3334 | 26.9 (23.4-30.7) | 3488 | 6.0 (4.5-8.0) | 3488 | 31.9 (26.8-37.3) | 15022 | 5.0 (2.8-8.8) | 921 |
| **<$5.50/day** | 52.8 (50.0-55.7) | 13374 | 11.9 (10.1-13.9) | 5743 | 28.2 (25.3-31.4) | 6939 | 5.3 (4.3-6.7) | 6939 | 29.5 (25.2-34.3) | 24256 | 8.1 (5.3-12.3) | 1398 |
| **>$5.50/day** | 59.0 (55.6-62.3) | 17105 | 18.0 (15.4-20.9) | 5086 | 44.9 (41.3-48.6) | 10890 | 13.4 (11.5-15.7) | 10890 | 15.9 (14.0-18.0) | 23808 | 21.2 (8.8-42.8) | 857 |

Table S15: Prevalences by 10-year age groups and poverty level, females only

| Poverty level | Hypertension | | Diabetes | | Overweight | | Obesity | | Smoking | | Dyslipidemia | |
|---|---|---|---|---|---|---|---|---|---|---|---|---|
| | **Prevalence** | **Obs.** | **Prevalence** | **Obs.** | **Prevalence** | **Obs.** | **Prevalence** | **Obs.** | **Prevalence** | **Obs.** | **Prevalence** | **Obs.** |
| ***All age groups*** | | | | | | | | | | | | |
| **Total** | 16.78 (16.3-17.3) | 815382 | 5.2 (4.8-5.6) | 726995 | 30.7 (30.0-31.4) | 1196911 | 10.2 (9.9-10.6) | 1196911 | 2.0 (1.8-2.3) | 579564 | 10.1 (8.0-12.6) | 40634 |
| **<$1.90/day** | 15.4 (14.7-16.2) | 165709 | 3.8 (3.4-4.3) | 160090 | 12.3 (11.8-12.9) | 257784 | 3.1 (2.9-3.4) | 257784 | 1.0 (0.9-1.2) | 130772 | 1.7 (1.1-2.5) | 10310 |
| **<$3.20/day** | 15.3 (14.8-15.9) | 430747 | 4.0 (3.7-4.3) | 415489 | 18.1 (17.6-18.6) | 604235 | 4.9 (4.7-5.1) | 604235 | 1.2 (1.0-1.5) | 244157 | 3.7 (2.7-5.2) | 18499 |
| **<$5.50/day** | 16.0 (15.5-16.6) | 639672 | 4.5 (4.2-4.8) | 606995 | 25.3 (24.7-25.9) | 899858 | 7.8 (7.5-8.1) | 899858 | 1.4 (1.1-1.7) | 367450 | 7.1 (5.5-9.2) | 26940 |
| **>$5.50/day** | 18.4 (17.6-19.2) | 175710 | 6.8 (6.0-7.7) | 120000 | 44.2 (42.8-45.5) | 297053 | 16.3 (15.5-17.1) | 297053 | 3.1 (2.8-3.5) | 212114 | 19.2 (15.0-24.3) | 13694 |
| ***15-24*** | | | | | | | | | | | | |
| **Total** | 4.4 (4.2-4.7) | 239784 | 2.5 (2.3-2.8) | 227985 | 13.6 (13.2-14.1) | 395462 | 3.7 (3.5-3.9) | 395462 | 0.8 (0.7-1.0) | 169254 | 1.3 (0.6-2.7) | 4953 |
| **<$1.90/day** | 4.9 (4.5-5.3) | 52415 | 2.1 (1.9-2.4) | 51701 | 5.9 (5.5-6.3) | 90693 | 1.7 (1.5-1.8) | 90693 | 0.3 (0.1-0.5) | 43833 | 0.5 (0.2-1.2) | 1573 |
| **<$3.20/day** | 4.7 (4.4-4.9) | 141316 | 2.0 (1.9-2.2) | 139228 | 7.8 (7.5-8.1) | 213548 | 1.9 (1.8-2.0) | 213548 | 0.2 (0.2-0.4) | 79938 | 0.8 (0.4-1.7) | 2598 |
| **<$5.50/day** | 4.6 (4.4-4.8) | 203976 | 2.3 (2.1-2.5) | 198985 | 11.0 (10.6-11.4) | 310681 | 2.8 (2.6-3.0) | 310681 | 0.4 (0.3-0.5) | 116640 | 0.8 (0.4-1.4) | 3418 |
| **>$5.50/day** | 4.1 (3.5-4.7) | 35808 | 3.4 (2.7-4.2) | 29000 | 22.8 (21.8-23.8) | 84781 | 6.7 (6.2-7.2) | 84781 | 1.8 (1.3-2.5) | 52614 | 3.3 (1.0-10.6) | 1535 |
| ***25-34*** | | | | | | | | | | | | |
| **Total** | 9.6 (9.3-10.0) | 228802 | 3.3 (3.0-3.6) | 210714 | 32.5 (31.8-33.3) | 350534 | 10.8 (10.4-11.2) | 350534 | 1.2 (1.1-1.4) | 154567 | 4.2 (2.5-6.9) | 11191 |
| **<$1.90/day** | 10.0 (9.4-10.7) | 48731 | 2.6 (2.3-3.0) | 47293 | 11.4 (10.8-12.0) | 77180 | 2.8 (2.5-3.0) | 77180 | 0.6 (0.5-0.8) | 37109 | 0.7 (0.3-1.3) | 3066 |
| **<$3.20/day** | 10.1 (9.7-10.5) | 124117 | 2.5 (2.4-2.8) | 120594 | 19.1 (18.5-19.8) | 177698 | 5.1 (4.8-5.4) | 177698 | 0.6 (0.5-0.8) | 68092 | 2.0 (1.0-3.8) | 5397 |
| **<$5.50/day** | 10.2 (9.9-10.6) | 184242 | 3.0 (2.8-3.3) | 176890 | 28.0 (27.2-28.7) | 266168 | 8.8 (8.4-9.3) | 266168 | 0.7 (0.6-0.9) | 101695 | 3.4 (1.9-5.8) | 7654 |
| **>$5.50/day** | 8.1 (7.2-9.0) | 44560 | 4.1 (3.4-4.9) | 33824 | 45.0 (43.3-46.6) | 84366 | 16.2 (15.3-17.2) | 84366 | 2.3 (2.1-2.6) | 52872 | 7.4 (3.9-13.4) | 3537 |
| ***35-44*** | | | | | | | | | | | | |
| **Total** | 18.3 (17.6-19.0) | 201413 | 5.2 (4.8-5.6) | 182330 | 40.6 (39.5-41.7) | 290234 | 15.0 (14.3-15.6) | 290234 | 2.3 (1.9-2.8) | 123288 | 10.8 (7.7-15.0) | 10089 |
| **<$1.90/day** | 18.2 (17.1-19.4) | 41096 | 3.6 (3.0-4.4) | 40030 | 16.9 (15.7-18.1) | 61955 | 4.2 (3.7-4.7) | 61955 | 1.2 (1.0-1.6) | 27693 | 1.5 (0.5-3.9) | 2347 |

| | | | | | | | | | | | | |
|---|---|---|---|---|---|---|---|---|---|---|---|---|
| **<$3.20/day** | 18.5 (17.7-19.3) | 105615 | 4.2 (3.8-4.7) | 102611 | 25.3 (24.4-26.2) | 145330 | 7.2 (6.8-7.8) | 145330 | 1.4 (1.1-1.8) | 51729 | 2.7 (1.6-4.5) | 4412 |
| **<$5.50/day** | 19.1 (18.4-19.9) | 157617 | 5.0 (4.6-5.4) | 151066 | 34.5 (33.5-35.4) | 217365 | 11.7 (11.2-12.3) | 217365 | 1.6 (1.3-2.1) | 77854 | 7.6 (5.2-11.1) | 6601 |
| **>$5.50/day** | 16.6 (15.2-18.1) | 43796 | 5.6 (4.8-6.6) | 31264 | 54.3 (52.0-56.5) | 72869 | 22.1 (20.7-23.6) | 72869 | 3.3 (2.6-4.2) | 45434 | 21.7 (13.8-32.6) | 3488 |
| ***45-54*** | | | | | | | | | | | | |
| **Total** | 30.0 (28.6-31.4) | 104741 | 8.2 (7.2-9.4) | 87698 | 44.6 (43.1-46.2) | 134051 | 15.5 (14.7-16.4) | 134051 | 4.6 (3.9-5.5) | 73264 | 21.3 (17.2-26.1) | 7810 |
| **<$1.90/day** | 29.4 (26.8-32.2) | 18841 | 5.4 (4.0-7.1) | 17899 | 22.3 (20.1-24.6) | 25796 | 5.7 (4.6-7.0) | 25796 | 3.3 (2.3-4.6) | 13806 | 3.4 (1.8-6.3) | 1744 |
| **<$3.20/day** | 28.9 (27.1-30.7) | 49167 | 6.5 (5.5-7.7) | 46500 | 28.7 (27.2-30.3) | 62346 | 8.4 (7.6-9.2) | 62346 | 3.9 (2.8-5.3) | 26839 | 10.2 (5.7-17.4) | 3258 |
| **<$5.50/day** | 30.1 (28.5-31.8) | 75032 | 7.4 (6.5-8.3) | 69313 | 38.0 (36.5-39.4) | 95016 | 11.9 (11.2-12.7) | 95016 | 4.0 (3.0-5.2) | 42064 | 16.5 (11.9-22.4) | 4954 |
| **>$5.50/day** | 29.8 (27.6-32.1) | 29709 | 9.7 (8.0-11.7) | 18385 | 56.0 (53.1-58.8) | 39035 | 21.6 (20.0-23.4) | 39035 | 5.4 (4.6-6.4) | 31200 | 30.0 (22.3-38.9) | 2856 |
| ***55-64*** | | | | | | | | | | | | |
| **Total** | 42.8 (40.2-45.3) | 24066 | 11.5 (9.8-13.4) | 12507 | 42.0 (39.3-44.9) | 16512 | 10.6 (9.4-11.9) | 16512 | 5.5 (4.3-6.9) | 34005 | 30.1 (24.6-36.3) | 5388 |
| **<$1.90/day** | 39.4 (33.9-45.2) | 3076 | 12.5 (8.8-17.4) | 2348 | 33.6 (26.9-41.1) | 1374 | 7.3 (4.6-11.7) | 1374 | 6.2 (4.1-9.2) | 4696 | 8.3 (5.0-13.6) | 1306 |
| **<$3.20/day** | 44.0 (40.0-48.1) | 6597 | 11.0 (8.8-13.7) | 4661 | 35.5 (31.1-40.0) | 3330 | 6.4 (4.8-8.6) | 3330 | 6.4 (4.4-9.3) | 10140 | 13.7 (9.1-19.9) | 2316 |
| **<$5.50/day** | 42.6 (39.5-45.7) | 11558 | 10.5 (8.8-12.5) | 7567 | 36.9 (33.5-40.4) | 6702 | 7.2 (5.8-8.8) | 6702 | 6.1 (4.3-8.5) | 17006 | 23.2 (17.6-30.0) | 3548 |
| **>$5.50/day** | 42.9 (39.4-46.5) | 12508 | 13.0 (10.4-16.1) | 4940 | 48.2 (44.3-52.1) | 9810 | 14.7 (12.8-16.8) | 9810 | 5.0 (3.9-6.3) | 16999 | 45.6 (34.0-57.7) | 1840 |
| ***>65*** | | | | | | | | | | | | |
| **Total** | 56.3 (53.1-59.4) | 16576 | 13.6 (11.7-15.7) | 5761 | 39.2 (36.2-42.3) | 10118 | 11.1 (9.6-12.8) | 10118 | 6.8 (5.5-8.4) | 25186 | 14.7 (9.0-23.1) | 1203 |
| **<$1.90/day** | 59.3 (52.8-65.6) | 1550 | 11.8 (8.3-16.5) | 819 | 32.0 (26.0-38.6) | 786 | 8.3 (5.3-12.8) | 786 | 8.2 (5.4-12.1) | 3635 | 4.1 (1.4-11.8) | 274 |
| **<$3.20/day** | 56.9 (52.1-61.5) | 3935 | 12.9 (10.2-16.3) | 1895 | 30.9 (26.5-35.6) | 1983 | 7.9 (5.8-10.8) | 1983 | 8.7 (6.3-12.0) | 7419 | 5.3 (2.3-11.9) | 518 |
| **<$5.50/day** | 56.3 (52.5-60.1) | 7247 | 11.8 (9.7-14.4) | 3174 | 32.8 (29.0-36.7) | 3926 | 7.1 (5.5-9.1) | 3926 | 7.8 (6.0-10.1) | 12191 | 11.2 (5.6-21.1) | 765 |
| **>$5.50/day** | 56.3 (52.2-60.3) | 9329 | 16.8 (13.7-20.6) | 2587 | 48.5 (43.9-53.2) | 6192 | 16.9 (14.3-19.9) | 6192 | 5.9 (4.3-7.9) | 12995 | 24.0 (7.7-54.4) | 438 |

Table S16: Prevalences by 10-year age groups and poverty level, males only

| Poverty level | Hypertension | | Diabetes | | Overweight | | Obesity | | Smoking | | Dyslipidemia | |
|---|---|---|---|---|---|---|---|---|---|---|---|---|
| | Prevalence | Obs. | Prevalence | Obs. | Prevalence | Obs. | Prevalence | Obs. | Prevalence | Obs. | Prevalence | Obs. |
| ***All age groups*** | | | | | | | | | | | | |
| **Total** | 23.1 (22.21-23.91) | 255618 | 6.7 (5.9-7.5) | 173991 | 34.1 (31.2-37.1) | 87035 | 7.2 (5.8-8.9) | 87035 | 44.7 (40.1-49.4) | 336931 | 6.6 (4.9-8.9) | 29957 |
| **<$1.90/day** | 22.1 (20.6-23.7) | 39320 | 4.6 (3.9-5.4) | 33464 | 24.1 (19.6-29.2) | 7448 | 2.6 (1.7-3.9) | 7448 | 34.2 (30.3-38.4) | 61274 | 1.0 (0.5-1.9) | 6541 |
| **<$3.20/day** | 21.6 (20.6-22.6) | 102949 | 4.8 (4.2-5.4) | 86925 | 24.7 (21.8-27.9) | 19110 | 3.5 (2.6-4.5) | 19110 | 44.2 (38.7-49.9) | 129142 | 1.5 (1.0-2.4) | 12804 |
| **<$5.50/day** | 22.6 (21.6-23.5) | 164687 | 5.3 (4.8-5.9) | 131807 | 26.6 (24.3-29.2) | 35908 | 4.2 (3.4-5.1) | 35908 | 47.1 (41.4-52.9) | 201288 | 3.0 (2.1-4.3) | 19239 |
| **>$5.50/day** | 23.7 (22.6-25.0) | 90931 | 8.7 (7.3-10.3) | 42184 | 40.1 (36.6-43.6) | 51127 | 9.7 (7.8-12.0) | 51127 | 42.0 (38.3-45.8) | 135643 | 15.2 (10.3-22.0) | 10718 |
| ***15-24*** | | | | | | | | | | | | |
| **Total** | 8.3 (7.5-9.2) | 54312 | 2.3 (1.8-2.9) | 41761 | 19.7 (16.1-23.9) | 12913 | 4.1 (3.0-5.6) | 12913 | 29.2 (24.7-34.2) | 86502 | 0.5 (0.2-1.1) | 3604 |
| **<$1.90/day** | 9.4 (7.3-12.1) | 9589 | 2.0 (1.6-2.7) | 8717 | 17.2 (9.5-29.2) | 1358 | 1.3 (0.7-2.4) | 1358 | 18.4 (14.9-22.5) | 18241 | 0.1 (0.0-0.4) | 1093 |
| **<$3.20/day** | 8.6 (7.4-9.9) | 26431 | 2.5 (1.9-3.4) | 23836 | 15.9 (11.4-21.8) | 3309 | 1.2 (0.8-1.8) | 3309 | 26.3 (21.1-32.3) | 37211 | 0.1 (0.1-0.3) | 1790 |
| **<$5.50/day** | 9.0 (8.0-10.1) | 40546 | 2.5 (1.9-3.2) | 34863 | 15.3 (11.9-19.6) | 6118 | 2.3 (1.4-3.8) | 6118 | 29.5 (24.1-35.6) | 56293 | 0.1 (0.1-0.3) | 2426 |
| **>$5.50/day** | 7.3 (6.0-8.8) | 13766 | 1.9 (1.2-3.0) | 6898 | 23.4 (19.0-28.3) | 6795 | 5.5 (4.1-7.5) | 6795 | 28.8 (24.1-34.0) | 30209 | 2.2 (0.9-5.2) | 1178 |
| ***25-34*** | | | | | | | | | | | | |
| **Total** | 15.5 (14.6-16.5) | 61175 | 3.2 (2.6-3.9) | 44903 | 35.0 (30.8-39.5) | 20779 | 8.4 (6.4-11.1) | 20779 | 45.9 (40.7-51.3) | 78872 | 4.2 (1.4-11.7) | 7894 |
| **<$1.90/day** | 17.4 (15.5-19.5) | 9996 | 2.5 (2.0-3.0) | 8749 | 21.9 (14.8-31.3) | 1911 | 3.6 (1.5-8.3) | 1911 | 36.9 (32.1-42.0) | 15161 | 0.2 (0.1-0.7) | 1912 |
| **<$3.20/day** | 16.5 (15.1-17.9) | 26224 | 2.9 (2.3-3.6) | 22903 | 22.4 (17.8-27.7) | 4712 | 3.6 (2.1-6.0) | 4712 | 47.3 (40.7-54.0) | 31141 | 0.3 (0.1-0.8) | 3670 |
| **<$5.50/day** | 16.4 (15.3-17.7) | 41565 | 2.9 (2.4-3.6) | 34787 | 25.3 (21.4-29.5) | 8547 | 4.6 (3.2-6.6) | 8547 | 49.8 (43.4-56.2) | 48234 | 0.6 (0.3-1.2) | 5330 |
| **>$5.50/day** | 14.2 (12.7-15.8) | 19610 | 3.8 (2.5-5.6) | 10116 | 41.0 (36.2-46.0) | 12232 | 10.8 (8.2-14.1) | 12232 | 41.7 (37.1-46.5) | 30638 | 14.6 (4.7-37.4) | 2564 |
| ***35-44*** | | | | | | | | | | | | |
| **Total** | 22.4 (20.9-24.0) | 57657 | 6.2 (5.3-7.2) | 40190 | 39.4 (35.7-43.3) | 19137 | 8.1 (6.3-10.4) | 19137 | 53.8 (49.1-58.4) | 67398 | 6.9 (4.7-10.1) | 7082 |
| **<$1.90/day** | 20.3 (17.9-22.8) | 8955 | 6.0 (4.2-8.5) | 7857 | 26.6 (17.9-37.7) | 1454 | 1.9 (1.1-3.4) | 1454 | 42.9 (37.6-48.4) | 11574 | 1.3 (0.6-3.0) | 1411 |

| | | | | | | | | | | | | |
|---|---|---|---|---|---|---|---|---|---|---|---|---|
| **<$3.20/day** | 21.5 (19.7-23.4) | 23291 | 6.3 (5.0-8.0) | 20071 | 32.8 (27.3-38.8) | 3984 | 4.2 (2.5-7.0) | 3984 | 54.1 (48.5-59.5) | 25065 | 2.8 (1.4-5.4) | 2933 |
| **<$5.50/day** | 22.4 (20.7-24.2) | 37233 | 5.7 (4.8-6.8) | 30498 | 33.9 (30.0-38.0) | 7555 | 4.9 (3.6-6.7) | 7555 | 56.5 (51.1-61.8) | 39938 | 3.6 (2.0-6.5) | 4510 |
| **>$5.50/day** | 22.4 (20.3-24.8) | 20424 | 6.8 (5.2-8.9) | 9692 | 43.7 (38.8-48.7) | 11582 | 10.5 (8.0-13.7) | 11582 | 50.7 (45.9-55.5) | 27460 | 14.2 (9.5-20.6) | 2572 |
| ***45-54*** | | | | | | | | | | | | |
| **Total** | 32.9 (31.0-34.7) | 48277 | 10.3 (8.6-12.3) | 32079 | 39.1 (35.7-42.6) | 15418 | 8.4 (6.7-10.4) | 15418 | 56.7 (52.1-61.2) | 50631 | 14.3 (10.5-19.2) | 6115 |
| **<$1.90/day** | 35.4 (30.9-40.3) | 7063 | 5.0 (3.4-7.2) | 5948 | 34.0 (25.5-43.6) | 1261 | 3.3 (1.4-7.6) | 1261 | 48.4 (43.2-53.6) | 8268 | 1.9 (0.7-5.2) | 1117 |
| **<$3.20/day** | 31.6 (28.8-34.6) | 18092 | 5.5 (4.3-6.9) | 15094 | 27.2 (22.2-32.7) | 3200 | 3.8 (2.2-6.3) | 3200 | 58.1 (52.6-63.4) | 17787 | 3.3 (1.8-6.0) | 2327 |
| **<$5.50/day** | 32.9 (30.6-35.3) | 29403 | 8.0 (6.7-9.4) | 23305 | 31.0 (27.3-34.9) | 6057 | 5.2 (3.7-7.4) | 6057 | 60.4 (55.0-65.5) | 28286 | 9.1 (4.9-16.3) | 3693 |
| **>$5.50/day** | 32.8 (30.3-35.4) | 18874 | 13.1 (10.4-16.5) | 8774 | 45.7 (41.5-49.8) | 9361 | 10.9 (8.6-13.7) | 9361 | 53.0 (48.3-57.5) | 22345 | 21.7 (14.1-31.9) | 2422 |
| ***55-64*** | | | | | | | | | | | | |
| **Total** | 41.7 (39.3-44.0) | 20294 | 12.0 (10.3-14.0) | 9990 | 34.8 (31.7-37.9) | 11077 | 6.4 (5.2-7.8) | 11077 | 52.0 (47.4-56.5) | 30650 | 13.9 (10.4-18.4) | 4210 |
| **<$1.90/day** | 39.3 (33.3-45.7) | 2458 | 10.7 (6.9-16.2) | 1642 | 22.4 (16.2-30.1) | 914 | 2.8 (1.0-7.4) | 914 | 50.4 (44.5-56.3) | 4577 | 6.6 (2.3-17.4) | 826 |
| **<$3.20/day** | 39.5 (35.6-43.6) | 5625 | 7.7 (5.9-10.1) | 3582 | 23.8 (19.8-28.4) | 2400 | 4.2 (2.5-6.7) | 2400 | 58.3 (52.1-64.2) | 10335 | 5.7 (2.7-11.9) | 1681 |
| **<$5.50/day** | 40.1 (37.0-43.3) | 9813 | 8.5 (7.0-10.4) | 5785 | 25.6 (22.5-29.0) | 4618 | 3.6 (2.5-5.2) | 4618 | 57.8 (52.1-63.2) | 16472 | 10.1 (6.7-14.8) | 2647 |
| **>$5.50/day** | 43.6 (40.4-46.8) | 10481 | 16.8 (14.0-19.9) | 4205 | 44.8 (40.5-49.1) | 6459 | 9.5 (7.7-11.7) | 6459 | 45.7 (41.4-50.0) | 14178 | 20.1 (12.3-30.9) | 1563 |
| ***>65*** | | | | | | | | | | | | |
| **Total** | 54.4 (51.3-57.4) | 13903 | 14.8 (12.6-17.2) | 5068 | 31.0 (27.7-34.5) | 7711 | 6.2 (5.0-7.8) | 7711 | 38.7 (34.1-43.4) | 22878 | 11.3 (5.8-20.8) | 1052 |
| **<$1.90/day** | 50.5 (43.0-58.0) | 1259 | 10.2 (5.4-18.4) | 551 | 19.5 (13.2-27.9) | 550 | 3.2 (1.2-8.0) | 550 | 49.0 (42.4-55.7) | 3453 | 5.3 (1.9-14.4) | 182 |
| **<$3.20/day** | 48.1 (43.0-53.2) | 3286 | 10.5 (7.4-14.7) | 1439 | 21.6 (16.6-27.7) | 1505 | 3.5 (1.9-6.3) | 1505 | 52.9 (47.1-58.6) | 7603 | 4.8 (2.1-10.3) | 403 |
| **<$5.50/day** | 48.8 (45.3-52.4) | 6127 | 12.0 (9.6-14.9) | 2569 | 22.8 (18.9-27.4) | 3013 | 3.3 (2.1-5.1) | 3013 | 49.1 (43.5-54.7) | 12065 | 5.3 (3.1-8.9) | 633 |
| **>$5.50/day** | 61.7 (56.9-66.2) | 7776 | 19.1 (15.6-23.1) | 2499 | 41.3 (36.8-45.9) | 4698 | 9.9 (7.7-12.5) | 4698 | 26.7 (23.1-30.6) | 10813 | 19.9 (7.6-42.9) | 419 |

Figure S5: Prevalences by 10-year age groups and poverty level[1]

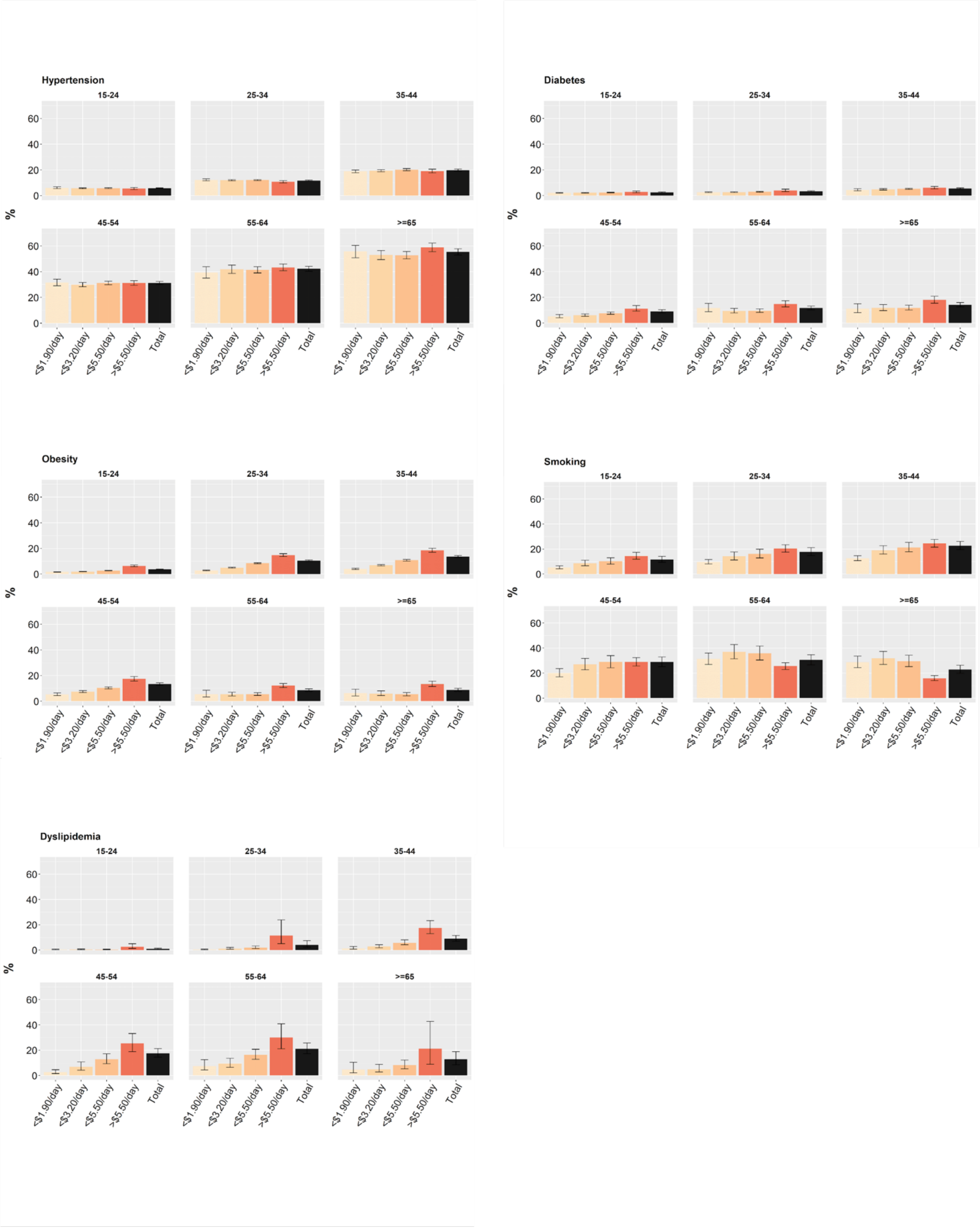

[1]Data are point estimates with 95% CIs.

Figure S6: Prevalences by 10-year age groups and poverty level, disaggregated by sex[1]

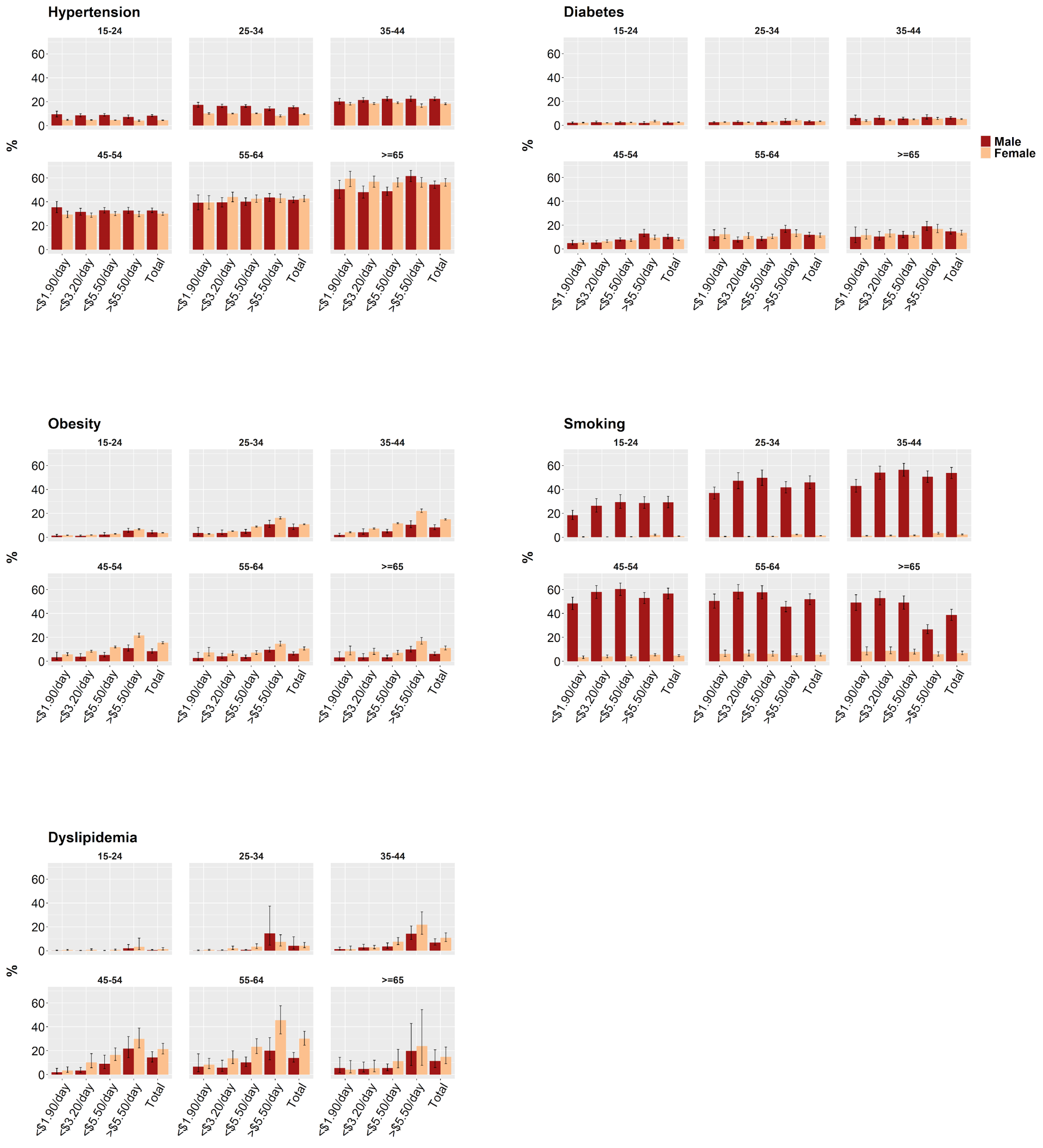

[1]Data are point estimates with 95% CIs.

Table S17: Prevalences by poverty level and region

| Poverty level | Hypertension | | Diabetes | | Overweight | | Obesity | | Smoking | | Dyslipidemia | |
|---|---|---|---|---|---|---|---|---|---|---|---|---|
| | Prevalence | Obs. | Prevalence | Obs. | Prevalence | Obs. | Prevalence | Obs. | Prevalence | Obs. | Prevalence | Obs. |
| *All regions* | | | | | | | | | | | | |
| **Total** | 19.1 (18.6-19.7) | 1071000 | 5.7 (5.2-6.2) | 900986 | 31.4 (30.3-32.3) | 1283946 | 9.6 (9.1-10.1) | 1283946 | 19.2 (16.2-22.2) | 634515 | 8.3 (6.4-10.2) | 70591 |
| **<$1.90/day** | 17.5 (16.7-18.3) | 205029 | 4.0 (3.6-4.5) | 193554 | 13.3 (12.6-14.1) | 265232 | 3.1 (2.8-3.3) | 265232 | 10.6 (9.0-12.3) | 192046 | 1.4 (0.9-1.9) | 16851 |
| **<$3.20/day** | 17.3 (16.7-17.9) | 533696 | 4.2 (3.9-4.5) | 502414 | 18.8 (18.2-19.4) | 623345 | 4.7 (4.5-4.9) | 623345 | 15.9 (12.8-18.9) | 373299 | 2.7 (1.9-3.5) | 31303 |
| **<$5.50/day** | 18.2 (17.6-18.8) | 804359 | 4.8 (4.5-5.1) | 738802 | 25.5 (24.9-26.1) | 935766 | 7.3 (7.0-7.6) | 935766 | 17.9 (14.3-21.4) | 568738 | 5.1 (3.8-6.4) | 46179 |
| **>$5.50/day** | 20.8 (20.0-21.6) | 266641 | 7.6 (6.5-8.7) | 162184 | 42.8 (41.1-44.6) | 348180 | 14.2 (13.2-15.2) | 348180 | 21.2 (18.7-23.7) | #REF! | 17.0 (12.6-21.4) | 24412 |
| *East Asia and Pacific* | | | | | | | | | | | | |
| **Total** | 23.1 (21.9-24.4) | 69852 | 6.0 (5.4-6.7) | 38965 | 28.3 (26.9-29.7) | 85031 | 4.5 (3.9-5.1) | 85031 | 34.9 (29.6-40.6) | 94481 | 35.2 (30.2-40.6) | 21809 |
| **<$1.90/day** | 26.4 (23.2-29.8) | 5131 | 7.0 (5.4-8.9) | 3324 | 25.5 (22.1-29.2) | 6889 | 3.7 (2.7-5.1) | 6889 | 37.2 (29.4-45.7) | 7875 | 18.0 (9.5-31.6) | 1796 |
| **<$3.20/day** | 25.2 (23.4-27.1) | 21070 | 6.3 (5.4-7.4) | 11549 | 25.9 (24.0-27.8) | 28398 | 4.1 (3.4-4.9) | 28398 | 41.7 (34.1-49.8) | 29496 | 26.4 (18.8-35.9) | 5701 |
| **<$5.50/day** | 24.6 (23.2-26.1) | 41876 | 5.7 (5.0-6.5) | 23147 | 27.1 (25.6-28.7) | 54142 | 4.3 (3.8-5.0) | 54142 | 39.8 (32.7-47.4) | 56492 | 29.2 (23.4-35.8) | 11511 |
| **>$5.50/day** | 21.5 (20.0-23.0) | 27976 | 6.4 (5.6-7.2) | 15818 | 29.5 (27.7-31.5) | 30889 | 4.6 (3.9-5.5) | 30889 | 30.1 (26.4-34.1) | 37989 | 42.6 (35.3-50.2) | 10298 |
| *Europe and Central Asia* | | | | | | | | | | | | |
| **Total** | 28.3 (26.5-30.2) | 14545 | 6.1 (5.4-6.9) | 7487 | 53.1 (52.2-54.0) | 50394 | 24.9 (24.1-25.7) | 50394 | 16.5 (15.4-17.7) | 50610 | 18.4 (13.7-24.3) | 3454 |
| **<$1.90/day** | 25.0 (16.9-35.4) | 262 | 11.4 (6.6-18.8) | 217 | 44.4 (35.9-53.2) | 807 | 20.9 (15.2-27.9) | 807 | 19.6 (13.6-27.6) | 913 | 19.6 (4.2-57.6) | 84 |
| **<$3.20/day** | 28.8 (24.2-34.0) | 1603 | 7.0 (5.1-9.5) | 1183 | 47.8 (44.7-50.9) | 5311 | 21.7 (19.1-24.4) | 5311 | 15.9 (13.1-19.3) | 6014 | 8.1 (3.2-18.7) | 663 |
| **<$5.50/day** | 30.9 (27.3-34.7) | 5508 | 4.9 (3.9-6.2) | 3284 | 47.5 (45.6-49.3) | 18745 | 22.1 (20.5-23.7) | 18745 | 13.3 (11.7-15.1) | 21440 | 11.8 (7.0-19.2) | 1981 |
| **>$5.50/day** | 27.2 (25.1-29.3) | 9037 | 6.7 (5.8-7.7) | 4203 | 54.5 (53.5-55.5) | 31649 | 25.6 (24.7-26.5) | 31649 | 18.3 (16.9-19.8) | 29170 | 23.7 (17.0-32.0) | 1473 |
| *Latin America and the Caribbean* | | | | | | | | | | | | |
| **Total** | 22.3 (20.9-23.9) | 97750 | 21.3 (17.5-25.7) | 6684 | 56.5 (53.2-59.7) | 174087 | 21.6 (20.2-23.1) | 174087 | 12.6 (12.2-13.0) | 175997 | 10.7 (9.4-12.2) | 5994 |

| | | | | | | | | | | | | |
|---|---|---|---|---|---|---|---|---|---|---|---|---|
| **<$1.90/day** | 21.1 (17.0-25.9) | 3903 | 22.7 (16.8-29.9) | 286 | 42.9 (35.4-50.7) | 12958 | 13.2 (9.7-17.7) | 12958 | 8.5 (7.3-9.9) | 11276 | 9.2 (5.4-15.4) | 252 |
| **<$3.20/day** | 19.6 (17.2-22.1) | 10254 | 16.7 (11.9-22.9) | 831 | 45.4 (39.1-51.9) | 29398 | 14.5 (11.6-18.1) | 29398 | 9.1 (8.3-10.1) | 25863 | 12.3 (8.7-17.2) | 731 |
| **<$5.50/day** | 20.0 (17.8-22.2) | 24201 | 19.9 (15.5-25.2) | 1999 | 49.2 (44.1-54.2) | 60164 | 17.0 (14.6-19.6) | 60164 | 11.1 (10.4-11.8) | 53892 | 9.6 (7.8-11.6) | 1742 |
| **>$5.50/day** | 23.0 (21.7-24.3) | 73549 | 21.9 (17.7-26.7) | 4685 | 58.8 (56.0-61.5) | 113923 | 23.1 (21.8-24.5) | 113923 | 13.0 (12.6-13.4) | 122105 | 11.2 (10.1-12.5) | 4252 |
| ***Middle East and North Africa*** | | | | | | | | | | | | |
| **Total** | 17.4 (16.9-17.8) | 49061 | 7.3 (6.9-7.7) | 22893 | 64.2 (63.2-65.2) | 81224 | 30.5 (29.5-31.5) | 81224 | 17.0 (16.6-17.4) | 169828 | 1.6 (1.2-2.2) | 3200 |
| **<$1.90/day** | 13.1 (8.7-19.4) | 315 | 8.9 (2.6-26.0) | 58 | 20.8 (17.1-25.0) | 4066 | 8.2 (6.5-10.3) | 4066 | 16.6 (15.1-18.4) | 11750 | - | 3[1] |
| **<$3.20/day** | 13.8 (12.4-15.3) | 3389 | 6.6 (3.9-11.0) | 507 | 54.4 (51.7-57.1) | 15989 | 27.9 (25.9-29.9) | 15989 | 17.1 (16.2-18.0) | 37497 | 4.0 (0.8-17.8) | 71 |
| **<$5.50/day** | 15.7 (14.9-16.5) | 14104 | 6.1 (5.0-7.4) | 3378 | 66.5 (64.8-68.2) | 37141 | 35.5 (33.9-37.1) | 37141 | 18.1 (17.5-18.8) | 77007 | 2.7 (1.4-5.3) | 631 |
| **>$5.50/day** | 18.5 (18.0-19.0) | 34957 | 7.4 (7.0-7.9) | 19515 | 62.1 (61.1-63.0) | 44083 | 26.0 (25.2-26.8) | 44083 | 16.3 (15.8-16.8) | 92821 | 1.6 (1.1-2.2) | 2569 |
| ***South Asia*** | | | | | | | | | | | | |
| **Total** | 13.4 (13.2-13.6) | 757750 | 5.0 (4.8-5.1) | 764537 | 6.8 (6.5-7.1) | 662316 | 7.1 (6.7-7.5) | 662316 | 7.2 (6.7-7.8) | 94297 | 1.4 (0.9-2.0) | 2690 |
| **<$1.90/day** | 10.8 (10.5-11.0) | 164482 | 2.7 (2.6-2.9) | 165361 | 13.8 (13.4-14.2) | 144423 | 1.0 (0.9-1.1) | 144423 | 13.3 (11.7-15.1) | 16453 | 3.6 (0.8-15.2) | 50 |
| **<$3.20/day** | 12.0 (11.8-12.2) | 446572 | 3.3 (3.2-3.4) | 449179 | 21.1 (20.6-21.7) | 390675 | 2.9 (2.7-3.1) | 390675 | 9.9 (9.1-10.8) | 50234 | 1.5 (0.7-3.4) | 370 |
| **<$5.50/day** | 13.0 (12.8-13.2) | 652834 | 4.3 (4.2-4.4) | 657588 | 44.0 (42.4-45.6) | 570903 | 5.5 (5.2-5.8) | 570903 | 8.0 (7.4-8.7) | 77927 | 1.4 (0.8-2.3) | 1135 |
| **>$5.50/day** | 15.6 (15.2-16.1) | 104916 | 8.7 (8.3-9.2) | 106949 | 24.7 (24.1-25.4) | 91413 | 15.5 (14.3-16.8) | 91413 | 3.7 (2.9-4.8) | 16370 | 1.4 (0.8-2.3) | 1555 |
| ***Sub-Saharan Africa*** | | | | | | | | | | | | |
| **Total** | 22.2 (21.5-23.0) | 82042 | 3.3 (3.0-3.5) | 60420 | 25.4 (24.8-26.0) | 230894 | 9.9 (9.6-10.2) | 230894 | 2.9 (2.8-3.1) | 331282 | 1.6 (1.1-2.3) | 33444 |
| **<$1.90/day** | 22.0 (21.0-23.1) | 30936 | 3.7 (3.3-4.1) | 24308 | 13.2 (12.6-13.7) | 96089 | 4.0 (3.8-4.3) | 96089 | 4.3 (4.1-4.7) | 49302 | 0.8 (0.6-1.2) | 14666 |
| **<$3.20/day** | 21.3 (20.5-22.1) | 50808 | 3.3 (3.0-3.7) | 39165 | 17.1 (16.6-17.6) | 153574 | 5.5 (5.3-5.8) | 153574 | 3.6 (3.4-3.9) | 143779 | 1.1 (0.8-1.6) | 23767 |
| **<$5.50/day** | 21.7 (20.9-22.5) | 65836 | 3.1 (2.9-3.4) | 49406 | 21.6 (21.0-22.1) | 194671 | 7.6 (7.3-7.9) | 194671 | 3.2 (3.0-3.4) | 224195 | 1.4 (0.9-2.2) | 29179 |
| **>$5.50/day** | 25.3 (23.6-27.0) | 16206 | 4.1 (3.5-4.8) | 11014 | 50.9 (49.7-52.2) | 36223 | 25.2 (24.1-26.3) | 36223 | 1.3 (1.1-1.5) | 281980 | 2.9 (1.7-4.8) | 4265 |

[1] No prevalence calculated when sample size was <50.

Table S18: Prevalences by poverty level and region, females only

| Poverty level | Hypertension | | Diabetes | | Overweight | | Obesity | | Smoking | | Dyslipidemia | |
|---|---|---|---|---|---|---|---|---|---|---|---|---|
| | Prevalence | Obs. | Prevalence | Obs. | Prevalence | Obs. | Prevalence | Obs. | Prevalence | Obs. | Prevalence | Obs. |
| ***All regions*** | | | | | | | | | | | | |
| **Total** | 16.8 (16.3-17.3) | 815382 | 5.2 (4.8-5.6) | 726995 | 30.7 (30.0-31.4) | 1196911 | 10.2 (9.9-10.6) | 1196911 | 2.0 (1.8-2.3) | 579564 | 10.1 (8.1-12.6) | 40634 |
| **<$1.90/day** | 15.4 (14.7-16.2) | 165709 | 3.8 (3.4-4.3) | 160090 | 12.3 (11.8-12.9) | 257784 | 3.1 (2.9-3.4) | 257784 | 1.0 (0.9-1.2) | 130772 | 1.7 (1.1-2.5) | 10310 |
| **<$3.20/day** | 15.3 (14.8-15.9) | 430747 | 4.0 (3.7-4.3) | 415489 | 18.1 (17.6-18.6) | 604235 | 4.9 (4.7-5.1) | 604235 | 1.2 (1.0-1.5) | 244157 | 3.7 (2.7-5.2) | 18499 |
| **<$5.50/day** | 16.0 (15.5-16.6) | 639672 | 4.5 (4.2-4.8) | 606995 | 25.3 (24.7-25.9) | 899858 | 7.8 (7.5-8.1) | 899858 | 1.4 (1.1-1.7) | 367450 | 7.1 (5.6-9.2) | 26940 |
| **>$5.50/day** | 18.4 (17.6-19.2) | 175710 | 6.8 (6.0-7.7) | 120000 | 44.2 (42.8-45.5) | 297053 | 16.3 (15.5-17.1) | 297053 | 3.1 (2.8-3.5) | 212114 | 19.2 (15.1-24.3) | 13694 |
| ***East Asia and Pacific*** | | | | | | | | | | | | |
| **Total** | 21.1 (19.9-22.4) | 38256 | 5.3 (4.6-6.1) | 21872 | 28.9 (26.9-30.9) | 56935 | 5.1 (4.3-6.1) | 56935 | 2.8 (2.1-3.7) | 54403 | 41.4 (34.6-48.5) | 12543 |
| **<$1.90/day** | 26.1 (22.4-30.1) | 2883 | 7.3 (5.3-9.8) | 1938 | 27.3 (23.7-31.4) | 4933 | 5.3 (3.8-7.5) | 4933 | 4.6 (2.9-7.1) | 4594 | 22.2 (10.8-40.3) | 1054 |
| **<$3.20/day** | 24.5 (22.6-26.5) | 11512 | 6.5 (5.3-7.9) | 6587 | 28.0 (25.8-30.2) | 19964 | 5.4 (4.4-6.6) | 19964 | 4.1 (2.8-6.0) | 17052 | 31.9 (23.8-41.2) | 3256 |
| **<$5.50/day** | 23.1 (21.5-24.8) | 22754 | 5.5 (4.6-6.5) | 13055 | 29.0 (27.0-31.1) | 37390 | 5.4 (4.5-6.5) | 37390 | 3.7 (2.6-5.3) | 32960 | 37.7 (31.0-44.8) | 6572 |
| **>$5.50/day** | 18.9 (17.4-20.5) | 15502 | 5.1 (4.3-6.1) | 8817 | 28.7 (26.2-31.2) | 19545 | 4.8 (3.8-6.0) | 19545 | 2.0 (1.5-2.7) | 21443 | 46.9 (37.1-56.9) | 5971 |
| ***Europe and Central Asia*** | | | | | | | | | | | | |
| **Total** | 22.0 (20.4-23.8) | 8563 | 5.7 (4.8-6.7) | 4613 | 52.0 (51.0-52.9) | 48695 | 24.5 (23.6-25.3) | 48695 | 8.1 (7.1-9.2) | 40306 | 22.0 (15.7-29.9) | 2048 |
| **<$1.90/day** | 15.1 (8.3-26.0) | 148 | 16.5 (8.3-30.2) | 124 | 39.8 (30.5-49.8) | 747 | 20.8 (14.5-28.8) | 747 | 12.3 (6.4-22.5) | 714 | - | 41[1] |
| **<$3.20/day** | 25.4 (20.1-31.6) | 1016 | 6.6 (4.1-10.4) | 770 | 46.6 (43.4-49.9) | 5116 | 22.0 (19.3-25.0) | 5116 | 6.8 (4.5-10.1) | 4853 | 9.3 (4.2-19.3) | 419 |
| **<$5.50/day** | 24.9 (21.9-28.1) | 3489 | 4.9 (3.6-6.5) | 2166 | 46.8 (44.9-48.7) | 18291 | 22.2 (20.5-23.9) | 18291 | 5.8 (4.5-7.4) | 17236 | 15.9 (9.0-26.3) | 1242 |
| **>$5.50/day** | 20.7 (18.9-22.7) | 5074 | 6.2 (5.0-7.5) | 2447 | 53.2 (52.1-54.2) | 30404 | 25.0 (24.1-26.0) | 30404 | 9.6 (8.3-11.1) | 23070 | 27.7 (19.6-37.7) | 806 |
| ***Latin America and the Caribbean*** | | | | | | | | | | | | |
| **Total** | 20.8 (19.2-22.5) | 52374 | 22.0 (17.3-27.6) | 3083 | 54.8 (52.2-57.5) | 143233 | 22.6 (21.7-23.5) | 143233 | 7.6 (7.2-8.0) | 117985 | 12.6 (9.7-16.3) | 2916 |

| | | | | | | | | | | | | |
|---|---|---|---|---|---|---|---|---|---|---|---|---|
| **<$1.90/day** | 18.0 (13.4-23.8) | 1779 | 25.4 (11.5-47.3) | 137 | 38.5 (33.0-44.4) | 11664 | 13.7 (9.8-18.6) | 11664 | 3.5 (2.6-4.7) | 8882 | 7.2 (4.1-12.3) | 129 |
| **<$3.20/day** | 16.9 (14.7-19.4) | 5033 | 18.3 (12.0-26.8) | 390 | 42.9 (37.1-48.8) | 26196 | 14.2 (11.5-17.5) | 26196 | 3.4 (2.8-4.1) | 19586 | 17.2 (8.3-32.2) | 366 |
| **<$5.50/day** | 19.0 (17.0-21.1) | 12285 | 21.1 (17.0-25.9) | 929 | 48.8 (43.5-54.2) | 52585 | 17.9 (16.0-19.9) | 52585 | 5.1 (4.6-5.7) | 39220 | 12.3 (8.9-16.9) | 859 |
| **>$5.50/day** | 21.3 (19.8-22.9) | 40089 | 22.5 (16.9-29.2) | 2154 | 57.0 (55.1-58.9) | 90648 | 24.3 (23.6-25.1) | 90648 | 8.3 (7.8-8.7) | 78765 | 12.8 (10.1-16.1) | 2057 |
| ***Middle East and North Africa*** | | | | | | | | | | | | |
| **Total** | 17.7 (17.1-18.3) | 25987 | 7.7 (7.1-8.3) | 12620 | 67.1 (66.0-68.3) | 67144 | 34.3 (33.2-35.4) | 67144 | 3.5 (3.2-3.8) | 84374 | 2.2 (1.6-3.1) | 2036 |
| **<$1.90/day** | 16.2 (10.9-23.5) | 163 | - | 28[1] | 20.3 (16.6-24.6) | 4034 | 8.2 (6.5-10.4) | 4034 | 8.6 (6.8-10.7) | 5662 | - | 2[1] |
| **<$3.20/day** | 14.5 (12.8-16.4) | 1810 | 4.9 (2.8-8.4) | 297 | 54.5 (51.7-57.2) | 15722 | 28.4 (26.4-30.4) | 15722 | 4.7 (3.9-5.6) | 18264 | - | 48[1] |
| **<$5.50/day** | 16.0 (15.0-17.1) | 7719 | 6.6 (5.2-8.3) | 2012 | 68.0 (66.3-69.7) | 35493 | 36.8 (35.2-38.5) | 35493 | 3.4 (3.0-3.9) | 38161 | 4.4 (2.2-8.6) | 438 |
| **>$5.50/day** | 18.8 (18.2-19.5) | 18268 | 7.8 (7.2-8.5) | 10608 | 66.0 (64.8-67.2) | 31651 | 31.0 (30.0-32.1) | 31651 | 3.5 (3.2-3.8) | 46213 | 2.0 (1.4-3.0) | 1598 |
| ***South Asia*** | | | | | | | | | | | | |
| **Total** | 12.4 (12.3-12.6) | 644245 | 4.8 (4.6-4.9) | 650202 | 24.7 (24.1-25.4) | 661249 | 7.1 (6.7-7.5) | 661249 | 0.3 (0.2-0.3) | 53381 | 1.9 (1.2-3.0) | 1644 |
| **<$1.90/day** | 10.4 (10.1-10.7) | 142687 | 2.7 (2.5-2.9) | 143468 | 6.8 (6.5-7.1) | 144409 | 1.0 (0.9-1.1) | 144409 | 1.4 (1.0-1.8) | 8888 | 6- | 37[1] |
| **<$3.20/day** | 11.3 (11.2-11.5) | 382620 | 3.2 (3.1-3.3) | 384907 | 13.8 (13.4-14.2) | 390568 | 2.9 (2.7-3.1) | 390568 | 0.6 (0.5-0.7) | 27408 | 1.6 (0.6-4.1) | 265 |
| **<$5.50/day** | 12.1 (11.9-12.3) | 556476 | 4.1 (4.0-4.2) | 560606 | 21.1 (20.6-21.7) | 570480 | 5.5 (5.2-5.8) | 570480 | 0.3 (0.3-0.4) | 43438 | 1.5 (0.8-2.8) | 719 |
| **>$5.50/day** | 14.3 (13.8-14.8) | 87769 | 8.4 (8.0-8.8) | 89596 | 44.0 (42.4-45.6) | 90769 | 15.5 (14.3-16.8) | 90769 | 0.1 (0.1-0.2) | 9943 | 2.2 (1.2-3.9) | 925 |
| ***Sub-Saharan Africa*** | | | | | | | | | | | | |
| **Total** | 21.7 (20.9-22.6) | 45957 | 3.3 (3.0-3.6) | 34605 | 25.6 (25.0-26.1) | 219655 | 10.0 (9.7-10.4) | 219655 | 0.3 (0.2-0.4) | 229115 | 1.9 (1.2-3.1) | 19447 |
| **<$1.90/day** | 21.0 (19.8-22.3) | 18049 | 3.7 (3.2-4.2) | 14395 | 13.1 (12.6-13.7) | 91997 | 4.0 (3.8-4.3) | 91997 | 0.4 (0.3-0.6) | 102032 | 1.0 (0.7-1.6) | 9047 |
| **<$3.20/day** | 20.6 (19.7-21.6) | 28756 | 3.3 (2.9-3.6) | 22538 | 17.1 (16.6-17.6) | 146669 | 5.6 (5.3-5.8) | 146669 | 0.4 (0.3-0.5) | 156994 | 1.6 (1.0-2.5) | 14145 |
| **<$5.50/day** | 21.3 (20.4-22.2) | 36949 | 3.1 (2.8-3.5) | 28227 | 21.7 (21.1-22.2) | 185619 | 7.7 (7.4-8.0) | 185619 | 0.3 (0.2-0.4) | 196435 | 1.8 (1.1-3.2) | 17110 |
| **>$5.50/day** | 24.3 (22.3-26.4) | 9008 | 4.6 (3.7-5.5) | 6378 | 51.5 (50.2-52.8) | 34036 | 25.7 (24.6-26.8) | 34036 | 0.1 (0.1-0.2) | 32680 | 2.6 (1.3-5.0) | 2337 |

[1] No prevalence calculated when sample size was <50.

Table S19: Prevalences by poverty level and region, males only

| Poverty level | Hypertension | | Diabetes | | Overweight | | Obesity | | Smoking | | Dyslipidemia | |
|---|---|---|---|---|---|---|---|---|---|---|---|---|
| | Prevalence | Obs. | Prevalence | Obs. | Prevalence | Obs. | Prevalence | Obs. | Prevalence | Obs. | Prevalence | Obs. |
| ***All regions*** | | | | | | | | | | | | |
| **Total** | 23.1 (22.2-23.9) | 255618 | 6.7 (5.9-7.5) | 173991 | 34.1 (31.2-37.1) | 87035 | 7.2 (5.8-8.9) | 87035 | 44.7 (40.1-49.4) | 336931 | 6.6 (4.9-8.9) | 29957 |
| **<$1.90/day** | 22.1 (20.6-23.7) | 39320 | 4.6 (3.9-5.4) | 33464 | 24.1 (19.6-29.2) | 7448 | 2.6 (1.7-3.9) | 7448 | 34.2 (30.3-38.4) | 61274 | 1.0 (0.6-2.0) | 14919 |
| **<$3.20/day** | 21.6 (20.6-22.6) | 102949 | 4.8 (4.2-5.4) | 86925 | 24.7 (21.8-27.9) | 19110 | 3.5 (2.6-4.5) | 19110 | 44.2 (38.7-49.9) | 129142 | 1.5 (1.0-2.4) | 17179 |
| **<$5.50/day** | 22.6 (21.6-23.6) | 164687 | 5.3 (4.8-5.9) | 131807 | 26.6 (24.3-29.2) | 35908 | 4.2 (3.4-5.1) | 35908 | 47.1 (41.4-52.9) | 201288 | 3.0 (2.1-4.3) | 19239 |
| **>$5.50/day** | 23.8 (22.6-25.0) | 90931 | 8.7 (7.3-10.3) | 42184 | 40.1 (36.6-43.6) | 51127 | 9.7 (7.8-12.0) | 51127 | 42.0 (38.3-45.8) | 135643 | 15.2 (10.3-22.0) | 10718 |
| ***East Asia and Pacific*** | | | | | | | | | | | | |
| **Total** | 25.3 (23.7-26.9) | 31596 | 6.8 (6.1-7.6) | 17093 | 27.6 (25.8-29.5) | 28096 | 3.8 (3.2-4.4) | 28096 | 59.8 (54.2-65.1) | 40078 | 28.0 (20.3-37.3) | 9266 |
| **<$1.90/day** | 26.8 (22.7-31.4) | 2248 | 6.6 (4.6-9.4) | 1386 | 23.1 (18.0-29.2) | 1956 | 1.6 (0.8-3.3) | 1956 | 61.3 (50.9-70.7) | 3281 | 8.0 (2.7-21.2) | 742 |
| **<$3.20/day** | 26.0 (23.6-28.6) | 9558 | 6.1 (4.9-7.6) | 4962 | 23.4 (20.4-26.7) | 8434 | 2.5 (1.8-3.5) | 8434 | 67.2 (59.6-73.9) | 12444 | 17.7 (9.1-31.7) | 2445 |
| **<$5.50/day** | 26.3 (24.3-28.4) | 19122 | 6.0 (5.1-7.0) | 10092 | 24.9 (22.7-27.3) | 16752 | 3.1 (2.5-3.9) | 16752 | 66.0 (59.2-72.1) | 23532 | 17.3 (11.4-25.5) | 4939 |
| **>$5.50/day** | 24.1 (22.2-26.2) | 12474 | 7.7 (6.6-8.9) | 7001 | 30.5 (28.2-32.8) | 11344 | 4.5 (3.7-5.4) | 11344 | 53.5 (48.9-57.9) | 16546 | 38.5 (23.8-55.7) | 4327 |
| ***Europe and Central Asia*** | | | | | | | | | | | | |
| **Total** | 34.1 (31.3-37.1) | 5982 | 6.4 (5.4-7.6) | 2874 | 67.4 (64.4-70.3) | 1699 | 30.3 (27.6-33.1) | 1699 | 37.4 (35.0-40.0) | 10304 | 15.9 (9.9-24.4) | 1406 |
| **<$1.90/day** | 33.0 (20.6-48.3) | 114 | 7.3 (3.1-16.4) | 93 | 57.4 (39.5-73.6) | 60 | 21.1 (10.3-38.4) | 60 | 31.7 (20.0-46.4) | 199 | - | 43[1] |
| **<$3.20/day** | 31.9 (24.9-40.0) | 587 | 7.4 (4.8-11.2) | 413 | 55.0 (45.1-64.5) | 195 | 19.5 (13.0-28.2) | 195 | 41.0 (33.6-48.7) | 1161 | 7.1 (1.3-30.6) | 244 |
| **<$5.50/day** | 36.6 (30.5-43.2) | 2019 | 5.0 (3.7-6.8) | 1118 | 54.1 (47.4-60.6) | 454 | 21.3 (16.6-26.9) | 454 | 40.4 (36.0-44.9) | 4204 | 8.5 (3.1-21.1) | 739 |
| **>$5.50/day** | 33.1 (29.8-36.5) | 3963 | 7.2 (5.9-8.8) | 1756 | 71.3 (67.9-74.4) | 1245 | 32.9 (29.8-36.1) | 1245 | 36.3 (33.4-39.4) | 6100 | 21.2 (12.7-33.4) | 667 |
| ***Latin America and the Caribbean*** | | | | | | | | | | | | |
| **Total** | 23.7 (22.1-25.4) | 45376 | 20.9 (17.5-24.8) | 3601 | 59.1 (55.4-62.7) | 30854 | 20.1 (17.2-23.3) | 30854 | 19.2 (18.4-19.9) | 58012 | 9.4 (8.4-10.5) | 3078 |
| **<$1.90/day** | 23.3 (17.4-30.4) | 2124 | 20.6 (16.7-25.0) | 149 | 54.1 (44.7-63.2) | 1294 | 11.9 (8.0-17.2) | 1294 | 19.4 (16.3-22.8) | 2394 | 11.2 (3.5-30.4) | 123 |

| | | | | | | | | | | | | |
|---|---|---|---|---|---|---|---|---|---|---|---|---|
| **<$3.20/day** | 21.8 (18.2-25.8) | 5221 | 15.5 (11.7-20.2) | 441 | 51.7 (45.5-57.9) | 3202 | 15.2 (11.5-19.8) | 3202 | 20.2 (18.1-22.4) | 6277 | 7.5 (3.7-14.6) | 365 |
| **<$5.50/day** | 20.9 (18.3-23.7) | 11916 | 18.9 (13.9-25.0) | 1070 | 49.9 (45.2-54.5) | 7579 | 14.9 (10.9-19.9) | 7579 | 20.8 (19.4-22.3) | 14672 | 6.7 (4.7-9.2) | 883 |
| **>$5.50/day** | 24.4 (23.0-25.9) | 33460 | 21.6 (18.1-25.5) | 2531 | 61.3 (57.8-64.7) | 23275 | 21.4 (18.5-24.5) | 23275 | 18.8 (18.0-19.6) | 43340 | 10.3 (9.0-11.7) | 2195 |
| ***Middle East and North Africa*** | | | | | | | | | | | | |
| **Total** | 17.1 (16.5-17.7) | 23074 | 6.8 (6.2-7.4) | 10273 | 52.0 (51.0-53.1) | 14080 | 14.8 (14.1-15.5) | 14080 | 31.3 (30.6-32.0) | 85454 | 0.9 (0.5-1.9) | 1164 |
| **<$1.90/day** | 9.6 (4.7-18.7) | 152 | - | 30[1] | - | 32[1] | - | 32[1] | 24.6 (22.6-26.7) | 6088 | - | 1[1] |
| **<$3.20/day** | 13.0 (11.1-15.3) | 1579 | 8.9 (4.2-18.1) | 210 | 50.6 (43.2-57.9) | 267 | 9.1 (5.7-14.2) | 267 | 29.8 (28.2-31.4) | 19233 | - | 23[1] |
| **<$5.50/day** | 15.3 (14.2-16.4) | 6385 | 5.5 (4.0-7.6) | 1366 | 36.6 (33.7-39.6) | 1648 | 8.3 (6.9-10.0) | 1648 | 34.3 (33.0-35.6) | 38846 | 0.1 (0.0-0.7) | 193 |
| **>$5.50/day** | 18.2 (17.5-18.9) | 16689 | 7.0 (6.3-7.6) | 8907 | 54.0 (53.0-55.1) | 12432 | 15.7 (14.9-16.4) | 12432 | 29.7 (28.9-30.5) | 46608 | 1.0 (0.5-2.0) | 971 |
| ***South Asia*** | | | | | | | | | | | | |
| **Total** | 17.9 (17.5-18.4) | 113505 | 6.0 (5.7-6.2) | 114335 | 26.3 (22.2-30.8) | 1067 | 4.3 (3.0-6.3) | 1067 | 27.2 (25.9-28.5) | 40916 | 1.0 (0.5-1.9) | 1046 |
| **<$1.90/day** | 13.1 (12.4-13.9) | 21795 | 3.0 (2.7-3.3) | 21893 | - | 14[1] | - | 14[1] | 33.4 (30.8-36.2) | 7565 | - | 13[1] |
| **<$3.20/day** | 15.3 (14.8-15.8) | 63952 | 4.0 (3.7-4.3) | 64272 | 13.8 (7.4-24.4) | 107 | 3.2 (0.8-12.4) | 107 | 30.8 (29.2-32.5) | 22826 | 1.4 (0.3-6.3) | 105 |
| **<$5.50/day** | 17.2 (16.8-17.7) | 96358 | 5.1 (4.9-5.4) | 96982 | 19.0 (14.1-25.1) | 423 | 2.3 (1.1-4.9) | 423 | 28.2 (26.8-29.5) | 34489 | 1.2 (0.6-2.8) | 416 |
| **>$5.50/day** | 21.7 (20.4-23.0) | 17147 | 10.3 (9.4-11.2) | 17353 | 30.2 (25.5-35.5) | 644 | 5.4 (3.7-7.8) | 644 | 20.3 (16.7-24.5) | 6427 | 0.8 (0.3-2.2) | 630 |
| ***Sub-Saharan Africa*** | | | | | | | | | | | | |
| **Total** | 22.7 (21.6-23.7) | 36085 | 3.2 (2.9-3.6) | 25815 | 19.2 (17.5-21.0) | 11239 | 5.0 (4.2-5.9) | 11239 | 13.8 (13.2-14.6) | 102167 | 1.3 (0.7-2.2) | 13997 |
| **<$1.90/day** | 23.2 (21.8-24.6) | 12887 | 3.6 (3.1-4.2) | 9913 | 15.1 (13.1-17.4) | 4092 | 4.0 (3.1-5.2) | 4092 | 19.0 (17.8-20.3) | 41747 | 0.6 (0.3-1.3) | 13997 |
| **<$3.20/day** | 22.0 (20.8-23.1) | 22052 | 3.4 (3.0-3.9) | 16627 | 16.7 (14.9-18.7) | 6905 | 4.6 (3.7-5.6) | 6905 | 16.5 (15.6-17.5) | 67201 | 0.7 (0.4-1.1) | 13997 |
| **<$5.50/day** | 22.0 (21.0-23.2) | 28887 | 3.2 (2.8-3.6) | 21179 | 17.7 (15.9-19.6) | 9052 | 4.8 (4.0-5.8) | 9052 | 15.2 (14.5-16.1) | 85545 | 1.1 (0.5-2.2) | 12069 |
| **>$5.50/day** | 26.3 (23.9-28.9) | 7198 | 3.6 (2.9-4.7) | 4636 | 27.7 (23.6-32.2) | 2187 | 5.8 (4.2-8.1) | 2187 | 5.6 (4.9-6.5) | 16622 | 3.1 (1.6-5.9) | 1928 |

[1] No prevalence calculated when sample size was <50.

Figure S7: Prevalences by poverty level and region[1,2]

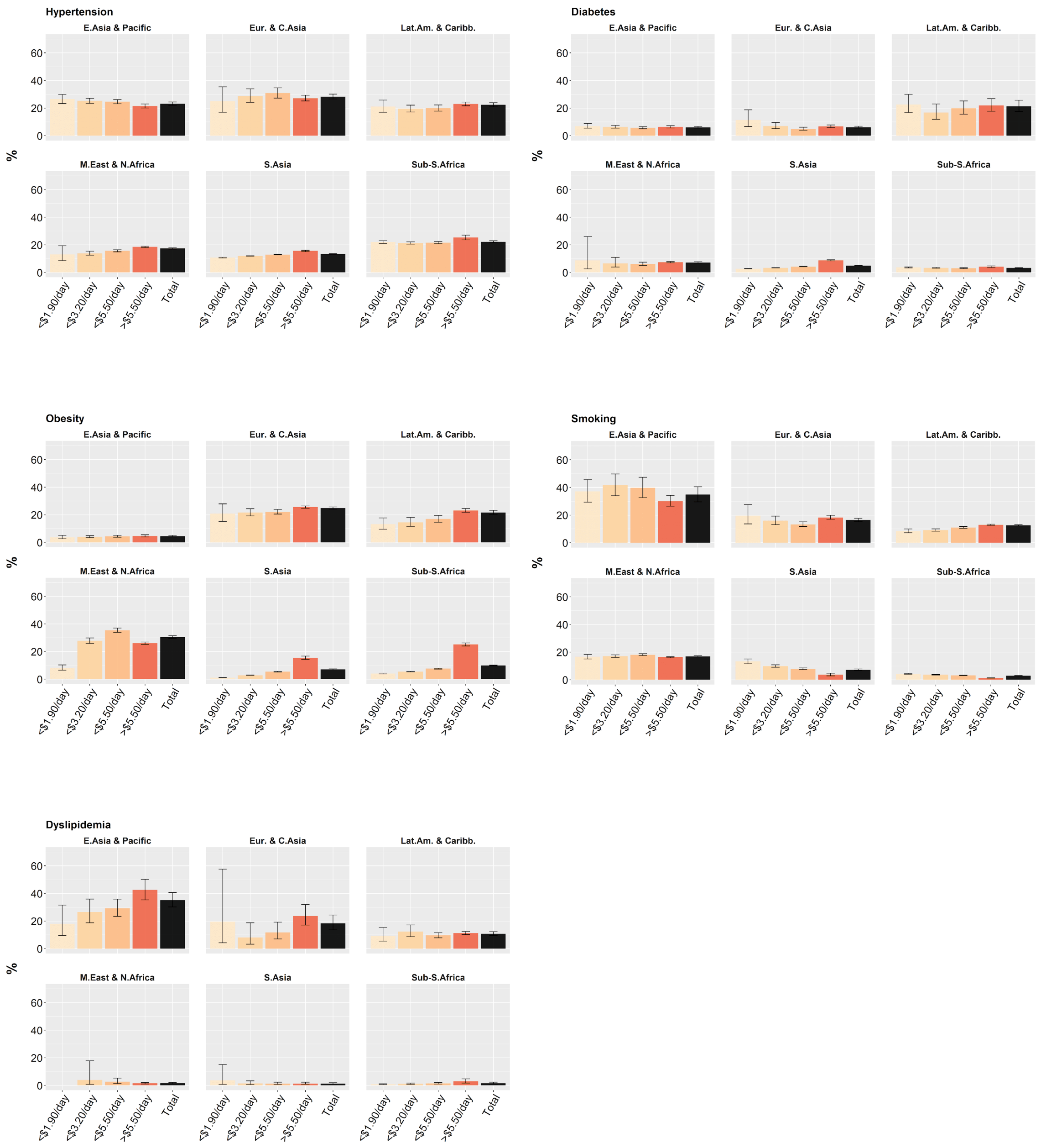

[1]No prevalence calculated when sample size was <50.
[2]Data are point estimates with 95% CIs.

Figure S8: Prevalences by poverty level and region, disaggregated by sex[1,2]

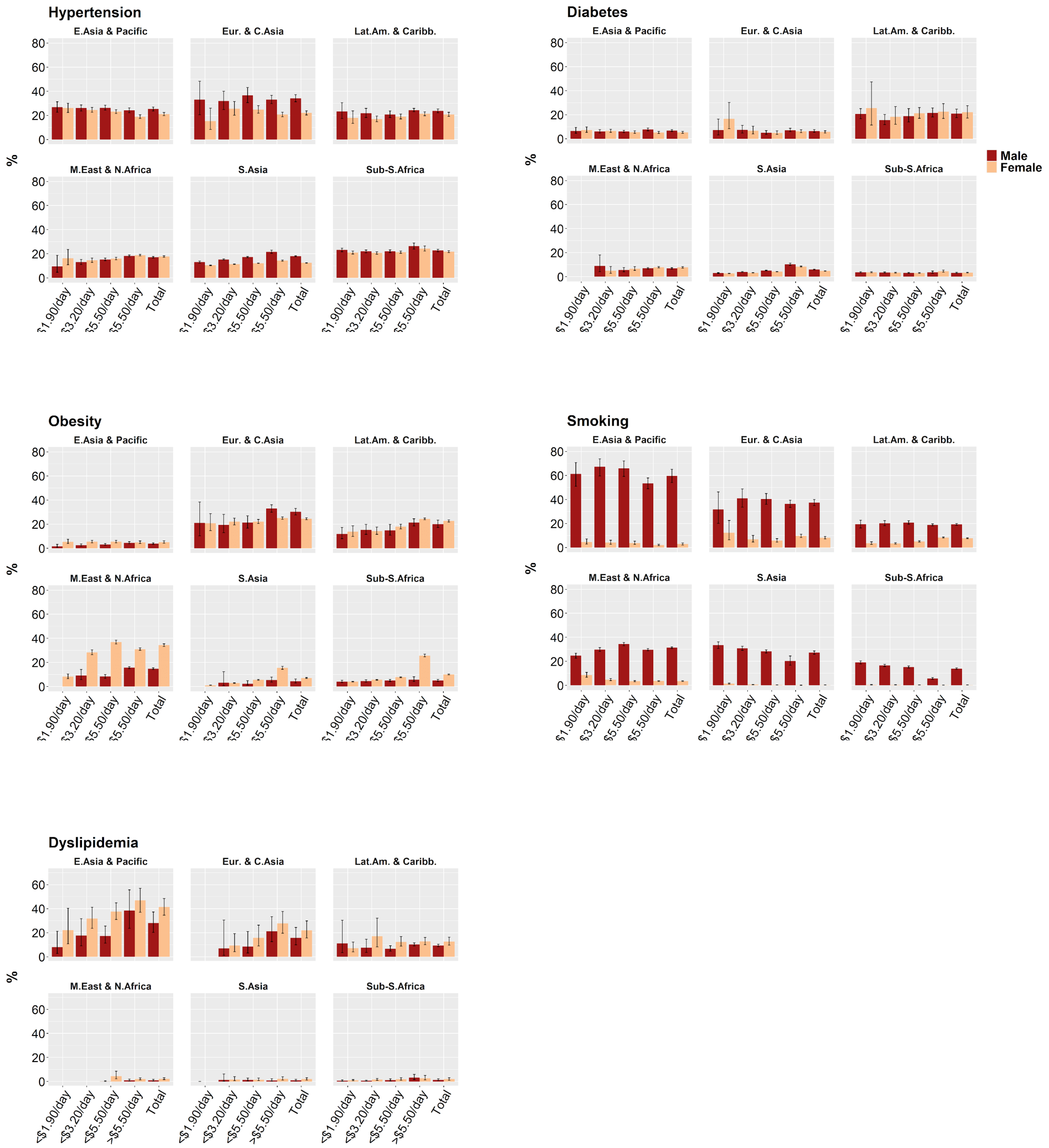

[1]No prevalence calculated when sample size was <50.
[2]Data are point estimates with 95% CIs.

Table S20: Prevalences by poverty level and World Bank country classification

| Poverty level | Hypertension | | Diabetes | | Overweight | | Obesity | | Smoking | | Dyslipidemia | |
|---|---|---|---|---|---|---|---|---|---|---|---|---|
| | Prevalence | Obs. | Prevalence | Obs. | Prevalence | Obs. | Prevalence | Obs. | Prevalence | Obs. | Prevalence | Obs. |
| ***All poverty levels*** | | | | | | | | | | | | |
| **Total** | 19.1 (18.6-19.7) | 1071000 | 5.7 (5.2-6.2) | 900986 | 31.4 (30.3-32.3) | 1283946 | 9.6 (9.1-10.1) | 1283946 | 19.2 (16.2-22.2) | 910915 | 8.3 (6.4-10.2) | 70591 |
| **<$1.90/day** | 17.5 (16.7-18.3) | 205029 | 4.0 (3.6-4.5) | 193554 | 13.3 (12.6-14.1) | 265232 | 3.1 (2.8-3.3) | 265232 | 10.6 (9.0-12.3) | 192036 | 1.4 (0.9-1.9) | 16851 |
| **<$3.20/day** | 17.3 (16.7-17.9) | 533696 | 4.2 (3.9-4.5) | 502414 | 18.8 (18.2-19.4) | 623345 | 4.7 (4.5-4.9) | 623345 | 15.9 (12.8-18.9) | 373200 | 2.7 (1.9-3.5) | 31303 |
| **<$5.50/day** | 18.2 (17.6-18.8) | 804359 | 4.8 (4.5-5.1) | 738802 | 25.5 (24.9-26.1) | 935766 | 7.3 (7.0-7.6) | 935766 | 17.9 (14.3-21.4) | 568193 | 5.1 (3.8-6.4) | 46179 |
| **>$5.50/day** | 20.8 (20.0-21.6) | 266641 | 7.6 (6.5-8.7) | 162184 | 42.8 (41.1-44.6) | 348180 | 14.2 (13.2-15.2) | 348180 | 21.2 (18.7-23.7) | 342722 | 17.0 (12.6-21.4) | 24412 |
| ***Low income countries*** | | | | | | | | | | | | |
| **Total** | 21.6 (20.5-22.7) | 26652 | 3.6 (3.2-4.0) | 21141 | 15.2 (14.6-15.9) | 133201 | 5.3 (5-5.6.0) | 133201 | 4.9 (4.5-5.3) | 244213 | 1.0 (0.8-1.3) | 14953 |
| **<$1.90/day** | 23.1 (21.7-24.6) | 13668 | 4.4 (3.8-5.0) | 11299 | 10.1 (9.7-10.6) | 62553 | 3.7 (3.4-4.0) | 62553 | 5.5 (5.0-6.0) | 105415 | 0.7 (0.4-1.3) | 7166 |
| **<$3.20/day** | 21.2 (20.0-22.4) | 20574 | 3.8 (3.4-4.3) | 16785 | 11.3 (10.8-11.8) | 97770 | 3.7 (3.5-4.0) | 97770 | 5.2 (4.8-5.7) | 170283 | 0.9 (0.6-1.2) | 11452 |
| **<$5.50/day** | 21.3 (20.2-22.4) | 24431 | 3.6 (3.2-4.0) | 19639 | 13.7 (13.1-14.3) | 120347 | 4.6 (4.3-4.9) | 120347 | 5.0 (4.6-5.4) | 216124 | 0.9 (0.7-1.2) | 13696 |
| **>$5.50/day** | 25.5 (22.1-29.1) | 2221 | 4.5 (3.3-6.2) | 1502 | 38.7 (36.8-40.7) | 12854 | 15.7 (14.5-17.1) | 12854 | 3.4 (2.6-4.4) | 28089 | 1.8 (0.9-3.2) | 1257 |
| ***Lower middle income countries*** | | | | | | | | | | | | |
| **Total** | 16.1 (15.5-16.7) | 900946 | 5.2 (4.8-5.6) | 841439 | 30.0 (29.6-30.5) | 950853 | 10.1 (9.8-10.4) | 950853 | 18.3 (12.9-25.4) | 408569 | 13.6 (9.9-18.2) | 39986 |
| **<$1.90/day** | 1.03 (12.6-13.4) | 183282 | 2.9 (2.7-3.2) | 178094 | 9.8 (9.4-10.1) | 193178 | 2.0 (1.9-2.2) | 193178 | 9.5 (7.0-12.7) | 75809 | 1.7 (0.9-3.1) | 9029 |
| **<$3.20/day** | 14.1 (13.6-14.6) | 492770 | 3.5 (3.3-3.9) | 475905 | 17.1 (16.7-17.5) | 499269 | 4.4 (4.3-4.7) | 499269 | 16.1 (11.3-22.4) | 172527 | 4.2 (2.7-6.4) | 17842 |
| **<$5.50/day** | 15.2 (14.7-15.8) | 735628 | 4.4 (4.1-4.7) | 701342 | 25.1 (24.6-25.6) | 753347 | 7.9 (7.6-8.2) | 753347 | 17.6 (12.2-24.7) | 276437 | 8.9 (6.4-12.3) | 27510 |
| **>$5.50/day** | 19.1 (18.5-19.8) | 165318 | 8.0 (7.4-8.7) | 140097 | 48.2 (47.0-49.5) | 197506 | 18.2 (17.2-19.2) | 197506 | 20.0 (14.4-27.0) | 132132 | 27.9 (20.1-37.4) | 12476 |
| ***Upper middle income countries*** | | | | | | | | | | | | |
| **Total** | 22.6 (21.5-23.8) | 143402 | 6.7 (5.6-8.0) | 38406 | 36.5 (34.0-39.0) | 199892 | 9.7 (8.5-11.1) | 199892 | 23.7 (22.4-25.2) | 258133 | 12.3 (10.8-14.0) | 15652 |
| **<$1.90/day** | 25.4 (22.4-28.8) | 8079 | 6.9 (5.3-8.9) | 4161 | 30.3 (26.8-34.1) | 9501 | 6.3 (5.0-7.9) | 9501 | 23.3 (18.7-28.6) | 10812 | 10.9 (6.1-18.7) | 656 |

| | | | | | | | | | | | | |
|---|---|---|---|---|---|---|---|---|---|---|---|---|
| **<$3.20/day** | 24.7 (22.8-26.8) | 20352 | 6.3 (5.3-7.5) | 9724 | 30.4 (28.0-32.9) | 26306 | 6.3 (5.3-7.4) | 26306 | 25.6 (22.6-28.9) | 30390 | 12.8 (9.3-17.3) | 2009 |
| **<$5.50/day** | 24.0 (22.4-25.7) | 44300 | 5.9 (5.1-6.9) | 17821 | 31.6 (29.4-33.8) | 62072 | 6.8 (5.8-7.8) | 62072 | 25.6 (23.7-27.6) | 75632 | 11.7 (9.3-14.7) | 4973 |
| **>$5.50/day** | 21.6 (20.4-22.8) | 99102 | 7.5 (6.0-9.4) | 20585 | 40.0 (37.2-42.9) | 137820 | 11.9 (10.4-13.5) | 137820 | 22.5 (20.8-24.2) | 182501 | 12.6 (11.2-14.1) | 10679 |

Table S21: Prevalences by poverty level and World Bank country classification, females only

| Poverty level | Hypertension | | Diabetes | | Overweight | | Obesity | | Smoking | | Dyslipidemia | |
|---|---|---|---|---|---|---|---|---|---|---|---|---|
| | Prevalence | Obs. | Prevalence | Obs. | Prevalence | Obs. | Prevalence | Obs. | Prevalence | Obs. | Prevalence | Obs. |
| ***All poverty levels*** | | | | | | | | | | | | |
| **Total** | 16.8 (16.3-17.3) | 815382 | 5.2 (4.8-5.6) | 726995 | 30.7 (30.0-31.4) | 1196911 | 10.2 (9.9-10.6) | 1196911 | 2.0 (1.8-2.3) | 576618 | 10.1 (8.1-12.6) | 40634 |
| **<$1.90/day** | 15.4 (14.7-16.2) | 165709 | 3.8 (3.4-4.3) | 160090 | 12.3 (11.8-12.9) | 257784 | 3.1 (2.9-3.4) | 257784 | 1.0 (0.9-1.2) | 130770 | 1.7 (1.1-2.5) | 10310 |
| **<$3.20/day** | 15.3 (14.8-15.9) | 430747 | 4.0 (3.7-4.3) | 415489 | 18.1 (17.6-18.6) | 604235 | 4.9 (4.7-5.1) | 604235 | 1.2 (1.0-1.5) | 244132 | 3.7 (2.7-5.2) | 18499 |
| **<$5.50/day** | 16.0 (15.5-16.6) | 639672 | 4.5 (4.2-4.8) | 606995 | 25.3 (24.7-25.9) | 899858 | 7.8 (7.5-8.1) | 899858 | 1.4 (1.1-1.7) | 367233 | 7.1 (5.6-9.2) | 26940 |
| **>$5.50/day** | 18.4 (17.6-19.2) | 175710 | 6.8 (6.0-7.7) | 120000 | 44.2 (42.8-45.5) | 297053 | 16.3 (15.5-17.1) | 297053 | 3.1 (2.8-3.5) | 209385 | 19.2 (15.1-24.3) | 13694 |
| ***Low income countries*** | | | | | | | | | | | | |
| **Total** | 20.5 (19.2-21.8) | 14531 | 3.4 (2.9-3.8) | 11636 | 15.0 (14.4-15.7) | 132398 | 5.2 (4.9-5.5) | 132398 | 0.7 (0.6-1.0) | 145784 | 1.2 (0.9-1.6) | 8662 |
| **<$1.90/day** | 21.8 (20.1-23.6) | 7965 | 4.2 (3.5-5.0) | 6615 | 9.8 (9.4-10.3) | 62122 | 3.6 (3.3-3.9) | 62122 | 0.9 (0.6-1.3) | 67745 | 0.8 (0.5-1.2) | 4504 |
| **<$3.20/day** | 20.1 (18.7-21.5) | 11579 | 3.5 (3.0-4.0) | 9526 | 11.0 (10.6-11.5) | 97100 | 3.6 (3.4-3.9) | 97100 | 0.8 (0.6-1.1) | 105133 | 1.1 (0.8-1.5) | 6898 |
| **<$5.50/day** | 20.3 (19.0-21.6) | 13504 | 3.3 (2.9-3.8) | 10937 | 13.4 (12.9-14.0) | 119571 | 4.5 (4.2-4.8) | 119571 | 0.8 (0.6-1.0) | 130540 | 1.2 (0.9-1.6) | 8076 |
| **>$5.50/day** | 23.2 (18.3-29.0) | 1027 | 4.4 (2.6-7.3) | 699 | 38.7 (36.7-40.6) | 12827 | 15.7 (14.5-17.1) | 12827 | 0.4 (0.2-0.8) | 15244 | 1.7 (0.8-3.6) | 586 |
| ***Lower middle income countries*** | | | | | | | | | | | | |
| **Total** | 14.6 (14.0-15.2) | 721839 | 5.1 (4.8-5.4) | 693632 | 30.0 (29.4-30.7) | 904049 | 10.3 (10.0-10.6) | 904049 | 1.2 (0.9-1.7) | 281672 | 16.9 (12.7-22.0) | 22818 |
| **<$1.90/day** | 11.9 (11.5-12.3) | 153431 | 2.9 (2.7-3.2) | 150977 | 9.8 (9.4-10.2) | 188072 | 2.1 (1.9-2.2) | 188072 | 0.9 (0.7-1.2) | 56572 | 2.7 (1.4-5.1) | 5380 |
| **<$3.20/day** | 12.8 (12.4-13.3) | 408113 | 3.5 (3.2-3.8) | 400263 | 17.2 (16.7-17.7) | 485770 | 4.5 (4.3-4.8) | 485770 | 1.0 (0.8-1.5) | 120638 | 6.2 (4.1-9.2) | 10327 |
| **<$5.50/day** | 13.9 (13.3-14.4) | 601913 | 4.4 (4.1-4.7) | 585705 | 25.5 (24.9-26.0) | 729444 | 8.1 (7.8-8.4) | 729444 | 1.1 (0.8-1.6) | 191146 | 12.7 (9.4-17.0) | 15784 |
| **>$5.50/day** | 17.4 (16.7-18.1) | 119926 | 8.2 (7.7-8.8) | 107927 | 49.1 (48.2-50.0) | 174605 | 19.4 (18.7-20.2) | 174605 | 1.6 (1.3-2.0) | 90526 | 30.1 (21.1-41.0) | 7034 |
| ***Upper middle income countries*** | | | | | | | | | | | | |
| **Total** | 20.3 (19.2-21.5) | 79012 | 5.5 (4.5-6.8) | 21727 | 37.6 (35.5-39.7) | 160464 | 11.6 (10.6-12.7) | 160464 | 3.5 (3.0-4.1) | 149162 | 14.0 (11.7-16.7) | 9154 |
| **<$1.90/day** | 24.8 (21.2-28.7) | 4313 | 7.0 (5.1-9.5) | 2498 | 32.4 (28.8-36.3) | 7590 | 8.9 (7.1-11.1) | 7590 | 2.4 (1.5-3.6) | 6453 | 7.8 (4.7-12.7) | 426 |

| | | | | | | | | | | | | |
|---|---|---|---|---|---|---|---|---|---|---|---|---|
| **<$3.20/day** | 23.6 (21.5-25.9) | 11055 | 6.2 (5.1-7.7) | 5700 | 32.0 (29.6-34.6) | 21365 | 8.2 (7.0-9.6) | 21365 | 2.1 (1.4-3.2) | 18361 | 17.1 (8.8-30.5) | 1274 |
| **<$5.50/day** | 22.0 (20.3-23.7) | 24255 | 5.3 (4.4-6.4) | 10353 | 33.5 (31.2-35.9) | 50843 | 8.7 (7.6-9.8) | 50843 | 2.5 (1.9-3.4) | 45547 | 14.6 (11.1-19.0) | 3080 |
| **>$5.50/day** | 19.0 (17.8-20.3) | 54757 | 5.8 (4.4-7.5) | 11374 | 40.4 (38.2-42.7) | 109621 | 13.7 (12.5-14.9) | 109621 | 4.2 (3.6-4.8) | 103615 | 13.7 (11.6-16.1) | 6074 |

Table S22: Prevalences by poverty level and World Bank country classification, males only

| Poverty level | Hypertension | | Diabetes | | Overweight | | Obesity | | Smoking | | Dyslipidemia | |
|---|---|---|---|---|---|---|---|---|---|---|---|---|
| | Prevalence | Obs. | Prevalence | Obs. | Prevalence | Obs. | Prevalence | Obs. | Prevalence | Obs. | Prevalence | Obs. |
| ***All poverty levels*** | | | | | | | | | | | | |
| **Total** | 23.1 (22.2-23.9) | 255618 | 6.7 (5.9-7.5) | 173991 | 34.1 (31.2-37.1) | 87035 | 7.2 (5.8-8.9) | 87035 | 44.7 (40.1-49.4) | 334297 | 6.6 (4.9-8.9) | 23666 |
| **<$1.90/day** | 22.1 (20.6-23.7) | 39320 | 4.6 (3.9-5.4) | 33464 | 24.1 (19.6-29.2) | 7448 | 2.6 (1.7-3.9) | 7448 | 34.2 (30.3-38.4) | 61266 | 1.0 (0.6-2.0) | 6541 |
| **<$3.20/day** | 21.6 (20.6-22.6) | 102949 | 4.8 (4.2-5.4) | 86925 | 24.7 (21.8-27.9) | 19110 | 3.5 (2.6-4.5) | 19110 | 44.2 (38.7-49.9) | 129068 | 1.5 (1.0-2.4) | 12804 |
| **<$5.50/day** | 22.6 (21.6-23.6) | 164687 | 5.3 (4.8-5.9) | 131807 | 26.6 (24.3-29.2) | 35908 | 4.2 (3.4-5.1) | 35908 | 47.1 (41.4-52.9) | 200960 | 3.0 (2.1-4.3) | 19239 |
| **>$5.50/day** | 23.8 (22.6-25.0) | 90931 | 8.7 (7.3-10.3) | 42184 | 40.1 (36.6-43.6) | 51127 | 9.7 (7.8-12) | 51127 | 42.0 (38.3-45.8) | 133337 | 15.2 (10.3-22.0) | 10718 |
| ***Low income countries*** | | | | | | | | | | | | |
| **Total** | 22.6 (21.2-24.1) | 12121 | 3.9 (3.3-4.5) | 9505 | 42.4 (38.0-46.9) | 803 | 16.3 (13.3-19.7) | 803 | 17.2 (15.8-18.7) | 98429 | 0.8 (0.5-1.3) | 9732 |
| **<$1.90/day** | 24.4 (22.5-26.5) | 5703 | 4.5 (3.8-5.4) | 4684 | 43.4 (38.1-48.8) | 431 | 17.1 (13.7-21.1) | 431 | 21.0 (18.9-23.4) | 37670 | 0.7 (0.2-2.1) | 2662 |
| **<$3.20/day** | 22.3 (20.7-23.9) | 8995 | 4.1 (3.5-4.8) | 7259 | 42.5 (38.2-46.8) | 670 | 15.9 (13.2-19.1) | 670 | 18.2 (16.6-19.9) | 65150 | 0.7 (0.3-1.3) | 4554 |
| **<$5.50/day** | 22.2 (20.7-23.8) | 10927 | 3.8 (3.3-4.5) | 8702 | 42.0 (37.5-46.5) | 776 | 16.3 (13.3-19.8) | 776 | 17.6 (16.1-19.2) | 85584 | 0.7 (0.4-1.2) | 5620 |
| **>$5.50/day** | 27.1 (22.9-31.7) | 1194 | 4.6 (3.0-7.1) | 803 | - | 27[1] | - | 27[1] | 12.6 (9.7-16.2) | 12845 | 1.8 (0.7-4.4) | 671 |
| ***Lower middle income countries*** | | | | | | | | | | | | |
| **Total** | 20.3 (19.7-20.9) | 179107 | 5.4 (4.9-6.0) | 147807 | 30.0 (26.5-33.7) | 46804 | 7.1 (5.8-8.6) | 46804 | 51.5 (41.9-61.0) | 126897 | 10.2 (6.3-16.0) | 17168 |
| **<$1.90/day** | 17.3 (16.4-18.3) | 29851 | 3.0 (2.6-3.4) | 27117 | 9.7 (8.1-11.6) | 5106 | 1.6 (1.1-2.4) | 5106 | 44.2 (36.4-52.3) | 19237 | 0.7 (0.3-1.5) | 3649 |
| **<$3.20/day** | 18.2 (17.5-18.9) | 84657 | 3.9 (3.5-4.4) | 75642 | 13.7 (11.8-15.8) | 13499 | 2.4 (1.9-3.0) | 13499 | 54.0 (44.4-63.4) | 51889 | 2.1 (1.1-4.1) | 7515 |
| **<$5.50/day** | 19.4 (18.8-20.1) | 133715 | 4.7 (4.2-5.1) | 115637 | 17.9 (16.3-19.6) | 23903 | 3.4 (2.9-4.0) | 23903 | 54.4 (44.5-64.0) | 85291 | 5.0 (2.9-8.3) | 11726 |
| **>$5.50/day** | 22.5 (21.8-23.2) | 45392 | 7.6 (6.6-8.7) | 32170 | 43.5 (39.7-47.4) | 22901 | 11.3 (9.7-13.1) | 22901 | 46.4 (37.4-55.7) | 41606 | 25.8 (14.7-41.1) | 5442 |
| ***Upper middle income countries*** | | | | | | | | | | | | |
| **Total** | 25.0 (23.5-26.6) | 64390 | 8.0 (6.7-9.5) | 16679 | 35.0 (31.7-38.5) | 39428 | 7.2 (5.6-9.3) | 39428 | 44.1 (41.5-46.7) | 108971 | 11.0 (9.0-13.4) | 6498 |
| **<$1.90/day** | 26.2 (22.2-30.6) | 3766 | 6.8 (4.7-9.6) | 1663 | 27.4 (21.9-33.7) | 1911 | 2.6 (1.5-4.3) | 1911 | 45.0 (36.0-54.4) | 4359 | 13.8 (5.1-32.3) | 230 |

| | | | | | | | | | | | | |
|---|---|---|---|---|---|---|---|---|---|---|---|---|
| **<$3.20/day** | 26.0 (23.4-28.9) | 9297 | 6.4 (5.0-8.2) | 4024 | 28.2 (24.8-31.8) | 4941 | 3.7 (2.6-5.1) | 4941 | 49.7 (44.4-55.1) | 12029 | 8.4 (4.3-15.6) | 735 |
| **<$5.50/day** | 26.3 (24.1-28.6) | 20045 | 6.6 (5.6-7.9) | 7468 | 29.1 (26.5-31.9) | 11229 | 4.3 (3.3-5.6) | 11229 | 49.8 (46.0-53.6) | 30085 | 8.6 (5.7-12.9) | 1893 |
| **>$5.50/day** | 24.1 (22.5-25.8) | 44345 | 9.2 (7.4-11.4) | 9211 | 39.4 (35.3-43.7) | 28199 | 9.4 (7.2-12.2) | 28199 | 40.4 (37.5-43.5) | 78886 | 11.8 (9.6-14.3) | 4605 |

[1] No prevalence calculated when sample size was <50.

Figure S9: Prevalences by poverty level and WB country classification, disaggregated by sex[1,2]

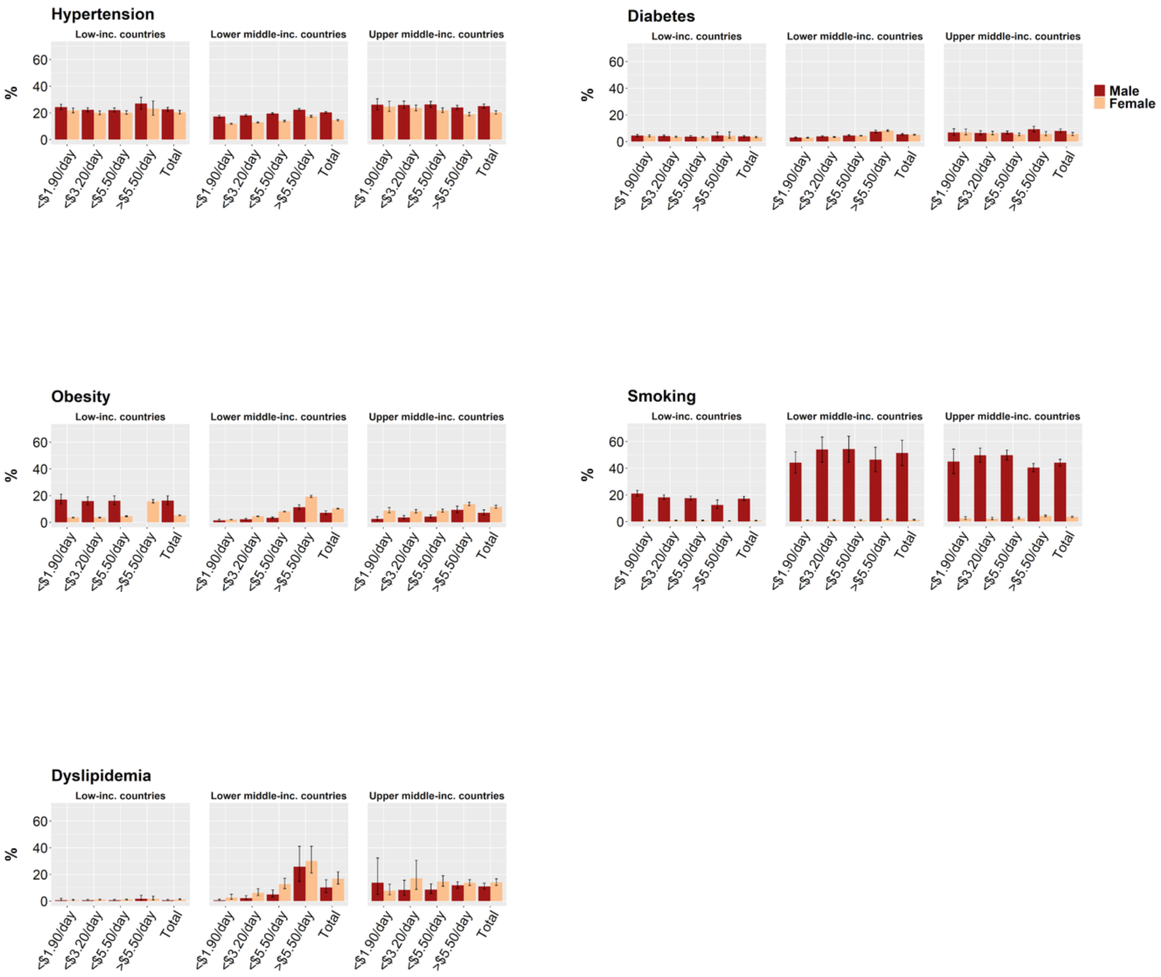

[1]No prevalence calculated when sample size was <50.
[2]Data are point estimates with 95% CIs.

Table S23: Prevalences by 10-year age groups and region (<$1.90/day only)

| Region | Hypertension | | Diabetes | | Overweight | | Obesity | | Smoking | | Dyslipidemia | |
|---|---|---|---|---|---|---|---|---|---|---|---|---|
| ***Total*** | 10.6 (10.2 - 11.0) | Obs. | 12.5 (11.5 - 13.5) | Obs. | 14.9 (14.5 - 15.4) | Obs. | 3.0 (2.8 - 3.2) | Obs. | 3.1 (2.9 - 3.3) | Obs. | 2.8 (1.9 - 3.8) | Obs. |
| ***15-24*** | | | | | | | | | | | | |
| **Total** | 5.8 (5.4 - 6.1) | 57810 | 2.1 (1.9 - 2.3) | 56387 | 5.6 (5.3 - 6.0) | 85563 | 1.8 (1.6 - 2.0) | 85365 | 4.6 (3.6 - 6.0) | 56976 | 0.7 (0.4 - 1.3) | 2394 |
| **East Asia and Pacific** | 5.1 (3.0 - 8.6) | 561 | 0.2 (0.0 - 1.1) | 266 | 5.0 (3.2 - 7.7) | 1420 | 1.3 (0.7 - 2.5) | 1420 | 50.2 (36.0 - 64.3) | 1606 | 0 (0 - 0) | 234 |
| **Europe and Central Asia** | 15.1 (6.1 - 32.8) | 94 | - | 40[1] | 25.4 (12.9 - 43.8) | 217 | 8.3 (2.3 - 25.5) | 217 | 15.5 (6.7 - 32.0) | 244 | - | 29[1] |
| **Latin America and the Caribbean** | 6.8 (2.9 - 15.0) | 419 | - | 23[1] | 22.5 (18.8 - 26.7) | 3662 | 4.9 (3.9 - 6.1) | 3662 | 5.3 (3.4 - 8.2) | 2700 | - | 18[1] |
| **Middle East and North Africa** | 6.2 (3.1 - 11.8) | 162 | - | 6[1] | 17.1 (11.6 - 24.3) | 1632 | 4.1 (2.5 - 6.9) | 1632 | 10.5 (8.1 - 13.5) | 3812 | - | 6[1] |
| **South Asia** | 4.5 (4.2 - 4.7) | 52118 | 1.9 (1.8 - 2.1) | 52472 | 2.4 (2.2 - 2.6) | 47781 | 0.5 (0.4 - 0.6) | 47781 | 4.6 (3.4 - 6.2) | 2861 | - | 48[1] |
| **Sub-Saharan Africa** | 11.9 (10.4 - 13.6) | 4456 | 3.0 (2.4 - 3.7) | 3580 | 8.2 (7.4 - 9.0) | 30851 | 3.1 (2.7 - 3.5) | 30653 | 2.3 (1.8 - 2.8) | 45753 | 0.8 (0.4 - 1.4) | 2059 |
| ***25-34*** | | | | | | | | | | | | |
| **Total** | 12.2 (11.6 - 12.8) | 54791 | 2.7 (2.4 - 3.0) | 52433 | 10.8 (10.3 - 11.3) | 73874 | 2.8 (2.6 - 3.1) | 73721 | 13.5 (11.7 - 15.5) | 48570 | 0.8 (0.4 - 1.4) | 4469 |
| **East Asia and Pacific** | 6.2 (4.2 - 9.2) | 931 | 6.3 (2.1 - 17.7) | 629 | 11.8 (7.5 - 18.1) | 1571 | 2.9 (1.3 - 6.4) | 1571 | 63.7 (49.1 - 76.2) | 1679 | 0.4 (0.1 - 1.6) | 573 |
| **Europe and Central Asia** | 16.2 (5.7 - 38.0) | 64 | - | 43[1] | 31.4 (16.5 - 51.4) | 171 | 5.8 (1.2 - 24.2) | 171 | 31.1 (15.9 - 51.9) | 187 | - | 39[1] |
| **Latin America and the Caribbean** | 10.3 (6.8 - 15.3) | 648 | - | 38[1] | 40.7 (35.9 - 45.6) | 2858 | 11.7 (9.2 - 14.8) | 2858 | 13.4 (10.3 - 17.2) | 2304 | - | 35[1] |
| **Middle East and North Africa** | 10.4 (4.9 - 20.7) | 101 | - | 14[1] | 36.8 (30.5 - 43.6) | 1193 | 18.5 (13.9 - 24.2) | 1193 | 19.8 (16.8 - 23.2) | 2424 | - | 14[1] |
| **South Asia** | 9.9 (9.5 - 10.3) | 45073 | 2.6 (2.3 - 2.8) | 45305 | 7.2 (6.8 - 7.7) | 41566 | 1.1 (0.9 - 1.2) | 41566 | 14.0 (12.0 - 16.3) | 4586 | 4.2 (1.5 - 11.3) | 100 |
| **Sub-Saharan Africa** | 17.5 (15.8 - 19.3) | 7974 | 2.5 (2.0 - 3.1) | 6404 | 12.6 (11.7 - 13.6) | 26515 | 3.9 (3.5 - 4.4) | 26362 | 5.1 (4.6 - 5.8) | 37390 | 0.6 (0.3 - 1.4) | 3708 |
| ***35-44*** | | | | | | | | | | | | |
| **Total** | 18.8 (18.1 - 19.6) | 46651 | 3.6 (3.3 - 4.0) | 44726 | 14.6 (13.9 - 15.3) | 59107 | 4.1 (3.7 - 4.5) | 58994 | 16.9 (15.1 - 18.8) | 36555 | 1.6 (0.9 - 2.9) | 3594 |
| **East Asia and Pacific** | 17.6 (9.4 - 30.7) | 805 | 5.1 (2.0 - 12.7) | 550 | 13.8 (8.8 - 21.1) | 1269 | 3.3 (1.6 - 6.6) | 1269 | 56.3 (42.1 - 69.6) | 1376 | 0.6 (0.2 - 1.4) | 529 |

| | | | | | | | | | | | | |
|---|---|---|---|---|---|---|---|---|---|---|---|---|
| **Europe and Central Asia** | 21.4 (10.9 - 37.7) | 100 | 15.8 (6.1 - 35.4) | 72 | 72.3 (51.5 - 86.6) | 233 | 33.7 (17.0 - 55.9) | 233 | 24.1 (10.6 - 45.9) | 243 | 4.9 (0.6 - 29.0) | 59 |
| **Latin America and the Caribbean** | 20.1 (13.7 - 28.6) | 650 | - | 33[1] | 48.2 (42.1 - 54.4) | 2282 | 12.9 (9.1 - 18.2) | 2282 | 14.2 (10.4 - 19.0) | 2173 | - | 29[1] |
| **Middle East and North Africa** | 19.8 (13.6 -28.0) | 115 | - | 20[1] | 50.3 (44.6 - 56.1) | 961 | 29.1 (23.9 - 34.9) | 961 | 22.6 (19.5 - 26.1) | 1773 | - | 19[1] |
| **South Asia** | 16.4 (15.8 - 17.0) | 38790 | 3.4 (3.1 - 3.7) | 38938 | 10.1 (9.6 - 10.6) | 34976 | 1.5 (1.3 - 1.8) | 34976 | 17.6 (15.7 - 19.7) | 3865 | 10.7 (5.2 - 20.9) | 127 |
| **Sub-Saharan Africa** | 25.0 (23.1 - 27.1) | 6191 | 3.7 (3.0 - 4.7) | 5108 | 17.3 (16.0 - 18.8) | 19386 | 6.0 (5.3 - 6.8) | 19273 | 6.7 (5.9 - 7.7) | 27125 | 0.9 (0.4 - 2.1) | 2831 |
| ***45-54*** | | | | | | | | | | | | |
| **Total** | 29.8 (28.5 - 31.2) | 23926 | 5.5 (4.9 - 6.2) | 22216 | 19.1 (17.7 - 20.6) | 25205 | 5.3 (4.7 - 6.0) | 25123 | 19.4 (17.2 - 21.8) | 20650 | 6.5 (3.3 - 12.4) | 2851 |
| **East Asia and Pacific** | 33.2 (24.8 - 42.9) | 719 | 4.7 (1.6 - 13.1) | 496 | 33.6 (18.9 - 52.4) | 844 | 2.7 (1.3 - 5.8) | 844 | 67.5 (55.0 - 77.9) | 943 | 26.5 (8.2 - 59.3) | 491 |
| **Europe and Central Asia** | 36.3 (21.2 - 54.7) | 95 | 22.6 (9.5 - 45.0) | 71 | 86.5 (64.3 - 95.8) | 165 | 65.8 (43.9 - 82.5) | 165 | 19.5 (6.9 - 44.1) | 183 | - | 33[1] |
| **Latin America and the Caribbean** | 34.0 (28.2 - 40.2) | 751 | 36.2 (14.8 - 64.9) | 86 | 48.6 (40.8 - 56.3) | 1238 | 16.3 (11.1 - 23.4) | 1238 | 17.1 (12.6 - 22.7) | 1406 | 12.8 (5.7 - 26.0) | 77 |
| **Middle East and North Africa** | 30.7 (20.7 - 42.9) | 104 | - | 13[1] | 58.6 (50.4 - 66.4) | 360 | 36.0 (27.5 - 45.4) | 360 | 27.8 (22.9 - 33.2) | 1454 | - | 13[1] |
| **South Asia** | 24.0 (23.0 - 24.9) | 17765 | 5.2 (4.7 - 5.8) | 17844 | 11.8 (10.9 - 12.8) | 14528 | 1.7 (1.4 - 2.1) | 14528 | 19.8 (17.4 - 22.5) | 2637 | 17.5 (10.3 - 28.1) | 111 |
| **Sub-Saharan Africa** | 39.4 (36.1 - 42.7) | 4492 | 4.9 (4.0 - 5.9) | 3702 | 21.5 (19.7 - 23.5) | 8070 | 7.9 (6.7 - 9.3) | 7988 | 8.4 (7.2 - 9.8) | 14027 | 1.6 (0.9 - 2.7) | 2126 |
| ***55-64*** | | | | | | | | | | | | |
| **Total** | 47.9 (45.0 - 50.8) | 4877 | 5.5 (4.5 -6.8) | 3679 | 36.0 (27.3 - 45.5) | 2208 | 13.0 (9.3 - 18.0) | 2176 | 21.4 (18.7 - 24.3) | 8717 | 10.5 (6.0 - 18.0) | 2285 |
| **East Asia and Pacific** | 41.5 (32.7 - 50.9) | 618 | 3.0 (1.1 - 7.8) | 509 | 15.0 (6.5 - 30.9) | 441 | 0.8 (0.3 - 2.5) | 441 | 58.3 (42.3 - 72.7) | 500 | 29.2 (8.7 - 64.0) | 462 |
| **Europe and Central Asia** | 67.8 (49.2 - 82.1) | 88 | 20.0 (9.1 - 38.5) | 81 | 70.8 (46.7 - 87.0) | 84 | 44.2 (23.5 - 67.1) | 84 | 57.1 (34.9 - 76.8) | 94 | - | 29[1] |
| **Latin America and the Caribbean** | 43.2 (32.9 - 54.1) | 539 | 26.6 (16.2 - 40.3) | 59 | 52.9 (41.4 - 64.1) | 382 | 15.3 (10.7 - 21.6) | 382 | 21.0 (14.0 - 30.4) | 717 | 20.8 (11.9 - 33.8) | 56 |
| **Middle East and North Africa** | - | 42[1] | - | 16[1] | - | 18[1] | - | 18[1] | 16.6 (13.0 - 20.9) | 990 | - | 17[1] |
| **South Asia** | 48.0 (40.9 - 55.3) | 287 | 6.1 (3.8 - 9.7) | 278 | 21.0 (3.9 - 63.6) | 133 | 2.6 (0.3 - 18.1) | 133 | 21.4 (18.6 - 24.5) | 2004 | 14.5 (7.2 - 27.3) | 87 |
| **Sub-Saharan Africa** | 48.2 (45.0 - 51.4) | 3303 | 4.9 (3.9 - 6.2) | 2734 | 49.8 (43.6 - 56.0) | 1150 | 25.5 (20.3 - 31.5) | 1118 | 9.9 (7.3 - 13.4) | 4412 | 5.4 (2.6 - 10.9) | 1634 |
| ***>65*** | | | | | | | | | | | | |

| | | | | | | | | | | | | |
|---|---|---|---|---|---|---|---|---|---|---|---|---|
| **Total** | 51.9 (46.8 - 57.0) | 2027 | 8.9 (5.4 - 14.1) | 1018 | 23.9 (15.9 - 34.3) | 1151 | 9.0 (5.6 - 14.1) | 1149 | 19.6 (16.9 - 22.7) | 6517 | 7.4 (3.4 - 15.6) | 417 |
| **East Asia and Pacific** | 57.4 (45.0 - 69.0) | 366 | 10.2 (3.1 - 28.9) | 217 | 3.6 (1.6 - 8.0) | 283 | 0.4 (0.1 - 1.7) | 283 | 43.9 (31.1 - 57.5) | 322 | 0.1 (0.0 - 0.4) | 131 |
| **Europe and Central Asia** | - | 11[1] | - | 10[1] | - | 4[1] | - | 4[1] | - | 5[1] | - | 9[1] |
| **Latin America and the Caribbean** | 45.1 (38.3 - 52.9) | 631 | - | 43[1] | 42.4 (33.6 - 51.7) | 385 | 10.9 (6.7 - 17.3) | 385 | 21.5 (15.8 - 28.6) | 497 | - | 41[1] |
| **Middle East and North Africa** | - | 17[1] | - | 22[1] | - | 17[1] | - | 17[1] | 19.9 (14.2 - 27.2) | 1015 | - | 22[1] |
| **South Asia** | 53.9 (45.6 - 61.9) | 214 | 6.4 (3.4 - 11.5) | 205 | 2.4 (0.6 - 9.5) | 121 | 0 (0 - 0) | 121 | 19.5 (16.7 - 22.7) | 1853 | - | 29[1] |
| **Sub-Saharan Africa** | 46.5 (40.9 - 52.2) | 788 | 7.4 (5.2 - 10.6) | 521 | 54.3 (46.4 - 62.0) | 341 | 26.8 (19.6 - 35.4) | 339 | 4.7 (2.8 - 7.7) | 2825 | 7.4 (3.9 - 13.6) | 185 |

[1] No prevalence calculated when sample size was <50

Figure S10: Prevalences by 10-year age groups and region (<$1.90/day only)[1,2]

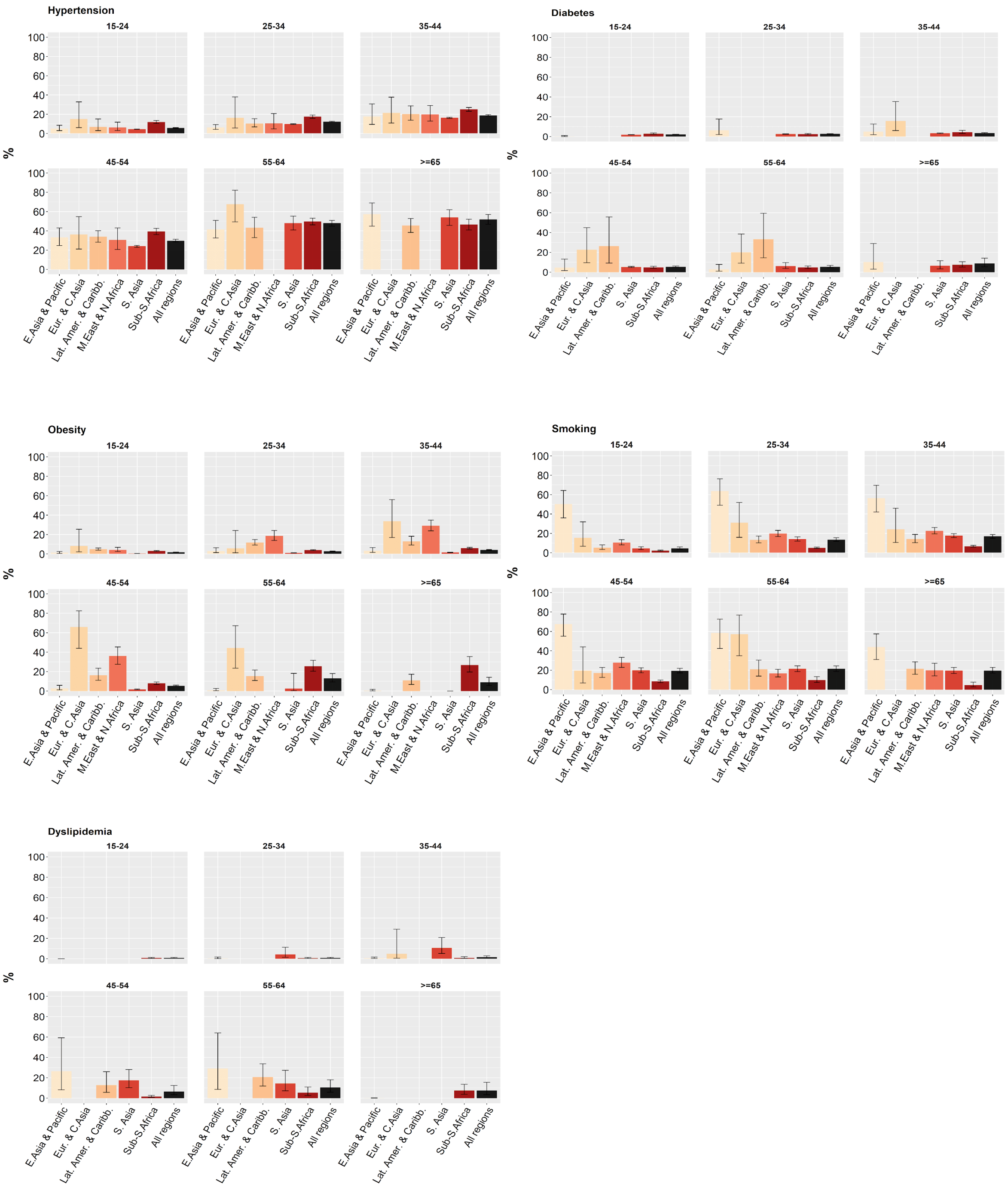

[1]No prevalence calculated when sample size was <50.
[2]Data are point estimates with 95% CIs.

Table S24: Prevalences for each risk factor by country[1]

| | Hypertension | | | | | Diabetes | | | | | Overweight | | | | |
|---|---|---|---|---|---|---|---|---|---|---|---|---|---|---|---|
| **Country** | **<$1.90/day** | **<$3.20/day** | **<$5.50/day** | **>$5.50/day** | **total** | **<$1.90/day** | **<$3.20/day** | **<$5.50/day** | **>$5.50/day** | **total** | **<$1.90/day** | **<$3.20/day** | **<$5.50/day** | **>$5.50/day** | **total** |
| **Albania** | - | 26.4 (20.3-33.6) | 25.9 (22.9-29.1) | 21.0 (19.0-23.1) | 22.5 (20.7-24.4) | - | - | - | - | - | 40.1 (33.9-46.7) | 45.3 (41.5-49.2) | 49.3 (47.3-51.3) | 51.0 (49.3-52.7) | 50.6 (49.2-52.0) |
| **Angola** | - | - | - | - | - | - | - | - | - | - | - | - | - | - | - |
| **Armenia** | - | - | - | - | - | - | - | - | - | - | - | 42.8 (37.7-48.2) | 42.9 (40.5-45.4) | 42.1 (40.1-44.1) | 42.4 (40.8-44.0) |
| **Bangladesh** | 24.8 (22.2-27.6) | 26.8 (25.5-28) | 27.9 (26.8-29.0) | 32.7 (30.2-35.4) | 28.6 (27.6-29.6) | 2.9 (2.1-4.1) | 4.2 (3.6-4.9) | 5.7 (5.1-6.3) | 15.5 (13.5-17.7) | 7.1 (6.5-7.7) | 13.9 (12.5-15.5) | 21.1 (20.0-22.2) | 27.3 (26.2-28.3) | 56.4 (53.7-58.9) | 31.4 (30.2-32.6) |
| **Benin** | 24.2 (19.7-29.4) | 25.5 (21.8-29.7) | 26.0 (22.6-29.8) | 22.8 (13.5-35.8) | 25.7 (22.0-29.7) | 3.4 (2.4-4.7) | 5.0 (3.5-7.0) | 5.0 (3.6-7.0) | 6.8 (3.5-13.0) | 5.2 (3.8-7.2) | 17.3 (14.1-21.1) | 19.9 (16.8-23.5) | 20.7 (17.8-23.9) | 30.7 (18.3-46.9) | 21.8 (18.5-25.5) |
| **Bhutan** | - | 43.0 (35.4-51.0) | 37.9 (33.1-42.9) | 31.1 (27.8-34.7) | 33.6 (30.7-36.6) | - | - | 1.5 (0.9-2.4) | 2.4 (1.5-3.7) | 2.0 (1.4-2.9) | - | 23.5 (18.0-30.2) | 25.7 (22.0-29.9) | 35.9 (32.3-39.6) | 32.1 (29.0-35.4) |
| **Bolivia** | - | - | - | - | - | - | - | - | - | - | 40.6 (37.3-44) | 43.2 (40.8-45.7) | 47.6 (45.9-49.4) | 54.6 (53.4-55.8) | 52.2 (51.2-53.3) |
| **Botswana** | 27.6 (21.6-34.6) | 27.7 (23.2-32.8) | 26.5 (23.2-30.2) | 29.5 (26.0-33.4) | 27.9 (25.3-30.7) | - | 2.0 (1.2-3.1) | 2.0 (1.3-2.8) | 2.1 (1.3-3.2) | 2.0 (1.5-2.7) | 25.3 (19.6-32.0) | 22.8 (19.0-27.2) | 22.4 (19.5-25.7) | 34.1 (30.5-38.0) | 27.7 (25.3-30.2) |
| **Brazil** | 19.5 (16.8-22.5) | 22.6 (20.5-24.8) | 25.3 (23.9-26.8) | 25.4 (24.6-26.1) | 25.4 (24.7-26.1) | - | - | - | - | - | 37.1 (32.9-41.6) | 40.8 (38.1-43.5) | 45.1 (43.4-46.9) | 56.0 (55.0-57.0) | 54.5 (53.6-55.4) |
| **Burkina Faso** | - | - | - | - | - | - | - | - | - | - | 7.3 (6.4-8.3) | 9.3 (8.4-10.3) | 11.8 (10.7-12.9) | 33.3 (29.2-37.8) | 13.4 (12.1-14.8) |
| **Burundi** | - | - | - | - | - | - | - | - | - | - | 4.0 (3.3-4.7) | 5.7 (5.0-6.6) | 7.1 (6.2-8.0) | 34.7 (28.4-41.7) | 7.8 (6.8-8.8) |
| **Cabo Verde** | - | 40.4 (31.6-50.0) | 40.2 (32.8-48.1) | 34.5 (29.4-40.0) | 36.9 (30.6-43.6) | - | - | 7.5 (3.2-16.5) | - | 4.8 (2.0-10.9) | - | 26.4 (20.1-33.8) | 30.0 (23.6-37.2) | 37.7 (31.2-44.7) | 34.5 (28.8-40.7) |
| **Cameroon** | - | - | - | - | - | - | - | - | - | - | 12.1 (9.9-14.9) | 18.9 (16.4-21.7) | 27.2 (24.7-29.8) | 53.0 (49.8-56.2) | 35.8 (33.2-38.5) |
| **Chad** | - | - | - | - | - | - | - | - | - | - | 8.5 (7.4-9.8) | 8.1 (7.2-9.2) | 8.9 (8-9.9) | 27.0 (24.0-30.2) | 11.4 (10.3-12.5) |
| **China** | 27.1 (23.5-31.0) | 26.0 (23.8-28.3) | 25.0 (23.2-26.9) | 21.1 (19.5-22.9) | 23.1 (21.6-24.6) | 6.8 (5.1-8.9) | 6.3 (5.2-7.5) | 5.5 (4.8-6.3) | 6.1 (5.4-6.9) | 5.8 (5.3-6.4) | 26.7 (23.0-30.8) | 26.7 (24.4-29.2) | 27.3 (25.4-29.2) | 28.2 (26.3-30.2) | 27.8 (26.2-29.4) |
| **Comoros** | 25.2 (22.1-28.5) | 24.4 (21.2-27.9) | 24.0 (20.5-28.0) | 25.7 (21.9-29.9) | 24.6 (21.2-28.4) | - | - | 3.4 (2.5-4.7) | 4.3 (2.7-6.7) | 3.7 (2.9-4.8) | 31.8 (27.2-36.7) | 34.1 (31.2-37.2) | 37.1 (34.6-39.6) | 45.4 (42.6-48.3) | 39.9 (37.9-42.0) |
| **Congo** | - | - | - | - | - | - | - | - | - | - | 14.5 (12.7-16.6) | 18.3 (14.5-22.8) | 26.0 (22.7-29.5) | 47.0 (42.0-52.1) | 33.0 (29.9-36.2) |

| | | | | | | | | | | | | | | | |
|---|---|---|---|---|---|---|---|---|---|---|---|---|---|---|---|
| **Congo Democratic Republic** | - | - | - | - | - | - | - | - | - | - | 9.5 (7.6-11.9) | 12.9 (11.0-15.1) | 14.7 (12.8-16.8) | 36.4 (27.2-46.6) | 15.2 (13.3-17.3) |
| **Costa Rica** | - | - | - | - | - | - | - | - | - | - | - | - | - | - | - |
| **Côte d'Ivoire** | - | - | - | - | - | - | - | - | - | - | 26.3 (22.9-29.8) | 28.1 (25.5-30.6) | 33.2 (31.0-35.3) | 50.3 (44.8-55.8) | 37.1 (35.0-39.2) |
| **Dominican Republic** | - | - | - | - | - | - | - | - | - | - | - | 37.6 (32.5-43.0) | 45.5 (42.6-48.4) | 50.0 (48.2-51.8) | 49.0 (47.5-50.6) |
| **Ecuador** | 22.0 (15.0-31.2) | 20.7 (16.6-25.4) | 17.9 (15.5-20.6) | 16.0 (14.4-17.7) | 16.4 (15.1-17.9) | - | 7.2 (4.6-11.2) | 6.9 (5.2-9.3) | 6.1 (5.2-7.2) | 6.3 (5.4-7.3) | 57.7 (46.8-68.0) | 60.1 (54.3-65.6) | 60.4 (56.5-64.1) | 60.9 (58.6-63.2) | 60.8 (58.8-62.7) |
| **Egypt** | 14.2 (8.4-22.8) | 13.5 (12.0-15.2) | 14.8 (14.0-15.8) | 16.3 (14.9-17.9) | 15.3 (14.5-16.0) | - | - | - | - | - | 77.4 (70.3-83.3) | 80.3 (78.6-81.9) | 82.8 (81.7-83.8) | 87.5 (85.8-89.1) | 83.8 (82.9-84.7) |
| **Eswatini**[2] | 21.9 (18.7-25.4) | 20.8 (18.6-23.1) | 21.4 (19.4-23.6) | 20.5 (16.5-25.3) | 21.2 (19.4-23.2) | 3.0 (1.9-4.6) | 3.9 (2.6-5.6) | 4.0 (3.0-5.4) | 4.0 (2.4-6.7) | 4.0 (3.0-5.0) | 40.5 (35.7-45.3) | 41.6 (38.1-45.1) | 41.1 (37.9-44.3) | 39.3 (34.6-43.9) | 40.6 (38.1-43.1) |
| **Ethiopia** | 14.4 (12.1-17.0) | 14.0 (12.3-15.8) | 14.9 (13.3-16.6) | 24.5 (19.6-30.2) | 15.6 (14.0-17.3) | 2.4 (1.5-3.7) | 1.7 (1.1-2.4) | 1.7 (1.2-2.3) | 6.1 (3.7-9.7) | 2.0 (1.5-2.6) | 2.5 (1.9-3.4) | 3.9 (3.3-4.5) | 6.4 (5.4-7.6) | 32.8 (26.4-40.0) | 7.2 (6.1-8.6) |
| **Gabon** | - | - | - | - | - | - | - | - | - | - | - | 18.4 (14.3-23.2) | 26.1 (22.6-29.8) | 47.1 (44.4-50.0) | 45.2 (42.6-48.0) |
| **Gambia** | 26.2 (21.7-31.3) | 26.0 (22.8-29.3) | 24.5 (21.4-27.9) | 24.5 (19.2-30.7) | 24.5 (21.6-27.6) | - | - | - | - | - | 26.9 (22.6-31.5) | 27.0 (24.5-29.7) | 32.3 (30.1-34.6) | 43.7 (38.8-48.7) | 35.8 (33.5-38.1) |
| **Georgia**[2] | 36.2 (25.6-48.2) | 36.0 (29.8-42.7) | 37.4 (33.3-41.8) | 30.4 (27.1-33.8) | 32.9 (30.4-35.6) | - | 5.6 (3.78-8.4) | 4.4 (3.2-5.9) | 4.7 (3.7-6.0) | 4.6 (3.7-5.4) | 48.7 (35.7-61.6) | 59.1 (51.9-66.3) | 64.6 (60.0-69.1) | 60.1 (56.8-64.5) | 62.1 (59.1-65.0) |
| **Ghana**[2] | 31.6 (23.9-40.5) | 31.4 (25.8-37.5) | 39.1 (34.4-44.0) | 43.5 (37.7-49.5) | 40.8 (37.0-44.7) | - | - | - | - | - | 9.6 (6.7-12.4) | 13.1 (10.5-15.7) | 20.8 (18.5-23.0) | 52.5 (50.1-54.9) | 38.7 (36.6-40.9) |
| **Guatemala** | - | - | - | - | - | - | - | - | - | - | 35.1 (32.0-38.3) | 38.3 (36.5-40.2) | 42.7 (41.3-44.1) | 57.2 (56.1-58.3) | 50.0 (49.0-51.1) |
| **Guinea** | - | - | - | - | - | - | - | - | - | - | 15.6 (13.5-18.0) | 20.1 (18.2-22.2) | 24.1 (22.1-26.2) | 36.6 (31.5-41.9) | 25.5 (23.5-27.6) |
| **Haiti** | - | - | - | - | - | - | - | - | - | - | 16.9 (14.9-19.1) | 18.8 (17.2-20.5) | 24.8 (23.5-26.2) | 44.0 (41.6-46.4) | 30.6 (29.2-32.1) |
| **Honduras** | - | - | - | - | - | - | - | - | - | - | 38.7 (36.7-40.7) | 42.1 (40.5-43.6) | 46.6 (45.4-47.9) | 59.4 (58.1-60.6) | 54.2 (53.3-55.2) |
| **India** | 10.0 (9.8-10.2) | 10.6 (10.4-10.7) | 11.4 (11.3-11.6) | 13.8 (13.4-14.2) | 11.8 (11.7-11.9) | 2.7 (2.6-2.9) | 3.2 (3.1-3.3) | 4.2 (4.0-4.3) | 8.1 (7.7-8.5) | 4.8 (4.6-4.9) | 6.1 (5.9-6.3) | 11.4 (11.2-11.6) | 16.9 (16.7-17.0) | 35.2 (34.6-35.9) | 19.6 (19.4-19.8) |
| **Indonesia** | 23.0 (19.4-27.0) | 22.8 (20.7-25.0) | 22.9 (21.3-24.6) | 22.2 (21.0-23.5) | 22.7 (21.3-24.1) | 9.9 (5.5-17) | 7.2 (4.6-11.2) | 6.8 (4.6-10.0) | 8.8 (5.7-13.3) | 7.5 (5.1-10.9) | 16.7 (12.7-21.6) | 23.6 (22.0-25.4) | 27.5 (25.9-29.1) | 38.9 (37.3-40.5) | 31.4 (29.5-33.3) |
| **Iran** | - | 15.3 (12.2-19.0) | 19.5 (18.1-21.0) | 19.4 (18.9-20.0) | 19.4 (18.9-20.0) | - | 6.6 (3.9-11.2) | 5.8 (4.7-7.2) | 7.3 (6.8-7.8) | 7.1 (6.7-7.6) | - | 56.3 (51.1-61.3) | 43.4 (41.2-45.6) | 58.8 (58.0-59.6) | 56.9 (56.1-57.7) |
| **Jordan** | - | 26.5 (14.8-42.7) | 24.7 (19.8-30.3) | 16.2 (14.7-17.8) | 16.8 (15.4-18.4) | - | - | 11.6 (8.2-16.1) | 8.8 (7.6-10.3) | 9.0 (7.8-10.4) | - | 52.4 (42.4-62.2) | 54.1 (50.9-57.3) | 51.4 (49.3-53.4) | 51.6 (49.8-53.5) |

| | | | | | | | | | | | | | | | |
|---|---|---|---|---|---|---|---|---|---|---|---|---|---|---|---|
| **Kenya** | 20.7 (18.0-23.7) | 21.6 (19.3-24.0) | 21.6 (19.4-24.0) | 22.3 (17.3-28.3) | 21.7 (19.5-24.1) | 0.9 (0.5-1.5) | 1.4 (1.0-2.1) | 1.5 (1.1-2.1) | - | 1.6 (1.1-2.2) | 15.6 (14.3-17) | 20.9 (19.7-22.2) | 26.9 (25.5-28.4) | 50.3 (47.4-53.1) | 31.6 (30.2-33.0) |
| **Kiribati** | 30.3 (17.6-47.0) | 27.4 (20.9-35.0) | 33.0 (26.4-40.4) | 37.1 (31.5-43.1) | 34.2 (29.9-38.8) | - | 10.9 (7.4-16) | 12.9 (9.3-17.6) | 20.4 (12.9-30.7) | 15.4 (13.0-18.3) | 77.8 (63.1-87.7) | 77.6 (69.9-83.8) | 79.0 (71.4-85.1) | 86.0 (76.4-92.0) | 81.0 (73.2-87.0) |
| **Kyrgyz Republic** | - | 41.5 (35.4-47.8) | 41.4 (37.6-45.4) | 40.3 (33.5-47.4) | 41.0 (36.9-45.2) | - | 4.0 (2.2-7.0) | 4.7 (3.3-6.8) | 3.9 (2.6-6.0) | 4.4 (3.5-5.6) | 44.7 (33.8-56.2) | 39.8 (35.8-44.0) | 37.5 (35.6-39.4) | 33.1 (30.5-35.8) | 35.5 (33.9-37.2) |
| **Laos** | 10.3 (7.1-14.7) | 12.3 (10.1-14.9) | 14.7 (12.8-16.8) | 21.3 (17.8-25.3) | 16.3 (14.4-18.4) | - | 3.3 (2.1-5.0) | 3.9 (2.9-5.2) | 3.9 (2.5-6.0) | 3.9 (3.0-5.0) | 11.0 (7.1-16.6) | 15.0 (12.3-18.2) | 19.0 (16.5-21.7) | 34.6 (29.0-40.8) | 22.8 (20.1-25.6) |
| **Lesotho** | 12.4 (10.8-14.3) | 13.7 (12.2-15.4) | 14.0 (12.7-15.3) | 21.6 (19.1-24.4) | 16.1 (14.9-17.5) | - | - | - | - | - | 18.1 (16.2-20.1) | 21.3 (19.7-23.0) | 23.9 (22.5-25.4) | 39.8 (37.1-42.6) | 28.5 (27.0-29.9) |
| **Liberia** | 26.6 (23.7-29.8) | 26.1 (23.6-28.8) | 25.6 (23.2-28.1) | - | 25.5 (23.2-27.9) | 12.6 (10.3-15.2) | 13.1 (10.9-15.7) | 12.7 (10.6-15.1) | - | 12.5 (10.5-14.8) | 51.9 (48.1-55.7) | 50.8 (47.4-54.2) | 50.7 (47.4-54.0) | - | 50.9 (47.7-54.1) |
| **Madagascar** | - | - | - | - | - | - | - | - | - | - | 10.3 (9.2-11.4) | 11.5 (10.5-12.5) | 12.3 (11.3-13.5) | 20.7 (13.7-30.0) | 12.5 (11.4-13.6) |
| **Malawi** | 36.6 (33.5-39.8) | 35.9 (33.0-39.0) | 36.1 (33.2-39.1) | - | 35.5 (32.6-38.4) | - | - | - | - | - | 14.9 (13.7-16.3) | 17.4 (16.2-18.7) | 19.2 (17.9-20.6) | 44.5 (38.7-50.6) | 20.1 (18.8-21.5) |
| **Mexico** | 22.5 (13.2-31.9) | 17.6 (14.0-21.1) | 16.6 (15.3-17.9) | 19.4 (18.7-20.1) | 18.5 (17.8-19.3) | 24.1 (17.3-31.0) | 17.9 (12.1-23.6) | 21.7 (16.4-27.0) | 25.3 (22.8-27.9) | 24.2 (21.1-27.3) | 69.2 (57.4-80.7) | 65.1 (55.0-74.4) | 62.2 (55.0-70.0) | 67.7 (65.4-70.0) | 66.1 (62.0-70.2) |
| **Mongolia** | - | 25.4 (18.8-33.4) | 26.4 (23.8-29.3) | 20.2 (18.7-21.8) | 21.7 (20.3-23.1) | - | - | 8.2 (6.4-10.4) | 8.3 (7.2-9.5) | 8.3 (7.3-9.3) | | 50.6 (40.9-60.2) | 49.2 (45.2-53.2) | 52.2 (50.1-54.2) | 51.5 (49.6-53.4) |
| **Morocco** | - | - | - | - | - | - | - | - | - | - | - | - | - | - | - |
| **Mozambique** | 28.5 (23.2-34.3) | 29.2 (24.0-35.0) | 29.4 (24.3-35.1) | | 29.6 (24.5-35.2) | - | 0.8 (0.4-1.6) | 0.9 (0.5-1.6) | - | 0.9 (0.5-1.7) | 10.6 (9.7-11.7) | 14.6 (13.5-15.8) | 16.3 (15.1-17.6) | 48.1 (43.5-52.6) | 17.5 (16.2-18.9) |
| **Myanmar** | 17.1 (12.1-23.6) | 24.3 (20.0-29.1) | 27.3 (22.0-33.3) | 29.0 (26.1-32.1) | 28.0 (24.0-32.4) | - | 3.0 (1.8-4.8) | 4.4 (2.7-7.1) | 6.3 (4.8-8.2) | 5.2 (3.6-7.4) | 9.4 (6.4-13.7) | 14.9 (13.0-17.0) | 18.9 (17.5-20.3) | 30.1 (28.2-32.1) | 23.5 (22.2-24.9) |
| **Namibia** | 29.3 (25.5-33.4) | 34.2 (30.9-37.6) | 35.9 (33.2-38.6) | 47.1 (43.8-50.4) | 40.7 (38.5-42.9) | - | 2.7 (1.8-4.1) | 3.1 (2.3-4.3) | 9.2 (7.4-11.5) | 5.7 (4.7-6.9) | 20.2 (17.5-23.3) | 24.2 (21.9-26.6) | 28.9 (26.8-31.1) | 55.7 (53.5-57.9) | 41.7 (39.6-43.9) |
| **Niger** | 34.0 (31.7-36.2) | 35.3 (33.2-37.3) | 35.6 (33.7-37.6) | - | 35.7 (33.7-37.6) | 22.1 (20.1-24.2) | 22.4 (20.6-24.3) | 22.1 (20.4-23.9) | - | 22.1 (20.4-23.9) | 20.5 (18.5-22.7) | 23.3 (21.4-25.2) | 25.9 (24.0-27.9) | 52.1 (44.6-59.5) | 26.5 (24.6-28.5) |
| **Nigeria** | - | - | - | - | - | - | - | - | - | - | 12.4 (11.2-13.7) | 18.8 (17.7-20.0) | 24.3 (23.2-25.4) | 52.0 (47.7-56.3) | 26.8 (25.6-28.1) |
| **Pakistan**[2] | - | - | - | - | - | - | - | - | - | - | 12.5 (5.7-19.3) | 29.6 (25.7-33.4) | 44.1 (40.8-47.4) | 71.8 (67.3-76.3) | 50.5 (47.3-53.6) |
| **Panama** | - | - | - | - | - | - | - | - | - | - | - | - | - | - | - |
| **Peru** | 19.3 (16.4-22.6) | 18.0 (16.2-19.8) | 16.4 (15.3-17.5) | 22.9 (22.0-23.9) | 21.5 (20.7-22.3) | - | - | - | - | - | 36.9 (32.7-41.2) | 41.7 (39.1-44.4) | 45.1 (43.3-46.9) | 57.7 (56.6-58.7) | 55.3 (54.4-56.3) |
| **Philippines** | - | - | - | - | - | - | - | - | - | - | - | - | - | - | - |

| | | | | | | | | | | | | | | | |
|---|---|---|---|---|---|---|---|---|---|---|---|---|---|---|---|
| **Romania** | - | 19.3 (13.9-26.2) | 17.9 (14.1-22.4) | 25.3 (23.2-27.6) | 23.7 (21.8-25.7) | - | - | 8.1 (5.7-11.5) | 9.5 (8.2-11.0) | 9.2 (8-10.6) | - | 50.3 (42.2-58.4) | 49.0 (43.4-54.6) | 64.0 (61.3-66.7) | 60.8 (58.3-63.2) |
| **Rwanda** | 14.2 (12.5-16.0) | 14.3 (12.8-15.9) | 14.8 (13.3-16.3) | 20.6 (16.2-25.8) | 15.2 (13.8-16.7) | 1.3 (0.8-2.1) | 1.2 (0.8-1.9) | 1.3 (0.8-2.1) | - | 1.5 (0.9-2.4) | 14.1 (12.8-15.4) | 16.8 (15.6-18.0) | 18.9 (17.7-20.2) | 41.8 (37.4-46.4) | 20.7 (19.3-22.1) |
| **Samoa** | - | 19.6 (11.0-32.5) | 17.7 (13.1-23.5) | 21.3 (18.2-24.8) | 20.2 (17.4-23.3) | - | 24.1 (14.1-38.1) | 21.9 (14.6-31.4) | 19.4 (16.7-22.4) | 20.2 (17.7-22.9) | - | 79.6 (67.2-88.2) | 78.9 (73.4-83.5) | 85.9 (78.9-90.8) | 83.8 (79.6-87.2) |
| **São Tomé and Principe** | 36.5 (34.1-38.8) | 37.7 (35.0-40.4) | 38.1 (35.9-40.2) | 40.5 (26.5-54.5) | 38.3 (35.6-41.0) | 3.6 (2.4-5.4) | 2.8 (1.8-4.5) | 2.9 (1.9-4.5) | - | 2.9 (1.7-4.0) | 28.6 (26.5-30.7) | 30.1 (28.5-31.7) | 32.7 (29.2-36.1) | 42.2 (29.4-55.0) | 33.4 (29.4-37.3) |
| **Senegal** | - | - | - | - | - | - | - | - | - | - | 26.0 (22.7-29.5) | 26.6 (24.4-28.9) | 28.4 (26.5-30.4) | 41.1 (35.5-46.9) | 31.4 (29.2-33.8) |
| **Solomon Islands** | 19.2 (15.0-24.3) | 19.9 (16.6-23.7) | 19.0 (16.1-22.3) | 18.3 (13.1-24.9) | 18.9 (16.2-21.9) | 6.0 (3.5-10.0) | 4.8 (3.2-7.2) | 5.0 (3.5-7.0) | - | 5.2 (3.7-7.1) | 46.5 (40.3-52.9) | 55.9 (51.2-60.4) | 57.0 (53.2-60.8) | 69.2 (59.4-77.4) | 58.9 (55.2-62.6) |
| **South Africa** | 9.0 (7.7-10.4) | 9.5 (8.4-10.6) | 10.6 (9.7-11.6) | 16.8 (15.3-18.4) | 13.2 (12.3-14.1) | 1.5 (1.0-2.1) | 1.6 (1.2-2.1) | 1.7 (1.4-2.1) | 4.4 (3.7-5.2) | 2.8 (2.5-3.2) | 54.6 (50.7-58.5) | 58.1 (54.9-61.3) | 61.2 (58.7-63.6) | 69.0 (65.8-72.0) | 64.3 (62.3-66.3) |
| **Tajikistan** | - | 38.9 (26.8-52.6) | 40.8 (32.1-50.0) | 32.4 (25.6-40.2) | 36.0 (30.7-41.6) | - | - | 2.2 (1.3-3.9) | - | 2.1 (1.2-3.5) | 22.1 (12.8-35.6) | 32.4 (29.4-35.7) | 31.9 (30.1-33.9) | 38.7 (37.1-40.3) | 35.1 (33.7-36.5) |
| **Tanzania** | 22.7 (20.2-25.4) | 24.9 (22.9-27.0) | 24.4 (22.3-26.7) | 26.0 (19.5-33.8) | 24.6 (22.5-26.8) | 2.4 (1.6-3.6) | 2.5 (1.9-3.2) | 2.3 (1.8-3.0) | - | 2.5 (1.9-3.2) | 14.6 (13.3-15.9) | 20.3 (19.0-21.6) | 24.6 (23.1-26.2) | 52.9 (48.8-56.9) | 27.4 (25.6-29.2) |
| **Timor-Leste** | 14.3 (8.9-22.1) | 18.2 (15.6-21.1) | 29.0 (21.7-37.7) | 22.1 (16.1-29.4) | 28.7 (21.4-37.2) | 1.8 (0.8-4.3) | 1.4 (0.6-3.2) | 1.2 (0.4-3.1) | - | 1.2 (0.1-2.2) | 5.2 (4.0-6.3) | 6.9 (6.2-7.6) | 8.9 (8.2-9.6) | 16.8 (13.6-20.0) | 9.8 (9.0-10.6) |
| **Togo** | 18.0 (15.4-20.9) | 19.1 (16.8-21.7) | 19.4 (17.3-21.8) | 13.9 (10.4-18.3) | 18.8 (16.9-21.0) | 2.0 (1.3-3.0) | 1.9 (1.4-2.7) | 2.2 (1.7-2.9) | - | 2.4 (1.9-3.1) | 15.2 (13.3-17.3) | 20.9 (19.0-22.9) | 26.7 (24.6-28.8) | 49.2 (44.2-54.1) | 29.4 (27.3-31.6) |
| **Tonga** | - | 41.7 (31.8-52.2) | 38.4 (34.4-42.5) | 33.7 (31.2-36.3) | 35.0 (32.5-37.5) | - | - | - | - | - | - | 92.4 (86.5-95.8) | 91.3 (88.7-93.3) | 93.2 (92.2-94.2) | 92.7 (91.8-93.5) |
| **Turkey** | - | - | - | - | - | - | - | - | - | - | - | 56.4 (49.5-63.1) | 55.8 (52.1-59.5) | 54.7 (53.5-56.0) | 54.9 (53.7-56.0) |
| **Tuvalu** | - | - | 51.3 (41.0-61.5) | 40.4 (28.0-54.2) | 44.4 (33.0-56.4) | - | - | - | - | 10.2 (8.9-11.8) | - | - | - | - | 86.6 (70.9-94.5) |
| **Uganda** | 20.1 (16.6-24.1) | 19.2 (16.6-22.2) | 20.7 (18.2-23.4) | 27.8 (20.6-36.3) | 21.7 (19.3-24.3) | - | - | - | - | 1.6 (0.9-3.0) | 9.4 (8.1-10.9) | 14.5 (13.2-16.0) | 18.5 (16.9-20.2) | 44.4 (40.3-48.6) | 22.6 (20.9-24.5) |
| **Uruguay** | - | - | - | - | - | - | - | - | - | | | | | | |
| **Vanuatu** | 29.5 (24.4-35.1) | 25.8 (22.2-29.8) | 24.3 (21.3-27.6) | 26.4 (22.7-30.4) | 24.9 (22.4-27.6) | 14.3 (10.2-19.5) | 11.9 (8.8-15.9) | 10.2 (7.9-13.1) | 7.4 (5.8-9.3) | 9.4 (7.6-11.6) | 29.6 (24.9-34.8) | 37.4 (33.6-41.2) | 42.9 (39.5-46.4) | 59.5 (55.7-63.1) | 47.4 (44.2-50.6) |
| **Vietnam** | - | - | - | - | - | - | - | - | - | - | - | - | - | - | - |
| **Yemen** | - | - | - | - | - | - | - | - | - | - | 6.4 (5.4-7.6) | 11.8 (10.8-13.0) | 17.9 (16.8-19.2) | 37.4 (35.6-39.2) | 22.6 (21.2-24.0) |
| **Zambia** | 18.4 (16.0-21.1) | 17.8 (15.7-20.1) | 18.4 (16.4-20.6) | 27.1 (19.1-37.0) | 19.4 (17.3-21.7) | 6.3 (4.9-8.0) | 6.0 (4.8-7.5) | 5.9 (4.7-7.3) | - | 5.7 (4.6-7.1) | 16.3 (14.2-18.6) | 18.5 (16.3-21.0) | 21.1 (18.8-23.7) | 37.9 (30.6-45.7) | 23.1 (20.8-25.5) |

| | | | | | | | | | | | | | | | |
|---|---|---|---|---|---|---|---|---|---|---|---|---|---|---|---|
| **Zanzibar** | 27.9 (23.6-32.6) | 28.8 (24.9-33.0) | 29.2 (25.4-33.3) | 28.3 (20.8-37.1) | 29.1 (25.4-33.1) | 2.8 (1.7-4.7) | 3.2 (2.2-4.7) | 3.4 (2.4-4.8) | - | 3.2 (2.2-4.5) | 33.0 (28.7-37.5) | 35.1 (31.5-38.8) | 36.4 (33.1-39.8) | 30.0 (22.7-38.6) | 35.9 (32.8-39.1) |
| **Zimbabwe** | - | - | - | - | - | - | - | - | - | - | 21.0 (19.2-22.9) | 25.2 (23.7-26.8) | 30.2 (28.7-31.8) | 49.5 (46.6-52.3) | 33.8 (32.1-35.5) |

[1] We excluded countries for which either the sample with non-missing data for the respective risk factor was <100 or samples in which a risk factor was prevalent among less than 20 respondents.
[2] Country has sampling weight=1.

Table S24: Prevalences for each risk factor by country (cont.)[1]

| | Obesity | | | | | Smoking | | | | | Dyslipidemia | | | | |
|---|---|---|---|---|---|---|---|---|---|---|---|---|---|---|---|
| **Country** | **<$1.90/day** | **<$3.20/day** | **<$5.50/day** | **>$5.50/day** | **total** | **<$1.90/day** | **<$3.20/day** | **<$5.50/day** | **>$5.50/day** | **total** | **<$1.90/day** | **<$3.20/day** | **<$5.50/day** | **>$5.50/day** | **total** |
| **Albania** | 10.4 (6.0-17.5) | 17.2 (14.5-20.2) | 20.3 (18.9-21.8) | 21.6 (20.3-23.0) | 21.3 (20.2-22.4) | - | - | - | - | - | - | - | - | - | - |
| **Angola** | - | - | - | - | - | 6.6 (5.6-7.7) | 5.9 (5.1-6.7) | 5.1 (4.4-5.8) | 2.0 (1.3-3.1) | 4.4 (3.9-5.0) | - | - | - | - | - |
| **Armenia** | - | 15.7 (12.2-19.9) | 15.2 (13.6-17.0) | 13.0 (11.6-14.5) | 13.8 (12.6-14.9) | - | - | - | - | - | - | - | - | - | - |
| **Bangladesh** | 1.4 (0.9-2.1) | 2.8 (2.5-3.2) | 4.6 (4.2-5.0) | 16.9 (15.3-18.7) | 6.3 (5.8-6.9) | 22.8 (19.8-26.2) | 19.1 (17.7-20.5) | 17.5 (16.5-18.5) | 12.3 (9.7-15.5) | 16.8 (15.8-17.8) | - | - | - | - | - |
| **Benin** | 5.4 (4.0-7.2) | 5.8 (4.7-7.3) | 6.4 (5.2-7.8) | 11.1 (6.2-19.2) | 6.9 (5.6-8.6) | 4.2 (3.2-5.5) | 4.6 (3.7-5.8) | 4.9 (4.0-5.9) | 5.2 (2.5-10.3) | 4.9 (4.0-5.9) | 1.8 (1.3-2.6) | 2.6 (1.9-3.5) | 2.7 (2.0-3.6) | 6.8 (3.6-12.6) | 3.1 (2.4-4.2) |
| **Bhutan** | - | 4.4 (2.6-7.6) | 4.8 (3.5-6.6) | 6.6 (5.3-8.2) | 6.0 (4.9-7.2) | - | - | 4.9 (3.0-7.8) | 10.1 (8-12.7) | 8.2 (6.5-10.2) | - | - | 1.4 (0.8-2.3) | 1.4 (0.8-2.3) | 1.4 (0.9-2.0) |
| **Bolivia** | 11.6 (9.4-14.1) | 12.3 (10.9-13.9) | 15.7 (14.4-17) | 25.4 (24.4-26.4) | 22.1 (21.2-22.9) | 2.2 (1.5-3.3) | 1.7 (1.2-2.5) | 1.8 (1.4-2.3) | 4.5 (3.9-5.2) | 3.6 (3.1-4.1) | - | - | - | - | - |
| **Botswana** | 8.0 (5.4-11.8) | 7.4 (5.6-9.6) | 7.9 (6.4-9.7) | 13.3 (11.3-15.7) | 10.4 (9.1-11.8) | 18.1 (14.8-21.9) | 17.3 (15.0-19.9) | 15.9 (13.9-18.1) | 11.7 (9.8-13.8) | 14.0 (12.6-15.6) | - | 1.4 (0.8-2.5) | 1.1 (0.7-1.9) | 3.0 (1.9-4.9) | 2.0 (1.3-3.0) |
| **Brazil** | 8.3 (6.4-10.7) | 11.8 (10.4-13.5) | 14.7 (13.6-15.8) | 20.6 (19.9-21.3) | 19.7 (19.1-20.4) | 24.1 (20.7-27.9) | 23.9 (21.7-26.2) | 21.9 (20.5-23.3) | 13.4 (12.8-14.0) | 14.6 (14.1-15.1) | - | - | - | - | - |
| **Burkina Faso** | 3.7 (3.0-4.4) | 4.2 (3.6-4.9) | 5.1 (4.5-5.8) | 15.8 (12.5-19.8) | 5.9 (5.2-6.8) | - | - | - | - | - | - | - | - | - | - |
| **Burundi** | - | 0.7 (0.5-1.0) | 1.2 (0.9-1.6) | 17.3 (13.0-22.7) | 1.6 (1.2-2.1) | - | - | - | - | - | - | - | - | - | - |
| **Cabo Verde** | - | 7.4 (4.4-12.2) | 8.1 (5.3-12.2) | 11.7 (7.0-18.7) | 10.2 (6.9-14.9) | - | 9.0 (4.2-18.1) | 10.1 (5.6-17.8) | 8.0 (6.4-9.8) | 8.9 (6.3-12.3) | - | - | - | - | 3.5 (1.4-8.8) |
| **Cameroon** | 2.5 (1.7-3.6) | 4.7 (3.8-5.8) | 8.4 (7.2-9.9) | 21.0 (18.6-23.7) | 12.6 (11.2-14.3) | - | - | - | - | 0.2 (0.1-0.3) | - | - | - | - | - |
| **Chad** | 0.9 (0.7-1.3) | 0.9 (0.7-1.2) | 1.2 (1.0-1.6) | 7.9 (6.5-9.6) | 2.1 (1.8-2.6) | 9.0 (7.8-10.4) | 7.0 (6.1-8.1) | 6.1 (5.3-7.0) | 4.7 (3.9-5.7) | 5.9 (5.2-6.7) | - | - | - | - | - |
| **China** | 3.7 (2.5-5.3) | 3.7 (2.9-4.8) | 3.7 (3.1-4.4) | 3.8 (3.2-4.6) | 3.8 (3.3-4.3) | 27.7 (21.7-34.7) | 28.7 (25.2-32.5) | 28.4 (26.1-30.8) | 27.0 (24.9-29.1) | 27.6 (26.0-29.4) | - | - | - | - | - |
| **Comoros** | 15.0 (11.5-19.3) | 14.7 (12.6-17.2) | 16.5 (14.5-18.6) | 19.4 (17.1-21.9) | 17.5 (15.9-19.2) | - | - | - | - | - | - | - | - | - | - |
| **Congo** | 5.7 (4.6-7.1) | 7.9 (6.2-10.1) | 13.7 (11.3-16.4) | 24.4 (20.4-28.9) | 17.2 (14.9-19.8) | - | - | - | - | - | - | - | - | - | - |

| | | | | | | | | | | | | | | | |
|---|---|---|---|---|---|---|---|---|---|---|---|---|---|---|---|
| **Congo Democratic Republic** | 1.1 (0.6-1.8) | 2.2 (1.7-3.0) | 2.9 (2.3-3.7) | 13.4 (8.6-20.1) | 3.2 (2.5-4.0) | 10.5 (9.5-11.6) | 9.7 (8.8-10.6) | 9.1 (8.3-10.0) | - | 9.0 (8.2-9.9) | - | - | - | - | - |
| **Costa Rica** | - | - | - | - | - | - | 11.5 (7.3-17.6) | 9.6 (7.0-13.0) | 9.0 (8.1-10.0) | 9.0 (8.1-10.0) | - | - | - | - | - |
| **Côte d’Ivoire** | 15.8 (12.6-19.5) | 17.0 (14.6-19.6) | 19.6 (17.5-21.8) | 31.0 (25.6-37.0) | 22.2 (20.0-24.4) | - | - | - | - | - | - | - | - | - | - |
| **Dominican Republic** | - | 13.7 (10.3-17.9) | 18.3 (16.4-20.4) | 20.4 (19.2-21.6) | 19.9 (18.9-21.0) | - | - | - | - | 0.2 (0.2-0.4) | - | - | - | - | - |
| **Ecuador** | 24.1 (15.8-35.1) | 23.3 (18.5-29.0) | 22.4 (19.2-25.8) | 24.5 (22.6-26.4) | 24.0 (22.4-25.8) | - | 11.7 (8.3-16.1) | 13.8 (11.1-17.0) | 14.6 (13.0-16.5) | 14.4 (13.0-16.1) | - | 10.7 (7.7-14.8) | 10.9 (8.7-13.7) | 10.4 (9.1-11.8) | 10.5 (9.4-11.7) |
| **Egypt** | 35.7 (26.4-46.4) | 44.0 (41.7-46.4) | 46.6 (45.0-48.2) | 47.3 (44.9-49.6) | 46.8 (45.3-48.2) | 12.9 (8.3-19.5) | 20.2 (18.5-22.0) | 20.5 (19.4-21.5) | 22.9 (21.4-24.5) | 21.1 (20.3-22.0) | - | - | - | - | - |
| **Eswatini**[2] | 18.0 (14.9-21.6) | 18.7 (16.0-21.6) | 18.0 (15.8-20.4) | 18.2 (15.1-21.8) | 18.0 (16.0-20.0) | 7.1 (5.0-10.1) | 6.0 (4.4-8.0) | 5.9 (4.6-7.5) | 5.4 (3.3-8.6) | 5.8 (4.5-7.0) | 1.6 (0.8-3.3) | 1.8 (1.2-2.9) | 1.6 (1.1-2.4) | 1.7 (0.9-3.1) | 1.6 (1.2-2.2) |
| **Ethiopia** | 0.3 (0.1-0.5) | 0.5 (0.3-0.7) | 1.2 (0.8-1.8) | 8.3 (6.1-11.1) | 1.4 (1.0-2.1) | 3.3 (1.9-5.7) | 3.5 (2.4-5.1) | 3.4 (2.4-4.8) | 3.1 (1.7-5.7) | 3.4 (2.5-4.7) | 0.6 (0.3-1.0) | 0.8 (0.5-1.3) | 0.8 (0.5-1.2) | - | 0.8 (0.6-1.2) |
| **Gabon** | - | 5.7 (3.8-8.4) | 9.1 (7.2-11.5) | 22.5 (20.5-24.7) | 21.3 (19.4-23.4) | - | - | - | - | - | - | - | - | - | - |
| **Gambia** | 16.6 (13.2-20.8) | 16.9 (14.7-19.3) | 20.8 (18.5-23.3) | 30.2 (25.6-35.1) | 23.7 (21.4-26.1) | 11.2 (9.5-13.0) | 8.3 (7.3-9.5) | 8.2 (7.3-9.1) | 6.8 (5.4-8.5) | 7.7 (7.0-8.5) | - | - | - | - | - |
| **Georgia**[2] | 23.0 (15.2-33.3) | 27.8 (22.7-33.6) | 31.8 (28.4-35.4) | 30.0 (27.0-33.1) | 30.6 (28.1-33.1) | 39.2 (26.7-53.3) | 35.6 (29.1-42.8) | 30.5 (26.4-34.9) | 32.1 (28.6-35.7) | 31.5 (28.7-34.2) | - | - | - | - | - |
| **Ghana**[2] | - | 2.2 (1.4-3.5) | 4.8 (3.8-6.0) | 21.8 (19.5-24.3) | 14.4 (12.8-15.9) | 6.7 (4.9-9.1) | 5.1 (4.1-6.5) | 3.7 (3.1-4.5) | 0.9 (0.6-1.4) | 2.1 (1.8-2.5) | - | - | - | - | - |
| **Guatemala** | 8.2 (6.9-9.8) | 9.9 (8.9-11) | 13.4 (12.6-14.3) | 24.1 (23.1-25.2) | 18.9 (18.0-19.7) | - | - | - | - | - | - | - | - | - | - |
| **Guinea** | 3.1 (2.3-4.1) | 5.4 (4.6-6.4) | 7.1 (6.1-8.1) | 15.3 (12.1-19.3) | 8.0 (7.0-9.1) | - | - | 0.6 (0.3-1.0) | - | 0.5 (0.3-0.9) | - | - | - | - | - |
| **Haiti** | 2.9 (2.2-3.9) | 4.0 (3.4-4.7) | 7.4 (6.7-8.2) | 17.7 (15.8-19.8) | 10.6 (9.6-11.6) | - | - | - | - | - | - | - | - | - | - |
| **Honduras** | 15.8 (14.3-17.4) | 17.0 (15.9-18.2) | 20.6 (19.6-21.7) | 34.0 (32.8-35.2) | 28.6 (27.7-29.6) | - | - | - | - | - | - | - | - | - | - |
| **India** | 1.0 (0.9-1.1) | 2.1 (2-2.2) | 3.9 (3.8-4) | 10.7 (10.4-11.1) | 4.9 (4.8-5.0) | 12.8 (11.8-13.9) | 11.2 (10.5-11.8) | 10.0 (9.5-10.5) | 5.7 (4.8-6.6) | 9.5 (9.0-9.9) | - | - | - | - | - |
| **Indonesia** | 4.0 (2.6-6.1) | 5.5 (4.9-6.3) | 6.9 (6.1-7.8) | 10.3 (9.4-11.2) | 8.1 (7.2-9.0) | 90.4 (86.3-93.3) | 90.2 (87.4-92.3) | 89.4 (87.3-91.2) | 85.6 (82.7-88.1) | 88.3 (86.3-90.1) | - | - | - | - | - |
| **Iran** | - | 17.8 (14.4-21.7) | 14.7 (13.3-16.1) | 22.2 (21.6-22.8) | 21.3 (20.7-21.9) | - | 7.6 (5.3-10.8) | 14.5 (13.1-16.1) | 14.1 (13.6-14.6) | 14.2 (13.7-14.7) | - | - | - | - | - |
| **Jordan** | - | 24.6 (18.6-31.6) | 24.7 (22.0-27.8) | 19.3 (17.9-20.8) | 19.9 (18.6-21.3) | - | 29.0 (26.6-31.6) | 30.4 (28.9-31.9) | 32.1 (31.3-32.9) | 31.9 (31.2-32.6) | - | - | - | 1.6 (1.1-2.2) | 1.6 (1.2-2.2) |

| | | | | | | | | | | | | | | | |
|---|---|---|---|---|---|---|---|---|---|---|---|---|---|---|---|
| **Kenya** | 2.7 (2.2-3.3) | 4.7 (4.2-5.2) | 6.9 (6.3-7.6) | 19.7 (17.5-22.2) | 9.5 (8.8-10.3) | - | - | - | - | - | - | - | - | - | - |
| **Kiribati** | 51.6 (36.1-66.8) | 47.6 (38.8-56.6) | 44.1 (35.9-52.6) | 49.9 (41.4-58.4) | 45.8 (40.8-50.8) | 42.2 (32.6-52.5) | 54.1 (47.0-61.0) | 52.4 (46.8-57.9) | 40.2 (31.3-49.8) | 48.1 (43.0-53.2) | - | - | 4.1 (1.3-12.0) | - | 5.4 (2.5-11.5) |
| **Kyrgyz Republic** | 17.6 (13.5-22.7) | 16.3 (13.6-19.4) | 14.2 (12.8-15.8) | 12.6 (10.8-14.7) | 13.5 (12.3-14.8) | 10.7 (6.5-17.0) | 9.8 (8.3-11.4) | 9.7 (8.9-10.6) | 15.2 (13.8-16.7) | 12.1 (11.2-13.1) | - | - | 2.0 (1.4-2.7) | 4.1 (2.9-5.8) | 2.9 (2.2-3.6) |
| **Laos** | - | 2.7 (1.7-4.2) | 4.0 (3.0-5.2) | 8.2 (5.6-12.1) | 5.0 (4.0-6.3) | 37.9 (26.6-50.7) | 38.5 (31.2-46.3) | 34.6 (28.0-41.8) | 27.5 (19.6-37.2) | 33.1 (27.2-39.6) | - | 2.3 (1.5-3.4) | 3.1 (2.3-4.2) | 7.0 (4.8-10.2) | 4.0 (3.1-5.2) |
| **Lesotho** | 5.2 (4.2-6.6) | 6.9 (5.8-8.1) | 8.6 (7.6-9.7) | 17.1 (14.9-19.5) | 11.1 (10.1-12.1) | 22.8 (20.8-24.9) | 22.8 (21.1-24.6) | 21.8 (20.4-23.3) | 15.9 (13.9-18.1) | 20.1 (18.9-21.4) | - | - | - | - | - |
| **Liberia** | 25.1 (21.9-28.5) | 24.3 (21.5-27.4) | 23.9 (21.0-27.0) | - | 24.0 (21.1-27.1) | 10.0 (8.2-12.1) | 9.1 (7.6-10.9) | 8.8 (7.3-10.5) | - | 8.8 (7.4-10.5) | - | - | - | - | - |
| **Madagascar** | 7.5 (6.6-8.5) | 7.7 (6.8-8.6) | 8.1 (7.2-9.0) | 10.9 (6.5-17.7) | 8.1 (7.2-9.1) | 0.7 (0.5-1.0) | 1.1 (0.8-1.4) | 1.2 (0.9-1.6) | - | 1.3 (1.0-1.7) | - | - | - | - | - |
| **Malawi** | 3.1 (2.6-3.8) | 3.9 (3.4-4.6) | 4.8 (4.1-5.4) | 20.8 (15.6-27.2) | 5.3 (4.7-6.1) | 37.6 (35.7-39.6) | 38.3 (36.5-40.1) | 38.3 (36.5-40.1) | 40.9 (34.2-47.9) | 38.4 (36.6-40.2) | - | - | - | - | - |
| **Mexico** | 25.3 (12.6-38.0) | 23.2 (15.5-30.8) | 22.5 (16.8-28.1) | 27.1 (25.6-28.6) | 25.8 (23.4-28.1) | 8.6 (4.6-12.5) | 7.8 (5.4-10.2) | 11.6 (9.8-13.4) | 18.4 (17.2-19.6) | 17.3 (16.2-18.3) | 9.3 (4.1-14.5) | 12.5 (7.9-17.2) | 9.3 (7.1-11.5) | 11.4 (10.1-12.7) | 10.8 (9.1-12.5) |
| **Mongolia** | - | 18.8 (12.6-27) | 17.3 (14.8-20.1) | 18.9 (17.5-20.5) | 18.5 (17.2-20.0) | | 20.9 (14.7-28.7) | 22.5 (19.8-25.4) | 26.3 (24.5-28.2) | 25.4 (23.8-27.0) | | 5.2 (2.7-9.7) | 4.0 (3.0-5.3) | 5.5 (4.7-6.5) | 5.2 (4.4-6.0) |
| **Morocco** | - | - | - | - | - | - | - | - | - | - | - | - | - | - | - |
| **Mozambique** | 2.6 (2.1-3.1) | 4.2 (3.7-4.9) | 5.1 (4.4-5.8) | 21.6 (18.5-25.2) | 5.7 (5.0-6.4) | 0.9 (0.6-1.2) | 0.7 (0.5-1.0) | 0.7 (0.5-1.0) | - | 0.7 (0.5-1.0) | - | - | - | - | - |
| **Myanmar** | | 2.6 (2.0-3.6) | 3.8 (3.2-4.4) | 7.3 (6.4-8.3) | 5.2 (4.6-5.9) | 9.0 (6.2-13.0) | 5.8 (4.4-7.4) | 3.0 (2.3-3.8) | 0.3 (0.1-0.5) | 1.8 (1.4-2.4) | - | 39.6 (28.3-52.2) | 35.5 (27.6-44.3) | 48.6 (40.8-56.5) | 41.6 (35.6-47.9) |
| **Namibia** | 10.7 (8.7-13.1) | 12.8 (11.2-14.7) | 15.9 (14.3-17.6) | 36.6 (34.1-39.2) | 25.8 (23.9-27.9) | 11.2 (9.4-13.2) | 11.0 (9.6-12.6) | 10.3 (9.2-11.6) | 9.9 (8.7-11.2) | 10.1 (9.2-11.1) | - | - | - | - | - |
| **Niger** | 12.6 (11.0-14.5) | 12.9 (11.4-14.4) | 14.4 (13.0-16.0) | 33.0 (26.0-41.0) | 14.9 (13.4-16.4) | 4.5 (3.8-5.3) | 5.0 (4.4-5.7) | 5.3 (4.7-6.0) | 9.7 (6.8-13.6) | 5.4 (4.8-6.1) | - | - | - | - | - |
| **Nigeria** | 2.8 (2.3-3.3) | 5.2 (4.6-5.8) | 7.7 (7.1-8.3) | 25.0 (21.8-28.4) | 9.3 (8.6-10.0) | - | - | - | - | - | - | - | - | - | - |
| **Pakistan**[2] | - | 10.2 (8.2-12.5) | 16.8 (14.6-19.2) | 33.0 (28.8-37.6) | 20.5 (18.2-22.8) | - | - | - | - | - | - | - | - | - | - |
| **Panama** | - | - | - | - | - | 6.4 (3.0-13.0) | 5.8 (3.8-8.6) | 5.6 (4.1-7.5) | 6.1 (4.6-8.1) | 6.1 (4.6-8.0) | - | - | - | - | - |
| **Peru** | 8.8 (6.8-11.3) | 10.0 (8.5-11.7) | 11.8 (10.7-12.9) | 23.7 (22.7-24.7) | 21.5 (20.6-22.4) | - | - | - | - | - | - | - | - | - | - |
| **Philippines** | - | - | - | - | - | 27.1 (22.7-32.1) | 28.4 (26.0-30.9) | 26.3 (24.7-28.0) | 18.8 (17.4-20.3) | 22.3 (21.2-23.4) | - | - | - | - | - |

| | | | | | | | | | | | | | | | |
|---|---|---|---|---|---|---|---|---|---|---|---|---|---|---|---|
| **Romania** | - | 25.2 (19.0-32.6) | 23.1 (18.9-28.0) | 31.0 (28.6-33.5) | 29.3 (27.2-31.5) | - | 31.0 (24.0-39.0) | 34.4 (29.2-40.0) | 28.1 (25.7-30.7) | 29.5 (27.2-31.8) | - | - | - | - | - |
| **Rwanda** | 1.2 (0.9-1.7) | 1.8 (1.4-2.3) | 2.9 (2.4-3.4) | 12.6 (9.8-15.9) | 3.6 (3.1-4.2) | 8.9 (8.3-9.6) | 7.6 (7.1-8.1) | 7.3 (6.8-7.7) | 2.6 (2.0-3.5) | 7.0 (6.5-7.4) | - | - | 0.4 (0.2-0.6) | - | 0.5 (0.3-0.8) |
| **Samoa** | - | 41.7 (36.3-47.4) | 50.9 (45.1-56.8) | 57.4 (52.4-62.2) | 55.5 (52.9-58.0) | - | 30.3 (19.0-44.5) | 26.6 (20.3-34.1) | 26.1 (21.5-31.3) | 26.3 (22.1-30.9) | - | - | - | 1.8 (1.0-3.2) | 2.1 (1.5-3.0) |
| **São Tomé and Principe** | 9.9 (7.4-13.1) | 9.3 (6.9-12.3) | 10.5 (7.9-13.8) | 15.2 (7.5-28.4) | 10.8 (7.9-13.8) | 8.5 (6.9-10.4) | 7.6 (6.5-8.9) | 6.9 (5.9-8.0) | | 6.9 (5.9-7.9) | - | - | - | - | 0.6 (0.3-1.4) |
| **Senegal** | 18.0 (14.9-21.6) | 17.4 (15.3-19.7) | 17.1 (15.3-19.0) | 23.8 (19.2-29.1) | 18.7 (16.9-20.6) | 6.7 (5.2-8.6) | 5.9 (4.8-7.3) | 5.5 (4.5-6.6) | 3.4 (2.0-5.8) | 5.2 (4.3-6.2) | - | - | - | - | - |
| **Solomon Islands** | 14.6 (10.9-19.2) | 20.1 (17.2-23.4) | 21.7 (19.0-24.7) | 31.1 (24.8-38.2) | 23.2 (20.5-26.1) | 43.8 (37.8-50.0) | 38.4 (34.4-42.6) | 38.0 (34.4-41.9) | 34.0 (28.0-40.5) | 37.3 (33.6-41.2) | 4.0 (2.5-6.6) | 5.5 (4.2-7.2) | 6.1 (4.9-7.7) | 9.8 (5.8-16.2) | 6.7 (5.5-8.2) |
| **South Africa** | 27.9 (24.2-31.8) | 30.3 (27.6-33.1) | 33.7 (31.4-36.2) | 44.0 (40.9-47.2) | 37.9 (35.9-39.9) | - | - | - | - | - | - | - | - | - | - |
| **Tajikistan** | - | 10.1 (8.3-12.2) | 10.5 (9.5-11.5) | 14.1 (12.9-15.2) | 12.2 (11.4-13.0) | - | - | - | - | - | - | - | - | 1.9 (0.7-4.9) | 1.4 (0.6-3.1) |
| **Tanzania** | 2.8 (2.3-3.4) | 5.4 (4.8-6.1) | 7.8 (7.0-8.0) | 24.7 (21.4-28.4) | 9.5 (8.5-10.6) | - | - | - | - | - | 1.3 (0.7-2.3) | 2.6 (1.3-5.3) | 3.3 (1.5-6.8) | - | 3.3 (1.6-6.7) |
| **Timor-Leste** | - | 1.0 (0.7-1.3) | 1.4 (1.2-1.7) | 3.4 (2.3-5.2) | 1.6 (1.3-1.9) | - | - | - | - | - | - | - | - | - | - |
| **Togo** | 3.4 (2.6-4.5) | 5.6 (4.8-6.6) | 8.8 (7.7-10.0) | 22.3 (18.4-26.9) | 10.5 (9.3-11.7) | 1.0 (0.7-1.5) | 0.6 (0.4-1.0) | 0.5 (0.3-0.8) | | 0.4 (0.3-0.7) | - | - | - | - | 0.9 (0.4-1.9) |
| **Tonga** | - | 76.0 (69.5-81.6) | 71.7 (68.7-74.4) | 78.6 (76.9-80.3) | 76.7 (74.9-78.4) | - | 24.1 (19.7-29.2) | 27.0 (22.9-31.5) | 25.2 (22.8-27.7) | 25.7 (23.9-27.6) | - | 10.8 (6.4-17.5) | 9.8 (6.9-13.6) | 9.4 (7.7-11.5) | 9.5 (7.6-11.9) |
| **Turkey** | - | 27.6 (22.0-34.0) | 29.7 (26.4-33.2) | 26.1 (25.0-27.2) | 26.5 (25.4-27.6) | - | - | - | - | - | - | - | - | - | - |
| **Tuvalu** | - | - | 65.1 (53.6-75.0) | 57.7 (50.1-64.9) | 60.3 (53.5-66.7) | - | - | 46.4 (34.6-58.7) | 25.9 (21.1-31.4) | 33.2 (24.0-43.8) | - | - | - | - | - |
| **Uganda** | 1.2 (0.8-2) | 2.7 (2.1-3.4) | 4.3 (3.6-5.2) | 18.3 (15.4-21.6) | 6.6 (5.7-7.6) | - | - | - | - | - | - | - | - | - | 1.0 (0.6-1.7) |
| **Uruguay** | - | - | - | - | - | - | - | 43.6 (34.6-53.1) | 26.8 (24.9-28.9) | 27.5 (25.6-29.5) | - | - | - | - | |
| **Vanuatu** | 8.6 (5.8-12.7) | 10.1 (8.1-12.7) | 13.8 (11.7-16.1) | 23.2 (20.1-26.6) | 16.3 (14.3-18.6) | 23.1 (18.3-28.8) | 26.0 (22.4-29.9) | 25.2 (22.5-28.1) | 26.3 (23.1-29.7) | 25.5 (23.2-27.9) | 8.7 (5.8-12.8) | 9.0 (6.9-11.7) | 8.1 (6.5-10.1) | 8.5 (6.4-11.3) | 8.2 (6.8-10.0) |
| **Vietnam** | - | - | - | - | - | 34.6 (26.7-43.4) | 28.8 (23.9-34.3) | 27.0 (24.3-29.9) | 20.4 (18.9-22.0) | 22.4 (21.0-23.8) | - | - | - | - | - |
| **Yemen** | 1.7 (1.2-2.4) | 3.1 (2.6-3.6) | 5.3 (4.8-5.8) | 14.8 (13.2-16.7) | 7.6 (6.8-8.4) | 17.3 (15.6-19.1) | 16.7 (15.6-17.8) | 16.1 (15.2-17.0) | 15.4 (14.2-16.6) | 15.9 (15.1-16.7) | - | - | - | - | - |
| **Zambia** | 3.6 (2.6-5.0) | 6.0 (4.6-7.7) | 6.9 (5.5-8.5) | 12.4 (8.4-17.8) | 7.5 (6.1-9.2) | 0.3 (0.2-0.5) | 0.3 (0.2-0.4) | 0.3 (0.2-0.4) | - | 0.3 (0.2-0.4) | 1.1 (0.6-2.0) | 1.0 (0.6-1.7) | 1.1 (0.7-1.7) | - | 1.3 (0.9-2.0) |

| | | | | | | | | | | | | | | | |
|---|---|---|---|---|---|---|---|---|---|---|---|---|---|---|---|
| **Zanzibar** | 14.9 (11.6-19.0) | 14.2 (11.5-17.5) | 14.8 (12.2-17.8) | 8.9 (4.6-16.5) | 14.4 (11.8-17.4) | 8.9 (6.2-12.6) | 8.1 (6.0-10.9) | 8.0 (6.0-10.5) | - | 7.7 (5.8-10.1) | 5.5 (3.9-7.8) | 4.7 (3.3-6.4) | 4.3 (3.1-6.0) | - | 4.2 (3.1-5.8) |
| **Zimbabwe** | 4.2 (3.4-5.0) | 6.4 (5.7-7.2) | 9.4 (8.5-10.3) | 23.0 (20.7-25.5) | 11.9 (10.9-13.0) | 8.6 (7.7-9.5) | 7.8 (7.1-8.5) | 7.4 (6.9-8.0) | 4.5 (3.8-5.4) | 6.9 (6.4-7.4) | - | - | - | - | - |

[1] We excluded countries for which either the sample with non-missing data for the respective risk factor was <100 or samples in which a risk factor was prevalent among less than 20 respondents.
[2] Country has sampling weight=1.

Figure S11: GDP per capita and poverty intensity among the moderately poor (<$3.20/day)

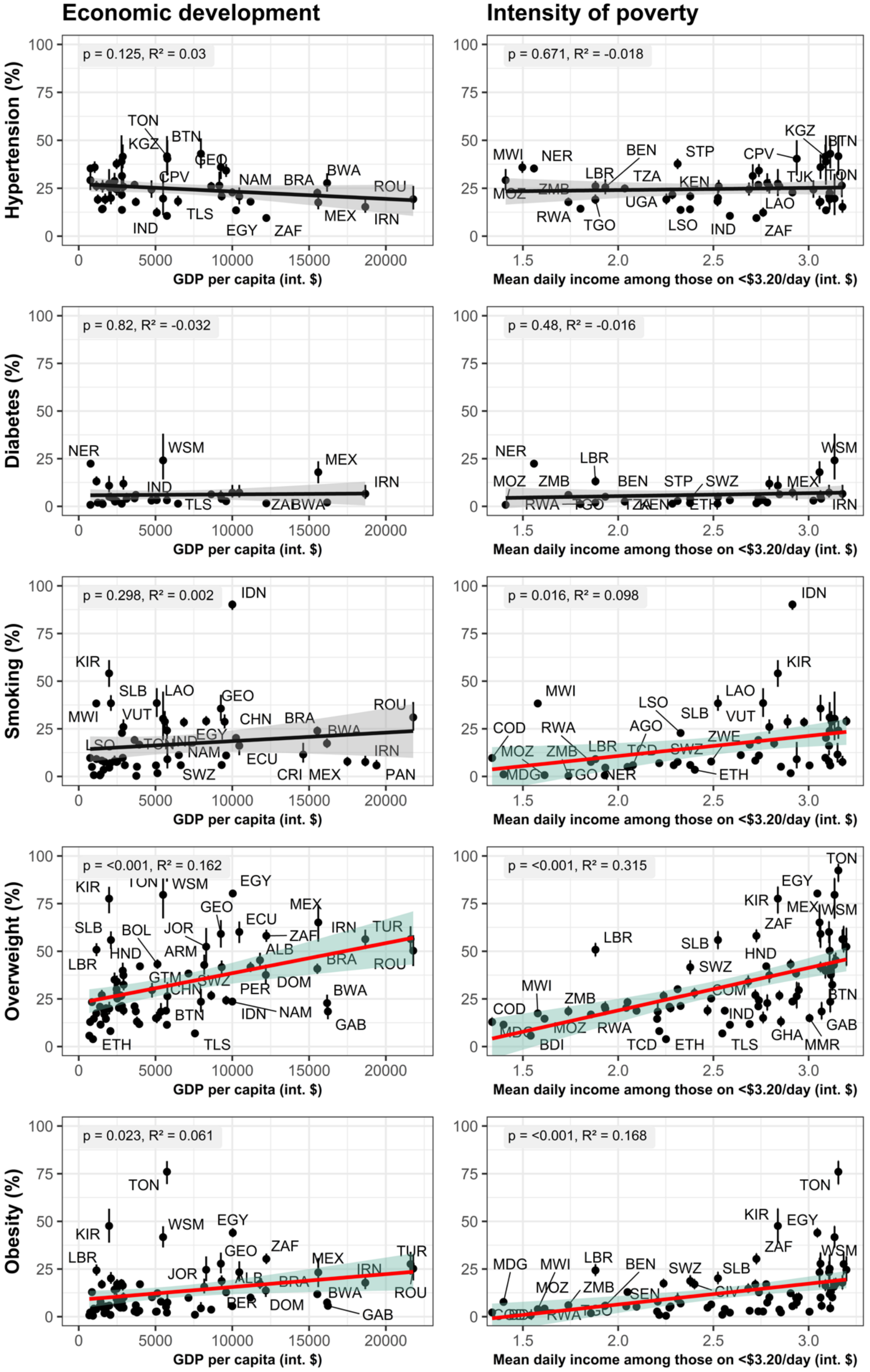

Figure S12: GDP per capita and poverty intensity among the moderately poor (<$5.50/day)

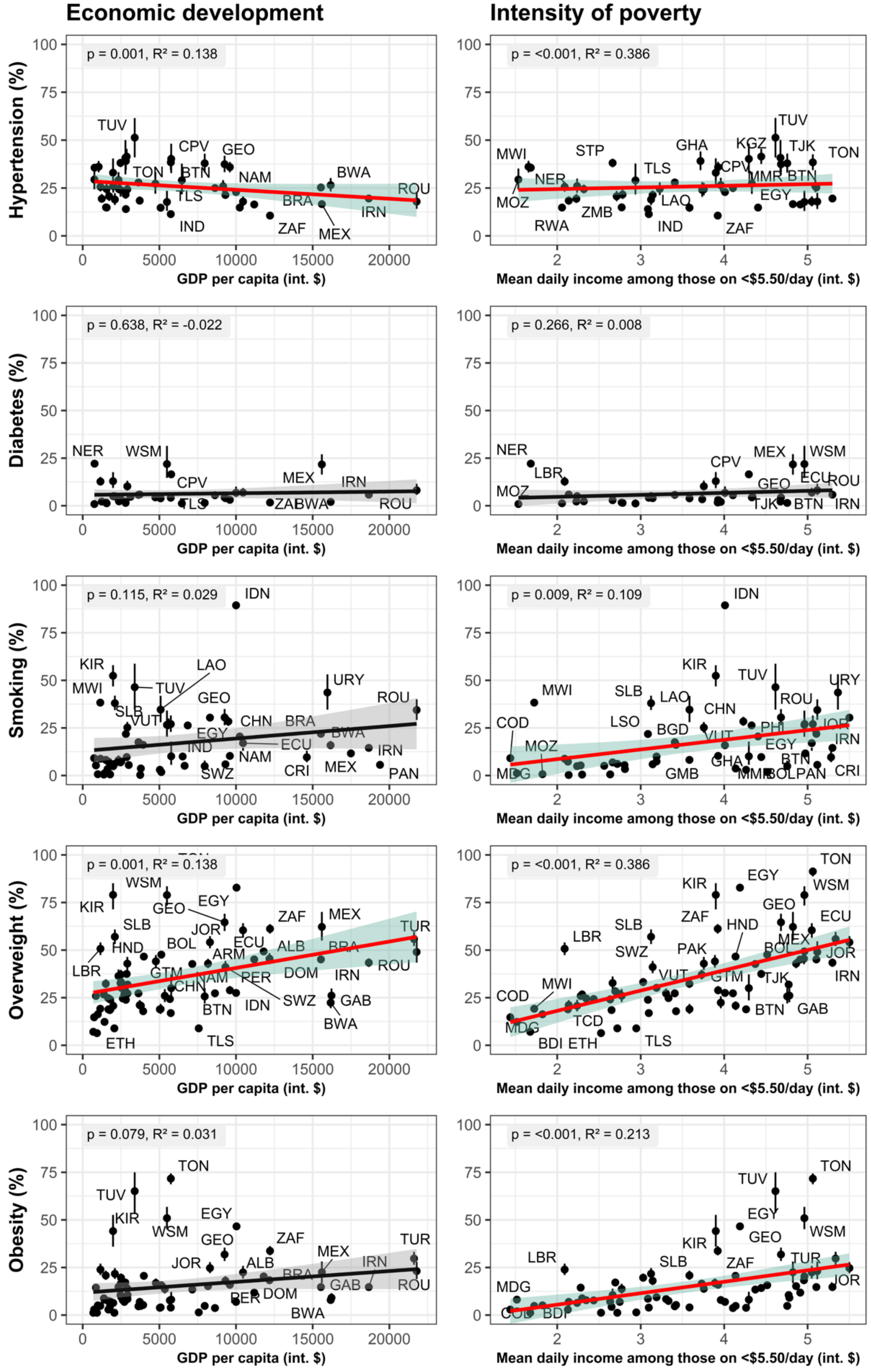

Figure S13: GDP per capita and poverty levels, adjusted for poverty intensity and age

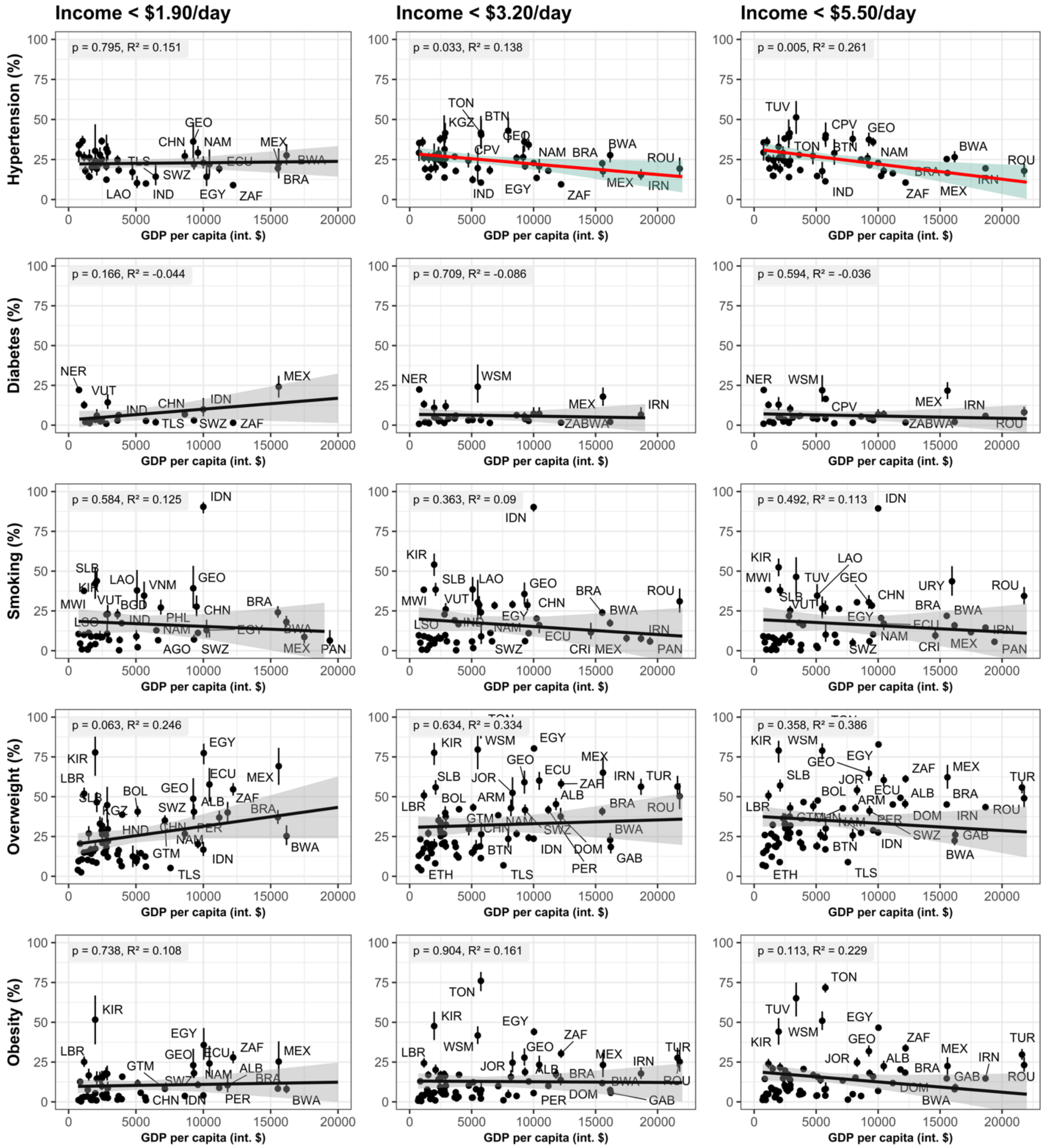

Figure S14: Variation in CVD risk factor prevalence among those living in extreme poverty (<$1.90/day) by rural-urban residency, sex, and education (absolute marginal effect)[1]

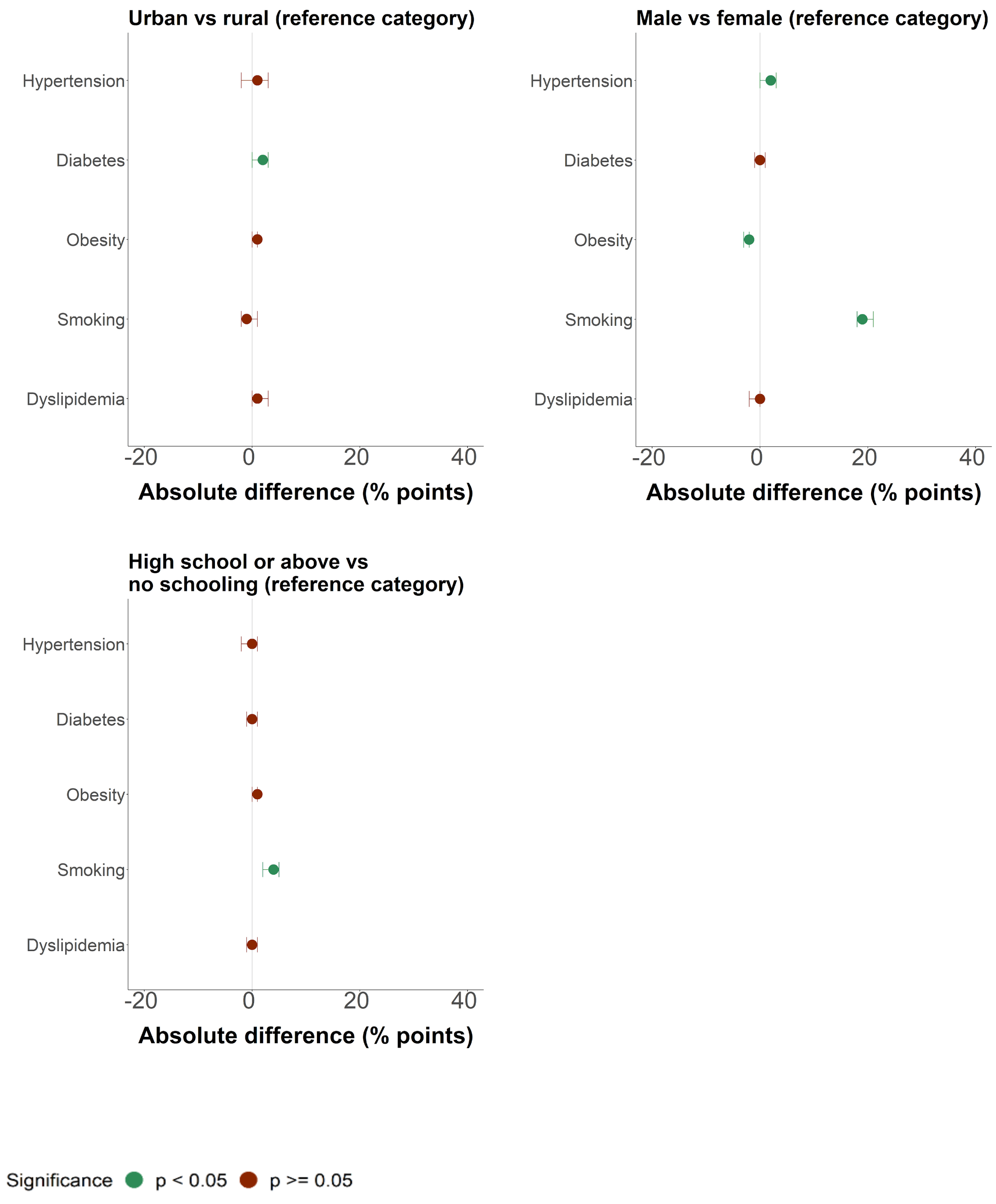

[1]Absolute difference (% points) is equivalent to absolute marginal effect (AME).
Data are point estimates with 95% CIs.

Figure S15: Variation in CVD risk factor prevalence among those living in moderate poverty (<$3.20/day) by rural-urban residency, sex, and education[1]

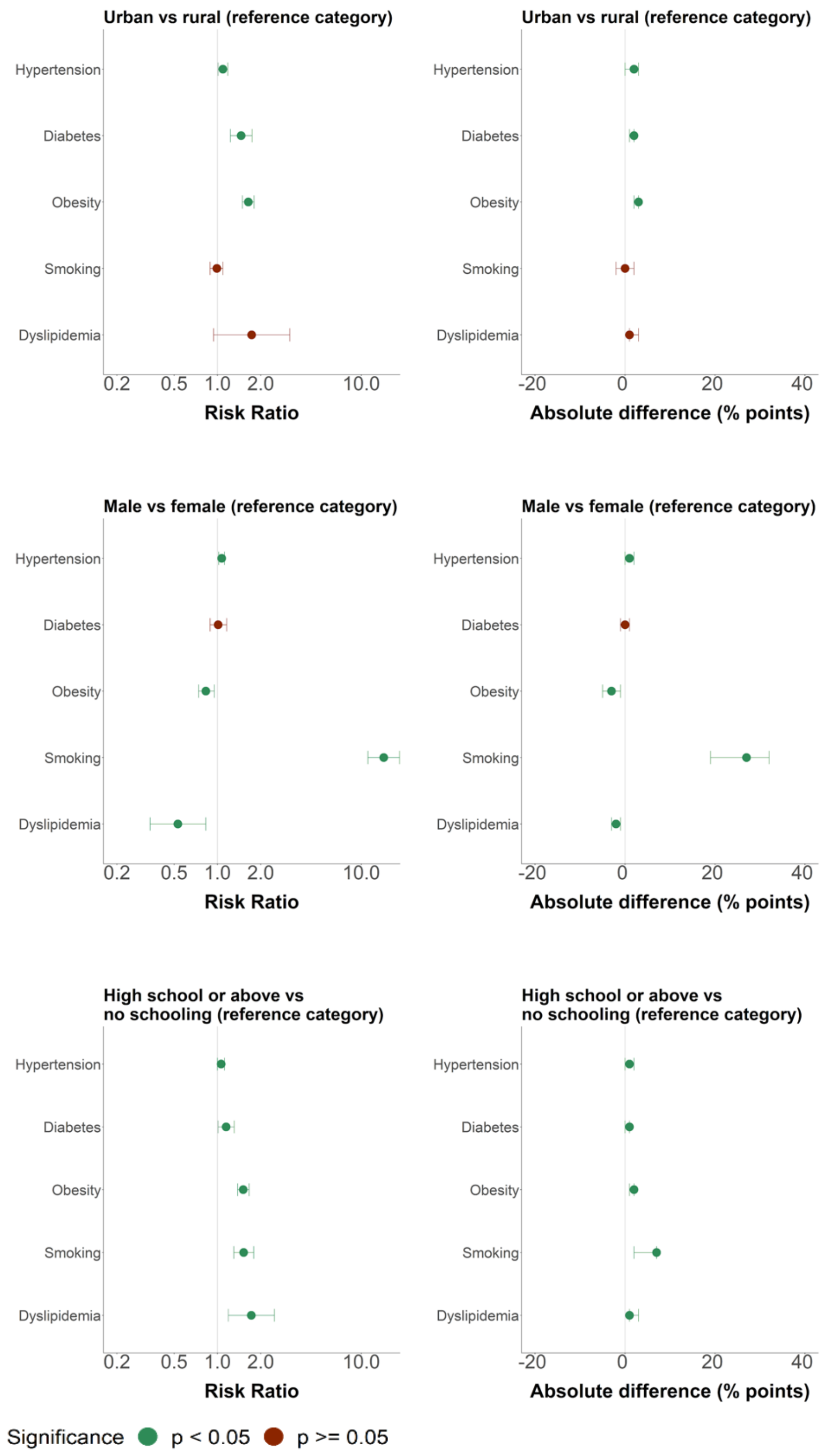

[1]Absolute difference (% points) is equivalent to absolute marginal effect (AME).
Data are point estimates with 95% CIs.

Figure S16: Variation in CVD risk factor prevalence among those living in moderate poverty (<$5.50/day) by rural-urban residency, sex, and education[1]

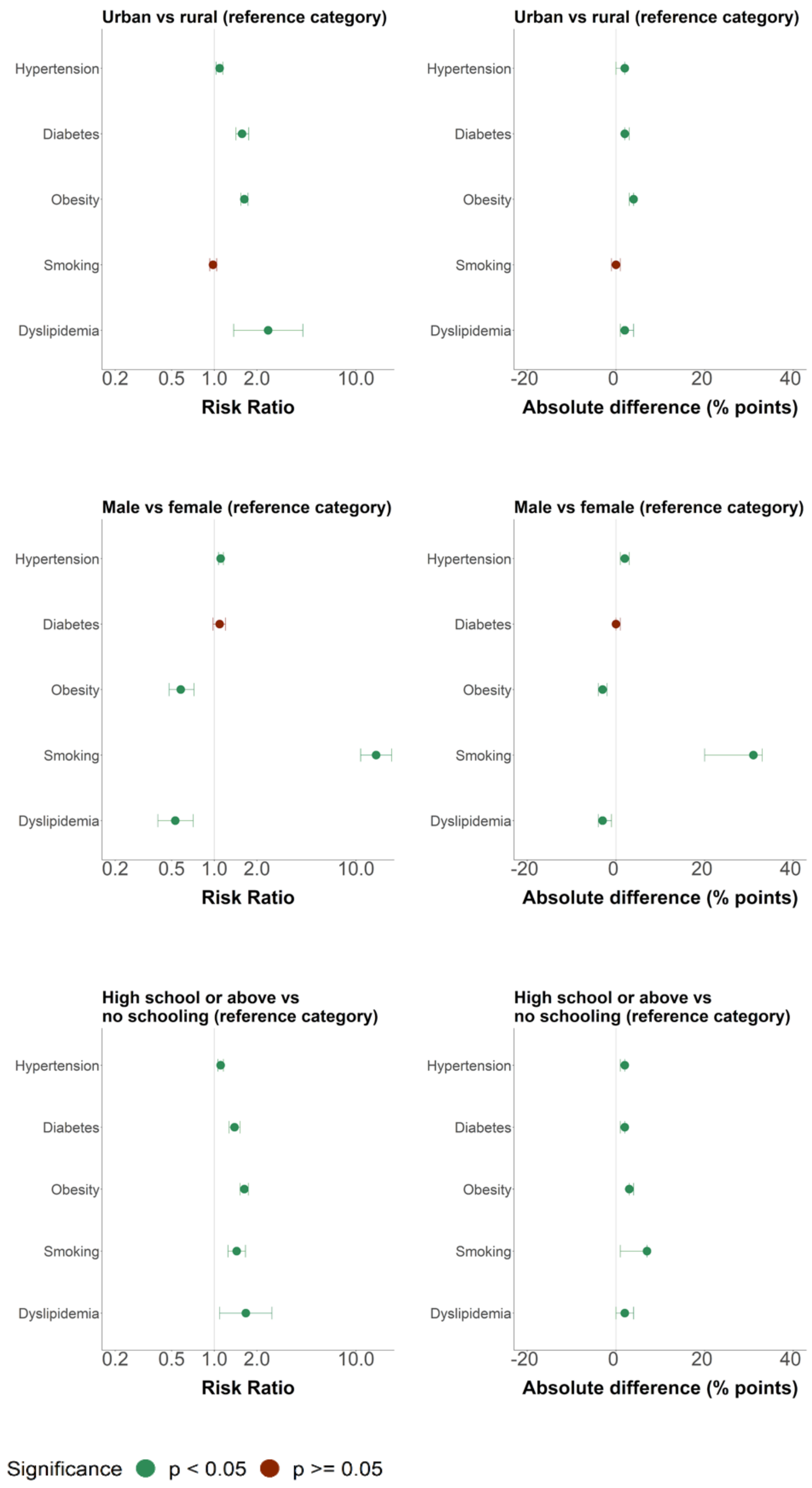

[1]Absolute difference (% points) is equivalent to absolute marginal effect (AME).
Data are point estimates with 95% CIs.

Figure S17: Variation in CVD risk factor prevalence among those living on >$5.50/day by rural-urban residency, sex, and education[1]

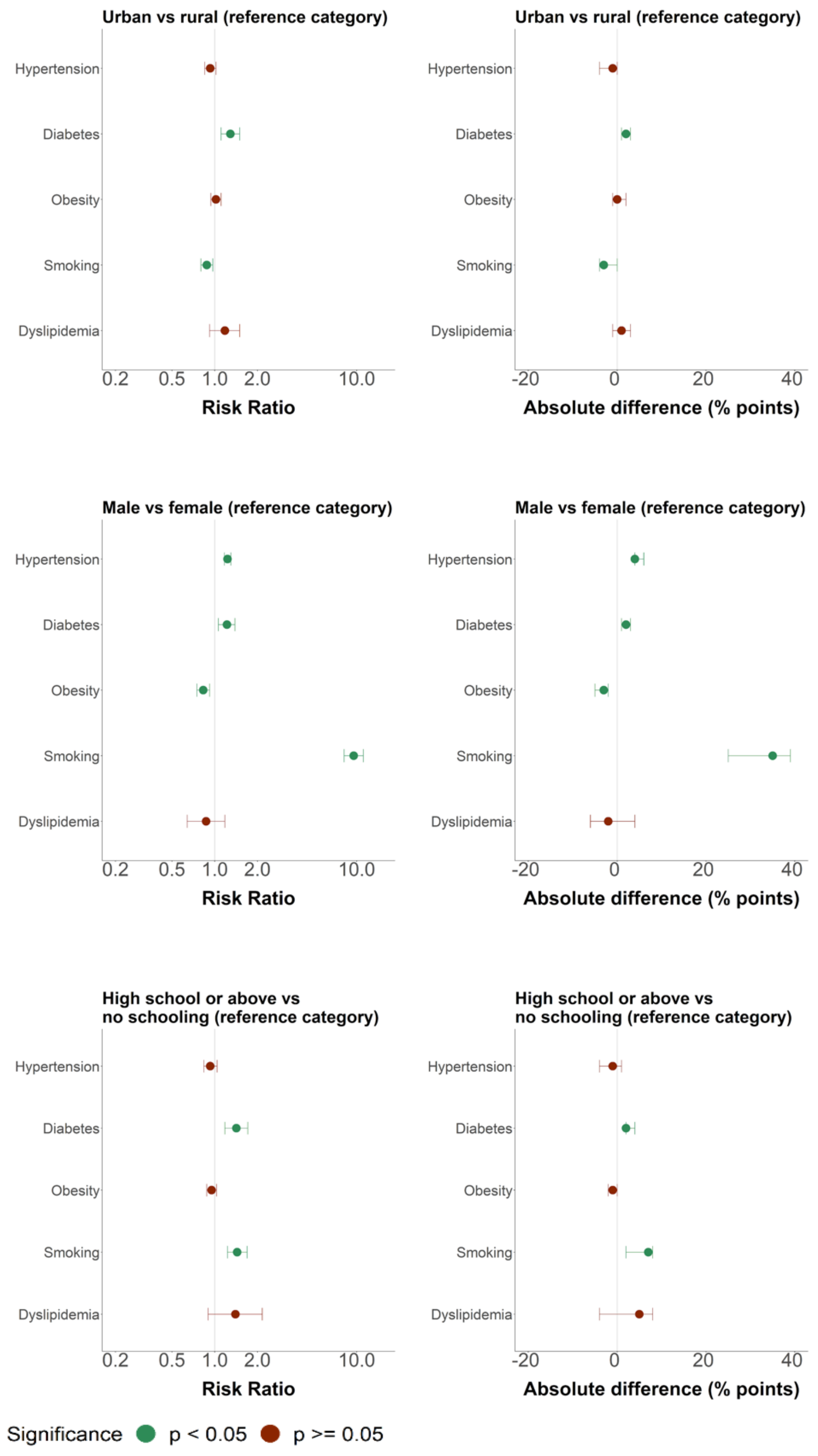

[1]Absolute difference (% points) is equivalent to absolute marginal effect (AME).
Data are point estimates with 95% CIs.

Figure S18: Variation in CVD risk factor prevalence by rural-urban residency, sex, and education, all poverty groups[1]

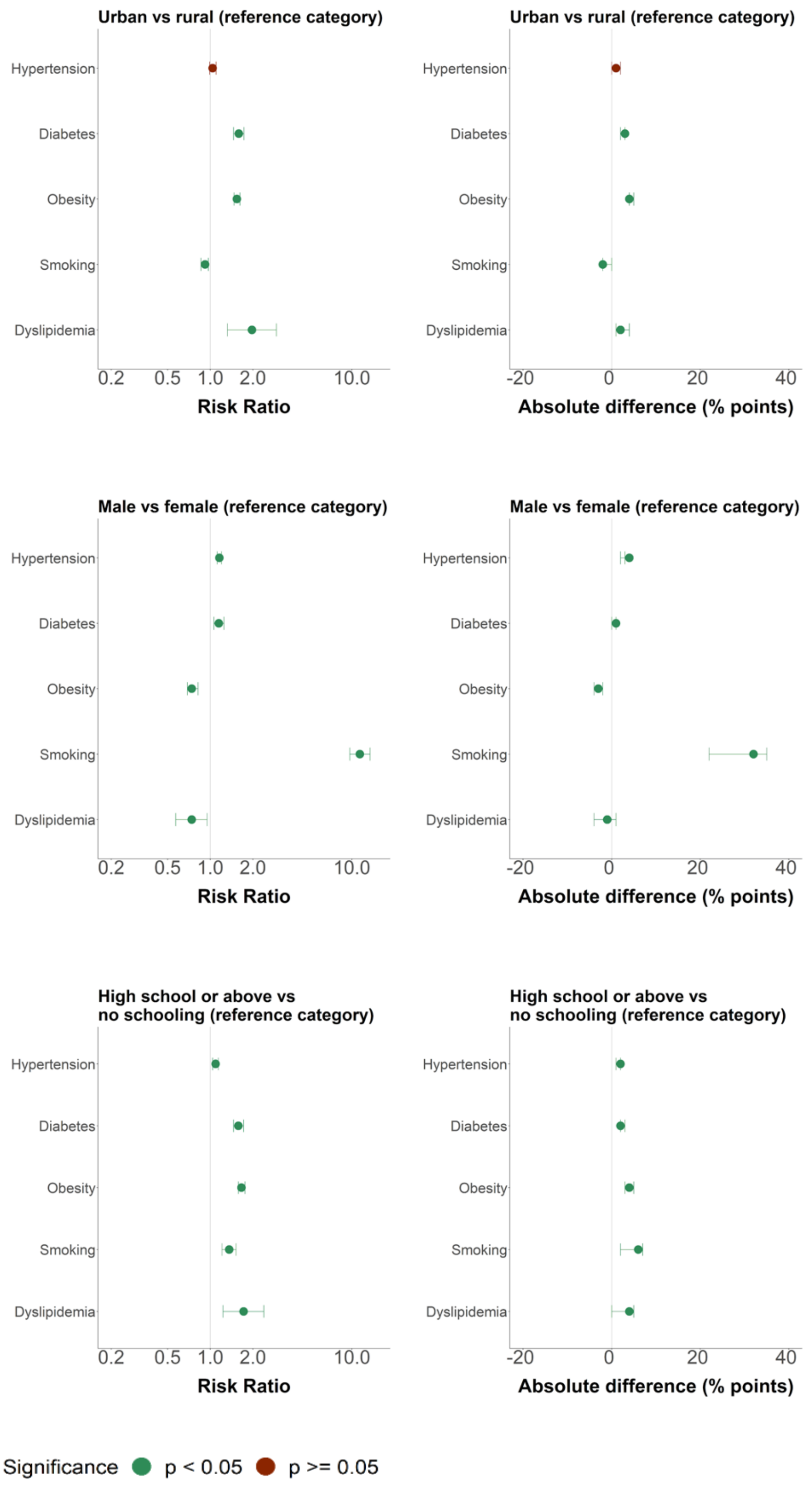

[1]Absolute difference (% points) is equivalent to absolute marginal effect (AME).
Data are point estimates with 95% CIs.

Figure S19: Variation within regions by sex[1]

### A. Hypertension

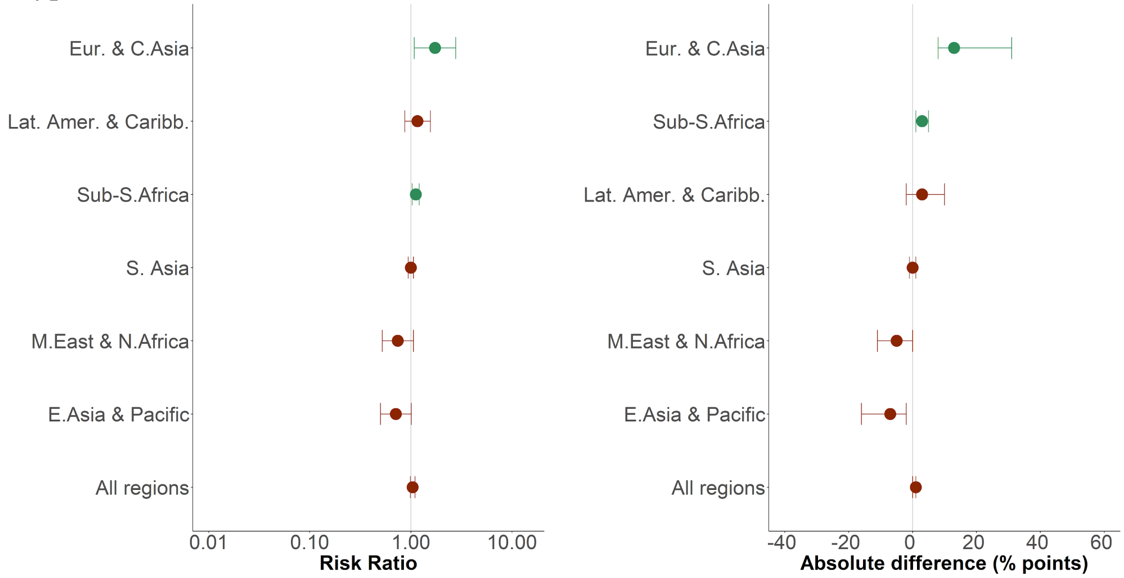

### B. Diabetes[2]

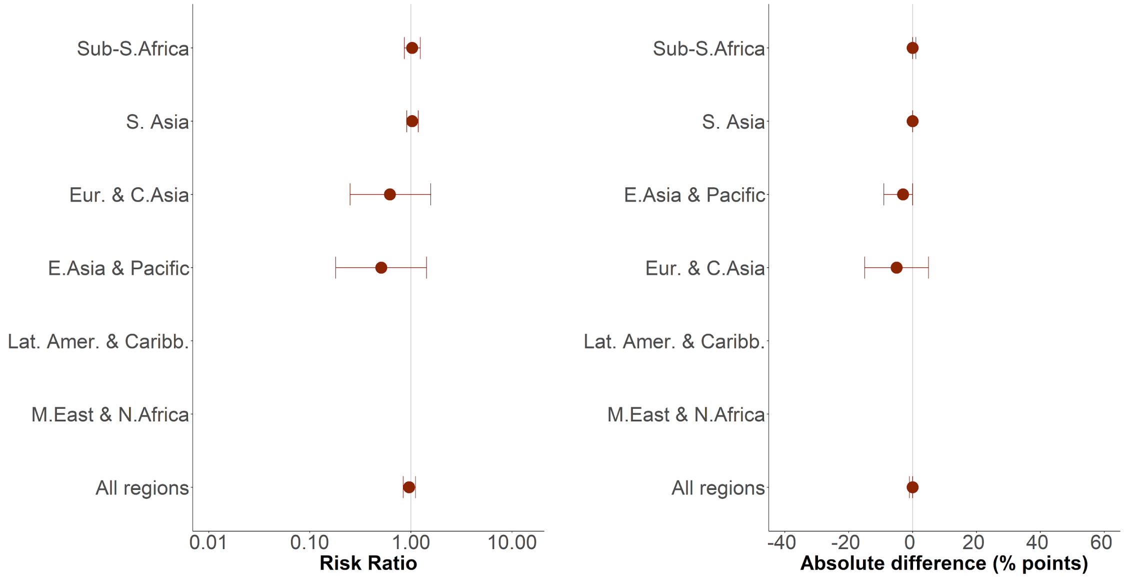

C. Overweight[3]

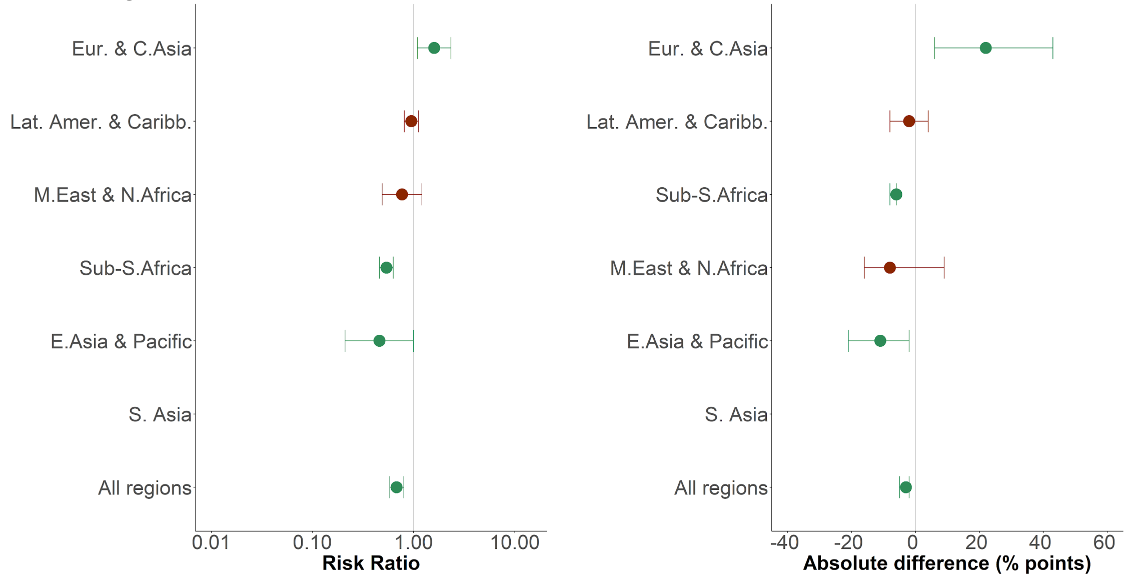

D. Obesity[4]

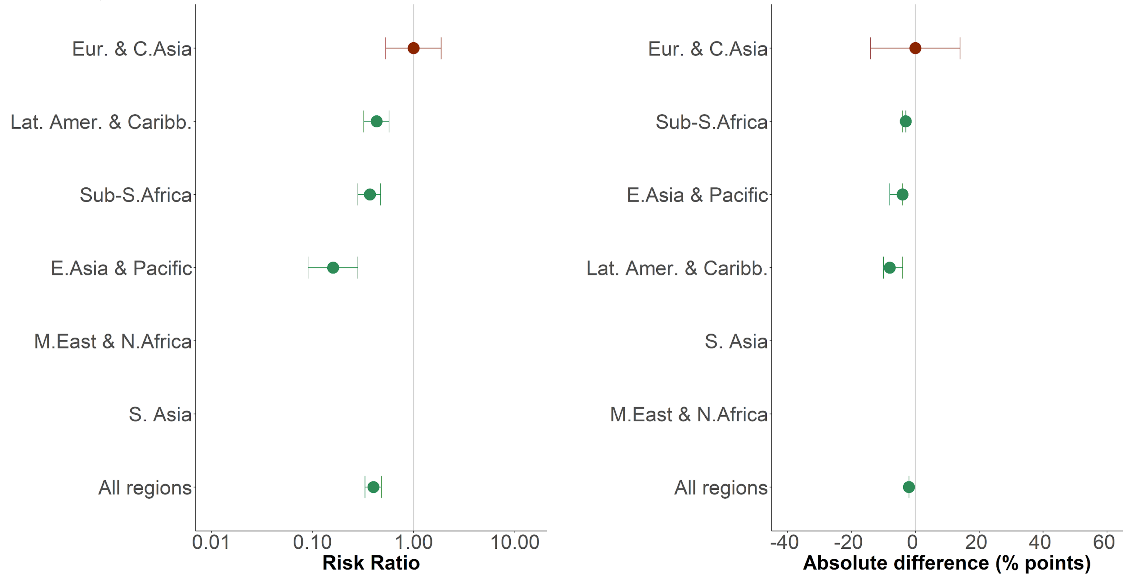

### E. Smoking

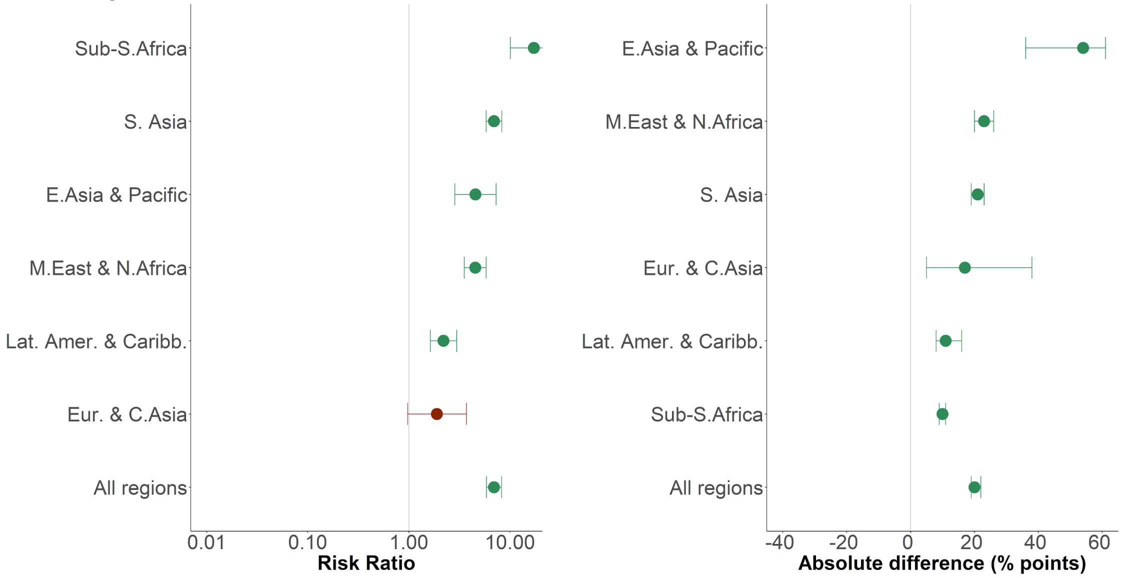

### F. Dyslipidemia

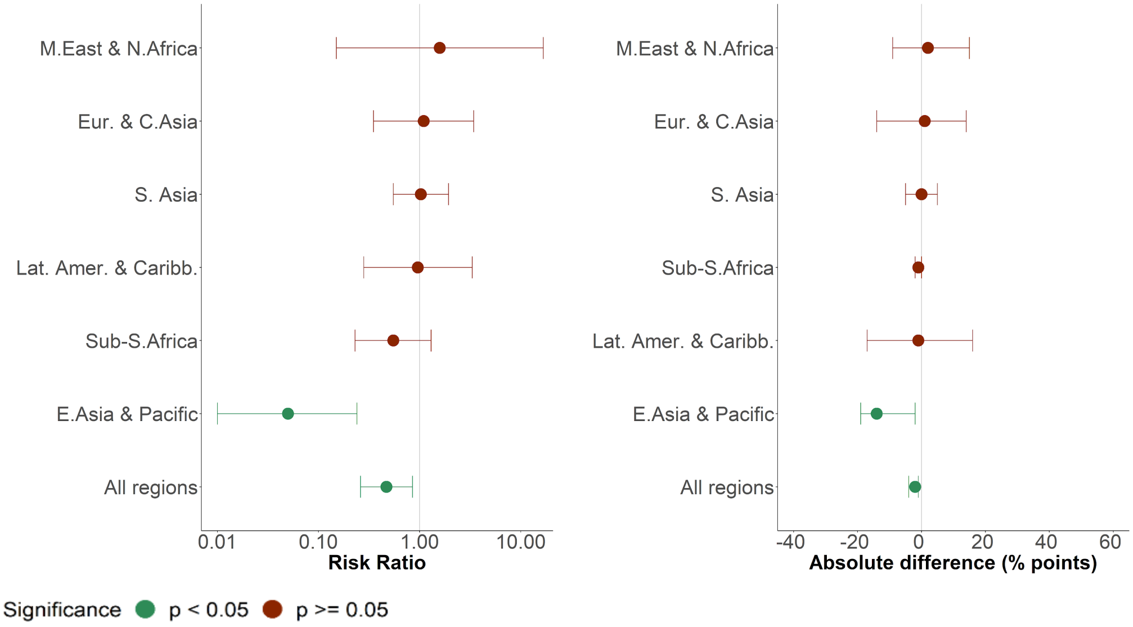

[1]For all comparisons, women are the reference group; risk ratio > 1 represents a statistically significantly greater chance of having a particular risk factor in men. Absolute difference (% points) is equivalent to absolute marginal effect (AME).
[2]Samples were too small in Latin America and the Caribbean (n=612) and Middle East and North Africa (n=91) to conduct regressions for diabetes.
[3]South Asian men were highly underrepresented in the sample (n=12); as such there was only one overweight male in this extremely small group, therefore regressions could not be conducted for sex.
[4]South Asian men were highly underrepresented in the sample (n=12); as such there were no obese males in this extremely small group. The sample of Middle East and North Africa has only four obese males, therefore regressions could not be conducted for sex for these two regions.
Data are point estimates with 95% CIs. Abbreviations: E.Asia & Pacific=East Asia and Pacific. Eur. & C.Asia=Europe and Central Asia. Lat. Amer. & Caribb.=Latin America and the Caribbean. M.East & N.Africa=Middle East and North Africa. S. Asia=South Asia. Sub-S.Africa=Sub-Saharan Africa.

Figure S20: Variation within regions by place of residence[1]

A. Hypertension

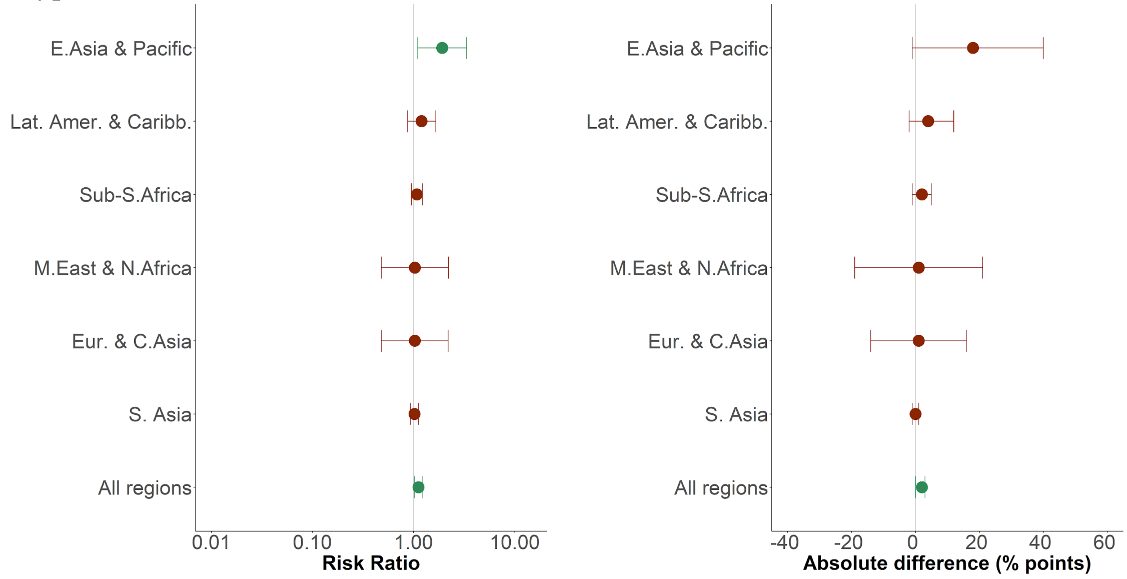

B. Diabetes[2]

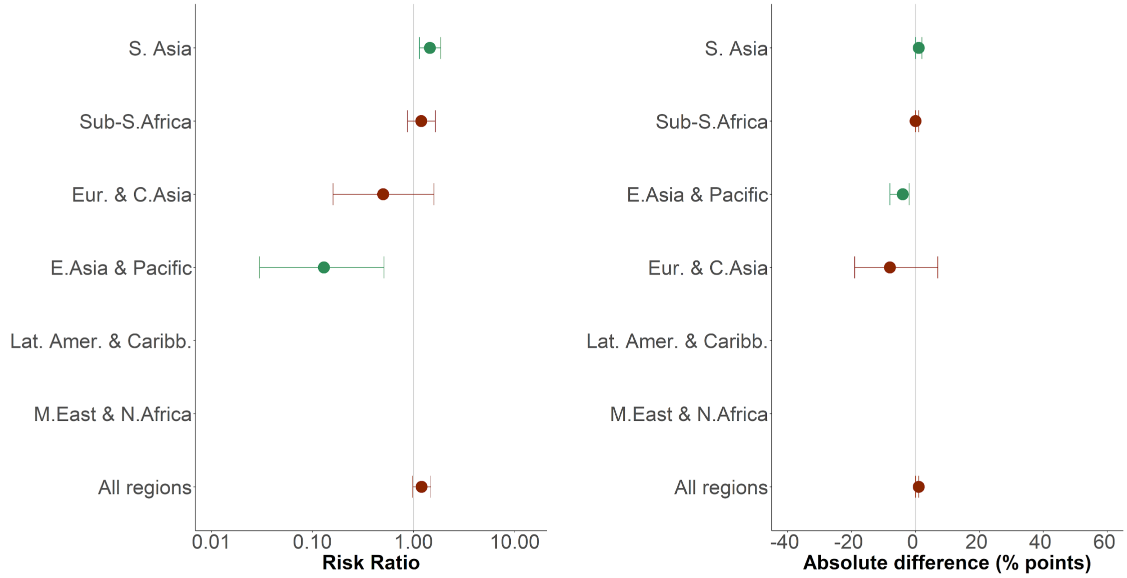

### C. Overweight

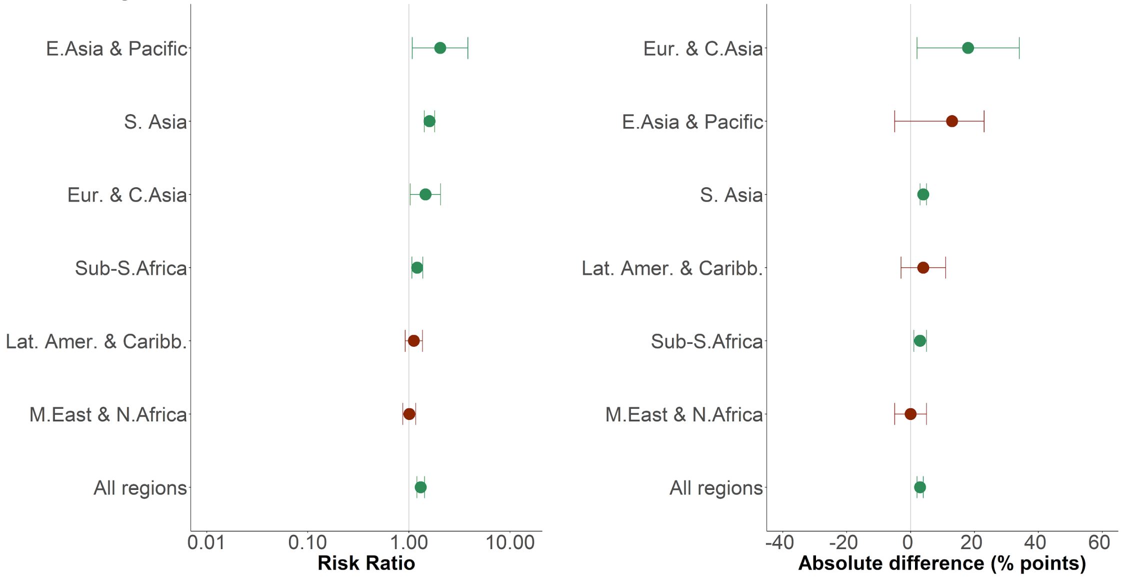

### D. Obesity

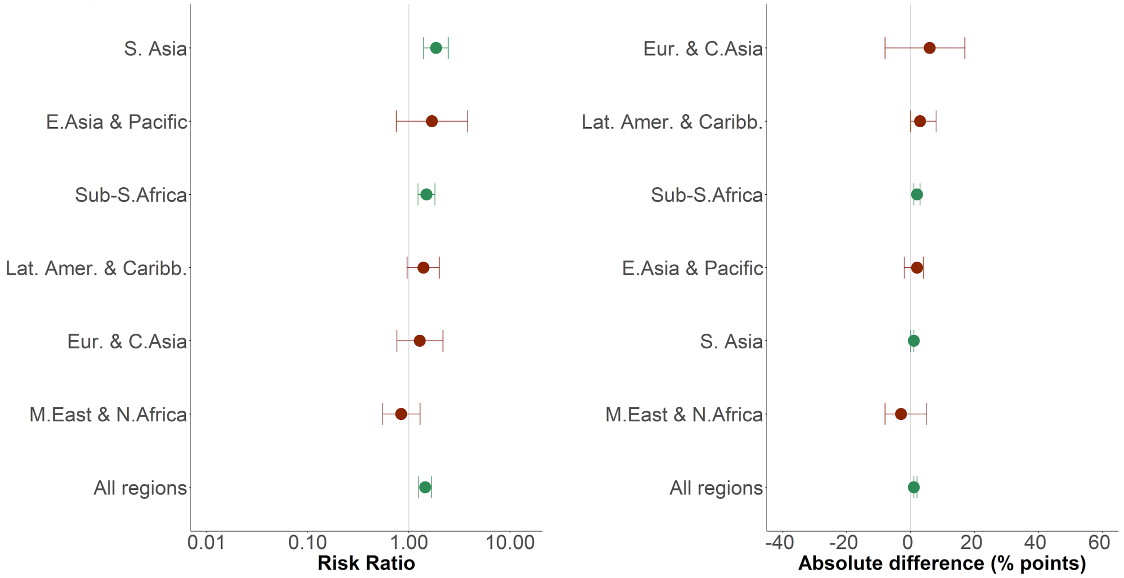

### E. Smoking

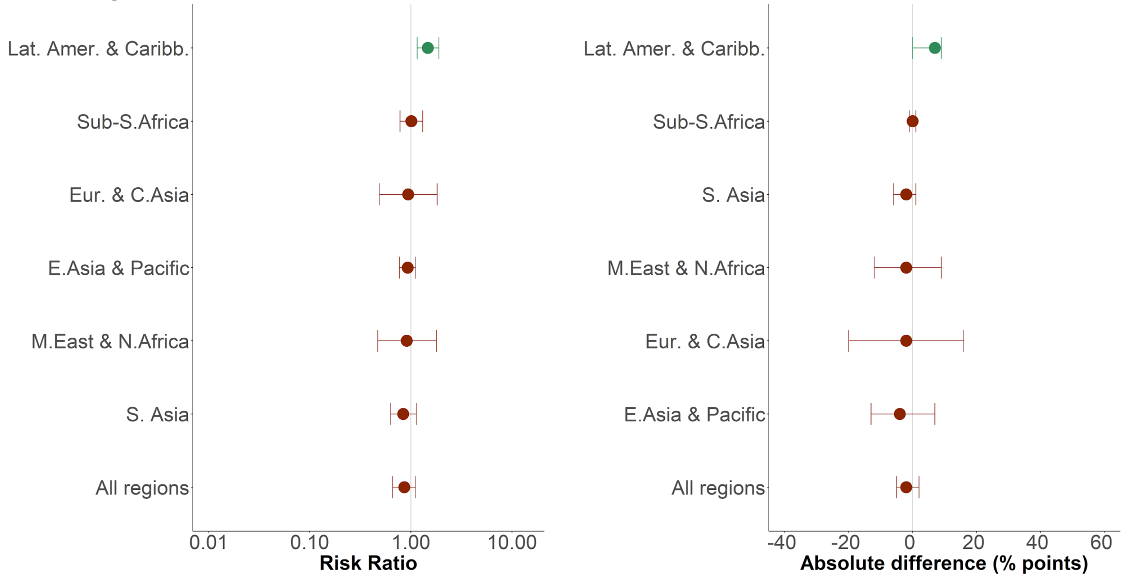

### F. Dyslipidemia[3]

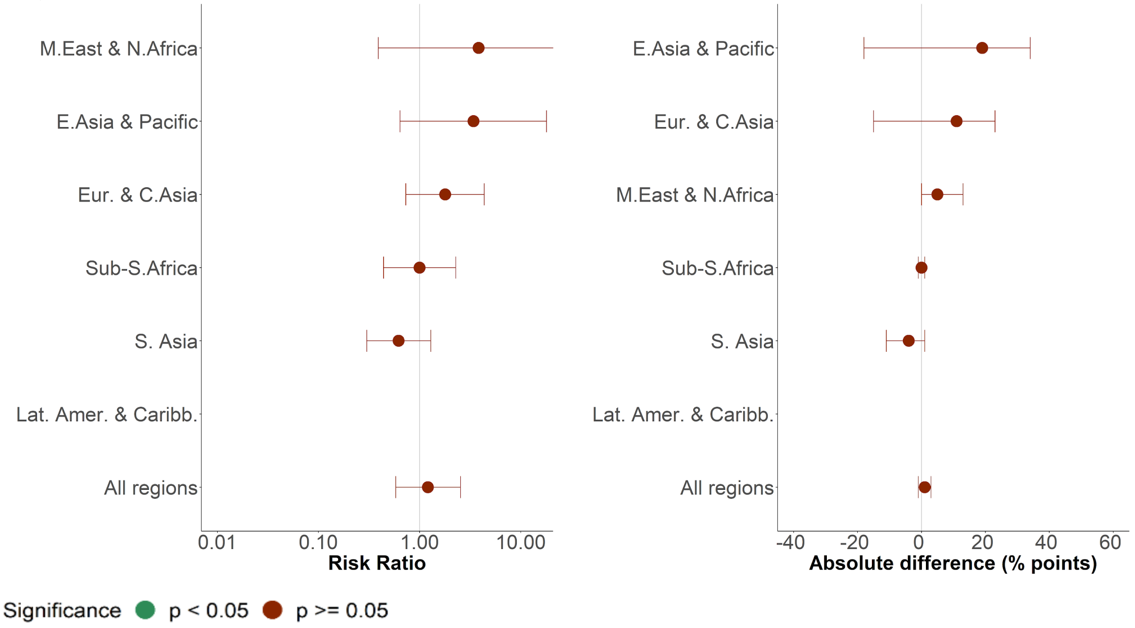

[1]For all comparisons, rural dwellers are the reference group; risk ratio > 1 represents a statistically significantly greater chance of having a particular risk factor in urban dwellers. Absolute difference (% points) is equivalent to absolute marginal effect (AME).
[2]Diabetics were highly underrepresented in the sample; as such there were only diabetics in one country (Mexico), therefore regression could not be conducted. For Middle East and North Africa, the sample was too small (n=91) to allow a regression.
[3]No surveys in Latin America and the Caribbean that measured dyslipidemia asked for place of residence, therefore regression could not be conducted.
Data are point estimates with 95% CIs. Abbreviations: E.Asia & Pacific=East Asia and Pacific. Eur. & C.Asia=Europe and Central Asia. Lat. Amer. & Caribb.=Latin America and the Caribbean. M.East & N.Africa=Middle East and North Africa. S. Asia=South Asia. Sub-S.Africa=Sub-Saharan Africa

Figure S21: Variation within regions by level of education[1]

A. Hypertension

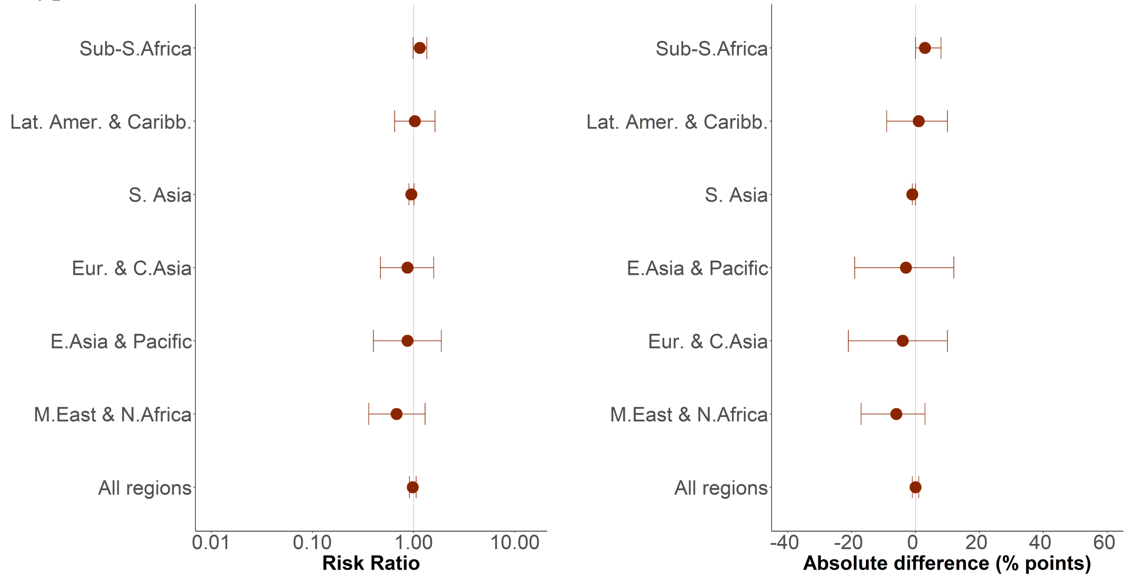

B. Diabetes[2]

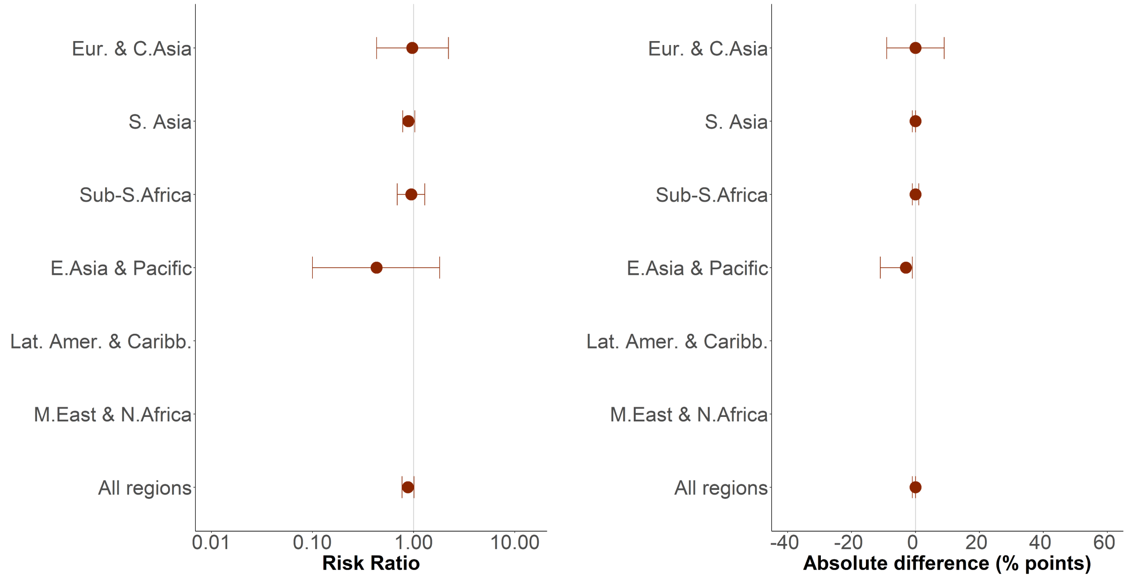

C. Overweight

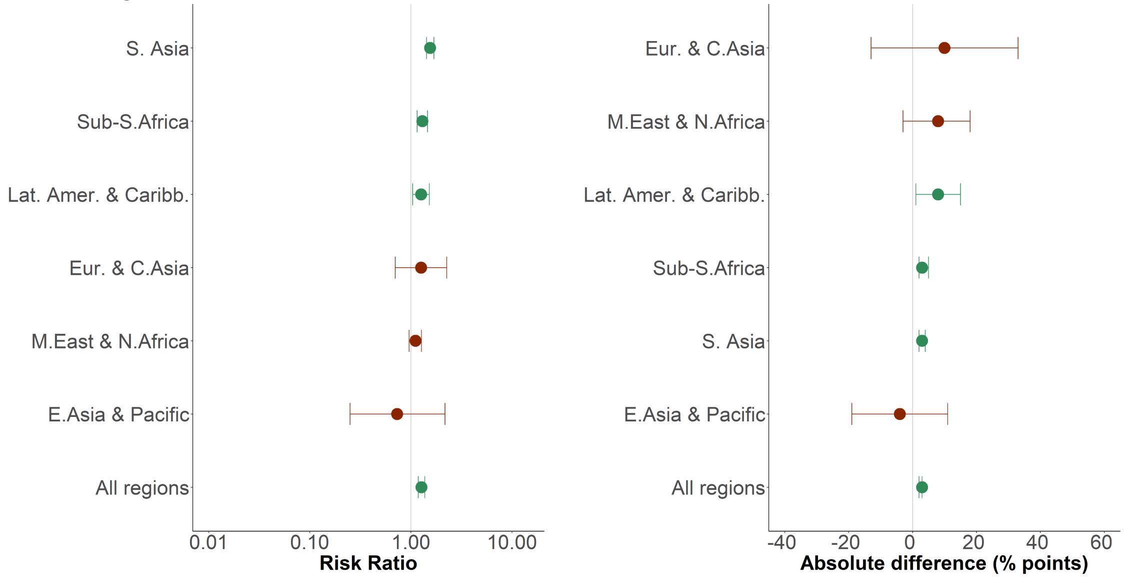

D. Obesity

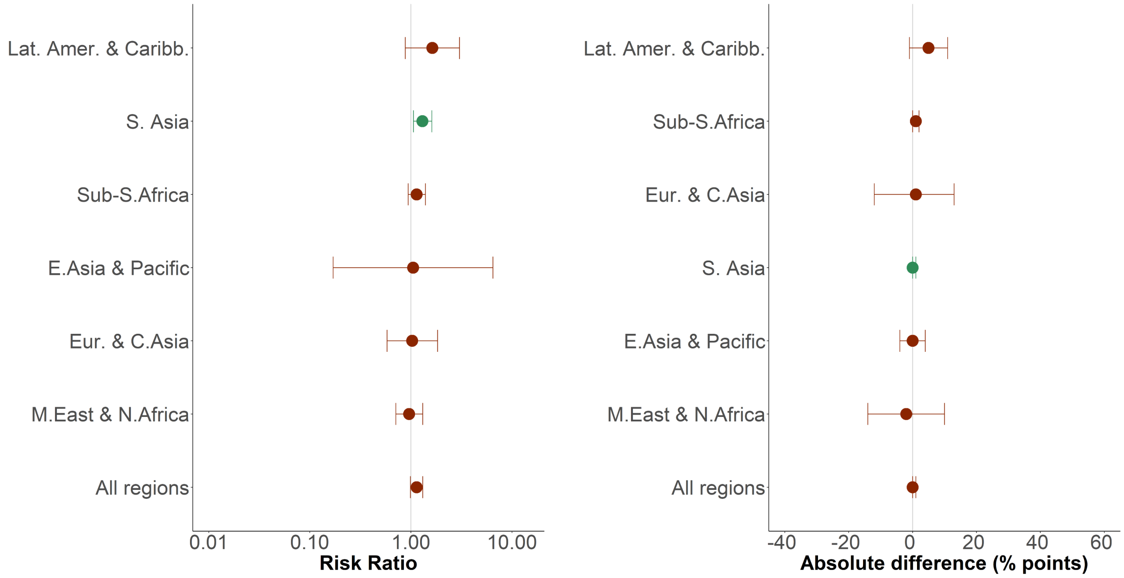

### E. Smoking

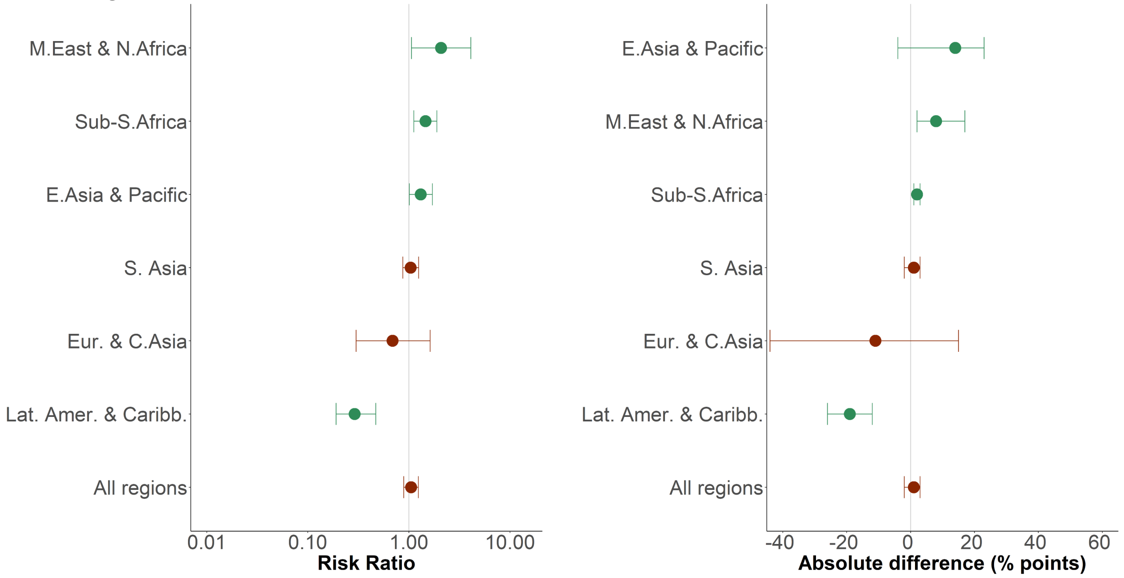

### F. Dyslipidemia

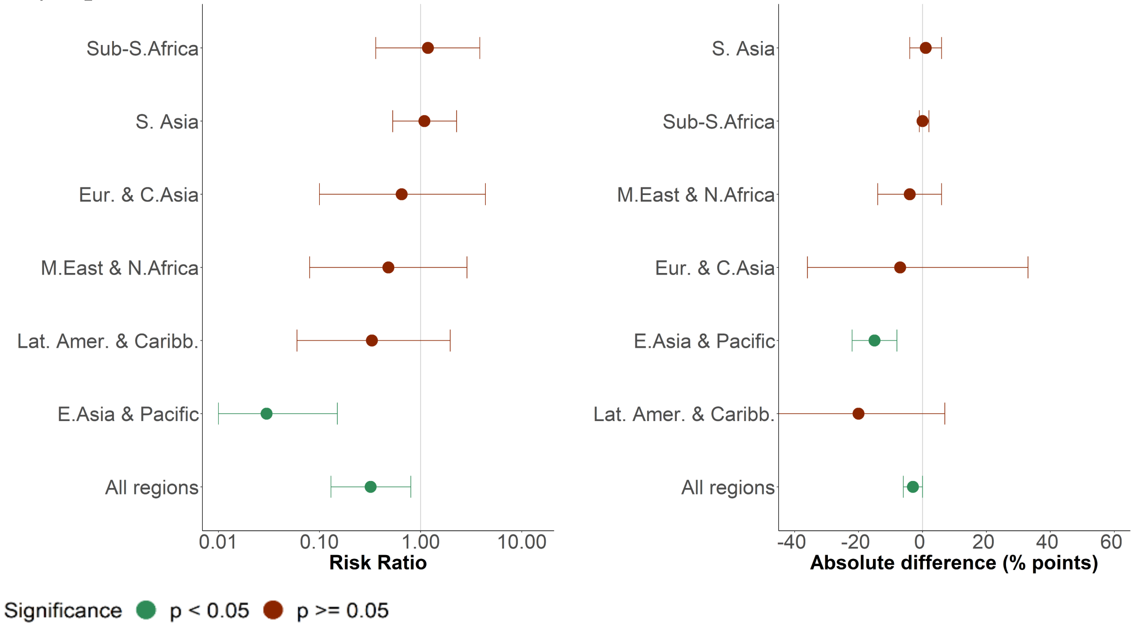

[1]For all comparisons, individuals with no schooling are the reference group; risk ratio > 1 represents a statistically significantly greater chance of having a particular risk factor in those with a high school education or above. Absolute difference (% points) is equivalent to absolute marginal effect (AME).

[2]Samples were too small in Latin America and the Caribbean (n=612) and Middle East and North Africa (n=91) to conduct regressions for diabetes.

Data are point estimates with 95% CIs. Abbreviations: E.Asia & Pacific=East Asia and Pacific. Eur. & C.Asia=Europe and Central Asia. Lat. Amer. & Caribb.=Latin America and the Caribbean. M.East & N.Africa=Middle East and North Africa. S. Asia=South Asia. Sub-S.Africa=Sub-Saharan Africa.

Table S25: Prevalences of the care cascade by World Bank country income group for hypertension, diabetes and statins, both sexes[1,2]

| World Bank country income group | Percentage using blood pressure lowering medication ("Hypt treated") | Percentage using blood pressure lowering medication and normal BP ("Hypt controlled") | Percentage using medication for diabetes ("Dia treated") | Percentage using statins for primary prevention ("Statin primary") | Percentage using statins for secondary prevention ("Statin secondary") |
|---|---|---|---|---|---|
| **All World Bank country income groups** | | | | | |
| Total | 24.6 (23.2-26.0) | 9.3 (8.6-10.0) | 39.1 (36.7-41.5) | 13.3 (11.2-15.3) | 14.2 (12.1-16.4) |
| <$1.90/day | 15.2 (13.3-17.1) | 5.7 (4.7-6.7) | 19.7 (15.3-24.1) | - | 1.1 (0.4-1.9) |
| <$3.20/day | 18.2 (16.7-20.0) | 7.0 (6.3-7.7) | 22.8 (19.8-25.7) | 0.5 (0.0-1.2) | 2.1 (0.8-3.3) |
| <$5.50/day | 20.1 (19.3-22.0) | 7.7 (7.1-8.4) | 26.7 (24.4-29.0) | 3.1 (1.7-4.4) | 5.8 (3.8-7.8) |
| >$5.50/day | 30.9 (28.7-33.0) | 11.9 (10.7-13.0) | 51.4 (47.4-55.7) | 18.2 (15.7-20.7) | 24.1 (19.5-28.8) |
| **Low-income** | | | | | |
| Total | 4.8 (4.0-5.7) | 1.5 (1.1-1.9) | 3.6 (2.4-5.5) | - | 2.5 (1.0-6.3) |
| <$1.90/day | 4.3 (3.4-5.5) | 1.3 (0.9-1.8) | 1.7 (1.2-2.5) | - | - |
| <$3.20/day | 4.5 (3.7-5.4) | 1.4 (1.0-1.9) | 1.9 (1.2-3.1) | - | - |
| <$5.50/day | 4.6 (3.8-5.5) | 1.4 (1.0-1.8) | 2.6 (1.6-4.0) | - | - |
| >$5.50/day | 8.0 (5.4-11.7) | 2.7 (1.4-5.3) | 26.0 (11.6-48.5) | - | - |
| **Lower middle-income** | | | | | |
| Total | 19.0 (18.0-20.0) | 9.1 (8.3-9.8) | 32.2 (29.7-34.8) | 15.0 (12.5-17.9) | 19.4 (16.0-23.4) |
| <$1.90/day | 11.3 (10.6-12.0) | 6.5 (6.0-7.1) | 19.8 (16.2-24.0) | - | 2.9 (1.1-7.3) |
| <$3.20/day | 14.2 (13.5-15.0) | 7.6 (7.0-8.1) | 22.4 (19.4-25.9) | 0.9 (0.2-3.6) | 5.2 (2.4-11.0) |
| <$5.50/day | 17.1 (16.2-18.0) | 8.4 (7.8-9.1) | 27.5 (24.8-30.4) | 3.8 (2.3-6.3) | 9.8 (6.8-14.1) |
| >$5.50/day | 24.3 (22.5-26.2) | 10.8 (9.5-12.2) | 41.6 (38.1-45.1) | 19.6 (16.7-22.7) | 26.9 (20.9-33.8) |
| **Upper middle-income** | | | | | |
| Total | 31.4 (29.0-33.9) | 10.3 (9.1-11.6) | 46.7 (42.1-51.4) | 14.6 (11.0-19.0) | 19.2 (14.8-24.6) |
| <$1.90/day | 27.3 (22.2-33.0) | 7.2 (4.8-10.8) | 27.7 (18.4-39.4) | - | - |
| <$3.20/day | 28.8 (25.2-32.8) | 7.8 (6.2-10.0) | 28.4 (22.4-35.2) | - | 7.3 (3.2-15.7) |
| <$5.50/day | 28.4 (25.6-31.3) | 7.8 (6.5-9.4) | 28.8 (24.5-33.4) | 7.7 (3.9-14.9) | 10.3 (5.1-19.6) |
| >$5.50/day | 34.1 (31.2-37.1) | 12.4 (11.0-14.1) | 55.7 (49.0-62.2) | 16.1 (12.0-21.3) | 22.1 (16.8-28.6) |

[1]Prevalences were not calculated for samples with less than 50 individuals.
[2]According to WHO guidelines, statin use is recommended for individuals aged 40 years or older. Statin treatment is therefore not calculated for younger age groups.

Table S26: Prevalences of the care cascade by World Bank country income group for hypertension, diabetes and statins, females only[1,2]

| World Bank country income group | Percentage using blood pressure lowering medication ("Hypt treated") | Percentage using blood pressure lowering medication and normal BP ("Hypt controlled") | Percentage using medication for diabetes ("Dia treated") | Percentage using statins for primary prevention ("Statin primary") | Percentage using statins for secondary prevention ("Statin secondary") |
|---|---|---|---|---|---|
| **All World Bank country income groups** | | | | | |
| Total | 28.8 (27.3-30.3) | 11.8 (10.9-12.7) | 40.3 (37.8-42.8) | 16.3 (13.6-19.0) | 10.4 (8.1-12.7) |
| <$1.90/day | 17.4 (15.4-19.4) | 7.4 (6.1-8.7) | 22.9 (17.2-28.5) | - | 1.3 (0.3-2.2) |
| <$3.20/day | 21.9 (20.2-23.4) | 9.1 (8.2-10.1) | 25.6 (22.1-29.2) | 0.4 (0.0-0.8) | 2.3 (1.0-3.6) |
| <$5.50/day | 24.6 (23.2-26.1) | 10.0 (9.2-10.9) | 29.5 (26.7-32.2) | 4.5 (2.4-6.7) | 5.0 (2.6-7.4) |
| >$5.50/day | 36.9 (34.5-39.3) | 15.2 (13.7-16.7) | 53.6 (49.4-57.8) | 21.1 (17.9-24.3) | 18.3 (12.6-24.0) |
| **Low-income** | | | | | |
| Total | 7.9 (6.5-9.7) | 2.7 (2.0-3.6) | 3.6 (1.9-6.8) | - | - |
| <$1.90/day | 6.8 (5.1-8.8) | 2.2 (1.6-3.2) | 2.2 (1.1-4.4) | - | - |
| <$3.20/day | 7.3 (5.8-9.1) | 2.5 (1.8-3.4) | 2.6 (1.2-5.6) | - | - |
| <$5.50/day | 7.7 (6.2-9.4) | 2.4 (1.8-3.3) | 2.8 (1.4-5.4) | - | - |
| >$5.50/day | 11.7 (6.8-19.5) | 6.9 (3.3-13.9) | 21.4 (4.2-62.5) | - | - |
| **Lower middle-income** | | | | | |
| Total | 22.4 (21.5-23.3) | 11.2 (10.4-12.0) | 32.2 (29.7-34.8) | 17.7 (14.6-21.3) | 16.1 (12.4-20.8) |
| <$1.90/day | 13.5 (12.7-14.4) | 8.0 (7.4-8.7) | 20.3 (16.1-25.2) | - | 2.9 (1.1-7.3) |
| <$3.20/day | 17.1 (16.4-17.9) | 9.4 (8.8-10.0) | 23.0 (19.9-26.4) | - | 5.2 (2.4-11.0) |
| <$5.50/day | 20.3 (19.5-21.2) | 10.4 (9.7-11.1) | 28.4 (25.6-31.4) | 5.0 (2.9-8.5) | 9.8 (6.8-14.1) |
| >$5.50/day | 29.1 (27.4-30.8) | 13.7 (12.2-15.2) | 40.4 (37.0-43.9) | 22.5 (19.0-26.4) | 26.9 (20.9-33.8) |
| **Upper middle-income** | | | | | |
| Total | 39.2 (36.2-42.2) | 13.5 (11.8-15.4) | 53.1 (47.4-58.7) | 15.3 (11.0-20.8) | 10.1 (6.6-15.1) |
| <$1.90/day | 30.9 (24.5-38.2) | 9.0 (5.5-14.4) | 35.6 (22.0-51.8) | - | - |
| <$3.20/day | 36.6 (31.8-41.6) | 10.3 (7.7-13.7) | 36.5 (27.6-46.5) | - | - |
| <$5.50/day | 36.3 (32.6-40.2) | 10.6 (8.4-13.2) | 35.8 (29.2-42.9) | 10.3 (5.4-18.8) | 10.3 (5.1-19.6) |
| >$5.50/day | 42.0 (38.5-45.5) | 16.3 (14.2-18.6) | 62.2 (54.5-69.4) | 16.4 (11.5-22.9) | 22.1 (16.8-28.6) |

[1]Prevalences were not calculated for samples with less than 50 individuals.
[2]According to WHO guidelines, statin use is recommended for individuals aged 40 years or older. Statin treatment is therefore not calculated for younger age groups.

Figure S22: Hypertension treatment, hypertension control, diabetes and statin treatment by World Bank country income groups, females only[1,2]

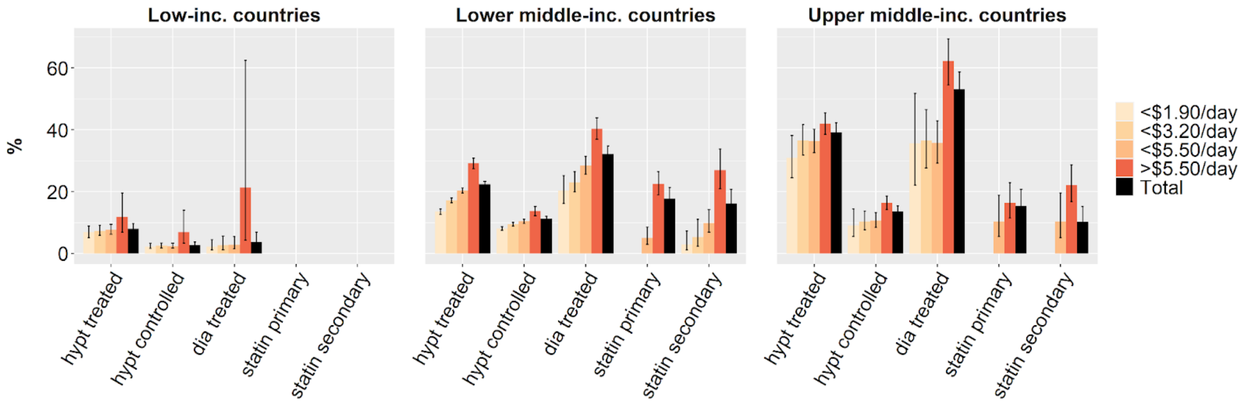

[1]Prevalences were not calculated for samples with less than 50 individuals.
[2]According to WHO guidelines, statin use is recommended for individuals aged 40 years or older. Statin treatment is therefore not calculated for younger age groups.

Table S27: Prevalences of the care cascade by World Bank country income group for hypertension, diabetes and statins, males only[1,2]

| World Bank country income group | Percentage using blood pressure lowering medication ("Hypt treated") | Percentage using blood pressure lowering medication and normal BP ("Hypt controlled") | Percentage using medication for diabetes ("Dia treated") | Percentage using statins for primary prevention ("Statin primary") | Percentage using statins for secondary prevention ("Statin secondary") |
|---|---|---|---|---|---|
| **All World Bank country income groups** | | | | | |
| Total | 19.3 (17.6-21.1) | 6.3 (5.5-7.1) | 37.3 (34.0-40.7) | 9.7 (7.5-11.8) | 19.1 (15.6-22.7) |
| <$1.90/day | 11.7 (8.6-14.7) | 3.1 (1.6-4.6) | 13.5 (7.8-19.2) | - | - |
| <$3.20/day | 12.3 (10.3-14.4) | 3.7 (2.7-4.7) | 17.3 (12.8-21.8) | - | 1.6 (0.0-4.1) |
| <$5.50/day | 14.7 (13.0-16.3) | 4.3 (3.5-5.2) | 21.6 (18.2-25.9) | 1.8 (0.2-3.3) | 7.1 (3.5-10.7) |
| >$5.50/day | 25.1 (22.4-27.8) | 8.7 (7.4-10.0) | 49.0 (43.5-54.5) | 14.3 (11.3-17.5) | 29.7 (23.5-35.8) |
| **Low-income** | | | | | |
| Total | 2.2 (1.7-2.9) | 0.5 (0.3-0.8) | 3.7 (1.9-6.9) | - | - |
| <$1.90/day | 2.1 (1.4-3.0) | 0.4 (0.2-0.9) | 1.4 (0.6-3.2) | - | - |
| <$3.20/day | 2.1 (1.5-2.8) | 0.5 (0.3-0.8) | 1.2 (0.3-4.4) | - | - |
| <$5.50/day | 2.0 (1.5-2.6) | 0.5 (0.3-0.8) | 2.4 (1.1-5.1) | - | - |
| >$5.50/day | 5.7 (3.1-10.2) | 0.1 (0.0-0.3) | 29.6 (11.0-59.0) | - | - |
| **Lower middle-income** | | | | | |
| Total | 12.2 (11.2-13.3) | 4.8 (4.2-5.5) | 32.2 (28.3-36.4) | 11.3 (8.7-14.4) | 23.7 (19.0-29.1) |
| <$1.90/day | 5.6 (4.6-6.8) | 2.6 (2.0-3.5) | 20.6 (15.0-27.7) | - | - |
| <$3.20/day | 7.3 (6.5-8.2) | 3.2 (2.7-3.8) | 18.0 (13.2-24.0) | - | - |
| <$5.50/day | 9.9 (9.1-10.9) | 4.1 (3.5-4.7) | 24.4 (20.5-28.7) | 2.4 (0.8-6.6) | 10.0 (5.4-17.8) |
| >$5.50/day | 17.2 (15.2-19.3) | 6.6 (5.4-7.9) | 44.2 (37.8-50.9) | 15.2 (12.0-19.2) | 32.5 (25.5-40.3) |
| **Upper middle-income** | | | | | |
| Total | 24.9 (22.3-27.6) | 7.6 (6.4-8.9) | 41.1 (36.4-46.1) | 13.8 (8.8-21.1) | 27.6 (20.3-36.3) |
| <$1.90/day | 23.2 (16.6-31.4) | 5.3 (2.6-10.7) | 18.9 (12.1-28.2) | - | - |
| <$3.20/day | 20.8 (16.6-25.5) | 5.2 (3.3-8.1) | 17.1 (8.3-32.1) | - | 6.6 (2.2-18.2) |
| <$5.50/day | 21.1 (18.1-24.3) | 5.3 (3.9-7.1) | 22.2 (17.0-28.6) | - | 14.9 (5.7-33.6) |
| >$5.50/day | 28.0 (24.6-31.6) | 9.4 (7.9-11.2) | 50.2 (43.4-57.0) | - | 30.8 (22.4-40.5) |

[1]Prevalences were not calculated for samples with less than 50 individuals.
[2]According to WHO guidelines, statin use is recommended for individuals aged 40 years or older. Statin treatment is therefore not calculated for younger age groups.

Figure S23: Hypertension treatment, hypertension control, diabetes and statin treatment by World Bank country income groups, males only[1,2]

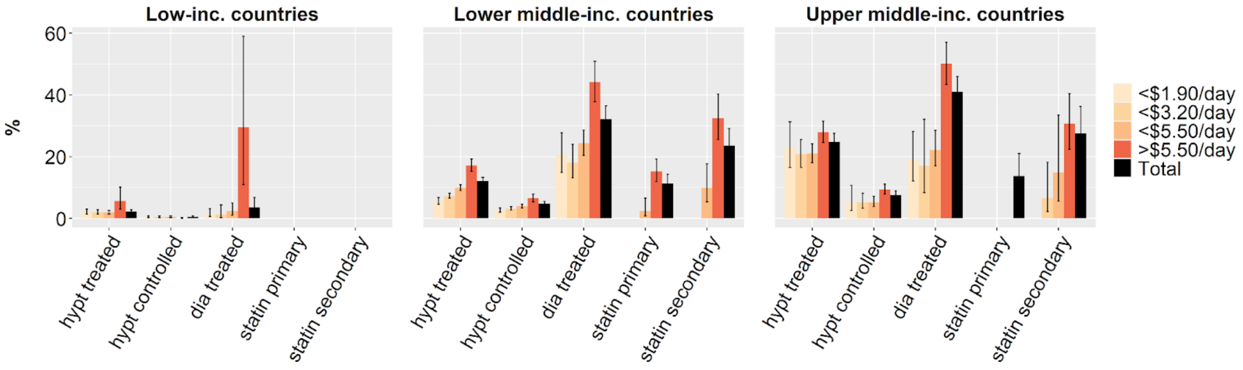

[1]Prevalences were not calculated for samples with less than 50 individuals.
[2]According to WHO guidelines, statin use is recommended for individuals aged 40 years or older. Statin treatment is therefore not calculated for younger age groups.

Table S28: Prevalences of the care cascade by age group for hypertension, diabetes and statins, both sexes[1,2]

| Age group | Percentage using blood pressure lowering medication ("Hypt treated") | Percentage using blood pressure lowering medication and normal BP ("Hypt controlled") | Percentage using medication for diabetes ("Dia treated") | Percentage using statins for primary prevention ("Statin primary") | Percentage using statins for secondary prevention ("Statin secondary") |
|---|---|---|---|---|---|
| **All age groups** | | | | | |
| Total | 24.6 (23.2-26.0) | 9.3 (8.6-10.0) | 39.0 (36.7-41.5) | 13.3 (11.2-15.3) | 14.3 (12.1-16.4) |
| <$1.90/day | 15.2 (13.3-17.1) | 5.7 (4.7-6.7) | 19.7 (15.4-24.1) | - | 1.1 (0.4-1.9) |
| <$3.20/day | 18.2 (16.7-19.7) | 7.0 (6.3-7.7) | 22.8 (19.8-25.7) | 0.5 (0.0-1.2) | 2.1 (0.8-3.3) |
| <$5.50/day | 20.6 (19.3-21.9) | 7.7 (7.1-8.4) | 26.7 (24.4-29.0) | 3.1 (1.8-4.5) | 5.8 (3.8-7.8) |
| >$5.50/day | 30.9 (28.7-33.0) | 11.9 (10.7-13.0) | 51.1 (47.1-55.7) | 18.2 (15.7-20.7) | 24.1 (19.5-28.8) |
| **15-24** | | | | | |
| Total | 7.7 (6.9-8.6) | 6.6 (5.9-7.4) | 7.9 (5.0-12.4) | - | - |
| <$1.90/day | 7.3 (5.9-9.0) | 6.2 (5.0-7.6) | 8.9 (6.7-11.8) | - | - |
| <$3.20/day | 8.2 (7.3-9.3) | 7.4 (6.5-8.4) | 7.2 (5.7-8.9) | - | - |
| <$5.50/day | 8.2 (7.3-9.2) | 7.2 (6.4-8.0) | 9.3 (5.6-15.2) | - | - |
| >$5.50/day | 6.5 (5.1-8.3) | 5.1 (4.0-6.6) | 3.8 (2.4-6.0) | - | - |
| **25-34** | | | | | |
| Total | 11.3 (10.3-12.3) | 7.1 (6.5-7.8) | 15.2 (12.2-18.9) | - | - |
| <$1.90/day | 6.6 (5.8-7.6) | 4.5 (3.9-5.2) | 11.4 (8.9-14.5) | - | - |
| <$3.20/day | 9.6 (8.8-10.4) | 6.6 (6.0-7.2) | 13.0 (11.1-15.1) | - | - |
| <$5.50/day | 10.8 (10.0-11.7) | 6.9 (6.3-7.4) | 14.4 (12.7-16.3) | - | - |
| >$5.50/day | 12.3 (10.2-14.8) | 7.7 (6.2-9.7) | 16.7 (10.0-26.5) | - | - |
| **35-44** | | | | | |
| Total | 17.7 (16.3-19.2) | 8.1 (7.3-9.1) | 32.4 (29.0-36.0) | 6.8 (4.3-10.6) | 4.6 (2.6-8.2) |
| <$1.90/day | 11.7 (9.3-14.5) | 5.4 (4.0-7.1) | 19.0 (12.2-28.3) | - | - |
| <$3.20/day | 14.3 (12.7-16.0) | 6.1 (5.3-7.1) | 26.0 (20.6-32.3) | - | - |
| <$5.50/day | 16.2 (14.8-17.6) | 7.3 (6.4-8.2) | 26.4 (22.8-30.3) | - | 1.8 (0.6-5.1) |
| >$5.50/day | 20.5 (17.9-23.4) | 9.6 (8.0-11.4) | 40.7 (34.5-47.2) | - | 9.1 (4.5-17.4) |
| **45-54** | | | | | |

| | | | | | |
|---|---|---|---|---|---|
| Total | 26.4 (24.5-28.4) | 9.3 (8.3-10.5) | 45.4 (41.6-49.3) | 11.2 (8.8-14.2) | 10.1 (7.6-13.3) |
| <$1.90/day | 14.7 (11.3-18.8) | 6.8 (4.5-10.2) | 30.9 (20.1-44.4) | - | - |
| <$3.20/day | 18.3 (15.9-21.0) | 7.1 (5.7-8.8) | 28.1 (22.2-34.8) | - | 1.2 (0.5-2.9) |
| <$5.50/day | 22.6 (20.6-24.8) | 7.6 (6.5-8.8) | 36.9 (32.3-41.7) | 3.4 (1.5-7.4) | 4.6 (2.1-9.8) |
| >$5.50/day | 31.6 (28.7-34.5) | 11.7 (10.0-13.6) | 52.1 (46.3-57.9) | 14.6 (11.6-18.3) | 16.5 (12.1-22.2) |
| **55-64** | | | | | |
| Total | 38.4 (35.4-41.5) | 12.1 (10.4-14.1) | 55.9 (51.1-60.6) | 15.3 (12.7-18.4) | 25.4 (20.3-31.1) |
| <$1.90/day | 24.8 (19.2-31.4) | 5.2 (2.8-9.3) | 25.3 (15.4-38.6) | - | - |
| <$3.20/day | 29.4 (25.1-34.2) | 7.7 (5.5-10.6) | 29.4 (21.4-38.9) | - | 5.0 (1.7-13.5) |
| <$5.50/day | 31.1 (27.8-34.7) | 8.6 (6.8-10.8) | 32.3 (26.4-39.0) | 3.7 (2.1-6.3) | 12.1 (7.9-18.2) |
| >$5.50/day | 47.2 (43.3-51.2) | 16.5 (14.0-19.3) | 69.6 (64.0-74.7) | 20.2 (16.9-24.0) | 38.1 (30.9-45.9) |
| **>65** | | | | | |
| Total | 46.2 (42.6-49.9) | 12.8 (11.1-14.8) | 57.8 (52.7-62.6) | 25.7 (19.8-32.7) | 28.8 (19.6-40.1) |
| <$1.90/day | 33.5 (26.8-40.9) | 6.6 (3.7-11.6) | 22.2 (11.9-37.6) | - | - |
| <$3.20/day | 37.6 (32.1-43.5) | 8.9 (6.5-12.1) | 32.5 (23.0-43.7) | - | 3.4 (1.1-10.5) |
| <$5.50/day | 40.3 (35.7-45.1) | 10.1 (8.1-12.7) | 38.3 (30.8-46.3) | 3.0 (1.2-7.2) | 3.4 (1.4-8.1) |
| >$5.50/day | 53.7 (49.4-57.9) | 16.3 (13.8-19.2) | 70.1 (64.6-75.0) | 34.0 (26.6-42.3) | 53.2 (40.1-65.8) |

[1]Prevalences were not calculated for samples with less than 50 individuals.
[2]According to WHO guidelines, statin use is recommended for individuals aged 40 years or older. Statin treatment is therefore not calculated for younger age groups.

Table S29: Prevalences of the care cascade by age group for hypertension, diabetes and statins, females only[1,2]

| Age group[3] | Percentage using blood pressure lowering medication ("Hypt treated") | Percentage using blood pressure lowering medication and normal BP ("Hypt controlled") | Percentage using medication for diabetes ("Dia treated") | Percentage using statins for primary prevention ("Statin primary") | Percentage using statins for secondary prevention ("Statin secondary") |
|---|---|---|---|---|---|
| **All age groups** | | | | | |
| Total | 28.8 (27.3-30.3) | 11.8 (10.9-12.7) | 40.3 (37.8-42.8) | 16.3 (13.6-19.0) | 10.4 (8.1-12.7) |
| <$1.90/day | 17.4 (15.4-19.5) | 7.4 (6.1-8.7) | 22.9 (17.3-28.6) | - | 1.3 (0.4-2.3) |
| <$3.20/day | 21.9 (20.2-23.5) | 9.1 (8.2-10.1) | 25.6 (22.1-29.2) | - | 2.3 (1.0-3.6) |
| <$5.50/day | 24.6 (23.2-26.1) | 10.0 (9.2-10.9) | 29.5 (26.7-32.2) | 4.6 (2.4-6.7) | 5.0 (2.6-7.4) |
| >$5.50/day | 36.9 (34.5-39.0) | 15.2 (13.7-16.7) | 53.6 (49.4-57.8) | 21.1 (17.9-24.3) | 18.3 (12.6-24.0) |
| **15-24** | | | | | |
| Total | 12.1 (11.1-13.3) | 10.6 (9.6-11.7) | 9.6 (5.8-15.4) | - | - |
| <$1.90/day | 11.0 (9.2-13.3) | 9.4 (7.9-11.3) | 10.0 (7.3-13.6) | - | - |
| <$3.20/day | 11.7 (10.5-13.1) | 10.7 (9.5-11.9) | 8.4 (6.8-10.4) | - | - |
| <$5.50/day | 12.5 (11.4-13.7) | 10.9 (9.9-12.1) | 11.4 (6.6-19.0) | - | - |
| >$5.50/day | 10.7 (8.3-13.6) | 9.4 (7.2-12.1) | 4.1 (2.6-6.4) | - | - |
| **25-34** | | | | | |
| Total | 15.4 (14.5-16.4) | 10.6 (9.8-11.4) | 18.8 (14.9-23.5) | - | - |
| <$1.90/day | 10.3 (9.1-11.6) | 7.0 (6.2-8.0) | 14.3 (10.9-18.5) | - | - |
| <$3.20/day | 13.9 (12.9-15.0) | 9.8 (9.0-10.7) | 15.9 (13.6-18.5) | - | - |
| <$5.50/day | 15.2 (14.3-16.2) | 10.2 (9.5-11.0) | 17.3 (15.1-19.6) | - | - |
| >$5.50/day | 16.0 (13.7-18.5) | 11.6 (9.8-13.8) | 21.9 (12.8-34.8) | - | - |
| **35-44** | | | | | |
| Total | 22.1 (20.5-23.8) | 10.8 (9.7-12.0) | 37.0 (33.5-40.6) | 12.7 (7.9-19.6) | 3.1 (1.5-6.3) |
| <$1.90/day | 12.6 (10.5-15.0) | 6.1 (5.3-7.1) | 26.8 (16.9-39.7) | - | - |
| <$3.20/day | 17.6 (15.8-19.7) | 8.0 (7.1-9.0) | 31.0 (25.0-37.9) | - | - |
| <$5.50/day | 20.2 (18.6-21.9) | 9.3 (8.3-10.4) | 31.4 (27.7-35.3) | - | 2.6 (0.9-7.4) |
| >$5.50/day | 26.4 (23.1-29.9) | 14.1 (11.7-17.0) | 46.2 (39.2-53.5) | 17.1 (10.1-27.4) | 4.0 (1.3-11.9) |
| **45-54** | | | | | |

| | | | | | |
|---|---|---|---|---|---|
| Total | 30.9 (28.4-33.4) | 12.1 (10.7-13.7) | 48.1 (43.5-52.7) | 14.6 (11.0-19.0) | 8.2 (5.2-12.8) |
| <$1.90/day | 17.2 (13.1-22.3) | 9.4 (6.0-14.6) | 33.2 (19.8-49.9) | - | - |
| <$3.20/day | 22.0 (18.8-25.5) | 8.7 (6.9-10.9) | 31.0 (23.7-39.5) | - | 1.9 (0.7-5.0) |
| <$5.50/day | 26.0 (23.4-28.8) | 9.9 (8.4-11.7) | 40.9 (35.2-46.8) | 4.3 (1.9-9.5) | 5.3 (1.8-14.6) |
| >$5.50/day | 38.5 (34.7-42.3) | 15.4 (13.1-18.1) | 55.3 (48.2-62.1) | 18.6 (14.1-24.1) | 11.7 (7.3-18.2) |
| **55-64** | | | | | |
| Total | 44.5 (40.6-48.4) | 13.9 (11.6-16.6) | 58.0 (51.7-63.9) | 17.0 (13.7-20.9) | 18.6 (12.7-26.5) |
| <$1.90/day | 23.2 (16.3-31.8) | 4.4 (1.9-9.9) | 27.2 (14.3-45.5) | - | - |
| <$3.20/day | 33.9 (28.5-39.7) | 9.6 (6.7-13.6) | 32.6 (22.7-44.4) | - | 3.9 (1.3-10.8) |
| <$5.50/day | 37.7 (33.2-42.4) | 10.6 (8.1-13.9) | 36.2 (28.0-45.2) | 4.6 (2.4-8.6) | 7.4 (4.4-11.9) |
| >$5.50/day | 53.4 (48.5-58.2) | 18.3 (14.9-22.3) | 73.3 (66.0-79.6) | 21.7 (17.8-26.3) | 35.6 (24.9-48.1) |
| **>65** | | | | | |
| Total | 49.0 (45.1-53.0) | 13.2 (11.0-15.8) | 61.5 (55.2-67.4) | 26.9 (20.6-34.4) | 21.4 (11.3-36.6) |
| <$1.90/day | 39.9 (31.6-48.8) | 8.5 (4.4-15.7) | 27.0 (13.5-46.9) | - | - |
| <$3.20/day | 42.9 (36.3-49.7) | 10.4 (7.0-15.1) | 35.5 (23.9-49.2) | - | - |
| <$5.50/day | 44.6 (39.1-50.3) | 10.6 (7.9-14.1) | 40.0 (30.6-50.1) | 4.3 (1.5-11.7) | 3.7 (1.2-10.7) |
| >$5.50/day | 55.8 (51.0-60.4) | 17.2 (13.8-21.2) | 75.0 (67.9-81.0) | 35.4 (27.2-44.6) | 53.5 (35.2-71) |

[1]Prevalences were not calculated for samples with less than 50 individuals.
[2]According to WHO guidelines, statin use is recommended for individuals aged 40 years or older. Statin treatment is therefore not calculated for younger age groups.

Figure S24: Hypertension treatment, hypertension control, diabetes and statin treatment by poverty level (age groups 45-54; 55-64; 65+)[1,2,3]

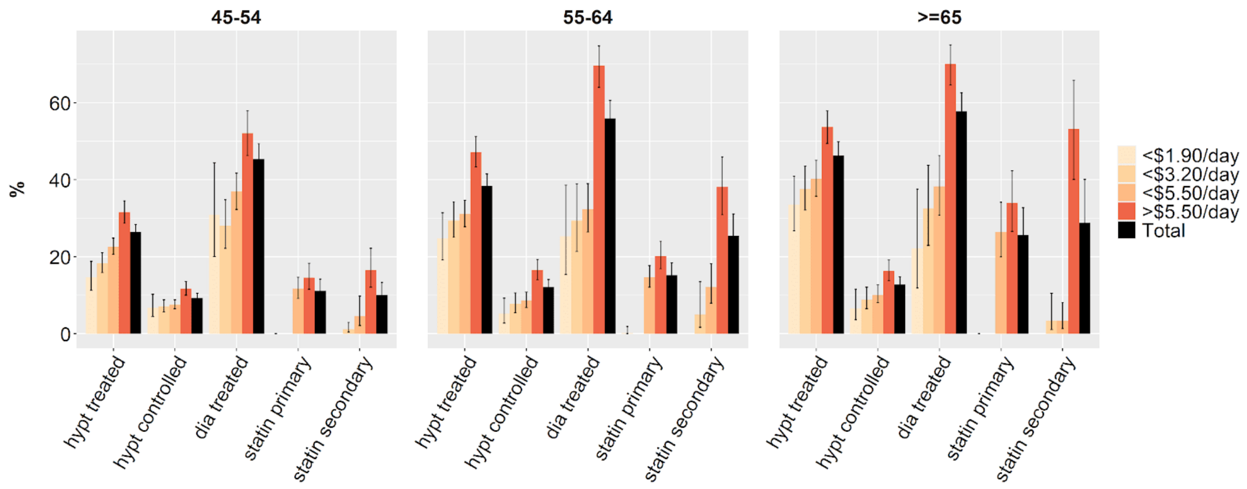

[1]Prevalences were not calculated for samples with less than 50 individuals.
[2]According to WHO guidelines, statin use is recommended for individuals aged 40 years or older. Statin treatment is therefore not calculated for younger age groups.
[3]Data are point estimates with 95% CIs.

Figure S25: Hypertension treatment, hypertension control, diabetes and statin treatment by poverty level (all age groups)[1,2,3]

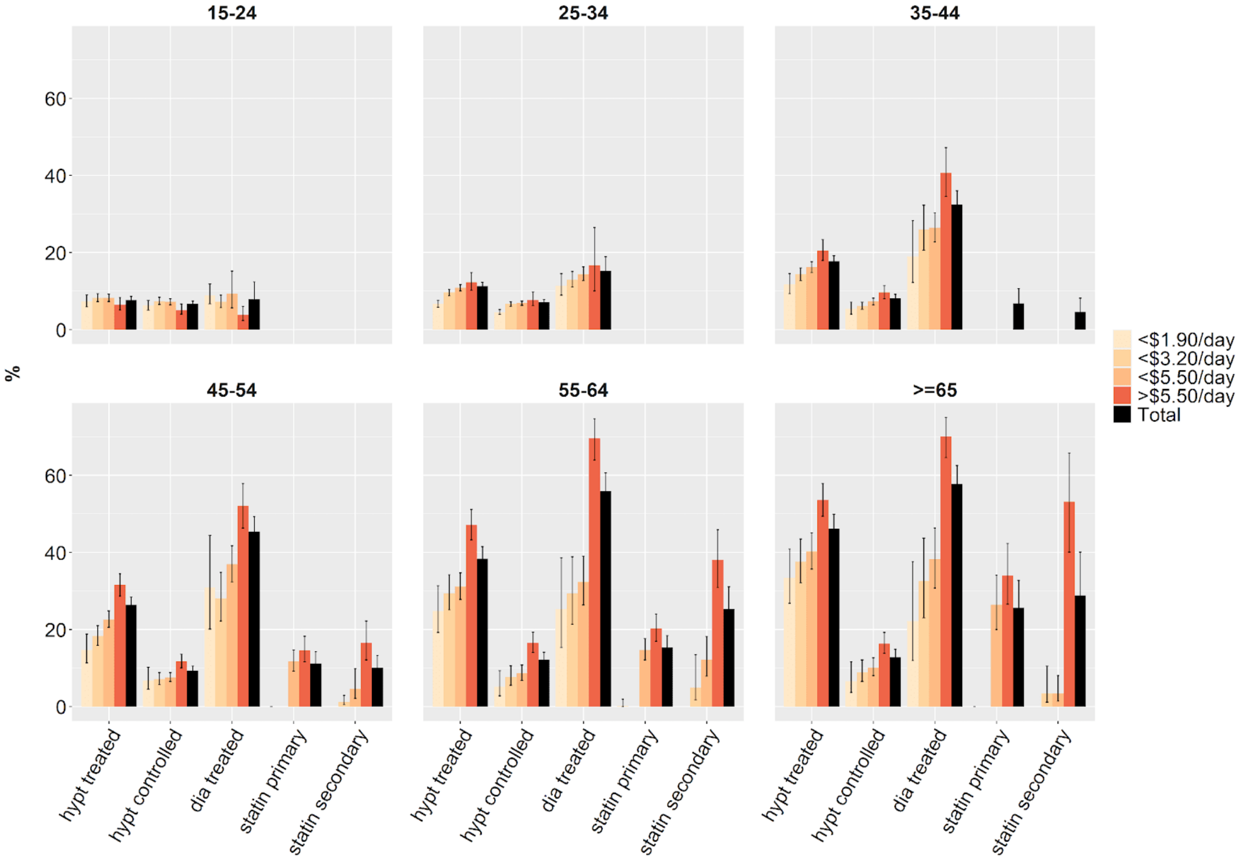

[1]Prevalences were not calculated for samples with less than 50 individuals.
[2]According to WHO guidelines, statin use is recommended for individuals aged 40 years or older. Statin treatment is therefore not calculated for younger age groups.
[3]Data are point estimates with 95% CIs.

Figure S26: Hypertension treatment, hypertension control, diabetes and statin treatment by poverty level (age groups 45-54; 55-64; 65+), females only[1,2,3]

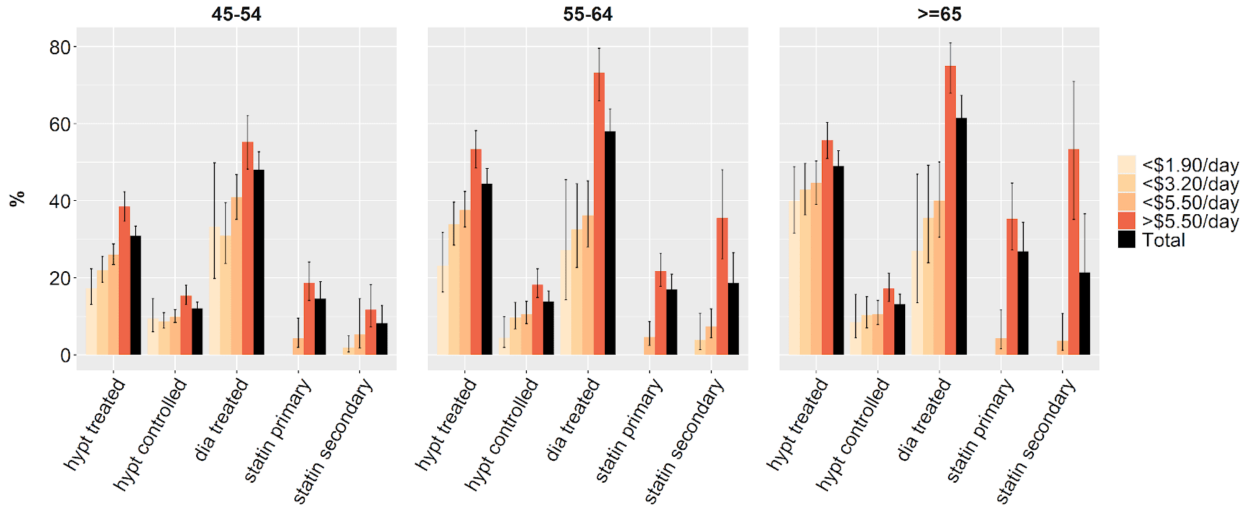

[1]Prevalences were not calculated for samples with less than 50 individuals.
[2]According to WHO guidelines, statin use is recommended for individuals aged 40 years or older. Statin treatment is therefore not calculated for younger age groups.
[3]Data are point estimates with 95% CIs.

Figure S27: Hypertension treatment, hypertension control, diabetes and statin treatment by poverty level (all age groups), females only[1,2,3]

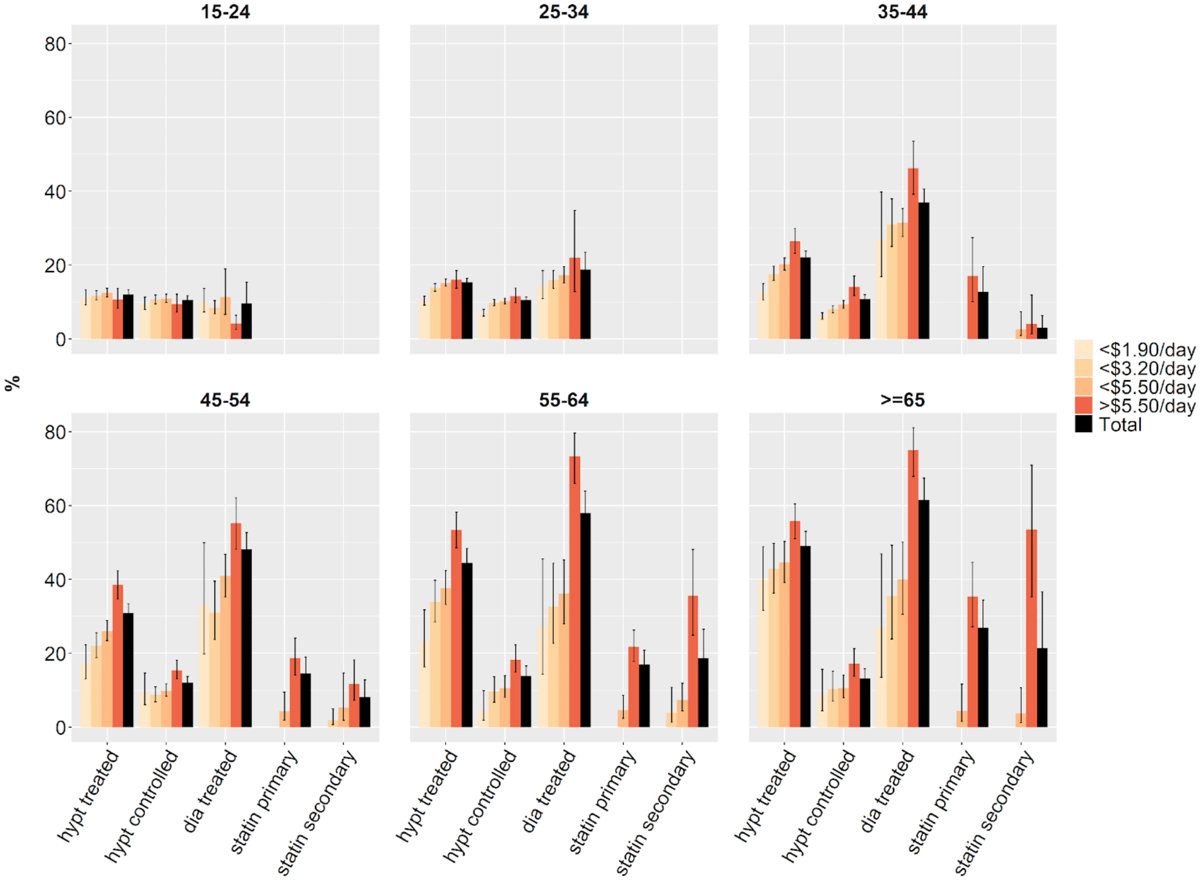

[1]Prevalences were not calculated for samples with less than 50 individuals.
[2]According to WHO guidelines, statin use is recommended for individuals aged 40 years or older. Statin treatment is therefore not calculated for younger age groups.
[3]Data are point estimates with 95% CIs.

Table S30: Prevalences of the care cascade for hypertension, diabetes and statins, males only[1,2]

| Age group[3] | Percentage using blood pressure lowering medication ("Hypt treated") | Percentage using blood pressure lowering medication and normal BP ("Hypt controlled") | Percentage using medication for diabetes ("Dia treated") | Percentage using statins for primary prevention ("Statin primary") | Percentage using statins for secondary prevention ("Statin secondary") |
|---|---|---|---|---|---|
| **All age groups** | | | | | |
| Total | 19.4 (17.6-21.1) | 6.3 (5.5-7.1) | 37.5 (34.2-40.9) | 11.7 (9.2-14.2) | 19.1 (15.6-22.7) |
| <$1.90/day | 11.7 (8.6-14.8) | 3.1 (1.6-4.6) | 13.5 (7.8-19.2) | - | - |
| <$3.20/day | 12.3 (10.3-14.4) | 3.7 (2.6-4.7) | 17.3 (12.8-21.8) | - | 1.6 (0.0-4.1) |
| <$5.50/day | 14.7 (13.1-16.4) | 4.4 (3.5-5.2) | 21.8 (18.4-25.2) | 1.7 (0.2-3.3) | 7.1 (3.4-10.7) |
| >$5.50/day | 25.1 (22.5-27.8) | 8.7 (7.4-9.9) | 49.0 (43.5-54.5) | 14.4 (11.2-17.4) | 29.7 (23.5-35.8) |
| **15-24** | | | | | |
| Total | 2.7 (2.1-3.5) | 2.0 (1.5-2.7) | 3.3 (2.0 -5.2) | - | - |
| <$1.90/day | 1.6 (0.8-3.1) | 1.4 (0.7-2.8) | 3.9 (2.3-6.6) | - | - |
| <$3.20/day | 2.4 (1.7-3.5) | 2.0 (1.3-3.0) | 5.3 (2.9-9.7) | - | - |
| <$5.50/day | 2.2 (1.6-3.0) | 1.9 (1.3-2.6) | 3.3 (2.1-5.3) | - | - |
| >$5.50/day | 3.7 (2.4-5.6) | 2.2 (1.3-3.7) | 3.1 (1.0 -9.2) | - | - |
| **25-34** | | | | | |
| Total | 6.3 (4.8-8.3) | 3.0 (2.1-4.4) | 7.3 (4.9-10.9) | - | - |
| <$1.90/day | 1.6 (0.9-2.5) | 1.0 (0.5-1.8) | 6.4 (4.2-9.6) | - | - |
| <$3.20/day | 2.8 (2.1-3.6) | 1.8 (1.3-2.4) | 3.8 (2.4-6.1) | - | - |
| <$5.50/day | 4.3 (3.1-5.9) | 1.9 (1.5-2.5) | 7.1 (5.1-9.7) | - | - |
| >$5.50/day | 9.6 (6.4-14.0) | 4.8 (2.8-8.4) | 7.7 (3.4-16.5) | - | - |
| **35-44** | | | | | |
| Total | 11.6 (9.5-14.1) | 4.4 (3.3-5.8) | 25.2 (19.4-32.1) | - | 7.1 (3.1-15.4) |
| <$1.90/day | 9.8 (4.8-18.9) | 3.9 (1.2-11.5) | 18.0 (9.6-31.2) | - | - |
| <$3.20/day | 7.6 (5.0-11.6) | 2.5 (1.3-4.7) | 7.1 (3.3-14.8) | - | - |
| <$5.50/day | 9.1 (6.8-11.9) | 3.7 (2.4-5.7) | 16.8 (10.7-25.4) | - | - |
| >$5.50/day | 14.8 (11.5-18.9) | 5.2 (3.7-7.3) | 34.0 (24.2-45.3) | - | 14.6 (6.5-29.6) |
| **45-54** | | | | | |

| | | | | | |
|---|---|---|---|---|---|
| Total | 20.9 (18.3-23.8) | 6.0 (4.8-7.5) | 42.2 (36.6-48.1) | 9.6 (6.5-13.9) | 12.4 (8.3-18.0) |
| <$1.90/day | 10.9 (6.2-18.3) | 2.9 (0.9-8.5) | 21.4 (14.1-31.1) | - | - |
| <$3.20/day | 12.6 (9.3-16.9) | 4.7 (2.7-8.0) | 26.0 (11.4-48.9) | - | - |
| <$5.50/day | 17.8 (14.8-21.3) | 4.2 (3.0-6.0) | 30.3 (23.2-38.4) | - | 3.5 (1.1-10.8) |
| >$5.50/day | 24.4 (20.6-28.6) | 7.9 (5.9-10.5) | 49.1 (41.4-56.9) | 9.6 (6.5-13.9) | 22.6 (14.5-33.3) |
| **55-64** | | | | | |
| Total | 31.7 (28.2-35.4) | 10.2 (8.3-12.4) | 53.7 (47.6-59.7) | 13.1 (9.8-17.2) | 32.3 (25.3-40.2) |
| <$1.90/day | 26.8 (18.4-37.3) | 6.0 (2.4-14.2) | 24.2 (14.4-37.7) | - | - |
| <$3.20/day | 23.9 (18.4-30.5) | 5.2 (2.7-9.7) | 22.4 (9.5-44.1) | - | - |
| <$5.50/day | 23.5 (19.5-28.0) | 6.2 (4.2-9.1) | 27.0 (19.3-36.4) | 2.7 (0.9-7.6) | 20.0 (11.2-33.2) |
| >$5.50/day | 41.0 (35.9-46.3) | 14.6 (11.6-18.3) | 66.1 (59.0 -72.6) | 18.1 (13.4-24.0) | 39.8 (30.8-49.5) |
| **>65** | | | | | |
| Total | 43.0 (38.4-47.7) | 12.4 (10.2-15.2) | 56.6 (49.8-63.1) | 32.3 (21.1-46.1) | 38.7 (25.5-53.8) |
| <$1.90/day | 22.4 (14.3-33.1) | 3.4 (1.0-10.8) | 36.6 (18.4-59.8) | - | - |
| <$3.20/day | 29.8 (22.7-37.9) | 6.6 (3.7-11.6) | - | - | - |
| <$5.50/day | 34.5 (28.8-40.7) | 9.5 (6.6-13.7) | 41.7 (30.4-53.9) | - | 2.4 (0.7-8.0) |
| >$5.50/day | 51.8 (46.0-57.5) | 15.5 (12.2-19.3) | 64.2 (56.9-71.0 ) | 32.3 (21.0-46.1) | 53.0 (34.2-70.9) |

[1]Prevalences were not calculated for samples with less than 50 individuals.
[2]According to WHO guidelines, statin use is recommended for individuals aged 40 years or older. Statin treatment is therefore not calculated for younger age groups.

Figure S28: Hypertension treatment, hypertension control, diabetes and statin treatment by poverty level (age groups 45-54; 55-64; 65+), males only[1,2,3]

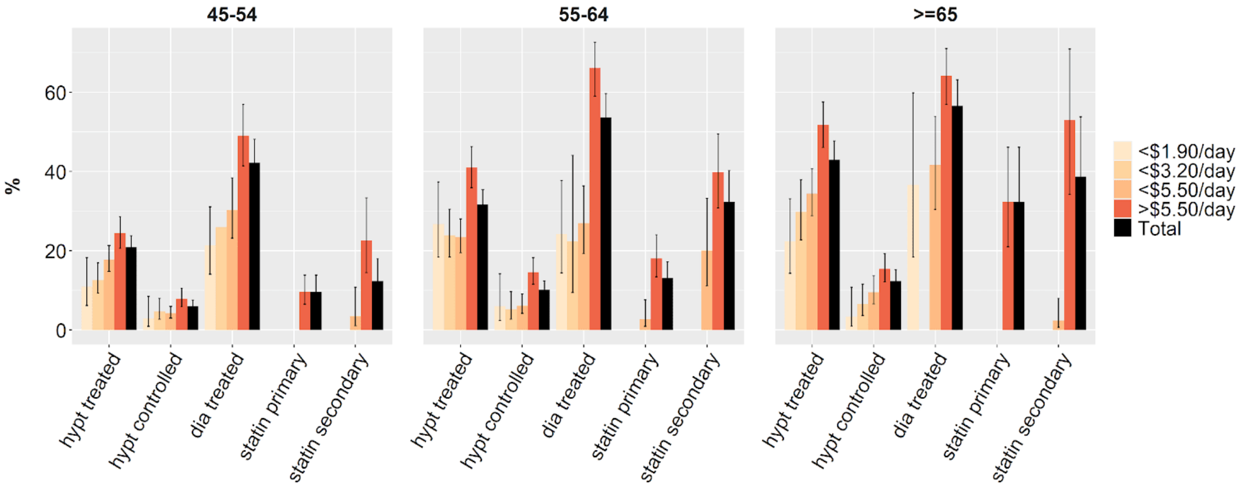

[1]Prevalences were not calculated for samples with less than 50 individuals.
[2]According to WHO guidelines, statin use is recommended for individuals aged 40 years or older. Statin treatment is therefore not calculated for younger age groups.
[3]Data are point estimates with 95% CIs.

Figure S29: Hypertension treatment, hypertension control, diabetes and statin treatment by poverty level (all age groups), males only[1,2,3]

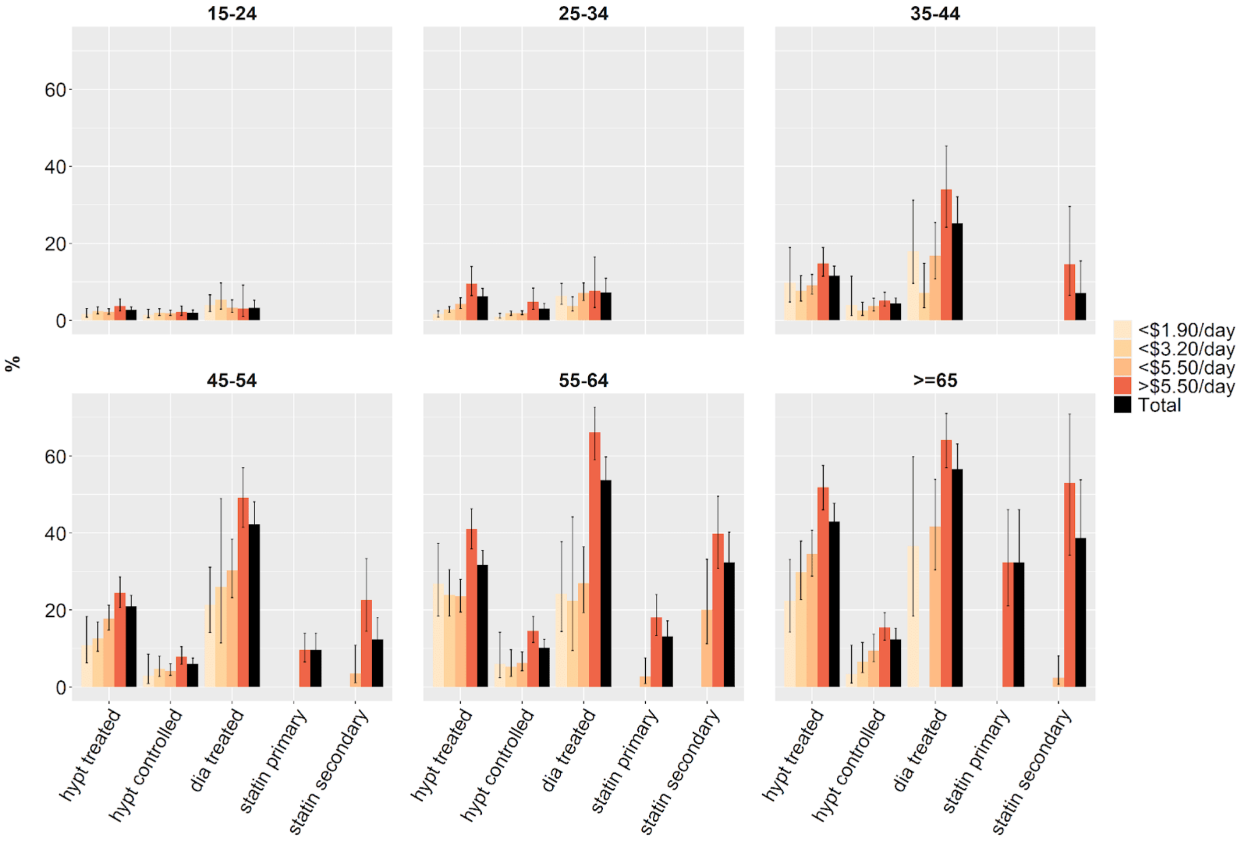

[1]Prevalences were not calculated for samples with less than 50 individuals.
[2]According to WHO guidelines, statin use is recommended for individuals aged 40 years or older. Statin treatment is therefore not calculated for younger age groups.
[3]Data are point estimates with 95% CIs.

Figure S30: Hypertension treatment, hypertension control, and diabetes treatment poor vs non poor[1]

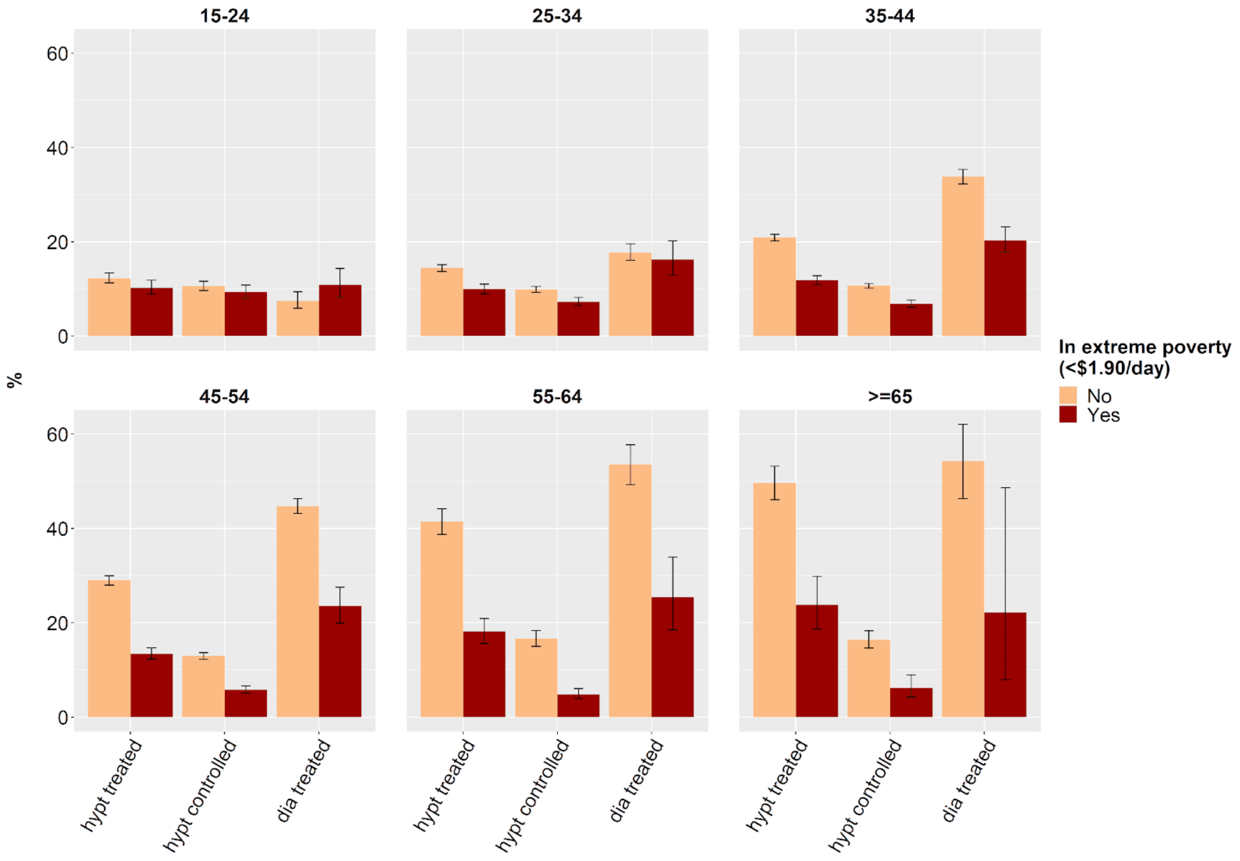

[1]Data are point estimates with 95% CIs.

Figure S31: Hypertension treatment, hypertension control, and diabetes treatment for females, poor vs non poor[1]

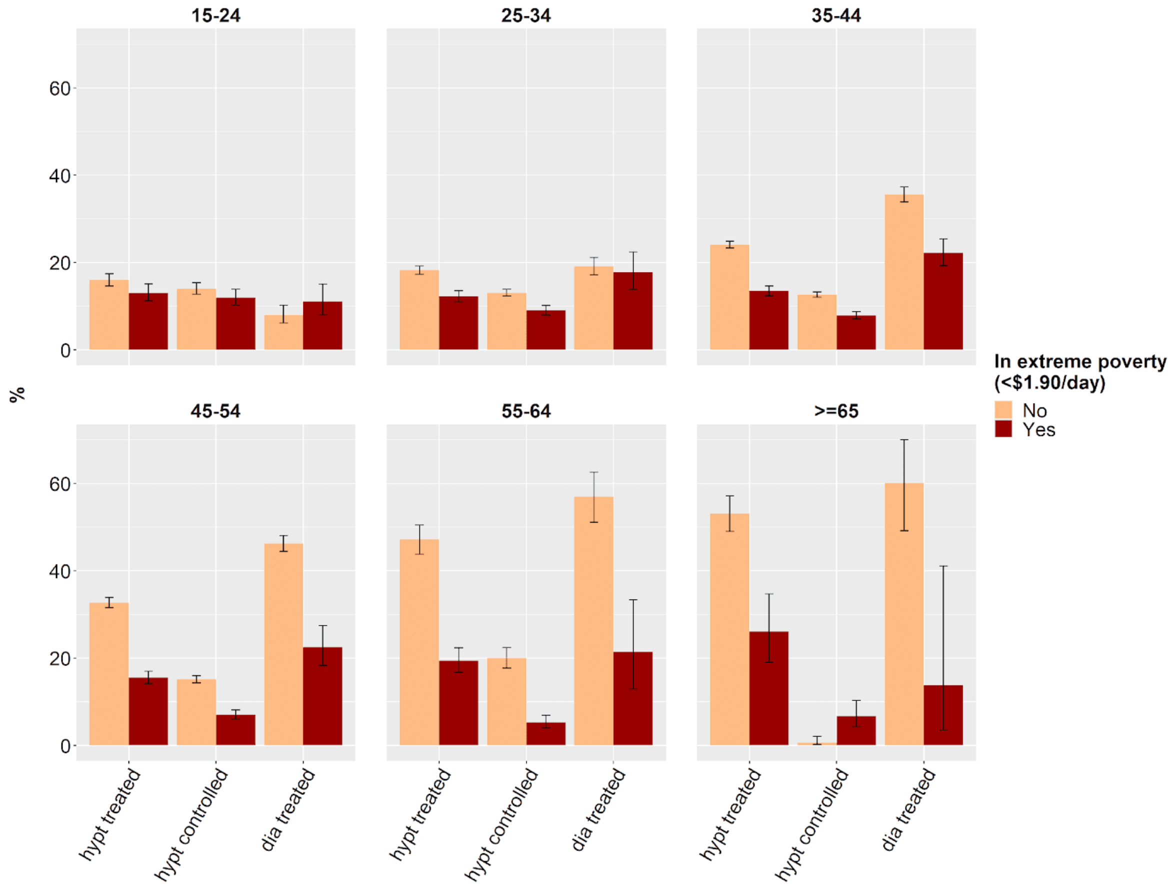

[1]Data are point estimates with 95% CIs.

Figure S32: Hypertension treatment, hypertension control, and diabetes treatment for males, poor vs non poor[1, 2]

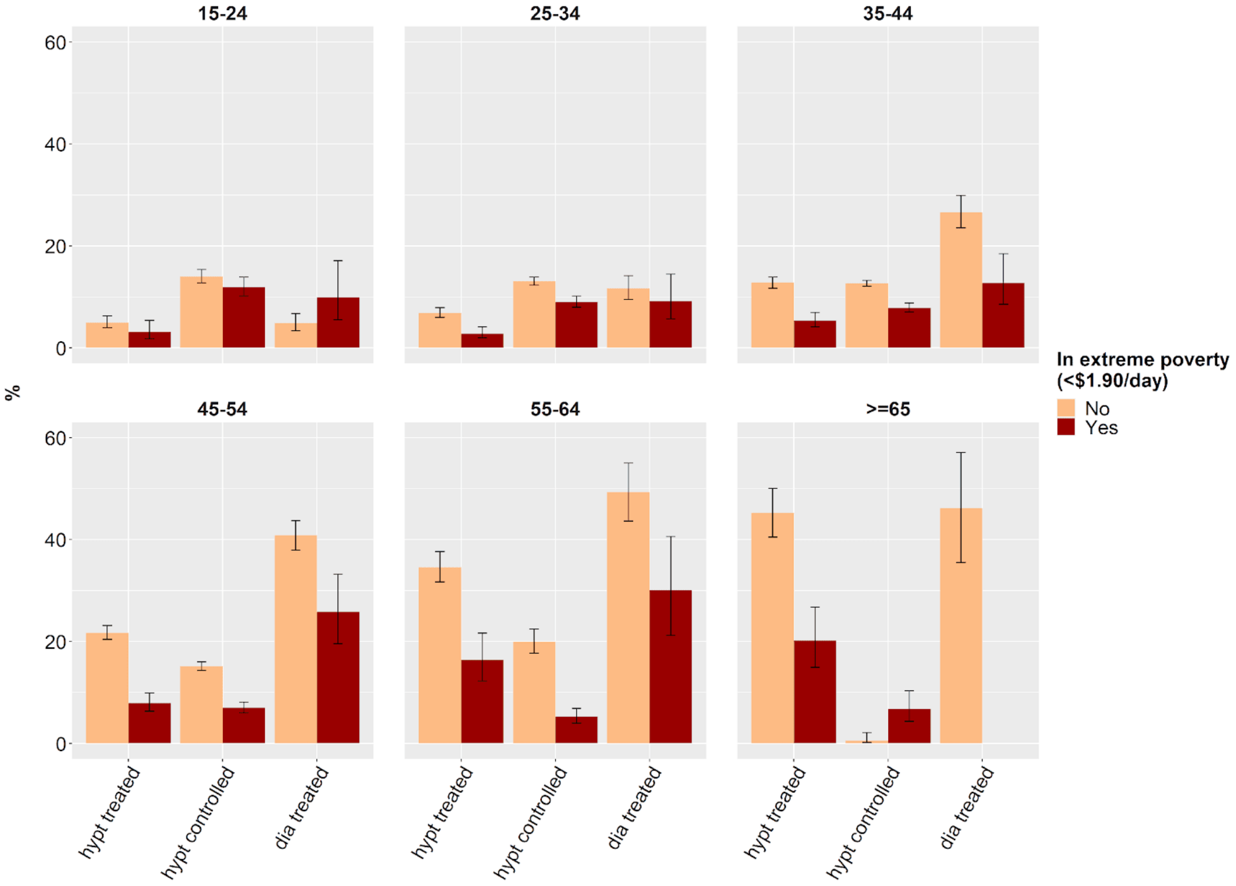

[1]Sample size for diabetes treatment for men in poverty aged 65 or older was insufficient for calculations, as samples of <50 individuals were excluded.
[2]Data are point estimates with 95% CIs.

Table S31: Poisson regressions for hypertension onto individual-level characteristics (with survey weights), urban sample: Poverty headcount ratio <$1.90/day to poverty headcount ratio <$5.50/day [c]

| | <$1.90/day | | | | | <$3.20/day | | | | | <$5.50/day | | | | |
|---|---|---|---|---|---|---|---|---|---|---|---|---|---|---|---|
| **Variable** | **RR (95% CI)** | ***P* value** | **AME (95% CI)** | ***P* value** | **Obs** | **RR (95% CI)** | ***P* value** | **AME (95% CI)** | ***P* value** | **Obs** | **RR (95% CI)** | ***P* value** | **AME (95% CI)** | ***P* value** | **Obs** |
| **Countries** | **29** | | | | | **29** | | | | | **29** | | | | |
| **Covariate-unadjusted regressions[a]** | | | | | | | | | | | | | | | |
| Sex | | | | | | | | | | | | | | | |
| Female | 1 (ref.) | | 1 (ref.) | | 159318 | 1 (ref.) | | 1 (ref.) | | 416972 | 1 (ref.) | | 1 (ref.) | | 614304 |
| Male | 1.08 (1.00-1.17) | 0.058 | 0.01 (0.00-0.02) | 0.0625 | 34840 | 1.06 (1.01-1.12) | 0.018 | 0.01 (0.00-0.02) | 0.020 | 92803 | 1.12 (1.07-1.17) | 0.000 | 0.02 (0.01-0.03) | 0.000 | 145307 |
| Place of residence | | | | | | | | | | | | | | | |
| Rural | 1 (ref.) | | 1 (ref.) | | 179212 | 1 (ref.) | | 1 (ref.) | | 435224 | 1 (ref.) | | 1 (ref.) | | 568471 |
| Urban | 1.03 (0.88-1.21) | 0.678 | 0.01 (-0.02-0.03) | 0.6820 | 14946 | 1.09 (1.01-1.18) | 0.024 | 0.02 (0.00-0.03) | 0.028 | 74551 | 1.09 (1.03-1.15) | 0.003 | 0.02 (0.00-0.02) | 0.003 | 191140 |
| Education | | | | | | | | | | | | | | | |
| No schooling | 1 (ref.) | | 1 (ref.) | | 92230 | 1 (ref.) | | 1 (ref.) | | 175761 | 1 (ref.) | | 1 (ref.) | | 209979 |
| Primary education | 1.08 (0.99-1.17) | 0.075 | 0.01 (0.00-0.02) | 0.0738 | 42301 | 1.08 (1.02-1.15) | 0.005 | 0.01 (0.00-0.02) | 0.005 | 103310 | 1.11 (1.05-1.16) | 0.000 | 0.02 (0.01-0.02) | 0.000 | 143198 |
| Secondary education or further | 0.98 (0.87-1.10) | 0.678 | 0.00 (-0.02-0.02) | 0.6769 | 59206 | 1.07 (1.01-1.13) | 0.018 | 0.01 (0.00-0.02) | 0.018 | 229555 | 1.13 (1.08-1.18) | 0.000 | 0.02 (0.01-0.03) | 0.000 | 404628 |
| **Covariate-adjusted regressions[b]** | | | | | | | | | | | | | | | |
| Sex | | | | | | | | | | | | | | | |
| Female | 1 (ref.) | | 1 (ref.) | | 159023 | 1 (ref.) | | 1 (ref.) | | 416143 | 1 (ref.) | | 1 (ref.) | | 612998 |
| Male | 1.08 (1.00-1.18) | 0.065 | 0.01 (0.00-0.03) | 0.0696 | 34714 | 1.06 (1.00-1.11) | 0.054 | 0.01 (0.00-0.02) | 0.058 | 92483 | 1.11 (1.05-1.16) | 0.000 | 0.02 (0.01-0.02) | 0.000 | 144807 |
| Place of residence | | | | | | | | | | | | | | | |
| Rural | 1 (ref.) | | 1 (ref.) | | 178832 | 1 (ref.) | | 1 (ref.) | | 434322 | 1 (ref.) | | 1 (ref.) | | 190607 |
| Urban | 1.04 (0.89-1.22) | 0.608 | 0.01 (-0.02-0.03) | 0.6138 | 14905 | 1.09 (1.01-1.18) | 0.028 | 0.01 (0.00-0.03) | 0.032 | 74304 | 1.08 (1.02-1.15) | 0.007 | 0.01 (0.00-0.02) | 0.008 | 567198 |
| Education | | | | | | | | | | | | | | | |

| | | | | | | | | | | | | | | | |
|---|---|---|---|---|---|---|---|---|---|---|---|---|---|---|---|
| No schooling | 1 (ref.) | | 1 (ref.) | | 92230 | 1 (ref.) | | 1 (ref.) | | 175761 | 1 (ref.) | | 1 (ref.) | | 209979 |
| Primary education | 1.06 (0.97-1.15) | 0.210 | 0.01 (-0.01-0.02) | 0.2082 | 42301 | 1.07 (1.01-1.13) | 0.023 | 0.01 (0.00-0.02) | 0.023 | 103310 | 1.08 (1.03-1.14) | 0.002 | 0.01 (0.00-0.02) | 0.002 | 143198 |
| Secondary education or further | 0.95 (0.84-1.07) | 0.364 | -0.01 (-0.03-0.01) | 0.3598 | 59206 | 1.04 (0.98-1.11) | 0.170 | 0.01 (0.00-0.02) | 0.169 | 229555 | 1.08 (1.03-1.13) | 0.003 | 0.01 (0.00-0.02) | 0.003 | 404628 |
| Observations[d] | 193,737 | | | | | 508,626 | | | | | 757,805 | | | | |

Abbreviations: RR=risk ratio; AME=average marginal effect. RRs and AMEs are shown with 95 CIs which are adjusted for clustering at the levels of the primary and secondary sampling unit.

[a] Included only one of the variables shown in the table and a binary indicator for each country (country-level fixed effects).

[b] Included age, sex, education, place of residence (urban sample only) and a binary indicator for each country (country-level fixed effects).

[c] All regressions are age adjusted with age as continuous variable with restricted cubic splines with five knots (placed at the fifth, 27.5th, 50th, 72.5th, and 95th percentiles).

[d] Number of observations in covariate-adjusted regression model.

Table S32: Poisson regressions for hypertension onto individual-level characteristics (with survey weights), urban sample: Poverty headcount ratio >$5.50/day to all poverty levels[c]

| | **>$5.50/day** | | | | | **Total** | | | | |
|---|---|---|---|---|---|---|---|---|---|---|
| **Variable** | **RR (95% CI)** | ***P* value** | **AME (95% CI)** | ***P* value** | **Obs.** | **RR (95% CI)** | ***P* value** | **AME (95% CI)** | ***P* value** | **Obs.** |
| **Countries** | **29** | | | | | **29** | | | | |
| **Covariate-unadjusted regressions[a]** | | | | | | | | | | |
| Sex | | | | | | | | | | |
| Female | 1 (ref.) | | 1 (ref.) | | 159566 | 1 (ref.) | | 1 (ref.) | | 773870 |
| Male | 1.23 (1.17-1.31) | 0.000 | 0.04 (0.04-0.06) | 0.000 | 76147 | 1.16 (1.12-1.21) | 0.000 | 0.03 (0.02-0.03) | 0.000 | 221454 |
| Place of residence | | | | | | | | | | |
| Rural | 1 (ref.) | | 1 (ref.) | | 59082 | 1 (ref.) | | 1 (ref.) | | 627553 |
| Urban | 0.93 (0.85-1.02) | 0.125 | -0.01 (-0.04-0.00) | 0.130 | 176631 | 1.04 (0.99-1.10) | 0.132 | 0.01 (0.00-0.02) | 0.132 | 367771 |
| Education | | | | | | | | | | |
| No schooling | 1 (ref.) | | 1 (ref.) | | 15337 | 1 (ref.) | | 1 (ref.) | | 225316 |
| Primary education | 0.99 (0.90-1.10) | 0.900 | 0.00 (-0.02-0.02) | 0.900 | 44100 | 1.11 (1.06-1.16) | 0.000 | 0.02 (0.01-0.03) | 0.000 | 187298 |
| Secondary education or further | 0.92 (0.82-1.04) | 0.188 | -0.02 (-0.04-0.01) | 0.202 | 175454 | 1.09 (1.04-1.15) | 0.000 | 0.02 (0.01-0.02) | 0.000 | 580082 |
| **Covariate-adjusted regressions[b]** | | | | | | | | | | |
| Sex | | | | | | | | | | |
| Female | 1 (ref.) | | 1 (ref.) | | 159063 | 1 (ref.) | | 1 (ref.) | | 772061 |
| Male | 1.26 (1.18-1.33) | 0.000 | 0.05 (0.04-0.07) | 0.000 | 75828 | 1.16 (1.11-1.21) | 0.000 | 0.03 (0.02-0.03) | 0.000 | 220635 |

| | | | | | | | | | | |
|---|---|---|---|---|---|---|---|---|---|---|
| Place of residence | | | | | | | | | | |
| Rural | 1 (ref.) | | 1 (ref.) | | 58842 | 1 (ref.) | | 1 (ref.) | | 626040 |
| Urban | 0.96 (0.88-1.05) | 0.360 | -0.01 (-0.03-0.01) | 0.363 | 176049 | 1.04 (0.99-1.10) | 0.121 | 0.01 (0.00-0.02) | 0.121 | 366656 |
| Education | | | | | | | | | | |
| No schooling | 1 (ref.) | | 1 (ref.) | | 15337 | 1 (ref.) | | 1 (ref.) | | 225316 |
| Primary education | 0.95 (0.86-1.06) | 0.357 | -0.01 (-0.04-0.01) | 0.366 | 44100 | 1.08 (1.03-1.13) | 0.001 | 0.01 (0.00-0.02) | 0.001 | 187298 |
| Secondary education or further | 0.87 (0.77-0.98) | 0.020 | -0.03 (-0.06--0.01) | 0.028 | 175454 | 1.04 (0.99-1.09) | 0.168 | 0.01 (0.00-0.02) | 0.163 | 580082 |
| Observations[d] | 234,891 | | | | | 992,696 | | | | |

Abbreviations: RR=risk ratio; AME=average marginal effect; NA=not applicable. RRs and AMEs are shown with 95 CIs which are adjusted for clustering at the levels of the primary and secondary sampling unit.

[a] Included only one of the variables shown in the table and a binary indicator for each country (country-level fixed effects).

[b] Included age, sex, education, place of residence (urban sample only) and a binary indicator for each country (country-level fixed effects).

[c] All regressions are age adjusted with age as continuous variable with restricted cubic splines with five knots (placed at the fifth, 27.5$^{th}$, 50$^{th}$, 72.5$^{th}$, and 95$^{th}$ percentiles).

[d] Number of observations in covariate-adjusted regression model.

Table S33: Poisson regressions for hypertension onto individual-level characteristics (with survey weights), non-urban sample: Poverty headcount ratio <$1.90/day to poverty headcount ratio <$5.50/day [c]

| | <$1.90/day | | | | | <$3.20/day | | | | | <$5.50/day | | | | |
|---|---|---|---|---|---|---|---|---|---|---|---|---|---|---|---|
| **Variable** | **RR (95% CI)** | ***P* value** | **AME (95% CI)** | ***P* value** | **Obs.** | **RR (95% CI)** | ***P* value** | **AME (95% CI)** | ***P* value** | **Obs.** | **RR (95% CI)** | ***P* value** | **AME (95% CI)** | ***P* value** | **Obs.** |
| **Countries** | **50** | | | | | **50** | | | | | **50** | | | | |
| **Covariate-unadjusted regressions[a]** | | | | | | | | | | | | | | | |
| Sex | | | | | | | | | | | | | | | |
| Female | 1 (ref.) | | 1 (ref.) | | 165709 | 1 (ref.) | | 1 (ref.) | | 430747 | 1 (ref.) | | 1 (ref.) | | 639672 |
| Male | 1.09 (1.01-1.17) | 0.025 | 0.02 (0.00-0.03) | 0.0276 | 39320 | 1.07 (1.02-1.12) | 0.008 | 0.01 (0.00-0.02) | 0.009 | 102949 | 1.11 (1.07-1.16) | 0.000 | 0.02 (0.01-0.03) | 0.000 | 164687 |
| Education | | | | | | | | | | | | | | | |
| No schooling | 1 (ref.) | | 1 (ref.) | | 95643 | 1 (ref.) | | 1 (ref.) | | 181894 | 1 (ref.) | | 1 (ref.) | | 218779 |
| Primary education | 1.08 (0.99-1.16) | 0.072 | 0.01 (0.00-0.02) | 0.0712 | 47173 | 1.08 (1.02-1.14) | 0.005 | 0.01 (0.00-0.02) | 0.005 | 113986 | 1.10 (1.05-1.15) | 0.000 | 0.02 (0.01-0.02) | 0.000 | 161630 |
| Secondary education or further | 0.97 (0.87-1.09) | 0.620 | 0.00 (-0.02-0.01) | 0.6182 | 61759 | 1.06 (1.00-1.12) | 0.037 | 0.01 (0.00-0.02) | 0.037 | 236569 | 1.11 (1.06-1.16) | 0.000 | 0.02 (0.01-0.02) | 0.000 | 422010 |
| **Covariate-adjusted regressions[b]** | | | | | | | | | | | | | | | |
| Sex | | | | | | | | | | | | | | | |
| Female | 1 (ref.) | | 1 (ref.) | | 165392 | 1 (ref.) | | 1 (ref.) | | 429853 | 1 (ref.) | | 1 (ref.) | | 638280 |
| Male | 1.09 (1.01-1.18) | 0.026 | 0.02 (0.00-0.03) | 0.0288 | 39183 | 1.06 (1.01-1.11) | 0.029 | 0.01 (0.00-0.02) | 0.031 | 102596 | 1.10 (1.05-1.15) | 0.000 | 0.02 (0.01-0.02) | 0.000 | 164139 |
| Education | | | | | | | | | | | | | | | |
| No schooling | 1 (ref.) | | 1 (ref.) | | 95643 | 1 (ref.) | | 1 (ref.) | | 181894 | 1 (ref.) | | 1 (ref.) | | 218779 |
| Primary education | 1.05 (0.97-1.14) | 0.212 | 0.01 (-0.01-0.02) | 0.2104 | 47173 | 1.07 (1.01-1.13) | 0.018 | 0.01 (0.00-0.02) | 0.018 | 113986 | 1.08 (1.03-1.13) | 0.001 | 0.01 (0.00-0.02) | 0.001 | 161630 |
| Secondary education or further | 0.95 (0.85-1.06) | 0.323 | -0.01 (-0.03-0.01) | 0.3186 | 61759 | 1.04 (0.98-1.10) | 0.156 | 0.01 (0.00-0.02) | 0.154 | 236569 | 1.08 (1.04-1.13) | 0.001 | 0.01 (0.00-0.02) | 0.000 | 422010 |
| Observations[d] | 204,575 | | | | | 532,449 | | | | | 802,419 | | | | |

Abbreviations: RR=risk ratio; AME=average marginal effect; NA=not applicable. RRs and AMEs are shown with 95 CIs which are adjusted for clustering at the levels of the primary and secondary sampling unit.

[a] Included only one of the variables shown in the table and a binary indicator for each country (country-level fixed effects).

[b] Included age, sex, education, place of residence (urban sample only) and a binary indicator for each country (country-level fixed effects).

[c] All regressions are age adjusted with age as continuous variable with restricted cubic splines with five knots (placed at the fifth, 27.5$^{th}$, 50$^{th}$, 72.5$^{th}$, and 95$^{th}$ percentiles).

[d] Number of observations in covariate-adjusted regression model.

Table S34: Poisson regressions for hypertension onto individual-level characteristics (with survey weights), non-urban sample: Poverty headcount ratio >$5.50/day to all poverty levels[c]

| | >$5.50/day | | | | | Total | | | | |
|---|---|---|---|---|---|---|---|---|---|---|
| **Variable** | **RR (95% CI)** | ***P* value** | **AME (95% CI)** | ***P* value** | **Obs.** | **RR (95% CI)** | ***P* value** | **AME (95% CI)** | ***P* value** | **Obs.** |
| **Countries** | **50** | | | | | **50** | | | | |
| **Covariate-unadjusted regressions[a]** | | | | | | | | | | |
| Sex | | | | | | | | | | |
| Female | 1 (ref.) | | 1 (ref.) | | 175710 | 1 (ref.) | | 1 (ref.) | | 815382 |
| Male | 1.23 (1.17-1.30) | 0.000 | 0.04 (0.04-0.06) | 0.000 | 90931 | 1.16 (1.12-1.20) | 0.000 | 0.03 (0.02-0.03) | 0.000 | 255618 |
| Education | | | | | | | | | | |
| No schooling | 1 (ref.) | | 1 (ref.) | | 17042 | 1 (ref.) | | 1 (ref.) | | 235821 |
| Primary education | 0.99 (0.90-1.09) | 0.792 | 0.00 (-0.02-0.02) | 0.793 | 52494 | 1.10 (1.06-1.15) | 0.000 | 0.02 (0.01-0.02) | 0.000 | 214124 |
| Secondary education or further | 0.93 (0.84-1.04) | 0.199 | -0.01 (-0.04-0.01) | 0.211 | 196258 | 1.09 (1.04-1.14) | 0.000 | 0.02 (0.01-0.02) | 0.000 | 618268 |
| **Covariate-adjusted regressions[b]** | | | | | | | | | | |
| Sex | | | | | | | | | | |
| Female | 1 (ref.) | | 1 (ref.) | | 175189 | 1 (ref.) | | 1 (ref.) | | 813469 |
| Male | 1.25 (1.19-1.32) | 0.000 | 0.05 (0.04-0.07) | 0.000 | 90605 | 1.15 (1.11-1.20) | 0.000 | 0.03 (0.02-0.03) | 0.000 | 254744 |
| Education | | | | | | | | | | |
| No schooling | 1 (ref.) | | 1 (ref.) | | 17042 | 1 (ref.) | | 1 (ref.) | | 235821 |
| Primary education | 0.95 (0.86-1.04) | 0.255 | -0.01 (-0.04-0.01) | 0.266 | 52494 | 1.07 (1.03-1.12) | 0.001 | 0.01 (0.00-0.02) | 0.001 | 214124 |

| | | | | | | | | | | |
|---|---|---|---|---|---|---|---|---|---|---|
| Secondary education or further | 0.87 (0.78-0.97) | 0.013 | -0.03 (-0.06--0.01) | 0.019 | 196258 | 1.05 (1.00-1.09) | 0.063 | 0.01 (0.00-0.02) | 0.059 | 618268 |
| Observations[d] | 265,794 | | | | | 1,068,213 | | | | |

Abbreviations: RR=risk ratio; AME=average marginal effect; NA=not applicable. RRs and AMEs are shown with 95 CIs which are adjusted for clustering at the levels of the primary and secondary sampling unit.

[a] Included only one of the variables shown in the table and a binary indicator for each country (country-level fixed effects).

[b] Included age, sex, education, place of residence (urban sample only) and a binary indicator for each country (country-level fixed effects).

[c] All regressions are age adjusted with age as continuous variable with restricted cubic splines with five knots (placed at the fifth, 27.5$^{th}$, 50$^{th}$, 72.5$^{th}$, and 95$^{th}$ percentiles).

[d] Number of observations in covariate-adjusted regression model.

Table S35: Poisson regressions for diabetes onto individual-level characteristics (with survey weights), urban sample: Poverty headcount ratio <$1.90/day to poverty headcount ratio <$5.50/day [c]

| | **<$1.90/day** | | | | | **<$3.20/day** | | | | | **<$5.50/day** | | | | |
|---|---|---|---|---|---|---|---|---|---|---|---|---|---|---|---|
| **Variable** | **RR (95% CI)** | ***P* value** | **AME (95% CI)** | ***P* value** | **Obs.** | **RR (95% CI)** | ***P* value** | **AME (95% CI)** | ***P* value** | **Obs.** | **RR (95% CI)** | ***P* value** | **AME (95% CI)** | ***P* value** | **Obs.** |
| **Countries** | **24** | | | | | **24** | | | | | **24** | | | | |
| **Covariate-unadjusted regressions[a]** | | | | | | | | | | | | | | | |
| Sex | | | | | | | | | | | | | | | |
| Female | 1 (ref.) | | 1 (ref.) | | 154666 | 1 (ref.) | | 1 (ref.) | | 405169 | 1 (ref.) | | 1 (ref.) | | 591225 |
| Male | 0.98 (0.76-1.25) | 0.845 | 0.00 (-0.01-0.01) | 0.844 | 29709 | 1.03 (0.89-1.20) | 0.679 | 0.00 (0.00-0.01) | 0.681 | 79511 | 1.13 (1.01-1.26) | 0.033 | 0.01 (0.00-0.01) | 0.036 | 120209 |
| Place of residence | | | | | | | | | | | | | | | |
| Rural | 1 (ref.) | | 1 (ref.) | | 171279 | 1 (ref.) | | 1 (ref.) | | 417518 | 1 (ref.) | | 1 (ref.) | | 172925 |
| Urban | 1.47 (1.08-2.00) | 0.014 | 0.02 (0.00-0.03) | 0.029 | 13096 | 1.46 (1.23-1.74) | 0.000 | 0.02 (0.01-0.02) | 0.000 | 67162 | 1.57 (1.42-1.75) | 0.000 | 0.02 (0.02-0.03) | 0.000 | 538509 |
| Education | | | | | | | | | | | | | | | |
| No schooling | 1 (ref.) | | 1 (ref.) | | 89499 | 1 (ref.) | | 1 (ref.) | | 170149 | 1 (ref.) | | 1 (ref.) | | 200761 |
| Primary education | 1.25 (1.00-1.56) | 0.051 | 0.01 (0.00-0.01) | 0.057 | 37148 | 1.09 (0.95-1.26) | 0.218 | 0.00 (0.00-0.01) | 0.219 | 90867 | 1.21 (1.09-1.35) | 0.001 | 0.01 (0.00-0.01) | 0.001 | 119923 |
| Secondary education or further | 1.12 (0.84-1.48) | 0.442 | 0.00 (-0.01-0.01) | 0.452 | 57319 | 1.19 (1.04-1.37) | 0.011 | 0.01 (0.00-0.01) | 0.011 | 222448 | 1.42 (1.30-1.57) | 0.000 | 0.02 (0.01-0.02) | 0.000 | 388920 |
| **Covariate-adjusted regressions[b]** | | | | | | | | | | | | | | | |
| Sex | | | | | | | | | | | | | | | |
| Female | 1 (ref.) | | 1 (ref.) | | 154372 | 1 (ref.) | | 1 (ref.) | | 404284 | 1 (ref.) | | 1 (ref.) | | 589887 |
| Male | 0.94 (0.74-1.20) | 0.635 | 0.00 (-0.01-0.01) | 0.632 | 29594 | 1.00 (0.86-1.16) | 0.963 | 0.00 (-0.01-0.01) | 0.963 | 79180 | 1.08 (0.97-1.21) | 0.152 | 0.00 (0.00-0.01) | 0.158 | 119717 |
| Place of residence | | | | | | | | | | | | | | | |
| Rural | 1 (ref.) | | 1 (ref.) | | 170916 | 1 (ref.) | | 1 (ref.) | | 416580 | 1 (ref.) | | 1 (ref.) | | 537218 |
| Urban | 1.48 (1.09-2.00) | 0.013 | 0.02 (0.00-0.03) | 0.026 | 13050 | 1.46 (1.23-1.73) | 0.000 | 0.02 (0.01-0.02) | 0.000 | 66884 | 1.53 (1.38-1.70) | 0.000 | 0.02 (0.02-0.03) | 0.000 | 172386 |

| | | | | | | | | | | | | | | | |
|---|---|---|---|---|---|---|---|---|---|---|---|---|---|---|---|
| Education | | | | | | | | | | | | | | | |
| No schooling | 1 (ref.) | | 1 (ref.) | | 89499 | 1 (ref.) | | 1 (ref.) | | 170149 | 1 (ref.) | | 1 (ref.) | | 200761 |
| Primary education | 1.26 (1.01-1.56) | 0.036 | 0.01 (0.00-0.01) | 0.043 | 37148 | 1.08 (0.94-1.25) | 0.275 | 0.00 (0.00-0.01) | 0.276 | 90867 | 1.17 (1.05-1.30) | 0.005 | 0.01 (0.00-0.01) | 0.006 | 119923 |
| Secondary education or further | 1.10 (0.84-1.43) | 0.499 | 0.00 (-0.01-0.01) | 0.507 | 57319 | 1.14 (1.00-1.30) | 0.045 | 0.01 (0.00-0.01) | 0.043 | 222448 | 1.27 (1.16-1.39) | 0.000 | 0.01 (0.01-0.01) | 0.000 | 388920 |
| Observations[d] | 183,966 | | | | | 483,464 | | | | | 709,604 | | | | |

Abbreviations: RR=risk ratio; AME=average marginal effect. RRs and AMEs are shown with 95 CIs which are adjusted for clustering at the levels of the primary and secondary sampling unit.

[a] Included only one of the variables shown in the table and a binary indicator for each country (country-level fixed effects).

[b] Included age, sex, education, place of residence (urban sample only) and a binary indicator for each country (country-level fixed effects).

[c] All regressions are age adjusted with age as continuous variable with restricted cubic splines with five knots (placed at the fifth, 27.5$^{th}$, 50$^{th}$, 72.5$^{th}$, and 95$^{th}$ percentiles).

[d] Number of observations in covariate-adjusted regression model.

Table S36: Poisson regressions for diabetes onto individual-level characteristics (with survey weights), urban sample: Poverty headcount ratio >$5.50/day to all poverty levels[c]

| | >$5.50/day | | | | | Total | | | | |
|---|---|---|---|---|---|---|---|---|---|---|
| **Variable** | **RR (95% CI)** | ***P* value** | **AME (95% CI)** | ***P* value** | **Obs.** | **RR (95% CI)** | ***P* value** | **AME (95% CI)** | ***P* value** | **Obs.** |
| **Countries** | **24** | | | | | **24** | | | | |
| **Covariate-unadjusted regressions[a]** | | | | | | | | | | |
| Sex | | | | | | | | | | |
| Female | 1 (ref.) | | 1 (ref.) | | 112588 | 1 (ref.) | | 1 (ref.) | | 703813 |
| Male | 1.32 (1.15-1.51) | 0.000 | 0.02 (0.02-0.04) | 0.000 | 35988 | 1.22 (1.12-1.32) | 0.000 | 0.01 (0.01-0.02) | 0.000 | 156197 |
| Place of residence | | | | | | | | | | |
| Rural | 1 (ref.) | | 1 (ref.) | | 46635 | 1 (ref.) | | 1 (ref.) | | 585144 |
| Urban | 1.29 (1.11-1.50) | 0.001 | 0.02 (0.01-0.03) | 0.001 | 101941 | 1.59 (1.46-1.73) | 0.000 | 0.03 (0.02-0.03) | 0.000 | 274866 |
| Education | | | | | | | | | | |
| No schooling | 1 (ref.) | | 1 (ref.) | | 10078 | 1 (ref.) | | 1 (ref.) | | 210839 |
| Primary education | 1.31 (1.05-1.63) | 0.015 | 0.02 (0.01-0.03) | 0.013 | 16498 | 1.31 (1.19-1.45) | 0.000 | 0.01 (0.01-0.02) | 0.000 | 136421 |
| Secondary education or further | 1.45 (1.19-1.76) | 0.000 | 0.02 (0.02-0.04) | 0.000 | 121400 | 1.63 (1.50-1.77) | 0.000 | 0.02 (0.02-0.03) | 0.000 | 510320 |
| **Covariate-adjusted regressions[b]** | | | | | | | | | | |
| Sex | | | | | | | | | | |
| Female | 1 (ref.) | | 1 (ref.) | | 112192 | 1 (ref.) | | 1 (ref.) | | 702079 |
| Male | 1.30 (1.12-1.50) | 0.000 | 0.02 (0.01-0.03) | 0.001 | 35784 | 1.16 (1.07-1.26) | 0.001 | 0.01 (0.00-0.01) | 0.001 | 155501 |

| | | | | | | | | | | |
|---|---|---|---|---|---|---|---|---|---|---|
| Place of residence | | | | | | | | | | |
| Rural | 1 (ref.) | | 1 (ref.) | | 46428 | 1 (ref.) | | 1 (ref.) | | 273934 |
| Urban | 1.27 (1.09-1.48) | 0.002 | 0.02 (0.01-0.03) | 0.002 | 101548 | 1.51 (1.39-1.65) | 0.000 | 0.02 (0.02-0.03) | 0.000 | 583646 |
| Education | | | | | | | | | | |
| No schooling | 1 (ref.) | | 1 (ref.) | | 10078 | 1 (ref.) | | 1 (ref.) | | 210839 |
| Primary education | 1.21 (0.97-1.52) | 0.089 | 0.01 (0.00-0.03) | 0.080 | 16498 | 1.24 (1.12-1.37) | 0.000 | 0.01 (0.01-0.01) | 0.000 | 136421 |
| Secondary education or further | 1.21 (0.99-1.49) | 0.065 | 0.01 (0.00-0.03) | 0.048 | 121400 | 1.37 (1.26-1.49) | 0.000 | 0.02 (0.01-0.02) | 0.000 | 510320 |
| Observations[d] | 147,976 | | | | | 857,580 | | | | |

Abbreviations: RR=risk ratio; AME=average marginal effect; NA=not applicable. RRs and AMEs are shown with 95 CIs which are adjusted for clustering at the levels of the primary and secondary sampling unit.

[a] Included only one of the variables shown in the table and a binary indicator for each country (country-level fixed effects).

[b] Included age, sex, education, place of residence (urban sample only) and a binary indicator for each country (country-level fixed effects).

[c] All regressions are age adjusted with age as continuous variable with restricted cubic splines with five knots (placed at the fifth, 27.5$^{th}$, 50$^{th}$, 72.5$^{th}$, and 95$^{th}$ percentiles).

[d] Number of observations in covariate-adjusted regression model.

Table S37: Poisson regressions for diabetes onto individual-level characteristics (with survey weights), non-urban sample: Poverty headcount ratio <$1.90/day to poverty headcount ratio <$5.50/day [c]

| | **<$1.90/day** | | | | | **<$3.20/day** | | | | | **<$5.50/day** | | | | |
|---|---|---|---|---|---|---|---|---|---|---|---|---|---|---|---|
| **Variable** | **RR (95% CI)** | ***P* value** | **AME (95% CI)** | ***P* value** | **Obs.** | **RR (95% CI)** | ***P* value** | **AME (95% CI)** | ***P* value** | **Obs.** | **RR (95% CI)** | ***P* value** | **AME (95% CI)** | ***P* value** | **Obs.** |
| **Countries** | **42** | | | | | **42** | | | | | **42** | | | | |
| **Covariate-unadjusted regressions[a]** | | | | | | | | | | | | | | | |
| Sex | | | | | | | | | | | | | | | |
| Female | 1 (ref.) | | 1 (ref.) | | 160090 | 1 (ref.) | | 1 (ref.) | | 415489 | 1 (ref.) | | 1 (ref.) | | 606995 |
| Male | 0.95 (0.78-1.16) | 0.630 | 0.00 (-0.01-0.01) | 0.628 | 33464 | 1.01 (0.89-1.16) | 0.851 | 0.00 (-0.01-0.01) | 0.851 | 86925 | 1.09 (0.98-1.20) | 0.109 | 0.00 (0.00-0.01) | 0.113 | 131807 |
| Education | | | | | | | | | | | | | | | |
| No schooling | 1 (ref.) | | 1 (ref.) | | 92487 | 1 (ref.) | | 1 (ref.) | | 174863 | 1 (ref.) | | 1 (ref.) | | 206707 |
| Primary education | 1.21 (1.01-1.47) | 0.044 | 0.01 (0.00-0.01) | 0.049 | 41389 | 1.09 (0.96-1.24) | 0.202 | 0.00 (0.00-0.01) | 0.203 | 99645 | 1.21 (1.10-1.34) | 0.000 | 0.01 (0.00-0.01) | 0.000 | 133597 |
| Secondary education or further | 1.04 (0.80-1.33) | 0.787 | 0.00 (-0.01-0.01) | 0.789 | 59239 | 1.15 (1.01-1.31) | 0.031 | 0.01 (0.00-0.01) | 0.031 | 226608 | 1.39 (1.27-1.52) | 0.000 | 0.02 (0.01-0.02) | 0.000 | 396549 |
| **Covariate-adjusted regressions[b]** | | | | | | | | | | | | | | | |
| Sex | | | | | | | | | | | | | | | |
| Female | 1 (ref.) | | 1 (ref.) | | 159776 | 1 (ref.) | | 1 (ref.) | | 414550 | 1 (ref.) | | 1 (ref.) | | 605581 |
| Male | 0.93 (0.77-1.13) | 0.465 | 0.00 (-0.01-0.01) | 0.461 | 33339 | 0.98 (0.86-1.12) | 0.779 | 0.00 (-0.01-0.00) | 0.778 | 86566 | 1.03 (0.93-1.14) | 0.554 | 0.00 (0.00-0.01) | 0.556 | 131272 |
| Education | | | | | | | | | | | | | | | |
| No schooling | 1 (ref.) | | 1 (ref.) | | 92487 | 1 (ref.) | | 1 (ref.) | | 174863 | 1 (ref.) | | 1 (ref.) | | 206707 |
| Primary education | 1.23 (1.03-1.48) | 0.026 | 0.01 (0.00-0.01) | 0.031 | 41389 | 1.09 (0.96-1.24) | 0.190 | 0.00 (0.00-0.01) | 0.191 | 99645 | 1.21 (1.09-1.33) | 0.000 | 0.01 (0.00-0.01) | 0.000 | 133597 |
| Secondary education or further | 1.06 (0.83-1.35) | 0.653 | 0.00 (-0.01-0.01) | 0.657 | 59239 | 1.16 (1.02-1.32) | 0.024 | 0.01 (0.00-0.01) | 0.025 | 226608 | 1.38 (1.26-1.50) | 0.000 | 0.01 (0.01-0.02) | 0.000 | 396549 |
| Observations[d] | 193,115 | | | | | 501,116 | | | | | 736,853 | | | | |

Abbreviations: RR=risk ratio; AME=average marginal effect; NA=not applicable. RRs and AMEs are shown with 95 CIs which are adjusted for clustering at the levels of the primary and secondary sampling unit.

[a] Included only one of the variables shown in the table and a binary indicator for each country (country-level fixed effects).

[b] Included age, sex, education, place of residence (urban sample only) and a binary indicator for each country (country-level fixed effects).

[c] All regressions are age adjusted with age as continuous variable with restricted cubic splines with five knots (placed at the fifth, 27.5$^{th}$, 50$^{th}$, 72.5$^{th}$, and 95$^{th}$ percentiles).

[d] Number of observations in covariate-adjusted regression model.

Table S38: Poisson regressions for diabetes onto individual-level characteristics (with survey weights), non-urban sample: Poverty headcount ratio >$5.50/day to all poverty levels[c]

| | >$5.50/day | | | | | Total | | | | |
|---|---|---|---|---|---|---|---|---|---|---|
| **Variable** | **RR (95% CI)** | ***P* value** | **AME (95% CI)** | ***P* value** | **Obs.** | **RR (95% CI)** | ***P* value** | **AME (95% CI)** | ***P* value** | **Obs.** |
| **Countries** | **42** | | | | | **42** | | | | |
| **Covariate-unadjusted regressions[a]** | | | | | | | | | | |
| Sex | | | | | | | | | | |
| Female | 1 (ref.) | | 1 (ref.) | | 120000 | 1 (ref.) | | 1 (ref.) | | 726995 |
| Male | 1.22 (1.06-1.39) | 0.004 | 0.02 (0.01-0.03) | 0.005 | 42184 | 1.15 (1.06-1.25) | 0.001 | 0.01 (0.00-0.01) | 0.001 | 173991 |
| Education | | | | | | | | | | |
| No schooling | 1 (ref.) | | 1 (ref.) | | 11062 | 1 (ref.) | | 1 (ref.) | | 217769 |
| Primary education | 1.25 (1.01-1.56) | 0.042 | 0.01 (0.00-0.03) | 0.036 | 21577 | 1.29 (1.18-1.41) | 0.000 | 0.01 (0.01-0.02) | 0.000 | 155174 |
| Secondary education or further | 1.42 (1.18-1.71) | 0.000 | 0.02 (0.02-0.04) | 0.000 | 128919 | 1.58 (1.46-1.72) | 0.000 | 0.02 (0.02-0.03) | 0.000 | 525468 |
| **Covariate-adjusted regressions[b]** | | | | | | | | | | |
| Sex | | | | | | | | | | |
| Female | 1 (ref.) | | 1 (ref.) | | 119586 | 1 (ref.) | | 1 (ref.) | | 725167 |
| Male | 1.18 (1.03-1.35) | 0.014 | 0.01 (0.01-0.03) | 0.016 | 41972 | 1.08 (1.00-1.17) | 0.040 | 0.00 (0.00-0.01) | 0.042 | 173244 |
| Education | | | | | | | | | | |
| No schooling | 1 (ref.) | | 1 (ref.) | | 11062 | 1 (ref.) | | 1 (ref.) | | 217769 |
| Primary education | 1.21 (0.98-1.51) | 0.080 | 0.01 (0.00-0.03) | 0.071 | 21577 | 1.28 (1.17-1.40) | 0.000 | 0.01 (0.01-0.02) | 0.000 | 155174 |

| | | | | | | | | | | |
|---|---|---|---|---|---|---|---|---|---|---|
| Secondary education or further | 1.35 (1.13-1.62) | 0.001 | 0.02 (0.01-0.04) | 0.000 | 128919 | 1.55 (1.43-1.68) | 0.000 | 0.02 (0.02-0.03) | 0.000 | 525468 |
| Observations[d] | 161,558 | | | | | 898,411 | | | | |

Abbreviations: RR=risk ratio; AME=average marginal effect; NA=not applicable. RRs and AMEs are shown with 95 CIs which are adjusted for clustering at the levels of the primary and secondary sampling unit.

[a] Included only one of the variables shown in the table and a binary indicator for each country (country-level fixed effects).

[b] Included age, sex, education, place of residence (urban sample only) and a binary indicator for each country (country-level fixed effects).

[c] All regressions are age adjusted with age as continuous variable with restricted cubic splines with five knots (placed at the fifth, 27.5$^{th}$, 50$^{th}$, 72.5$^{th}$, and 95$^{th}$ percentiles).

[d] Number of observations in covariate-adjusted regression model.

Table S39: Poisson regressions for overweight onto individual-level characteristics (with survey weights), urban sample: Poverty headcount ratio <$1.90/day to poverty headcount ratio <$5.50/day [c]

| | **<$1.90/day** | | | | | **<$3.20/day** | | | | | **<$5.50/day** | | | | |
|---|---|---|---|---|---|---|---|---|---|---|---|---|---|---|---|
| **Variable** | **RR (95% CI)** | ***P* value** | **AME (95% CI)** | ***P* value** | **Obs.** | **RR (95% CI)** | ***P* value** | **AME (95% CI)** | ***P* value** | **Obs.** | **RR (95% CI)** | ***P* value** | **AME (95% CI)** | ***P* value** | **Obs.** |
| **Countries** | **58** | | | | | **58** | | | | | **58** | | | | |
| **Covariate-unadjusted regressions[a]** | | | | | | | | | | | | | | | |
| Sex | | | | | | | | | | | | | | | |
| Female | 1 (ref.) | | 1 (ref.) | | 255036 | 1 (ref.) | | 1 (ref.) | | 598173 | 1 (ref.) | | 1 (ref.) | | 889846 |
| Male | 0.84 (0.66-1.05) | 0.127 | -0.02 (-0.04-0.01) | 0.100 | 5484 | 0.84 (0.73-0.96) | 0.012 | -0.03 (-0.05-0.00) | 0.007 | 14620 | 0.85 (0.77-0.95) | 0.003 | -0.04 (-0.06--0.01) | 0.001 | 28033 |
| Place of residence | | | | | | | | | | | | | | | |
| Rural | 1 (ref.) | | 1 (ref.) | | 241069 | 1 (ref.) | | 1 (ref.) | | 524415 | 1 (ref.) | | 1 (ref.) | | 688496 |
| Urban | 1.22 (1.05-1.41) | 0.009 | 0.03 (0.00-0.05) | 0.015 | 19451 | 1.41 (1.33-1.50) | 0.000 | 0.07 (0.05-0.08) | 0.000 | 88378 | 1.45 (1.40-1.50) | 0.000 | 0.10 (0.08-0.10) | 0.000 | 229383 |
| Education | | | | | | | | | | | | | | | |
| No schooling | 1 (ref.) | | 1 (ref.) | | 119425 | 1 (ref.) | | 1 (ref.) | | 216072 | 1 (ref.) | | 1 (ref.) | | 258024 |
| Primary education | 1.19 (1.09-1.30) | 0.000 | 0.02 (0.01-0.03) | 0.000 | 74745 | 1.29 (1.23-1.36) | 0.000 | 0.04 (0.03-0.05) | 0.000 | 154373 | 1.35 (1.30-1.40) | 0.000 | 0.07 (0.05-0.07) | 0.000 | 214997 |
| Secondary education or further | 1.28 (1.15-1.42) | 0.000 | 0.03 (0.01-0.04) | 0.000 | 62065 | 1.44 (1.37-1.52) | 0.000 | 0.07 (0.05-0.07) | 0.000 | 229929 | 1.58 (1.52-1.63) | 0.000 | 0.11 (0.09-0.11) | 0.000 | 424807 |
| **Covariate-adjusted regressions[b]** | | | | | | | | | | | | | | | |
| Sex | | | | | | | | | | | | | | | |
| Female | 1 (ref.) | | 1 (ref.) | | 250771 | 1 (ref.) | | 1 (ref.) | | 585835 | 1 (ref.) | | 1 (ref.) | | 869971 |
| Male | 0.81 (0.64-1.02) | 0.075 | -0.03 (-0.05-0.00) | 0.051 | 5464 | 0.81 (0.70-0.93) | 0.003 | -0.04 (-0.05--0.01) | 0.001 | 14539 | 0.82 (0.74-0.91) | 0.000 | -0.05 (-0.06--0.02) | 0.000 | 27857 |
| Place of residence | | | | | | | | | | | | | | | |
| Rural | 1 (ref.) | | 1 (ref.) | | 236899 | 1 (ref.) | | 1 (ref.) | | 512707 | 1 (ref.) | | 1 (ref.) | | 672041 |
| Urban | 1.19 (1.03-1.39) | 0.022 | 0.02 (0.00-0.04) | 0.032 | 19336 | 1.37 (1.29-1.45) | 0.000 | 0.06 (0.04-0.07) | 0.000 | 87667 | 1.37 (1.32-1.42) | 0.000 | 0.08 (0.06-0.09) | 0.000 | 225787 |
| Education | | | | | | | | | | | | | | | |

| | | | | | | | | | | | | | | | |
|---|---|---|---|---|---|---|---|---|---|---|---|---|---|---|---|
| No schooling | 1 (ref.) | | 1 (ref.) | | 119425 | 1 (ref.) | | 1 (ref.) | | 216072 | 1 (ref.) | | 1 (ref.) | | 258024 |
| Primary education | 1.21 (1.10-1.32) | 0.000 | 0.02 (0.01-0.03) | 0.000 | 74745 | 1.28 (1.22-1.35) | 0.000 | 0.04 (0.03-0.05) | 0.000 | 154373 | 1.32 (1.27-1.37) | 0.000 | 0.06 (0.05-0.06) | 0.000 | 214997 |
| Secondary education or further | 1.28 (1.14-1.44) | 0.000 | 0.03 (0.01-0.04) | 0.000 | 62065 | 1.40 (1.34-1.47) | 0.000 | 0.06 (0.04-0.06) | 0.000 | 229929 | 1.48 (1.43-1.53) | 0.000 | 0.09 (0.08-0.09) | 0.000 | 424807 |
| Observations[d] | 256,235 | | | | | 600,374 | | | | | 897,828 | | | | |

Abbreviations: RR=risk ratio; AME=average marginal effect. RRs and AMEs are shown with 95 CIs which are adjusted for clustering at the levels of the primary and secondary sampling unit.

[a] Included only one of the variables shown in the table and a binary indicator for each country (country-level fixed effects).

[b] Included age, sex, education, place of residence (urban sample only) and a binary indicator for each country (country-level fixed effects).

[c] All regressions are age adjusted with age as continuous variable with restricted cubic splines with five knots (placed at the fifth, 27.5$^{th}$, 50$^{th}$, 72.5$^{th}$, and 95$^{th}$ percentiles).

[d] Number of observations in covariate-adjusted regression model.

Table S40: Poisson regressions for overweight onto individual-level characteristics (with survey weights), urban sample: Poverty headcount ratio >$5.50/day to all poverty levels[c]

| | **>$5.50/day** | | | | | **Total** | | | | |
|---|---|---|---|---|---|---|---|---|---|---|
| **Variable** | **RR (95% CI)** | ***P* value** | **AME (95% CI)** | ***P* value** | **Obs.** | **RR (95% CI)** | ***P* value** | **AME (95% CI)** | ***P* value** | **Obs.** |
| **Countries** | **58** | | | | | **58** | | | | |
| **Covariate-unadjusted regressions[a]** | | | | | | | | | | |
| Sex | | | | | | | | | | |
| Female | 1 (ref.) | | 1 (ref.) | | 289221 | 1 (ref.) | | 1 (ref.) | | 1179067 |
| Male | 1.05 (0.98-1.11) | 0.147 | 0.02 (0.00-0.05) | 0.152 | 43103 | 0.97 (0.90-1.03) | 0.292 | -0.01 (-0.03-0.01) | 0.287 | 71136 |
| Place of residence | | | | | | | | | | |
| Rural | 1 (ref.) | | 1 (ref.) | | 76974 | 1 (ref.) | | 1 (ref.) | | 765470 |
| Urban | 1.03 (0.98-1.09) | 0.260 | 0.01 (-0.01-0.04) | 0.257 | 255350 | 1.38 (1.33-1.43) | 0.000 | 0.10 (0.09-0.11) | 0.000 | 484733 |
| Education | | | | | | | | | | |
| No schooling | 1 (ref.) | | 1 (ref.) | | 17475 | 1 (ref.) | | 1 (ref.) | | 275499 |
| Primary education | 1.02 (0.96-1.08) | 0.544 | 0.01 (-0.01-0.03) | 0.543 | 61276 | 1.36 (1.32-1.41) | 0.000 | 0.08 (0.07-0.09) | 0.000 | 276273 |
| Secondary education or further | 1.00 (0.94-1.05) | 0.870 | 0.00 (-0.03-0.02) | 0.870 | 248045 | 1.57 (1.52-1.62) | 0.000 | 0.12 (0.12-0.13) | 0.000 | 672852 |
| **Covariate-adjusted regressions[b]** | | | | | | | | | | |
| Sex | | | | | | | | | | |
| Female | 1 (ref.) | | 1 (ref.) | | 283944 | 1 (ref.) | | 1 (ref.) | | 1153915 |
| Male | 1.05 (0.99-1.12) | 0.123 | 0.02 (0.00-0.05) | 0.127 | 42852 | 0.94 (0.88-1.01) | 0.072 | -0.02 (-0.04-0.00) | 0.067 | 70709 |

| | | | | | | | | | | |
|---|---|---|---|---|---|---|---|---|---|---|
| Place of residence | | | | | | | | | | |
| Rural | 1 (ref.) | | 1 (ref.) | | 75692 | 1 (ref.) | | 1 (ref.) | | 747733 |
| Urban | 1.04 (0.98-1.10) | 0.184 | 0.02 (0.00-0.04) | 0.180 | 251104 | 1.29 (1.25-1.33) | 0.000 | 0.08 (0.07-0.09) | 0.000 | 476891 |
| Education | | | | | | | | | | |
| No schooling | 1 (ref.) | | 1 (ref.) | | 17475 | 1 (ref.) | | 1 (ref.) | | 275499 |
| Primary education | 1.01 (0.95-1.07) | 0.699 | 0.00 (-0.02-0.03) | 0.698 | 61276 | 1.32 (1.28-1.37) | 0.000 | 0.07 (0.06-0.08) | 0.000 | 276273 |
| Secondary education or further | 0.98 (0.93-1.03) | 0.460 | -0.01 (-0.03-0.01) | 0.463 | 248045 | 1.44 (1.39-1.49) | 0.000 | 0.10 (0.09-0.11) | 0.000 | 672852 |
| Observations[d] | 326,796 | | | | | 1,224,624 | | | | |

Abbreviations: RR=risk ratio; AME=average marginal effect; NA=not applicable. RRs and AMEs are shown with 95 CIs which are adjusted for clustering at the levels of the primary and secondary sampling unit.

[a] Included only one of the variables shown in the table and a binary indicator for each country (country-level fixed effects).

[b] Included age, sex, education, place of residence (urban sample only) and a binary indicator for each country (country-level fixed effects).

[c] All regressions are age adjusted with age as continuous variable with restricted cubic splines with five knots (placed at the fifth, 27.5$^{th}$, 50$^{th}$, 72.5$^{th}$, and 95$^{th}$ percentiles).

[d] Number of observations in covariate-adjusted regression model.

Table S41: Poisson regressions for overweight onto individual-level characteristics (with survey weights), non-urban sample: Poverty headcount ratio <$1.90/day to poverty headcount ratio <$5.50/day [c]

| | **<$1.90/day** | | | | | **<$3.20/day** | | | | | **<$5.50/day** | | | | |
|---|---|---|---|---|---|---|---|---|---|---|---|---|---|---|---|
| **Variable** | **RR (95% CI)** | ***P* value** | **AME (95% CI)** | ***P* value** | **Obs.** | **RR (95% CI)** | ***P* value** | **AME (95% CI)** | ***P* value** | **Obs.** | **RR (95% CI)** | ***P* value** | **AME (95% CI)** | ***P* value** | **Obs.** |
| **Countries** | **71** | | | | | **71** | | | | | **71** | | | | |
| **Covariate-unadjusted regressions[a]** | | | | | | | | | | | | | | | |
| Sex | | | | | | | | | | | | | | | |
| Female | 1 (ref.) | | 1 (ref.) | | 257784 | 1 (ref.) | | 1 (ref.) | | 604235 | 1 (ref.) | | 1 (ref.) | | 899858 |
| Male | 0.84 (0.68-1.03) | 0.095 | -0.02 (-0.04-0.01) | 0.0730 | 7448 | 0.83 (0.74-0.95) | 0.005 | -0.03 (-0.05--0.01) | 0.003 | 19110 | 0.85 (0.78-0.94) | 0.001 | -0.04 (-0.05--0.01) | 0.000 | 35908 |
| Education | | | | | | | | | | | | | | | |
| No schooling | 1 (ref.) | | 1 (ref.) | | 120215 | 1 (ref.) | | 1 (ref.) | | 217569 | 1 (ref.) | | 1 (ref.) | | 260096 |
| Primary education | 1.19 (1.09-1.29) | 0.000 | 0.02 (0.01-0.03) | 0.0001 | 76809 | 1.28 (1.22-1.35) | 0.000 | 0.04 (0.03-0.05) | 0.000 | 159234 | 1.34 (1.29-1.39) | 0.000 | 0.06 (0.05-0.07) | 0.000 | 223021 |
| Secondary education or further | 1.27 (1.14-1.40) | 0.000 | 0.03 (0.01-0.04) | 0.0000 | 63894 | 1.44 (1.37-1.51) | 0.000 | 0.07 (0.05-0.07) | 0.000 | 234065 | 1.56 (1.51-1.62) | 0.000 | 0.11 (0.09-0.11) | 0.000 | 432517 |
| **Covariate-adjusted regressions[b]** | | | | | | | | | | | | | | | |
| Sex | | | | | | | | | | | | | | | |
| Female | 1 (ref.) | | 1 (ref.) | | 253499 | 1 (ref.) | | 1 (ref.) | | 591859 | 1 (ref.) | | 1 (ref.) | | 879937 |
| Male | 0.81 (0.65-1.00) | 0.048 | -0.03 (-0.05-0.00) | 0.0308 | 7419 | 0.80 (0.71-0.91) | 0.001 | -0.04 (-0.05--0.01) | 0.000 | 19009 | 0.81 (0.74-0.89) | 0.000 | -0.05 (-0.06--0.02) | 0.000 | 35697 |
| Education | | | | | | | | | | | | | | | |
| No schooling | 1 (ref.) | | 1 (ref.) | | 120215 | 1 (ref.) | | 1 (ref.) | | 217569 | 1 (ref.) | | 1 (ref.) | | 260096 |
| Primary education | 1.21 (1.10-1.32) | 0.000 | 0.02 (0.01-0.03) | 0.0000 | 76809 | 1.29 (1.23-1.36) | 0.000 | 0.04 (0.03-0.05) | 0.000 | 159234 | 1.34 (1.29-1.40) | 0.000 | 0.07 (0.05-0.07) | 0.000 | 223021 |
| Secondary education or further | 1.30 (1.16-1.44) | 0.000 | 0.03 (0.02-0.05) | 0.0000 | 63894 | 1.46 (1.39-1.53) | 0.000 | 0.07 (0.05-0.07) | 0.000 | 234065 | 1.58 (1.52-1.64) | 0.000 | 0.11 (0.09-0.11) | 0.000 | 432517 |

| | | | |
|---|---|---|---|
| Observations[d] | 260,918 | 610,868 | 915,634 |

Abbreviations: RR=risk ratio; AME=average marginal effect; NA=not applicable. RRs and AMEs are shown with 95 CIs which are adjusted for clustering at the levels of the primary and secondary sampling unit.

[a] Included only one of the variables shown in the table and a binary indicator for each country (country-level fixed effects).

[b] Included age, sex, education, place of residence (urban sample only) and a binary indicator for each country (country-level fixed effects).

[c] All regressions are age adjusted with age as continuous variable with restricted cubic splines with five knots (placed at the fifth, 27.5$^{th}$, 50$^{th}$, 72.5$^{th}$, and 95$^{th}$ percentiles).

[d] Number of observations in covariate-adjusted regression model.

Table S42: Poisson regressions for overweight onto individual-level characteristics (with survey weights), non-urban sample: Poverty headcount ratio >$5.50/day to all poverty levels[c]

| | **>$5.50/day** | | | | | **Total** | | | | |
|---|---|---|---|---|---|---|---|---|---|---|
| **Variable** | **RR (95% CI)** | ***P* value** | **AME (95% CI)** | ***P* value** | **Obs.** | **RR (95% CI)** | ***P* value** | **AME (95% CI)** | ***P* value** | **Obs.** |
| **Countries** | **71** | | | | | **71** | | | | |
| **Covariate-unadjusted regressions[a]** | | | | | | | | | | |
| Sex | | | | | | | | | | |
| Female | 1 (ref.) | | 1 (ref.) | | 297053 | 1 (ref.) | | 1 (ref.) | | 1196911 |
| Male | 1.04 (0.99-1.10) | 0.129 | 0.02 (0.00-0.05) | 0.132 | 51127 | 0.97 (0.91-1.02) | 0.244 | -0.01 (-0.03-0.01) | 0.239 | 87035 |
| Education | | | | | | | | | | |
| No schooling | 1 (ref.) | | 1 (ref.) | | 18117 | 1 (ref.) | | 1 (ref.) | | 278213 |
| Primary education | 1.02 (0.96-1.07) | 0.552 | 0.01 (-0.01-0.03) | 0.551 | 65657 | 1.35 (1.31-1.4) | 0.000 | 0.12 (0.12-0.13) | 0.000 | 288678 |
| Secondary education or further | 1.00 (0.95-1.05) | 0.939 | 0.00 (-0.02-0.02) | 0.939 | 258860 | 1.55 (1.50-1.60) | 0.000 | 0.12 (0.12-0.13) | 0.000 | 691377 |
| **Covariate-adjusted regressions[b]** | | | | | | | | | | |
| Sex | | | | | | | | | | |
| Female | 1 (ref.) | | 1 (ref.) | | 291761 | 1 (ref.) | | 1 (ref.) | | 1171698 |
| Male | 1.04 (0.99-1.10) | 0.119 | 0.02 (0.00-0.05) | 0.122 | 50873 | 0.94 (0.88-0.99) | 0.027 | -0.02 (-0.04-0.00) | 0.024 | 86570 |
| Education | | | | | | | | | | |
| No schooling | 1 (ref.) | | 1 (ref.) | | 18117 | 1 (ref.) | | 1 (ref.) | | 278213 |
| Primary education | 1.01 (0.96-1.07) | 0.653 | 0.01 (-0.02-0.03) | 0.652 | 65657 | 1.36 (1.31-1.40) | 0.000 | 0.08 (0.07-0.09) | 0.000 | 288678 |

| | | | | | | | | | | |
|---|---|---|---|---|---|---|---|---|---|---|
| Secondary education or further | 0.99 (0.94-1.05) | 0.768 | 0.00 (-0.03-0.02) | 0.768 | 258860 | 1.56 (1.51-1.61) | 0.000 | 0.13 (0.12-0.13) | 0.000 | 691377 |
| Observations[d] | 342,634 | | | | | 1,258,268 | | | | |

Abbreviations: RR=risk ratio; AME=average marginal effect; NA=not applicable. RRs and AMEs are shown with 95 CIs which are adjusted for clustering at the levels of the primary and secondary sampling unit.

[a] Included only one of the variables shown in the table and a binary indicator for each country (country-level fixed effects).

[b] Included age, sex, education, place of residence (urban sample only) and a binary indicator for each country (country-level fixed effects).

[c] All regressions are age adjusted with age as continuous variable with restricted cubic splines with five knots (placed at the fifth, $27.5^{th}$, $50^{th}$, $72.5^{th}$, and $95^{th}$ percentiles).

[d] Number of observations in covariate-adjusted regression model.

Table S43: Poisson regressions for obesity onto individual-level characteristics (with survey weights), urban sample: Poverty headcount ratio <$1.90/day to poverty headcount ratio <$5.50/day [c]

| Variable | <$1.90/day RR (95% CI) | *P* value | AME (95% CI) | *P* value | Obs. | <$3.20/day RR (95% CI) | *P* value | AME (95% CI) | *P* value | Obs. | <$5.50/day RR (95% CI) | *P* value | AME (95% CI) | *P* value | Obs. |
|---|---|---|---|---|---|---|---|---|---|---|---|---|---|---|---|
| **Countries** | **58** | | | | | **58** | | | | | **58** | | | | |
| **Covariate-unadjusted regressions[a]** | | | | | | | | | | | | | | | |
| Sex | | | | | | | | | | | | | | | |
| Female | 1 (ref.) | | 1 (ref.) | | 255036 | 1 (ref.) | | 1 (ref.) | | 598173 | 1 (ref.) | | 1 (ref.) | | 889846 |
| Male | 0.33 (0.19-0.58) | 0.000 | -0.02 (-0.03--0.01) | 0.000 | 5484 | 0.46 (0.34-0.63) | 0.000 | -0.03 (-0.03--0.02) | 0.000 | 14620 | 0.55 (0.44-0.69) | 0.000 | -0.03 (-0.04--0.02) | 0.000 | 28033 |
| Place of residence | | | | | | | | | | | | | | | |
| Rural | 1 (ref.) | | 1 (ref.) | | 241069 | 1 (ref.) | | 1 (ref.) | | 524415 | 1 (ref.) | | 1 (ref.) | | 688496 |
| Urban | 1.24 (0.99-1.57) | 0.063 | 0.01 (0.00-0.01) | 0.081 | 19451 | 1.64 (1.49-1.80) | 0.000 | 0.03 (0.02-0.03) | 0.000 | 88378 | 1.63 (1.54-1.73) | 0.000 | 0.04 (0.03-0.04) | 0.000 | 229383 |
| Education | | | | | | | | | | | | | | | |
| No schooling | 1 (ref.) | | 1 (ref.) | | 119425 | 1 (ref.) | | 1 (ref.) | | 216072 | 1 (ref.) | | 1 (ref.) | | 258024 |
| Primary education | 1.12 (0.94-1.33) | 0.203 | 0.00 (0.00-0.01) | 0.207 | 74745 | 1.38 (1.25-1.52) | 0.000 | 0.01 (0.01-0.02) | 0.000 | 154373 | 1.44 (1.34-1.55) | 0.000 | 0.02 (0.02-0.03) | 0.000 | 214997 |
| Secondary education or further | 1.18 (0.94-1.48) | 0.155 | 0.00 (0.00-0.01) | 0.170 | 62065 | 1.52 (1.38-1.67) | 0.000 | 0.02 (0.01-0.02) | 0.000 | 229929 | 1.65 (1.54-1.76) | 0.000 | 0.03 (0.03-0.04) | 0.000 | 424807 |
| **Covariate-adjusted regressions[b]** | | | | | | | | | | | | | | | |
| Sex | | | | | | | | | | | | | | | |
| Female | 1 (ref.) | | 1 (ref.) | | 250771 | 1 (ref.) | | 1 (ref.) | | 585835 | 1 (ref.) | | 1 (ref.) | | 869971 |
| Male | 0.32 (0.18-0.56) | 0.000 | -0.02 (-0.03--0.01) | 0.000 | 5464 | 0.45 (0.33-0.61) | 0.000 | -0.03 (-0.03--0.02) | 0.000 | 14539 | 0.53 (0.42-0.66) | 0.000 | -0.04 (-0.04--0.02) | 0.000 | 27857 |
| Place of residence | | | | | | | | | | | | | | | |
| Rural | 1 (ref.) | | 1 (ref.) | | 236899 | 1 (ref.) | | 1 (ref.) | | 512707 | 1 (ref.) | | 1 (ref.) | | 672041 |
| Urban | 1.22 (0.97-1.53) | 0.090 | 0.01 (0.00-0.01) | 0.109 | 19336 | 1.58 (1.44-1.74) | 0.000 | 0.02 (0.02-0.03) | 0.000 | 87667 | 1.54 (1.45-1.63) | 0.000 | 0.03 (0.03-0.04) | 0.000 | 225787 |
| Education | | | | | | | | | | | | | | | |

| | | | | | | | | | | | | | | | |
|---|---|---|---|---|---|---|---|---|---|---|---|---|---|---|---|
| No schooling | 1 (ref.) | | 1 (ref.) | | 119425 | 1 (ref.) | | 1 (ref.) | | 216072 | 1 (ref.) | | 1 (ref.) | | 258024 |
| Primary education | 1.18 (0.99-1.40) | 0.064 | 0.00 (0.00-0.01) | 0.070 | 74745 | 1.38 (1.25-1.52) | 0.000 | 0.01 (0.01-0.02) | 0.000 | 154373 | 1.39 (1.29-1.50) | 0.000 | 0.02 (0.02-0.03) | 0.000 | 214997 |
| Secondary education or further | 1.24 (1.00-1.56) | 0.055 | 0.01 (0.00-0.01) | 0.069 | 62065 | 1.48 (1.34-1.62) | 0.000 | 0.02 (0.01-0.02) | 0.000 | 229929 | 1.52 (1.42-1.62) | 0.000 | 0.03 (0.02-0.03) | 0.000 | 424807 |
| Observations[d] | 256,235 | | | | | 600,374 | | | | | 897,828 | | | | |

Abbreviations: RR=risk ratio; AME=average marginal effect. RRs and AMEs are shown with 95 CIs which are adjusted for clustering at the levels of the primary and secondary sampling unit.

[a] Included only one of the variables shown in the table and a binary indicator for each country (country-level fixed effects).

[b] Included age, sex, education, place of residence (urban sample only) and a binary indicator for each country (country-level fixed effects).

[c] All regressions are age adjusted with age as continuous variable with restricted cubic splines with five knots (placed at the fifth, 27.5$^{th}$, 50$^{th}$, 72.5$^{th}$, and 95$^{th}$ percentiles).

[d] Number of observations in covariate-adjusted regression model.

Table S44: Poisson regressions for obesity onto individual-level characteristics (with survey weights), urban sample: Poverty headcount ratio >$5.50/day to all poverty levels[c]

| | **>$5.50/day** | | | | | **Total** | | | | |
|---|---|---|---|---|---|---|---|---|---|---|
| **Variable** | **RR (95% CI)** | ***P* value** | **AME (95% CI)** | ***P* value** | **Obs.** | **RR (95% CI)** | ***P* value** | **AME (95% CI)** | ***P* value** | **Obs.** |
| **Countries** | **58** | | | | | **58** | | | | |
| **Covariate-unadjusted regressions[a]** | | | | | | | | | | |
| Sex | | | | | | | | | | |
| Female | 1 (ref.) | | 1 (ref.) | | 289221 | 1 (ref.) | | 1 (ref.) | | 1179067 |
| Male | 0.80 (0.72-0.88) | 0.000 | -0.03 (-0.05--0.03) | 0.000 | 43103 | 0.71 (0.63-0.80) | 0.000 | -0.03 (-0.04--0.02) | 0.000 | 71136 |
| Place of residence | | | | | | | | | | |
| Rural | 1 (ref.) | | 1 (ref.) | | 76974 | 1 (ref.) | | 1 (ref.) | | 765470 |
| Urban | 1.02 (0.94-1.11) | 0.603 | 0.00 (-0.01-0.02) | 0.601 | 255350 | 1.54 (1.47-1.62) | 0.000 | 0.04 (0.04-0.05) | 0.000 | 484733 |
| Education | | | | | | | | | | |
| No schooling | 1 (ref.) | | 1 (ref.) | | 17475 | 1 (ref.) | | 1 (ref.) | | 275499 |
| Primary education | 1.04 (0.95-1.13) | 0.427 | 0.00 (0.00-0.02) | 0.423 | 61276 | 1.48 (1.40-1.57) | 0.000 | 0.03 (0.03-0.04) | 0.000 | 276273 |
| Secondary education or further | 0.94 (0.87-1.03) | 0.191 | -0.01 (-0.02-0.00) | 0.201 | 248045 | 1.69 (1.60-1.79) | 0.000 | 0.04 (0.05-0.05) | 0.000 | 672852 |
| **Covariate-adjusted regressions[b]** | | | | | | | | | | |
| Sex | | | | | | | | | | |
| Female | 1 (ref.) | | 1 (ref.) | | 283944 | 1 (ref.) | | 1 (ref.) | | 1153915 |
| Male | 0.80 (0.72-0.89) | 0.000 | -0.03 (-0.05--0.03) | 0.000 | 42852 | 0.69 (0.62-0.78) | 0.000 | -0.03 (-0.04--0.02) | 0.000 | 70709 |

| | | | | | | | | | | |
|---|---|---|---|---|---|---|---|---|---|---|
| Place of residence | | | | | | | | | | |
| Rural | 1 (ref.) | | 1 (ref.) | | 75692 | 1 (ref.) | | 1 (ref.) | | 747733 |
| Urban | 1.04 (0.95-1.13) | 0.423 | 0.00 (0.00-0.02) | 0.418 | 251104 | 1.43 (1.36-1.50) | 0.000 | 0.03 (0.03-0.04) | 0.000 | 476891 |
| Education | | | | | | | | | | |
| No schooling | 1 (ref.) | | 1 (ref.) | | 17475 | 1 (ref.) | | 1 (ref.) | | 275499 |
| Primary education | 1.05 (0.96-1.14) | 0.304 | 0.01 (0.00-0.02) | 0.297 | 61276 | 1.42 (1.33-1.50) | 0.000 | 0.03 (0.03-0.04) | 0.000 | 276273 |
| Secondary education or further | 0.96 (0.88-1.04) | 0.318 | -0.01 (-0.02-0.00) | 0.326 | 248045 | 1.50 (1.42-1.59) | 0.000 | 0.03 (0.03-0.04) | 0.000 | 672852 |
| Observations[d] | 326,796 | | | | | 1,224,624 | | | | |

Abbreviations: RR=risk ratio; AME=average marginal effect; NA=not applicable. RRs and AMEs are shown with 95 CIs which are adjusted for clustering at the levels of the primary and secondary sampling unit.

[a] Included only one of the variables shown in the table and a binary indicator for each country (country-level fixed effects).

[b] Included age, sex, education, place of residence (urban sample only) and a binary indicator for each country (country-level fixed effects).

[c] All regressions are age adjusted with age as continuous variable with restricted cubic splines with five knots (placed at the fifth, 27.5$^{th}$, 50$^{th}$, 72.5$^{th}$, and 95$^{th}$ percentiles).

[d] Number of observations in covariate-adjusted regression model.

Table S45: Poisson regressions for obesity onto individual-level characteristics (with survey weights), non-urban sample: Poverty headcount ratio <$1.90/day to poverty headcount ratio <$5.50/day [c]

| | **<$1.90/day** | | | | | **<$3.20/day** | | | | | **<$5.50/day** | | | | |
|---|---|---|---|---|---|---|---|---|---|---|---|---|---|---|---|
| **Variable** | **RR (95% CI)** | ***P* value** | **AME (95% CI)** | ***P* value** | **Obs.** | **RR (95% CI)** | ***P* value** | **AME (95% CI)** | ***P* value** | **Obs.** | **RR (95% CI)** | ***P* value** | **AME (95% CI)** | ***P* value** | **Obs.** |
| **Countries** | **71** | | | | | **71** | | | | | **71** | | | | |
| **Covariate-unadjusted regressions[a]** | | | | | | | | | | | | | | | |
| Sex | | | | | | | | | | | | | | | |
| Female | 1 (ref.) | | 1 (ref.) | | 257784 | 1 (ref.) | | 1 (ref.) | | 604235 | 1 (ref.) | | 1 (ref.) | | 899858 |
| Male | 0.36 (0.24-0.54) | 0.000 | -0.02 (-0.03--0.02) | 0.000 | 7448 | 0.50 (0.38-0.64) | 0.000 | -0.03 (-0.03--0.02) | 0.000 | 19110 | 0.58 (0.48-0.72) | 0.000 | -0.03 (-0.04--0.02) | 0.000 | 35908 |
| Education | | | | | | | | | | | | | | | |
| No schooling | 1 (ref.) | | 1 (ref.) | | 120215 | 1 (ref.) | | 1 (ref.) | | 217569 | 1 (ref.) | | 1 (ref.) | | 260096 |
| Primary education | 1.13 (0.95-1.33) | 0.159 | 0.00 (0.00-0.01) | 0.162 | 76809 | 1.37 (1.25-1.51) | 0.000 | 0.01 (0.01-0.02) | 0.000 | 159234 | 1.42 (1.32-1.53) | 0.000 | 0.02 (0.02-0.03) | 0.000 | 223021 |
| Secondary education or further | 1.20 (0.97-1.49) | 0.101 | 0.01 (0.00-0.01) | 0.111 | 63894 | 1.51 (1.38-1.66) | 0.000 | 0.02 (0.01-0.02) | 0.000 | 234065 | 1.63 (1.52-1.74) | 0.000 | 0.03 (0.03-0.04) | 0.000 | 432517 |
| **Covariate-adjusted regressions[b]** | | | | | | | | | | | | | | | |
| Sex | | | | | | | | | | | | | | | |
| Female | 1 (ref.) | | 1 (ref.) | | 253499 | 1 (ref.) | | 1 (ref.) | | 591859 | 1 (ref.) | | 1 (ref.) | | 879937 |
| Male | 0.35 (0.23-0.52) | 0.000 | -0.02 (-0.03--0.02) | 0.000 | 7419 | 0.47 (0.37-0.61) | 0.000 | -0.03 (-0.03--0.02) | 0.000 | 19009 | 0.56 (0.45-0.68) | 0.000 | -0.03 (-0.04--0.02) | 0.000 | 35697 |
| Education | | | | | | | | | | | | | | | |
| No schooling | 1 (ref.) | | 1 (ref.) | | 120215 | 1 (ref.) | | 1 (ref.) | | 217569 | 1 (ref.) | | 1 (ref.) | | 260096 |
| Primary education | 1.19 (1.01-1.41) | 0.035 | 0.01 (0.00-0.01) | 0.038 | 76809 | 1.39 (1.27-1.53) | 0.000 | 0.01 (0.01-0.02) | 0.000 | 159234 | 1.43 (1.33-1.54) | 0.000 | 0.02 (0.02-0.03) | 0.000 | 223021 |
| Secondary education or further | 1.31 (1.06-1.62) | 0.011 | 0.01 (0.00-0.02) | 0.016 | 63894 | 1.57 (1.43-1.72) | 0.000 | 0.02 (0.02-0.02) | 0.000 | 234065 | 1.66 (1.55-1.77) | 0.000 | 0.04 (0.03-0.04) | 0.000 | 432517 |

| | | | |
|---|---|---|---|
| Observations[d] | 260,918 | 610,868 | 915,634 |

Abbreviations: RR=risk ratio; AME=average marginal effect; NA=not applicable. RRs and AMEs are shown with 95 CIs which are adjusted for clustering at the levels of the primary and secondary sampling unit.

[a] Included only one of the variables shown in the table and a binary indicator for each country (country-level fixed effects).

[b] Included age, sex, education, place of residence (urban sample only) and a binary indicator for each country (country-level fixed effects).

[c] All regressions are age adjusted with age as continuous variable with restricted cubic splines with five knots (placed at the fifth, 27.5$^{th}$, 50$^{th}$, 72.5$^{th}$, and 95$^{th}$ percentiles).

[d] Number of observations in covariate-adjusted regression model.

Table S46: Poisson regressions for obesity onto individual-level characteristics (with survey weights), non-urban sample: Poverty headcount ratio >$5.50/day to all poverty levels[c]

| | >$5.50/day | | | | | Total | | | | |
|---|---|---|---|---|---|---|---|---|---|---|
| **Variable** | **RR (95% CI)** | ***P* value** | **AME (95% CI)** | ***P* value** | **Obs.** | **RR (95% CI)** | ***P* value** | **AME (95% CI)** | ***P* value** | **Obs.** |
| **Countries** | **71** | | | | | **71** | | | | |
| **Covariate-unadjusted regressions[a]** | | | | | | | | | | |
| Sex | | | | | | | | | | |
| Female | 1 (ref.) | | 1 (ref.) | | 297053 | 1 (ref.) | | 1 (ref.) | | 1196911 |
| Male | 0.83 (0.75-0.92) | 0.000 | -0.03 (-0.05--0.02) | 0.000 | 51127 | 0.74 (0.69-0.82) | 0.000 | -0.03 (-0.04--0.02) | 0.000 | 87035 |
| Education | | | | | | | | | | |
| No schooling | 1 (ref.) | | 1 (ref.) | | 18117 | 1 (ref.) | | 1 (ref.) | | 278213 |
| Primary education | 1.03 (0.95-1.12) | 0.424 | 0.01 (0.00-0.02) | 0.419 | 65657 | 1.47 (1.38-1.55) | 0.000 | 0.03 (0.03-0.04) | 0.000 | 288678 |
| Secondary education or further | 0.95 (0.88-1.03) | 0.241 | -0.01 (-0.02-0.00) | 0.250 | 258860 | 1.66 (1.58-1.76) | 0.000 | 0.04 (0.03-0.05) | 0.000 | 691377 |
| **Covariate-adjusted regressions[b]** | | | | | | | | | | |
| Sex | | | | | | | | | | |
| Female | 1 (ref.) | | 1 (ref.) | | 291761 | 1 (ref.) | | 1 (ref.) | | 1171698 |
| Male | 0.83 (0.75-0.92) | 0.001 | -0.03 (-0.05--0.02) | 0.000 | 50873 | 0.73 (0.64-0.82) | 0.000 | -0.03 (-0.04--0.02) | 0.000 | 86570 |
| Education | | | | | | | | | | |
| No schooling | 1 (ref.) | | 1 (ref.) | | 18117 | 1 (ref.) | | 1 (ref.) | | 278213 |
| Primary education | 1.05 (0.96-1.14) | 0.298 | 0.01 (0.00-0.02) | 0.291 | 65657 | 1.47 (1.39-1.56) | 0.000 | 0.03 (0.03-0.04) | 0.000 | 288678 |

| | | | | | | | | | | |
|---|---|---|---|---|---|---|---|---|---|---|
| Secondary education or further | 0.97 (0.89-1.05) | 0.406 | 0.00 (-0.02-0.00) | 0.413 | 258860 | 1.68 (1.59-1.78) | 0.000 | 0.04 (0.05-0.06) | 0.000 | 691377 |
| Observations[d] | 342,634 | | | | | 1,258,268 | | | | |

Abbreviations: RR=risk ratio; AME=average marginal effect; NA=not applicable. RRs and AMEs are shown with 95 CIs which are adjusted for clustering at the levels of the primary and secondary sampling unit.

[a] Included only one of the variables shown in the table and a binary indicator for each country (country-level fixed effects).

[b] Included age, sex, education, place of residence (urban sample only) and a binary indicator for each country (country-level fixed effects).

[c] All regressions are age adjusted with age as continuous variable with restricted cubic splines with five knots (placed at the fifth, 27.5th, 50th, 72.5th, and 95th percentiles).

[d] Number of observations in covariate-adjusted regression model.

Table S47: Poisson regressions for smoking onto individual-level characteristics (with survey weights), urban sample: Poverty headcount ratio <$1.90/day to poverty headcount ratio <$5.50/day [c]

| | **<$1.90/day** | | | | | **<$3.20/day** | | | | | **<$5.50/day** | | | | |
|---|---|---|---|---|---|---|---|---|---|---|---|---|---|---|---|
| **Variable** | **RR (95% CI)** | ***P* value** | **AME (95% CI)** | ***P* value** | **Obs.** | **RR (95% CI)** | ***P* value** | **AME (95% CI)** | ***P* value** | **Obs.** | **RR (95% CI)** | ***P* value** | **AME (95% CI)** | ***P* value** | **Obs.** |
| **Countries** | **49** | | | | | **49** | | | | | **49** | | | | |
| **Covariate-unadjusted regressions[a]** | | | | | | | | | | | | | | | |
| Sex | | | | | | | | | | | | | | | |
| Female | 1 (ref.) | | 1 (ref.) | | 114943 | 1 (ref.) | | 1 (ref.) | | 204842 | 1 (ref.) | | 1 (ref.) | | 297501 |
| Male | 12.39 (9.56-16.04) | 0.000 | 0.18 (0.17-0.20) | 0.000 | 56593 | 12.57 (9.53-16.56) | 0.000 | 0.26 (0.17-0.31) | 0.000 | 114975 | 12.2 (9.29-16.02) | 0.000 | 0.30 (0.18-0.32) | 0.000 | 174733 |
| Place of residence | | | | | | | | | | | | | | | |
| Rural | 1 (ref.) | | 1 (ref.) | | 154133 | 1 (ref.) | | 1 (ref.) | | 261920 | 1 (ref.) | | 1 (ref.) | | 336952 |
| Urban | 0.92 (0.81-1.05) | 0.219 | -0.01 (-0.02-0.01) | 0.213 | 17403 | 0.99 (0.89-1.09) | 0.766 | 0.00 (-0.02-0.02) | 0.765 | 57897 | 0.98 (0.93-1.04) | 0.560 | 0.00 (-0.01-0.01) | 0.560 | 135282 |
| Education | | | | | | | | | | | | | | | |
| No schooling | 1 (ref.) | | 1 (ref.) | | 56868 | 1 (ref.) | | 1 (ref.) | | 87983 | 1 (ref.) | | 1 (ref.) | | 108320 |
| Primary education | 1.41 (1.25-1.60) | 0.000 | 0.03 (0.02-0.04) | 0.000 | 71222 | 1.38 (1.24-1.54) | 0.000 | 0.05 (0.02-0.05) | 0.000 | 117227 | 1.33 (1.20-1.48) | 0.000 | 0.05 (0.01-0.05) | 0.000 | 156332 |
| Secondary education or further | 1.46 (1.22-1.74) | 0.000 | 0.04 (0.02-0.05) | 0.000 | 31636 | 1.52 (1.29-1.79) | 0.000 | 0.06 (0.02-0.06) | 0.000 | 80751 | 1.43 (1.23-1.66) | 0.000 | 0.07 (0.01-0.07) | 0.000 | 152898 |
| **Covariate-adjusted regressions[b]** | | | | | | | | | | | | | | | |
| Sex | | | | | | | | | | | | | | | |
| Female | 1 (ref.) | | 1 (ref.) | | 109188 | 1 (ref.) | | 1 (ref.) | | 188425 | 1 (ref.) | | 1 (ref.) | | 270903 |
| Male | 13.74 (10.44-18.09) | 0.000 | 0.19 (0.17-0.20) | 0.000 | 50538 | 13.61 (10.17-18.21) | 0.000 | 0.26 (0.17-0.30) | 0.000 | 97536 | 12.95 (9.75-17.2) | 0.000 | 0.31 (0.17-0.31) | 0.000 | 146647 |
| Place of residence | | | | | | | | | | | | | | | |
| Rural | 1 (ref.) | | 1 (ref.) | | 142583 | 1 (ref.) | | 1 (ref.) | | 229596 | 1 (ref.) | | 1 (ref.) | | 291645 |
| Urban | 0.96 (0.85-1.09) | 0.533 | 0.00 (-0.02-0.01) | 0.530 | 17143 | 0.99 (0.91-1.07) | 0.733 | 0.00 (-0.02-0.01) | 0.733 | 56365 | 0.99 (0.94-1.04) | 0.662 | 0.00 (-0.01-0.01) | 0.662 | 125905 |

| | | | | | | | | | | | | | | | |
|---|---|---|---|---|---|---|---|---|---|---|---|---|---|---|---|
| Education | | | | | | | | | | | | | | | |
| No schooling | 1 (ref.) | | 1 (ref.) | | 56868 | 1 (ref.) | | 1 (ref.) | | 87983 | 1 (ref.) | | 1 (ref.) | | 108320 |
| Primary education | 0.94 (0.84-1.05) | 0.262 | -0.01 (-0.02-0.01) | 0.269 | 71222 | 0.87 (0.81-0.94) | 0.001 | -0.03 (-0.03-0.00) | 0.001 | 117227 | 0.84 (0.79-0.90) | 0.000 | -0.04 (-0.04-0.00) | 0.000 | 156332 |
| Secondary education or further | 0.78 (0.69-0.89) | 0.000 | -0.03 (-0.04--0.01) | 0.000 | 31636 | 0.84 (0.76-0.93) | 0.001 | -0.03 (-0.04-0.00) | 0.001 | 80751 | 0.81 (0.74-0.87) | 0.000 | -0.05 (-0.05--0.01) | 0.000 | 152898 |
| Observations[d] | 159,726 | | | | | 285,961 | | | | | 417,550 | | | | |

Abbreviations: RR=risk ratio; AME=average marginal effect. RRs and AMEs are shown with 95 CIs which are adjusted for clustering at the levels of the primary and secondary sampling unit.

[a] Included only one of the variables shown in the table and a binary indicator for each country (country-level fixed effects).

[b] Included age, sex, education, place of residence (urban sample only) and a binary indicator for each country (country-level fixed effects).

[c] All regressions are age adjusted with age as continuous variable with restricted cubic splines with five knots (placed at the fifth, 27.5$^{th}$, 50$^{th}$, 72.5$^{th}$, and 95$^{th}$ percentiles).

[d] Number of observations in covariate-adjusted regression model.

Table S48: Poisson regressions for smoking onto individual-level characteristics (with survey weights), urban sample: Poverty headcount ratio >$5.50/day to all poverty levels[c]

| | **>$5.50/day** | | | | | **Total** | | | | |
|---|---|---|---|---|---|---|---|---|---|---|
| **Variable** | **RR (95% CI)** | ***P* value** | **AME (95% CI)** | ***P* value** | **Obs.** | **RR (95% CI)** | ***P* value** | **AME (95% CI)** | ***P* value** | **Obs.** |
| **Countries** | **45** | | | | | **49** | | | | |
| **Covariate-unadjusted regressions[a]** | | | | | | | | | | |
| Sex | | | | | | | | | | |
| Female | 1 (ref.) | | 1 (ref.) | | 164102 | 1 (ref.) | | 1 (ref.) | | 465883 |
| Male | 9.32 (7.94-10.95) | 0.000 | 0.35 (0.25-0.39) | 0.000 | 119509 | 10.56 (8.85-12.6) | 0.000 | 0.32 (0.21-0.34) | 0.000 | 294668 |
| Place of residence | | | | | | | | | | |
| Rural | 1 (ref.) | | 1 (ref.) | | 64954 | 1 (ref.) | | 1 (ref.) | | 402958 |
| Urban | 0.88 (0.80-0.97) | 0.007 | -0.03 (-0.04-0.00) | 0.008 | 218657 | 0.92 (0.86-0.97) | 0.004 | -0.02 (-0.02-0.00) | 0.004 | 357593 |
| Education | | | | | | | | | | |
| No schooling | 1 (ref.) | | 1 (ref.) | | 17798 | 1 (ref.) | | 1 (ref.) | | 126412 |
| Primary education | 1.50 (1.30-1.72) | 0.000 | 0.08 (0.03-0.08) | 0.000 | 58510 | 1.34 (1.22-1.48) | 0.000 | 0.06 (0.02-0.07) | 0.000 | 215617 |
| Secondary education or further | 1.45 (1.23-1.72) | 0.000 | 0.07 (0.02-0.08) | 0.000 | 192995 | 1.34 (1.19-1.52) | 0.000 | 0.06 (0.01-0.07) | 0.000 | 349518 |
| **Covariate-adjusted regressions[b]** | | | | | | | | | | |
| Sex | | | | | | | | | | |
| Female | 1 (ref.) | | 1 (ref.) | | 156901 | 1 (ref.) | | 1 (ref.) | | 432072 |
| Male | 9.48 (8.06-11.16) | 0.000 | 0.35 (0.25-0.39) | 0.000 | 112402 | 10.94 (9.13-13.11) | 0.000 | 0.32 (0.21-0.34) | 0.000 | 259475 |

| | | | | | | | | | | |
|---|---|---|---|---|---|---|---|---|---|---|
| Place of residence | | | | | | | | | | |
| Rural | 1 (ref.) | | 1 (ref.) | | 61499 | 1 (ref.) | | 1 (ref.) | | 354193 |
| Urban | 0.93 (0.86-1.00) | 0.053 | -0.02 (-0.03-0.01) | 0.057 | 207804 | 0.94 (0.90-0.99) | 0.020 | -0.01 (-0.02-0.00) | 0.021 | 337354 |
| Education | | | | | | | | | | |
| No schooling | 1 (ref.) | | 1 (ref.) | | 17798 | 1 (ref.) | | 1 (ref.) | | 126412 |
| Primary education | 0.98 (0.86-1.11) | 0.713 | -0.01 (-0.04-0.03) | 0.715 | 58510 | 0.88 (0.83-0.93) | 0.000 | -0.03 (-0.03-0.00) | 0.000 | 215617 |
| Secondary education or further | 0.82 (0.74-0.92) | 0.000 | -0.05 (-0.06-0.00) | 0.001 | 192995 | 0.79 (0.74-0.85) | 0.000 | -0.05 (-0.05--0.01) | 0.000 | 349518 |
| Observations[d] | 269,303 | | | | | 691,547 | | | | |

Abbreviations: RR=risk ratio; AME=average marginal effect; NA=not applicable. RRs and AMEs are shown with 95 CIs which are adjusted for clustering at the levels of the primary and secondary sampling unit.

[a] Included only one of the variables shown in the table and a binary indicator for each country (country-level fixed effects).

[b] Included age, sex, education, place of residence (urban sample only) and a binary indicator for each country (country-level fixed effects).

[c] All regressions are age adjusted with age as continuous variable with restricted cubic splines with five knots (placed at the fifth, 27.5$^{th}$, 50$^{th}$, 72.5$^{th}$, and 95$^{th}$ percentiles).

[d] Number of observations in covariate-adjusted regression model.

Table S49: Poisson regressions for smoking onto individual-level characteristics (with survey weights), non-urban sample: Poverty headcount ratio <$1.90/day to poverty headcount ratio <$5.50/day [c]

| | <$1.90/day | | | | | <$3.20/day | | | | | <$5.50/day | | | | |
|---|---|---|---|---|---|---|---|---|---|---|---|---|---|---|---|
| **Variable** | **RR (95% CI)** | ***P* value** | **AME (95% CI)** | ***P* value** | **Obs.** | **RR (95% CI)** | ***P* value** | **AME (95% CI)** | ***P* value** | **Obs.** | **RR (95% CI)** | ***P* value** | **AME (95% CI)** | ***P* value** | **Obs.** |
| **Countries** | **61** | | | | | **61** | | | | | **61** | | | | |
| **Covariate-unadjusted regressions**[a] | | | | | | | | | | | | | | | |
| Sex | | | | | | | | | | | | | | | |
| Female | 1 (ref.) | | 1 (ref.) | | 119051 | 1 (ref.) | | 1 (ref.) | | 216646 | 1 (ref.) | | 1 (ref.) | | 319455 |
| Male | 13.16 (10.37-16.70) | 0.000 | 0.19 (0.18-0.21) | 0.000 | 59628 | 14.33 (11.12-18.47) | 0.000 | 0.27 (0.19-0.32) | 0.000 | 124278 | 13.86 (10.78-17.83) | 0.000 | 0.31 (0.20-0.33) | 0.000 | 192821 |
| Education | | | | | | | | | | | | | | | |
| No schooling | 1 (ref.) | | 1 (ref.) | | 57752 | 1 (ref.) | | 1 (ref.) | | 90370 | 1 (ref.) | | 1 (ref.) | | 112623 |
| Primary education | 1.41 (1.25-1.60) | 0.000 | 0.03 (0.02-0.04) | 0.000 | 73405 | 1.39 (1.25-1.55) | 0.000 | 0.05 (0.02-0.05) | 0.000 | 122608 | 1.35 (1.22-1.49) | 0.000 | 0.05 (0.01-0.05) | 0.000 | 165820 |
| Secondary education or further | 1.45 (1.22-1.74) | 0.000 | 0.04 (0.02-0.05) | 0.000 | 33424 | 1.52 (1.30-1.79) | 0.000 | 0.07 (0.02-0.07) | 0.000 | 85874 | 1.44 (1.25-1.66) | 0.000 | 0.07 (0.01-0.07) | 0.000 | 165682 |
| **Covariate-adjusted regressions**[b] | | | | | | | | | | | | | | | |
| Sex | | | | | | | | | | | | | | | |
| Female | 1 (ref.) | | 1 (ref.) | | 111960 | 1 (ref.) | | 1 (ref.) | | 195656 | 1 (ref.) | | 1 (ref.) | | 285594 |
| Male | 13.59 (10.37-17.80) | 0.000 | 0.19 (0.17-0.21) | 0.000 | 52621 | 13.73 (10.31-18.28) | 0.000 | 0.27 (0.17-0.31) | 0.000 | 103196 | 13.41 (10.16-17.69) | 0.000 | 0.31 (0.19-0.32) | 0.000 | 158531 |
| Education | | | | | | | | | | | | | | | |
| No schooling | 1 (ref.) | | 1 (ref.) | | 57752 | 1 (ref.) | | 1 (ref.) | | 90370 | 1 (ref.) | | 1 (ref.) | | 112623 |
| Primary education | 0.94 (0.84-1.04) | 0.228 | -0.01 (-0.02-0.01) | 0.235 | 73405 | 0.87 (0.81-0.94) | 0.000 | -0.03 (-0.03-0.00) | 0.001 | 122608 | 0.84 (0.79-0.90) | 0.000 | -0.04 (-0.04-0.00) | 0.000 | 165820 |
| Secondary education or further | 0.78 (0.69-0.88) | 0.000 | -0.03 (-0.04--0.01) | 0.000 | 33424 | 0.84 (0.76-0.92) | 0.000 | -0.03 (-0.04-0.00) | 0.001 | 85874 | 0.80 (0.74-0.86) | 0.000 | -0.05 (-0.05--0.01) | 0.000 | 165682 |
| Observations[d] | 164,581 | | | | | 298,852 | | | | | 444,125 | | | | |

Abbreviations: RR=risk ratio; AME=average marginal effect; NA=not applicable. RRs and AMEs are shown with 95 CIs which are adjusted for clustering at the levels of the primary and secondary sampling unit.

[a] Included only one of the variables shown in the table and a binary indicator for each country (country-level fixed effects).

[b] Included age, sex, education, place of residence (urban sample only) and a binary indicator for each country (country-level fixed effects).

[c] All regressions are age adjusted with age as continuous variable with restricted cubic splines with five knots (placed at the fifth, 27.5$^{th}$, 50$^{th}$, 72.5$^{th}$, and 95$^{th}$ percentiles).

[d] Number of observations in covariate-adjusted regression model.

Table S50: Poisson regressions for smoking onto individual-level characteristics (with survey weights), non-urban sample: Poverty headcount ratio >$5.50/day to all poverty levels[c]

| | >$5.50/day | | | | | Total | | | | |
|---|---|---|---|---|---|---|---|---|---|---|
| **Variable** | **RR (95% CI)** | ***P* value** | **AME (95% CI)** | ***P* value** | **Obs.** | **RR (95% CI)** | ***P* value** | **AME (95% CI)** | ***P* value** | **Obs.** |
| **Countries** | **57** | | | | | **61** | | | | |
| **Covariate-unadjusted regressions**[a] | | | | | | | | | | |
| Sex | | | | | | | | | | |
| Female | 1 (ref.) | | 1 (ref.) | | 176446 | 1 (ref.) | | 1 (ref.) | | 500181 |
| Male | 9.52 (8.15-11.13) | 0.000 | 0.35 (0.25-0.39) | 0.000 | 129466 | 11.39 (9.66-13.44) | 0.000 | 0.32 (0.22-0.35) | 0.000 | 322713 |
| Education | | | | | | | | | | |
| No schooling | 1 (ref.) | | 1 (ref.) | | 19098 | 1 (ref.) | | 1 (ref.) | | 132015 |
| Primary education | 1.49 (1.30-1.70) | 0.000 | 0.08 (0.03-0.08) | 0.000 | 63116 | 1.36 (1.24-1.49) | 0.000 | 0.06 (0.02-0.07) | 0.000 | 229711 |
| Secondary education or further | 1.44 (1.23-1.69) | 0.000 | 0.07 (0.02-0.08) | 0.000 | 205360 | 1.36 (1.21-1.52) | 0.000 | 0.06 (0.02-0.07) | 0.000 | 374667 |
| **Covariate-adjusted regressions**[b] | | | | | | | | | | |
| Sex | | | | | | | | | | |
| Female | 1 (ref.) | | 1 (ref.) | | 167033 | 1 (ref.) | | 1 (ref.) | | 456895 |
| Male | 9.58 (8.16-11.24) | 0.000 | 0.35 (0.26-0.39) | 0.000 | 120541 | 11.19 (9.38-13.34) | 0.000 | 0.32 (0.22-0.34) | 0.000 | 279498 |
| Education | | | | | | | | | | |
| No schooling | 1 (ref.) | | 1 (ref.) | | 19098 | 1 (ref.) | | 1 (ref.) | | 132015 |
| Primary education | 0.98 (0.87-1.10) | 0.689 | -0.01 (-0.04-0.03) | 0.691 | 63116 | 0.88 (0.83-0.93) | 0.000 | -0.03 (-0.03-0.00) | 0.000 | 229711 |

| | | | | | | | | | | |
|---|---|---|---|---|---|---|---|---|---|---|
| Secondary education or further | 0.81 (0.73-0.89) | 0.000 | -0.05 (-0.06--0.01) | 0.000 | 205360 | 0.78 (0.73-0.83) | 0.000 | -0.06 (-0.06--0.02) | 0.000 | 374667 |
| Observations[d] | 287,574 | | | | | 736,393 | | | | |

Abbreviations: RR=risk ratio; AME=average marginal effect; NA=not applicable. RRs and AMEs are shown with 95 CIs which are adjusted for clustering at the levels of the primary and secondary sampling unit.

[a] Included only one of the variables shown in the table and a binary indicator for each country (country-level fixed effects).

[b] Included age, sex, education, place of residence (urban sample only) and a binary indicator for each country (country-level fixed effects).

[c] All regressions are age adjusted with age as continuous variable with restricted cubic splines with five knots (placed at the fifth, 27.5$^{th}$, 50$^{th}$, 72.5$^{th}$, and 95$^{th}$ percentiles).

[d] Number of observations in covariate-adjusted regression model.

Table S51: Poisson regressions for dyslipidemia onto individual-level characteristics (with survey weights), urban sample: Poverty headcount ratio <$1.90/day to poverty headcount ratio <$5.50/day [c]

| | **<$1.90/day** | | | | | **<$3.20/day** | | | | | **<$5.50/day** | | | | |
|---|---|---|---|---|---|---|---|---|---|---|---|---|---|---|---|
| **Variable** | **RR (95% CI)** | ***P* value** | **AME (95% CI)** | ***P* value** | **Obs.** | **RR (95% CI)** | ***P* value** | **AME (95% CI)** | ***P* value** | **Obs.** | **RR (95% CI)** | ***P* value** | **AME (95% CI)** | ***P* value** | **Obs.** |
| **Countries** | **14** | | | | | **16** | | | | | **16** | | | | |
| **Covariate-unadjusted regressions[a]** | | | | | | | | | | | | | | | |
| Sex | | | | | | | | | | | | | | | |
| Female | 1 (ref.) | | 1 (ref.) | | 6290 | 1 (ref.) | | 1 (ref.) | | 10635 | 1 (ref.) | | 1 (ref.) | | 14935 |
| Male | 0.60 (0.26-1.40) | 0.239 | 0.00 (-0.02-0.00) | 0.207 | 4032 | 0.44 (0.22-0.84) | 0.014 | -0.01 (-0.03--0.01) | 0.021 | 7487 | 0.57 (0.29-1.11) | 0.098 | -0.01 (-0.03-0.00) | 0.102 | 10684 |
| Place of residence | | | | | | | | | | | | | | | |
| Rural | 1 (ref.) | | 1 (ref.) | | 7925 | 1 (ref.) | | 1 (ref.) | | 13099 | 1 (ref.) | | 1 (ref.) | | 17375 |
| Urban | 1.73 (0.57-5.24) | 0.334 | 0.01 (0.00-0.03) | 0.408 | 2397 | 1.73 (0.94-3.18) | 0.080 | 0.01 (0.01-0.03) | 0.124 | 5023 | 2.40 (1.37-4.22) | 0.002 | 0.02 (0.01-0.04) | 0.019 | 8244 |
| Education | | | | | | | | | | | | | | | |
| No schooling | 1 (ref.) | | 1 (ref.) | | 3813 | 1 (ref.) | | 1 (ref.) | | 6402 | 1 (ref.) | | 1 (ref.) | | 8148 |
| Primary education | 1.83 (0.77-4.36) | 0.170 | 0.01 (0.01-0.02) | 0.189 | 4743 | 1.63 (0.90-2.94) | 0.106 | 0.01 (0.01-0.02) | 0.136 | 7900 | 1.65 (0.94-2.87) | 0.079 | 0.01 (0.00-0.02) | 0.125 | 10285 |
| Secondary education or further | 0.92 (0.37-2.31) | 0.859 | 0.00 (-0.01-0.00) | 0.859 | 1743 | 1.32 (0.70-2.52) | 0.392 | 0.00 (0.00-0.01) | 0.416 | 3795 | 2.93 (1.25-6.84) | 0.013 | 0.02 (0.01-0.06) | 0.081 | 7156 |
| **Covariate-adjusted regressions[b]** | | | | | | | | | | | | | | | |
| Sex | | | | | | | | | | | | | | | |
| Female | 1 (ref.) | | 1 (ref.) | | 6277 | 1 (ref.) | | 1 (ref.) | | 10620 | 1 (ref.) | | 1 (ref.) | | 14916 |
| Male | 0.54 (0.26-1.12) | 0.098 | -0.01 (-0.02--0.01) | 0.086 | 4022 | 0.40 (0.21-0.75) | 0.005 | -0.01 (-0.03--0.02) | 0.008 | 7477 | 0.47 (0.26-0.85) | 0.013 | -0.01 (-0.03--0.01) | 0.019 | 10673 |
| Place of residence | | | | | | | | | | | | | | | |
| Rural | 1 (ref.) | | 1 (ref.) | | 7903 | 1 (ref.) | | 1 (ref.) | | 13076 | 1 (ref.) | | 1 (ref.) | | 17350 |
| Urban | 1.50 (0.57-3.97) | 0.410 | 0.00 (0.00-0.02) | 0.461 | 2396 | 1.47 (0.82-2.62) | 0.194 | 0.01 (0.00-0.02) | 0.232 | 5021 | 1.97 (1.20-3.22) | 0.007 | 0.01 (0.01-0.03) | 0.026 | 8239 |
| Education | | | | | | | | | | | | | | | |

| | | | | | | | | | | | | | | | |
|---|---|---|---|---|---|---|---|---|---|---|---|---|---|---|---|
| No schooling | 1 (ref.) | | 1 (ref.) | | 3813 | 1 (ref.) | | 1 (ref.) | | 6402 | 1 (ref.) | | 1 (ref.) | | 8148 |
| Primary education | 2.17 (0.93-5.09) | 0.074 | 0.01 (0.01-0.03) | 0.102 | 4743 | 1.96 (1.07-3.59) | 0.028 | 0.01 (0.01-0.03) | 0.047 | 7900 | 1.80 (1.03-3.16) | 0.040 | 0.01 (0.01-0.02) | 0.068 | 10285 |
| Secondary education or further | 1.09 (0.44-2.72) | 0.846 | 0.00 (0.00-0.01) | 0.846 | 1743 | 1.72 (0.84-3.55) | 0.141 | 0.01 (0.01-0.02) | 0.195 | 3795 | 2.96 (1.45-6.04) | 0.003 | 0.02 (0.02-0.05) | 0.039 | 7156 |
| Observations[d] | 10,299 | | | | | 18,097 | | | | | 25,589 | | | | |

Abbreviations: RR=risk ratio; AME=average marginal effect. RRs and AMEs are shown with 95 CIs which are adjusted for clustering at the levels of the primary and secondary sampling unit.

[a] Included only one of the variables shown in the table and a binary indicator for each country (country-level fixed effects).

[b] Included age, sex, education, place of residence (urban sample only) and a binary indicator for each country (country-level fixed effects).

[c] All regressions are age adjusted with age as continuous variable with restricted cubic splines with five knots (placed at the fifth, 27.5th, 50th, 72.5th, and 95th percentiles).

[d] Number of observations in covariate-adjusted regression model.

Table S52: Poisson regressions for dyslipidemia onto individual-level characteristics (with survey weights), urban sample: Poverty headcount ratio >$5.50/day to all poverty levels[c]

| | >$5.50/day | | | | | Total | | | | |
|---|---|---|---|---|---|---|---|---|---|---|
| **Variable** | **RR (95% CI)** | ***P* value** | **AME (95% CI)** | ***P* value** | **Obs.** | **RR (95% CI)** | ***P* value** | **AME (95% CI)** | ***P* value** | **Obs.** |
| **Countries** | **16** | | | | | **16** | | | | |
| **Covariate-unadjusted regressions[a]** | | | | | | | | | | |
| Sex | | | | | | | | | | |
| Female | 1 (ref.) | | 1 (ref.) | | 6503 | 1 (ref.) | | 1 (ref.) | | 21438 |
| Male | 1.07 (0.85-1.35) | 0.542 | 0.01 (-0.01-0.02) | 0.544 | 4840 | 0.73 (0.47-1.13) | 0.157 | -0.01 (-0.02-0.00) | 0.164 | 15524 |
| Place of residence | | | | | | | | | | |
| Rural | 1 (ref.) | | 1 (ref.) | | 3781 | 1 (ref.) | | 1 (ref.) | | 21156 |
| Urban | 1.18 (0.92-1.50) | 0.188 | 0.01 (-0.01-0.03) | 0.183 | 7562 | 1.97 (1.32-2.93) | 0.001 | 0.02 (0.01-0.04) | 0.004 | 15806 |
| Education | | | | | | | | | | |
| No schooling | 1 (ref.) | | 1 (ref.) | | 1434 | 1 (ref.) | | 1 (ref.) | | 9582 |
| Primary education | 0.76 (0.34-1.66) | 0.484 | -0.02 (-0.06-0.04) | 0.523 | 3386 | 1.51 (0.89-2.55) | 0.126 | 0.01 (0.00-0.02) | 0.171 | 13671 |
| Secondary education or further | 1.29 (0.57-2.93) | 0.539 | 0.02 (-0.04-0.07) | 0.495 | 6516 | 2.88 (1.57-5.26) | 0.001 | 0.02 (0.02-0.05) | 0.004 | 13672 |
| **Covariate-adjusted regressions[b]** | | | | | | | | | | |
| Sex | | | | | | | | | | |
| Female | 1 (ref.) | | 1 (ref.) | | 6501 | 1 (ref.) | | 1 (ref.) | | 21417 |
| Male | 1.13 (0.86-1.47) | 0.385 | 0.01 (-0.01-0.03) | 0.389 | 4835 | 0.68 (0.45-1.04) | 0.075 | -0.01 (-0.02-0) | 0.084 | 15508 |

| | | | | | | | | | | |
|---|---|---|---|---|---|---|---|---|---|---|
| Place of residence | | | | | | | | | | |
| Rural | 1 (ref.) | | 1 (ref.) | | 3780 | 1 (ref.) | | 1 (ref.) | | 21130 |
| Urban | 1.20 (0.92-1.58) | 0.183 | 0.01 (-0.01-0.03) | 0.179 | 7556 | 1.61 (1.12-2.32) | 0.011 | 0.01 (0.01-0.03) | 0.021 | 15795 |
| Education | | | | | | | | | | |
| No schooling | 1 (ref.) | | 1 (ref.) | | 1434 | 1 (ref.) | | 1 (ref.) | | 9582 |
| Primary education | 0.72 (0.32-1.58) | 0.408 | -0.02 (-0.07-0.04) | 0.460 | 3386 | 1.56 (0.91-2.68) | 0.108 | 0.01 (0-0.02) | 0.148 | 13671 |
| Secondary education or further | 1.18 (0.51-2.69) | 0.702 | 0.01 (-0.05-0.07) | 0.682 | 6516 | 2.62 (1.56-4.40) | 0.000 | 0.02 (0.02-0.05) | 0.002 | 13672 |
| Observations[d] | 24,423 | | | | | 36,925 | | | | |

Abbreviations: RR=risk ratio; AME=average marginal effect; NA=not applicable. RRs and AMEs are shown with 95 CIs which are adjusted for clustering at the levels of the primary and secondary sampling unit.

[a] Included only one of the variables shown in the table and a binary indicator for each country (country-level fixed effects).

[b] Included age, sex, education, place of residence (urban sample only) and a binary indicator for each country (country-level fixed effects).

[c] All regressions are age adjusted with age as continuous variable with restricted cubic splines with five knots (placed at the fifth, 27.5th, 50th, 72.5th, and 95th percentiles).

[d] Number of observations in covariate-adjusted regression model.

Table S53: Poisson regressions for dyslipidemia onto individual-level characteristics (with survey weights), non-urban sample: Poverty headcount ratio <$1.90/day to poverty headcount ratio <$5.50/day [c]

| | <$1.90/day | | | | | <$3.20/day | | | | | <$5.50/day | | | | |
|---|---|---|---|---|---|---|---|---|---|---|---|---|---|---|---|
| **Variable** | **RR (95% CI)** | ***P* value** | **AME (95% CI)** | ***P* value** | **Obs.** | **RR (95% CI)** | ***P* value** | **AME (95% CI)** | ***P* value** | **Obs.** | **RR (95% CI)** | ***P* value** | **AME (95% CI)** | ***P* value** | **Obs.** |
| **Countries** | **28** | | | | | **32** | | | | | **32** | | | | |
| **Covariate-unadjusted regressions**[a] | | | | | | | | | | | | | | | |
| Sex | | | | | | | | | | | | | | | |
| Female | 1 (ref.) | | 1 (ref.) | | 10265 | 1 (ref.) | | 1 (ref.) | | 18499 | 1 (ref.) | | 1 (ref.) | | 26940 |
| Male | 0.75 (0.39-1.42) | 0.374 | 0.00 (-0.02-0.00) | 0.348 | 6494 | 0.53 (0.34-0.83) | 0.005 | -0.02 (-0.03--0.01) | 0.004 | 12804 | 0.53 (0.40-0.71) | 0.000 | -0.03 (-0.04--0.01) | 0.000 | 19239 |
| Education | | | | | | | | | | | | | | | |
| No schooling | 1 (ref.) | | 1 (ref.) | | 5097 | 1 (ref.) | | 1 (ref.) | | 8676 | 1 (ref.) | | 1 (ref.) | | 11019 |
| Primary education | 1.55 (0.82-2.93) | 0.174 | 0.01 (0.00-0.02) | 0.178 | 8464 | 1.58 (1.00-2.49) | 0.048 | 0.01 (0.00-0.02) | 0.046 | 15286 | 1.66 (1.17-2.35) | 0.005 | 0.02 (0.00-0.03) | 0.002 | 21170 |
| Secondary education or further | 1.12 (0.54-2.35) | 0.758 | 0.00 (-0.01-0.01) | 0.763 | 3165 | 1.72 (1.19-2.49) | 0.004 | 0.01 (0.01-0.03) | 0.010 | 7264 | 1.67 (1.09-2.55) | 0.019 | 0.02 (0.00-0.04) | 0.041 | 13876 |
| **Covariate-adjusted regressions**[b] | | | | | | | | | | | | | | | |
| Sex | | | | | | | | | | | | | | | |
| Female | 1 (ref.) | | 1 (ref.) | | 10247 | 1 (ref.) | | 1 (ref.) | | 18451 | 1 (ref.) | | 1 (ref.) | | 26870 |
| Male | 0.69 (0.35-1.36) | 0.288 | 0.00 (-0.02-0.00) | 0.264 | 6479 | 0.50 (0.32-0.79) | 0.003 | -0.02 (-0.03--0.01) | 0.002 | 12775 | 0.51 (0.39-0.68) | 0.000 | -0.03 (-0.04--0.01) | 0.000 | 19195 |
| Education | | | | | | | | | | | | | | | |
| No schooling | 1 (ref.) | | 1 (ref.) | | 5097 | 1 (ref.) | | 1 (ref.) | | 8676 | 1 (ref.) | | 1 (ref.) | | 11019 |
| Primary education | 1.68 (0.86-3.27) | 0.127 | 0.01 (0.00-0.02) | 0.130 | 8464 | 1.77 (1.08-2.89) | 0.023 | 0.01 (0.01-0.03) | 0.022 | 15286 | 1.76 (1.22-2.54) | 0.002 | 0.02 (0.01-0.03) | 0.001 | 21170 |
| Secondary education or further | 1.29 (0.63-2.67) | 0.489 | 0.00 (0.00-0.01) | 0.507 | 3165 | 1.93 (1.28-2.92) | 0.002 | 0.02 (0.01-0.03) | 0.004 | 7264 | 1.83 (1.15-2.91) | 0.011 | 0.03 (0.00-0.05) | 0.033 | 13876 |

| | | | |
|---|---|---|---|
| Observations[d] | 16,726 | 31,226 | 46,065 |

Abbreviations: RR=risk ratio; AME=average marginal effect; NA=not applicable. RRs and AMEs are shown with 95 CIs which are adjusted for clustering at the levels of the primary and secondary sampling unit.

[a] Included only one of the variables shown in the table and a binary indicator for each country (country-level fixed effects).

[b] Included age, sex, education, place of residence (urban sample only) and a binary indicator for each country (country-level fixed effects).

[c] All regressions are age adjusted with age as continuous variable with restricted cubic splines with five knots (placed at the fifth, 27.5$^{th}$, 50$^{th}$, 72.5$^{th}$, and 95$^{th}$ percentiles).

[d] Number of observations in covariate-adjusted regression model.

Table S54: Poisson regressions for dyslipidemia onto individual-level characteristics (with survey weights), non-urban sample: Poverty headcount ratio >$5.50/day to all poverty levels[c]

| | **>$5.50/day** | | | | | **Total** | | | | |
|---|---|---|---|---|---|---|---|---|---|---|
| **Variable** | **RR (95% CI)** | ***P* value** | **AME (95% CI)** | ***P* value** | **Obs.** | **RR (95% CI)** | ***P* value** | **AME (95% CI)** | ***P* value** | **Obs.** |
| **Countries** | **32** | | | | | **32** | | | | |
| **Covariate-unadjusted regressions[a]** | | | | | | | | | | |
| Sex | | | | | | | | | | |
| Female | 1 (ref.) | | 1 (ref.) | | 13694 | 1 (ref.) | | 1 (ref.) | | 40634 |
| Male | 0.87 (0.64-1.18) | 0.378 | -0.02 (-0.06-0.04) | 0.369 | 10718 | 0.74 (0.57-0.95) | 0.017 | -0.03 (-0.04-0.01) | 0.017 | 29957 |
| Education | | | | | | | | | | |
| No schooling | 1 (ref.) | | 1 (ref.) | | 1970 | 1 (ref.) | | 1 (ref.) | | 12989 |
| Primary education | 1.17 (0.73-1.90) | 0.510 | 0.02 (-0.05-0.07) | 0.486 | 7305 | 1.46 (1.08-1.98) | 0.014 | 0.03 (0-0.03) | 0.007 | 28475 |
| Secondary education or further | 1.40 (0.90-2.16) | 0.136 | 0.05 (-0.04-0.08) | 0.076 | 15106 | 1.72 (1.23-2.39) | 0.001 | 0.04 (0-0.05) | 0.000 | 28982 |
| **Covariate-adjusted regressions[b]** | | | | | | | | | | |
| Sex | | | | | | | | | | |
| Female | 1 (ref.) | | 1 (ref.) | | 13675 | 1 (ref.) | | 1 (ref.) | | 40545 |
| Male | 0.85 (0.64-1.14) | 0.276 | -0.03 (-0.06-0.04) | 0.266 | 10706 | 0.72 (0.56-0.92) | 0.010 | -0.03 (-0.04-0) | 0.010 | 29901 |
| Education | | | | | | | | | | |
| No schooling | 1 (ref.) | | 1 (ref.) | | 1970 | 1 (ref.) | | 1 (ref.) | | 12989 |
| Primary education | 1.19 (0.73-1.96) | 0.482 | 0.02 (-0.05-0.08) | 0.453 | 7305 | 1.5 (1.09-2.07) | 0.012 | 0.03 (0-0.04) | 0.006 | 28475 |

| | | | | | | | | | | |
|---|---|---|---|---|---|---|---|---|---|---|
| Secondary education or further | 1.44 (0.92-2.26) | 0.111 | 0.06 (-0.03-0.08) | 0.054 | 15106 | 1.81 (1.28-2.55) | 0.001 | 0.04 (0.01-0.05) | 0.000 | 28982 |
| Observations[d] | 24,381 | | | | | 70,446 | | | | |

Abbreviations: RR=risk ratio; AME=average marginal effect; NA=not applicable. RRs and AMEs are shown with 95 CIs which are adjusted for clustering at the levels of the primary and secondary sampling unit.

[a] Included only one of the variables shown in the table and a binary indicator for each country (country-level fixed effects).

[b] Included age, sex, education, place of residence (urban sample only) and a binary indicator for each country (country-level fixed effects).

[c] All regressions are age adjusted with age as continuous variable with restricted cubic splines with five knots (placed at the fifth, 27.5[th], 50[th], 72.5[th], and 95[th] percentiles).

[d] Number of observations in covariate-adjusted regression model.
